# Causal Mediation Analysis with Multiple Causally Ordered and Non-ordered Mediators based on Summarized Genetic Data

**DOI:** 10.1101/2021.01.07.21249415

**Authors:** Lei Hou, Yuanyuan Yu, Xiaoru Sun, Xinhui Liu, Yifan Yu, Ran Yan, Hongkai Li, Fuzhong Xue

**Author notes:** Corresponding author: Fuzhong Xue, Address: School of public health, Shandong University, 44 Wenhua West Road, Jinan, Shandong province, China, Hongkai Li, Address: School of public health, Shandong University, 44 Wenhua West Road, Jinan, Shandong province, China. Fuzhong Xue and Hongkai Li contributed equally to this work. **Conflicts of Interest** None declared. **Availability of data and materials** GWAS summary data for BMI, Lipids and CVD are publicly available at https://portals.broadinstitute.org/collaboration/giant/index.php/GIANT_consortium_data_files, http://lipidgenetics.org/ and http://www.cardiogramplusc4d.org/, respectively. Code to implement the method and reproduce all simulations and analyses is available on Github (https://github.com/hhoulei/PSEMR). **Ethics approval and consent to participate** Ethical approval was not sought, because this study involved analysis of publicly available summary-level data from GWASs, and no individual-level data were used. **Authors’ contributions** HL and FX conceived the study. LH, HL contributed to theoretical derivation with assistance from YY, XL and XS. LH, RY and YY contributed to the data simulation. LH, HL and SS contributed to the application. LH and HL wrote the manuscript with input from all other authors. All authors reviewed and approved the final manuscript.

## Abstract

Causal mediation analysis aims to investigate the mechanism linking an exposure and an outcome. Dealing with the impact of unobserved confounders among the exposure, mediator and outcome has always been an issue of great concern. Moreover, when multiple mediators exist, this causal pathway intertwines with other causal pathways, making it more difficult to estimate of path-specific effects (PSEs). In this article, we propose a method (PSE-MR) to identify and estimate PSEs of an exposure on an outcome through multiple causally ordered and non-ordered mediators using Mendelian Randomization, when there are unmeasured confounders among the exposure, mediators and outcome. Additionally, PSE-MR can be used when pleiotropy exists, and can be implemented using only summarized genetic data. We also conducted simulations to evaluate the finite sample performances of our proposed estimators in different scenarios. The results show that the causal estimates of PSEs are almost unbiased with good coverage and Type I error properties. We illustrate the utility of our method through a study of exploring the mediation effects of lipids in the causal pathways from body mass index to cardiovascular disease.

**Author summary:** A new method (PSE-MR) is proposed to identify and estimate PSEs of an exposure on an outcome through multiple causally ordered and non-ordered mediators using summarized genetic data, when there are unmeasured confounders among the exposure, mediators and outcome. Lipids play important roles in the causal pathways from body mass index to cardiovascular disease

## 1 Introduction

Mediation analyses help to uncover the mechanisms underlying causal relationships between an exposure and an outcome by using mediator variables [1]. In mediation analyses, the total effect of an exposure on an outcome is partitioned into indirect and direct effects. Indirect effects act through mediators of interest, whereas direct effects are determined by fixing the mediator at a specified level. Estimating direct and indirect effects via existing methods typically requires a stringent sequential ignorability assumption [2] that no unmeasured confounders exist among the exposure, mediators and outcome [3]. However, this assumption may not hold in practice and omitting important confounders will necessarily bias results [4]. When multiple intermediate variables (*M*_1_ and *M*_2_) are involved in a study, three types of mediators with respect to *M*_1_ and *M*_2_ may arise, as shown in Figure 1. In Figure 1A, *M*_1_ is conditionally independent of *M*_2_ given the treatment (*X*) and measured covariates [5]. In Figure 1B, *M*_1_ and *M*_2_ are not causally ordered because they are independent of each other, conditional upon the treatment (*X*) and measured covariates [6]. In Figure 1C, mediators are causally ordered, and *M*_1_ is treated as a mediator-outcome confounder affected by the treatment. If we are interested in the mediator *M*_2_, we get a two-way decomposition into an indirect effect through *M*_2_ and a direct effect (not through *M*_2_). Imai and Yamamoto [7] proposed an approach for all the three types of mediators under a linear structural equation model. Daniel et al. [8] considered the finest possible decomposition of the total effect when there are two causally ordered mediators, and evaluated each path-specific effect (PSE) under the counterfactual framework. Additionally, VanderWeele and Vansteelandt [9] regarded the multiple mediators simultaneously as joint mediators, and defined the “joint” natural direct and indirect effects as extensions of the usual two-way decomposition of the total effect using regression-based approach and weighting approach. Several methods [10-16] have been developed to relax the sequential ignorability assumption. However, none of them allowed for the simultaneous existence of unmeasured confounders among the exposure, mediators and the outcome.

**Figure 1.**
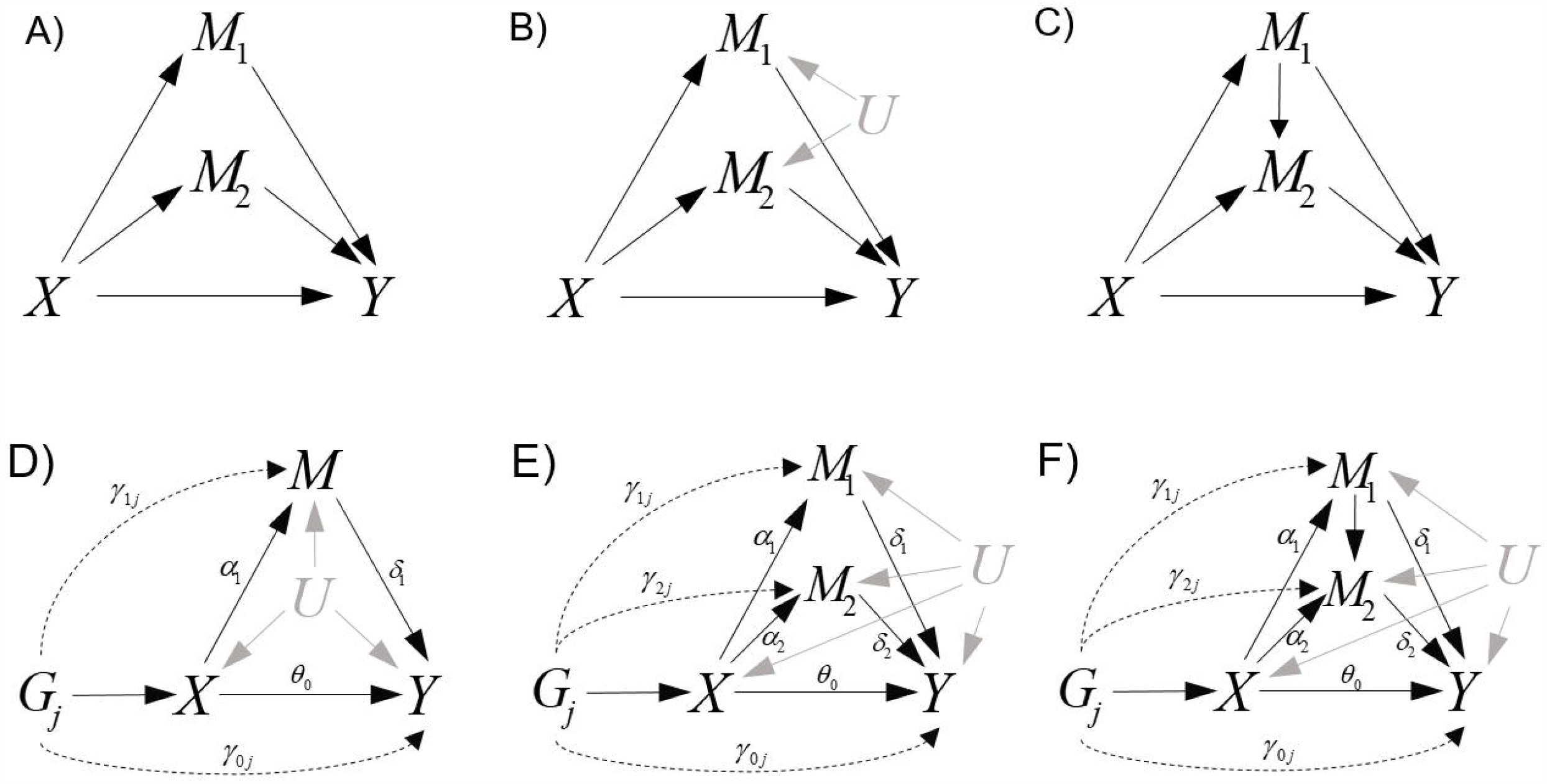
Three types of settings with two mediators, *M*_1_ and *M*_2_ are shown in (A) where *M*_1_ is independent of *M*_2_; (B) where *M*_1_ is related to *M*_2_, but not causally; and (C) where *M*_1_ is causally related to *M*_2_ (causally-ordered mediators). Graphical diagrams for PSE-MR are given in settings with one mediator (D), two non-ordered mediators (E), and two ordered mediators (F). *X*: the exposure, *M*_1_ and *M*_2_: two mediators, *Y*: outcome, *G*: instrumental variables (genetic variants).

Mendelian randomization (MR) analyses [17] using summarized data have recently become popular due to the increase in public availability of suitable data in large sample sizes from recently published genome-wide association studies [18]. For instance, Tikkanen E et al. (2019) performed a two-sample MR to evaluate independent causal roles of body components (fat-free mass and fat mass) on atrial fibrillation (AF) [19]. Firstly, univariate MR was used to estimate the causal effect of fat-free mass on AF by leveraging genetic variants (instrumental variables). Some genetic variants may be associated with both fat-free mass and fat mass, which is problematic because fat mass is also associated with AF. These genetic variants are invalid because they violate the assumption of exclusion restriction, since – they unlock the pathway from genetic variants to AF not via fat-free mass. This phenomenon is called horizontal pleiotropy, and fat mass is considered a pleiotropic trait [20]. In order to eliminate the effect of pleiotropy on causal estimation, multivariable MR [21] was performed to evaluate the causal role of fat-free mass on AF independent of fat mass. Similarly, we can obtain the causal effect of fat mass on AF independent of fat-free mass.

Risk factors associated with genetic variants may not always be pleiotropic traits, rather they may be mediators in the causal pathway from the exposure to the outcome (Figure 1D). In this case, these genetic variants are still valid instruments and MR can be used for mediation analysis. Burgess S et al. (2017) showed that total and direct effects in a single mediator setting can be estimated by univariate and multivariable MR analyses, respectively [22]. We will review this in Section 2.1. In Section 2.2, we extend the analysis from a single mediator setting to a multiple mediators setting (PSE-MR) for both causally ordered and non-ordered mediators. Then in Section 3, we apply our method to estimate PSEs from body mass index (BMI) to cardiovascular disease (CVD) through lipids mediators. In Section 4, we conduct simulations to compare the performance of PSE-MR in different scenarios. Finally, we discuss the methods and results of this study and its potential for application. R package *PSEMR* for implementing PSE-MR is provided in Github (https://github.com/hhoulei/PSEMR).

## 2 Methods

Throughout, we let *X, Y, M* and *G* denote the exposure, outcome, mediator and genetic variant, respectively. *U* denotes a set of baseline covariates and potential confounders of the mediators, exposure and outcome relationships. We also let θ_0_, α_1_ and δ_1_ denote the effect of *X* on *Y, X* on *M* and *M* on *Y*, respectively. The subscript *j* (*j =* 1,…, *J*), denotes the *j*-th genetic variant. Increasingly, MR analyses are implemented using summarized data on the associations of each genetic variant with the exposure, mediator and outcome, obtained from linear regressions on non-overlapping data consortia. This included the beta-coefficients 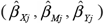 and their standard errors 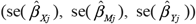. If the exposure *X* or the outcome *Y* is binary, then these summarized association estimates may be replaced with association estimates (log(OR)) obtained from logistic regression.

Initially, we consider the indirect (through *M*) and direct (not through the above mediators) effects of an exposure *X* on an outcome *Y* using genetic variants *G*. Then we declare several assumptions. We assume all genetic variants are uncorrelated (not in linkage disequilibrium). We also assume all variables are continuous, and relationships between variables (the genetic associations with the exposure *X*, mediator *M*, and outcome *Y*, and the causal effects of *X* and *M* on *Y* as well as *X* on the *M*) are linear with homogeneity across the population. In other words, interactions between the exposure (*X*) and mediator (*M*) are not allowed unless individual data is available. We also assume that the consistency and composition assumptions in causal mediation analyses hold [24] (see S1 Appendix, Section 1). Note that we relax the assumption of no unmeasured confounders among the exposure *X*, mediator *M*, and outcome *Y*, which is required in most studies.

### 2.1 PSE-MR in one mediator setting

In a single mediator setting (Figure 1D), a valid instrumental variable *G*_*j*_ must satisfy the following three assumptions:

**Assumption I**. For each *j*(*j* = 1,…, *J*), the instrumental variable *G*_*j*_ is associated with the exposure *X*.

This assumption requires that *G*_*j*_ should be strongly associated with *X*, otherwise, weak instrumental variable bias will exist [25]. The “rule of thumb” advocates that the *F* statistic of each instrumental variable should be at least 10 to avoid this bias [26-27] (see S1 Appendix, Section 2.3).

**Assumption II**. For each *j*(*j* = 1,…, *J*), *G*_*j*_ ⊥ *U*, and these three unmeasured confounders satisfy the following criteria:

1. There is no additive *X* −*U* interaction on *M* and *Y*.
2. There is no additive *M* −*U* interaction on *Y*.
3. There is no confounders of *M*-*Y* relationship induced by *X*..

In this assumption, we posit that there is no confounders of *M*-*Y* relationship induced by *X*, nor any interactions between *X* (or *M*) and these confounders [17]. When the interactions between *M* and *U* exist, the direct effect of *X* on *Y* can be identified (see S1 Appendix, Section 4). Swanson S and VanderWeele T [28] suggested that the E-value can be used to examine the independence between *G*_*j*_ and *U*, that is, to evaluate the sensitivity of estimates to confounders between *G*_*j*_ and *Y* (see S1 Appendix, Section 5).

**Assumption III**. For each *j*(*j* = 1,…, *J*), *G*_*j*_ ⊥ *Y* | (*X, U*), *G*_*j*_ ⊥ *M* | (*X, U*).

This assumption means that there is no pleiotropy. In other words, *G*_*j*_ must affect *Y* through *X*, and the pathways *G*_*j*_ → *M* → *Y* or *G*_*j*_ → *Y* (not via *X*) are not allowed. We examine and relax this assumption in Section 2.1.2.

#### 2.1.1 PSE-MR based on IVW (PSE-IVW)

For each *j*(*j* = 1,…, *J*), we do not allow for direct effects between *G*_*j*_ and *M* (*γ*_1 *j*_ =0) as well as *G*_*j*_ and *Y* (*γ* _0 *j*_ =0) (Figure 1D). Based on above three assumptions, the inverse-variance weighting method (IVW) can provide an estimate of the total effect *θ*_*T*_ of *X* on *Y* by the following weighted regression with the intercept set to zero

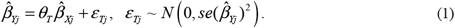

The total effect *θ*_*T*_ between *X* and *Y* can be decomposed into a direct effect (*θ*_*T*_ =*θ*_*I*_ *+θ*_*D*_ = *α*_1_ ×*δ*_1_ *+θ*_0_) and an indirect effect via *M*.

Under the framework of multivariable MR, the weighted regression model can be expanded by including genetic associations with the mediator

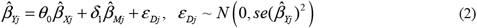

where 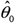 provides an estimate of the direct effect *θ*_*D*_. The indirect effect *θ*_*I*_ of exposure on the outcome can be calculated as *θ*_*ID*_ =*θ*_*T*_ −*θ*_*D*_ (difference indirect effect). It is equivalent to *θ*_*IP*_ =*α*_1_ *×δ*_1_ (product indirect effect), where *δ*_1_ can be estimated by equation (2) and *α*_*1*_, can be estimated by the following weighted regression with the intercept set to zero

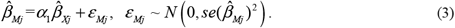

The standard error of the difference and product indirect effects are presented in S1 Appendix, Section 5. The total effect can also be estimated from individual-level data using the two-stage least squares (2SLS) method. The direct effect can also be estimated using 2SLS by regressing the outcome on fitted values of the exposure, and further on fitted values of the mediator [22].

#### 2.1.2 PSE-MR of a single mediator based on MR-Egger (PSE-Egger)

The method proposed by Burgess et al. (2017) has some limitations. This method cannot be used if Assumption III is violated, that is, direct effects of *G*_*j*_ on *M* (*M* simultaneously plays the role of a pleiotropic trait) or *G*_*j*_ on *Y* (pleiotropic pathway) exist. Thus, we relax the Assumption III by allowing for direct effects between *G*_*j*_ and *M* (*γ*_1 *j*_ ≠ 0) as well as *G*_*j*_ and *Y* (*γ* _0 *j*_ ≠ 0) (Figure 1D). Without the limitation of intercept set to zero, the causal effect of *X* on *Y* can be obtained by MR-Egger regression. To satisfy the InSIDE assumption [23] for MR-Egger, we require

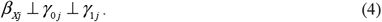

The total effect *θ*_*T*_ can be estimated by the following weighted linear regression

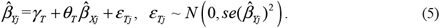

*θ*_*T*_ can also be decomposed into the direct effect *θ*_*D*_ = *θ*_0_ and the product indirect effect *θ*_*IP*_, where *θ*_*D*_ can be obtained by multivariable MR-Egger regression:

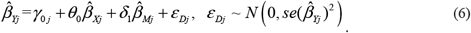

The intercept term *γ*_0 *j*_ that differs from zero is an indicator of direct effect between *G*_*j*_ and *Y*, which is called directional pleiotropy. For product indirect effect *θ*_*IP*_, *δ*_1_ can be estimated by above equation (6), and *α*_1_ can also be obtained by the following multivariable MR-Egger regression:

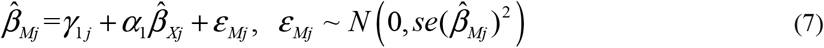

Where *γ*_1 *j*_ that differs from zero is an indicator of direct effect between *G*_*j*_ and *M*. The estimation of standard error for difference and product indirect effect is presented in the S1 Appendix.

### 2.2 Extending PSE-MR to multiple mediators setting

In this section, we extend the PSE-MR method to a multiple mediators setting. If there are *n* mediators *M*_1_, *M*_2_,…, *M*_*n*_ in the causal pathway from *X* to *Y*, PSEs can be identified. In the multiple mediators setting, we consider two relationships among mediators: causally non-ordered and causally ordered, respectively. In both cases, a valid instrumental variable must satisfy Assumption I mentioned in Section 2.1, and the following Assumption II^*^ and III^*^, which extend from the Assumption II and III.

**Assumption II**^**\***^. For each *i, j*(*i* =1,…, *n, j* = 1,…, *J*), *G*_*j*_ ⊥ *U*.

1. There is no additive *X* −*U* interaction on *M*_*i*_ and *Y*.
2. There is no additive *M*_*i*_ −*U* interaction on *Y*.
3. There is no confounders of *M*_*i*_ − *Y* relationship induced by *X*.

**Assumption III**^**\***^. For each *i, j*(*i* =1,…, *n, j* = 1,…, *J*), *G*_*j*_ ⊥ *Y* | (*X, U*), *G*_*j*_ ⊥ *M*_*i*_ | (*X, U*).

The illustrations and examinations for Assumptions I and III can also be extended to the multiple mediators setting.

#### 2.2.1 PSE-MR for causally non-ordered mediators

Firstly, we consider causally non-ordered mediators (Figure 2A, B), where *n* mediators are independent of each other, conditional on *X*. Total effect *θ*_*T*_ can also be estimated by equation (1). The direct effect (*θ*_*D*_ =*θ*_0_) and product indirect effect 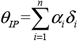 can be estimated by the following weighted regressions with the intercept set to zero:

**Figure 2.**
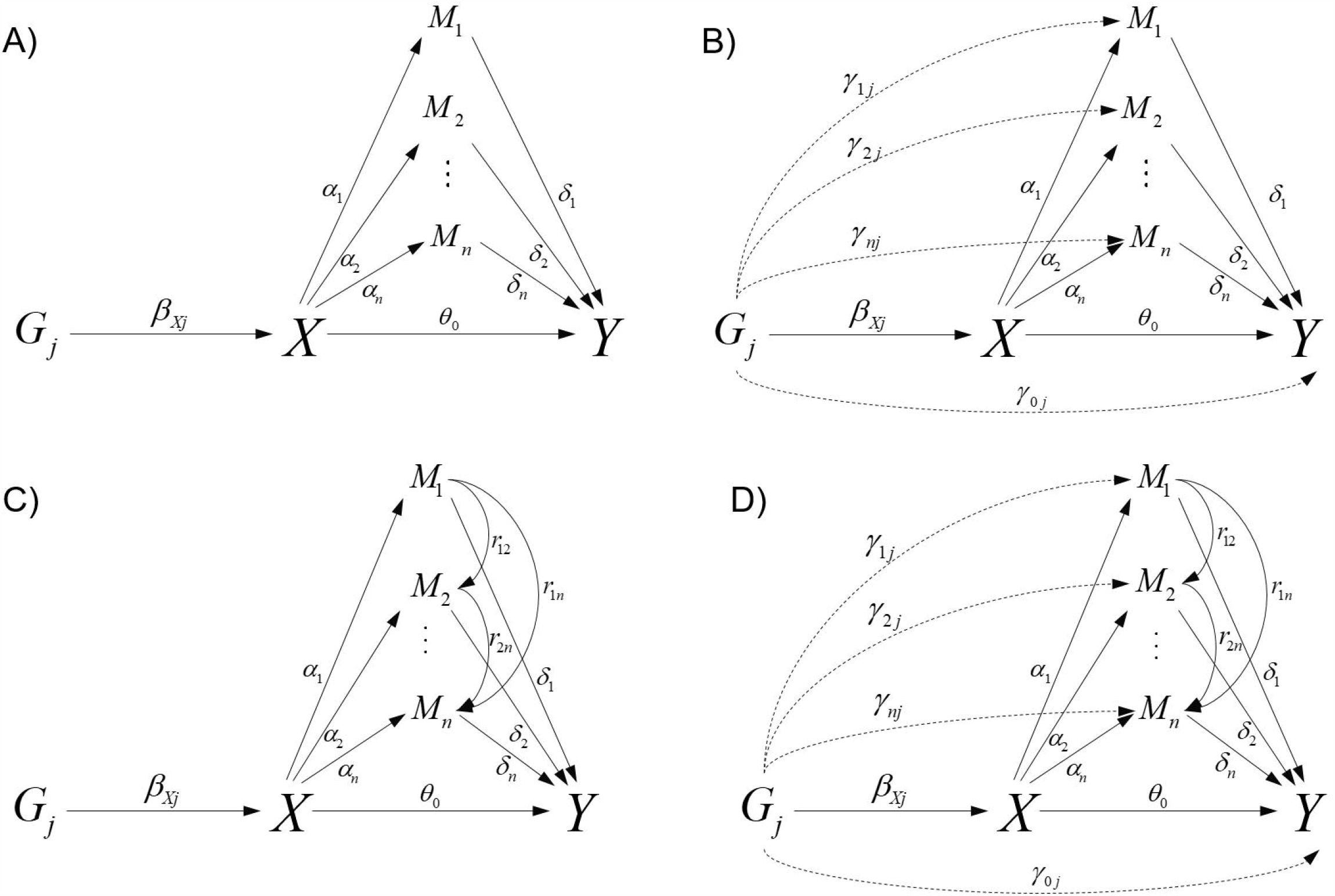
Graphical diagrams of relationships between the exposure (*X*), causally non-ordered mediators (*M*_1_, …, *M*_n_), outcome (*Y*), and instrumental variables (*G*), which omits the confounders among *X, M* and *Y*, are shown as analyzed with (A) PSE-IVW and (B) PSE-Egger. Graphical diagrams of relationships between exposure (*X*), causally ordered mediators (*M*_1_, …, *M*_n_), outcome (*Y*), and instrumental variables (*G*), which omits the confounders (*U*) among *X, M*_1_, …, *M*_n_ and *Y* are shown, as analyzed with (C) PSE-IVW and PSE-Egger.

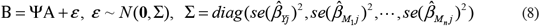

where 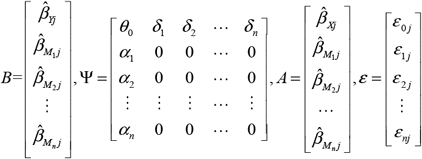. These estimations can also be obtained from individual-level data using 2SLS method.

Similarly, we relax Assumption III^*^ by allowing for the direct effect between the instrumental variable *G*_*j*_ and mediators *M*_*i*_ (*γ*_1 *j*_, *γ* _2 *j*_, …, *γ* _*nj*_), as well as *G*_*j*_ and *Y* (*γ*_0 *j*_ ≠ 0) (Figure 2B). Under the InSIDE assumption *β*_*Xj*_ ⊥ *γ*_1 *j*_ ⊥ *γ* _2 *j*_ ⊥ … ⊥ *γ* _*nj*_ ⊥ *γ* _0 *j*_, the total effect *θ*_*T*_ can also be estimated by equation (5). The direct effect (*θ*_*D*_ =*θ*_0_) and product indirect effect (*θ*_*IP*_) can also be estimated by the following linear regression equations:

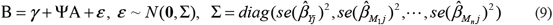

where

*γ* = [*γ*_0 *j*_ *γ*_1 *j*_ *γ*_2 *j*_ *γ*_*n j*_]^*T*^. Intercept terms *γ*_*0j*_and *γ*_*ij*_ (*i* = 1,…, *n*) that differ from zero are indicators of direct effect between *G*_*j*_ and *Y*, as well as *G*_*j*_ and *M*_*i*_, respectively. Detailed theoretical derivations are presented in S1 Appendix, section 3.

#### 2.2.2 PSE-MR for causally ordered mediators

When all the mediators are causally ordered (Figure 2C, D), we let *r*_*pq*_ denote the direct effect of *M* _*p*_ on *M*_*q*_, *p, q* ∈(1, 2,…, *n*), *p* ≠ *q*. The total effect *θ*_*T*_ can also be estimated by equation (5). The direct effect (*θ*_*D*_ =*θ*_0_) and product indirect effect

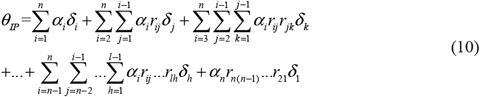

can be estimated by the weighted regressions in equation (8) and (9) by substitutingΨ * for Ψ, where

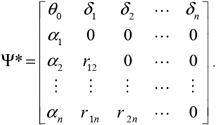

The causal effect *r*_*pq*_ from *M* _*p*_ to *M*_*q*_, *p, q* ∈(1, 2,…, *n*), *p* ≠ *q* can be identified. Details of theoretical derivation are presented in S1 Appendix, section 3. In practice, we can use Mendelian randomization to justify the causal direction of any two mediators. Then we combine the results of causal relationships of any two mediators to obtain the ordering of multiple mediators.

## 3 Application

We attempted to reveal the causal mechanism from body mass index (BMI) to cardiovascular disease (CVD) as an illustrative example. CVD, which includes coronary heart disease, stroke and heart failure, is the leading cause of death worldwide [29]. High BMI is an important risk factor of CVD [30]. Furthermore, dyslipidaemia in obesity is characterized by increased levels of very low density lipoprotein (VLDL) cholesterol, triacylglycerols (TG) and total cholesterol (TC), and lower high density lipoprotein (HDL) cholesterol levels levels [31].

Previous studies suggested that a variety of alterations in cardiac structure and function occur in the individual as adipose tissue accumulates excessively [32]. However, Van Gaal LF et al. found little evidence that LDL cholesterol is enhanced in obesity [31]. Hence, we aim to examine whether BMI affects CVD through its influence on HDL and TG.

Genetic associations with BMI in 694,649 participants from European were obtained from the Genetic Investigation of ANthropometric Traits (GIANT) [33]. Genetic associations with TG and HDL in 188,577 participants were obtained from the Global Lipids Genetics Consortium (GLGC) [34]. Genetic associations with CVD risk in 22,233 cases and 64,762 controls of European descent were obtained from the CARDIoGRAMplusC4D Consortium [35]. We identified 285 single-nucleotide polymorphisms (SNPs) associated with BMI as a genetic instrument with *F* statistics greater than 10 (explaining 2.89% of exposure variance), by extracting the effect sizes for SNP associated with BMI (*P* ≤ 5 ×10^−8^) from summary statistics. As the extracted SNPs for BMI might be correlated with each other, we pruned the variants by linkage disequilibrium (LD) (*r* ^2^ < 0.01, clumping window = 10000 kbp). Then we tested whether these SNPs violate the exclusion restriction assumption. Firstly we plotted funnel plot (Figure 3) and found three SNPs were outliers. After removing them, the funnel plots were more symmetric. The Egger test revealed no significant effects of the mediators, HDL (*P* = 0.204), TG (*P* = 0.349) and the outcome CVD (*P* = 0.071). These results indicate the absence of directional pleiotropy. Details of the SNPs are listed in ***S1 Appendix***.

**Figure 3.**
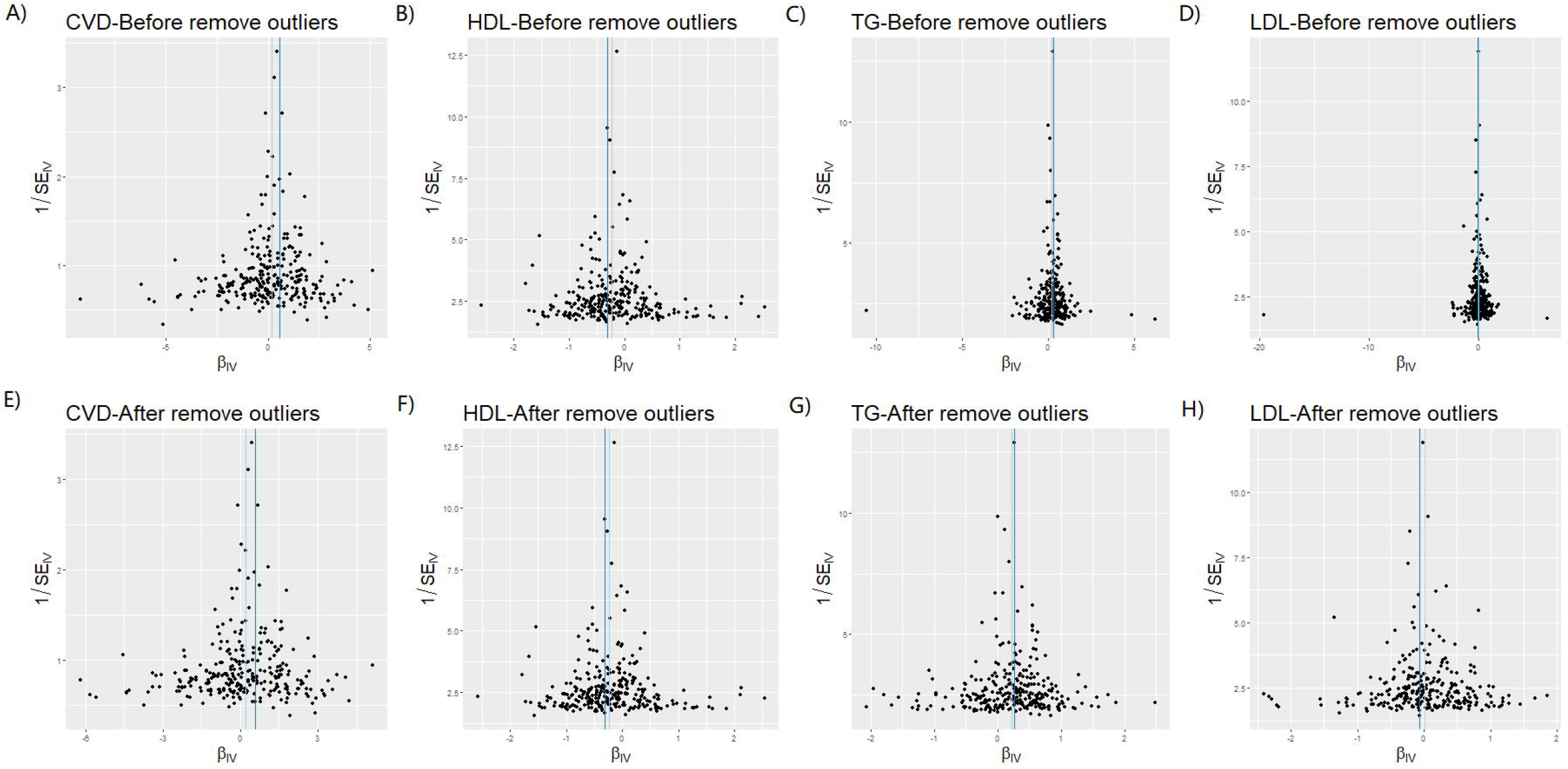
Funnel plots before (A-D) and after (E-H) removing outliers.

Firstly, we performed a single mediator analysis for the mediators (HDL and TG) via PSE-MR. Table 1 suggests TG and HDL are mediators in the causal pathway from BMI to CVD. Then we performed PSE-MR analysis with multiple mediators to test whether BMI has indirect effects on CVD risk through HDL and TG. Although a higher BMI increase the risk of CVD, no significant direct effect was obtained after adjusting for genetic associations with TG and HDL. Indirect effects through TG and HDL explained a large proportion of causal effect from BMI to CVD, and their total mediation proportion (MP) is 93.44%. In conclusion, three pathways exist from BMI to CVD: BMI→HDL→CVD (MP: 27.1% [17.1, 38.2]), BMI→ TG→CVD (MP: 24.9% [16.3, 34.7]) and BMI→TG→HDL→CVD (MP: 23.7% [2.5, 49.3]). These results (Figure 4) are consistent with results from a pooled analysis of 97 prospective cohorts with 1.8 million participants [37] and previously described biological mechanisms [36, 38].

**Table 1.** Causal effect of each pathway between BMI and CVD

|  | IVW |  | MR-Egger |  |
| --- | --- | --- | --- | --- |
|  | OR[95% CI] | Power | OR[95% CI] | Power |
| Total effect | 1.216[1.034,1.431] | 1 | 1.772[1.208,2.600] | 1 |
| Direct effect | 1.013[0.850,1.208] | - | 1.391[0.945,2.048] | - |
| BMI→HDL→CVD | 1.054[1.034,1.101] | 0.97 | 1.097[1.040,1.125] | 1 |
| BMI→TG→CVD | 1.050[1.033,1.096] | 0.97 | 1.068[1.027,1.137] | 1 |
| BMI→TG→HDL→CVD | 1.047[1.005,1.101] | 0.7 | 1.052[1.000,1.125] | 0.8 |

BMI: body mass index; CVD: cardiovascular disease; TG: triacylglycerols; HDL: high density lipoprotein; LDL: low density lipoprotein.

**Figure 4.**
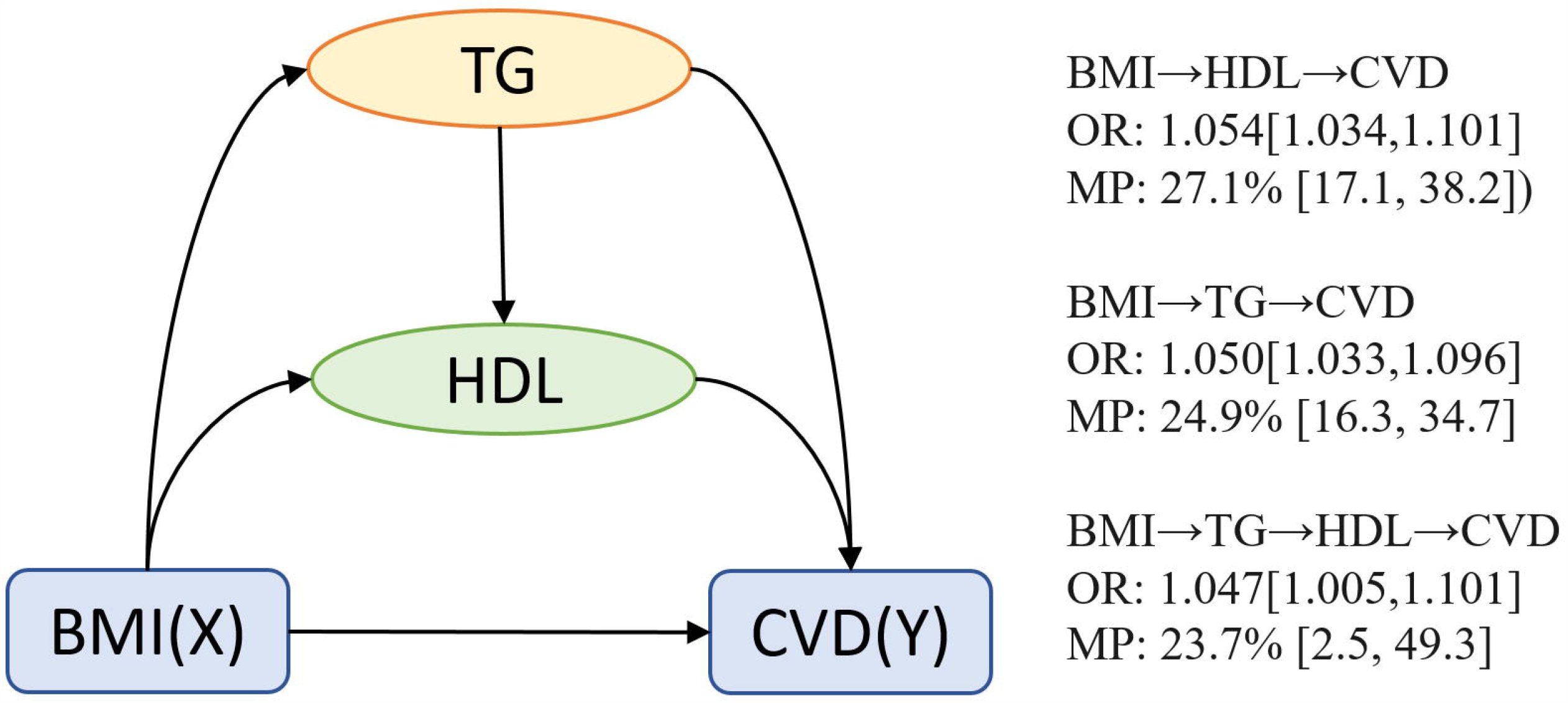
Diagrams of the causal pathway from BMI to CVD. BMI, body mass index; CVD, cardiovascular disease; TG, triacylglycerol; HDL, high-density lipoprotein.

## 4 Simulation

### 4.1 Settings

To validate the utility of the PSE-MR method for estimating PSEs, we designed six scenarios: when Assumption III is satisfied (PSE-IVW) or violated (PSE-Egger) for settings with one mediator (simulations A, B), multiple causally non-ordered (simulation C, D) and multiple causally ordered mediators (simulation E, F).

We generated data on 25 genetic variants, an exposure (*X*), mediators (*M*), and outcome (*Y*) for 20,000 individuals. Briefly, we specified different values of the parameters *θ*_*D*_ (the direct effect of *X* on *Y*) and *θ*_*I*_ (the indirect effect of *X* on *M*) to observe performances of our methods. According to the specification of *θ*_*D*_ and *θ*_*I*_, simulations from A to F included four settings: no direct effect, no indirect effect, a direct effect along with a directionally concordant indirect effect, and a direct effect and a directionally discordant indirect effect. For PSE-Egger, the data were simulated to consider the following three cases:

**Case (a)**: Balanced pleiotropy, InSIDE assumption satisfied;

**Case (b)**: Directional pleiotropy, InSIDE assumption satisfied;

**Case (c)**: Directional pleiotropy, InSIDE assumption not satisfied.

We also performed additional simulations for sensitivity analyses, where bidirectional causal effects between the exposure and mediators, population homogeneity assumption is violated, the causal order is misspecified and one of the mediators is missing. In addition, we also consider the performance of PSE-MR when the exposure and outcome are time varying. We also find the optimal number of genetic variants when we consider multiple mediators. Details of the simulation are presented in S2 Appendix.

We used the following metrics to evaluate performance of our methods: mean bias, standard errors (SE), mean square error (MSE), type I error rate for a null causal effect and empirical power to detect a non-null effect (i.e., the proportion of confidence intervals excluding zero).

### 4.2 Results

We varied the sample size, the number of instrumental variables, and simulated four scenarios for different sets of parameter values. We found that causal estimates of direct and indirect effects were unbiased with good Type I error properties. As the sample size increased, bias and standard errors decreased, while power improved. Higher power and lower bias were observed as the number of instrumental variables increased (see S2 Appendix, Section 1, 3 and 5).

For two non-ordered mediators, PSE-IVW showed good performance of in standard MR when estimating the total, direct and indirect effects as well as three PSEs (Table 2). As the sample size and the number of genetic variants increased, the bias was smaller and the type I error was more stable at approximately 0.05 (see S2 Appendix, Section 3). The performance of PSE-MR based on IVW and MR-Egger with two non-ordered mediators in Case (a) and (b), are listed in eTables 9 to12 (see S2 Appendix, section 4). In Case (a), we observed that the bias was close to zero and Type I error rates was around 0.05 in PSE-MR. PSE-Egger had less bias and more stable Type I error rates than IVW when directional pleiotropy existed in at least one pathway from *G* to *Y* (Case (b)). MR-Egger performed better than IVW in term of bias, even when the InSIDE assumption was not satisfied (Case (c)). When the pleiotropic effects through confounders (violating the InSIDE assumption) were 2.5 times larger than the direct pleiotropic effects (satisfying InSIDE), estimates from PSE-Egger were much less biased and rejection rates of the causal null hypothesis were much closer to the nominal 5% rate than those from PSE-IVW were. In all cases, PSE-Egger had smaller MSE and more stable Type I error rates (0.05) than PSE-IVW when the PSE was zero. Estimators of indirect effects based on product method had more stable Type I error rates (0.05) than those based on the difference method. Results for the two ordered multiple mediators were similar to those of two non-ordered mediators (Table 3 and eTables 17-24 in S2 Appendix, section 6). In addition, the magnitude of *r*_*qp*_ does not influence the performances of PSE-MR. Details are presented in eTable 15 (see S2 Appendix, section 5).

**Table 2.** Simulation of PSE-IVW with two non-ordered mediators in standard MR

| True |  |  |  |  | Estimates |  |  |  |  |  | MSE |  |  |  |  |  | Power/Type I error |  |  |  |  |  |
| --- | --- | --- | --- | --- | --- | --- | --- | --- | --- | --- | --- | --- | --- | --- | --- | --- | --- | --- | --- | --- | --- | --- |
| TE | DE | IE | IE1 | IE2 | TE | DE | IE_d | IE_p | IE1 | IE2 | TE | DE | IE_d | IE_p | IE1 | IE2 | TE | DE | IE_d | IE_p | IE1 | IE2 |
| 1.6 | 0.8 | 0.8 | 0.5 | 0.3 | 1.61 | 0.75 | 0.85 | 0.85 | 0.53 | 0.32 | 0.00 | 0.00 | 0.01 | 0.01 | 0.00 | 0.00 | 1.00 | 1.00 | 1.00 | 1.00 | 1.00 | 1.00 |
| 0.6 | 0.8 | -0.2 | -0.5 | 0.3 | 0.60 | 0.75 | -0.15 | -0.15 | -0.47 | 0.32 | 0.00 | 0.00 | 0.01 | 0.01 | 0.00 | 0.00 | 1.00 | 1.00 | 0.26 | 0.29 | 1.00 | 1.00 |
| 0.7 | 0.5 | 0.2 | 0.5 | -0.3 | 0.70 | 0.45 | 0.25 | 0.25 | 0.54 | -0.29 | 0.00 | 0.00 | 0.01 | 0.01 | 0.00 | 0.00 | 1.00 | 1.00 | 0.74 | 0.75 | 1.00 | 1.00 |
| -0.3 | 0.5 | -0.8 | -0.5 | -0.3 | -0.30 | 0.46 | -0.76 | -0.76 | -0.47 | -0.28 | 0.00 | 0.00 | 0.01 | 0.01 | 0.00 | 0.00 | 1.00 | 1.00 | 1.00 | 1.00 | 1.00 | 1.00 |
| 0 | -0.8 | 0.8 | 0.5 | 0.3 | 0.01 | -0.84 | 0.85 | 0.85 | 0.53 | 0.32 | 0.01 | 0.00 | 0.01 | 0.01 | 0.00 | 0.00 | 0.07 | 1.00 | 1.00 | 1.00 | 1.00 | 1.00 |
| -1.6 | -0.8 | -0.8 | -0.5 | -0.3 | -1.60 | -0.85 | -0.76 | -0.76 | -0.47 | -0.29 | 0.00 | 0.00 | 0.01 | 0.01 | 0.00 | 0.00 | 1.00 | 1.00 | 1.00 | 1.00 | 1.00 | 1.00 |
| 0.8 | 0 | 0.8 | 0.5 | 0.3 | 0.81 | -0.04 | 0.85 | 0.85 | 0.53 | 0.32 | 0.00 | 0.00 | 0.01 | 0.01 | 0.00 | 0.00 | 1.00 | 0.00 | 1.00 | 1.00 | 1.00 | 1.00 |
| 1.3 | 0.8 | 0.5 | 0.5 | 0 | 1.31 | 0.75 | 0.55 | 0.55 | 0.54 | 0.02 | 0.00 | 0.00 | 0.01 | 0.01 | 0.00 | 0.00 | 1.00 | 1.00 | 1.00 | 1.00 | 1.00 | 0.00 |
| 1.3 | 0.8 | 0.5 | 0.5 | 0 | 1.31 | 0.77 | 0.54 | 0.54 | 0.54 | 0.00 | 0.00 | 0.00 | 0.01 | 0.01 | 0.00 | 0.00 | 1.00 | 1.00 | 1.00 | 1.00 | 1.00 | 0.07 |
| 0.8 | 0.8 | 0 | 0 | 0 | 0.80 | 0.75 | 0.05 | 0.05 | 0.03 | 0.02 | 0.00 | 0.00 | 0.00 | 0.00 | 0.00 | 0.00 | 1.00 | 1.00 | 0.01 | 0.02 | 0.01 | 0.01 |

TE: total effect; DE: direct effect; IE: indirect effect; IE1: $X \rightarrow M_1 \rightarrow Y$ ; IE2: $X \rightarrow M_2 \rightarrow Y$ ; IE\_d: indirect effect calculated by difference method; IE\_p: indirect effect calculated by product method.

**Table 3.** Simulation of PSE-IVW with two ordered mediators in standard MR

| True |  |  |  |  |  | Estimates |  |  |  |  |  |  | MSE |  |  |  |  |  |  | Power/Type I error |  |  |  |  |  |  |
| --- | --- | --- | --- | --- | --- | --- | --- | --- | --- | --- | --- | --- | --- | --- | --- | --- | --- | --- | --- | --- | --- | --- | --- | --- | --- | --- |
| TE | DE | IE | IE1 | IE2 | IE3 | TE | DE | IE_d | IE_p | IE1 | IE2 | IE3 | TE | DE | IE_d | IE_p | IE1 | IE2 | IE3 | TE | DE | IE_d | IE_p | IE1 | IE2 | IE3 |
| 1.8 | 0.8 | 1 | 0.5 | 0.3 | 0.2 | 1.81 | 0.75 | 1.06 | 1.06 | 0.52 | 0.28 | 0.25 | 0.01 | 0.00 | 0.01 | 0.01 | 0.00 | 0.02 | 0.02 | 1.00 | 1.00 | 1.00 | 1.00 | 0.97 | 0.64 | 0.62 |
| 0.8 | 0.8 | 0 | -0.5 | 0.3 | 0.2 | 0.81 | 0.75 | 0.05 | 0.05 | -0.48 | 0.29 | 0.24 | 0.00 | 0.00 | 0.01 | 0.01 | 0.00 | 0.02 | 0.02 | 1.00 | 1.00 | 0.01 | 0.00 | 1.00 | 0.70 | 0.60 |
| 1.2 | 0.8 | 0.4 | 0.5 | -0.3 | 0.2 | 1.20 | 0.75 | 0.45 | 0.45 | 0.54 | -0.26 | 0.16 | 0.01 | 0.00 | 0.01 | 0.01 | 0.01 | 0.01 | 0.01 | 1.00 | 1.00 | 0.98 | 0.72 | 1.00 | 0.69 | 0.41 |
| 0.2 | 0.8 | -0.6 | -0.5 | -0.3 | 0.2 | 0.20 | 0.76 | -0.56 | -0.56 | -0.46 | -0.25 | 0.16 | 0.00 | 0.00 | 0.01 | 0.01 | 0.00 | 0.01 | 0.01 | 0.97 | 1.00 | 1.00 | 1.00 | 1.00 | 0.68 | 0.38 |
| 0.2 | -0.8 | 1 | 0.5 | 0.3 | 0.2 | 0.21 | -0.84 | 1.05 | 1.06 | 0.52 | 0.29 | 0.25 | 0.01 | 0.00 | 0.01 | 0.01 | 0.00 | 0.02 | 0.02 | 0.71 | 1.00 | 1.00 | 1.00 | 1.00 | 0.67 | 0.62 |
| -1.8 | -0.8 | -1 | -0.5 | -0.3 | -0.2 | -1.80 | -0.85 | -0.96 | -0.96 | -0.48 | -0.25 | -0.23 | 0.01 | 0.00 | 0.01 | 0.01 | 0.00 | 0.02 | 0.01 | 1.00 | 1.00 | 1.00 | 1.00 | 1.00 | 0.66 | 0.64 |
| 1 | 0 | 1 | 0.5 | 0.3 | 0.2 | 1.01 | -0.05 | 1.05 | 1.05 | 0.52 | 0.29 | 0.25 | 0.01 | 0.00 | 0.01 | 0.01 | 0.00 | 0.02 | 0.02 | 1.00 | 0.00 | 1.00 | 1.00 | 0.99 | 0.67 | 0.61 |
| 1.3 | 0.8 | 0.5 | 0 | 0.3 | 0.2 | 1.31 | 0.75 | 0.56 | 0.56 | 0.02 | 0.29 | 0.25 | 0.00 | 0.00 | 0.01 | 0.01 | 0.00 | 0.02 | 0.02 | 1.00 | 1.00 | 1.00 | 0.89 | 0.00 | 0.67 | 0.62 |
| 1.5 | 0.8 | 0.7 | 0.5 | 0 | 0.2 | 1.51 | 0.77 | 0.74 | 0.74 | 0.52 | -0.03 | 0.25 | 0.01 | 0.00 | 0.01 | 0.01 | 0.00 | 0.02 | 0.02 | 1.00 | 1.00 | 1.00 | 0.97 | 0.99 | 0.07 | 0.61 |
| 1 | 0.8 | 0.2 | 0 | 0 | 0.2 | 1.01 | 0.77 | 0.24 | 0.24 | 0.02 | -0.03 | 0.25 | 0.00 | 0.00 | 0.01 | 0.01 | 0.00 | 0.02 | 0.02 | 1.00 | 1.00 | 0.41 | 0.01 | 0.00 | 0.08 | 0.61 |
| 0 | -1 | 1 | 0.5 | 0.3 | 0.2 | 0.01 | -1.05 | 1.06 | 1.06 | 0.52 | 0.29 | 0.25 | 0.01 | 0.00 | 0.01 | 0.01 | 0.00 | 0.02 | 0.02 | 0.05 | 1.00 | 1.00 | 1.00 | 1.00 | 0.69 | 0.62 |

TE: total effect; DE: direct effect; IE: indirect effect; IE1: $X \rightarrow M_1 \rightarrow Y$ ; IE2: $X \rightarrow M_2 \rightarrow Y$ ; IE3: $X \rightarrow M_1 \rightarrow M_2 \rightarrow Y$ ; IE\_d: indirect effect calculated by difference method; IE\_p: indirect effect calculated by product method.

The estimation of direct effect is unbiased regardless of whether bidirectional causal effects between exposure and mediators exist, or the causal order is misspecified, though the estimation of PSEs is biased. Heterogeneous populations sometimes introduce bias of causal estimation for non-ordered and ordered mediators. Note that if we are missing upstream mediators (e.g. *M*_1_), *M*_1_ is the confounder of *M*_2_ and Y and it is affected by *X* (i.e. *X*–induced unmeasured confounder of *M*_2_ and *Y*). Thus the assumption of cross-world independence is violated. In addition, if we can obtain the information in each time points, PSE-MR can be applied into time varying exposure and mediators and it can also deal with the bi-directional relationship between exposure and mediators (see S1 Appendix, section 7-13). Performance of PSE-MR with different number of SNPs and mediators are listed in the eTable 41-42 and eFigure 9-10.

## 5 Discussion

In this paper, we develop a method PSE-MR to identify and estimate PSEs from an exposure on an outcome through the mediator(s) using MR when there are unmeasured confounders among the exposure, mediators and the outcome. We extend PSE-MR from a single mediator setting to the multiple mediator setting for both causally ordered and non-ordered mediators, and outline the assumptions required to obtain causal effect. PSE-IVW can be used to explore the role of multiple mediators in the causal pathways between the exposure and outcome. The PSE-Egger can be viewed as a sensitivity analysis to provide robustness against both measured and unmeasured pleiotropy and to strengthen the evidence from the PSE-IVW analysis.

PSE-MR can estimate the direct effects between the exposure and outcome and indirect effects through mediators when the sequential ignorability assumption [39] in mediation analyses is relaxed. We compared the assumptions of PSE-MR with traditional mediation analysis methods in Table 4. Our method requires other independent assumptions. While Assumptions I and III are testable, there is no accepted method to test for the Assumption II. Several sensitivity analyses can be performed to examine this assumption, such as the E-value [28] and heterogeneity test. The validity of multiple mediators PSE-Egger and its ability to estimate consistent causal effects rely on the InSIDE assumption [21] being satisfied. When the direct genetic associations with the exposure are independent of the direct genetic associations with mediators and outcome, the InSIDE assumption is satisfied. Whereas the InSIDE assumption is plausible in some cases, it sometimes will not always be valid. For example, heterogeneous populations and misspecification of the multiple mediators would bias the mediation effect estimation. When *γ* _*kj*_ is not independent from each other or *γ* _0 *j*_ is not independent with *γ* _*kj*_ for *k* = 1,…,*n* (e.g. we are missing one of multiple mediators), the direct effect is downward-biased and the indirect effect is upward-biased.

**Table 4.** Comparison of the assumption in PSE-MR and typical causal mediation analysis

| Methods | PSE-MR | Typical causal mediation analysis | Solution for violating assumptions |
| --- | --- | --- | --- |
| Common | (1) Consistency assumption: $M(x)=M$ and $Y(x)=Y$ if $X=x$; $Y(x,m)=Y$ if $X=x, M=m$. | | |
| | (2) Composition assumption: $Y(x)=Y(x,M(x))$ if $X=x$. | | |
| | (3) A cross-world independence assumption: $Y(x,m) \perp M(x^*) \mid C$ for all $(x, x^*, m)$. | | [10] [44] [45] [51] |
| | (4) There is no additive interaction of the exposure and the mediators on the outcome ($Y$). | | [10] [46] [48] [49] |
|  | (5) Linearity. |  | [10] [47] |
| Different | (1) The IV $G_j$ is associated with the exposure $X$ but independent of all the unmeasured confounders. | (1) No-unmeasured confounders of the $X$-$Y$ relation, that is, $Y(x,m) \perp X \mid C$ for all $(x,m)$. | [10] [15] |
| | (2) The Exclusion restriction assumption or the InSIDE assumption. | (2) No-unmeasured confounders of the $M$-$Y$ relation, that is, $Y(x,m) \perp M(x) \mid X=x, C$ for all $(x,m)$. | [10] [11] [12] [13] [14] [15][16] |
| | (3) There is no additive interaction of the exposure or the mediators and confounders on the mediator ($M$) and the outcome ($Y$). | (3) No-unmeasured confounders of the $X$-$M$ relation, that is, $M(x) \perp X \mid C$ for all $x$. | [10] [15] |
|  | (4) Data for exposure, mediators and outcome can from different datasets with homogeneous population. | (4) Data for exposure, mediators and outcome must from the same dataset. |  |
|  |  | (5) Exposure, mediators and confounders not vary with time. | [51] [52] [53] |

According to our simulation, we find that PSE-IVW is more robust in estimating causal effect than PSE-Egger for heterogeneous populations and misspecified multiple mediators. However, PSE-Egger can be applied to test directional pleiotropy, and it can give less biased estimates when the InSIDE assumption is violated.

For the multiple causally ordered mediator settings, PSE-MR can be widely used in time-varying exposure and mediators. Labrecque and Swanson (2019) [40] suggested that if the genetic associations of the exposure and mediators were time-varying, the lifetime effect estimate could be biased if we obtained the information of the exposure and mediators only at one time point. However, if we can obtain the information of the exposure and mediators at different time points, PSE-MR can provide unbiased estimates of the lifetime effects of the exposure and mediators on the outcome and other PSEs (see S2 Appendix, section 9). Thus PSE-MR can estimate each PSEs, including the causal relationships (which may potentially be bi-directional) in a non-experimental setting.

In conclusion, we propose a method of causal mediation analysis with causally ordered and non-ordered mediators based on summarized genetic data and provides a new perspective for mediation analysis.

## Data Availability

GWAS summary data for BMI, Lipids and CVD are publicly available at https://portals.broadinstitute.org/collaboration/giant/index.php/GIANT_consortium_data_files, http://lipidgenetics.org/ and http://www.cardiogramplusc4d.org/, respectively. Code to implement the method and reproduce all simulations and analyses is available on Github (https://github.com/hhoulei/PSEMR).

## Acknowledgements

We would like to thank Editage (www.editage.com) for English language editing.

## Supplementary Digital Content

**S1 Appendix**. Supplemental methods.

**S2 Appendix**. Supplemental simulations.

