## Supplementary material for "Causal Mediation Analysis with Multiple Causally Ordered and Non-ordered Mediators based on Summarized Genetic Data": S1 Appendix

**Supplemental methods**

**Supplemental Methods**

### 1 Mediation analysis

Let *Y* denote observed outcome for an individual, *X* denote an exposure, *C* denote a set of confounders that may affect the treatment, mediators, and/or outcome, and ***M****=*(*M*_1_*, M*_2_*, …, M_n_*) denote a set of mediators that may be on the pathway from the exposure *X* to the outcome *Y*. A causal mediation analysis is defined under the potential outcome framework. Let *Y*(*x*) and *M*(*x*) denote the potential outcome and potential mediator, respectively, that would be observed if, possibly contrary to the fact, *X* was set to *x*. Likewise, let *Y*(*x,m*) denote the potential outcome that would be observed if, possibly contrary to the fact, *X* was set to *x* and *M* was set to *m*. The simple linear models are:

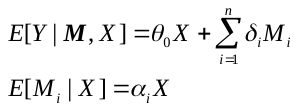

There are several assumptions which must be satisfied:

**Assumption 1 (Consistency assumption)**

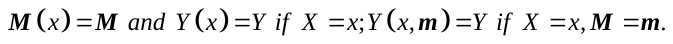

**Assumption 2 (Composition assumption)**

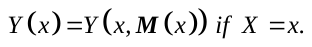

**Assumption 3 (Sequential ignorability assumption)** [1-2]

a) No-unmeasured confounders of the *X-Y* relation, that is,
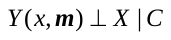
 for all (*x,****m***).
b) No-unmeasured confounders of the ***M****-Y* relation, that is,
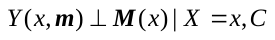
 for all (*x,****m***).
c) No-unmeasured confounders of the *X-****M*** relation, that is,
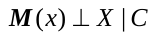
 for all *x*.

**Assumption 4 (A cross-world independence assumption)**

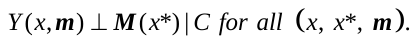

Under the above assumptions, we have the following decompositions:

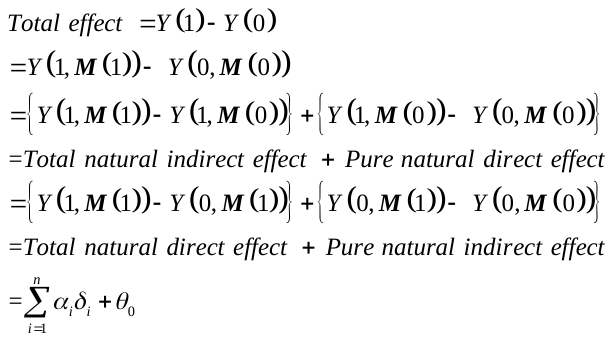

In addition, we always use summary statistics to perform MR. Using instrumental variables ***G*** which satisfy
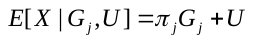
, we can obtain the association of *j*-th genetic variant with the risk factor, mediator and the outcome from above simple linear models:

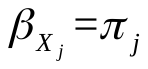

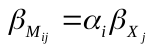

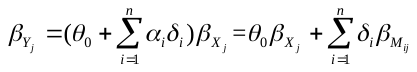
.

In PSE-MR, we mainly relax the Sequential ignorability assumption, that is, we allow the existence of unmeasured confounders (*U*) among the exposure, mediator and the outcome. Instead, we use the genetic variants as instrumental variables (*G*), which must satisfy several additional assumptions, to estimate total, direct and indirect causal effects of the exposure on the outcome. In addition, PSE-MR can be applied to estimate PSEs from the exposure to the outcome.

### 2 Mendelian randomization

Instrumental variable (IV) analysis is the exploitation of a natural experiment to obtain a causal association of an exposure on an outcome. A valid instrumental variable must satisfy the following three assumptions: (1) Relevance – IV (*G*) is robustly associated with the exposure (*X*); (2) Exchangeability – IV (*G*) is not associated with any confounder (*U*) of the exposure–outcome relationship; (3) Exclusion restriction – IV (*G*) is independent of the outcome (*Y*) conditional on the exposure (*X*) and all confounders of the exposure-outcome relationship (i.e. the only path between the instrument and the outcome is via the exposure). A genetic variant (e.g. SNP – single-nucleotide polymorphism) is a section of genetic code that differs between individuals. Genetic variants are good candidate instrumental variables: the function of many genes is known and well characterized; genetic variants are fixed at conception, and so do not change due to environmental factors, thus avoiding reverse causation; and genetic variants are generally inherited independently, meaning that they tend to be specific in their associations. The use of genetic variants as instrumental variables in observational data has been termed “Mendelian randomization (MR)”. MR uses an identification strategy that requires no assumptions about exposure-outcome confounding.

#### 2.1 Linkage disequilibrium (LD)

Linkage disequilibrium (LD) is the correlation between allelic states at different loci within the population. In Mendelian randomization studies, exchangeability is violated if the genetic variant being used as an instrument is in linkage disequilibrium with another genetic variant which is related to the confounders or outcome. This is shown in the Figure 2 (b) in the main text.

#### 2.2 Pleiotropy

Pleiotropy refers to a genetic variant having multiple functions. If a variant is associated with outcome via a pleiotropic trait (*X*_1_) other than via the modifiable exposure of interest (*X*), the exclusion restriction is violated. This is shown in the Figure 2 (c) in the main text.

#### 2.3 Weak instrumental variable bias

Angrist and Krueger (1995) [3] proved that in split-sample (two-sample) IV estimation, when the instruments (*G*) were very weakly correlated with the exposure (*X*), the causal estimator appeared to be biased towards zero.

*Proof:*

Considering the following model:

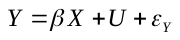

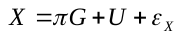

where *G*, *X* and *Y* represent instrumental variable, exposure and outcome respectively.
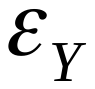
 and
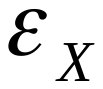
 are random variables from normal distribution with mean zero and variance
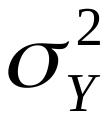
 and
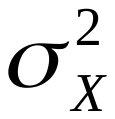
.
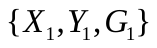
 and
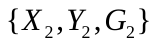
 are two datasets from two sample denoted subscript 1 and 2. The causal estimator can be written as:

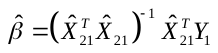

where
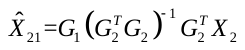
 is the cross-sample fitted value. Assuming that
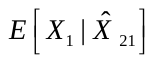
 is linear. We know that
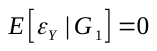
 and substitute
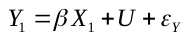
 in above estimator, then

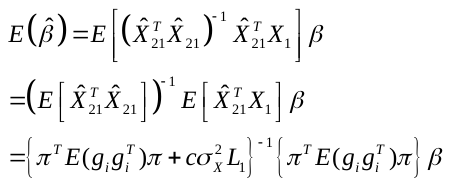

Where
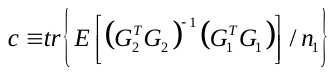
 and
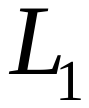
 is a constant matrix. What we are interested is when the vector of reduced-form coefficients,
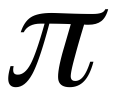
, is near 0. In this case, it is apparent from above equation that the causal estimate of
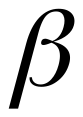
 has expectation near 0.

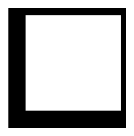

Note that

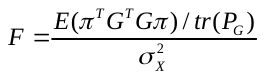

is the *F* statistic in the first stage regression. And the bias of the two-sample IV estimator to zero is

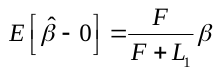
.

Suppose the coefficient
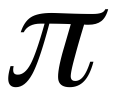
 is near 0, then F is 0. Furthermore,
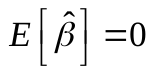
. If *F* is big enough to even infinity, then
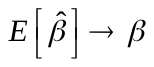
. The “rule of thumb” advocates that the *F* statistic should be at least 10 to avoid bias [4-5].

#### 2.4 Inverse-variance weighted method

Inverse-variance weighted (IVW) estimator can be calculated from summarized data when all the three assumptions are satisfied. To some extent, IVW method is a fixed-effect meta-analysis, where the IV-specific causal estimates
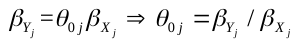
 (Wald ratio for *j*-th IV) are the study-specific estimates, and the weights are the inverse-variance weights. The causal estimate from the IVW method (
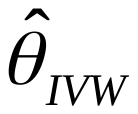
) is calculated by the following equation:

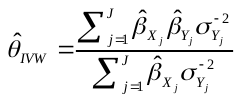

Inverse-variance weighted estimator can also be obtained by following weighted regression with the intercept set to zero

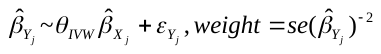

The standard error is estimated by:

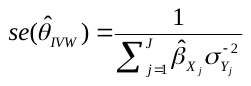

where *J* is the total number of genetic variants,
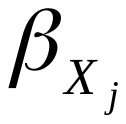
 is the effects of *j*-th genetic variant on the exposure,
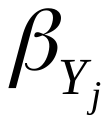
 is the *j*-th genetic variant on the outcome, and
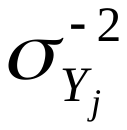
 is the inverse of standard error of
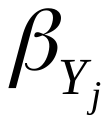
.

#### 2.5 MR-Egger method

For each IV, Wald ratio is no more the unbiased estimate of causal effect of exposure on outcome when there exists a direct effect (
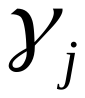
) of the *j*-th IV on the outcome, which also called pleiotropic pathway

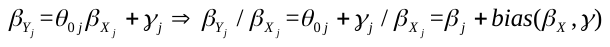
.

And in this case, the IVW estimator is also biased:

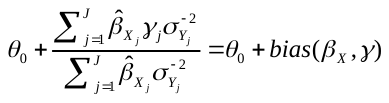
.

Thus, MR-Egger allows that the intercept is not equal to zero in the weighted regression of IVW:

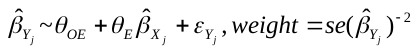
.

The slope represents the causal effect of exposure on outcome. And the intercept represents the average pleiotropic effect across the genetic variants (the average direct effect of an IV on the outcome). The estimate coefficients from Egger regression are calculated from the following equations:

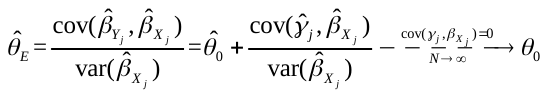
,

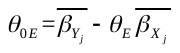
.

We really need to assume that the standard error of
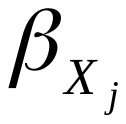
 estimated from linear regression does not decay to zero at a faster rate than the numerator of the expression involving a covariance of an estimated parameter. Note that MR-Egger method assumes that the correlation between the genetic associations with the exposure (
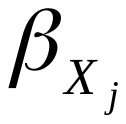
) and the direct effects of the genetic variants on the outcome (
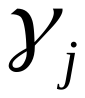
) is zero (
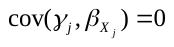
), which refers to InSIDE (Instrument Strength Independent of Direct Effect) assumption. In the limit as both the sample size and the number of genetic variants increase to infinity, the InSIDE condition ensures that
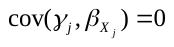
 and therefore
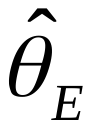
 is a consistent estimate of the causal effect
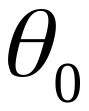
.

Overall, the IVW is used when there are no direct effects between (1) the IV and mediator (2) the IV and outcome (e.g. exclusion restriction holds). Then, MR-Egger is used when there are direct effects (e.g. exclusion restriction does not hold).

### 3 Detailed theoretical derivation for PSE-MR

#### 3.1 PSE-MR for one mediator setting

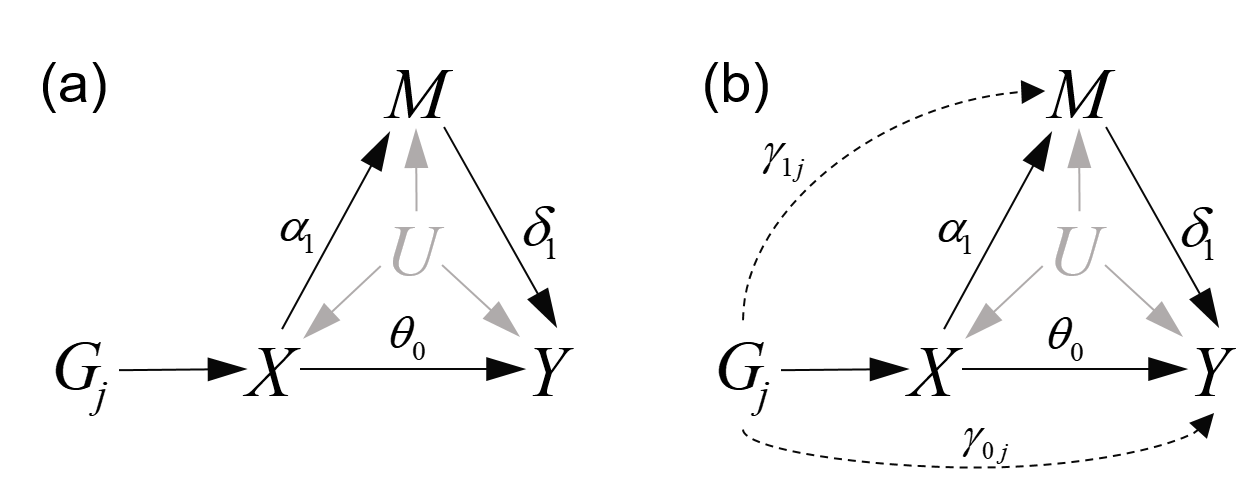

**eFigure 2.** Graphical diagram of relationships between a risk factor (*X*), a mediator (*M*), an outcome (*Y*), and instrumental variable (*G*), and omit the confounders among *X*, *M* and *Y*. (a) Single-mediator PSE-IVW; (b) Single-mediator PSE-Egger.

We add the genetic variants as instrumental variables to estimate the casual effect of the exposure on the outcome in the presence of unmeasured confounders (*U*) based on the traditional mediation analysis model (eFigure 2). We consider the following models (eFigure *2* (a)):

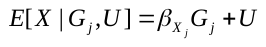

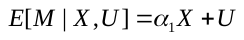

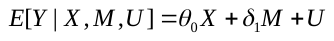
,

where *X*, *M* and *Y* denote the risk factor, mediator and outcome. *U* denotes a set of baseline covariates and potential confounders of the mediators, exposure and outcome relationships. Our aim is to estimate the total (
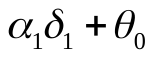
), direct (
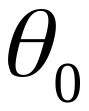
) and indirect (
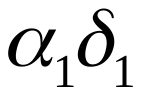
) effect.

We can obtain the association of *j*-th genetic variant with the mediator and the outcome:

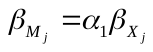

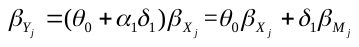
.

Based on the **Assumption Ⅰ** mentioned in the main text**,** the IVW method could provide an estimate of the total effect
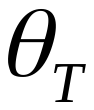
 of the risk factor *X* on the outcome *Y* by the following weighted regression with the intercept set to zero

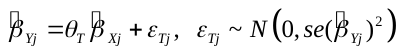
.

The total effect
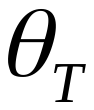
 between the risk factor *X* and the outcome *Y* can be decomposed into an indirect effect via the mediator *M* and a direct effect (
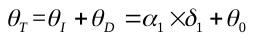
).

Under the framework of multivariable MR, the weighted regression model can be expanded by including genetic associations with the mediator

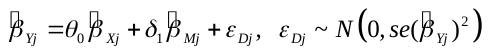

to provide an estimate of the direct effect
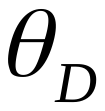
. The indirect effect
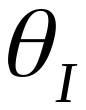
 of risk factor on the outcome can be calculated as
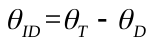
 (difference indirect effect). It is equivalent to
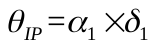
 (product indirect effect), of which
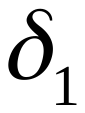
 can be estimated by equation (2) and
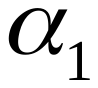
 can be estimated by the following weighted regression with the intercept set to zero

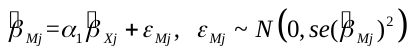
.

The standard error of the difference indirect eﬀect is the square root of the sum of the squared standard errors for the total eﬀect and the direct eﬀect

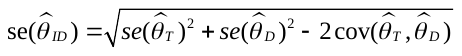
.

And the standard error of the product indirect eﬀect is

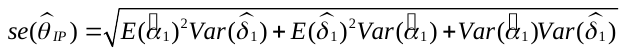
.

The method we mentioned above is the prior work by *Burgess et al. (2017)*. We call this PSE-IVW.

PSE-IVW is unavailable if there existed a direct effect of genetic variants on the mediator (this mediator simultaneously plays a role of a pleiotropic trait) or genetic variants on the outcome (pleiotropic pathway), that is, the **Assumption Ⅲ** is violated. We propose following method PSE-Egger. We relax the **Assumption Ⅲ** by allowing the direct effect between (i) instrumental variable
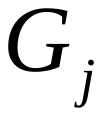
 and mediator *M* (
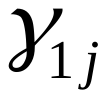
) (ii) instrumental variable
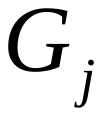
 and outcome *Y* (
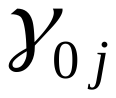
) (*Fig S2 (b)*). The association of *j*-th genetic variant with the mediator and outcome are:

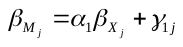

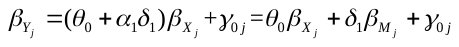
.

Without the limitation of intercept set to zero, the causal effect of the risk factor *X* on the outcome *Y* can be obtained by MR-Egger regression. To satisfy the InSIDE assumption for MR-Egger, we require

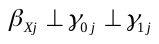

The total effect
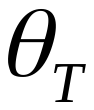
 can be estimated by the following linear regression

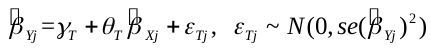

with

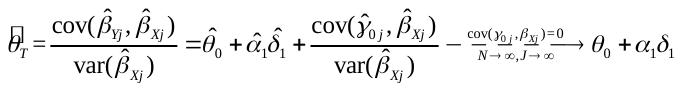
.

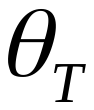
 can also be decomposed into the direct effect
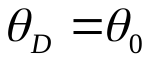
 and indirect effect
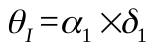
via the mediator *M*, where direct effect
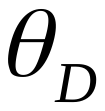
 can be obtained by multivariable MR-Egger regression:

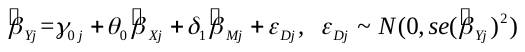

with

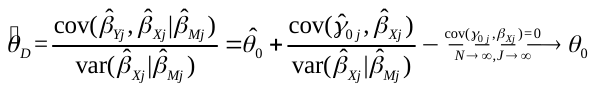
.

The intercept term
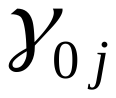
 that differs from zero is indicative of direct effect between instrumental variable
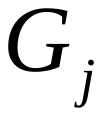
 and outcome *Y*, which is called directional pleiotropy. For indirect effect,
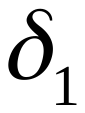
 can be estimated by above equation (9)：

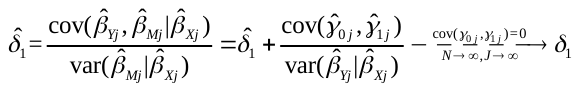
.

And can also be obtained by the following multivariable MR-Egger regression:

with

where that differs from zero is indicative of direct effect between instrumental variable and mediator *M*. The estimation of standard error for difference and product indirect effect is the same as equation (4) and (5). We need to assume that the standard error of  estimated from linear regression doesn’t decay to zero at a faster rate than the numerator of the expression involving a covariance of estimated parameters.

#### 3.2 PSE-MR for two mediators setting

##### 3.2.1 Causally non-ordered mediators

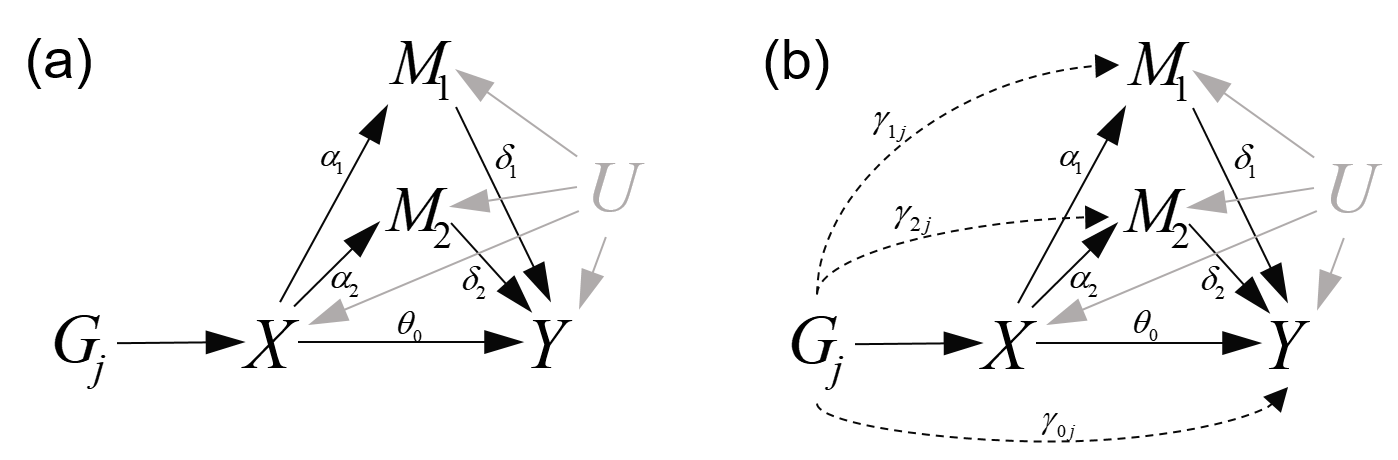

**eFigure 3.** Graphical diagram of relationships between risk factor (*X*), causally non-ordered mediators (*M_1_*, *M_2_*), outcome (*Y*), and genetic variant (*G*), and omit the confounders among *X*, *M_1_*, *M_2_* and *Y*. (a) Double-mediators PSE-IVW; (b) Double-mediators PSE-Egger

We consider the following models (*eFigure 3 (a)*):

,

where *X*, *M_1_*, *M_2_*, *Y* and denote the risk factor, mediator, outcome and unmeasured confounders. Then we obtain the association of *j*-th genetic variant with the mediators and outcome

.

If there are two independent mediators (,) in the causal pathway from risk factor *X* to outcome *Y*, both direct and indirect effect can also be identified (*eFigure 3 (a)*). In a double mediator IVW analysis, each IV must satisfy the **Assumption** **Ⅰ**and **Assumption Ⅱ^*^-Ⅲ^*^** mentioned in the main text. Similar to PSE-IVW for one mediator setting, total effect can be estimated by the following weighted regression with the intercept set to zero:

.

Direct effect () and indirect effect () can also be estimated by the following weighted regression with the intercept set to zero:

The estimation of standard error for difference indirect effect is the same as equation (4). And the estimation of standard error for difference indirect effect is

Next, we also relax the **Assumption Ⅲ^*^** by allowing the direct effect between (i) instrumental variable and mediators *M* (,..) (ii) instrumental variable and outcome *Y* () (*eFigure 3 (b)*). The association of *j*-th genetic variant with the mediators and outcome are

.

The causal effect of the risk factor *X* on the outcome *Y* can be obtained by MR-Egger regression without the limitation of intercept set to zero. To satisfy the InSIDE assumption for MR-Egger, we require

The total effect can be estimated by the following linear regression

with

It can also be decomposed into the direct effect and indirect effect via mediators and , where direct effect can be obtained by multivariable MR-Egger regression:

with

The intercept term that differs from zero is indicative of direct effect between instrumental variable and outcome *Y*, which is called directional pleiotropy. For indirect effect, and can be estimated by above equation (20). And and can also be obtained by the following multivariable MR-Egger regression:

where intercept terms and that differs from zero are indicative of direct effect between instrumental variable and mediator , instrumental variable and mediator , respectively. The estimation of standard error for difference indirect effect and difference indirect effect are the same as equation (4) and (16), respectively.

##### 3.2.2 Causally ordered mediators

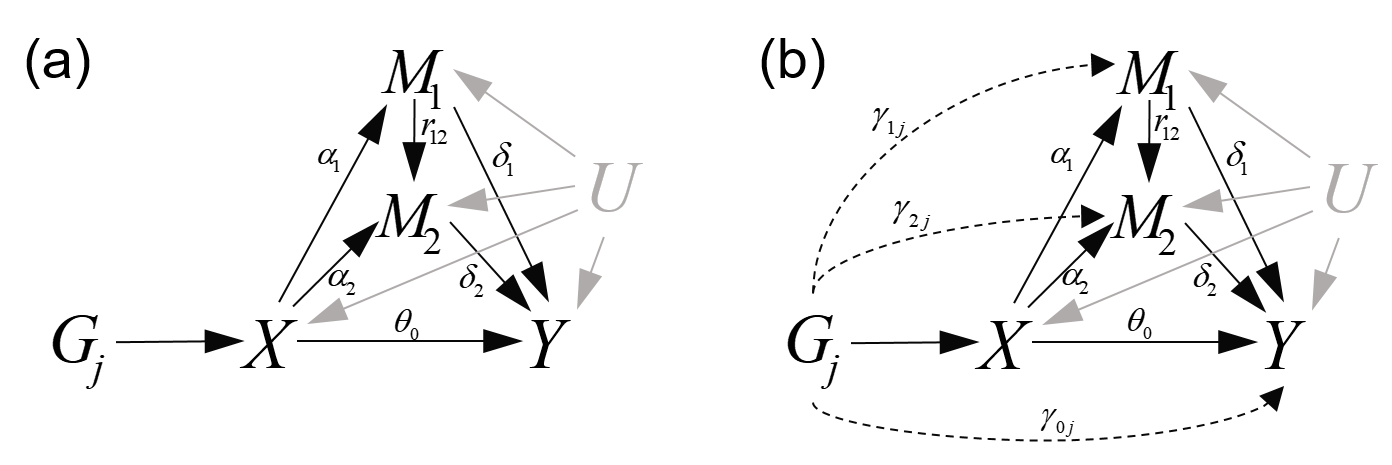

**eFigure 4.** Graphical diagram of relationships between risk factor (*X*), causally ordered mediators (*M_1_*, *M_2_*), outcome (*Y*), and genetic variant (*G*), and omit the confounders among *X*, *M_1_*, *M_2_* and *Y*. (a) Double-mediators PSE-IVW; (b) Double-mediators PSE-Egger.

We consider the following model (*eFigure 4 (a)*):

,

where *X*, *M_1_*, *M_2_*, *Y* and denote the risk factor, mediator, outcome and unmeasured confounders. Then we obtain the association of *j*-th genetic variant with mediators and outcome

If there are two correlated mediators (,) in the causal pathway from risk factor *X* to outcome *Y*, both direct and indirect effect can also be identified (*eFigure 4 (a)*). In a double mediator IVW analysis, each IV must satisfy the **Assumption** **Ⅰs**and **Ⅱ^*^-Ⅲ^*^** mentioned in the main text.

Similar to PSE-IVW for one mediator setting, total effect can be estimated by the following weighted regression with the intercept set to zero:

Direct effect () and indirect effect () can also be estimated by the following weighted regression with the intercept set to zero:

,

where indicates the causal effect of on . The estimation of standard error for difference indirect effect is the same as equation (4). And the estimation of standard error for difference indirect effect is

Next, we also relax the **Assumption Ⅲ^*^** by allowing the direct effect between (i) instrumental variable and mediators *M* (,.) (ii) instrumental variable and outcome *Y* () (*Fig S4 (b)*). The association of *j*-th genetic variant with mediators and outcome are

The causal effect of the risk factor *X* on the outcome *Y* can be obtained by MR-Egger regression without the limitation of intercept set to zero. To satisfy the InSIDE assumption for MR-Egger, we require

The total effect can be estimated by the following linear regression

with

It can also be decomposed into the direct effect and indirect effect via mediators and , where direct effect can be obtained by multivariable MR-Egger regression:

with

The intercept term that differs from zero is indicative of direct effect between instrumental variable and outcome *Y*, which is called directional pleiotropy. For indirect effect, and can be estimated by above equation (29). And , and can also be obtained by the following multivariable MR-Egger regression:

where intercept terms and that differs from zero are indicative of direct effect between instrumental variable and mediator , instrumental variable and mediator , respectively. The estimation of standard error for difference indirect effect and difference indirect effect are the same as equation (4) and (25), respectively.

#### 3.3 PSE-MR for multiple mediators setting

##### 3.3.1 Causally non-ordered mediators

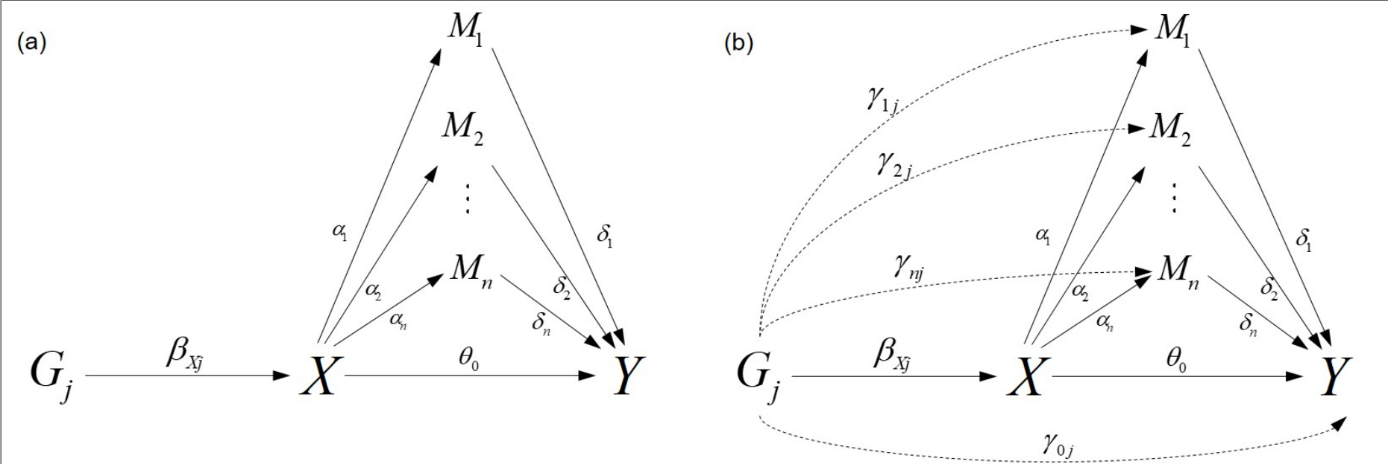

**eFigure 5.** Graphical diagram of relationships between risk factor (*X*), causally non-ordered mediators (*M*), outcome (*Y*), and genetic variant (*G*), and omit the confounders among *X*, *M* and *Y*. (a) multiple mediators PSE-IVW; (b) multiple mediators PSE-Egger. Unmeasured confounders (*U*) are omitted in the DAGs.

We consider the following model (*eFigure 5 (a)*):

……

,

where *X*, *M_1_*, …, *M_n_*, *Y* and denote the risk factor, *n* mediators, outcome and unmeasured confounders. Then we obtain the association of *j*-th genetic variant with mediators and outcome

……

,

Expanding to multi-mediator PSE-IVW (*eFigure 5 (a****)***), there are *n* causally non-ordered mediators () in the causal pathway from risk factor *X* to outcome *Y*. For all mediators (), a valid instrumental variable must satisfy the **Assumption** **Ⅰs**and **Ⅱ^*^-Ⅲ^*^** mentioned in the main text.

Total effect can be estimated by the following weighted regression with the intercept set to zero:

Direct effect () and indirect effect

can also be estimated by the following weighted regression with the intercept set to zero:

These estimation can also be obtained from individual-level data using 2SLS method.

Next, we also relax the **Assumption Ⅲ^*^** by allowing the direct effect between (i) instrumental variable and mediators *M* (,, …, ) (ii) instrumental variable and outcome *Y* () (*eFigure 5 (b)*). The association of *j*-th genetic variant with mediators and outcome are

……

,

To satisfy the InSIDE assumption for MR-Egger, we require

Total effect can be estimated by the following linear regression:

Direct effect () and indirect effect ( is the same as the multi-mediator PSE-IVW) can also be estimated by the following linear regression:

where intercept terms and that differs from zero are indicative of direct effect between instrumental variable and mediator *Y*, instrumental variable and mediators , respectively.

##### 3.3.2 Causally ordered mediators

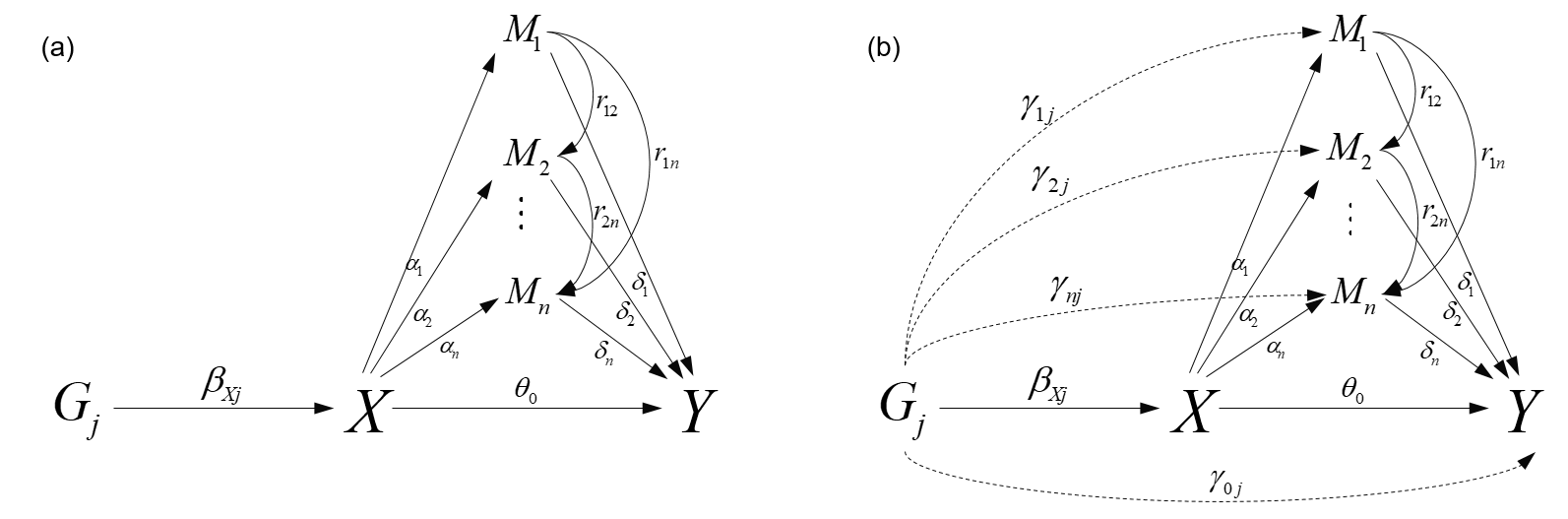

**eFigure 6.** Graphical diagram of relationships between risk factor (*X*), causally ordered mediators (*M*), outcome (*Y*), and genetic variant (*G*), and omit the confounders among *X*, *M* and *Y*. (a) multiple mediators PSE-IVW; (b) multiple mediators PSE-Egger. Unmeasured confounders (*U*) are omitted in the DAGs.

We consider the following model (*eFigure 6 (a)*):

……

,

where *X*, *M_1_*, …, *M_n_*, *Y* and denote the risk factor, *n* mediators, outcome and unmeasured confounders. Then we obtain the association of *j*-th genetic variant with mediators and outcome

……

,

where

Expanding to multi-mediator PSE-IVW (*eFigure 6 (a)*), there are *n* causally ordered mediators () in the causal pathway from risk factor *X* to outcome *Y*. For all mediators () , a valid instrumental variable must satisfy the **Assumption** **Ⅱ** and **Ⅲ^*^** mentioned in the main text.

Total effect can be estimated by the following weighted regression with the intercept set to zero:

Direct effect () and indirect effect

can also be estimated by the following weighted regression with the intercept set to zero:

The causal effect of on can be identified. These estimation can also be obtained from individual-level data using 2SLS method.To satisfy the InSIDE assumption for MR-Egger, we require

Total effect can be estimated by the following linear regression:

Direct effect () and indirect effect ( is the same as the multi-mediator PSE-IVW) can also be estimated by the following linear regression:

where intercept terms and that differs from zero are indicative of direct effect between instrumental variable and mediator *Y*, instrumental variable and mediator , respectively. The causal effect of on can be identified.

### 4 Interactions in PSE-MR

#### 4.1 Interaction between exposure and mediator(s)

When there is interaction between exposure and mediator(s), our method is unavailable for summary data but still available for individual data. Now we provide theoretical proof as follows.

Take eFigure 2(a) as an example, firstly we only consider the case of one mediator by the following model

,

where *U* is the set of ,,. is the interaction parameter of exposure and mediators. Then we obtain the association of *j*-th genetic variant with the outcome

Thus, we can divide the whole population into several subgroups according to *M* from small to large and estimate total or direct effect in different subgroups, respectively. Because the causal effect between exposure and outcome is not a constant at different values of *M*. Total and direct effects can also be obtained from equation (1) and (2) in the main text, respectively. When pleiotropy exists or in the multiple mediators setting, estimation process is similar as the case of no interaction and the only difference is that total and direct effects need to be estimated in different subgroups.

#### 4.2 Interaction between exposure and confounders

When there is interaction between exposure and confounders, our method is unavailable. Similar as the previous section, we consider the case of one mediator by the following model

,

where *U* is the set of ,,. is the interaction parameter of exposure and confounders. Then we obtain the association of *j*-th genetic variant with the mediator and outcome

The causal effect between exposure and outcome is not a constant at different values of *U*, which makes total and direct effects unspecified because of the uncertainty of the confounders *U*.

#### 4.3 Interaction between mediator(s) and confounders

When there is interaction between mediator(s) and confounders, total effect is unspecified but direct effect can be estimated. We consider the case of one mediator by the following model

.

where *U* is the set of ,,. is the interaction parameter of mediators and confounders. Then we obtain the association of *j*-th genetic variant with the mediator and outcome

Because of the uncertainty of the confounders *U*, total effect is unspecified. However, we can obtain the estimate of direct effect () from exposure to outcome by multivariable weighted regression

where .

#### 4.4 Interaction between mediators

When there is interaction between mediators, our method is unavailable for summary data but still available for individual data. We consider the case of two mediator by the following model

.

where *U* is the set of ,,. is the interaction parameter of mediators. Then we obtain the association of *j*-th genetic variant with the mediators and outcome

or

Thus, we can divide the whole population into several subgroups according to *M_1_* or *M_2_* from small to large and estimate total or direct effect in different subgroups, respectively. Because the causal effect between exposure and outcome is not a constant at different values of *M_1_* or *M_2_*. Total and direct effects can also be obtained from equation (1) and (2) in the main text, respectively. When pleiotropy exists or in the multiple mediators setting, estimation process is similar as the case of no interaction and the only difference is that total and direct effects need to be estimated in different subgroups.

#### 4.5 Exposure-induced mediator-outcome confounders (arrow from *X* to *U_MY_*)

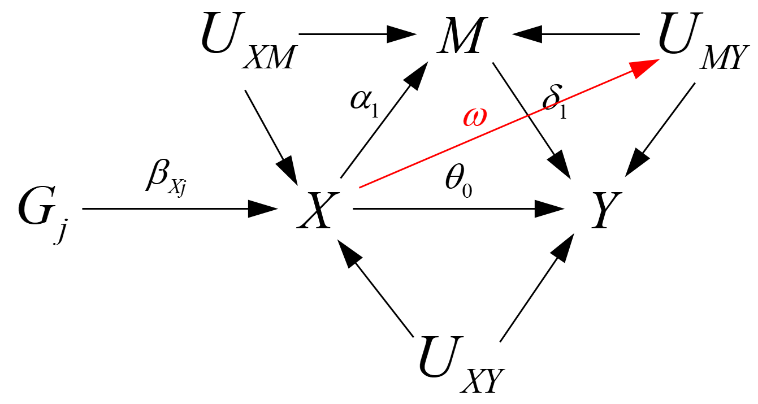

**eFigure 7.** Graphical diagram of exposure-induced mediator-outcome confounders

When there exist exposure-induced mediator-outcome confounders (arrow from *X* to *U_MY_*), our method is also unavailable. Similar as the previous section, we consider the case of one mediator by the following model

,

where *U* is the set of ,,. is the causal effect of exposure *X* on confounders *U_MY_*. Then we obtain the association of *j*-th genetic variant with the mediator and outcome

Because of the uncertainty of the confounders *U*, is unspecified. Thus total and direct effects are unspecified.

### 5 E-value

The E-value is defined as the minimum strength of association on the risk ratio scale that an unmeasured confounder would need to have with both the exposure and the outcome, conditional on the measured covariates, to completely explain away an observed exposure-outcome association. It is straightforward to calculate, and for an observed risk ratio of magnitude *RR_obs_*, it can be obtained by

If the initial risk ratio is less than 1, then it is inverted first before applying the formula. Swanson S and VanderWeele T suggested that the E-value can be used to examine the independence between genetic variants and confounders, that is, to evaluate the sensitivity of estimates to confounders between the genetic variant and the outcome. If some genetic variants are associated with confounders, then there must exist confounders between these genetic variants and outcome.

### 6 R package: *PSEMR*

We developed an R package called *PSEMR*. It can implement PSE-MR in one and multiple mediators setting with causal estimates, standard error, 95% CI and P value. In addition, *PSEMR* can calculate each PSEs and provide visual tools to demonstrate DAGs among the exposure, mediators and outcome. R package PSEMR can be download in Github (<https://github.com/hhoulei/PSEMR>).

### 7 SNPs used as IVs for BMI

| SNP | effect_allele | other_allele | beta | se | pval | samplesize |
| --- | --- | --- | --- | --- | --- | --- |
| rs10779751 | A | G | 0.0131 | 0.0018 | 2.66E-13 | 806531 |
| rs3766160 | A | G | -0.0107 | 0.0018 | 4.63E-09 | 806782 |
| rs2271928 | A | G | -0.0102 | 0.0016 | 4.44E-10 | 806702 |
| rs11577094 | T | C | 0.0186 | 0.003 | 3.28E-10 | 791496 |
| rs657452 | A | G | 0.0188 | 0.0016 | 3.17E-30 | 798331 |
| rs17114036 | A | G | -0.0158 | 0.0028 | 1.37E-08 | 805725 |
| rs2481665 | T | C | 0.0161 | 0.0016 | 3.24E-23 | 806470 |
| rs7513441 | A | G | -0.0109 | 0.0019 | 7.25E-09 | 806796 |
| rs12566985 | A | G | -0.0194 | 0.0016 | 2.80E-33 | 803962 |
| rs2815325 | T | G | 0.0136 | 0.002 | 8.28E-12 | 793230 |
| rs9787306 | T | C | -0.0155 | 0.002 | 9.85E-15 | 806775 |
| rs17391694 | T | C | 0.032 | 0.0025 | 1.59E-38 | 781664 |
| rs4970712 | A | C | 0.014 | 0.002 | 2.75E-12 | 806729 |
| rs11165468 | T | C | -0.0093 | 0.0016 | 1.12E-08 | 794997 |
| rs11165643 | T | C | 0.0185 | 0.0016 | 4.49E-30 | 805410 |
| rs17024393 | T | C | -0.0644 | 0.0049 | 7.11E-39 | 782554 |
| rs1546924 | T | C | 0.0139 | 0.0016 | 2.34E-17 | 794849 |
| rs7534091 | A | G | -0.012 | 0.0018 | 6.65E-11 | 803017 |
| rs10923724 | T | C | -0.0126 | 0.0016 | 1.14E-14 | 806787 |
| rs11577179 | A | G | -0.012 | 0.0017 | 6.85E-13 | 805734 |
| rs10733051 | A | G | 0.0093 | 0.0016 | 6.96E-09 | 794799 |
| rs12564992 | A | G | -0.0189 | 0.0026 | 2.57E-13 | 806818 |
| rs10489219 | A | G | 0.0148 | 0.0027 | 3.01E-08 | 806707 |
| rs543874 | A | G | -0.0479 | 0.002 | 3.06E-125 | 806688 |
| rs10920678 | A | G | 0.0149 | 0.0016 | 7.15E-20 | 805921 |
| rs11119208 | A | G | 0.0108 | 0.0017 | 6.94E-11 | 806749 |
| rs6661316 | T | C | 0.012 | 0.0016 | 1.72E-13 | 806610 |
| rs11118308 | A | G | 0.0098 | 0.0016 | 1.55E-09 | 806492 |
| rs4653942 | A | G | -0.0113 | 0.002 | 2.25E-08 | 806528 |
| rs6548221 | A | G | 0.0145 | 0.002 | 1.06E-13 | 806578 |
| rs2685263 | C | G | 0.0102 | 0.0016 | 2.84E-10 | 799491 |
| rs13021737 | A | G | -0.0578 | 0.0021 | 2.89E-161 | 802967 |
| rs10929925 | A | C | -0.0142 | 0.0016 | 3.05E-18 | 801209 |
| rs10182181 | A | G | -0.0327 | 0.0016 | 2.45E-91 | 806439 |
| rs12468863 | T | C | -0.0148 | 0.0016 | 6.63E-20 | 806266 |
| rs1260326 | T | C | -0.0107 | 0.0017 | 1.16E-10 | 799274 |
| rs7567655 | A | G | -0.0242 | 0.0043 | 2.19E-08 | 794748 |
| rs4372836 | T | C | 0.0125 | 0.0017 | 7.33E-13 | 806681 |
| rs17327461 | T | C | 0.0122 | 0.0016 | 7.65E-14 | 806655 |
| rs13432055 | T | C | -0.0117 | 0.0018 | 5.72E-11 | 805940 |
| rs6545714 | A | G | -0.0194 | 0.0016 | 4.01E-32 | 806783 |
| rs13417156 | T | C | -0.0135 | 0.0017 | 6.90E-16 | 767912 |
| rs11884795 | A | G | 0.0112 | 0.0018 | 1.57E-09 | 806699 |
| rs4988235 | A | G | 0.0124 | 0.0017 | 7.09E-13 | 796039 |
| rs6710871 | A | G | 0.0193 | 0.0024 | 3.07E-16 | 752446 |
| rs12692596 | T | C | 0.012 | 0.0017 | 1.03E-12 | 806739 |
| rs12692738 | T | C | -0.0131 | 0.0019 | 5.24E-12 | 806581 |
| rs10930502 | A | G | 0.0128 | 0.0018 | 2.74E-13 | 806321 |
| rs7588437 | A | G | -0.0165 | 0.0017 | 2.32E-22 | 806379 |
| rs10497870 | A | G | 0.0121 | 0.0016 | 1.97E-13 | 806733 |
| rs11692326 | T | C | 0.0147 | 0.0019 | 1.69E-14 | 801583 |
| rs16825005 | A | G | 0.0124 | 0.0018 | 3.51E-12 | 806435 |
| rs7599312 | A | G | -0.0182 | 0.0018 | 1.52E-23 | 806704 |
| rs4480977 | T | G | 0.009 | 0.0016 | 2.51E-08 | 806152 |
| rs7607369 | A | G | 0.0121 | 0.0016 | 8.20E-14 | 801568 |
| rs17535749 | A | G | 0.0159 | 0.0027 | 3.46E-09 | 793365 |
| rs10510419 | T | G | -0.0168 | 0.0023 | 2.23E-13 | 800535 |
| rs6804842 | A | G | -0.0141 | 0.0016 | 7.57E-18 | 806143 |
| rs7374289 | T | C | 0.0092 | 0.0016 | 2.29E-08 | 806622 |
| rs10460960 | A | G | 0.0216 | 0.0025 | 5.92E-18 | 805284 |
| rs6785669 | T | C | 0.0116 | 0.0017 | 1.46E-11 | 806722 |
| rs2365389 | T | C | -0.0168 | 0.0016 | 6.49E-25 | 801448 |
| rs1911746 | T | C | 0.0113 | 0.0019 | 4.17E-09 | 806670 |
| rs1452075 | T | C | 0.0128 | 0.0018 | 2.67E-12 | 794919 |
| rs925018 | C | G | -0.013 | 0.0017 | 4.33E-14 | 806677 |
| rs2371767 | C | G | 0.0107 | 0.0018 | 2.08E-09 | 787408 |
| rs12633819 | A | G | -0.0095 | 0.0017 | 2.68E-08 | 805793 |
| rs6781254 | T | C | 0.0105 | 0.0018 | 4.56E-09 | 794430 |
| rs9818122 | T | C | -0.0228 | 0.002 | 3.97E-30 | 806655 |
| rs7640424 | T | C | -0.0135 | 0.0018 | 1.23E-14 | 805792 |
| rs2124499 | C | G | -0.0115 | 0.0017 | 7.25E-12 | 802110 |
| rs7621331 | A | G | -0.0095 | 0.0017 | 3.77E-08 | 806741 |
| rs7621025 | T | C | -0.0187 | 0.0019 | 7.54E-24 | 806690 |
| rs1199334 | A | G | 0.0145 | 0.0021 | 1.50E-12 | 806644 |
| rs11924032 | A | G | 0.0151 | 0.0018 | 2.70E-16 | 806805 |
| rs6443750 | T | C | -0.0152 | 0.0021 | 7.25E-13 | 792002 |
| rs9816226 | A | T | -0.0315 | 0.0021 | 1.45E-50 | 794122 |
| rs13072095 | T | C | 0.0094 | 0.0017 | 4.38E-08 | 806761 |
| rs6850639 | T | C | 0.0112 | 0.002 | 2.84E-08 | 802057 |
| rs10938397 | A | G | -0.0322 | 0.0016 | 2.42E-86 | 805635 |
| rs6819344 | A | C | 0.0102 | 0.0016 | 5.08E-10 | 806778 |
| rs13107325 | T | C | 0.0468 | 0.0032 | 3.81E-47 | 806141 |
| rs10516497 | A | C | 0.0125 | 0.002 | 8.88E-10 | 806786 |
| rs4834272 | T | C | -0.0108 | 0.0017 | 3.46E-10 | 806658 |
| rs4864201 | T | C | 0.0137 | 0.0017 | 4.30E-16 | 806465 |
| rs11724872 | T | C | -0.0095 | 0.0016 | 8.34E-09 | 806480 |
| rs17019336 | A | T | -0.0128 | 0.0019 | 2.45E-11 | 804991 |
| rs3914628 | T | C | 0.0165 | 0.0023 | 6.91E-13 | 806780 |
| rs13110266 | A | G | -0.0124 | 0.0016 | 3.96E-14 | 806749 |
| rs12189178 | T | C | 0.0348 | 0.0046 | 4.30E-14 | 805880 |
| rs4865796 | A | G | -0.0096 | 0.0018 | 4.21E-08 | 800901 |
| rs2112347 | T | G | 0.0276 | 0.0017 | 1.17E-61 | 806699 |
| rs6870983 | T | C | -0.0208 | 0.0019 | 4.90E-27 | 806785 |
| rs7713317 | A | G | -0.0166 | 0.0018 | 1.96E-20 | 802425 |
| rs459552 | A | T | -0.0133 | 0.0019 | 8.36E-12 | 797086 |
| rs13174863 | A | G | -0.0197 | 0.0023 | 1.94E-17 | 793550 |
| rs7715256 | T | G | -0.0158 | 0.0016 | 3.98E-22 | 806764 |
| rs7734385 | A | G | -0.0101 | 0.0016 | 6.08E-10 | 805693 |
| rs12519652 | T | C | 0.0097 | 0.0017 | 1.58E-08 | 801200 |
| rs17695092 | T | G | 0.0114 | 0.0018 | 1.40E-10 | 790186 |
| rs6556301 | T | G | -0.0113 | 0.0017 | 8.14E-11 | 767716 |
| rs9463175 | T | C | -0.0115 | 0.0017 | 2.24E-11 | 794719 |
| rs2228213 | A | G | -0.0144 | 0.0017 | 5.50E-17 | 806782 |
| rs3806114 | A | G | -0.012 | 0.0018 | 1.46E-11 | 804965 |
| rs2229768 | T | C | 0.0134 | 0.0019 | 2.09E-12 | 806796 |
| rs2178899 | A | T | 0.0243 | 0.0025 | 5.44E-23 | 797281 |
| rs7748777 | A | G | 0.0107 | 0.0016 | 4.18E-11 | 806317 |
| rs6901756 | T | C | 0.0141 | 0.0024 | 7.50E-09 | 806733 |
| rs1358980 | T | C | -0.0129 | 0.0017 | 5.15E-15 | 797930 |
| rs2206277 | T | C | 0.0408 | 0.0021 | 1.82E-83 | 806768 |
| rs9688431 | T | C | 0.0212 | 0.0034 | 3.62E-10 | 801216 |
| rs9294260 | A | G | 0.014 | 0.0016 | 8.16E-18 | 806412 |
| rs1324110 | C | G | -0.0097 | 0.0016 | 3.01E-09 | 806168 |
| rs17789218 | T | C | -0.0111 | 0.0019 | 4.78E-09 | 806769 |
| rs768023 | A | G | 0.0161 | 0.0016 | 1.13E-22 | 806650 |
| rs2357760 | A | G | 0.0143 | 0.0017 | 2.11E-16 | 806795 |
| rs2246012 | T | C | -0.0161 | 0.0022 | 1.15E-13 | 806786 |
| rs11754747 | T | C | 0.0123 | 0.0019 | 1.18E-10 | 804870 |
| rs2185027 | A | C | -0.015 | 0.0018 | 1.40E-17 | 803718 |
| rs1557100 | A | C | -0.0179 | 0.0025 | 8.43E-13 | 806776 |
| rs394487 | T | C | 0.0098 | 0.0018 | 4.30E-08 | 806403 |
| rs13191362 | A | G | 0.0235 | 0.0025 | 4.08E-21 | 806582 |
| rs9364687 | T | G | -0.0097 | 0.0016 | 3.03E-09 | 806594 |
| rs9356132 | T | C | -0.0099 | 0.0018 | 1.79E-08 | 806398 |
| rs6461115 | A | G | 0.0134 | 0.0019 | 1.47E-12 | 806292 |
| rs6463489 | T | C | 0.0167 | 0.0026 | 2.50E-10 | 805923 |
| rs9638713 | A | G | 0.0298 | 0.0052 | 9.24E-09 | 779808 |
| rs6968554 | A | G | -0.0107 | 0.0017 | 2.34E-10 | 806326 |
| rs849135 | A | G | 0.0106 | 0.0016 | 4.74E-11 | 805299 |
| rs2717926 | T | C | 0.01 | 0.0017 | 7.76E-09 | 806770 |
| rs217433 | T | C | -0.0115 | 0.0021 | 3.00E-08 | 805664 |
| rs10269783 | A | G | 0.0126 | 0.0017 | 2.26E-14 | 804361 |
| rs10499694 | A | G | 0.013 | 0.0016 | 1.29E-15 | 806696 |
| rs10237317 | A | G | -0.0112 | 0.0017 | 1.26E-11 | 805984 |
| rs2245368 | T | C | -0.0238 | 0.0023 | 1.72E-25 | 690355 |
| rs2299383 | T | C | 0.0162 | 0.0016 | 6.89E-23 | 801620 |
| rs11973097 | T | C | -0.023 | 0.004 | 1.12E-08 | 787944 |
| rs1899689 | T | C | 0.012 | 0.0017 | 4.20E-13 | 806684 |
| rs972283 | A | G | 0.0089 | 0.0016 | 4.24E-08 | 806776 |
| rs1700082 | C | G | 0.0094 | 0.0017 | 3.38E-08 | 803933 |
| rs4240673 | T | C | 0.0175 | 0.0016 | 1.58E-26 | 806735 |
| rs11781222 | T | C | 0.0167 | 0.0024 | 3.25E-12 | 806346 |
| rs17446091 | T | C | -0.0127 | 0.002 | 3.73E-10 | 806546 |
| rs7828890 | A | G | -0.0152 | 0.0028 | 4.48E-08 | 794853 |
| rs12334435 | T | C | -0.0101 | 0.0018 | 3.71E-08 | 806631 |
| rs6468188 | T | C | 0.0093 | 0.0016 | 1.14E-08 | 805422 |
| rs17405819 | T | C | 0.0211 | 0.0018 | 6.04E-33 | 806765 |
| rs16907751 | T | C | -0.0194 | 0.0029 | 1.63E-11 | 792432 |
| rs12680842 | A | G | 0.0142 | 0.0017 | 3.41E-16 | 800300 |
| rs3808434 | A | G | 0.0115 | 0.0016 | 1.91E-12 | 806715 |
| rs11781699 | T | C | -0.0141 | 0.0021 | 1.56E-11 | 795827 |
| rs12675063 | A | T | -0.0158 | 0.0025 | 2.70E-10 | 804382 |
| rs4740619 | T | C | 0.0189 | 0.0016 | 3.15E-31 | 806567 |
| rs1412235 | C | G | 0.0237 | 0.0017 | 2.28E-42 | 804074 |
| rs10757826 | A | G | 0.0102 | 0.0017 | 5.37E-09 | 794739 |
| rs7043482 | A | C | 0.0097 | 0.0017 | 1.65E-08 | 802925 |
| rs10990303 | T | C | -0.0112 | 0.002 | 1.25E-08 | 806728 |
| rs420158 | T | C | -0.0107 | 0.0019 | 9.41E-09 | 806778 |
| rs7024334 | T | G | 0.0135 | 0.002 | 4.71E-12 | 799813 |
| rs6477694 | T | C | -0.0126 | 0.0017 | 6.57E-14 | 806728 |
| rs1928295 | T | C | 0.0134 | 0.0016 | 2.23E-16 | 806659 |
| rs10818810 | A | G | 0.0131 | 0.0017 | 7.71E-15 | 787901 |
| rs10733682 | A | G | 0.0148 | 0.0016 | 1.76E-19 | 805407 |
| rs1993414 | T | G | 0.0189 | 0.0034 | 1.73E-08 | 794602 |
| rs7899106 | A | G | -0.0327 | 0.0037 | 1.72E-18 | 806450 |
| rs17094222 | T | C | -0.0173 | 0.002 | 4.04E-18 | 806450 |
| rs7083450 | T | C | 0.0165 | 0.0022 | 1.56E-13 | 806791 |
| rs10883759 | A | G | -0.0118 | 0.0018 | 1.96E-11 | 806644 |
| rs12413409 | A | G | 0.0267 | 0.0029 | 4.73E-20 | 806514 |
| rs7903146 | T | C | -0.0178 | 0.0018 | 1.67E-23 | 806810 |
| rs10886017 | A | C | 0.0146 | 0.0019 | 4.47E-15 | 806301 |
| rs17636031 | T | C | -0.0154 | 0.0018 | 3.87E-17 | 798189 |
| rs4256980 | C | G | -0.0187 | 0.0017 | 8.63E-29 | 804708 |
| rs11023948 | T | C | -0.0121 | 0.002 | 8.41E-10 | 806624 |
| rs1557765 | T | C | -0.0122 | 0.0017 | 2.61E-13 | 800622 |
| rs17309825 | T | C | -0.0301 | 0.0042 | 6.14E-13 | 802521 |
| rs7127273 | T | C | 0.024 | 0.004 | 2.80E-09 | 785832 |
| rs2862996 | T | G | -0.0216 | 0.0017 | 3.60E-35 | 806675 |
| rs10742752 | T | C | -0.0123 | 0.0017 | 1.20E-13 | 806700 |
| rs7124681 | A | C | 0.0257 | 0.0016 | 3.96E-55 | 806656 |
| rs6591407 | A | C | -0.0124 | 0.0021 | 3.58E-09 | 806782 |
| rs7938117 | A | G | 0.0107 | 0.0017 | 9.68E-10 | 806777 |
| rs592483 | T | C | -0.0137 | 0.0017 | 1.53E-16 | 802791 |
| rs7123876 | T | C | -0.0118 | 0.0019 | 2.92E-10 | 806649 |
| rs2605603 | A | G | -0.0103 | 0.0016 | 2.04E-10 | 806679 |
| rs12286929 | A | G | -0.0177 | 0.0016 | 1.93E-27 | 806583 |
| rs693701 | T | C | 0.0108 | 0.0019 | 1.04E-08 | 806706 |
| rs329651 | T | G | 0.016 | 0.0021 | 2.13E-14 | 799668 |
| rs12364470 | T | G | -0.0187 | 0.0022 | 2.18E-17 | 799515 |
| rs11611246 | T | G | 0.0223 | 0.002 | 2.04E-28 | 793063 |
| rs11170468 | A | C | 0.013 | 0.0019 | 1.12E-11 | 806636 |
| rs11181001 | A | G | 0.0136 | 0.0016 | 1.17E-16 | 805907 |
| rs2269828 | A | G | -0.0097 | 0.0018 | 3.98E-08 | 806232 |
| rs7138803 | A | G | 0.0297 | 0.0017 | 3.10E-71 | 806772 |
| rs4759075 | T | C | 0.0118 | 0.0016 | 4.49E-13 | 806786 |
| rs11115176 | T | C | 0.0131 | 0.0019 | 6.79E-12 | 805743 |
| rs11105839 | A | T | -0.0113 | 0.0017 | 1.25E-11 | 801313 |
| rs3825393 | T | C | -0.0102 | 0.0017 | 1.87E-09 | 806785 |
| rs6606686 | C | G | -0.0156 | 0.0017 | 3.57E-19 | 806671 |
| rs11066188 | A | G | -0.0114 | 0.0016 | 3.05E-12 | 805492 |
| rs7133378 | A | G | 0.0127 | 0.0017 | 2.93E-13 | 802968 |
| rs9595908 | T | C | 0.0154 | 0.0017 | 3.73E-20 | 806589 |
| rs7987314 | T | G | -0.0148 | 0.0026 | 7.44E-09 | 806251 |
| rs12429545 | A | G | 0.0313 | 0.0024 | 1.42E-37 | 797614 |
| rs2322622 | T | C | -0.0098 | 0.0017 | 4.64E-09 | 806789 |
| rs9540493 | A | G | 0.0129 | 0.0017 | 7.85E-15 | 803641 |
| rs10132280 | A | C | -0.0214 | 0.0018 | 2.28E-33 | 806477 |
| rs12885454 | A | C | -0.018 | 0.0017 | 5.44E-26 | 805503 |
| rs225883 | A | G | -0.0138 | 0.0024 | 6.65E-09 | 800934 |
| rs10483389 | T | C | 0.0334 | 0.0041 | 3.04E-16 | 794037 |
| rs1954494 | T | C | 0.0097 | 0.0016 | 2.43E-09 | 806720 |
| rs6573463 | A | C | 0.0101 | 0.0017 | 2.41E-09 | 806229 |
| rs1275691 | A | G | -0.0091 | 0.0016 | 2.01E-08 | 806683 |
| rs10146527 | T | C | 0.0126 | 0.0017 | 2.38E-13 | 793469 |
| rs7144011 | T | G | 0.0263 | 0.002 | 2.37E-40 | 806721 |
| rs4517716 | C | G | -0.0117 | 0.002 | 4.27E-09 | 806706 |
| rs9989141 | T | C | 0.0168 | 0.0017 | 8.16E-23 | 769164 |
| rs2010281 | A | G | -0.0155 | 0.0017 | 4.97E-20 | 806796 |
| rs12899905 | T | C | -0.0113 | 0.0018 | 6.16E-10 | 806664 |
| rs1559677 | A | G | -0.0115 | 0.0016 | 2.80E-12 | 806738 |
| rs6493498 | T | C | 0.0137 | 0.0016 | 4.84E-17 | 805967 |
| rs16965225 | T | G | 0.022 | 0.0033 | 5.32E-11 | 794658 |
| rs340025 | T | C | -0.0127 | 0.0016 | 1.40E-14 | 806687 |
| rs12595158 | T | C | -0.0382 | 0.0052 | 1.30E-13 | 786284 |
| rs2241423 | A | G | -0.0298 | 0.0019 | 3.60E-54 | 806797 |
| rs7164727 | T | C | 0.0171 | 0.0017 | 2.38E-23 | 805992 |
| rs12914489 | A | G | 0.0172 | 0.0027 | 8.78E-11 | 806429 |
| rs2290573 | A | G | 0.0101 | 0.0016 | 4.57E-10 | 806107 |
| rs11866815 | T | C | -0.0153 | 0.0019 | 2.21E-16 | 806553 |
| rs12448257 | A | G | 0.0161 | 0.002 | 9.04E-16 | 797946 |
| rs1876359 | T | C | 0.0128 | 0.0017 | 2.98E-14 | 806681 |
| rs977540 | A | G | 0.0134 | 0.0019 | 1.93E-12 | 806171 |
| rs12446632 | A | G | -0.0353 | 0.0024 | 3.12E-50 | 801438 |
| rs11074446 | T | C | 0.0214 | 0.0024 | 3.67E-19 | 794991 |
| rs7498665 | A | G | -0.0285 | 0.0017 | 1.14E-66 | 804216 |
| rs4889606 | A | G | 0.0209 | 0.0017 | 2.90E-36 | 806567 |
| rs9935222 | A | C | 0.0096 | 0.0017 | 1.75E-08 | 806550 |
| rs6500208 | A | G | 0.0146 | 0.002 | 3.21E-13 | 794515 |
| rs1477199 | A | G | -0.0221 | 0.0023 | 5.15E-21 | 806707 |
| rs7206790 | C | G | -0.0606 | 0.0017 | 1.00E-200 | 802720 |
| rs8063946 | T | C | -0.0238 | 0.0033 | 1.09E-12 | 802815 |
| rs8061518 | A | G | 0.0138 | 0.0017 | 7.05E-16 | 794179 |
| rs11076022 | A | G | 0.0109 | 0.0016 | 3.02E-11 | 806593 |
| rs7217226 | T | G | -0.0137 | 0.0017 | 9.43E-16 | 800625 |
| rs9893197 | T | C | -0.0106 | 0.0018 | 6.71E-09 | 805668 |
| rs4986044 | T | C | -0.0177 | 0.0016 | 1.27E-27 | 802591 |
| rs12150665 | T | C | 0.0168 | 0.0016 | 1.74E-24 | 806685 |
| rs17599948 | A | G | 0.0126 | 0.0022 | 7.38E-09 | 792995 |
| rs10491182 | T | C | 0.02 | 0.0031 | 9.09E-11 | 794959 |
| rs8071182 | A | G | 0.0125 | 0.0022 | 1.26E-08 | 784689 |
| rs9910424 | T | C | -0.0092 | 0.0016 | 1.44E-08 | 806002 |
| rs12325866 | A | G | -0.0136 | 0.0018 | 5.84E-14 | 806775 |
| rs12602912 | T | C | 0.0166 | 0.002 | 2.90E-16 | 788746 |
| rs312750 | A | G | 0.0097 | 0.0016 | 2.57E-09 | 806739 |
| rs12939549 | A | G | 0.018 | 0.0016 | 3.68E-28 | 806064 |
| rs1365466 | T | C | -0.012 | 0.0018 | 6.04E-11 | 806724 |
| rs7239883 | A | G | -0.0125 | 0.0017 | 5.58E-14 | 806589 |
| rs954018 | A | G | -0.0127 | 0.0018 | 3.57E-13 | 806752 |
| rs7239114 | A | G | 0.012 | 0.0017 | 6.24E-13 | 794467 |
| rs7243357 | T | G | 0.0198 | 0.0021 | 1.31E-20 | 806787 |
| rs6567160 | T | C | -0.0552 | 0.0019 | 7.82E-184 | 806638 |
| rs17066842 | A | G | -0.0644 | 0.0042 | 6.32E-54 | 781236 |
| rs8088123 | A | C | -0.0247 | 0.0033 | 3.78E-14 | 752439 |
| rs2012927 | A | G | 0.0144 | 0.0017 | 4.02E-17 | 806712 |
| rs11150911 | A | C | 0.0118 | 0.0018 | 3.72E-11 | 802197 |
| rs17724992 | A | G | 0.0172 | 0.0018 | 5.23E-21 | 804268 |
| rs4808934 | T | C | -0.0175 | 0.0022 | 3.85E-15 | 752265 |
| rs12971734 | T | C | 0.0211 | 0.0038 | 2.07E-08 | 794104 |
| rs2304130 | A | G | 0.017 | 0.0029 | 4.32E-09 | 795031 |
| rs29938 | T | C | -0.0158 | 0.0017 | 3.81E-20 | 803927 |
| rs7254892 | A | G | 0.0265 | 0.0046 | 1.18E-08 | 731144 |
| rs11672660 | T | C | -0.0338 | 0.0021 | 6.83E-60 | 781854 |
| rs3810291 | A | G | 0.0262 | 0.0017 | 2.36E-51 | 780941 |
| rs1884897 | A | G | -0.0184 | 0.0017 | 2.70E-28 | 806711 |
| rs6056413 | A | T | 0.0125 | 0.0022 | 1.81E-08 | 764178 |
| rs17272434 | A | G | -0.0097 | 0.0018 | 4.27E-08 | 806748 |
| rs8123881 | A | G | -0.0177 | 0.0024 | 1.05E-13 | 805952 |
| rs4911442 | A | G | 0.0143 | 0.0024 | 2.44E-09 | 806465 |
| rs16989232 | A | G | 0.0115 | 0.0017 | 6.19E-12 | 805861 |
| rs2426424 | T | C | 0.0185 | 0.0028 | 5.73E-11 | 794898 |
| rs6010784 | T | C | 0.0106 | 0.0016 | 6.91E-11 | 803088 |
| rs2832283 | A | G | 0.0115 | 0.002 | 4.72E-09 | 806772 |
| rs2836961 | A | C | -0.0103 | 0.0017 | 9.11E-10 | 806380 |
| rs8126575 | T | G | 0.0152 | 0.0025 | 5.88E-10 | 775870 |
| rs427943 | A | C | -0.0177 | 0.0017 | 3.60E-25 | 742360 |
