## Supplementary material for "Causal Mediation Analysis with Multiple Causally Ordered and Non-ordered Mediators based on Summarized Genetic Data": S2 Appendix

### Supplemental simulation

To validate the utility of the multivariable Mendelian randomization method for estimating the direct causal and indirect effect, we performed a simulation analysis. We generated 25 genetic variants, a risk factor (*X*), mediators (*M*), and outcome(*Y*) for 20,000 individuals in a one-sample Mendelian randomization context. Briefly, we considered several different sets of values of the parameters
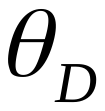
 (the direct effect of *X* on *Y*) and
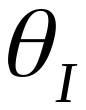
 (the indirect effect of *X* on *M*). We included scenarios where there is no direct effect, no indirect effect, a direct effect and a directionally concordant indirect effect, and a direct effect and a directionally discordant indirect effect. In all simulations, the
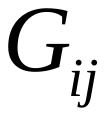
 were generated from a trinomial distribution, taking values (0, 1, 2) with probabilities (0.49, 0.42, 0.09) – this was equivalent to a single nucleotide polymorphism with minor allele frequency 0.3. And the combined F statistics of all genetic variables is above 3000.

We used the following metrics to evaluate the performance of the method: bias, standard deviations (SE), mean squared error (MSE), type I error rate for a null causal effect and empirical power to detect a non-null effect (that is, the proportion of confidence intervals excluding zero).

#### 1 Simulation A: PSE-IVW in one mediator setting

The data-generating models were

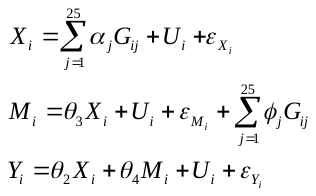

Error variables
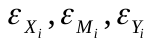
 were independently generated from a
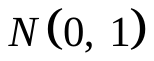
 distribution; and instrument strength parameters
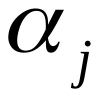
 were generated from a
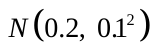
 distribution. The
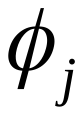
 parameters were generated from a normal distribution
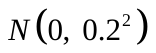
. These effects were included in the simulation model to ensure that the direct and indirect effect was identified.

The result were listed in **eTable 1-3**. We varied the sample size, the number of instrumental variables and simulated eight different sets of values of the parameters. We saw that causal estimates of direct and indirect effects were close to unbiased. It also had good coverage and Type 1 error properties. As the sample size increased, bias and standard deviations decreased, with increased power. With increasing of numbers of instrumental variables, the higher the proportion of variation explained by exposure, the higher power, and the smaller bias.

**eTable 1**. Simulation of PSE-IVW in one mediator setting with different numbers of genetic variants.

|  | BIAS | | | | Power | | | |
| --- | --- | --- | --- | --- | --- | --- | --- | --- |
| g | TE | DE | IE_d | IE_p | TE | DE | IE_d | IE_p |
| 3 | 0.002 | -0.031 | 0.033 | 0.033 | 0.793 | 0.000 | 0.006 | 0.127 |
| 5 | 0.004 | -0.020 | 0.024 | 0.024 | 1.000 | 0.478 | 0.161 | 0.534 |
| 10 | 0.006 | -0.016 | 0.022 | 0.022 | 1.000 | 0.982 | 0.836 | 0.961 |
| 15 | 0.007 | -0.018 | 0.025 | 0.025 | 1.000 | 1.000 | 0.984 | 0.996 |
| 20 | 0.008 | -0.017 | 0.025 | 0.025 | 1.000 | 1.000 | 1.000 | 1.000 |
| 30 | 0.008 | -0.015 | 0.023 | 0.023 | 1.000 | 1.000 | 1.000 | 1.000 |
| 50 | 0.007 | -0.017 | 0.023 | 0.024 | 1.000 | 1.000 | 1.000 | 1.000 |
| 100 | 0.005 | -0.017 | 0.022 | 0.022 | 1.000 | 1.000 | 1.000 | 1.000 |
| 150 | 0.005 | -0.017 | 0.022 | 0.022 | 1.000 | 1.000 | 1.000 | 1.000 |

TE: total effect; DE: direct effect; IE: indirect effect; IE_d: indirect effect calculated by difference method; IE_p: indirect effect calculated by product method.

e**Table** **2. Simulation of PSE-IVW in one mediator setting with different effects.**

| TRUE | | | | | BIAS | | | | SE | | | | MSE | | | | Power/Type I error | | | |
| --- | --- | --- | --- | --- | --- | --- | --- | --- | --- | --- | --- | --- | --- | --- | --- | --- | --- | --- | --- | --- |
| TE | DE | IE | $\theta_{4}$ | $\theta_{3}$ | TE | DE | IE_d | IE_p | TE | DE | IE_d | IE_p | TE | DE | IE_d | IE_p | TE | DE | IE_d | IE_p |
| 0.8 | 0.5 | 0.3 | 1 | 0.3 | 0.014 | -0.028 | 0.042 | 0.042 | 0.057 | 0.037 | 0.061 | 0.061 | 0.003 | 0.002 | 0.006 | 0.006 | 1.000 | 1.000 | 0.997 | 1.000 |
| 0.2 | 0.5 | -0.3 | -1 | 0.3 | 0.002 | -0.016 | 0.018 | 0.018 | 0.041 | 0.028 | 0.046 | 0.046 | 0.002 | 0.001 | 0.002 | 0.002 | 0.997 | 1.000 | 1.000 | 1.000 |
| -0.2 | -0.5 | 0.3 | 1 | 0.3 | 0.008 | -0.018 | 0.026 | 0.027 | 0.049 | 0.027 | 0.051 | 0.051 | 0.002 | 0.001 | 0.003 | 0.003 | 0.967 | 1.000 | 1.000 | 1.000 |
| -0.8 | -0.5 | -0.3 | 1 | -0.3 | 0.004 | 0.022 | -0.019 | -0.019 | 0.050 | 0.028 | 0.054 | 0.054 | 0.002 | 0.001 | 0.003 | 0.003 | 1.000 | 1.000 | 1.000 | 1.000 |
| 0.5 | 0.5 | 0 | 1 | 0 | 0.007 | 0.002 | 0.005 | 0.005 | 0.047 | 0.015 | 0.045 | 0.045 | 0.002 | 0.000 | 0.002 | 0.002 | 1.000 | 1.000 | 0.006 | 0.056 |
| 0.5 | 0.5 | 0 | 0 | 0.3 | 0.003 | -0.018 | 0.020 | 0.020 | 0.015 | 0.027 | 0.024 | 0.024 | 0.000 | 0.001 | 0.001 | 0.001 | 1.000 | 1.000 | 0.004 | 0.041 |
| 0.3 | 0 | 0.3 | 1 | 0.3 | 0.009 | -0.017 | 0.026 | 0.026 | 0.050 | 0.028 | 0.054 | 0.054 | 0.003 | 0.001 | 0.004 | 0.004 | 1.000 | 0.000 | 1.000 | 1.000 |
| 0 | 0.3 | -0.3 | 1 | -0.3 | 0.007 | 0.022 | -0.016 | -0.016 | 0.049 | 0.026 | 0.051 | 0.051 | 0.002 | 0.001 | 0.003 | 0.003 | 0.055 | 1.000 | 1.000 | 1.000 |

TE: total effect; DE: direct effect; IE: indirect effect; IE_d: indirect effect calculated by difference method; IE_p: indirect effect calculated by product method.

**eTable 3.** Simulation of PSE-IVW in one mediator setting with different sample size.

|  | BIAS | | | | SE | | | | MSE | | | | Power | | | |
| --- | --- | --- | --- | --- | --- | --- | --- | --- | --- | --- | --- | --- | --- | --- | --- | --- |
| N | TE | DE | IE_d | IE_p | TE | DE | IE_d | IE_p | TE | DE | IE_d | IE_p | TE | DE | IE_d | IE_p |
| 2000 | 0.055 | -0.070 | 0.124 | 0.124 | 0.090 | 0.068 | 0.099 | 0.099 | 0.011 | 0.009 | 0.025 | 0.025 | 1.000 | 0.845 | 0.638 | 0.990 |
| 4000 | 0.028 | -0.053 | 0.081 | 0.081 | 0.074 | 0.055 | 0.079 | 0.079 | 0.006 | 0.006 | 0.013 | 0.013 | 1.000 | 0.994 | 0.899 | 0.999 |
| 6000 | 0.017 | -0.041 | 0.058 | 0.058 | 0.060 | 0.047 | 0.067 | 0.067 | 0.004 | 0.004 | 0.008 | 0.008 | 1.000 | 1.000 | 0.971 | 0.999 |
| 8000 | 0.013 | -0.034 | 0.047 | 0.047 | 0.060 | 0.041 | 0.063 | 0.063 | 0.004 | 0.003 | 0.006 | 0.006 | 1.000 | 1.000 | 0.996 | 0.999 |
| 10000 | 0.010 | -0.030 | 0.040 | 0.040 | 0.055 | 0.038 | 0.060 | 0.060 | 0.003 | 0.002 | 0.005 | 0.005 | 1.000 | 1.000 | 1.000 | 1.000 |
| 12000 | 0.009 | -0.026 | 0.034 | 0.034 | 0.053 | 0.035 | 0.058 | 0.058 | 0.003 | 0.002 | 0.005 | 0.005 | 1.000 | 1.000 | 1.000 | 1.000 |
| 14000 | 0.008 | -0.023 | 0.031 | 0.031 | 0.052 | 0.033 | 0.056 | 0.056 | 0.003 | 0.002 | 0.004 | 0.004 | 1.000 | 1.000 | 1.000 | 1.000 |
| 16000 | 0.004 | -0.021 | 0.026 | 0.026 | 0.050 | 0.031 | 0.054 | 0.054 | 0.003 | 0.001 | 0.004 | 0.004 | 1.000 | 1.000 | 1.000 | 1.000 |
| 18000 | 0.005 | -0.019 | 0.024 | 0.024 | 0.047 | 0.029 | 0.050 | 0.050 | 0.002 | 0.001 | 0.003 | 0.003 | 1.000 | 1.000 | 1.000 | 1.000 |
| 20000 | 0.004 | -0.017 | 0.021 | 0.021 | 0.047 | 0.028 | 0.050 | 0.050 | 0.002 | 0.001 | 0.003 | 0.003 | 1.000 | 1.000 | 1.000 | 1.000 |

TE: total effect; DE: direct effect; IE: indirect effect; IE_d: indirect effect calculated by difference method; IE_p: indirect effect calculated by product method.

#### 2 Simulation B: PSE-Egger in one mediator setting

The data-generating models were

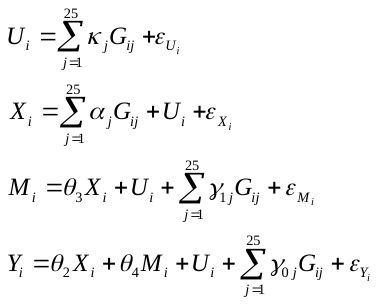

Error variables
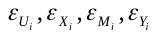
 were independently generated from a
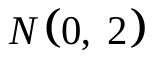
 distribution; and instrument strength parameters
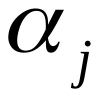
 were generated from a
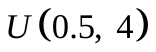
 distribution. This model additionally allowed genetic variant
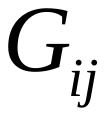
 to be associated with confounders variable
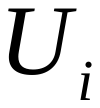
. This occurred when
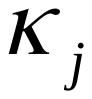
 was non-zero (a violation of the **Assumption Ⅱ**). Simulation case (a)–(c) were implemented by additionally specifying values for
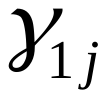
 ( the direct effect of genetic variables
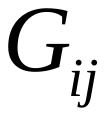
 on mediator *M*) and
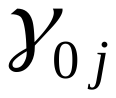
 ( the direct effect of genetic variables
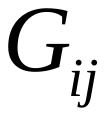
 on outcome *Y*) as below:

**eTable 4.** Settings for simulation of PSE-Egger in one mediators setting

|  | Pleiotropy | InSIDE | 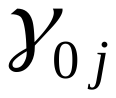 | 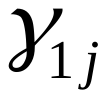 | 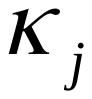 |  |
| --- | --- | --- | --- | --- | --- | --- |
| case(a) | Balanced | Satisfied | 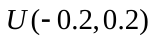 | 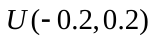 | 0 | sc2 |
|  |  |  | 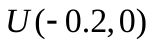 | 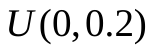 | 0 | sc21 |
| case(b) | Directional | Satisfied | 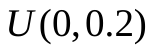 | 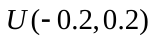 | 0 | sc31 |
|  |  |  | 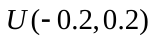 | 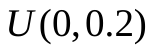 | 0 | sc32 |
|  |  |  | 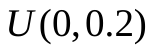 | 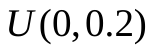 | 0 | sc3 |
| case(c) | Directional | Not satisfied | 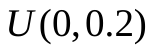 | 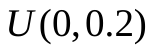 | 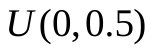 | Sc4 |

In case of each pathway had balanced pleiotropy (sc2), we saw that bias and Type 1 error properties were almost identical between the two methods. However, MR-Egger had less bias and great Type 1 error properties than IVW in case where at least one pathway had directional pleiotropy (sc21 and case b). MR-Egger also had better performance than IVW in bias when InSIDE assumption was not satisfied (case c). In all cases, MR-Egger had larger standard deviations than IVW, with larger CI width. Two methods estimating the indirect effect had similar bias and standard deviations. While product method had better Type 1 error properties than difference method. As sample size and the numbers of instrumental variables increased, Single-mediator MR-Egger had similar performance with Single-mediator IVW. Results were listed in **eTable 5**.

**eTable 5.** Simulation of PSE-Egger in one mediator setting with different effects.

|  |  | TRUE | | | | | BIAS | | | | SE | | | | MSE | | | | Power/Type I error | | | | | |
| --- | --- | --- | --- | --- | --- | --- | --- | --- | --- | --- | --- | --- | --- | --- | --- | --- | --- | --- | --- | --- | --- | --- | --- | --- |
|  |  | TE | DE | IE | $\theta_{4}$ | $\theta_{3}$ | TE | DE | IE_d | IE_p | TE | DE | IE_d | IE_p | TE | DE | IE_d | IE_p | TE | DE | IE_d | IE_p | GM | GY |
| Sc2 | IVW | 0.8 | 0.5 | 0.3 | 1 | 0.3 | 0.000 | -0.010 | 0.011 | 0.011 | 0.014 | 0.062 | 0.062 | 0.062 | 0.000 | 0.004 | 0.004 | 0.004 | 1.000 | 1.000 | 0.997 | 0.998 |  |  |
|  |  | 0.2 | 0.5 | -0.3 | -1 | 0.3 | 0.001 | -0.010 | 0.010 | 0.010 | 0.013 | 0.065 | 0.065 | 0.065 | 0.000 | 0.004 | 0.004 | 0.004 | 1.000 | 1.000 | 0.994 | 0.994 |  |  |
|  |  | -0.2 | -0.5 | 0.3 | 1 | 0.3 | 0.000 | -0.014 | 0.014 | 0.015 | 0.014 | 0.062 | 0.062 | 0.062 | 0.000 | 0.004 | 0.004 | 0.004 | 1.000 | 1.000 | 0.999 | 1.000 |  |  |
|  |  | -0.8 | -0.5 | -0.3 | 1 | -0.3 | 0.000 | 0.011 | -0.011 | -0.011 | 0.015 | 0.064 | 0.064 | 0.063 | 0.000 | 0.004 | 0.004 | 0.004 | 1.000 | 1.000 | 0.999 | 0.999 |  |  |
|  |  | 0.5 | 0.5 | 0 | 1 | 0 | 0.000 | 0.000 | 0.000 | 0.000 | 0.014 | 0.010 | 0.010 | 0.010 | 0.000 | 0.000 | 0.000 | 0.000 | 1.000 | 1.000 | 0.001 | 0.060 |  |  |
|  |  | 0.5 | 0.5 | 0 | 0 | 0.3 | 0.000 | -0.013 | 0.014 | 0.014 | 0.010 | 0.062 | 0.061 | 0.062 | 0.000 | 0.004 | 0.004 | 0.004 | 1.000 | 1.000 | 0.052 | 0.063 |  |  |
|  |  | 0.3 | 0 | 0.3 | 1 | 0.3 | 0.001 | -0.009 | 0.011 | 0.011 | 0.015 | 0.066 | 0.066 | 0.066 | 0.000 | 0.004 | 0.004 | 0.004 | 1.000 | 0.056 | 0.996 | 0.998 |  |  |
|  |  | 0 | 0.3 | -0.3 | 1 | -0.3 | 0.000 | 0.009 | -0.009 | -0.010 | 0.014 | 0.065 | 0.066 | 0.066 | 0.000 | 0.004 | 0.004 | 0.004 | 0.051 | 0.997 | 0.998 | 0.998 |  |  |
|  | Egger | 0.8 | 0.5 | 0.3 | 1 | 0.3 | 0.002 | -0.010 | 0.012 | 0.012 | 0.035 | 0.068 | 0.067 | 0.067 | 0.001 | 0.005 | 0.005 | 0.005 | 1.000 | 1.000 | 0.993 | 0.996 | 0.052 | 0.044 |
|  |  | 0.2 | 0.5 | -0.3 | -1 | 0.3 | 0.000 | -0.010 | 0.010 | 0.009 | 0.034 | 0.071 | 0.070 | 0.070 | 0.001 | 0.005 | 0.005 | 0.005 | 0.999 | 1.000 | 0.987 | 0.994 | 0.041 | 0.051 |
|  |  | -0.2 | -0.5 | 0.3 | 1 | 0.3 | 0.002 | -0.014 | 0.015 | 0.015 | 0.034 | 0.066 | 0.068 | 0.068 | 0.001 | 0.005 | 0.005 | 0.005 | 0.998 | 1.000 | 0.994 | 0.996 | 0.047 | 0.038 |
|  |  | -0.8 | -0.5 | -0.3 | 1 | -0.3 | 0.000 | 0.010 | -0.011 | -0.011 | 0.036 | 0.071 | 0.069 | 0.069 | 0.001 | 0.005 | 0.005 | 0.005 | 1.000 | 1.000 | 0.996 | 0.999 | 0.048 | 0.056 |
|  |  | 0.5 | 0.5 | 0 | 1 | 0 | 0.001 | 0.001 | 0.001 | 0.000 | 0.035 | 0.024 | 0.027 | 0.027 | 0.001 | 0.001 | 0.001 | 0.001 | 1.000 | 1.000 | 0.003 | 0.068 | 0.050 | 0.048 |
|  |  | 0.5 | 0.5 | 0 | 0 | 0.3 | 0.001 | -0.013 | 0.013 | 0.013 | 0.025 | 0.067 | 0.064 | 0.064 | 0.001 | 0.005 | 0.004 | 0.004 | 1.000 | 1.000 | 0.041 | 0.073 | 0.051 | 0.047 |
|  |  | 0.3 | 0 | 0.3 | 1 | 0.3 | 0.002 | -0.008 | 0.010 | 0.010 | 0.035 | 0.071 | 0.070 | 0.070 | 0.001 | 0.005 | 0.005 | 0.005 | 1.000 | 0.065 | 0.993 | 0.995 | 0.044 | 0.043 |
|  |  | 0 | 0.3 | -0.3 | 1 | -0.3 | 0.001 | 0.011 | -0.010 | -0.010 | 0.035 | 0.071 | 0.072 | 0.072 | 0.001 | 0.005 | 0.005 | 0.005 | 0.049 | 0.990 | 0.996 | 0.999 | 0.065 | 0.053 |
| Sc21 | IVW | 0.8 | 0.5 | 0.3 | 1 | 0.3 | 0.001 | 0.034 | -0.034 | -0.034 | 0.008 | 0.070 | 0.070 | 0.070 | 0.000 | 0.006 | 0.006 | 0.006 | 1.000 | 1.000 | 0.894 | 0.897 |  |  |
|  |  | 0.2 | 0.5 | -0.3 | -1 | 0.3 | -0.074 | 0.032 | -0.106 | -0.106 | 0.009 | 0.070 | 0.071 | 0.071 | 0.006 | 0.006 | 0.016 | 0.016 | 1.000 | 1.000 | 0.998 | 0.998 |  |  |
|  |  | -0.2 | -0.5 | 0.3 | 1 | 0.3 | 0.000 | 0.034 | -0.034 | -0.034 | 0.008 | 0.069 | 0.069 | 0.069 | 0.000 | 0.006 | 0.006 | 0.006 | 1.000 | 1.000 | 0.965 | 0.965 |  |  |
|  |  | -0.8 | -0.5 | -0.3 | 1 | -0.3 | 0.000 | -0.089 | 0.089 | 0.089 | 0.008 | 0.053 | 0.052 | 0.052 | 0.000 | 0.011 | 0.011 | 0.011 | 1.000 | 1.000 | 0.950 | 0.957 |  |  |
|  |  | 0.5 | 0.5 | 0 | 1 | 0 | 0.000 | -0.030 | 0.030 | 0.030 | 0.008 | 0.009 | 0.009 | 0.009 | 0.000 | 0.001 | 0.001 | 0.001 | 1.000 | 1.000 | 0.704 | 0.958 |  |  |
|  |  | 0.5 | 0.5 | 0 | 0 | 0.3 | -0.037 | 0.031 | -0.068 | -0.068 | 0.006 | 0.067 | 0.067 | 0.067 | 0.001 | 0.005 | 0.009 | 0.009 | 1.000 | 1.000 | 0.176 | 0.198 |  |  |
|  |  | 0.3 | 0 | 0.3 | 1 | 0.3 | 0.000 | 0.026 | -0.026 | -0.026 | 0.008 | 0.067 | 0.066 | 0.066 | 0.000 | 0.005 | 0.005 | 0.005 | 1.000 | 0.059 | 0.965 | 0.967 |  |  |
|  |  | 0 | 0.3 | -0.3 | 1 | -0.3 | 0.000 | -0.088 | 0.088 | 0.088 | 0.008 | 0.054 | 0.054 | 0.054 | 0.000 | 0.011 | 0.011 | 0.011 | 0.045 | 0.958 | 0.967 | 0.969 |  |  |
|  | Egger | 0.8 | 0.5 | 0.3 | 1 | 0.3 | 0.002 | -0.030 | 0.032 | 0.032 | 0.020 | 0.068 | 0.068 | 0.068 | 0.000 | 0.006 | 0.006 | 0.006 | 1.000 | 1.000 | 0.959 | 0.970 | 0.410 | 0.798 |
|  |  | 0.2 | 0.5 | -0.3 | -1 | 0.3 | 0.000 | -0.031 | 0.031 | 0.030 | 0.018 | 0.066 | 0.066 | 0.066 | 0.000 | 0.005 | 0.005 | 0.005 | 1.000 | 1.000 | 0.979 | 0.980 | 0.763 | 0.829 |
|  |  | -0.2 | -0.5 | 0.3 | 1 | 0.3 | 0.001 | -0.029 | 0.030 | 0.030 | 0.019 | 0.066 | 0.066 | 0.066 | 0.000 | 0.005 | 0.005 | 0.005 | 1.000 | 1.000 | 0.993 | 0.993 | 0.784 | 0.813 |
|  |  | -0.8 | -0.5 | -0.3 | 1 | -0.3 | -0.001 | 0.033 | -0.034 | -0.034 | 0.021 | 0.068 | 0.069 | 0.069 | 0.000 | 0.006 | 0.006 | 0.006 | 1.000 | 0.998 | 0.988 | 0.991 | 0.592 | 0.816 |
|  |  | 0.5 | 0.5 | 0 | 1 | 0 | 0.000 | -0.001 | 0.001 | 0.001 | 0.020 | 0.014 | 0.015 | 0.015 | 0.000 | 0.000 | 0.000 | 0.000 | 1.000 | 1.000 | 0.001 | 0.066 | 0.639 | 0.824 |
|  |  | 0.5 | 0.5 | 0 | 0 | 0.3 | 0.000 | -0.032 | 0.033 | 0.033 | 0.013 | 0.064 | 0.063 | 0.063 | 0.000 | 0.005 | 0.005 | 0.005 | 1.000 | 1.000 | 0.062 | 0.074 | 0.771 | 0.818 |
|  |  | 0.3 | 0 | 0.3 | 1 | 0.3 | 0.000 | -0.035 | 0.035 | 0.035 | 0.020 | 0.065 | 0.065 | 0.065 | 0.000 | 0.005 | 0.006 | 0.006 | 1.000 | 0.052 | 0.994 | 0.994 | 0.725 | 0.803 |
|  |  | 0 | 0.3 | -0.3 | 1 | -0.3 | 0.001 | 0.035 | -0.034 | -0.034 | 0.019 | 0.064 | 0.064 | 0.064 | 0.000 | 0.005 | 0.005 | 0.005 | 0.036 | 0.996 | 0.997 | 0.997 | 0.788 | 0.805 |
| Sc3 | IVW | 0.8 | 0.5 | 0.3 | 1 | 0.3 | 0.075 | -0.083 | 0.158 | 0.158 | 0.009 | 0.063 | 0.064 | 0.064 | 0.006 | 0.011 | 0.029 | 0.029 | 1.000 | 0.998 | 0.999 | 0.999 |  |  |
|  |  | 0.2 | 0.5 | -0.3 | -1 | 0.3 | 0.000 | -0.082 | 0.082 | 0.082 | 0.007 | 0.065 | 0.065 | 0.066 | 0.000 | 0.011 | 0.011 | 0.011 | 1.000 | 1.000 | 0.904 | 0.906 |  |  |
|  |  | -0.2 | -0.5 | 0.3 | 1 | 0.3 | 0.075 | -0.080 | 0.155 | 0.155 | 0.010 | 0.064 | 0.065 | 0.065 | 0.006 | 0.011 | 0.028 | 0.028 | 1.000 | 1.000 | 1.000 | 1.000 |  |  |
|  |  | -0.8 | -0.5 | -0.3 | 1 | -0.3 | 0.074 | 0.130 | -0.056 | -0.056 | 0.009 | 0.051 | 0.052 | 0.052 | 0.006 | 0.020 | 0.006 | 0.006 | 1.000 | 1.000 | 1.000 | 1.000 |  |  |
|  |  | 0.5 | 0.5 | 0 | 1 | 0 | 0.074 | 0.024 | 0.050 | 0.050 | 0.010 | 0.009 | 0.011 | 0.011 | 0.006 | 0.001 | 0.003 | 0.003 | 1.000 | 1.000 | 0.994 | 0.999 |  |  |
|  |  | 0.5 | 0.5 | 0 | 0 | 0.3 | 0.037 | -0.083 | 0.120 | 0.120 | 0.006 | 0.065 | 0.065 | 0.065 | 0.001 | 0.011 | 0.019 | 0.019 | 1.000 | 1.000 | 0.451 | 0.465 |  |  |
|  |  | 0.3 | 0 | 0.3 | 1 | 0.3 | 0.074 | -0.081 | 0.155 | 0.155 | 0.009 | 0.065 | 0.065 | 0.065 | 0.006 | 0.011 | 0.028 | 0.028 | 1.000 | 0.186 | 1.000 | 1.000 |  |  |
|  |  | 0 | 0.3 | -0.3 | 1 | -0.3 | 0.074 | 0.131 | -0.057 | -0.057 | 0.009 | 0.050 | 0.050 | 0.050 | 0.006 | 0.020 | 0.006 | 0.006 | 1.000 | 1.000 | 0.999 | 0.999 |  |  |
|  | Egger | 0.8 | 0.5 | 0.3 | 1 | 0.3 | 0.002 | -0.033 | 0.035 | 0.035 | 0.019 | 0.064 | 0.064 | 0.064 | 0.000 | 0.005 | 0.005 | 0.005 | 1.000 | 1.000 | 0.958 | 0.965 | 0.142 | 0.814 |
|  |  | 0.2 | 0.5 | -0.3 | -1 | 0.3 | 0.000 | -0.031 | 0.031 | 0.031 | 0.018 | 0.066 | 0.066 | 0.066 | 0.000 | 0.005 | 0.005 | 0.005 | 1.000 | 1.000 | 0.979 | 0.981 | 0.567 | 0.835 |
|  |  | -0.2 | -0.5 | 0.3 | 1 | 0.3 | 0.002 | -0.030 | 0.032 | 0.032 | 0.020 | 0.064 | 0.065 | 0.065 | 0.000 | 0.005 | 0.005 | 0.005 | 1.000 | 1.000 | 0.995 | 0.996 | 0.570 | 0.794 |
|  |  | -0.8 | -0.5 | -0.3 | 1 | -0.3 | 0.002 | 0.035 | -0.033 | -0.033 | 0.020 | 0.066 | 0.068 | 0.068 | 0.000 | 0.006 | 0.006 | 0.006 | 1.000 | 1.000 | 0.993 | 0.996 | 0.436 | 0.800 |
|  |  | 0.5 | 0.5 | 0 | 1 | 0 | 0.001 | 0.000 | 0.001 | 0.001 | 0.021 | 0.014 | 0.015 | 0.015 | 0.000 | 0.000 | 0.000 | 0.000 | 1.000 | 1.000 | 0.001 | 0.064 | 0.378 | 0.809 |
|  |  | 0.5 | 0.5 | 0 | 0 | 0.3 | 0.001 | -0.032 | 0.033 | 0.033 | 0.014 | 0.066 | 0.065 | 0.065 | 0.000 | 0.005 | 0.005 | 0.005 | 1.000 | 1.000 | 0.071 | 0.097 | 0.541 | 0.811 |
|  |  | 0.3 | 0 | 0.3 | 1 | 0.3 | 0.001 | -0.028 | 0.030 | 0.030 | 0.020 | 0.063 | 0.063 | 0.063 | 0.000 | 0.005 | 0.005 | 0.005 | 1.000 | 0.038 | 0.998 | 1.000 | 0.551 | 0.817 |
|  |  | 0 | 0.3 | -0.3 | 1 | -0.3 | 0.001 | 0.034 | -0.032 | -0.032 | 0.020 | 0.063 | 0.064 | 0.064 | 0.000 | 0.005 | 0.005 | 0.005 | 0.050 | 0.994 | 0.996 | 0.996 | 0.567 | 0.816 |
| Sc31 | IVW | 0.8 | 0.5 | 0.3 | 1 | 0.3 | 0.038 | 0.029 | 0.009 | 0.009 | 0.012 | 0.040 | 0.042 | 0.042 | 0.002 | 0.002 | 0.002 | 0.002 | 1.000 | 1.000 | 1.000 | 1.000 |  |  |
|  |  | 0.2 | 0.5 | -0.3 | -1 | 0.3 | 0.038 | 0.028 | 0.009 | 0.009 | 0.012 | 0.043 | 0.043 | 0.043 | 0.002 | 0.003 | 0.002 | 0.002 | 1.000 | 1.000 | 1.000 | 1.000 |  |  |
|  |  | -0.2 | -0.5 | 0.3 | 1 | 0.3 | 0.037 | 0.026 | 0.011 | 0.011 | 0.012 | 0.040 | 0.041 | 0.041 | 0.002 | 0.002 | 0.002 | 0.002 | 1.000 | 1.000 | 1.000 | 1.000 |  |  |
|  |  | -0.8 | -0.5 | -0.3 | 1 | -0.3 | 0.037 | 0.047 | -0.010 | -0.011 | 0.012 | 0.040 | 0.041 | 0.041 | 0.002 | 0.004 | 0.002 | 0.002 | 1.000 | 1.000 | 1.000 | 1.000 |  |  |
|  |  | 0.5 | 0.5 | 0 | 1 | 0 | 0.037 | 0.037 | 0.000 | 0.000 | 0.012 | 0.006 | 0.010 | 0.010 | 0.002 | 0.001 | 0.000 | 0.000 | 1.000 | 1.000 | 0.011 | 0.052 |  |  |
|  |  | 0.5 | 0.5 | 0 | 0 | 0.3 | 0.037 | 0.029 | 0.009 | 0.009 | 0.006 | 0.041 | 0.040 | 0.040 | 0.001 | 0.002 | 0.002 | 0.002 | 1.000 | 1.000 | 0.057 | 0.064 |  |  |
|  |  | 0.3 | 0 | 0.3 | 1 | 0.3 | 0.037 | 0.029 | 0.009 | 0.009 | 0.012 | 0.040 | 0.041 | 0.041 | 0.002 | 0.002 | 0.002 | 0.002 | 1.000 | 0.093 | 1.000 | 1.000 |  |  |
|  |  | 0 | 0.3 | -0.3 | 1 | -0.3 | 0.037 | 0.046 | -0.009 | -0.010 | 0.012 | 0.041 | 0.042 | 0.042 | 0.002 | 0.004 | 0.002 | 0.002 | 0.840 | 1.000 | 1.000 | 1.000 |  |  |
|  | Egger | 0.8 | 0.5 | 0.3 | 1 | 0.3 | 0.001 | -0.007 | 0.009 | 0.009 | 0.029 | 0.037 | 0.043 | 0.043 | 0.001 | 0.001 | 0.002 | 0.002 | 1.000 | 1.000 | 1.000 | 1.000 | 0.439 | 0.052 |
|  |  | 0.2 | 0.5 | -0.3 | -1 | 0.3 | 0.000 | -0.009 | 0.009 | 0.009 | 0.026 | 0.038 | 0.044 | 0.044 | 0.001 | 0.002 | 0.002 | 0.002 | 1.000 | 1.000 | 1.000 | 1.000 | 0.805 | 0.040 |
|  |  | -0.2 | -0.5 | 0.3 | 1 | 0.3 | 0.001 | -0.011 | 0.012 | 0.012 | 0.028 | 0.038 | 0.046 | 0.046 | 0.001 | 0.002 | 0.002 | 0.002 | 1.000 | 1.000 | 1.000 | 1.000 | 0.800 | 0.047 |
|  |  | -0.8 | -0.5 | -0.3 | 1 | -0.3 | 0.002 | 0.011 | -0.010 | -0.010 | 0.029 | 0.038 | 0.043 | 0.043 | 0.001 | 0.002 | 0.002 | 0.002 | 1.000 | 1.000 | 1.000 | 1.000 | 0.677 | 0.055 |
|  |  | 0.5 | 0.5 | 0 | 1 | 0 | 0.003 | 0.001 | 0.001 | 0.001 | 0.029 | 0.014 | 0.026 | 0.026 | 0.001 | 0.000 | 0.001 | 0.001 | 1.000 | 1.000 | 0.010 | 0.065 | 0.636 | 0.050 |
|  |  | 0.5 | 0.5 | 0 | 0 | 0.3 | 0.001 | -0.008 | 0.009 | 0.009 | 0.013 | 0.038 | 0.036 | 0.036 | 0.000 | 0.002 | 0.001 | 0.001 | 1.000 | 1.000 | 0.027 | 0.073 | 0.785 | 0.045 |
|  |  | 0.3 | 0 | 0.3 | 1 | 0.3 | 0.002 | -0.008 | 0.010 | 0.010 | 0.028 | 0.038 | 0.044 | 0.044 | 0.001 | 0.002 | 0.002 | 0.002 | 1.000 | 0.038 | 1.000 | 1.000 | 0.768 | 0.044 |
|  |  | 0 | 0.3 | -0.3 | 1 | -0.3 | 0.001 | 0.010 | -0.009 | -0.009 | 0.029 | 0.038 | 0.043 | 0.043 | 0.001 | 0.002 | 0.002 | 0.002 | 0.064 | 1.000 | 1.000 | 1.000 | 0.803 | 0.050 |
| Sc32 | IVW | 0.8 | 0.5 | 0.3 | 1 | 0.3 | 0.037 | -0.024 | 0.062 | 0.062 | 0.007 | 0.027 | 0.028 | 0.028 | 0.001 | 0.001 | 0.005 | 0.005 | 1.000 | 1.000 | 1.000 | 1.000 |  |  |
|  |  | 0.2 | 0.5 | -0.3 | -1 | 0.3 | -0.037 | -0.025 | -0.012 | -0.012 | 0.011 | 0.111 | 0.111 | 0.111 | 0.001 | 0.013 | 0.012 | 0.012 | 1.000 | 0.984 | 0.801 | 0.802 |  |  |
|  |  | -0.2 | -0.5 | 0.3 | 1 | 0.3 | 0.038 | -0.024 | 0.062 | 0.062 | 0.012 | 0.109 | 0.109 | 0.109 | 0.002 | 0.013 | 0.016 | 0.016 | 1.000 | 0.994 | 0.896 | 0.900 |  |  |
|  |  | -0.8 | -0.5 | -0.3 | 1 | -0.3 | 0.038 | 0.021 | 0.017 | 0.017 | 0.012 | 0.084 | 0.084 | 0.084 | 0.002 | 0.007 | 0.007 | 0.007 | 1.000 | 1.000 | 0.895 | 0.899 |  |  |
|  |  | 0.5 | 0.5 | 0 | 1 | 0 | 0.038 | -0.002 | 0.040 | 0.040 | 0.012 | 0.016 | 0.014 | 0.014 | 0.002 | 0.000 | 0.002 | 0.002 | 1.000 | 1.000 | 0.536 | 0.906 |  |  |
|  |  | 0.5 | 0.5 | 0 | 0 | 0.3 | 0.000 | -0.025 | 0.025 | 0.025 | 0.010 | 0.111 | 0.110 | 0.110 | 0.000 | 0.013 | 0.013 | 0.013 | 1.000 | 0.976 | 0.063 | 0.070 |  |  |
|  |  | 0.3 | 0 | 0.3 | 1 | 0.3 | 0.038 | -0.024 | 0.062 | 0.062 | 0.012 | 0.108 | 0.108 | 0.108 | 0.002 | 0.012 | 0.015 | 0.015 | 1.000 | 0.053 | 0.895 | 0.893 |  |  |
|  |  | 0 | 0.3 | -0.3 | 1 | -0.3 | 0.037 | 0.017 | 0.020 | 0.020 | 0.012 | 0.087 | 0.088 | 0.088 | 0.001 | 0.008 | 0.008 | 0.008 | 0.861 | 0.947 | 0.897 | 0.904 |  |  |
|  | Egger | 0.8 | 0.5 | 0.3 | 1 | 0.3 | 0.002 | -0.031 | 0.032 | 0.032 | 0.016 | 0.029 | 0.032 | 0.032 | 0.000 | 0.002 | 0.002 | 0.002 | 1.000 | 1.000 | 0.998 | 0.998 | 0.000 | 0.794 |
|  |  | 0.2 | 0.5 | -0.3 | -1 | 0.3 | 0.002 | -0.033 | 0.035 | 0.035 | 0.028 | 0.120 | 0.118 | 0.119 | 0.001 | 0.016 | 0.015 | 0.015 | 1.000 | 0.962 | 0.592 | 0.633 | 0.054 | 0.835 |
|  |  | -0.2 | -0.5 | 0.3 | 1 | 0.3 | 0.000 | -0.029 | 0.029 | 0.029 | 0.028 | 0.119 | 0.119 | 0.119 | 0.001 | 0.015 | 0.015 | 0.015 | 1.000 | 0.985 | 0.769 | 0.793 | 0.059 | 0.845 |
|  |  | -0.8 | -0.5 | -0.3 | 1 | -0.3 | 0.001 | 0.035 | -0.033 | -0.034 | 0.029 | 0.121 | 0.119 | 0.119 | 0.001 | 0.016 | 0.015 | 0.015 | 1.000 | 0.962 | 0.793 | 0.819 | 0.052 | 0.828 |
|  |  | 0.5 | 0.5 | 0 | 1 | 0 | 0.003 | 0.002 | 0.001 | 0.001 | 0.029 | 0.026 | 0.016 | 0.016 | 0.001 | 0.001 | 0.000 | 0.000 | 1.000 | 1.000 | 0.001 | 0.038 | 0.043 | 0.801 |
|  |  | 0.5 | 0.5 | 0 | 0 | 0.3 | -0.001 | -0.028 | 0.027 | 0.027 | 0.025 | 0.122 | 0.121 | 0.121 | 0.001 | 0.016 | 0.015 | 0.015 | 1.000 | 0.961 | 0.061 | 0.080 | 0.047 | 0.814 |
|  |  | 0.3 | 0 | 0.3 | 1 | 0.3 | 0.001 | -0.030 | 0.031 | 0.031 | 0.029 | 0.118 | 0.117 | 0.117 | 0.001 | 0.015 | 0.015 | 0.015 | 1.000 | 0.061 | 0.789 | 0.811 | 0.072 | 0.815 |
|  |  | 0 | 0.3 | -0.3 | 1 | -0.3 | 0.001 | 0.028 | -0.027 | -0.027 | 0.029 | 0.118 | 0.117 | 0.117 | 0.001 | 0.015 | 0.015 | 0.015 | 0.047 | 0.759 | 0.782 | 0.805 | 0.047 | 0.818 |
| Sc4 | IVW | 0.8 | 0.5 | 0.3 | 1 | 0.3 | 0.247 | -0.258 | 0.505 | 0.505 | 0.027 | 0.038 | 0.045 | 0.045 | 0.062 | 0.068 | 0.257 | 0.258 | 1.000 | 0.996 | 1.000 | 1.000 |  |  |
|  |  | 0.2 | 0.5 | -0.3 | -1 | 0.3 | 0.000 | -0.256 | 0.256 | 0.256 | 0.007 | 0.039 | 0.039 | 0.039 | 0.000 | 0.067 | 0.067 | 0.067 | 1.000 | 0.999 | 0.218 | 0.243 |  |  |
|  |  | -0.2 | -0.5 | 0.3 | 1 | 0.3 | 0.248 | -0.257 | 0.505 | 0.505 | 0.026 | 0.038 | 0.046 | 0.046 | 0.062 | 0.068 | 0.257 | 0.257 | 0.323 | 1.000 | 1.000 | 1.000 |  |  |
|  |  | -0.8 | -0.5 | -0.3 | 1 | -0.3 | 0.244 | 0.282 | -0.038 | -0.037 | 0.026 | 0.017 | 0.030 | 0.030 | 0.060 | 0.080 | 0.002 | 0.002 | 1.000 | 1.000 | 1.000 | 1.000 |  |  |
|  |  | 0.5 | 0.5 | 0 | 1 | 0 | 0.248 | 0.012 | 0.236 | 0.236 | 0.027 | 0.013 | 0.029 | 0.029 | 0.062 | 0.000 | 0.056 | 0.057 | 1.000 | 1.000 | 1.000 | 1.000 |  |  |
|  |  | 0.5 | 0.5 | 0 | 0 | 0.3 | 0.124 | -0.257 | 0.380 | 0.381 | 0.013 | 0.038 | 0.040 | 0.040 | 0.016 | 0.067 | 0.146 | 0.147 | 1.000 | 1.000 | 1.000 | 1.000 |  |  |
|  |  | 0.3 | 0 | 0.3 | 1 | 0.3 | 0.248 | -0.259 | 0.507 | 0.506 | 0.026 | 0.039 | 0.046 | 0.046 | 0.062 | 0.068 | 0.259 | 0.258 | 1.000 | 1.000 | 1.000 | 1.000 |  |  |
|  |  | 0 | 0.3 | -0.3 | 1 | -0.3 | 0.248 | 0.282 | -0.034 | -0.036 | 0.025 | 0.017 | 0.029 | 0.029 | 0.062 | 0.080 | 0.002 | 0.002 | 1.000 | 1.000 | 1.000 | 1.000 |  |  |
|  | Egger | 0.8 | 0.5 | 0.3 | 1 | 0.3 | 0.038 | -0.252 | 0.290 | 0.290 | 0.061 | 0.039 | 0.068 | 0.068 | 0.005 | 0.065 | 0.089 | 0.089 | 1.000 | 0.995 | 1.000 | 1.000 | 0.012 | 0.935 |
|  |  | 0.2 | 0.5 | -0.3 | -1 | 0.3 | 0.001 | -0.250 | 0.251 | 0.251 | 0.018 | 0.039 | 0.036 | 0.036 | 0.000 | 0.064 | 0.064 | 0.064 | 1.000 | 0.999 | 0.166 | 0.295 | 0.104 | 0.923 |
|  |  | -0.2 | -0.5 | 0.3 | 1 | 0.3 | 0.041 | -0.252 | 0.293 | 0.294 | 0.062 | 0.039 | 0.069 | 0.069 | 0.005 | 0.065 | 0.091 | 0.091 | 0.696 | 1.000 | 1.000 | 1.000 | 0.110 | 0.928 |
|  |  | -0.8 | -0.5 | -0.3 | 1 | -0.3 | 0.039 | 0.259 | -0.220 | -0.220 | 0.061 | 0.035 | 0.069 | 0.069 | 0.005 | 0.068 | 0.053 | 0.053 | 1.000 | 0.999 | 1.000 | 1.000 | 0.099 | 0.918 |
|  |  | 0.5 | 0.5 | 0 | 1 | 0 | 0.038 | 0.003 | 0.035 | 0.035 | 0.062 | 0.018 | 0.059 | 0.059 | 0.005 | 0.000 | 0.005 | 0.005 | 1.000 | 1.000 | 0.080 | 0.118 | 0.065 | 0.927 |
|  |  | 0.5 | 0.5 | 0 | 0 | 0.3 | 0.021 | -0.251 | 0.271 | 0.271 | 0.030 | 0.039 | 0.042 | 0.042 | 0.001 | 0.064 | 0.075 | 0.075 | 1.000 | 1.000 | 0.999 | 0.999 | 0.112 | 0.927 |
|  |  | 0.3 | 0 | 0.3 | 1 | 0.3 | 0.038 | -0.253 | 0.291 | 0.291 | 0.060 | 0.040 | 0.067 | 0.067 | 0.005 | 0.066 | 0.089 | 0.089 | 0.997 | 0.999 | 1.000 | 1.000 | 0.095 | 0.934 |
|  |  | 0 | 0.3 | -0.3 | 1 | -0.3 | 0.041 | 0.257 | -0.216 | -0.217 | 0.062 | 0.037 | 0.067 | 0.067 | 0.006 | 0.067 | 0.051 | 0.052 | 0.112 | 1.000 | 1.000 | 1.000 | 0.143 | 0.917 |

TE: total effect; DE: direct effect; IE: indirect effect; IE_d: indirect effect calculated by difference method; IE_p: indirect effect calculated by product method.

#### 3 Simulation C: Causally non-ordered mediators PSE-IVW in two mediators settings

The data-generating models were

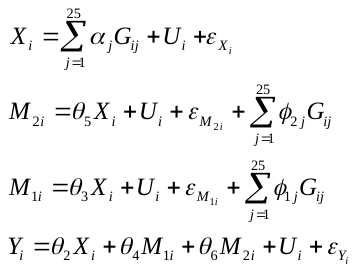

Error variables
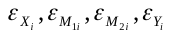
 were independently generated from a
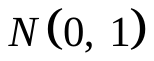
 distribution; and instrument strength parameters
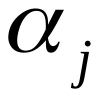
 were generated from a
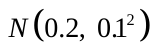
 distribution. The
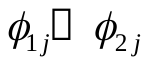
 parameters were generated from a normal distribution
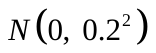
, independently. These effects were included in the simulation model to ensure that the direct and indirect effect was identified.

We also varied the sample size, the number of instrumental variables and simulated different sets of values of the parameters. We obtained similar performance with Single-mediator IVW. Results were listed in **eTable 6-7**.

**eTable 6**. Simulation of PSE-IVW in two non-ordered mediators setting with different numbers of genetic variants.

|  | BIAS | | | | | | SE | | | | | | MSE | | | | | | Power | | | | | |
| --- | --- | --- | --- | --- | --- | --- | --- | --- | --- | --- | --- | --- | --- | --- | --- | --- | --- | --- | --- | --- | --- | --- | --- | --- |
| g | TE | DE | IE_d | IE1 | IE2 | IE_p | TE | DE | IE_d | IE1 | IE2 | IE_p | TE | DE | IE_d | IE1 | IE2 | IE_p | TE | DE | IE_d | IE1 | IE2 | IE_p |
| 5 | 0.000 | -0.044 | 0.044 | 0.027 | 0.018 | 0.044 | 0.154 | 0.197 | 0.244 | 0.200 | 0.142 | 0.244 | 0.024 | 0.041 | 0.061 | 0.041 | 0.020 | 0.061 | 1.000 | 0.000 | 0.266 | 0.125 | 0.073 | 0.291 |
| 10 | 0.005 | -0.048 | 0.053 | 0.030 | 0.023 | 0.053 | 0.107 | 0.086 | 0.133 | 0.100 | 0.086 | 0.133 | 0.011 | 0.010 | 0.020 | 0.011 | 0.008 | 0.020 | 1.000 | 0.601 | 0.864 | 0.630 | 0.620 | 0.902 |
| 15 | 0.008 | -0.047 | 0.055 | 0.034 | 0.021 | 0.055 | 0.087 | 0.063 | 0.107 | 0.080 | 0.069 | 0.107 | 0.008 | 0.006 | 0.014 | 0.008 | 0.005 | 0.014 | 1.000 | 0.944 | 0.993 | 0.919 | 0.935 | 0.997 |
| 20 | 0.009 | -0.046 | 0.055 | 0.035 | 0.020 | 0.055 | 0.075 | 0.053 | 0.089 | 0.068 | 0.056 | 0.089 | 0.006 | 0.005 | 0.011 | 0.006 | 0.003 | 0.011 | 1.000 | 1.000 | 1.000 | 0.995 | 0.991 | 1.000 |
| 30 | 0.011 | -0.047 | 0.058 | 0.037 | 0.020 | 0.058 | 0.061 | 0.041 | 0.071 | 0.055 | 0.046 | 0.071 | 0.004 | 0.004 | 0.008 | 0.004 | 0.003 | 0.008 | 1.000 | 1.000 | 1.000 | 1.000 | 1.000 | 1.000 |
| 50 | 0.009 | -0.045 | 0.054 | 0.033 | 0.022 | 0.054 | 0.048 | 0.031 | 0.055 | 0.042 | 0.035 | 0.055 | 0.002 | 0.003 | 0.006 | 0.003 | 0.002 | 0.006 | 1.000 | 1.000 | 1.000 | 1.000 | 1.000 | 1.000 |
| 100 | 0.009 | -0.046 | 0.055 | 0.034 | 0.021 | 0.055 | 0.034 | 0.021 | 0.039 | 0.029 | 0.025 | 0.039 | 0.001 | 0.003 | 0.005 | 0.002 | 0.001 | 0.005 | 1.000 | 1.000 | 1.000 | 1.000 | 1.000 | 1.000 |
| 150 | 0.009 | -0.046 | 0.055 | 0.033 | 0.022 | 0.055 | 0.027 | 0.018 | 0.032 | 0.024 | 0.021 | 0.032 | 0.001 | 0.002 | 0.004 | 0.002 | 0.001 | 0.004 | 1.000 | 1.000 | 1.000 | 1.000 | 1.000 | 1.000 |
| 3 | -0.001 | -0.037 | 0.036 | 0.110 | -0.074 | 0.036 | 0.199 | 7.048 | 7.059 | 8.644 | 6.803 | 7.061 | 0.040 | 49.670 | 49.833 | 74.725 | 46.290 | 49.859 | 0.958 | NA | NA | NA | NA | NA |

TE: total effect; DE: direct effect; IE: indirect effect; IE_d: indirect effect calculated by difference method; IE_p: indirect effect calculated by product method; IE1: *X*→*M_1_*→*Y*; IE2: *X*→*M_2_*→*Y*.

**eTable 7.** Simulation of PSE-IVW in two non-ordered mediators setting with different sample sizes.

|  | BIAS | | | | | | SE | | | | | | MSE | | | | | | Power | | | | | |
| --- | --- | --- | --- | --- | --- | --- | --- | --- | --- | --- | --- | --- | --- | --- | --- | --- | --- | --- | --- | --- | --- | --- | --- | --- |
| N | TE | DE | IE_d | IE1 | IE2 | IE_p | TE | DE | IE_d | IE1 | IE2 | IE_p | TE | DE | IE_d | IE1 | IE2 | IE_p | TE | DE | IE_d | IE1 | IE2 | IE_p |
| 2000 | 0.071 | -0.172 | 0.243 | 0.143 | 0.100 | 0.243 | 0.129 | 0.095 | 0.150 | 0.113 | 0.092 | 0.150 | 0.022 | 0.039 | 0.082 | 0.033 | 0.018 | 0.082 | 1.000 | 0.108 | 0.926 | 0.450 | 0.456 | 0.955 |
| 4000 | 0.045 | -0.133 | 0.178 | 0.105 | 0.073 | 0.179 | 0.099 | 0.079 | 0.122 | 0.090 | 0.077 | 0.122 | 0.012 | 0.024 | 0.047 | 0.019 | 0.011 | 0.047 | 1.000 | 0.553 | 0.986 | 0.761 | 0.733 | 0.995 |
| 6000 | 0.029 | -0.106 | 0.135 | 0.083 | 0.052 | 0.136 | 0.090 | 0.072 | 0.111 | 0.084 | 0.070 | 0.111 | 0.009 | 0.016 | 0.031 | 0.014 | 0.008 | 0.031 | 1.000 | 0.838 | 0.995 | 0.890 | 0.877 | 0.998 |
| 8000 | 0.018 | -0.089 | 0.108 | 0.067 | 0.041 | 0.108 | 0.079 | 0.064 | 0.094 | 0.074 | 0.060 | 0.094 | 0.007 | 0.012 | 0.020 | 0.010 | 0.005 | 0.020 | 1.000 | 0.947 | 0.998 | 0.940 | 0.940 | 1.000 |
| 10000 | 0.019 | -0.079 | 0.098 | 0.060 | 0.038 | 0.098 | 0.080 | 0.059 | 0.097 | 0.073 | 0.060 | 0.097 | 0.007 | 0.010 | 0.019 | 0.009 | 0.005 | 0.019 | 1.000 | 0.981 | 1.000 | 0.984 | 0.968 | 1.000 |
| 12000 | 0.011 | -0.069 | 0.080 | 0.050 | 0.030 | 0.080 | 0.076 | 0.055 | 0.089 | 0.068 | 0.055 | 0.089 | 0.006 | 0.008 | 0.014 | 0.007 | 0.004 | 0.014 | 1.000 | 0.992 | 1.000 | 0.991 | 0.986 | 1.000 |
| 14000 | 0.014 | -0.059 | 0.073 | 0.045 | 0.028 | 0.073 | 0.071 | 0.054 | 0.083 | 0.063 | 0.052 | 0.083 | 0.005 | 0.006 | 0.012 | 0.006 | 0.003 | 0.012 | 1.000 | 0.999 | 1.000 | 0.992 | 0.996 | 1.000 |
| 16000 | 0.009 | -0.054 | 0.063 | 0.039 | 0.024 | 0.063 | 0.071 | 0.051 | 0.085 | 0.064 | 0.051 | 0.085 | 0.005 | 0.005 | 0.011 | 0.006 | 0.003 | 0.011 | 1.000 | 1.000 | 1.000 | 1.000 | 0.996 | 1.000 |
| 18000 | 0.009 | -0.049 | 0.058 | 0.038 | 0.020 | 0.058 | 0.070 | 0.051 | 0.081 | 0.061 | 0.052 | 0.081 | 0.005 | 0.005 | 0.010 | 0.005 | 0.003 | 0.010 | 1.000 | 1.000 | 1.000 | 0.999 | 0.997 | 1.000 |
| 20000 | 0.014 | -0.047 | 0.061 | 0.037 | 0.025 | 0.061 | 0.069 | 0.047 | 0.083 | 0.062 | 0.052 | 0.083 | 0.005 | 0.004 | 0.011 | 0.005 | 0.003 | 0.011 | 1.000 | 1.000 | 1.000 | 0.999 | 1.000 | 1.000 |

TE: total effect; DE: direct effect; IE: indirect effect; IE_d: indirect effect calculated by difference method; IE_p: indirect effect calculated by product method; IE1: *X*→*M_1_*→*Y*; IE2: *X*→*M_2_*→*Y*.

#### 4 Simulation D: Causally non-ordered mediators PSE-Egger in two mediators settings

The data-generating models were

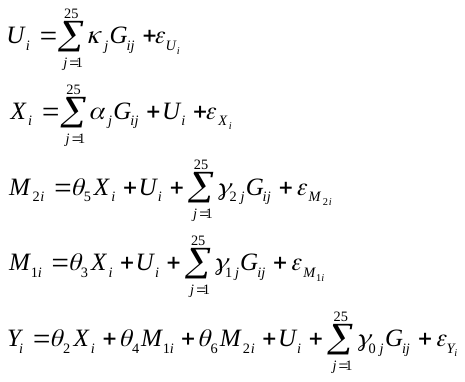

Error variables
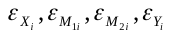
 were independently generated from a
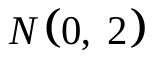
 distribution; and instrument strength parameters
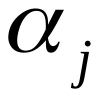
 were generated from a
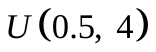
 distribution. This model additionally allowed genetic variant
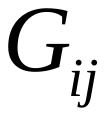
 to be associated with confounders variable
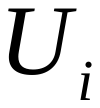
. This occurred when
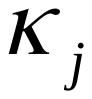
 was non-zero (a violation of the **Assumption Ⅱ**). Simulation scenarios (a)–(c) were implemented by additionally specifying values for
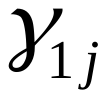
 ( the direct effect of genetic variables
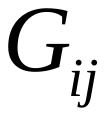
 on the mediator
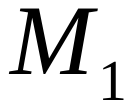
),
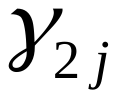
 ( the direct effect of genetic variables
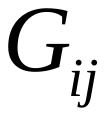
 on the mediator
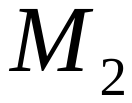
) and
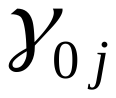
 ( the direct effect of genetic variables
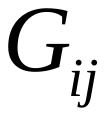
 on outcome *Y*) as below:

**eTable 8.** Settings for simulation of PSE-Egger in two non-ordered mediators setting

|  | Pleiotropy | InSIDE | 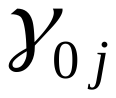 | 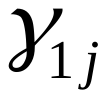 | 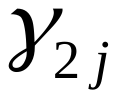 | 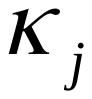 |  |
| --- | --- | --- | --- | --- | --- | --- | --- |
| case(a) | Balanced | Satisfied | 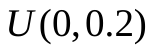 | 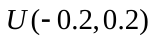 | 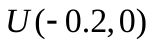 | 0 | sc21 |
|  |  |  | 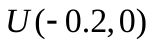 | 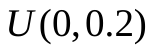 | 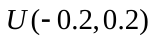 | 0 | sc22 |
|  |  |  | 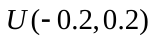 | 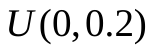 | 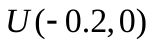 | 0 | sc23 |
|  |  |  | 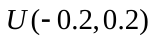 | 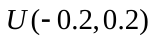 | 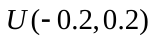 | 0 | sc2 |
| case(b) | Directional | Satisfied | 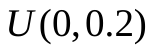 | 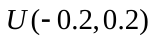 | 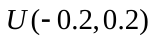 | 0 | sc31 |
|  |  |  | 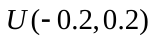 | 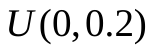 | 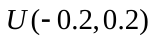 | 0 | sc32 |
|  |  |  | 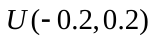 | 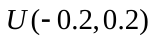 | 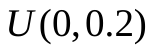 | 0 | sc33 |
|  |  |  | 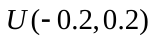 | 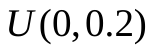 | 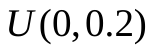 | 0 | sc34 |
|  |  |  | 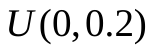 | 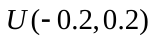 | 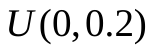 | 0 | sc35 |
|  |  |  | 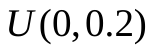 | 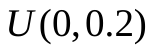 | 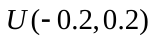 | 0 | sc36 |
|  |  |  | 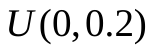 | 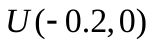 | 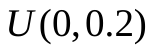 | 0 | sc37 |
|  |  |  | 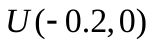 | 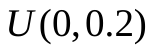 |  | 0 | sc38 |
|  |  |  |  |  |  | 0 | sc39 |
|  |  |  |  |  |  | 0 | sc3 |
| case(c) | Directional | Not satisfied |  |  |  |  | sc4 |

The performance was similar with Single-mediator MR-Egger. Results were listed in **eTable 9-12**.

**eTable 9.** Simulation of PSE-Egger in two non-ordered mediators setting with different effects (case(a)).

|  |  | TRUE | | | | | BIAS | | | | | | SE | | | | | | MSE | | | | | | Power/Type I error | | | | | | | | |
| --- | --- | --- | --- | --- | --- | --- | --- | --- | --- | --- | --- | --- | --- | --- | --- | --- | --- | --- | --- | --- | --- | --- | --- | --- | --- | --- | --- | --- | --- | --- | --- | --- | --- |
|  |  | TE | DE | IE | IE2 | IE1 | TE | DE | IE_d | IE1 | IE2 | IE_p | TE | DE | IE_d | IE1 | IE2 | IE_p | TE | DE | IE_d | IE1 | IE2 | IE_p | TE | DE | IE_d | IE1 | IE2 | IE_p | GY | GM1 | GM2 |
| sc2 | IVW | 1.6 | 0.8 | 0.8 | 0.5 | 0.3 | 0.001 | -0.019 | 0.019 | 0.014 | 0.006 | 0.020 | 0.018 | 0.124 | 0.126 | 0.107 | 0.066 | 0.126 | 0.000 | 0.016 | 0.016 | 0.012 | 0.004 | 0.016 | 1.000 | 0.996 | 0.999 | 0.962 | 0.957 | 0.997 |  |  |  |
|  |  | 0.6 | 0.8 | -0.2 | -0.5 | 0.3 | 0.000 | -0.028 | 0.028 | 0.020 | 0.008 | 0.028 | 0.016 | 0.125 | 0.125 | 0.111 | 0.063 | 0.125 | 0.000 | 0.016 | 0.016 | 0.013 | 0.004 | 0.016 | 1.000 | 1.000 | 0.291 | 0.989 | 0.997 | 0.295 |  |  |  |
|  |  | 0.7 | 0.5 | 0.2 | 0.5 | -0.3 | 0.000 | -0.020 | 0.020 | 0.011 | 0.009 | 0.020 | 0.017 | 0.122 | 0.123 | 0.108 | 0.065 | 0.123 | 0.000 | 0.015 | 0.015 | 0.012 | 0.004 | 0.015 | 1.000 | 0.960 | 0.439 | 0.992 | 0.997 | 0.425 |  |  |  |
|  |  | -0.3 | 0.5 | -0.8 | -0.5 | -0.3 | -0.001 | -0.028 | 0.027 | 0.016 | 0.011 | 0.027 | 0.017 | 0.127 | 0.128 | 0.106 | 0.064 | 0.128 | 0.000 | 0.017 | 0.017 | 0.012 | 0.004 | 0.017 | 1.000 | 0.950 | 1.000 | 0.994 | 0.992 | 1.000 |  |  |  |
|  |  | 0 | -0.8 | 0.8 | 0.5 | 0.3 | 0.000 | -0.032 | 0.031 | 0.018 | 0.013 | 0.032 | 0.018 | 0.124 | 0.124 | 0.109 | 0.066 | 0.124 | 0.000 | 0.016 | 0.016 | 0.012 | 0.004 | 0.017 | 0.056 | 1.000 | 1.000 | 0.994 | 0.998 | 1.000 |  |  |  |
|  |  | -1.6 | -0.8 | -0.8 | -0.5 | -0.3 | -0.001 | -0.020 | 0.020 | 0.016 | 0.004 | 0.019 | 0.017 | 0.124 | 0.124 | 0.104 | 0.066 | 0.124 | 0.000 | 0.016 | 0.016 | 0.011 | 0.004 | 0.016 | 1.000 | 0.999 | 0.999 | 0.976 | 0.978 | 1.000 |  |  |  |
|  |  | 0.8 | 0 | 0.8 | 0.5 | 0.3 | 0.001 | -0.029 | 0.030 | 0.019 | 0.012 | 0.031 | 0.018 | 0.121 | 0.122 | 0.109 | 0.064 | 0.122 | 0.000 | 0.015 | 0.016 | 0.012 | 0.004 | 0.016 | 1.000 | 0.045 | 1.000 | 0.994 | 0.998 | 1.000 |  |  |  |
|  |  | 1.3 | 0.8 | 0.5 | 0.5 | 0 | 0.000 | -0.020 | 0.021 | 0.014 | 0.007 | 0.021 | 0.014 | 0.123 | 0.123 | 0.109 | 0.062 | 0.123 | 0.000 | 0.016 | 0.016 | 0.012 | 0.004 | 0.016 | 1.000 | 0.999 | 0.971 | 0.988 | 0.021 | 0.973 |  |  |  |
|  |  | 1.3 | 0.8 | 0.5 | 0.5 | 0 | 0.000 | -0.018 | 0.018 | 0.019 | 0.000 | 0.018 | 0.018 | 0.105 | 0.104 | 0.104 | 0.010 | 0.104 | 0.000 | 0.011 | 0.011 | 0.011 | 0.000 | 0.011 | 1.000 | 0.999 | 0.982 | 0.982 | 0.070 | 0.982 |  |  |  |
|  |  | 0.8 | 0.8 | 0 | 0 | 0 | 0.000 | -0.023 | 0.023 | 0.014 | 0.009 | 0.023 | 0.010 | 0.120 | 0.120 | 0.105 | 0.064 | 0.120 | 0.000 | 0.015 | 0.015 | 0.011 | 0.004 | 0.015 | 1.000 | 1.000 | 0.065 | 0.059 | 0.057 | 0.065 |  |  |  |
|  | Egger | 1.6 | 0.8 | 0.8 | 0.5 | 0.3 | 0.003 | -0.018 | 0.020 | 0.014 | 0.006 | 0.020 | 0.043 | 0.129 | 0.133 | 0.113 | 0.073 | 0.133 | 0.002 | 0.017 | 0.018 | 0.013 | 0.005 | 0.018 | 1.000 | 0.991 | 0.997 | 0.956 | 0.940 | 0.996 | 0.002 | 0.063 | 0.049 |
|  |  | 0.6 | 0.8 | -0.2 | -0.5 | 0.3 | -0.002 | -0.030 | 0.027 | 0.018 | 0.009 | 0.027 | 0.042 | 0.132 | 0.134 | 0.116 | 0.071 | 0.134 | 0.002 | 0.018 | 0.019 | 0.014 | 0.005 | 0.019 | 1.000 | 0.999 | 0.256 | 0.985 | 0.993 | 0.281 | 0.053 | 0.057 | 0.043 |
|  |  | 0.7 | 0.5 | 0.2 | 0.5 | -0.3 | -0.001 | -0.022 | 0.021 | 0.012 | 0.009 | 0.021 | 0.042 | 0.126 | 0.129 | 0.113 | 0.070 | 0.129 | 0.002 | 0.016 | 0.017 | 0.013 | 0.005 | 0.017 | 1.000 | 0.947 | 0.389 | 0.989 | 0.995 | 0.393 | 0.051 | 0.060 | 0.054 |
|  |  | -0.3 | 0.5 | -0.8 | -0.5 | -0.3 | -0.001 | -0.028 | 0.027 | 0.016 | 0.011 | 0.027 | 0.041 | 0.133 | 0.134 | 0.110 | 0.071 | 0.134 | 0.002 | 0.018 | 0.019 | 0.012 | 0.005 | 0.019 | 1.000 | 0.931 | 1.000 | 0.990 | 0.987 | 1.000 | 0.040 | 0.044 | 0.036 |
|  |  | 0 | -0.8 | 0.8 | 0.5 | 0.3 | 0.003 | -0.031 | 0.034 | 0.021 | 0.014 | 0.034 | 0.044 | 0.129 | 0.132 | 0.113 | 0.072 | 0.132 | 0.002 | 0.018 | 0.019 | 0.013 | 0.005 | 0.019 | 0.042 | 1.000 | 1.000 | 0.994 | 0.996 | 1.000 | 0.040 | 0.050 | 0.043 |
|  |  | -1.6 | -0.8 | -0.8 | -0.5 | -0.3 | -0.001 | -0.021 | 0.020 | 0.016 | 0.004 | 0.019 | 0.043 | 0.130 | 0.133 | 0.111 | 0.071 | 0.133 | 0.002 | 0.017 | 0.018 | 0.013 | 0.005 | 0.018 | 1.000 | 0.998 | 0.999 | 0.966 | 0.966 | 1.000 | 0.007 | 0.054 | 0.051 |
|  |  | 0.8 | 0 | 0.8 | 0.5 | 0.3 | 0.002 | -0.031 | 0.032 | 0.020 | 0.012 | 0.032 | 0.043 | 0.127 | 0.131 | 0.114 | 0.069 | 0.131 | 0.002 | 0.017 | 0.018 | 0.013 | 0.005 | 0.018 | 1.000 | 0.034 | 1.000 | 0.993 | 0.995 | 1.000 | 0.036 | 0.031 | 0.046 |
|  |  | 1.3 | 0.8 | 0.5 | 0.5 | 0 | 0.002 | -0.019 | 0.021 | 0.014 | 0.007 | 0.021 | 0.035 | 0.128 | 0.128 | 0.115 | 0.064 | 0.128 | 0.001 | 0.017 | 0.017 | 0.013 | 0.004 | 0.017 | 1.000 | 0.998 | 0.958 | 0.985 | 0.016 | 0.965 | 0.014 | 0.040 | 0.055 |
|  |  | 1.3 | 0.8 | 0.5 | 0.5 | 0 | 0.001 | -0.017 | 0.018 | 0.019 | 0.000 | 0.018 | 0.044 | 0.109 | 0.111 | 0.110 | 0.024 | 0.111 | 0.002 | 0.012 | 0.013 | 0.012 | 0.001 | 0.013 | 1.000 | 0.999 | 0.973 | 0.979 | 0.049 | 0.974 | 0.009 | 0.036 | 0.056 |
|  |  | 0.8 | 0.8 | 0 | 0 | 0 | 0.001 | -0.022 | 0.022 | 0.013 | 0.009 | 0.022 | 0.025 | 0.127 | 0.125 | 0.109 | 0.066 | 0.125 | 0.001 | 0.017 | 0.016 | 0.012 | 0.004 | 0.016 | 1.000 | 1.000 | 0.057 | 0.066 | 0.059 | 0.065 | 0.051 | 0.047 | 0.059 |
| sc21 | IVW | 1.6 | 0.8 | 0.8 | 0.5 | 0.3 | 0.001 | 0.120 | -0.119 | -0.129 | 0.010 | -0.119 | 0.013 | 0.101 | 0.101 | 0.096 | 0.040 | 0.101 | 0.000 | 0.025 | 0.025 | 0.026 | 0.002 | 0.025 | 1.000 | 0.996 | 0.957 | 0.287 | 0.990 | 0.950 |  |  |  |
|  |  | 0.6 | 0.8 | -0.2 | -0.5 | 0.3 | 0.074 | 0.123 | -0.048 | -0.058 | 0.010 | -0.048 | 0.014 | 0.103 | 0.103 | 0.096 | 0.042 | 0.103 | 0.006 | 0.026 | 0.013 | 0.013 | 0.002 | 0.013 | 1.000 | 1.000 | 0.616 | 0.998 | 1.000 | 0.611 |  |  |  |
|  |  | 0.7 | 0.5 | 0.2 | 0.5 | -0.3 | 0.001 | 0.123 | -0.122 | -0.135 | 0.012 | -0.123 | 0.012 | 0.104 | 0.105 | 0.097 | 0.041 | 0.105 | 0.000 | 0.026 | 0.026 | 0.028 | 0.002 | 0.026 | 1.000 | 0.998 | 0.083 | 0.939 | 1.000 | 0.082 |  |  |  |
|  |  | -0.3 | 0.5 | -0.8 | -0.5 | -0.3 | 0.074 | 0.125 | -0.051 | -0.061 | 0.010 | -0.051 | 0.013 | 0.104 | 0.105 | 0.097 | 0.041 | 0.105 | 0.006 | 0.027 | 0.014 | 0.013 | 0.002 | 0.014 | 1.000 | 0.998 | 1.000 | 0.999 | 0.999 | 1.000 |  |  |  |
|  |  | 0 | -0.8 | 0.8 | 0.5 | 0.3 | 0.001 | 0.117 | -0.116 | -0.127 | 0.011 | -0.116 | 0.013 | 0.101 | 0.101 | 0.093 | 0.040 | 0.101 | 0.000 | 0.024 | 0.024 | 0.025 | 0.002 | 0.024 | 0.050 | 1.000 | 1.000 | 0.954 | 1.000 | 1.000 |  |  |  |
|  |  | -1.6 | -0.8 | -0.8 | -0.5 | -0.3 | 0.073 | 0.120 | -0.047 | -0.060 | 0.013 | -0.047 | 0.013 | 0.103 | 0.102 | 0.096 | 0.041 | 0.102 | 0.006 | 0.025 | 0.013 | 0.013 | 0.002 | 0.013 | 1.000 | 0.968 | 1.000 | 0.917 | 0.998 | 0.999 |  |  |  |
|  |  | 0.8 | 0 | 0.8 | 0.5 | 0.3 | 0.001 | 0.121 | -0.120 | -0.132 | 0.012 | -0.120 | 0.013 | 0.097 | 0.098 | 0.093 | 0.041 | 0.098 | 0.000 | 0.024 | 0.024 | 0.026 | 0.002 | 0.024 | 1.000 | 0.020 | 0.995 | 0.804 | 1.000 | 0.996 |  |  |  |
|  |  | 1.3 | 0.8 | 0.5 | 0.5 | 0 | 0.001 | 0.118 | -0.117 | -0.132 | 0.015 | -0.117 | 0.008 | 0.101 | 0.101 | 0.093 | 0.040 | 0.101 | 0.000 | 0.024 | 0.024 | 0.026 | 0.002 | 0.024 | 1.000 | 1.000 | 0.548 | 0.589 | 0.001 | 0.524 |  |  |  |
|  |  | 1.3 | 0.8 | 0.5 | 0.5 | 0 | 0.000 | 0.132 | -0.132 | -0.133 | 0.000 | -0.133 | 0.013 | 0.094 | 0.095 | 0.094 | 0.010 | 0.095 | 0.000 | 0.026 | 0.026 | 0.026 | 0.000 | 0.027 | 1.000 | 0.999 | 0.455 | 0.468 | 0.071 | 0.471 |  |  |  |
|  |  | 0.8 | 0.8 | 0 | 0 | 0 | 0.037 | 0.125 | -0.088 | -0.098 | 0.010 | -0.088 | 0.006 | 0.103 | 0.102 | 0.097 | 0.040 | 0.102 | 0.001 | 0.026 | 0.018 | 0.019 | 0.002 | 0.018 | 1.000 | 1.000 | 0.076 | 0.107 | 0.028 | 0.070 |  |  |  |
|  | Egger | 1.6 | 0.8 | 0.8 | 0.5 | 0.3 | 0.003 | -0.060 | 0.063 | 0.055 | 0.008 | 0.063 | 0.033 | 0.113 | 0.116 | 0.111 | 0.043 | 0.116 | 0.001 | 0.016 | 0.017 | 0.015 | 0.002 | 0.017 | 1.000 | 0.806 | 0.957 | 0.489 | 0.992 | 0.957 | 0.005 | 0.050 | 0.833 |
|  |  | 0.6 | 0.8 | -0.2 | -0.5 | 0.3 | 0.001 | -0.055 | 0.056 | 0.049 | 0.007 | 0.056 | 0.030 | 0.116 | 0.119 | 0.111 | 0.044 | 0.119 | 0.001 | 0.017 | 0.017 | 0.015 | 0.002 | 0.017 | 1.000 | 1.000 | 0.151 | 0.948 | 1.000 | 0.140 | 0.570 | 0.050 | 0.817 |
|  |  | 0.7 | 0.5 | 0.2 | 0.5 | -0.3 | 0.000 | -0.060 | 0.060 | 0.050 | 0.009 | 0.060 | 0.030 | 0.116 | 0.118 | 0.111 | 0.042 | 0.118 | 0.001 | 0.017 | 0.018 | 0.015 | 0.002 | 0.018 | 1.000 | 0.880 | 0.451 | 0.989 | 1.000 | 0.446 | 0.567 | 0.050 | 0.824 |
|  |  | -0.3 | 0.5 | -0.8 | -0.5 | -0.3 | -0.001 | -0.056 | 0.054 | 0.047 | 0.007 | 0.054 | 0.031 | 0.112 | 0.116 | 0.110 | 0.042 | 0.116 | 0.001 | 0.016 | 0.016 | 0.014 | 0.002 | 0.016 | 1.000 | 0.958 | 1.000 | 0.978 | 1.000 | 1.000 | 0.708 | 0.046 | 0.840 |
|  |  | 0 | -0.8 | 0.8 | 0.5 | 0.3 | 0.002 | -0.065 | 0.067 | 0.057 | 0.010 | 0.067 | 0.033 | 0.112 | 0.115 | 0.110 | 0.043 | 0.115 | 0.001 | 0.017 | 0.018 | 0.015 | 0.002 | 0.018 | 0.052 | 1.000 | 1.000 | 0.990 | 1.000 | 1.000 | 0.618 | 0.039 | 0.800 |
|  |  | -1.6 | -0.8 | -0.8 | -0.5 | -0.3 | 0.001 | -0.056 | 0.057 | 0.047 | 0.010 | 0.057 | 0.030 | 0.111 | 0.113 | 0.107 | 0.043 | 0.113 | 0.001 | 0.016 | 0.016 | 0.014 | 0.002 | 0.016 | 1.000 | 0.977 | 0.941 | 0.396 | 0.998 | 0.945 | 0.032 | 0.052 | 0.818 |
|  |  | 0.8 | 0 | 0.8 | 0.5 | 0.3 | 0.002 | -0.064 | 0.065 | 0.056 | 0.009 | 0.066 | 0.033 | 0.110 | 0.115 | 0.111 | 0.044 | 0.115 | 0.001 | 0.016 | 0.018 | 0.015 | 0.002 | 0.018 | 1.000 | 0.001 | 0.998 | 0.928 | 1.000 | 0.997 | 0.194 | 0.048 | 0.827 |
|  |  | 1.3 | 0.8 | 0.5 | 0.5 | 0 | 0.002 | -0.060 | 0.063 | 0.051 | 0.012 | 0.063 | 0.020 | 0.112 | 0.112 | 0.108 | 0.036 | 0.112 | 0.000 | 0.016 | 0.017 | 0.014 | 0.001 | 0.017 | 1.000 | 0.943 | 0.746 | 0.771 | 0.002 | 0.733 | 0.053 | 0.043 | 0.826 |
|  |  | 1.3 | 0.8 | 0.5 | 0.5 | 0 | 0.003 | -0.048 | 0.052 | 0.049 | 0.003 | 0.052 | 0.035 | 0.108 | 0.113 | 0.108 | 0.027 | 0.113 | 0.001 | 0.014 | 0.016 | 0.014 | 0.001 | 0.016 | 1.000 | 0.945 | 0.646 | 0.671 | 0.081 | 0.651 | 0.016 | 0.058 | 0.822 |
|  |  | 0.8 | 0.8 | 0 | 0 | 0 | 0.000 | -0.059 | 0.060 | 0.050 | 0.010 | 0.060 | 0.013 | 0.117 | 0.117 | 0.112 | 0.035 | 0.117 | 0.000 | 0.017 | 0.017 | 0.015 | 0.001 | 0.017 | 1.000 | 0.998 | 0.021 | 0.024 | 0.013 | 0.029 | 0.514 | 0.043 | 0.803 |
| sc22 | IVW | 1.6 | 0.8 | 0.8 | 0.5 | 0.3 | 0.001 | 0.011 | -0.010 | 0.020 | -0.030 | -0.010 | 0.013 | 0.095 | 0.095 | 0.071 | 0.069 | 0.095 | 0.000 | 0.009 | 0.009 | 0.005 | 0.006 | 0.009 | 1.000 | 0.991 | 0.996 | 0.988 | 0.267 | 0.998 |  |  |  |
|  |  | 0.6 | 0.8 | -0.2 | -0.5 | 0.3 | 0.000 | 0.019 | -0.019 | 0.018 | -0.037 | -0.019 | 0.012 | 0.092 | 0.093 | 0.068 | 0.070 | 0.093 | 0.000 | 0.009 | 0.009 | 0.005 | 0.006 | 0.009 | 1.000 | 1.000 | 0.625 | 1.000 | 0.950 | 0.594 |  |  |  |
|  |  | 0.7 | 0.5 | 0.2 | 0.5 | -0.3 | -0.074 | 0.005 | -0.078 | 0.025 | -0.104 | -0.079 | 0.013 | 0.095 | 0.096 | 0.069 | 0.069 | 0.096 | 0.006 | 0.009 | 0.015 | 0.005 | 0.016 | 0.015 | 1.000 | 0.993 | 0.204 | 1.000 | 0.999 | 0.201 |  |  |  |
|  |  | -0.3 | 0.5 | -0.8 | -0.5 | -0.3 | -0.075 | 0.006 | -0.081 | 0.024 | -0.106 | -0.081 | 0.013 | 0.093 | 0.094 | 0.066 | 0.070 | 0.094 | 0.006 | 0.009 | 0.016 | 0.005 | 0.016 | 0.016 | 1.000 | 0.998 | 1.000 | 1.000 | 1.000 | 1.000 |  |  |  |
|  |  | 0 | -0.8 | 0.8 | 0.5 | 0.3 | 0.000 | 0.013 | -0.013 | 0.022 | -0.034 | -0.013 | 0.013 | 0.092 | 0.092 | 0.070 | 0.065 | 0.092 | 0.000 | 0.009 | 0.009 | 0.005 | 0.005 | 0.009 | 0.045 | 1.000 | 1.000 | 1.000 | 0.949 | 1.000 |  |  |  |
|  |  | -1.6 | -0.8 | -0.8 | -0.5 | -0.3 | -0.074 | 0.006 | -0.081 | 0.024 | -0.105 | -0.081 | 0.013 | 0.097 | 0.098 | 0.067 | 0.070 | 0.098 | 0.006 | 0.009 | 0.016 | 0.005 | 0.016 | 0.016 | 1.000 | 0.999 | 1.000 | 0.992 | 0.877 | 1.000 |  |  |  |
|  |  | 0.8 | 0 | 0.8 | 0.5 | 0.3 | 0.000 | 0.011 | -0.011 | 0.023 | -0.034 | -0.011 | 0.013 | 0.095 | 0.096 | 0.070 | 0.071 | 0.096 | 0.000 | 0.009 | 0.009 | 0.005 | 0.006 | 0.009 | 1.000 | 0.003 | 1.000 | 1.000 | 0.778 | 1.000 |  |  |  |
|  |  | 1.3 | 0.8 | 0.5 | 0.5 | 0 | -0.038 | 0.014 | -0.052 | 0.020 | -0.071 | -0.051 | 0.012 | 0.094 | 0.095 | 0.068 | 0.070 | 0.095 | 0.002 | 0.009 | 0.012 | 0.005 | 0.010 | 0.012 | 1.000 | 0.999 | 0.854 | 1.000 | 0.008 | 0.836 |  |  |  |
|  |  | 1.3 | 0.8 | 0.5 | 0.5 | 0 | 0.001 | -0.050 | 0.051 | 0.022 | 0.030 | 0.051 | 0.013 | 0.068 | 0.069 | 0.068 | 0.009 | 0.069 | 0.000 | 0.007 | 0.007 | 0.005 | 0.001 | 0.007 | 1.000 | 1.000 | 1.000 | 0.998 | 0.438 | 1.000 |  |  |  |
|  |  | 0.8 | 0.8 | 0 | 0 | 0 | -0.037 | 0.017 | -0.053 | 0.018 | -0.071 | -0.053 | 0.006 | 0.093 | 0.092 | 0.068 | 0.066 | 0.092 | 0.001 | 0.009 | 0.011 | 0.005 | 0.009 | 0.011 | 1.000 | 1.000 | 0.052 | 0.039 | 0.124 | 0.047 |  |  |  |
|  | Egger | 1.6 | 0.8 | 0.8 | 0.5 | 0.3 | 0.001 | -0.048 | 0.049 | 0.016 | 0.034 | 0.049 | 0.031 | 0.088 | 0.091 | 0.066 | 0.067 | 0.091 | 0.001 | 0.010 | 0.011 | 0.005 | 0.006 | 0.011 | 1.000 | 0.979 | 0.996 | 0.995 | 0.457 | 0.998 | 0.001 | 0.813 | 0.051 |
|  |  | 0.6 | 0.8 | -0.2 | -0.5 | 0.3 | 0.001 | -0.042 | 0.043 | 0.015 | 0.028 | 0.043 | 0.030 | 0.085 | 0.087 | 0.064 | 0.069 | 0.087 | 0.001 | 0.009 | 0.009 | 0.004 | 0.006 | 0.009 | 1.000 | 1.000 | 0.312 | 1.000 | 0.990 | 0.309 | 0.640 | 0.839 | 0.047 |
|  |  | 0.7 | 0.5 | 0.2 | 0.5 | -0.3 | 0.001 | -0.051 | 0.052 | 0.020 | 0.032 | 0.052 | 0.031 | 0.084 | 0.089 | 0.066 | 0.064 | 0.089 | 0.001 | 0.010 | 0.011 | 0.005 | 0.005 | 0.011 | 1.000 | 0.990 | 0.689 | 1.000 | 0.954 | 0.668 | 0.599 | 0.807 | 0.059 |
|  |  | -0.3 | 0.5 | -0.8 | -0.5 | -0.3 | -0.002 | -0.050 | 0.047 | 0.017 | 0.030 | 0.047 | 0.030 | 0.088 | 0.091 | 0.065 | 0.068 | 0.091 | 0.001 | 0.010 | 0.011 | 0.004 | 0.005 | 0.011 | 1.000 | 0.995 | 1.000 | 1.000 | 0.973 | 1.000 | 0.727 | 0.824 | 0.048 |
|  |  | 0 | -0.8 | 0.8 | 0.5 | 0.3 | 0.001 | -0.046 | 0.047 | 0.017 | 0.031 | 0.048 | 0.033 | 0.082 | 0.088 | 0.066 | 0.064 | 0.088 | 0.001 | 0.009 | 0.010 | 0.005 | 0.005 | 0.010 | 0.049 | 1.000 | 1.000 | 1.000 | 0.993 | 1.000 | 0.619 | 0.828 | 0.049 |
|  |  | -1.6 | -0.8 | -0.8 | -0.5 | -0.3 | -0.003 | -0.050 | 0.047 | 0.015 | 0.031 | 0.047 | 0.031 | 0.090 | 0.093 | 0.065 | 0.067 | 0.093 | 0.001 | 0.011 | 0.011 | 0.005 | 0.005 | 0.011 | 1.000 | 0.998 | 0.995 | 0.991 | 0.270 | 0.997 | 0.019 | 0.787 | 0.048 |
|  |  | 0.8 | 0 | 0.8 | 0.5 | 0.3 | 0.002 | -0.049 | 0.051 | 0.020 | 0.031 | 0.051 | 0.032 | 0.088 | 0.092 | 0.066 | 0.068 | 0.092 | 0.001 | 0.010 | 0.011 | 0.005 | 0.006 | 0.011 | 1.000 | 0.002 | 1.000 | 1.000 | 0.923 | 1.000 | 0.195 | 0.819 | 0.048 |
|  |  | 1.3 | 0.8 | 0.5 | 0.5 | 0 | 0.000 | -0.044 | 0.044 | 0.014 | 0.030 | 0.044 | 0.029 | 0.085 | 0.088 | 0.064 | 0.065 | 0.088 | 0.001 | 0.009 | 0.010 | 0.004 | 0.005 | 0.010 | 1.000 | 0.998 | 0.960 | 1.000 | 0.000 | 0.963 | 0.081 | 0.805 | 0.042 |
|  |  | 1.3 | 0.8 | 0.5 | 0.5 | 0 | 0.003 | -0.015 | 0.019 | 0.018 | 0.001 | 0.019 | 0.032 | 0.062 | 0.067 | 0.065 | 0.015 | 0.067 | 0.001 | 0.004 | 0.005 | 0.004 | 0.000 | 0.005 | 1.000 | 1.000 | 0.996 | 0.998 | 0.016 | 0.999 | 0.029 | 0.818 | 0.049 |
|  |  | 0.8 | 0.8 | 0 | 0 | 0 | 0.000 | -0.040 | 0.041 | 0.013 | 0.028 | 0.041 | 0.013 | 0.087 | 0.086 | 0.061 | 0.066 | 0.086 | 0.000 | 0.009 | 0.009 | 0.004 | 0.005 | 0.009 | 1.000 | 1.000 | 0.021 | 0.022 | 0.020 | 0.027 | 0.529 | 0.804 | 0.050 |
| sc23 | IVW | 1.6 | 0.8 | 0.8 | 0.5 | 0.3 | 0.001 | -0.071 | 0.072 | 0.003 | 0.069 | 0.072 | 0.013 | 0.224 | 0.223 | 0.162 | 0.120 | 0.223 | 0.000 | 0.055 | 0.055 | 0.026 | 0.019 | 0.055 | 1.000 | 0.633 | 0.851 | 0.621 | 0.622 | 0.905 |  |  |  |
|  |  | 0.6 | 0.8 | -0.2 | -0.5 | 0.3 | 0.074 | -0.086 | 0.160 | 0.091 | 0.069 | 0.160 | 0.013 | 0.211 | 0.211 | 0.156 | 0.113 | 0.211 | 0.006 | 0.052 | 0.070 | 0.033 | 0.018 | 0.070 | 1.000 | 0.907 | 0.076 | 0.747 | 0.889 | 0.101 |  |  |  |
|  |  | 0.7 | 0.5 | 0.2 | 0.5 | -0.3 | -0.075 | -0.092 | 0.018 | 0.021 | -0.003 | 0.018 | 0.013 | 0.216 | 0.215 | 0.156 | 0.119 | 0.215 | 0.006 | 0.055 | 0.047 | 0.025 | 0.014 | 0.047 | 1.000 | 0.452 | 0.202 | 0.897 | 0.752 | 0.264 |  |  |  |
|  |  | -0.3 | 0.5 | -0.8 | -0.5 | -0.3 | 0.000 | -0.074 | 0.074 | 0.082 | -0.008 | 0.074 | 0.012 | 0.212 | 0.212 | 0.161 | 0.111 | 0.212 | 0.000 | 0.050 | 0.050 | 0.033 | 0.012 | 0.050 | 1.000 | 0.482 | 0.918 | 0.753 | 0.760 | 0.944 |  |  |  |
|  |  | 0 | -0.8 | 0.8 | 0.5 | 0.3 | 0.000 | -0.063 | 0.063 | -0.001 | 0.065 | 0.064 | 0.013 | 0.218 | 0.218 | 0.160 | 0.115 | 0.218 | 0.000 | 0.052 | 0.051 | 0.026 | 0.017 | 0.051 | 0.050 | 0.964 | 0.977 | 0.882 | 0.891 | 0.988 |  |  |  |
|  |  | -1.6 | -0.8 | -0.8 | -0.5 | -0.3 | 0.000 | -0.071 | 0.071 | 0.078 | -0.007 | 0.071 | 0.012 | 0.210 | 0.210 | 0.155 | 0.115 | 0.210 | 0.000 | 0.049 | 0.049 | 0.030 | 0.013 | 0.049 | 1.000 | 0.902 | 0.802 | 0.596 | 0.576 | 0.868 |  |  |  |
|  |  | 0.8 | 0 | 0.8 | 0.5 | 0.3 | 0.001 | -0.073 | 0.073 | 0.004 | 0.070 | 0.073 | 0.014 | 0.212 | 0.213 | 0.157 | 0.116 | 0.213 | 0.000 | 0.050 | 0.051 | 0.025 | 0.018 | 0.051 | 1.000 | 0.049 | 0.974 | 0.871 | 0.878 | 0.986 |  |  |  |
|  |  | 1.3 | 0.8 | 0.5 | 0.5 | 0 | -0.037 | -0.080 | 0.043 | 0.013 | 0.031 | 0.043 | 0.012 | 0.220 | 0.219 | 0.160 | 0.120 | 0.219 | 0.001 | 0.055 | 0.050 | 0.026 | 0.015 | 0.050 | 1.000 | 0.832 | 0.635 | 0.831 | 0.043 | 0.696 |  |  |  |
|  |  | 1.3 | 0.8 | 0.5 | 0.5 | 0 | 0.001 | -0.055 | 0.056 | 0.015 | 0.041 | 0.056 | 0.013 | 0.167 | 0.168 | 0.164 | 0.015 | 0.168 | 0.000 | 0.031 | 0.031 | 0.027 | 0.002 | 0.031 | 1.000 | 0.951 | 0.813 | 0.784 | 0.759 | 0.827 |  |  |  |
|  |  | 0.8 | 0.8 | 0 | 0 | 0 | 0.001 | -0.080 | 0.080 | 0.046 | 0.035 | 0.080 | 0.010 | 0.213 | 0.213 | 0.161 | 0.114 | 0.213 | 0.000 | 0.052 | 0.052 | 0.028 | 0.014 | 0.052 | 1.000 | 0.896 | 0.088 | 0.084 | 0.085 | 0.109 |  |  |  |
|  | Egger | 1.6 | 0.8 | 0.8 | 0.5 | 0.3 | 0.000 | -0.072 | 0.072 | 0.047 | 0.025 | 0.072 | 0.033 | 0.236 | 0.234 | 0.209 | 0.125 | 0.234 | 0.001 | 0.061 | 0.060 | 0.046 | 0.016 | 0.060 | 1.000 | 0.567 | 0.799 | 0.448 | 0.451 | 0.783 | 0.002 | 0.812 | 0.812 |
|  |  | 0.6 | 0.8 | -0.2 | -0.5 | 0.3 | 0.001 | -0.084 | 0.085 | 0.055 | 0.030 | 0.085 | 0.031 | 0.225 | 0.225 | 0.204 | 0.121 | 0.225 | 0.001 | 0.058 | 0.058 | 0.045 | 0.015 | 0.058 | 1.000 | 0.854 | 0.094 | 0.619 | 0.772 | 0.081 | 0.047 | 0.814 | 0.837 |
|  |  | 0.7 | 0.5 | 0.2 | 0.5 | -0.3 | 0.000 | -0.095 | 0.095 | 0.067 | 0.027 | 0.095 | 0.030 | 0.225 | 0.224 | 0.199 | 0.125 | 0.224 | 0.001 | 0.060 | 0.059 | 0.044 | 0.016 | 0.059 | 1.000 | 0.402 | 0.275 | 0.813 | 0.625 | 0.253 | 0.050 | 0.808 | 0.810 |
|  |  | -0.3 | 0.5 | -0.8 | -0.5 | -0.3 | 0.000 | -0.074 | 0.074 | 0.049 | 0.025 | 0.074 | 0.031 | 0.223 | 0.223 | 0.204 | 0.117 | 0.223 | 0.001 | 0.055 | 0.055 | 0.044 | 0.014 | 0.055 | 1.000 | 0.450 | 0.890 | 0.625 | 0.634 | 0.873 | 0.061 | 0.831 | 0.819 |
|  |  | 0 | -0.8 | 0.8 | 0.5 | 0.3 | 0.000 | -0.063 | 0.064 | 0.041 | 0.022 | 0.064 | 0.032 | 0.230 | 0.229 | 0.206 | 0.122 | 0.229 | 0.001 | 0.057 | 0.056 | 0.044 | 0.015 | 0.057 | 0.046 | 0.946 | 0.962 | 0.749 | 0.773 | 0.967 | 0.042 | 0.818 | 0.823 |
|  |  | -1.6 | -0.8 | -0.8 | -0.5 | -0.3 | 0.000 | -0.067 | 0.067 | 0.037 | 0.030 | 0.067 | 0.030 | 0.222 | 0.221 | 0.199 | 0.123 | 0.221 | 0.001 | 0.054 | 0.054 | 0.041 | 0.016 | 0.054 | 1.000 | 0.860 | 0.765 | 0.434 | 0.403 | 0.730 | 0.009 | 0.819 | 0.829 |
|  |  | 0.8 | 0 | 0.8 | 0.5 | 0.3 | 0.000 | -0.072 | 0.072 | 0.045 | 0.027 | 0.072 | 0.034 | 0.226 | 0.227 | 0.205 | 0.126 | 0.227 | 0.001 | 0.056 | 0.057 | 0.044 | 0.017 | 0.057 | 1.000 | 0.054 | 0.954 | 0.765 | 0.761 | 0.955 | 0.040 | 0.827 | 0.802 |
|  |  | 1.3 | 0.8 | 0.5 | 0.5 | 0 | 0.001 | -0.077 | 0.078 | 0.049 | 0.029 | 0.078 | 0.029 | 0.230 | 0.228 | 0.201 | 0.125 | 0.228 | 0.001 | 0.059 | 0.058 | 0.043 | 0.016 | 0.058 | 1.000 | 0.787 | 0.638 | 0.696 | 0.034 | 0.609 | 0.013 | 0.801 | 0.822 |
|  |  | 1.3 | 0.8 | 0.5 | 0.5 | 0 | 0.002 | -0.059 | 0.061 | 0.061 | 0.001 | 0.061 | 0.033 | 0.213 | 0.213 | 0.213 | 0.015 | 0.213 | 0.001 | 0.049 | 0.049 | 0.049 | 0.000 | 0.049 | 1.000 | 0.837 | 0.609 | 0.639 | 0.022 | 0.643 | 0.012 | 0.820 | 0.820 |
|  |  | 0.8 | 0.8 | 0 | 0 | 0 | 0.000 | -0.083 | 0.083 | 0.054 | 0.029 | 0.083 | 0.025 | 0.228 | 0.227 | 0.204 | 0.118 | 0.227 | 0.001 | 0.059 | 0.059 | 0.044 | 0.015 | 0.059 | 1.000 | 0.850 | 0.085 | 0.082 | 0.071 | 0.068 | 0.045 | 0.797 | 0.826 |

TE: total effect; DE: direct effect; IE: indirect effect; IE_d: indirect effect calculated by difference method; IE_p: indirect effect calculated by product method; IE1: *X*→*M_1_*→*Y*; IE2: *X*→*M_2_*→*Y*.

**eTable 10.** Simulation of PSE-Egger in two non-ordered mediators setting with different effects (case(b),sc3,sc31-33).

|  |  | TRUE | | | | | BIAS | | | | | | SE | | | | | | MSE | | | | | | Power/Type I error | | | | | | | | |
| --- | --- | --- | --- | --- | --- | --- | --- | --- | --- | --- | --- | --- | --- | --- | --- | --- | --- | --- | --- | --- | --- | --- | --- | --- | --- | --- | --- | --- | --- | --- | --- | --- | --- |
|  |  | TE | DE | IE | IE2 | IE1 | TE | DE | IE_d | IE1 | IE2 | IE_p | TE | DE | IE_d | IE1 | IE2 | IE_p | TE | DE | IE_d | IE1 | IE2 | IE_p | TE | DE | IE_d | IE1 | IE2 | IE_p | GY | GM1 | GM2 |
| Sc3 | IVW | 1.6 | 0.8 | 0.8 | 0.5 | 0.3 | 0.112 | -0.193 | 0.304 | 0.174 | 0.130 | 0.305 | 0.013 | 0.108 | 0.108 | 0.109 | 0.071 | 0.108 | 0.013 | 0.049 | 0.104 | 0.042 | 0.022 | 0.105 | 1.000 | 0.565 | 0.998 | 0.746 | 0.806 | 0.994 |  |  |  |
|  |  | 0.6 | 0.8 | -0.2 | -0.5 | 0.3 | 0.037 | -0.192 | 0.229 | 0.100 | 0.129 | 0.229 | 0.009 | 0.107 | 0.107 | 0.111 | 0.069 | 0.107 | 0.001 | 0.048 | 0.064 | 0.022 | 0.021 | 0.064 | 1.000 | 0.999 | 0.035 | 0.918 | 1.000 | 0.017 |  |  |  |
|  |  | 0.7 | 0.5 | 0.2 | 0.5 | -0.3 | 0.037 | -0.191 | 0.229 | 0.176 | 0.052 | 0.229 | 0.009 | 0.105 | 0.106 | 0.109 | 0.068 | 0.106 | 0.001 | 0.048 | 0.064 | 0.043 | 0.007 | 0.064 | 1.000 | 0.645 | 0.924 | 1.000 | 0.876 | 0.842 |  |  |  |
|  |  | -0.3 | 0.5 | -0.8 | -0.5 | -0.3 | -0.037 | -0.195 | 0.158 | 0.110 | 0.048 | 0.158 | 0.010 | 0.106 | 0.107 | 0.112 | 0.069 | 0.107 | 0.001 | 0.049 | 0.036 | 0.025 | 0.007 | 0.036 | 1.000 | 0.766 | 0.999 | 0.932 | 0.943 | 0.999 |  |  |  |
|  |  | 0 | -0.8 | 0.8 | 0.5 | 0.3 | 0.113 | -0.197 | 0.310 | 0.183 | 0.126 | 0.309 | 0.013 | 0.104 | 0.105 | 0.106 | 0.068 | 0.105 | 0.013 | 0.049 | 0.107 | 0.045 | 0.021 | 0.107 | 1.000 | 1.000 | 1.000 | 1.000 | 1.000 | 1.000 |  |  |  |
|  |  | -1.6 | -0.8 | -0.8 | -0.5 | -0.3 | -0.037 | -0.196 | 0.158 | 0.105 | 0.053 | 0.158 | 0.010 | 0.109 | 0.108 | 0.111 | 0.067 | 0.108 | 0.001 | 0.050 | 0.037 | 0.023 | 0.007 | 0.037 | 1.000 | 1.000 | 0.880 | 0.222 | 0.213 | 0.684 |  |  |  |
|  |  | 0.8 | 0 | 0.8 | 0.5 | 0.3 | 0.112 | -0.193 | 0.305 | 0.180 | 0.125 | 0.305 | 0.012 | 0.104 | 0.105 | 0.109 | 0.066 | 0.105 | 0.013 | 0.048 | 0.104 | 0.044 | 0.020 | 0.104 | 1.000 | 0.034 | 1.000 | 0.982 | 0.990 | 0.999 |  |  |  |
|  |  | 1.3 | 0.8 | 0.5 | 0.5 | 0 | 0.075 | -0.190 | 0.265 | 0.174 | 0.091 | 0.265 | 0.009 | 0.106 | 0.106 | 0.108 | 0.071 | 0.106 | 0.006 | 0.047 | 0.082 | 0.042 | 0.013 | 0.082 | 1.000 | 0.873 | 0.993 | 0.943 | 0.008 | 0.953 |  |  |  |
|  |  | 1.3 | 0.8 | 0.5 | 0.5 | 0 | 0.111 | -0.111 | 0.222 | 0.175 | 0.048 | 0.223 | 0.013 | 0.107 | 0.107 | 0.109 | 0.010 | 0.107 | 0.013 | 0.024 | 0.061 | 0.043 | 0.002 | 0.061 | 1.000 | 0.913 | 0.951 | 0.900 | 0.918 | 0.942 |  |  |  |
|  |  | 0.8 | 0.8 | 0 | 0 | 0 | 0.037 | -0.190 | 0.227 | 0.136 | 0.091 | 0.227 | 0.006 | 0.103 | 0.103 | 0.104 | 0.067 | 0.103 | 0.001 | 0.047 | 0.062 | 0.029 | 0.013 | 0.062 | 1.000 | 0.995 | 0.388 | 0.122 | 0.143 | 0.214 |  |  |  |
|  | Egger | 1.6 | 0.8 | 0.8 | 0.5 | 0.3 | 0.001 | -0.080 | 0.081 | 0.049 | 0.032 | 0.081 | 0.026 | 0.120 | 0.121 | 0.109 | 0.068 | 0.121 | 0.001 | 0.021 | 0.021 | 0.014 | 0.006 | 0.021 | 1.000 | 0.542 | 0.895 | 0.367 | 0.414 | 0.877 | 0.000 | 0.815 | 0.797 |
|  |  | 0.6 | 0.8 | -0.2 | -0.5 | 0.3 | 0.001 | -0.078 | 0.079 | 0.045 | 0.033 | 0.079 | 0.023 | 0.123 | 0.125 | 0.111 | 0.066 | 0.125 | 0.001 | 0.021 | 0.022 | 0.014 | 0.006 | 0.022 | 1.000 | 1.000 | 0.101 | 0.959 | 0.995 | 0.087 | 0.277 | 0.812 | 0.811 |
|  |  | 0.7 | 0.5 | 0.2 | 0.5 | -0.3 | 0.002 | -0.077 | 0.078 | 0.049 | 0.029 | 0.078 | 0.022 | 0.121 | 0.123 | 0.109 | 0.067 | 0.123 | 0.000 | 0.021 | 0.021 | 0.014 | 0.005 | 0.021 | 1.000 | 0.799 | 0.406 | 0.991 | 0.926 | 0.367 | 0.195 | 0.823 | 0.789 |
|  |  | -0.3 | 0.5 | -0.8 | -0.5 | -0.3 | -0.001 | -0.076 | 0.075 | 0.052 | 0.023 | 0.075 | 0.021 | 0.123 | 0.124 | 0.111 | 0.068 | 0.124 | 0.000 | 0.021 | 0.021 | 0.015 | 0.005 | 0.021 | 1.000 | 0.893 | 1.000 | 0.979 | 0.978 | 1.000 | 0.422 | 0.815 | 0.817 |
|  |  | 0 | -0.8 | 0.8 | 0.5 | 0.3 | 0.002 | -0.076 | 0.078 | 0.049 | 0.029 | 0.078 | 0.026 | 0.121 | 0.124 | 0.109 | 0.068 | 0.124 | 0.001 | 0.020 | 0.021 | 0.014 | 0.006 | 0.021 | 0.052 | 0.999 | 0.999 | 0.993 | 0.995 | 0.999 | 0.305 | 0.798 | 0.785 |
|  |  | -1.6 | -0.8 | -0.8 | -0.5 | -0.3 | 0.000 | -0.082 | 0.082 | 0.050 | 0.032 | 0.082 | 0.023 | 0.124 | 0.124 | 0.110 | 0.067 | 0.124 | 0.001 | 0.022 | 0.022 | 0.015 | 0.005 | 0.022 | 1.000 | 0.945 | 0.818 | 0.311 | 0.313 | 0.772 | 0.000 | 0.815 | 0.797 |
|  |  | 0.8 | 0 | 0.8 | 0.5 | 0.3 | 0.002 | -0.075 | 0.077 | 0.049 | 0.028 | 0.077 | 0.026 | 0.119 | 0.121 | 0.111 | 0.065 | 0.121 | 0.001 | 0.020 | 0.021 | 0.015 | 0.005 | 0.021 | 1.000 | 0.000 | 0.997 | 0.884 | 0.889 | 0.997 | 0.026 | 0.801 | 0.810 |
|  |  | 1.3 | 0.8 | 0.5 | 0.5 | 0 | 0.002 | -0.080 | 0.082 | 0.049 | 0.033 | 0.082 | 0.020 | 0.122 | 0.124 | 0.112 | 0.067 | 0.124 | 0.000 | 0.021 | 0.022 | 0.015 | 0.006 | 0.022 | 1.000 | 0.873 | 0.653 | 0.734 | 0.000 | 0.602 | 0.001 | 0.818 | 0.808 |
|  |  | 1.3 | 0.8 | 0.5 | 0.5 | 0 | 0.001 | -0.045 | 0.046 | 0.046 | 0.000 | 0.046 | 0.025 | 0.110 | 0.110 | 0.109 | 0.015 | 0.110 | 0.001 | 0.014 | 0.014 | 0.014 | 0.000 | 0.014 | 1.000 | 0.925 | 0.569 | 0.599 | 0.017 | 0.605 | 0.001 | 0.806 | 0.808 |
|  |  | 0.8 | 0.8 | 0 | 0 | 0 | 0.001 | -0.078 | 0.079 | 0.046 | 0.032 | 0.079 | 0.013 | 0.118 | 0.117 | 0.103 | 0.067 | 0.117 | 0.000 | 0.020 | 0.020 | 0.013 | 0.005 | 0.020 | 1.000 | 0.998 | 0.022 | 0.010 | 0.022 | 0.016 | 0.169 | 0.824 | 0.822 |
| Sc31 | IVW | 1.6 | 0.8 | 0.8 | 0.5 | 0.3 | 0.038 | 0.016 | 0.022 | 0.012 | 0.010 | 0.022 | 0.016 | 0.080 | 0.081 | 0.070 | 0.041 | 0.082 | 0.002 | 0.007 | 0.007 | 0.005 | 0.002 | 0.007 | 1.000 | 0.999 | 1.000 | 0.985 | 0.990 | 1.000 |  |  |  |
|  |  | 0.6 | 0.8 | -0.2 | -0.5 | 0.3 | 0.037 | 0.012 | 0.025 | 0.017 | 0.008 | 0.025 | 0.015 | 0.080 | 0.081 | 0.071 | 0.041 | 0.081 | 0.002 | 0.007 | 0.007 | 0.005 | 0.002 | 0.007 | 1.000 | 1.000 | 0.567 | 1.000 | 1.000 | 0.554 |  |  |  |
|  |  | 0.7 | 0.5 | 0.2 | 0.5 | -0.3 | 0.037 | 0.015 | 0.022 | 0.012 | 0.010 | 0.022 | 0.015 | 0.082 | 0.084 | 0.073 | 0.042 | 0.084 | 0.002 | 0.007 | 0.008 | 0.006 | 0.002 | 0.008 | 1.000 | 0.998 | 0.719 | 1.000 | 1.000 | 0.717 |  |  |  |
|  |  | -0.3 | 0.5 | -0.8 | -0.5 | -0.3 | 0.037 | 0.017 | 0.020 | 0.013 | 0.007 | 0.020 | 0.015 | 0.079 | 0.080 | 0.068 | 0.045 | 0.080 | 0.002 | 0.007 | 0.007 | 0.005 | 0.002 | 0.007 | 1.000 | 1.000 | 1.000 | 1.000 | 1.000 | 1.000 |  |  |  |
|  |  | 0 | -0.8 | 0.8 | 0.5 | 0.3 | 0.037 | 0.016 | 0.021 | 0.014 | 0.008 | 0.022 | 0.016 | 0.078 | 0.080 | 0.069 | 0.043 | 0.080 | 0.002 | 0.006 | 0.007 | 0.005 | 0.002 | 0.007 | 0.640 | 1.000 | 1.000 | 1.000 | 1.000 | 1.000 |  |  |  |
|  |  | -1.6 | -0.8 | -0.8 | -0.5 | -0.3 | 0.036 | 0.009 | 0.027 | 0.018 | 0.008 | 0.026 | 0.016 | 0.077 | 0.078 | 0.067 | 0.042 | 0.078 | 0.002 | 0.006 | 0.007 | 0.005 | 0.002 | 0.007 | 1.000 | 1.000 | 1.000 | 0.995 | 0.994 | 1.000 |  |  |  |
|  |  | 0.8 | 0 | 0.8 | 0.5 | 0.3 | 0.039 | 0.012 | 0.026 | 0.016 | 0.010 | 0.026 | 0.016 | 0.078 | 0.081 | 0.070 | 0.042 | 0.081 | 0.002 | 0.006 | 0.007 | 0.005 | 0.002 | 0.007 | 1.000 | 0.001 | 1.000 | 1.000 | 1.000 | 1.000 |  |  |  |
|  |  | 1.3 | 0.8 | 0.5 | 0.5 | 0 | 0.038 | 0.010 | 0.028 | 0.020 | 0.008 | 0.028 | 0.012 | 0.079 | 0.080 | 0.070 | 0.043 | 0.080 | 0.002 | 0.006 | 0.007 | 0.005 | 0.002 | 0.007 | 1.000 | 1.000 | 0.994 | 1.000 | 0.000 | 0.997 |  |  |  |
|  |  | 1.3 | 0.8 | 0.5 | 0.5 | 0 | 0.038 | 0.022 | 0.016 | 0.016 | 0.000 | 0.016 | 0.015 | 0.072 | 0.073 | 0.072 | 0.010 | 0.073 | 0.002 | 0.006 | 0.006 | 0.005 | 0.000 | 0.006 | 1.000 | 1.000 | 0.996 | 0.998 | 0.062 | 0.995 |  |  |  |
|  |  | 0.8 | 0.8 | 0 | 0 | 0 | 0.037 | 0.010 | 0.027 | 0.017 | 0.009 | 0.027 | 0.006 | 0.080 | 0.080 | 0.070 | 0.042 | 0.080 | 0.001 | 0.007 | 0.007 | 0.005 | 0.002 | 0.007 | 1.000 | 1.000 | 0.046 | 0.036 | 0.034 | 0.042 |  |  |  |
|  | Egger | 1.6 | 0.8 | 0.8 | 0.5 | 0.3 | 0.000 | -0.021 | 0.021 | 0.013 | 0.008 | 0.021 | 0.038 | 0.071 | 0.077 | 0.065 | 0.043 | 0.077 | 0.001 | 0.005 | 0.006 | 0.004 | 0.002 | 0.006 | 1.000 | 0.999 | 0.999 | 0.983 | 0.990 | 1.000 | 0.004 | 0.049 | 0.044 |
|  |  | 0.6 | 0.8 | -0.2 | -0.5 | 0.3 | 0.002 | -0.022 | 0.024 | 0.016 | 0.008 | 0.024 | 0.037 | 0.071 | 0.076 | 0.065 | 0.043 | 0.076 | 0.001 | 0.006 | 0.006 | 0.005 | 0.002 | 0.006 | 1.000 | 1.000 | 0.512 | 1.000 | 1.000 | 0.526 | 0.676 | 0.057 | 0.048 |
|  |  | 0.7 | 0.5 | 0.2 | 0.5 | -0.3 | 0.001 | -0.020 | 0.021 | 0.011 | 0.010 | 0.020 | 0.037 | 0.071 | 0.076 | 0.064 | 0.044 | 0.076 | 0.001 | 0.005 | 0.006 | 0.004 | 0.002 | 0.006 | 1.000 | 1.000 | 0.669 | 1.000 | 1.000 | 0.667 | 0.567 | 0.037 | 0.047 |
|  |  | -0.3 | 0.5 | -0.8 | -0.5 | -0.3 | 0.001 | -0.019 | 0.020 | 0.013 | 0.007 | 0.020 | 0.038 | 0.068 | 0.076 | 0.062 | 0.045 | 0.076 | 0.001 | 0.005 | 0.006 | 0.004 | 0.002 | 0.006 | 1.000 | 1.000 | 1.000 | 1.000 | 1.000 | 1.000 | 0.797 | 0.047 | 0.051 |
|  |  | 0 | -0.8 | 0.8 | 0.5 | 0.3 | 0.003 | -0.022 | 0.025 | 0.017 | 0.009 | 0.025 | 0.040 | 0.069 | 0.077 | 0.065 | 0.043 | 0.077 | 0.002 | 0.005 | 0.007 | 0.004 | 0.002 | 0.007 | 0.053 | 1.000 | 1.000 | 1.000 | 1.000 | 1.000 | 0.671 | 0.046 | 0.040 |
|  |  | -1.6 | -0.8 | -0.8 | -0.5 | -0.3 | -0.001 | -0.027 | 0.026 | 0.017 | 0.009 | 0.026 | 0.036 | 0.070 | 0.074 | 0.062 | 0.043 | 0.074 | 0.001 | 0.006 | 0.006 | 0.004 | 0.002 | 0.006 | 1.000 | 1.000 | 1.000 | 0.996 | 0.994 | 1.000 | 0.032 | 0.057 | 0.045 |
|  |  | 0.8 | 0 | 0.8 | 0.5 | 0.3 | 0.002 | -0.026 | 0.028 | 0.018 | 0.010 | 0.028 | 0.038 | 0.070 | 0.079 | 0.065 | 0.043 | 0.079 | 0.001 | 0.006 | 0.007 | 0.005 | 0.002 | 0.007 | 1.000 | 0.001 | 1.000 | 1.000 | 1.000 | 1.000 | 0.218 | 0.048 | 0.051 |
|  |  | 1.3 | 0.8 | 0.5 | 0.5 | 0 | 0.001 | -0.028 | 0.029 | 0.020 | 0.009 | 0.029 | 0.028 | 0.070 | 0.073 | 0.064 | 0.036 | 0.073 | 0.001 | 0.006 | 0.006 | 0.005 | 0.001 | 0.006 | 1.000 | 1.000 | 0.995 | 1.000 | 0.000 | 0.998 | 0.087 | 0.046 | 0.046 |
|  |  | 1.3 | 0.8 | 0.5 | 0.5 | 0 | 0.002 | -0.013 | 0.015 | 0.013 | 0.002 | 0.015 | 0.037 | 0.062 | 0.070 | 0.065 | 0.025 | 0.070 | 0.001 | 0.004 | 0.005 | 0.004 | 0.001 | 0.005 | 1.000 | 1.000 | 0.990 | 0.997 | 0.054 | 0.994 | 0.029 | 0.041 | 0.058 |
|  |  | 0.8 | 0.8 | 0 | 0 | 0 | 0.000 | -0.026 | 0.026 | 0.017 | 0.009 | 0.026 | 0.014 | 0.071 | 0.070 | 0.060 | 0.036 | 0.070 | 0.000 | 0.006 | 0.006 | 0.004 | 0.001 | 0.006 | 1.000 | 1.000 | 0.010 | 0.013 | 0.008 | 0.014 | 0.555 | 0.039 | 0.050 |
| Sc32 | IVW | 1.6 | 0.8 | 0.8 | 0.5 | 0.3 | 0.037 | -0.046 | 0.083 | 0.020 | 0.063 | 0.083 | 0.016 | 0.150 | 0.151 | 0.108 | 0.114 | 0.151 | 0.002 | 0.025 | 0.030 | 0.012 | 0.017 | 0.030 | 1.000 | 0.948 | 0.988 | 0.969 | 0.640 | 0.993 |  |  |  |
|  |  | 0.6 | 0.8 | -0.2 | -0.5 | 0.3 | 0.038 | -0.035 | 0.072 | 0.010 | 0.062 | 0.072 | 0.014 | 0.155 | 0.154 | 0.105 | 0.115 | 0.154 | 0.002 | 0.025 | 0.029 | 0.011 | 0.017 | 0.029 | 1.000 | 0.996 | 0.160 | 0.996 | 0.872 | 0.142 |  |  |  |
|  |  | 0.7 | 0.5 | 0.2 | 0.5 | -0.3 | -0.037 | -0.044 | 0.007 | 0.016 | -0.009 | 0.007 | 0.015 | 0.151 | 0.151 | 0.109 | 0.109 | 0.151 | 0.002 | 0.025 | 0.023 | 0.012 | 0.012 | 0.023 | 1.000 | 0.804 | 0.289 | 0.996 | 0.772 | 0.275 |  |  |  |
|  |  | -0.3 | 0.5 | -0.8 | -0.5 | -0.3 | -0.037 | -0.040 | 0.003 | 0.020 | -0.017 | 0.003 | 0.015 | 0.149 | 0.149 | 0.105 | 0.111 | 0.149 | 0.002 | 0.024 | 0.022 | 0.011 | 0.013 | 0.022 | 1.000 | 0.820 | 0.998 | 0.990 | 0.812 | 1.000 |  |  |  |
|  |  | 0 | -0.8 | 0.8 | 0.5 | 0.3 | 0.038 | -0.038 | 0.075 | 0.012 | 0.063 | 0.075 | 0.017 | 0.153 | 0.153 | 0.107 | 0.114 | 0.153 | 0.002 | 0.025 | 0.029 | 0.012 | 0.017 | 0.029 | 0.626 | 0.999 | 0.999 | 0.997 | 0.896 | 1.000 |  |  |  |
|  |  | -1.6 | -0.8 | -0.8 | -0.5 | -0.3 | -0.037 | -0.035 | -0.002 | 0.005 | -0.008 | -0.002 | 0.014 | 0.150 | 0.150 | 0.107 | 0.111 | 0.150 | 0.002 | 0.024 | 0.023 | 0.011 | 0.012 | 0.023 | 1.000 | 0.988 | 0.991 | 0.973 | 0.579 | 0.993 |  |  |  |
|  |  | 0.8 | 0 | 0.8 | 0.5 | 0.3 | 0.037 | -0.039 | 0.076 | 0.011 | 0.065 | 0.076 | 0.016 | 0.149 | 0.148 | 0.107 | 0.113 | 0.148 | 0.002 | 0.024 | 0.028 | 0.012 | 0.017 | 0.028 | 1.000 | 0.046 | 1.000 | 0.994 | 0.885 | 1.000 |  |  |  |
|  |  | 1.3 | 0.8 | 0.5 | 0.5 | 0 | 0.001 | -0.040 | 0.041 | 0.016 | 0.025 | 0.041 | 0.014 | 0.148 | 0.149 | 0.108 | 0.110 | 0.149 | 0.000 | 0.024 | 0.024 | 0.012 | 0.013 | 0.024 | 1.000 | 0.988 | 0.894 | 0.991 | 0.031 | 0.885 |  |  |  |
|  |  | 1.3 | 0.8 | 0.5 | 0.5 | 0 | 0.038 | -0.023 | 0.061 | 0.021 | 0.040 | 0.061 | 0.016 | 0.105 | 0.105 | 0.105 | 0.014 | 0.105 | 0.002 | 0.012 | 0.015 | 0.011 | 0.002 | 0.015 | 1.000 | 1.000 | 0.991 | 0.982 | 0.749 | 0.990 |  |  |  |
|  |  | 0.8 | 0.8 | 0 | 0 | 0 | 0.000 | -0.037 | 0.037 | 0.015 | 0.023 | 0.037 | 0.010 | 0.152 | 0.151 | 0.105 | 0.109 | 0.151 | 0.000 | 0.024 | 0.024 | 0.011 | 0.012 | 0.024 | 1.000 | 0.993 | 0.072 | 0.056 | 0.057 | 0.067 |  |  |  |
|  | Egger | 1.6 | 0.8 | 0.8 | 0.5 | 0.3 | 0.001 | -0.054 | 0.055 | 0.019 | 0.036 | 0.055 | 0.039 | 0.157 | 0.159 | 0.111 | 0.122 | 0.159 | 0.002 | 0.028 | 0.028 | 0.013 | 0.016 | 0.028 | 1.000 | 0.921 | 0.978 | 0.954 | 0.467 | 0.986 | 0.002 | 0.814 | 0.055 |
|  |  | 0.6 | 0.8 | -0.2 | -0.5 | 0.3 | 0.002 | -0.043 | 0.045 | 0.009 | 0.035 | 0.045 | 0.037 | 0.164 | 0.164 | 0.109 | 0.125 | 0.164 | 0.001 | 0.029 | 0.029 | 0.012 | 0.017 | 0.029 | 1.000 | 0.990 | 0.175 | 0.993 | 0.793 | 0.177 | 0.050 | 0.809 | 0.047 |
|  |  | 0.7 | 0.5 | 0.2 | 0.5 | -0.3 | 0.000 | -0.050 | 0.051 | 0.016 | 0.035 | 0.050 | 0.037 | 0.159 | 0.160 | 0.113 | 0.119 | 0.160 | 0.001 | 0.028 | 0.028 | 0.013 | 0.015 | 0.028 | 1.000 | 0.754 | 0.336 | 0.993 | 0.613 | 0.321 | 0.062 | 0.816 | 0.047 |
|  |  | -0.3 | 0.5 | -0.8 | -0.5 | -0.3 | -0.001 | -0.048 | 0.047 | 0.018 | 0.028 | 0.047 | 0.037 | 0.159 | 0.160 | 0.111 | 0.123 | 0.160 | 0.001 | 0.027 | 0.028 | 0.013 | 0.016 | 0.028 | 1.000 | 0.773 | 0.989 | 0.984 | 0.640 | 0.994 | 0.058 | 0.811 | 0.054 |
|  |  | 0 | -0.8 | 0.8 | 0.5 | 0.3 | 0.001 | -0.046 | 0.047 | 0.011 | 0.036 | 0.047 | 0.038 | 0.161 | 0.162 | 0.113 | 0.121 | 0.162 | 0.001 | 0.028 | 0.029 | 0.013 | 0.016 | 0.029 | 0.051 | 0.998 | 0.998 | 0.995 | 0.795 | 0.999 | 0.056 | 0.808 | 0.043 |
|  |  | -1.6 | -0.8 | -0.8 | -0.5 | -0.3 | -0.001 | -0.043 | 0.042 | 0.005 | 0.037 | 0.042 | 0.038 | 0.160 | 0.162 | 0.111 | 0.124 | 0.162 | 0.001 | 0.027 | 0.028 | 0.012 | 0.017 | 0.028 | 1.000 | 0.983 | 0.968 | 0.970 | 0.367 | 0.976 | 0.007 | 0.819 | 0.048 |
|  |  | 0.8 | 0 | 0.8 | 0.5 | 0.3 | 0.001 | -0.046 | 0.047 | 0.011 | 0.036 | 0.047 | 0.039 | 0.158 | 0.157 | 0.111 | 0.123 | 0.157 | 0.002 | 0.027 | 0.027 | 0.012 | 0.016 | 0.027 | 1.000 | 0.053 | 0.998 | 0.994 | 0.768 | 1.000 | 0.040 | 0.816 | 0.044 |
|  |  | 1.3 | 0.8 | 0.5 | 0.5 | 0 | 0.003 | -0.048 | 0.051 | 0.016 | 0.035 | 0.051 | 0.034 | 0.157 | 0.158 | 0.112 | 0.120 | 0.158 | 0.001 | 0.027 | 0.027 | 0.013 | 0.016 | 0.027 | 1.000 | 0.978 | 0.853 | 0.986 | 0.028 | 0.850 | 0.009 | 0.803 | 0.045 |
|  |  | 1.3 | 0.8 | 0.5 | 0.5 | 0 | 0.004 | -0.019 | 0.023 | 0.021 | 0.002 | 0.023 | 0.040 | 0.112 | 0.113 | 0.113 | 0.016 | 0.113 | 0.002 | 0.013 | 0.013 | 0.013 | 0.000 | 0.013 | 1.000 | 1.000 | 0.974 | 0.980 | 0.019 | 0.977 | 0.006 | 0.788 | 0.047 |
|  |  | 0.8 | 0.8 | 0 | 0 | 0 | 0.001 | -0.044 | 0.045 | 0.015 | 0.030 | 0.045 | 0.024 | 0.164 | 0.163 | 0.108 | 0.123 | 0.163 | 0.001 | 0.029 | 0.029 | 0.012 | 0.016 | 0.029 | 1.000 | 0.988 | 0.064 | 0.060 | 0.082 | 0.068 | 0.056 | 0.825 | 0.045 |
| Sc33 | IVW | 1.6 | 0.8 | 0.8 | 0.5 | 0.3 | 0.036 | -0.054 | 0.090 | 0.081 | 0.009 | 0.090 | 0.016 | 0.188 | 0.187 | 0.180 | 0.067 | 0.187 | 0.002 | 0.038 | 0.043 | 0.039 | 0.005 | 0.043 | 1.000 | 0.799 | 0.942 | 0.635 | 0.950 | 0.940 |  |  |  |
|  |  | 0.6 | 0.8 | -0.2 | -0.5 | 0.3 | -0.036 | -0.046 | 0.010 | 0.000 | 0.009 | 0.010 | 0.015 | 0.183 | 0.183 | 0.174 | 0.064 | 0.183 | 0.002 | 0.036 | 0.034 | 0.030 | 0.004 | 0.034 | 1.000 | 0.958 | 0.170 | 0.798 | 0.997 | 0.172 |  |  |  |
|  |  | 0.7 | 0.5 | 0.2 | 0.5 | -0.3 | 0.037 | -0.044 | 0.081 | 0.076 | 0.004 | 0.081 | 0.015 | 0.188 | 0.188 | 0.176 | 0.065 | 0.188 | 0.002 | 0.037 | 0.042 | 0.037 | 0.004 | 0.042 | 1.000 | 0.643 | 0.340 | 0.882 | 0.992 | 0.329 |  |  |  |
|  |  | -0.3 | 0.5 | -0.8 | -0.5 | -0.3 | -0.037 | -0.058 | 0.021 | 0.011 | 0.010 | 0.021 | 0.015 | 0.194 | 0.193 | 0.185 | 0.063 | 0.193 | 0.002 | 0.041 | 0.038 | 0.034 | 0.004 | 0.038 | 1.000 | 0.614 | 0.976 | 0.759 | 0.995 | 0.979 |  |  |  |
|  |  | 0 | -0.8 | 0.8 | 0.5 | 0.3 | 0.038 | -0.054 | 0.092 | 0.082 | 0.010 | 0.092 | 0.016 | 0.187 | 0.187 | 0.180 | 0.067 | 0.187 | 0.002 | 0.038 | 0.043 | 0.039 | 0.005 | 0.043 | 0.642 | 0.989 | 0.996 | 0.897 | 0.992 | 0.997 |  |  |  |
|  |  | -1.6 | -0.8 | -0.8 | -0.5 | -0.3 | -0.038 | -0.049 | 0.010 | 0.005 | 0.006 | 0.010 | 0.014 | 0.181 | 0.181 | 0.175 | 0.066 | 0.181 | 0.002 | 0.035 | 0.033 | 0.030 | 0.004 | 0.033 | 1.000 | 0.944 | 0.929 | 0.597 | 0.958 | 0.929 |  |  |  |
|  |  | 0.8 | 0 | 0.8 | 0.5 | 0.3 | 0.038 | -0.038 | 0.075 | 0.065 | 0.010 | 0.075 | 0.016 | 0.194 | 0.195 | 0.187 | 0.066 | 0.195 | 0.002 | 0.039 | 0.044 | 0.039 | 0.005 | 0.044 | 1.000 | 0.044 | 0.991 | 0.852 | 0.993 | 0.991 |  |  |  |
|  |  | 1.3 | 0.8 | 0.5 | 0.5 | 0 | 0.037 | -0.063 | 0.100 | 0.091 | 0.009 | 0.100 | 0.011 | 0.192 | 0.191 | 0.181 | 0.066 | 0.191 | 0.001 | 0.041 | 0.047 | 0.041 | 0.004 | 0.047 | 1.000 | 0.903 | 0.813 | 0.840 | 0.025 | 0.809 |  |  |  |
|  |  | 1.3 | 0.8 | 0.5 | 0.5 | 0 | 0.038 | -0.037 | 0.075 | 0.075 | 0.000 | 0.075 | 0.015 | 0.178 | 0.177 | 0.178 | 0.011 | 0.177 | 0.002 | 0.033 | 0.037 | 0.037 | 0.000 | 0.037 | 1.000 | 0.925 | 0.765 | 0.765 | 0.064 | 0.767 |  |  |  |
|  |  | 0.8 | 0.8 | 0 | 0 | 0 | 0.000 | -0.051 | 0.051 | 0.043 | 0.008 | 0.051 | 0.010 | 0.194 | 0.194 | 0.187 | 0.064 | 0.194 | 0.000 | 0.040 | 0.040 | 0.037 | 0.004 | 0.040 | 1.000 | 0.951 | 0.079 | 0.075 | 0.071 | 0.077 |  |  |  |
|  | Egger | 1.6 | 0.8 | 0.8 | 0.5 | 0.3 | -0.001 | -0.068 | 0.067 | 0.059 | 0.009 | 0.067 | 0.039 | 0.210 | 0.209 | 0.202 | 0.072 | 0.209 | 0.002 | 0.049 | 0.048 | 0.044 | 0.005 | 0.048 | 1.000 | 0.661 | 0.871 | 0.463 | 0.935 | 0.870 | 0.003 | 0.047 | 0.813 |
|  |  | 0.6 | 0.8 | -0.2 | -0.5 | 0.3 | 0.002 | -0.058 | 0.060 | 0.050 | 0.010 | 0.060 | 0.036 | 0.205 | 0.205 | 0.197 | 0.070 | 0.205 | 0.001 | 0.045 | 0.046 | 0.041 | 0.005 | 0.046 | 1.000 | 0.916 | 0.111 | 0.633 | 0.995 | 0.113 | 0.054 | 0.053 | 0.797 |
|  |  | 0.7 | 0.5 | 0.2 | 0.5 | -0.3 | 0.001 | -0.060 | 0.061 | 0.056 | 0.005 | 0.061 | 0.037 | 0.207 | 0.208 | 0.196 | 0.070 | 0.208 | 0.001 | 0.046 | 0.047 | 0.042 | 0.005 | 0.047 | 1.000 | 0.526 | 0.244 | 0.791 | 0.991 | 0.243 | 0.046 | 0.044 | 0.801 |
|  |  | -0.3 | 0.5 | -0.8 | -0.5 | -0.3 | -0.002 | -0.068 | 0.066 | 0.058 | 0.008 | 0.066 | 0.036 | 0.213 | 0.210 | 0.204 | 0.070 | 0.210 | 0.001 | 0.050 | 0.048 | 0.045 | 0.005 | 0.048 | 1.000 | 0.505 | 0.942 | 0.602 | 0.994 | 0.939 | 0.045 | 0.048 | 0.775 |
|  |  | 0 | -0.8 | 0.8 | 0.5 | 0.3 | 0.002 | -0.069 | 0.071 | 0.063 | 0.009 | 0.072 | 0.040 | 0.211 | 0.213 | 0.206 | 0.074 | 0.213 | 0.002 | 0.049 | 0.051 | 0.046 | 0.006 | 0.051 | 0.058 | 0.970 | 0.976 | 0.790 | 0.991 | 0.977 | 0.057 | 0.057 | 0.794 |
|  |  | -1.6 | -0.8 | -0.8 | -0.5 | -0.3 | -0.001 | -0.057 | 0.056 | 0.049 | 0.007 | 0.056 | 0.037 | 0.203 | 0.205 | 0.199 | 0.070 | 0.205 | 0.001 | 0.045 | 0.045 | 0.042 | 0.005 | 0.045 | 1.000 | 0.896 | 0.819 | 0.391 | 0.955 | 0.818 | 0.006 | 0.055 | 0.813 |
|  |  | 0.8 | 0 | 0.8 | 0.5 | 0.3 | 0.005 | -0.053 | 0.058 | 0.046 | 0.011 | 0.058 | 0.038 | 0.213 | 0.213 | 0.204 | 0.072 | 0.213 | 0.001 | 0.048 | 0.049 | 0.044 | 0.005 | 0.049 | 1.000 | 0.051 | 0.967 | 0.760 | 0.994 | 0.969 | 0.034 | 0.058 | 0.803 |
|  |  | 1.3 | 0.8 | 0.5 | 0.5 | 0 | -0.001 | -0.074 | 0.074 | 0.064 | 0.010 | 0.074 | 0.030 | 0.209 | 0.207 | 0.196 | 0.068 | 0.207 | 0.001 | 0.049 | 0.048 | 0.043 | 0.005 | 0.048 | 1.000 | 0.831 | 0.674 | 0.711 | 0.024 | 0.674 | 0.016 | 0.044 | 0.797 |
|  |  | 1.3 | 0.8 | 0.5 | 0.5 | 0 | 0.002 | -0.052 | 0.054 | 0.054 | 0.000 | 0.054 | 0.038 | 0.202 | 0.204 | 0.203 | 0.026 | 0.204 | 0.001 | 0.044 | 0.044 | 0.044 | 0.001 | 0.044 | 1.000 | 0.835 | 0.584 | 0.606 | 0.058 | 0.600 | 0.006 | 0.059 | 0.813 |
|  |  | 0.8 | 0.8 | 0 | 0 | 0 | 0.000 | -0.059 | 0.059 | 0.052 | 0.007 | 0.059 | 0.024 | 0.219 | 0.217 | 0.208 | 0.066 | 0.217 | 0.001 | 0.051 | 0.050 | 0.046 | 0.004 | 0.050 | 1.000 | 0.913 | 0.078 | 0.082 | 0.071 | 0.083 | 0.045 | 0.056 | 0.759 |

TE: total effect; DE: direct effect; IE: indirect effect; IE_d: indirect effect calculated by difference method; IE_p: indirect effect calculated by product method; IE1: *X*→*M_1_*→*Y*; IE2: *X*→*M_2_*→*Y*.

**eTable 11.** Simulation of PSE-Egger in two non-ordered mediators setting with different effects (case(b),sc34-37).

|  |  | TRUE | | | | | BIAS | | | | | | SE | | | | | | MSE | | | | | | Power/Type I error | | | | | | | | |
| --- | --- | --- | --- | --- | --- | --- | --- | --- | --- | --- | --- | --- | --- | --- | --- | --- | --- | --- | --- | --- | --- | --- | --- | --- | --- | --- | --- | --- | --- | --- | --- | --- | --- |
|  |  | TE | DE | IE | IE2 | IE1 | TE | DE | IE_d | IE1 | IE2 | IE_p | TE | DE | IE_d | IE1 | IE2 | IE_p | TE | DE | IE_d | IE1 | IE2 | IE_p | TE | DE | IE_d | IE1 | IE2 | IE_p | GY | GM1 | GM2 |
| Sc34 | IVW | 1.6 | 0.8 | 0.8 | 0.5 | 0.3 | 0.074 | -0.054 | 0.128 | 0.074 | 0.055 | 0.128 | 0.014 | 0.187 | 0.187 | 0.186 | 0.119 | 0.187 | 0.006 | 0.038 | 0.051 | 0.040 | 0.017 | 0.052 | 1.000 | 0.785 | 0.953 | 0.521 | 0.514 | 0.884 |  |  |  |
|  |  | 0.6 | 0.8 | -0.2 | -0.5 | 0.3 | 0.000 | -0.052 | 0.052 | -0.007 | 0.058 | 0.052 | 0.012 | 0.185 | 0.184 | 0.188 | 0.123 | 0.184 | 0.000 | 0.037 | 0.037 | 0.035 | 0.019 | 0.037 | 1.000 | 0.960 | 0.136 | 0.774 | 0.838 | 0.072 |  |  |  |
|  |  | 0.7 | 0.5 | 0.2 | 0.5 | -0.3 | 0.000 | -0.051 | 0.051 | 0.067 | -0.016 | 0.051 | 0.012 | 0.192 | 0.192 | 0.196 | 0.122 | 0.192 | 0.000 | 0.039 | 0.039 | 0.043 | 0.015 | 0.039 | 1.000 | 0.609 | 0.295 | 0.841 | 0.749 | 0.174 |  |  |  |
|  |  | -0.3 | 0.5 | -0.8 | -0.5 | -0.3 | -0.075 | -0.048 | -0.027 | -0.009 | -0.017 | -0.027 | 0.013 | 0.185 | 0.185 | 0.185 | 0.118 | 0.185 | 0.006 | 0.036 | 0.035 | 0.034 | 0.014 | 0.035 | 1.000 | 0.655 | 0.985 | 0.769 | 0.756 | 0.965 |  |  |  |
|  |  | 0 | -0.8 | 0.8 | 0.5 | 0.3 | 0.075 | -0.043 | 0.119 | 0.069 | 0.049 | 0.118 | 0.015 | 0.191 | 0.190 | 0.189 | 0.119 | 0.190 | 0.006 | 0.038 | 0.050 | 0.040 | 0.017 | 0.050 | 1.000 | 0.986 | 0.997 | 0.845 | 0.831 | 0.991 |  |  |  |
|  |  | -1.6 | -0.8 | -0.8 | -0.5 | -0.3 | -0.074 | -0.048 | -0.027 | -0.007 | -0.020 | -0.027 | 0.014 | 0.187 | 0.187 | 0.190 | 0.119 | 0.187 | 0.006 | 0.037 | 0.036 | 0.036 | 0.014 | 0.036 | 1.000 | 0.949 | 0.941 | 0.531 | 0.503 | 0.872 |  |  |  |
|  |  | 0.8 | 0 | 0.8 | 0.5 | 0.3 | 0.075 | -0.050 | 0.125 | 0.073 | 0.053 | 0.125 | 0.014 | 0.187 | 0.187 | 0.191 | 0.120 | 0.187 | 0.006 | 0.038 | 0.051 | 0.042 | 0.017 | 0.051 | 1.000 | 0.037 | 0.997 | 0.833 | 0.811 | 0.990 |  |  |  |
|  |  | 1.3 | 0.8 | 0.5 | 0.5 | 0 | 0.037 | -0.050 | 0.087 | 0.071 | 0.016 | 0.087 | 0.012 | 0.187 | 0.187 | 0.193 | 0.117 | 0.187 | 0.001 | 0.038 | 0.043 | 0.042 | 0.014 | 0.043 | 1.000 | 0.932 | 0.806 | 0.766 | 0.023 | 0.649 |  |  |  |
|  |  | 1.3 | 0.8 | 0.5 | 0.5 | 0 | 0.074 | -0.032 | 0.107 | 0.067 | 0.039 | 0.107 | 0.014 | 0.184 | 0.185 | 0.189 | 0.015 | 0.185 | 0.006 | 0.035 | 0.045 | 0.040 | 0.002 | 0.045 | 1.000 | 0.898 | 0.747 | 0.649 | 0.655 | 0.733 |  |  |  |
|  |  | 0.8 | 0.8 | 0 | 0 | 0 | 0.000 | -0.036 | 0.036 | 0.021 | 0.016 | 0.036 | 0.010 | 0.185 | 0.185 | 0.185 | 0.119 | 0.185 | 0.000 | 0.036 | 0.036 | 0.035 | 0.014 | 0.036 | 1.000 | 0.974 | 0.058 | 0.047 | 0.071 | 0.026 |  |  |  |
|  | Egger | 1.6 | 0.8 | 0.8 | 0.5 | 0.3 | 0.001 | -0.075 | 0.076 | 0.051 | 0.026 | 0.076 | 0.034 | 0.222 | 0.221 | 0.194 | 0.120 | 0.221 | 0.001 | 0.055 | 0.055 | 0.040 | 0.015 | 0.055 | 1.000 | 0.550 | 0.809 | 0.414 | 0.417 | 0.767 | 0.002 | 0.800 | 0.781 |
|  |  | 0.6 | 0.8 | -0.2 | -0.5 | 0.3 | 0.000 | -0.079 | 0.079 | 0.048 | 0.031 | 0.079 | 0.030 | 0.225 | 0.224 | 0.200 | 0.125 | 0.225 | 0.001 | 0.057 | 0.057 | 0.042 | 0.017 | 0.057 | 1.000 | 0.865 | 0.085 | 0.633 | 0.770 | 0.075 | 0.051 | 0.836 | 0.777 |
|  |  | 0.7 | 0.5 | 0.2 | 0.5 | -0.3 | 0.000 | -0.077 | 0.077 | 0.047 | 0.030 | 0.077 | 0.030 | 0.231 | 0.230 | 0.208 | 0.121 | 0.230 | 0.001 | 0.059 | 0.059 | 0.046 | 0.015 | 0.059 | 1.000 | 0.449 | 0.245 | 0.772 | 0.624 | 0.224 | 0.046 | 0.814 | 0.788 |
|  |  | -0.3 | 0.5 | -0.8 | -0.5 | -0.3 | 0.000 | -0.080 | 0.080 | 0.049 | 0.031 | 0.080 | 0.030 | 0.224 | 0.223 | 0.199 | 0.116 | 0.223 | 0.001 | 0.056 | 0.056 | 0.042 | 0.014 | 0.056 | 1.000 | 0.450 | 0.870 | 0.622 | 0.622 | 0.856 | 0.055 | 0.809 | 0.811 |
|  |  | 0 | -0.8 | 0.8 | 0.5 | 0.3 | 0.002 | -0.073 | 0.075 | 0.053 | 0.022 | 0.075 | 0.034 | 0.228 | 0.226 | 0.198 | 0.121 | 0.226 | 0.001 | 0.057 | 0.057 | 0.042 | 0.015 | 0.057 | 0.057 | 0.951 | 0.963 | 0.798 | 0.749 | 0.958 | 0.052 | 0.804 | 0.792 |
|  |  | -1.6 | -0.8 | -0.8 | -0.5 | -0.3 | 0.000 | -0.081 | 0.081 | 0.051 | 0.030 | 0.081 | 0.032 | 0.223 | 0.222 | 0.200 | 0.120 | 0.222 | 0.001 | 0.057 | 0.056 | 0.043 | 0.015 | 0.056 | 1.000 | 0.854 | 0.717 | 0.372 | 0.351 | 0.678 | 0.006 | 0.823 | 0.800 |
|  |  | 0.8 | 0 | 0.8 | 0.5 | 0.3 | 0.001 | -0.077 | 0.078 | 0.052 | 0.026 | 0.078 | 0.034 | 0.222 | 0.222 | 0.202 | 0.120 | 0.222 | 0.001 | 0.055 | 0.055 | 0.043 | 0.015 | 0.055 | 1.000 | 0.039 | 0.955 | 0.765 | 0.728 | 0.952 | 0.051 | 0.814 | 0.799 |
|  |  | 1.3 | 0.8 | 0.5 | 0.5 | 0 | 0.001 | -0.080 | 0.080 | 0.054 | 0.026 | 0.080 | 0.029 | 0.219 | 0.218 | 0.199 | 0.118 | 0.218 | 0.001 | 0.054 | 0.054 | 0.042 | 0.015 | 0.054 | 1.000 | 0.791 | 0.633 | 0.681 | 0.024 | 0.592 | 0.020 | 0.809 | 0.810 |
|  |  | 1.3 | 0.8 | 0.5 | 0.5 | 0 | 0.001 | -0.045 | 0.046 | 0.046 | 0.000 | 0.046 | 0.033 | 0.200 | 0.200 | 0.200 | 0.016 | 0.200 | 0.001 | 0.042 | 0.042 | 0.042 | 0.000 | 0.042 | 1.000 | 0.824 | 0.532 | 0.551 | 0.021 | 0.548 | 0.006 | 0.822 | 0.802 |
|  |  | 0.8 | 0.8 | 0 | 0 | 0 | 0.001 | -0.068 | 0.069 | 0.042 | 0.027 | 0.069 | 0.025 | 0.223 | 0.222 | 0.196 | 0.117 | 0.222 | 0.001 | 0.054 | 0.054 | 0.040 | 0.014 | 0.054 | 1.000 | 0.873 | 0.060 | 0.055 | 0.069 | 0.055 | 0.055 | 0.807 | 0.815 |
| Sc35 | IVW | 1.6 | 0.8 | 0.8 | 0.5 | 0.3 | 0.075 | -0.158 | 0.233 | 0.226 | 0.007 | 0.233 | 0.014 | 0.109 | 0.111 | 0.106 | 0.041 | 0.111 | 0.006 | 0.037 | 0.067 | 0.063 | 0.002 | 0.067 | 1.000 | 0.675 | 0.997 | 0.916 | 0.978 | 0.998 |  |  |  |
|  |  | 0.6 | 0.8 | -0.2 | -0.5 | 0.3 | 0.000 | -0.152 | 0.153 | 0.148 | 0.004 | 0.153 | 0.012 | 0.114 | 0.114 | 0.108 | 0.040 | 0.114 | 0.000 | 0.036 | 0.036 | 0.034 | 0.002 | 0.036 | 1.000 | 0.996 | 0.052 | 0.878 | 1.000 | 0.054 |  |  |  |
|  |  | 0.7 | 0.5 | 0.2 | 0.5 | -0.3 | 0.074 | -0.161 | 0.235 | 0.229 | 0.006 | 0.235 | 0.013 | 0.111 | 0.111 | 0.103 | 0.039 | 0.111 | 0.006 | 0.038 | 0.068 | 0.063 | 0.002 | 0.067 | 1.000 | 0.703 | 0.914 | 1.000 | 1.000 | 0.913 |  |  |  |
|  |  | -0.3 | 0.5 | -0.8 | -0.5 | -0.3 | 0.000 | -0.153 | 0.153 | 0.146 | 0.006 | 0.153 | 0.012 | 0.109 | 0.110 | 0.103 | 0.040 | 0.110 | 0.000 | 0.035 | 0.035 | 0.032 | 0.002 | 0.035 | 1.000 | 0.852 | 0.999 | 0.915 | 1.000 | 0.999 |  |  |  |
|  |  | 0 | -0.8 | 0.8 | 0.5 | 0.3 | 0.075 | -0.157 | 0.232 | 0.225 | 0.006 | 0.232 | 0.014 | 0.109 | 0.110 | 0.105 | 0.039 | 0.109 | 0.006 | 0.037 | 0.066 | 0.062 | 0.002 | 0.066 | 1.000 | 1.000 | 1.000 | 0.999 | 1.000 | 1.000 |  |  |  |
|  |  | -1.6 | -0.8 | -0.8 | -0.5 | -0.3 | -0.001 | -0.158 | 0.158 | 0.153 | 0.005 | 0.157 | 0.012 | 0.112 | 0.112 | 0.107 | 0.040 | 0.112 | 0.000 | 0.038 | 0.037 | 0.035 | 0.002 | 0.037 | 1.000 | 0.993 | 0.890 | 0.177 | 0.997 | 0.887 |  |  |  |
|  |  | 0.8 | 0 | 0.8 | 0.5 | 0.3 | 0.075 | -0.156 | 0.231 | 0.226 | 0.005 | 0.231 | 0.014 | 0.110 | 0.110 | 0.106 | 0.039 | 0.109 | 0.006 | 0.036 | 0.065 | 0.062 | 0.002 | 0.065 | 1.000 | 0.026 | 1.000 | 0.997 | 0.999 | 1.000 |  |  |  |
|  |  | 1.3 | 0.8 | 0.5 | 0.5 | 0 | 0.074 | -0.162 | 0.237 | 0.232 | 0.005 | 0.237 | 0.009 | 0.107 | 0.107 | 0.101 | 0.038 | 0.107 | 0.006 | 0.038 | 0.068 | 0.064 | 0.001 | 0.068 | 1.000 | 0.910 | 0.974 | 0.982 | 0.000 | 0.971 |  |  |  |
|  |  | 1.3 | 0.8 | 0.5 | 0.5 | 0 | 0.075 | -0.155 | 0.229 | 0.229 | 0.000 | 0.229 | 0.014 | 0.107 | 0.107 | 0.107 | 0.011 | 0.107 | 0.006 | 0.035 | 0.064 | 0.064 | 0.000 | 0.064 | 1.000 | 0.901 | 0.974 | 0.973 | 0.069 | 0.972 |  |  |  |
|  |  | 0.8 | 0.8 | 0 | 0 | 0 | 0.037 | -0.163 | 0.200 | 0.193 | 0.007 | 0.200 | 0.006 | 0.111 | 0.111 | 0.107 | 0.039 | 0.111 | 0.001 | 0.039 | 0.052 | 0.049 | 0.002 | 0.052 | 1.000 | 0.994 | 0.313 | 0.314 | 0.021 | 0.288 |  |  |  |
|  | Egger | 1.6 | 0.8 | 0.8 | 0.5 | 0.3 | 0.003 | -0.062 | 0.066 | 0.055 | 0.011 | 0.066 | 0.031 | 0.113 | 0.117 | 0.112 | 0.043 | 0.117 | 0.001 | 0.017 | 0.018 | 0.016 | 0.002 | 0.018 | 1.000 | 0.730 | 0.947 | 0.400 | 0.984 | 0.940 | 0.000 | 0.058 | 0.808 |
|  |  | 0.6 | 0.8 | -0.2 | -0.5 | 0.3 | 0.002 | -0.059 | 0.060 | 0.052 | 0.009 | 0.060 | 0.029 | 0.118 | 0.119 | 0.112 | 0.044 | 0.119 | 0.001 | 0.017 | 0.018 | 0.015 | 0.002 | 0.018 | 1.000 | 1.000 | 0.163 | 0.963 | 1.000 | 0.160 | 0.440 | 0.036 | 0.771 |
|  |  | 0.7 | 0.5 | 0.2 | 0.5 | -0.3 | 0.001 | -0.065 | 0.066 | 0.057 | 0.008 | 0.066 | 0.032 | 0.112 | 0.115 | 0.108 | 0.043 | 0.115 | 0.001 | 0.017 | 0.018 | 0.015 | 0.002 | 0.018 | 1.000 | 0.853 | 0.411 | 0.986 | 1.000 | 0.393 | 0.290 | 0.052 | 0.789 |
|  |  | -0.3 | 0.5 | -0.8 | -0.5 | -0.3 | 0.000 | -0.056 | 0.056 | 0.047 | 0.009 | 0.055 | 0.031 | 0.113 | 0.116 | 0.109 | 0.044 | 0.116 | 0.001 | 0.016 | 0.017 | 0.014 | 0.002 | 0.017 | 1.000 | 0.961 | 1.000 | 0.976 | 1.000 | 1.000 | 0.558 | 0.047 | 0.796 |
|  |  | 0 | -0.8 | 0.8 | 0.5 | 0.3 | 0.002 | -0.060 | 0.062 | 0.053 | 0.009 | 0.062 | 0.034 | 0.111 | 0.113 | 0.107 | 0.043 | 0.113 | 0.001 | 0.016 | 0.017 | 0.014 | 0.002 | 0.017 | 0.064 | 1.000 | 1.000 | 0.993 | 1.000 | 1.000 | 0.420 | 0.053 | 0.788 |
|  |  | -1.6 | -0.8 | -0.8 | -0.5 | -0.3 | -0.001 | -0.062 | 0.061 | 0.054 | 0.007 | 0.061 | 0.029 | 0.116 | 0.119 | 0.111 | 0.043 | 0.119 | 0.001 | 0.017 | 0.018 | 0.015 | 0.002 | 0.018 | 1.000 | 0.963 | 0.914 | 0.321 | 0.998 | 0.911 | 0.007 | 0.047 | 0.796 |
|  |  | 0.8 | 0 | 0.8 | 0.5 | 0.3 | 0.003 | -0.059 | 0.062 | 0.053 | 0.009 | 0.062 | 0.034 | 0.117 | 0.119 | 0.113 | 0.046 | 0.119 | 0.001 | 0.017 | 0.018 | 0.016 | 0.002 | 0.018 | 1.000 | 0.002 | 0.999 | 0.897 | 1.000 | 0.998 | 0.061 | 0.049 | 0.783 |
|  |  | 1.3 | 0.8 | 0.5 | 0.5 | 0 | 0.001 | -0.064 | 0.065 | 0.056 | 0.008 | 0.065 | 0.020 | 0.110 | 0.111 | 0.107 | 0.036 | 0.111 | 0.000 | 0.016 | 0.016 | 0.015 | 0.001 | 0.016 | 1.000 | 0.935 | 0.727 | 0.758 | 0.000 | 0.729 | 0.013 | 0.047 | 0.795 |
|  |  | 1.3 | 0.8 | 0.5 | 0.5 | 0 | 0.001 | -0.054 | 0.056 | 0.056 | 0.000 | 0.056 | 0.033 | 0.114 | 0.117 | 0.114 | 0.026 | 0.117 | 0.001 | 0.016 | 0.017 | 0.016 | 0.001 | 0.017 | 1.000 | 0.924 | 0.610 | 0.630 | 0.068 | 0.633 | 0.004 | 0.055 | 0.800 |
|  |  | 0.8 | 0.8 | 0 | 0 | 0 | 0.000 | -0.064 | 0.064 | 0.055 | 0.009 | 0.064 | 0.013 | 0.116 | 0.116 | 0.113 | 0.036 | 0.116 | 0.000 | 0.018 | 0.018 | 0.016 | 0.001 | 0.018 | 1.000 | 0.995 | 0.023 | 0.025 | 0.013 | 0.025 | 0.296 | 0.046 | 0.797 |
| Sc36 | IVW | 1.6 | 0.8 | 0.8 | 0.5 | 0.3 | 0.076 | -0.090 | 0.166 | 0.012 | 0.155 | 0.166 | 0.014 | 0.089 | 0.090 | 0.066 | 0.066 | 0.090 | 0.006 | 0.016 | 0.036 | 0.005 | 0.028 | 0.036 | 1.000 | 0.972 | 1.000 | 0.986 | 0.904 | 1.000 |  |  |  |
|  |  | 0.6 | 0.8 | -0.2 | -0.5 | 0.3 | 0.075 | -0.089 | 0.164 | 0.010 | 0.154 | 0.164 | 0.013 | 0.093 | 0.094 | 0.066 | 0.070 | 0.094 | 0.006 | 0.017 | 0.036 | 0.004 | 0.029 | 0.036 | 1.000 | 1.000 | 0.040 | 1.000 | 1.000 | 0.043 |  |  |  |
|  |  | 0.7 | 0.5 | 0.2 | 0.5 | -0.3 | 0.001 | -0.092 | 0.093 | 0.009 | 0.084 | 0.093 | 0.012 | 0.087 | 0.088 | 0.065 | 0.064 | 0.088 | 0.000 | 0.016 | 0.016 | 0.004 | 0.011 | 0.016 | 1.000 | 0.976 | 0.829 | 1.000 | 0.834 | 0.824 |  |  |  |
|  |  | -0.3 | 0.5 | -0.8 | -0.5 | -0.3 | -0.001 | -0.093 | 0.092 | 0.011 | 0.082 | 0.092 | 0.012 | 0.091 | 0.092 | 0.066 | 0.065 | 0.092 | 0.000 | 0.017 | 0.017 | 0.004 | 0.011 | 0.017 | 1.000 | 0.979 | 1.000 | 1.000 | 0.893 | 1.000 |  |  |  |
|  |  | 0 | -0.8 | 0.8 | 0.5 | 0.3 | 0.074 | -0.092 | 0.167 | 0.012 | 0.155 | 0.167 | 0.014 | 0.087 | 0.087 | 0.064 | 0.066 | 0.087 | 0.006 | 0.016 | 0.035 | 0.004 | 0.028 | 0.035 | 0.999 | 1.000 | 1.000 | 1.000 | 1.000 | 1.000 |  |  |  |
|  |  | -1.6 | -0.8 | -0.8 | -0.5 | -0.3 | 0.000 | -0.094 | 0.094 | 0.009 | 0.085 | 0.094 | 0.012 | 0.090 | 0.090 | 0.065 | 0.066 | 0.090 | 0.000 | 0.017 | 0.017 | 0.004 | 0.012 | 0.017 | 1.000 | 1.000 | 0.994 | 0.994 | 0.183 | 0.995 |  |  |  |
|  |  | 0.8 | 0 | 0.8 | 0.5 | 0.3 | 0.075 | -0.094 | 0.170 | 0.010 | 0.160 | 0.170 | 0.014 | 0.087 | 0.087 | 0.064 | 0.063 | 0.087 | 0.006 | 0.016 | 0.036 | 0.004 | 0.030 | 0.036 | 1.000 | 0.005 | 1.000 | 1.000 | 0.996 | 1.000 |  |  |  |
|  |  | 1.3 | 0.8 | 0.5 | 0.5 | 0 | 0.037 | -0.090 | 0.127 | 0.005 | 0.122 | 0.127 | 0.012 | 0.093 | 0.093 | 0.068 | 0.067 | 0.093 | 0.002 | 0.017 | 0.025 | 0.005 | 0.019 | 0.025 | 1.000 | 0.997 | 0.986 | 0.998 | 0.027 | 0.989 |  |  |  |
|  |  | 1.3 | 0.8 | 0.5 | 0.5 | 0 | 0.075 | 0.016 | 0.059 | 0.008 | 0.051 | 0.059 | 0.014 | 0.065 | 0.066 | 0.065 | 0.011 | 0.066 | 0.006 | 0.004 | 0.008 | 0.004 | 0.003 | 0.008 | 1.000 | 1.000 | 1.000 | 1.000 | 0.974 | 1.000 |  |  |  |
|  |  | 0.8 | 0.8 | 0 | 0 | 0 | 0.037 | -0.096 | 0.133 | 0.013 | 0.120 | 0.133 | 0.006 | 0.088 | 0.088 | 0.063 | 0.065 | 0.088 | 0.001 | 0.017 | 0.025 | 0.004 | 0.019 | 0.026 | 1.000 | 1.000 | 0.184 | 0.019 | 0.316 | 0.167 |  |  |  |
|  | Egger | 1.6 | 0.8 | 0.8 | 0.5 | 0.3 | 0.004 | -0.045 | 0.048 | 0.019 | 0.030 | 0.049 | 0.032 | 0.086 | 0.090 | 0.068 | 0.067 | 0.090 | 0.001 | 0.009 | 0.011 | 0.005 | 0.005 | 0.011 | 1.000 | 0.974 | 0.998 | 0.988 | 0.398 | 0.999 | 0.000 | 0.810 | 0.051 |
|  |  | 0.6 | 0.8 | -0.2 | -0.5 | 0.3 | 0.001 | -0.044 | 0.046 | 0.014 | 0.032 | 0.045 | 0.031 | 0.087 | 0.090 | 0.065 | 0.068 | 0.090 | 0.001 | 0.010 | 0.010 | 0.004 | 0.006 | 0.010 | 1.000 | 1.000 | 0.247 | 1.000 | 0.988 | 0.230 | 0.399 | 0.782 | 0.047 |
|  |  | 0.7 | 0.5 | 0.2 | 0.5 | -0.3 | 0.002 | -0.046 | 0.048 | 0.015 | 0.032 | 0.047 | 0.031 | 0.086 | 0.090 | 0.067 | 0.065 | 0.090 | 0.001 | 0.009 | 0.010 | 0.005 | 0.005 | 0.010 | 1.000 | 0.986 | 0.645 | 1.000 | 0.948 | 0.631 | 0.348 | 0.797 | 0.048 |
|  |  | -0.3 | 0.5 | -0.8 | -0.5 | -0.3 | -0.001 | -0.043 | 0.042 | 0.014 | 0.028 | 0.042 | 0.031 | 0.089 | 0.091 | 0.064 | 0.066 | 0.091 | 0.001 | 0.010 | 0.010 | 0.004 | 0.005 | 0.010 | 1.000 | 0.992 | 1.000 | 1.000 | 0.971 | 1.000 | 0.583 | 0.817 | 0.055 |
|  |  | 0 | -0.8 | 0.8 | 0.5 | 0.3 | 0.002 | -0.045 | 0.047 | 0.015 | 0.032 | 0.047 | 0.033 | 0.088 | 0.093 | 0.066 | 0.069 | 0.093 | 0.001 | 0.010 | 0.011 | 0.005 | 0.006 | 0.011 | 0.064 | 1.000 | 1.000 | 1.000 | 0.992 | 1.000 | 0.412 | 0.799 | 0.051 |
|  |  | -1.6 | -0.8 | -0.8 | -0.5 | -0.3 | 0.000 | -0.049 | 0.049 | 0.013 | 0.036 | 0.049 | 0.031 | 0.088 | 0.090 | 0.064 | 0.067 | 0.090 | 0.001 | 0.010 | 0.010 | 0.004 | 0.006 | 0.010 | 1.000 | 0.997 | 0.996 | 0.995 | 0.316 | 0.997 | 0.003 | 0.822 | 0.052 |
|  |  | 0.8 | 0 | 0.8 | 0.5 | 0.3 | 0.002 | -0.048 | 0.050 | 0.014 | 0.036 | 0.050 | 0.032 | 0.082 | 0.085 | 0.064 | 0.064 | 0.085 | 0.001 | 0.009 | 0.010 | 0.004 | 0.005 | 0.010 | 1.000 | 0.000 | 1.000 | 0.999 | 0.914 | 1.000 | 0.054 | 0.807 | 0.041 |
|  |  | 1.3 | 0.8 | 0.5 | 0.5 | 0 | 0.001 | -0.044 | 0.045 | 0.012 | 0.034 | 0.045 | 0.028 | 0.089 | 0.091 | 0.066 | 0.066 | 0.091 | 0.001 | 0.010 | 0.010 | 0.004 | 0.005 | 0.010 | 1.000 | 0.999 | 0.948 | 0.999 | 0.001 | 0.951 | 0.017 | 0.812 | 0.033 |
|  |  | 1.3 | 0.8 | 0.5 | 0.5 | 0 | 0.002 | -0.012 | 0.013 | 0.012 | 0.001 | 0.014 | 0.032 | 0.062 | 0.067 | 0.065 | 0.016 | 0.067 | 0.001 | 0.004 | 0.005 | 0.004 | 0.000 | 0.005 | 1.000 | 1.000 | 0.998 | 0.999 | 0.025 | 0.999 | 0.004 | 0.810 | 0.045 |
|  |  | 0.8 | 0.8 | 0 | 0 | 0 | 0.000 | -0.048 | 0.049 | 0.016 | 0.033 | 0.049 | 0.013 | 0.087 | 0.087 | 0.061 | 0.065 | 0.087 | 0.000 | 0.010 | 0.010 | 0.004 | 0.005 | 0.010 | 1.000 | 1.000 | 0.027 | 0.011 | 0.028 | 0.023 | 0.334 | 0.802 | 0.050 |
| Sc37 | IVW | 1.6 | 0.8 | 0.8 | 0.5 | 0.3 | 0.038 | -0.101 | 0.138 | 0.212 | -0.073 | 0.138 | 0.010 | 0.129 | 0.129 | 0.109 | 0.054 | 0.129 | 0.002 | 0.027 | 0.036 | 0.057 | 0.008 | 0.036 | 1.000 | 0.574 | 0.945 | 0.892 | 0.331 | 0.968 |  |  |  |
|  |  | 0.6 | 0.8 | -0.2 | -0.5 | 0.3 | -0.037 | -0.100 | 0.063 | 0.139 | -0.076 | 0.063 | 0.009 | 0.131 | 0.130 | 0.111 | 0.053 | 0.130 | 0.001 | 0.027 | 0.021 | 0.031 | 0.009 | 0.021 | 1.000 | 0.999 | 0.154 | 0.885 | 0.973 | 0.189 |  |  |  |
|  |  | 0.7 | 0.5 | 0.2 | 0.5 | -0.3 | 0.112 | -0.095 | 0.207 | 0.205 | 0.002 | 0.207 | 0.012 | 0.131 | 0.131 | 0.108 | 0.055 | 0.131 | 0.013 | 0.026 | 0.060 | 0.054 | 0.003 | 0.060 | 1.000 | 0.698 | 0.745 | 0.999 | 0.996 | 0.796 |  |  |  |
|  |  | -0.3 | 0.5 | -0.8 | -0.5 | -0.3 | 0.037 | -0.104 | 0.141 | 0.140 | 0.001 | 0.141 | 0.009 | 0.130 | 0.130 | 0.106 | 0.055 | 0.130 | 0.001 | 0.028 | 0.037 | 0.031 | 0.003 | 0.037 | 1.000 | 0.834 | 0.998 | 0.914 | 0.998 | 1.000 |  |  |  |
|  |  | 0 | -0.8 | 0.8 | 0.5 | 0.3 | 0.037 | -0.102 | 0.139 | 0.211 | -0.073 | 0.139 | 0.011 | 0.135 | 0.135 | 0.112 | 0.053 | 0.135 | 0.002 | 0.028 | 0.037 | 0.057 | 0.008 | 0.037 | 0.911 | 0.999 | 0.999 | 0.999 | 0.973 | 1.000 |  |  |  |
|  |  | -1.6 | -0.8 | -0.8 | -0.5 | -0.3 | 0.037 | -0.103 | 0.140 | 0.139 | 0.001 | 0.141 | 0.009 | 0.124 | 0.124 | 0.101 | 0.053 | 0.124 | 0.001 | 0.026 | 0.035 | 0.030 | 0.003 | 0.035 | 1.000 | 0.975 | 0.791 | 0.224 | 0.886 | 0.868 |  |  |  |
|  |  | 0.8 | 0 | 0.8 | 0.5 | 0.3 | 0.037 | -0.103 | 0.141 | 0.215 | -0.074 | 0.141 | 0.010 | 0.133 | 0.133 | 0.112 | 0.053 | 0.133 | 0.002 | 0.028 | 0.038 | 0.058 | 0.008 | 0.038 | 1.000 | 0.009 | 0.996 | 0.995 | 0.844 | 1.000 |  |  |  |
|  |  | 1.3 | 0.8 | 0.5 | 0.5 | 0 | 0.074 | -0.103 | 0.178 | 0.215 | -0.037 | 0.178 | 0.010 | 0.128 | 0.128 | 0.105 | 0.052 | 0.128 | 0.006 | 0.027 | 0.048 | 0.057 | 0.004 | 0.048 | 1.000 | 0.854 | 0.850 | 0.972 | 0.000 | 0.908 |  |  |  |
|  |  | 1.3 | 0.8 | 0.5 | 0.5 | 0 | 0.038 | -0.141 | 0.179 | 0.211 | -0.033 | 0.178 | 0.011 | 0.101 | 0.102 | 0.103 | 0.009 | 0.102 | 0.002 | 0.030 | 0.042 | 0.055 | 0.001 | 0.042 | 1.000 | 0.923 | 0.954 | 0.961 | 0.567 | 0.952 |  |  |  |
|  |  | 0.8 | 0.8 | 0 | 0 | 0 | 0.037 | -0.105 | 0.142 | 0.177 | -0.035 | 0.142 | 0.006 | 0.132 | 0.132 | 0.108 | 0.051 | 0.132 | 0.001 | 0.028 | 0.038 | 0.043 | 0.004 | 0.038 | 1.000 | 0.994 | 0.123 | 0.252 | 0.047 | 0.147 |  |  |  |
|  | Egger | 1.6 | 0.8 | 0.8 | 0.5 | 0.3 | 0.002 | -0.078 | 0.080 | 0.051 | 0.029 | 0.080 | 0.026 | 0.125 | 0.126 | 0.112 | 0.070 | 0.126 | 0.001 | 0.022 | 0.022 | 0.015 | 0.006 | 0.022 | 1.000 | 0.593 | 0.907 | 0.432 | 0.418 | 0.899 | 0.000 | 0.829 | 0.795 |
|  |  | 0.6 | 0.8 | -0.2 | -0.5 | 0.3 | 0.000 | -0.077 | 0.076 | 0.051 | 0.025 | 0.076 | 0.023 | 0.121 | 0.121 | 0.113 | 0.067 | 0.121 | 0.001 | 0.021 | 0.021 | 0.015 | 0.005 | 0.021 | 1.000 | 0.999 | 0.111 | 0.965 | 0.987 | 0.094 | 0.406 | 0.813 | 0.758 |
|  |  | 0.7 | 0.5 | 0.2 | 0.5 | -0.3 | 0.002 | -0.073 | 0.075 | 0.045 | 0.030 | 0.075 | 0.023 | 0.124 | 0.124 | 0.108 | 0.069 | 0.124 | 0.001 | 0.021 | 0.021 | 0.014 | 0.006 | 0.021 | 1.000 | 0.737 | 0.357 | 0.974 | 0.898 | 0.312 | 0.269 | 0.826 | 0.774 |
|  |  | -0.3 | 0.5 | -0.8 | -0.5 | -0.3 | 0.000 | -0.081 | 0.081 | 0.053 | 0.028 | 0.081 | 0.022 | 0.121 | 0.123 | 0.110 | 0.069 | 0.123 | 0.001 | 0.021 | 0.022 | 0.015 | 0.006 | 0.022 | 1.000 | 0.901 | 1.000 | 0.976 | 0.977 | 1.000 | 0.526 | 0.820 | 0.801 |
|  |  | 0 | -0.8 | 0.8 | 0.5 | 0.3 | 0.001 | -0.075 | 0.076 | 0.046 | 0.030 | 0.076 | 0.027 | 0.124 | 0.126 | 0.110 | 0.070 | 0.126 | 0.001 | 0.021 | 0.022 | 0.014 | 0.006 | 0.022 | 0.053 | 0.999 | 0.999 | 0.992 | 0.989 | 0.999 | 0.397 | 0.836 | 0.811 |
|  |  | -1.6 | -0.8 | -0.8 | -0.5 | -0.3 | 0.000 | -0.078 | 0.078 | 0.050 | 0.028 | 0.078 | 0.023 | 0.117 | 0.117 | 0.105 | 0.067 | 0.118 | 0.001 | 0.020 | 0.020 | 0.013 | 0.005 | 0.020 | 1.000 | 0.957 | 0.879 | 0.355 | 0.374 | 0.849 | 0.004 | 0.839 | 0.818 |
|  |  | 0.8 | 0 | 0.8 | 0.5 | 0.3 | 0.000 | -0.081 | 0.081 | 0.054 | 0.028 | 0.081 | 0.025 | 0.121 | 0.121 | 0.107 | 0.066 | 0.121 | 0.001 | 0.021 | 0.021 | 0.014 | 0.005 | 0.021 | 1.000 | 0.000 | 0.997 | 0.916 | 0.913 | 1.000 | 0.057 | 0.808 | 0.784 |
|  |  | 1.3 | 0.8 | 0.5 | 0.5 | 0 | 0.002 | -0.079 | 0.080 | 0.057 | 0.023 | 0.080 | 0.020 | 0.121 | 0.120 | 0.109 | 0.062 | 0.120 | 0.000 | 0.021 | 0.021 | 0.015 | 0.004 | 0.021 | 1.000 | 0.865 | 0.661 | 0.759 | 0.000 | 0.610 | 0.006 | 0.810 | 0.812 |
|  |  | 1.3 | 0.8 | 0.5 | 0.5 | 0 | 0.001 | -0.042 | 0.043 | 0.043 | 0.000 | 0.043 | 0.025 | 0.106 | 0.108 | 0.107 | 0.015 | 0.108 | 0.001 | 0.013 | 0.014 | 0.013 | 0.000 | 0.014 | 1.000 | 0.946 | 0.601 | 0.623 | 0.016 | 0.624 | 0.004 | 0.817 | 0.824 |
|  |  | 0.8 | 0.8 | 0 | 0 | 0 | 0.001 | -0.079 | 0.079 | 0.048 | 0.031 | 0.079 | 0.013 | 0.125 | 0.124 | 0.111 | 0.063 | 0.124 | 0.000 | 0.022 | 0.022 | 0.015 | 0.005 | 0.022 | 1.000 | 0.995 | 0.028 | 0.017 | 0.019 | 0.019 | 0.306 | 0.826 | 0.798 |

TE: total effect; DE: direct effect; IE: indirect effect; IE_d: indirect effect calculated by difference method; IE_p: indirect effect calculated by product method; IE1: *X*→*M_1_*→*Y*; IE2: *X*→*M_2_*→*Y*.

**eTable 12.** Simulation of PSE-Egger in two non-ordered mediators setting with different effects (case(b),sc38-39,case(c)).

|  |  | TRUE | | | | | BIAS | | | | | | SE | | | | | | MSE | | | | | | Power/Type I error | | | | | | | | |
| --- | --- | --- | --- | --- | --- | --- | --- | --- | --- | --- | --- | --- | --- | --- | --- | --- | --- | --- | --- | --- | --- | --- | --- | --- | --- | --- | --- | --- | --- | --- | --- | --- | --- |
|  |  | TE | DE | IE | IE2 | IE1 | TE | DE | IE_d | IE1 | IE2 | IE_p | TE | DE | IE_d | IE1 | IE2 | IE_p | TE | DE | IE_d | IE1 | IE2 | IE_p | TE | DE | IE_d | IE1 | IE2 | IE_p | GY | GM1 | GM2 |
| Sc38 | IVW | 1.6 | 0.8 | 0.8 | 0.5 | 0.3 | 0.038 | 0.090 | -0.052 | -0.039 | -0.013 | -0.052 | 0.011 | 0.115 | 0.116 | 0.122 | 0.077 | 0.116 | 0.002 | 0.021 | 0.016 | 0.016 | 0.006 | 0.016 | 1.000 | 0.964 | 0.909 | 0.251 | 0.258 | 0.766 |  |  |  |
|  |  | 0.6 | 0.8 | -0.2 | -0.5 | 0.3 | -0.037 | 0.095 | -0.132 | -0.119 | -0.013 | -0.132 | 0.009 | 0.118 | 0.118 | 0.121 | 0.073 | 0.118 | 0.001 | 0.023 | 0.031 | 0.029 | 0.006 | 0.031 | 1.000 | 1.000 | 0.802 | 0.996 | 0.951 | 0.662 |  |  |  |
|  |  | 0.7 | 0.5 | 0.2 | 0.5 | -0.3 | -0.037 | 0.095 | -0.132 | -0.046 | -0.086 | -0.132 | 0.009 | 0.114 | 0.114 | 0.119 | 0.077 | 0.114 | 0.001 | 0.022 | 0.030 | 0.016 | 0.013 | 0.030 | 1.000 | 0.991 | 0.057 | 0.947 | 0.995 | 0.022 |  |  |  |
|  |  | -0.3 | 0.5 | -0.8 | -0.5 | -0.3 | -0.112 | 0.086 | -0.198 | -0.108 | -0.089 | -0.198 | 0.011 | 0.117 | 0.117 | 0.121 | 0.076 | 0.117 | 0.013 | 0.021 | 0.053 | 0.026 | 0.014 | 0.053 | 1.000 | 0.990 | 1.000 | 0.992 | 0.996 | 1.000 |  |  |  |
|  |  | 0 | -0.8 | 0.8 | 0.5 | 0.3 | 0.038 | 0.095 | -0.057 | -0.038 | -0.019 | -0.057 | 0.011 | 0.112 | 0.111 | 0.119 | 0.073 | 0.111 | 0.002 | 0.022 | 0.016 | 0.016 | 0.006 | 0.016 | 0.928 | 0.999 | 0.999 | 0.946 | 0.947 | 0.999 |  |  |  |
|  |  | -1.6 | -0.8 | -0.8 | -0.5 | -0.3 | -0.111 | 0.094 | -0.206 | -0.121 | -0.085 | -0.206 | 0.011 | 0.119 | 0.120 | 0.124 | 0.076 | 0.120 | 0.013 | 0.023 | 0.057 | 0.030 | 0.013 | 0.057 | 1.000 | 0.889 | 0.994 | 0.772 | 0.772 | 0.991 |  |  |  |
|  |  | 0.8 | 0 | 0.8 | 0.5 | 0.3 | 0.038 | 0.097 | -0.059 | -0.044 | -0.015 | -0.059 | 0.010 | 0.119 | 0.120 | 0.123 | 0.075 | 0.120 | 0.002 | 0.023 | 0.018 | 0.017 | 0.006 | 0.018 | 1.000 | 0.010 | 0.995 | 0.774 | 0.771 | 0.986 |  |  |  |
|  |  | 1.3 | 0.8 | 0.5 | 0.5 | 0 | 0.000 | 0.094 | -0.094 | -0.045 | -0.049 | -0.094 | 0.008 | 0.118 | 0.118 | 0.119 | 0.076 | 0.118 | 0.000 | 0.023 | 0.023 | 0.016 | 0.008 | 0.023 | 1.000 | 0.995 | 0.476 | 0.591 | 0.001 | 0.202 |  |  |  |
|  |  | 1.3 | 0.8 | 0.5 | 0.5 | 0 | 0.037 | 0.043 | -0.006 | -0.037 | 0.032 | -0.006 | 0.011 | 0.117 | 0.117 | 0.120 | 0.010 | 0.117 | 0.002 | 0.016 | 0.014 | 0.016 | 0.001 | 0.014 | 1.000 | 0.979 | 0.579 | 0.459 | 0.443 | 0.543 |  |  |  |
|  |  | 0.8 | 0.8 | 0 | 0 | 0 | -0.037 | 0.095 | -0.132 | -0.083 | -0.049 | -0.132 | 0.006 | 0.107 | 0.107 | 0.113 | 0.073 | 0.107 | 0.001 | 0.020 | 0.029 | 0.020 | 0.008 | 0.029 | 1.000 | 1.000 | 0.140 | 0.074 | 0.065 | 0.062 |  |  |  |
|  | Egger | 1.6 | 0.8 | 0.8 | 0.5 | 0.3 | 0.003 | -0.079 | 0.082 | 0.052 | 0.030 | 0.082 | 0.026 | 0.122 | 0.124 | 0.112 | 0.071 | 0.124 | 0.001 | 0.021 | 0.022 | 0.015 | 0.006 | 0.022 | 1.000 | 0.575 | 0.915 | 0.414 | 0.422 | 0.908 | 0.009 | 0.805 | 0.766 |
|  |  | 0.6 | 0.8 | -0.2 | -0.5 | 0.3 | 0.000 | -0.077 | 0.077 | 0.046 | 0.031 | 0.077 | 0.022 | 0.126 | 0.126 | 0.111 | 0.066 | 0.126 | 0.000 | 0.022 | 0.022 | 0.014 | 0.005 | 0.022 | 1.000 | 0.996 | 0.129 | 0.959 | 0.994 | 0.108 | 0.648 | 0.836 | 0.789 |
|  |  | 0.7 | 0.5 | 0.2 | 0.5 | -0.3 | 0.001 | -0.078 | 0.080 | 0.050 | 0.030 | 0.079 | 0.022 | 0.124 | 0.125 | 0.111 | 0.068 | 0.125 | 0.000 | 0.022 | 0.022 | 0.015 | 0.006 | 0.022 | 1.000 | 0.824 | 0.492 | 0.993 | 0.945 | 0.425 | 0.597 | 0.787 | 0.786 |
|  |  | -0.3 | 0.5 | -0.8 | -0.5 | -0.3 | -0.001 | -0.082 | 0.080 | 0.054 | 0.026 | 0.080 | 0.024 | 0.124 | 0.125 | 0.111 | 0.071 | 0.125 | 0.001 | 0.022 | 0.022 | 0.015 | 0.006 | 0.022 | 1.000 | 0.901 | 1.000 | 0.972 | 0.972 | 1.000 | 0.721 | 0.808 | 0.811 |
|  |  | 0 | -0.8 | 0.8 | 0.5 | 0.3 | 0.002 | -0.073 | 0.074 | 0.049 | 0.026 | 0.074 | 0.026 | 0.123 | 0.124 | 0.112 | 0.067 | 0.124 | 0.001 | 0.020 | 0.021 | 0.015 | 0.005 | 0.021 | 0.055 | 1.000 | 1.000 | 0.988 | 0.987 | 1.000 | 0.598 | 0.816 | 0.798 |
|  |  | -1.6 | -0.8 | -0.8 | -0.5 | -0.3 | -0.002 | -0.075 | 0.073 | 0.044 | 0.029 | 0.073 | 0.022 | 0.129 | 0.129 | 0.114 | 0.068 | 0.129 | 0.001 | 0.022 | 0.022 | 0.015 | 0.005 | 0.022 | 1.000 | 0.925 | 0.782 | 0.271 | 0.238 | 0.739 | 0.010 | 0.802 | 0.797 |
|  |  | 0.8 | 0 | 0.8 | 0.5 | 0.3 | 0.003 | -0.073 | 0.076 | 0.046 | 0.029 | 0.076 | 0.025 | 0.125 | 0.127 | 0.114 | 0.069 | 0.127 | 0.001 | 0.021 | 0.022 | 0.015 | 0.006 | 0.022 | 1.000 | 0.002 | 0.996 | 0.900 | 0.914 | 0.994 | 0.170 | 0.818 | 0.796 |
|  |  | 1.3 | 0.8 | 0.5 | 0.5 | 0 | 0.001 | -0.073 | 0.074 | 0.044 | 0.030 | 0.074 | 0.020 | 0.125 | 0.125 | 0.109 | 0.067 | 0.125 | 0.000 | 0.021 | 0.021 | 0.014 | 0.005 | 0.021 | 1.000 | 0.877 | 0.697 | 0.755 | 0.000 | 0.630 | 0.060 | 0.824 | 0.799 |
|  |  | 1.3 | 0.8 | 0.5 | 0.5 | 0 | 0.001 | -0.050 | 0.051 | 0.051 | 0.000 | 0.051 | 0.025 | 0.115 | 0.117 | 0.115 | 0.015 | 0.117 | 0.001 | 0.016 | 0.016 | 0.016 | 0.000 | 0.016 | 1.000 | 0.931 | 0.622 | 0.644 | 0.019 | 0.625 | 0.021 | 0.819 | 0.776 |
|  |  | 0.8 | 0.8 | 0 | 0 | 0 | 0.000 | -0.076 | 0.076 | 0.046 | 0.030 | 0.076 | 0.013 | 0.116 | 0.115 | 0.104 | 0.066 | 0.115 | 0.000 | 0.019 | 0.019 | 0.013 | 0.005 | 0.019 | 1.000 | 0.997 | 0.025 | 0.018 | 0.022 | 0.028 | 0.529 | 0.815 | 0.806 |
| Sc39 | IVW | 1.6 | 0.8 | 0.8 | 0.5 | 0.3 | 0.038 | -0.011 | 0.049 | -0.099 | 0.148 | 0.049 | 0.010 | 0.129 | 0.129 | 0.097 | 0.068 | 0.129 | 0.002 | 0.017 | 0.019 | 0.019 | 0.027 | 0.019 | 1.000 | 0.797 | 0.910 | 0.340 | 0.890 | 0.960 |  |  |  |
|  |  | 0.6 | 0.8 | -0.2 | -0.5 | 0.3 | 0.112 | -0.014 | 0.126 | -0.023 | 0.149 | 0.126 | 0.012 | 0.127 | 0.127 | 0.095 | 0.067 | 0.127 | 0.013 | 0.016 | 0.032 | 0.010 | 0.027 | 0.032 | 1.000 | 0.998 | 0.050 | 0.996 | 1.000 | 0.080 |  |  |  |
|  |  | 0.7 | 0.5 | 0.2 | 0.5 | -0.3 | -0.037 | -0.007 | -0.030 | -0.101 | 0.071 | -0.030 | 0.009 | 0.128 | 0.128 | 0.094 | 0.066 | 0.128 | 0.001 | 0.016 | 0.017 | 0.019 | 0.009 | 0.017 | 1.000 | 0.918 | 0.208 | 0.971 | 0.874 | 0.250 |  |  |  |
|  |  | -0.3 | 0.5 | -0.8 | -0.5 | -0.3 | 0.037 | -0.006 | 0.043 | -0.030 | 0.073 | 0.043 | 0.009 | 0.122 | 0.123 | 0.092 | 0.065 | 0.123 | 0.001 | 0.015 | 0.017 | 0.009 | 0.010 | 0.017 | 1.000 | 0.957 | 0.999 | 0.999 | 0.919 | 1.000 |  |  |  |
|  |  | 0 | -0.8 | 0.8 | 0.5 | 0.3 | 0.038 | -0.003 | 0.041 | -0.102 | 0.143 | 0.041 | 0.011 | 0.118 | 0.118 | 0.089 | 0.065 | 0.118 | 0.002 | 0.014 | 0.016 | 0.018 | 0.025 | 0.016 | 0.939 | 1.000 | 1.000 | 0.979 | 1.000 | 1.000 |  |  |  |
|  |  | -1.6 | -0.8 | -0.8 | -0.5 | -0.3 | 0.037 | -0.017 | 0.054 | -0.023 | 0.077 | 0.054 | 0.009 | 0.125 | 0.125 | 0.095 | 0.065 | 0.125 | 0.001 | 0.016 | 0.019 | 0.010 | 0.010 | 0.019 | 1.000 | 0.943 | 0.923 | 0.853 | 0.193 | 0.964 |  |  |  |
|  |  | 0.8 | 0 | 0.8 | 0.5 | 0.3 | 0.037 | -0.008 | 0.046 | -0.104 | 0.150 | 0.046 | 0.011 | 0.125 | 0.125 | 0.093 | 0.067 | 0.125 | 0.002 | 0.016 | 0.018 | 0.020 | 0.027 | 0.018 | 1.000 | 0.001 | 0.996 | 0.833 | 0.999 | 0.999 |  |  |  |
|  |  | 1.3 | 0.8 | 0.5 | 0.5 | 0 | 0.000 | -0.012 | 0.011 | -0.100 | 0.111 | 0.012 | 0.008 | 0.126 | 0.125 | 0.092 | 0.067 | 0.125 | 0.000 | 0.016 | 0.016 | 0.018 | 0.017 | 0.016 | 1.000 | 0.959 | 0.614 | 0.712 | 0.025 | 0.715 |  |  |  |
|  |  | 1.3 | 0.8 | 0.5 | 0.5 | 0 | 0.038 | 0.092 | -0.055 | -0.104 | 0.049 | -0.054 | 0.010 | 0.095 | 0.095 | 0.093 | 0.011 | 0.095 | 0.002 | 0.018 | 0.012 | 0.019 | 0.003 | 0.012 | 1.000 | 1.000 | 0.665 | 0.518 | 0.956 | 0.699 |  |  |  |
|  |  | 0.8 | 0.8 | 0 | 0 | 0 | 0.037 | -0.007 | 0.044 | -0.064 | 0.109 | 0.045 | 0.006 | 0.124 | 0.123 | 0.091 | 0.066 | 0.123 | 0.001 | 0.015 | 0.017 | 0.012 | 0.016 | 0.017 | 1.000 | 1.000 | 0.025 | 0.044 | 0.224 | 0.040 |  |  |  |
|  | Egger | 1.6 | 0.8 | 0.8 | 0.5 | 0.3 | 0.003 | -0.078 | 0.081 | 0.051 | 0.030 | 0.081 | 0.026 | 0.124 | 0.126 | 0.112 | 0.069 | 0.126 | 0.001 | 0.021 | 0.022 | 0.015 | 0.006 | 0.022 | 1.000 | 0.598 | 0.906 | 0.449 | 0.421 | 0.897 | 0.000 | 0.803 | 0.817 |
|  |  | 0.6 | 0.8 | -0.2 | -0.5 | 0.3 | 0.001 | -0.079 | 0.080 | 0.050 | 0.030 | 0.080 | 0.022 | 0.125 | 0.127 | 0.115 | 0.067 | 0.127 | 0.000 | 0.022 | 0.022 | 0.016 | 0.005 | 0.022 | 1.000 | 0.996 | 0.084 | 0.943 | 0.991 | 0.069 | 0.341 | 0.829 | 0.816 |
|  |  | 0.7 | 0.5 | 0.2 | 0.5 | -0.3 | 0.000 | -0.074 | 0.074 | 0.046 | 0.028 | 0.074 | 0.023 | 0.126 | 0.127 | 0.113 | 0.066 | 0.127 | 0.001 | 0.021 | 0.021 | 0.015 | 0.005 | 0.021 | 1.000 | 0.788 | 0.452 | 0.989 | 0.956 | 0.414 | 0.360 | 0.802 | 0.809 |
|  |  | -0.3 | 0.5 | -0.8 | -0.5 | -0.3 | -0.002 | -0.076 | 0.074 | 0.048 | 0.026 | 0.074 | 0.022 | 0.121 | 0.121 | 0.110 | 0.065 | 0.121 | 0.001 | 0.020 | 0.020 | 0.014 | 0.005 | 0.020 | 1.000 | 0.908 | 0.999 | 0.983 | 0.983 | 0.999 | 0.554 | 0.796 | 0.840 |
|  |  | 0 | -0.8 | 0.8 | 0.5 | 0.3 | 0.003 | -0.074 | 0.076 | 0.049 | 0.027 | 0.076 | 0.026 | 0.116 | 0.119 | 0.106 | 0.067 | 0.119 | 0.001 | 0.019 | 0.020 | 0.014 | 0.005 | 0.020 | 0.057 | 1.000 | 1.000 | 0.992 | 0.994 | 1.000 | 0.399 | 0.807 | 0.834 |
|  |  | -1.6 | -0.8 | -0.8 | -0.5 | -0.3 | 0.001 | -0.078 | 0.080 | 0.046 | 0.034 | 0.080 | 0.022 | 0.121 | 0.123 | 0.113 | 0.066 | 0.123 | 0.001 | 0.021 | 0.022 | 0.015 | 0.005 | 0.022 | 1.000 | 0.958 | 0.869 | 0.368 | 0.335 | 0.844 | 0.007 | 0.812 | 0.804 |
|  |  | 0.8 | 0 | 0.8 | 0.5 | 0.3 | 0.000 | -0.076 | 0.076 | 0.046 | 0.030 | 0.076 | 0.026 | 0.119 | 0.122 | 0.109 | 0.065 | 0.122 | 0.001 | 0.020 | 0.021 | 0.014 | 0.005 | 0.021 | 1.000 | 0.000 | 0.996 | 0.901 | 0.921 | 0.996 | 0.065 | 0.819 | 0.825 |
|  |  | 1.3 | 0.8 | 0.5 | 0.5 | 0 | -0.001 | -0.079 | 0.079 | 0.051 | 0.028 | 0.079 | 0.020 | 0.117 | 0.116 | 0.103 | 0.065 | 0.116 | 0.000 | 0.020 | 0.020 | 0.013 | 0.005 | 0.020 | 1.000 | 0.900 | 0.709 | 0.808 | 0.000 | 0.667 | 0.009 | 0.809 | 0.806 |
|  |  | 1.3 | 0.8 | 0.5 | 0.5 | 0 | 0.002 | -0.044 | 0.046 | 0.046 | 0.001 | 0.046 | 0.026 | 0.113 | 0.114 | 0.112 | 0.015 | 0.114 | 0.001 | 0.015 | 0.015 | 0.015 | 0.000 | 0.015 | 1.000 | 0.923 | 0.600 | 0.626 | 0.017 | 0.634 | 0.001 | 0.812 | 0.820 |
|  |  | 0.8 | 0.8 | 0 | 0 | 0 | 0.001 | -0.075 | 0.076 | 0.048 | 0.028 | 0.076 | 0.013 | 0.123 | 0.122 | 0.111 | 0.068 | 0.122 | 0.000 | 0.021 | 0.021 | 0.015 | 0.005 | 0.021 | 1.000 | 0.998 | 0.023 | 0.016 | 0.014 | 0.025 | 0.292 | 0.837 | 0.826 |
| Sc4 | IVW | 1.6 | 0.8 | 0.8 | 0.5 | 0.3 | 0.369 | -0.374 | 0.744 | 0.423 | 0.321 | 0.744 | 0.040 | 0.057 | 0.069 | 0.115 | 0.083 | 0.069 | 0.138 | 0.143 | 0.558 | 0.192 | 0.110 | 0.559 | 1.000 | 0.862 | 1.000 | 0.952 | 0.944 | 1.000 |  |  |  |
|  |  | 0.6 | 0.8 | -0.2 | -0.5 | 0.3 | 0.123 | -0.374 | 0.497 | 0.175 | 0.323 | 0.498 | 0.015 | 0.059 | 0.061 | 0.117 | 0.082 | 0.061 | 0.015 | 0.144 | 0.251 | 0.044 | 0.111 | 0.251 | 1.000 | 1.000 | 0.989 | 0.718 | 1.000 | 0.391 |  |  |  |
|  |  | 0.7 | 0.5 | 0.2 | 0.5 | -0.3 | 0.124 | -0.372 | 0.496 | 0.419 | 0.076 | 0.496 | 0.015 | 0.056 | 0.058 | 0.113 | 0.076 | 0.058 | 0.016 | 0.142 | 0.249 | 0.189 | 0.012 | 0.249 | 1.000 | 0.351 | 1.000 | 0.999 | 0.657 | 0.999 |  |  |  |
|  |  | -0.3 | 0.5 | -0.8 | -0.5 | -0.3 | -0.124 | -0.369 | 0.246 | 0.163 | 0.083 | 0.246 | 0.015 | 0.057 | 0.058 | 0.112 | 0.076 | 0.058 | 0.016 | 0.140 | 0.064 | 0.039 | 0.013 | 0.064 | 1.000 | 0.581 | 1.000 | 0.833 | 0.794 | 1.000 |  |  |  |
|  |  | 0 | -0.8 | 0.8 | 0.5 | 0.3 | 0.373 | -0.371 | 0.744 | 0.415 | 0.328 | 0.743 | 0.040 | 0.056 | 0.070 | 0.113 | 0.078 | 0.070 | 0.141 | 0.141 | 0.559 | 0.185 | 0.113 | 0.557 | 1.000 | 1.000 | 1.000 | 1.000 | 1.000 | 1.000 |  |  |  |
|  |  | -1.6 | -0.8 | -0.8 | -0.5 | -0.3 | -0.123 | -0.372 | 0.249 | 0.168 | 0.081 | 0.249 | 0.015 | 0.058 | 0.060 | 0.114 | 0.077 | 0.060 | 0.015 | 0.142 | 0.066 | 0.041 | 0.012 | 0.066 | 1.000 | 1.000 | 0.998 | 0.044 | 0.041 | 0.152 |  |  |  |
|  |  | 0.8 | 0 | 0.8 | 0.5 | 0.3 | 0.370 | -0.368 | 0.738 | 0.411 | 0.326 | 0.738 | 0.039 | 0.058 | 0.069 | 0.112 | 0.077 | 0.069 | 0.139 | 0.139 | 0.549 | 0.182 | 0.113 | 0.549 | 1.000 | 0.964 | 1.000 | 0.997 | 0.998 | 1.000 |  |  |  |
|  |  | 1.3 | 0.8 | 0.5 | 0.5 | 0 | 0.246 | -0.373 | 0.620 | 0.419 | 0.201 | 0.620 | 0.026 | 0.055 | 0.061 | 0.112 | 0.077 | 0.061 | 0.061 | 0.142 | 0.388 | 0.188 | 0.046 | 0.388 | 1.000 | 0.973 | 1.000 | 0.993 | 0.039 | 1.000 |  |  |  |
|  |  | 1.3 | 0.8 | 0.5 | 0.5 | 0 | 0.370 | -0.233 | 0.603 | 0.421 | 0.182 | 0.603 | 0.037 | 0.093 | 0.099 | 0.112 | 0.030 | 0.099 | 0.138 | 0.063 | 0.373 | 0.190 | 0.034 | 0.374 | 1.000 | 0.729 | 1.000 | 0.985 | 0.982 | 0.997 |  |  |  |
|  |  | 0.8 | 0.8 | 0 | 0 | 0 | 0.123 | -0.372 | 0.495 | 0.298 | 0.198 | 0.496 | 0.013 | 0.056 | 0.057 | 0.111 | 0.077 | 0.057 | 0.015 | 0.142 | 0.249 | 0.101 | 0.045 | 0.249 | 1.000 | 0.999 | 1.000 | 0.508 | 0.487 | 0.923 |  |  |  |
|  | Egger | 1.6 | 0.8 | 0.8 | 0.5 | 0.3 | 0.066 | -0.368 | 0.434 | 0.266 | 0.168 | 0.434 | 0.086 | 0.059 | 0.100 | 0.106 | 0.079 | 0.100 | 0.012 | 0.139 | 0.198 | 0.082 | 0.035 | 0.198 | 1.000 | 0.818 | 1.000 | 0.921 | 0.910 | 1.000 | 0.000 | 0.929 | 0.916 |
|  |  | 0.6 | 0.8 | -0.2 | -0.5 | 0.3 | 0.021 | -0.369 | 0.391 | 0.223 | 0.167 | 0.391 | 0.037 | 0.061 | 0.068 | 0.102 | 0.077 | 0.068 | 0.002 | 0.140 | 0.157 | 0.060 | 0.034 | 0.157 | 1.000 | 1.000 | 0.704 | 0.693 | 1.000 | 0.086 | 0.023 | 0.910 | 0.901 |
|  |  | 0.7 | 0.5 | 0.2 | 0.5 | -0.3 | 0.021 | -0.366 | 0.387 | 0.261 | 0.126 | 0.387 | 0.035 | 0.059 | 0.066 | 0.108 | 0.063 | 0.066 | 0.002 | 0.137 | 0.154 | 0.080 | 0.020 | 0.154 | 1.000 | 0.352 | 1.000 | 1.000 | 0.646 | 0.999 | 0.020 | 0.926 | 0.922 |
|  |  | -0.3 | 0.5 | -0.8 | -0.5 | -0.3 | -0.021 | -0.363 | 0.341 | 0.211 | 0.130 | 0.341 | 0.037 | 0.060 | 0.068 | 0.099 | 0.062 | 0.068 | 0.002 | 0.135 | 0.121 | 0.054 | 0.021 | 0.121 | 1.000 | 0.577 | 1.000 | 0.816 | 0.788 | 0.996 | 0.069 | 0.907 | 0.921 |
|  |  | 0 | -0.8 | 0.8 | 0.5 | 0.3 | 0.064 | -0.365 | 0.429 | 0.256 | 0.173 | 0.429 | 0.092 | 0.059 | 0.107 | 0.107 | 0.077 | 0.107 | 0.012 | 0.136 | 0.195 | 0.077 | 0.036 | 0.196 | 0.122 | 1.000 | 1.000 | 1.000 | 1.000 | 1.000 | 0.024 | 0.912 | 0.922 |
|  |  | -1.6 | -0.8 | -0.8 | -0.5 | -0.3 | -0.021 | -0.366 | 0.345 | 0.216 | 0.129 | 0.345 | 0.036 | 0.060 | 0.069 | 0.098 | 0.063 | 0.069 | 0.002 | 0.137 | 0.124 | 0.056 | 0.021 | 0.124 | 1.000 | 1.000 | 0.977 | 0.032 | 0.043 | 0.146 | 0.000 | 0.923 | 0.931 |
|  |  | 0.8 | 0 | 0.8 | 0.5 | 0.3 | 0.064 | -0.361 | 0.425 | 0.255 | 0.171 | 0.425 | 0.088 | 0.061 | 0.105 | 0.107 | 0.075 | 0.105 | 0.012 | 0.134 | 0.192 | 0.076 | 0.035 | 0.192 | 1.000 | 0.940 | 1.000 | 0.996 | 0.996 | 1.000 | 0.000 | 0.921 | 0.911 |
|  |  | 1.3 | 0.8 | 0.5 | 0.5 | 0 | 0.044 | -0.366 | 0.410 | 0.261 | 0.149 | 0.410 | 0.062 | 0.057 | 0.081 | 0.107 | 0.062 | 0.105 | 0.006 | 0.137 | 0.175 | 0.079 | 0.026 | 0.175 | 1.000 | 0.964 | 1.000 | 0.986 | 0.026 | 0.999 | 0.000 | 0.919 | 0.926 |
|  |  | 1.3 | 0.8 | 0.5 | 0.5 | 0 | 0.060 | -0.230 | 0.290 | 0.262 | 0.028 | 0.290 | 0.089 | 0.094 | 0.124 | 0.104 | 0.047 | 0.124 | 0.011 | 0.062 | 0.100 | 0.079 | 0.003 | 0.099 | 1.000 | 0.690 | 0.964 | 0.971 | 0.090 | 0.979 | 0.000 | 0.931 | 0.924 |
|  |  | 0.8 | 0.8 | 0 | 0 | 0 | 0.020 | -0.365 | 0.385 | 0.239 | 0.146 | 0.385 | 0.032 | 0.058 | 0.062 | 0.098 | 0.063 | 0.062 | 0.001 | 0.137 | 0.152 | 0.067 | 0.025 | 0.152 | 1.000 | 0.999 | 0.999 | 0.450 | 0.432 | 0.825 | 0.015 | 0.906 | 0.921 |

TE: total effect; DE: direct effect; IE: indirect effect; IE_d: indirect effect calculated by difference method; IE_p: indirect effect calculated by product method; IE1: *X*→*M_1_*→*Y*; IE2: *X*→*M_2_*→*Y*.

#### 5 Simulation E: Causally ordered mediators PSE-IVW in two mediators settings

The data-generating models were

Error variables were independently generated from a distribution; and instrument strength parameters were generated from a distribution. The parameters were generated from a normal distribution , independently. These effects were included in the simulation model to ensure that the direct and indirect effect was identified.

We also varied the sample size, the number of instrumental variables and simulated twenty different sets of values of the parameters. We obtained similar performance with Single-mediator IVW. Results were listed in **eTable 13-15**.

**eTable 13.** Simulation of PSE-IVW in two ordered mediators setting with different numbers of genetic variants.

|  | BIAS | | | | | | | SE | | | | | | | MSE | | | | | | | Power | | | | | | |
| --- | --- | --- | --- | --- | --- | --- | --- | --- | --- | --- | --- | --- | --- | --- | --- | --- | --- | --- | --- | --- | --- | --- | --- | --- | --- | --- | --- | --- |
|  | TE | DE | IE_d | IE1 | IE2 | IE3 | IE_p | TE | DE | IE_d | IE1 | IE2 | IE3 | IE_p | TE | DE | IE_d | IE1 | IE2 | IE3 | IE_p | TE | DE | IE_d | IE1 | IE2 | IE3 | IE_p |
| 5 | 0.017 | -0.038 | 0.055 | 0.011 | -0.008 | 0.052 | 0.055 | 0.180 | 0.200 | 0.265 | 0.209 | 0.423 | 0.414 | 0.265 | 0.033 | 0.041 | 0.073 | 0.044 | 0.179 | 0.174 | 0.073 | 1.000 | 0.000 | 0.336 | 0.032 | 0.001 | 0.000 | 0.041 |
| 10 | 0.003 | -0.052 | 0.055 | 0.020 | -0.014 | 0.050 | 0.055 | 0.126 | 0.092 | 0.153 | 0.108 | 0.221 | 0.204 | 0.153 | 0.016 | 0.011 | 0.026 | 0.012 | 0.049 | 0.044 | 0.027 | 1.000 | 0.445 | 0.931 | 0.317 | 0.078 | 0.057 | 0.691 |
| 15 | 0.008 | -0.043 | 0.051 | 0.020 | -0.014 | 0.046 | 0.052 | 0.105 | 0.062 | 0.118 | 0.082 | 0.169 | 0.161 | 0.118 | 0.011 | 0.006 | 0.016 | 0.007 | 0.029 | 0.028 | 0.017 | 1.000 | 0.893 | 0.995 | 0.684 | 0.306 | 0.286 | 0.962 |
| 20 | 0.010 | -0.049 | 0.059 | 0.023 | -0.008 | 0.044 | 0.059 | 0.092 | 0.051 | 0.102 | 0.070 | 0.144 | 0.132 | 0.102 | 0.009 | 0.005 | 0.014 | 0.005 | 0.021 | 0.019 | 0.014 | 1.000 | 0.987 | 1.000 | 0.901 | 0.539 | 0.484 | 0.998 |
| 30 | 0.011 | -0.045 | 0.056 | 0.020 | -0.008 | 0.044 | 0.057 | 0.076 | 0.041 | 0.083 | 0.053 | 0.111 | 0.104 | 0.083 | 0.006 | 0.004 | 0.010 | 0.003 | 0.012 | 0.013 | 0.010 | 1.000 | 1.000 | 1.000 | 0.996 | 0.778 | 0.680 | 1.000 |
| 50 | 0.008 | -0.046 | 0.054 | 0.019 | -0.015 | 0.050 | 0.054 | 0.057 | 0.032 | 0.063 | 0.043 | 0.090 | 0.084 | 0.063 | 0.003 | 0.003 | 0.007 | 0.002 | 0.008 | 0.010 | 0.007 | 1.000 | 1.000 | 1.000 | 1.000 | 0.909 | 0.889 | 1.000 |
| 100 | 0.008 | -0.046 | 0.054 | 0.021 | -0.014 | 0.048 | 0.054 | 0.040 | 0.022 | 0.045 | 0.029 | 0.060 | 0.057 | 0.045 | 0.002 | 0.003 | 0.005 | 0.001 | 0.004 | 0.006 | 0.005 | 1.000 | 1.000 | 1.000 | 1.000 | 0.998 | 0.992 | 1.000 |
| 150 | 0.011 | -0.046 | 0.058 | 0.024 | -0.013 | 0.048 | 0.058 | 0.033 | 0.018 | 0.037 | 0.024 | 0.049 | 0.046 | 0.037 | 0.001 | 0.002 | 0.005 | 0.001 | 0.003 | 0.004 | 0.005 | 1.000 | 1.000 | 1.000 | 1.000 | 1.000 | 1.000 | 1.000 |

TE: total effect; DE: direct effect; IE: indirect effect; IE_d: indirect effect calculated by difference method; IE_p: indirect effect calculated by product method; IE1: *X*→*M_1_*→*Y*; IE2: *X*→*M_2_*→*Y*; IE3: *X*→*M_1_*→*M_2_*→*Y*.

**eTable 14.** Simulation of PSE-IVW in two ordered mediators setting with different sample size.

|  | BIAS | | | | | | | SE | | | | | | | MSE | | | | | | | POWER | | | | | | |
| --- | --- | --- | --- | --- | --- | --- | --- | --- | --- | --- | --- | --- | --- | --- | --- | --- | --- | --- | --- | --- | --- | --- | --- | --- | --- | --- | --- | --- |
|  | TE | DE | IE_d | IE1 | IE2 | IE3 | IE_p | TE | DE | IE_d | IE1 | IE2 | IE3 | IE_p | TE | DE | IE_d | IE1 | IE2 | IE3 | IE_p | TE | DE | IE_d | IE1 | IE2 | IE3 | IE_p |
| 2000 | 0.087 | -0.166 | 0.253 | 0.091 | -0.079 | 0.241 | 0.253 | 0.145 | 0.095 | 0.161 | 0.117 | 0.156 | 0.159 | 0.161 | 0.029 | 0.037 | 0.090 | 0.022 | 0.030 | 0.083 | 0.090 | 1.000 | 0.019 | 0.977 | 0.020 | 0.012 | 0.136 | 0.658 |
| 4000 | 0.044 | -0.131 | 0.175 | 0.065 | -0.060 | 0.171 | 0.175 | 0.113 | 0.080 | 0.133 | 0.096 | 0.146 | 0.139 | 0.133 | 0.015 | 0.024 | 0.048 | 0.013 | 0.025 | 0.048 | 0.048 | 1.000 | 0.325 | 0.997 | 0.163 | 0.122 | 0.364 | 0.930 |
| 6000 | 0.031 | -0.108 | 0.139 | 0.050 | -0.035 | 0.125 | 0.139 | 0.105 | 0.071 | 0.123 | 0.086 | 0.138 | 0.130 | 0.123 | 0.012 | 0.017 | 0.034 | 0.010 | 0.020 | 0.032 | 0.034 | 1.000 | 0.635 | 1.000 | 0.353 | 0.285 | 0.475 | 0.987 |
| 8000 | 0.024 | -0.085 | 0.109 | 0.041 | -0.032 | 0.101 | 0.110 | 0.095 | 0.063 | 0.110 | 0.083 | 0.136 | 0.126 | 0.110 | 0.010 | 0.011 | 0.024 | 0.009 | 0.020 | 0.026 | 0.024 | 1.000 | 0.874 | 1.000 | 0.576 | 0.410 | 0.531 | 0.992 |
| 10000 | 0.018 | -0.076 | 0.094 | 0.037 | -0.028 | 0.085 | 0.094 | 0.096 | 0.060 | 0.111 | 0.077 | 0.129 | 0.122 | 0.111 | 0.009 | 0.009 | 0.021 | 0.007 | 0.017 | 0.022 | 0.021 | 1.000 | 0.946 | 1.000 | 0.733 | 0.516 | 0.600 | 0.999 |
| 12000 | 0.016 | -0.072 | 0.089 | 0.033 | -0.013 | 0.069 | 0.089 | 0.089 | 0.057 | 0.101 | 0.071 | 0.129 | 0.125 | 0.101 | 0.008 | 0.008 | 0.018 | 0.006 | 0.017 | 0.020 | 0.018 | 1.000 | 0.973 | 1.000 | 0.851 | 0.591 | 0.584 | 0.999 |
| 14000 | 0.008 | -0.061 | 0.069 | 0.025 | -0.018 | 0.062 | 0.069 | 0.085 | 0.053 | 0.098 | 0.070 | 0.123 | 0.116 | 0.098 | 0.007 | 0.006 | 0.014 | 0.006 | 0.015 | 0.017 | 0.014 | 1.000 | 0.991 | 1.000 | 0.896 | 0.608 | 0.614 | 1.000 |
| 16000 | 0.013 | -0.053 | 0.066 | 0.024 | -0.018 | 0.060 | 0.066 | 0.083 | 0.050 | 0.092 | 0.064 | 0.129 | 0.121 | 0.092 | 0.007 | 0.005 | 0.013 | 0.005 | 0.017 | 0.018 | 0.013 | 1.000 | 0.992 | 1.000 | 0.944 | 0.640 | 0.628 | 1.000 |
| 18000 | 0.012 | -0.051 | 0.062 | 0.027 | -0.018 | 0.054 | 0.062 | 0.082 | 0.047 | 0.092 | 0.063 | 0.126 | 0.119 | 0.092 | 0.007 | 0.005 | 0.012 | 0.005 | 0.016 | 0.017 | 0.012 | 1.000 | 0.999 | 1.000 | 0.963 | 0.661 | 0.623 | 1.000 |
| 20000 | 0.004 | -0.046 | 0.050 | 0.019 | -0.012 | 0.043 | 0.050 | 0.083 | 0.047 | 0.092 | 0.060 | 0.127 | 0.117 | 0.092 | 0.007 | 0.004 | 0.011 | 0.004 | 0.016 | 0.016 | 0.011 | 1.000 | 1.000 | 1.000 | 0.981 | 0.675 | 0.571 | 1.000 |

TE: total effect; DE: direct effect; IE: indirect effect; IE_d: indirect effect calculated by difference method; IE_p: indirect effect calculated by product method; IE1: *X*→*M_1_*→*Y*; IE2: *X*→*M_2_*→*Y*; IE3: *X*→*M_1_*→*M_2_*→*Y*.

**eTable 15.** Simulation of PSE-IVW in two ordered mediators setting with different *r*_12_.

|  | BIAS | | | | | | | SE | | | | | | | MSE | | | | | | | POWER | | | | | | |
| --- | --- | --- | --- | --- | --- | --- | --- | --- | --- | --- | --- | --- | --- | --- | --- | --- | --- | --- | --- | --- | --- | --- | --- | --- | --- | --- | --- | --- |
| *r*_12_ | TE | DE | IE_d | IE1 | IE2 | IE3 | IE_p | TE | DE | TE_d | IE1 | IE2 | IE3 | IE_p | TE | DE | TE_d | IE1 | IE2 | IE3 | IE_p | TE | DE | TE_d | IE1 | IE2 | IE3 | IE_p |
| -1 | 0.005 | -0.045 | 0.049 | 0.062 | -0.015 | 0.002 | 0.049 | 0.049 | 0.045 | 0.062 | 0.071 | 0.132 | 0.132 | 0.062 | 0.002 | 0.004 | 0.006 | 0.009 | 0.018 | 0.017 | 0.006 | 1.000 | 1.000 | 0.920 | 0.997 | 0.664 | 0.981 | 0.035 |
| -0.8 | 0.010 | -0.045 | 0.054 | 0.058 | -0.014 | 0.010 | 0.054 | 0.050 | 0.045 | 0.065 | 0.066 | 0.123 | 0.120 | 0.065 | 0.003 | 0.004 | 0.007 | 0.008 | 0.015 | 0.014 | 0.007 | 1.000 | 1.000 | 0.991 | 1.000 | 0.647 | 0.926 | 0.374 |
| -0.6 | 0.004 | -0.047 | 0.051 | 0.053 | -0.015 | 0.013 | 0.051 | 0.053 | 0.048 | 0.068 | 0.066 | 0.124 | 0.119 | 0.068 | 0.003 | 0.004 | 0.007 | 0.007 | 0.015 | 0.014 | 0.007 | 1.000 | 1.000 | 0.998 | 1.000 | 0.659 | 0.740 | 0.839 |
| -0.4 | 0.008 | -0.046 | 0.054 | 0.047 | -0.014 | 0.022 | 0.054 | 0.057 | 0.046 | 0.070 | 0.062 | 0.128 | 0.118 | 0.070 | 0.003 | 0.004 | 0.008 | 0.006 | 0.017 | 0.014 | 0.008 | 1.000 | 1.000 | 1.000 | 1.000 | 0.650 | 0.376 | 0.967 |
| -0.2 | 0.007 | -0.046 | 0.054 | 0.039 | -0.011 | 0.026 | 0.054 | 0.063 | 0.047 | 0.076 | 0.061 | 0.121 | 0.113 | 0.076 | 0.004 | 0.004 | 0.009 | 0.005 | 0.015 | 0.013 | 0.009 | 1.000 | 0.999 | 1.000 | 1.000 | 0.665 | 0.134 | 0.997 |
| 0 | 0.010 | -0.050 | 0.060 | 0.037 | -0.009 | 0.031 | 0.060 | 0.069 | 0.046 | 0.080 | 0.061 | 0.123 | 0.110 | 0.080 | 0.005 | 0.005 | 0.010 | 0.005 | 0.015 | 0.013 | 0.010 | 1.000 | 1.000 | 1.000 | 1.000 | 0.674 | 0.062 | 0.998 |
| 0.2 | 0.006 | -0.047 | 0.053 | 0.027 | -0.018 | 0.044 | 0.053 | 0.073 | 0.048 | 0.084 | 0.061 | 0.128 | 0.118 | 0.084 | 0.005 | 0.004 | 0.010 | 0.004 | 0.017 | 0.016 | 0.010 | 1.000 | 0.999 | 1.000 | 0.997 | 0.655 | 0.292 | 1.000 |
| 0.4 | 0.008 | -0.046 | 0.054 | 0.024 | -0.021 | 0.052 | 0.055 | 0.082 | 0.048 | 0.092 | 0.062 | 0.123 | 0.114 | 0.092 | 0.007 | 0.004 | 0.011 | 0.004 | 0.015 | 0.016 | 0.011 | 1.000 | 0.999 | 1.000 | 0.978 | 0.657 | 0.623 | 1.000 |
| 0.6 | 0.008 | -0.047 | 0.055 | 0.017 | -0.017 | 0.055 | 0.055 | 0.087 | 0.046 | 0.095 | 0.064 | 0.124 | 0.120 | 0.095 | 0.008 | 0.004 | 0.012 | 0.004 | 0.016 | 0.017 | 0.012 | 1.000 | 1.000 | 1.000 | 0.907 | 0.632 | 0.855 | 1.000 |
| 0.8 | 0.006 | -0.046 | 0.052 | 0.004 | -0.015 | 0.063 | 0.052 | 0.092 | 0.047 | 0.102 | 0.066 | 0.124 | 0.124 | 0.102 | 0.008 | 0.004 | 0.013 | 0.004 | 0.016 | 0.019 | 0.013 | 1.000 | 0.995 | 1.000 | 0.662 | 0.638 | 0.954 | 1.000 |
| 1 | 0.015 | -0.046 | 0.061 | 0.003 | -0.001 | 0.059 | 0.061 | 0.108 | 0.046 | 0.115 | 0.073 | 0.125 | 0.128 | 0.115 | 0.012 | 0.004 | 0.017 | 0.005 | 0.016 | 0.020 | 0.017 | 1.000 | 0.991 | 1.000 | 0.344 | 0.658 | 0.970 | 1.000 |

TE: total effect; DE: direct effect; IE: indirect effect; IE_d: indirect effect calculated by difference method; IE_p: indirect effect calculated by product method; IE1: *X*→*M_1_*→*Y*; IE2: *X*→*M_2_*→*Y*; IE3: *X*→*M_1_*→*M_2_*→*Y*.

#### 6 Simulation F: Causally ordered mediators PSE-Egger in two mediators settings

The data-generating models were

Error variables were independently generated from a distribution; and instrument strength parameters were generated from a distribution. This model additionally allows genetic variant to be associated with confounders variable . This occurred when was non-zero (a violation of the **Assumption Ⅱ**). Simulation scenarios (a)–(c) were implemented by additionally specifying values for ( the direct effect of genetic variables on the mediator ), ( the direct effect of genetic variables on the mediator ) and ( the direct effect of genetic variables on outcome *Y*) as below:

**eTable 16.** Settings for simulation of PSE-Egger in two ordered mediators setting

|  | Pleiotropy | InSIDE |  |  |  |  |  |
| --- | --- | --- | --- | --- | --- | --- | --- |
| case(a) | Balanced | Satisfied |  |  |  | 0 | sc21 |
|  |  |  |  |  |  | 0 | sc22 |
|  |  |  |  |  |  | 0 | sc23 |
|  |  |  |  |  |  | 0 | sc2 |
| case(b) | Directional | Satisfied |  |  |  | 0 | sc31 |
|  |  |  |  |  |  | 0 | sc32 |
|  |  |  |  |  |  | 0 | sc33 |
|  |  |  |  |  |  | 0 | sc34 |
|  |  |  |  |  |  | 0 | sc35 |
|  |  |  |  |  |  | 0 | sc36 |
|  |  |  |  |  |  | 0 | sc37 |
|  |  |  |  |  |  | 0 | sc38 |
|  |  |  |  |  |  | 0 | sc39 |
|  |  |  |  |  |  | 0 | sc3 |
| case(c) | Directional | Not satisfied |  |  |  |  | sc4 |

The performance is similar with Single-mediator MR-Egger. Results are listed in **eTable 17-24**.

**eTable 17** Simulation of PSE-Egger in two ordered mediators setting with different effects (case(a),sc2,sc21).

|  |  | true | | | | | | BIAS | | | | | | | SE | | | | | | | MSE | | | | | | | Power/Type I error | | | | | | | | | |
| --- | --- | --- | --- | --- | --- | --- | --- | --- | --- | --- | --- | --- | --- | --- | --- | --- | --- | --- | --- | --- | --- | --- | --- | --- | --- | --- | --- | --- | --- | --- | --- | --- | --- | --- | --- | --- | --- | --- |
|  |  | TE | DE | IE | IE1 | IE2 | IE3 | TE | DE | IE_d | IE1 | IE2 | IE3 | IE_p | TE | DE | IE_d | IE1 | IE2 | IE3 | IE_p | TE | DE | IE_d | IE1 | IE2 | IE3 | IE_p | TE | DE | IE_d | IE1 | IE2 | IE3 | IE_p | GY | GM1 | GM2 |
| Sc2 | IVW | 1.8 | 0.8 | 1 | 0.5 | 0.3 | 0.2 | 0.000 | -0.027 | 0.028 | 0.013 | -0.008 | 0.023 | 0.028 | 0.020 | 0.122 | 0.121 | 0.108 | 0.127 | 0.114 | 0.121 | 0.000 | 0.016 | 0.015 | 0.012 | 0.016 | 0.014 | 0.015 | 1.000 | 0.991 | 1.000 | 0.866 | 0.692 | 0.490 | 0.999 |  |  |  |
|  |  | 0.8 | 0.8 | 0 | -0.5 | 0.3 | 0.2 | 0.000 | -0.024 | 0.024 | 0.014 | -0.007 | 0.017 | 0.024 | 0.015 | 0.122 | 0.123 | 0.113 | 0.125 | 0.115 | 0.123 | 0.000 | 0.016 | 0.016 | 0.013 | 0.016 | 0.013 | 0.016 | 1.000 | 0.999 | 0.068 | 0.988 | 0.784 | 0.545 | 0.000 |  |  |  |
|  |  | 1.2 | 0.8 | 0.4 | 0.5 | -0.3 | 0.2 | 0.002 | -0.023 | 0.024 | 0.020 | 0.028 | -0.023 | 0.024 | 0.019 | 0.121 | 0.122 | 0.114 | 0.122 | 0.112 | 0.122 | 0.000 | 0.015 | 0.016 | 0.013 | 0.016 | 0.013 | 0.016 | 1.000 | 1.000 | 0.888 | 0.986 | 0.756 | 0.415 | 0.551 |  |  |  |
|  |  | 0.2 | 0.8 | -0.6 | -0.5 | -0.3 | 0.2 | 0.001 | -0.020 | 0.021 | 0.019 | 0.020 | -0.019 | 0.021 | 0.015 | 0.123 | 0.124 | 0.115 | 0.119 | 0.110 | 0.124 | 0.000 | 0.016 | 0.016 | 0.014 | 0.015 | 0.012 | 0.016 | 1.000 | 1.000 | 0.989 | 0.981 | 0.772 | 0.442 | 0.934 |  |  |  |
|  |  | 0.2 | -0.8 | 1 | 0.5 | 0.3 | 0.2 | 0.001 | -0.026 | 0.027 | 0.009 | -0.008 | 0.026 | 0.027 | 0.021 | 0.130 | 0.131 | 0.116 | 0.133 | 0.124 | 0.131 | 0.000 | 0.018 | 0.018 | 0.013 | 0.018 | 0.016 | 0.018 | 1.000 | 1.000 | 1.000 | 0.990 | 0.771 | 0.562 | 1.000 |  |  |  |
|  |  | -1.8 | -0.8 | -1 | -0.5 | -0.3 | -0.2 | -0.001 | -0.021 | 0.020 | 0.005 | 0.028 | -0.013 | 0.020 | 0.019 | 0.121 | 0.121 | 0.113 | 0.120 | 0.119 | 0.121 | 0.000 | 0.015 | 0.015 | 0.013 | 0.015 | 0.014 | 0.015 | 1.000 | 0.999 | 1.000 | 0.905 | 0.689 | 0.497 | 1.000 |  |  |  |
|  |  | 1 | 0 | 1 | 0.5 | 0.3 | 0.2 | 0.001 | -0.024 | 0.025 | 0.010 | -0.008 | 0.023 | 0.025 | 0.020 | 0.127 | 0.128 | 0.115 | 0.126 | 0.121 | 0.128 | 0.000 | 0.017 | 0.017 | 0.013 | 0.016 | 0.015 | 0.017 | 1.000 | 0.022 | 1.000 | 0.971 | 0.759 | 0.551 | 1.000 |  |  |  |
|  |  | 1.3 | 0.8 | 0.5 | 0 | 0.3 | 0.2 | 0.000 | -0.027 | 0.027 | 0.014 | -0.009 | 0.021 | 0.027 | 0.015 | 0.120 | 0.120 | 0.114 | 0.127 | 0.119 | 0.120 | 0.000 | 0.015 | 0.015 | 0.013 | 0.016 | 0.015 | 0.015 | 1.000 | 1.000 | 0.970 | 0.022 | 0.756 | 0.542 | 0.782 |  |  |  |
|  |  | 1.5 | 0.8 | 0.7 | 0.5 | 0 | 0.2 | 0.001 | -0.015 | 0.016 | 0.008 | -0.019 | 0.028 | 0.016 | 0.021 | 0.110 | 0.111 | 0.117 | 0.107 | 0.118 | 0.111 | 0.000 | 0.012 | 0.013 | 0.014 | 0.012 | 0.015 | 0.013 | 1.000 | 1.000 | 1.000 | 0.916 | 0.050 | 0.543 | 0.956 |  |  |  |
|  |  | 1 | 0.8 | 0.2 | 0 | 0 | 0.2 | 0.001 | -0.014 | 0.015 | 0.008 | -0.017 | 0.024 | 0.015 | 0.015 | 0.106 | 0.107 | 0.116 | 0.114 | 0.122 | 0.107 | 0.000 | 0.012 | 0.012 | 0.013 | 0.013 | 0.016 | 0.012 | 1.000 | 1.000 | 0.508 | 0.042 | 0.067 | 0.545 | 0.072 |  |  |  |
|  |  | 0 | -1 | 1 | 0.5 | 0.3 | 0.2 | 0.001 | -0.030 | 0.030 | 0.010 | -0.006 | 0.027 | 0.031 | 0.020 | 0.120 | 0.122 | 0.119 | 0.127 | 0.118 | 0.122 | 0.000 | 0.015 | 0.016 | 0.014 | 0.016 | 0.015 | 0.016 | 0.044 | 1.000 | 1.000 | 0.989 | 0.767 | 0.562 | 1.000 |  |  |  |
|  | Egger | 1.8 | 0.8 | 1 | 0.5 | 0.3 | 0.2 | 0.000 | -0.027 | 0.027 | 0.012 | -0.007 | 0.022 | 0.027 | 0.051 | 0.128 | 0.132 | 0.113 | 0.135 | 0.118 | 0.133 | 0.003 | 0.017 | 0.018 | 0.013 | 0.018 | 0.014 | 0.018 | 1.000 | 0.986 | 1.000 | 0.850 | 0.648 | 0.489 | 0.999 | 0.000 | 0.050 | 0.046 |
|  |  | 0.8 | 0.8 | 0 | -0.5 | 0.3 | 0.2 | 0.000 | -0.024 | 0.023 | 0.012 | -0.008 | 0.019 | 0.023 | 0.037 | 0.127 | 0.130 | 0.120 | 0.132 | 0.120 | 0.130 | 0.001 | 0.017 | 0.017 | 0.014 | 0.017 | 0.015 | 0.017 | 1.000 | 0.999 | 0.054 | 0.982 | 0.744 | 0.530 | 0.001 | 0.047 | 0.055 | 0.067 |
|  |  | 1.2 | 0.8 | 0.4 | 0.5 | -0.3 | 0.2 | 0.004 | -0.023 | 0.026 | 0.022 | 0.028 | -0.024 | 0.026 | 0.049 | 0.127 | 0.133 | 0.119 | 0.128 | 0.115 | 0.133 | 0.002 | 0.017 | 0.018 | 0.015 | 0.017 | 0.014 | 0.018 | 1.000 | 1.000 | 0.839 | 0.983 | 0.719 | 0.401 | 0.527 | 0.020 | 0.059 | 0.050 |
|  |  | 0.2 | 0.8 | -0.6 | -0.5 | -0.3 | 0.2 | 0.002 | -0.017 | 0.019 | 0.017 | 0.020 | -0.018 | 0.018 | 0.036 | 0.129 | 0.131 | 0.121 | 0.123 | 0.112 | 0.131 | 0.001 | 0.017 | 0.017 | 0.015 | 0.016 | 0.013 | 0.017 | 0.999 | 1.000 | 0.985 | 0.981 | 0.736 | 0.437 | 0.923 | 0.049 | 0.056 | 0.051 |
|  |  | 0.2 | -0.8 | 1 | 0.5 | 0.3 | 0.2 | 0.006 | -0.025 | 0.031 | 0.010 | -0.005 | 0.026 | 0.031 | 0.050 | 0.136 | 0.141 | 0.121 | 0.140 | 0.129 | 0.141 | 0.002 | 0.019 | 0.021 | 0.015 | 0.020 | 0.017 | 0.021 | 0.972 | 1.000 | 1.000 | 0.986 | 0.731 | 0.536 | 1.000 | 0.047 | 0.046 | 0.054 |
|  |  | -1.8 | -0.8 | -1 | -0.5 | -0.3 | -0.2 | -0.001 | -0.021 | 0.020 | 0.005 | 0.029 | -0.014 | 0.019 | 0.050 | 0.126 | 0.131 | 0.116 | 0.127 | 0.123 | 0.130 | 0.003 | 0.016 | 0.017 | 0.014 | 0.017 | 0.015 | 0.017 | 1.000 | 0.995 | 1.000 | 0.882 | 0.642 | 0.476 | 1.000 | 0.002 | 0.055 | 0.058 |
|  |  | 1 | 0 | 1 | 0.5 | 0.3 | 0.2 | 0.002 | -0.024 | 0.026 | 0.011 | -0.008 | 0.023 | 0.026 | 0.052 | 0.135 | 0.139 | 0.119 | 0.131 | 0.124 | 0.139 | 0.003 | 0.019 | 0.020 | 0.014 | 0.017 | 0.016 | 0.020 | 1.000 | 0.020 | 1.000 | 0.968 | 0.731 | 0.531 | 1.000 | 0.017 | 0.041 | 0.056 |
|  |  | 1.3 | 0.8 | 0.5 | 0 | 0.3 | 0.2 | 0.000 | -0.026 | 0.026 | 0.014 | -0.010 | 0.022 | 0.026 | 0.037 | 0.124 | 0.124 | 0.116 | 0.129 | 0.121 | 0.124 | 0.001 | 0.016 | 0.016 | 0.014 | 0.017 | 0.015 | 0.016 | 1.000 | 1.000 | 0.946 | 0.016 | 0.708 | 0.509 | 0.744 | 0.008 | 0.041 | 0.049 |
|  |  | 1.5 | 0.8 | 0.7 | 0.5 | 0 | 0.2 | 0.001 | -0.015 | 0.015 | 0.007 | -0.019 | 0.028 | 0.016 | 0.050 | 0.115 | 0.117 | 0.121 | 0.114 | 0.123 | 0.117 | 0.002 | 0.014 | 0.014 | 0.015 | 0.013 | 0.016 | 0.014 | 1.000 | 1.000 | 1.000 | 0.899 | 0.058 | 0.507 | 0.937 | 0.002 | 0.047 | 0.052 |
|  |  | 1 | 0.8 | 0.2 | 0 | 0 | 0.2 | 0.003 | -0.012 | 0.015 | 0.008 | -0.017 | 0.024 | 0.016 | 0.037 | 0.112 | 0.113 | 0.119 | 0.120 | 0.126 | 0.113 | 0.001 | 0.013 | 0.013 | 0.014 | 0.015 | 0.016 | 0.013 | 1.000 | 1.000 | 0.428 | 0.047 | 0.073 | 0.531 | 0.062 | 0.028 | 0.056 | 0.050 |
|  |  | 0 | -1 | 1 | 0.5 | 0.3 | 0.2 | 0.002 | -0.028 | 0.030 | 0.009 | -0.005 | 0.027 | 0.031 | 0.051 | 0.125 | 0.131 | 0.124 | 0.131 | 0.121 | 0.131 | 0.003 | 0.016 | 0.018 | 0.015 | 0.017 | 0.015 | 0.018 | 0.052 | 1.000 | 1.000 | 0.982 | 0.740 | 0.531 | 1.000 | 0.040 | 0.040 | 0.054 |
| Sc21 | IVW | 1.8 | 0.8 | 1 | 0.5 | 0.3 | 0.2 | -0.014 | 0.124 | -0.138 | -0.142 | -0.032 | 0.036 | -0.138 | 0.014 | 0.100 | 0.101 | 0.101 | 0.162 | 0.163 | 0.101 | 0.000 | 0.025 | 0.029 | 0.030 | 0.027 | 0.028 | 0.029 | 1.000 | 0.982 | 0.988 | 0.089 | 0.391 | 0.311 | 0.844 |  |  |  |
|  |  | 0.8 | 0.8 | 0 | -0.5 | 0.3 | 0.2 | 0.060 | 0.127 | -0.068 | -0.066 | -0.023 | 0.021 | -0.068 | 0.012 | 0.100 | 0.100 | 0.097 | 0.155 | 0.151 | 0.100 | 0.004 | 0.026 | 0.015 | 0.014 | 0.025 | 0.023 | 0.015 | 1.000 | 1.000 | 0.036 | 0.995 | 0.462 | 0.342 | 0.000 |  |  |  |
|  |  | 1.2 | 0.8 | 0.4 | 0.5 | -0.3 | 0.2 | -0.014 | 0.123 | -0.138 | -0.128 | 0.056 | -0.066 | -0.138 | 0.014 | 0.100 | 0.101 | 0.096 | 0.149 | 0.146 | 0.101 | 0.000 | 0.025 | 0.029 | 0.026 | 0.025 | 0.026 | 0.029 | 1.000 | 1.000 | 0.243 | 0.700 | 0.411 | 0.165 | 0.012 |  |  |  |
|  |  | 0.2 | 0.8 | -0.6 | -0.5 | -0.3 | 0.2 | 0.060 | 0.120 | -0.060 | -0.051 | 0.047 | -0.057 | -0.061 | 0.012 | 0.099 | 0.100 | 0.096 | 0.143 | 0.139 | 0.100 | 0.004 | 0.024 | 0.014 | 0.012 | 0.023 | 0.023 | 0.014 | 1.000 | 1.000 | 1.000 | 1.000 | 0.440 | 0.175 | 0.941 |  |  |  |
|  |  | 0.2 | -0.8 | 1 | 0.5 | 0.3 | 0.2 | -0.015 | 0.124 | -0.139 | -0.145 | -0.029 | 0.035 | -0.139 | 0.015 | 0.100 | 0.101 | 0.102 | 0.166 | 0.167 | 0.101 | 0.000 | 0.026 | 0.030 | 0.031 | 0.028 | 0.029 | 0.029 | 1.000 | 0.999 | 1.000 | 0.866 | 0.427 | 0.355 | 0.989 |  |  |  |
|  |  | -1.8 | -0.8 | -1 | -0.5 | -0.3 | -0.2 | 0.088 | 0.123 | -0.035 | -0.066 | 0.051 | -0.020 | -0.035 | 0.015 | 0.104 | 0.106 | 0.104 | 0.152 | 0.150 | 0.106 | 0.008 | 0.026 | 0.012 | 0.015 | 0.026 | 0.023 | 0.012 | 1.000 | 0.937 | 0.999 | 0.833 | 0.416 | 0.333 | 0.989 |  |  |  |
|  |  | 1 | 0 | 1 | 0.5 | 0.3 | 0.2 | -0.014 | 0.120 | -0.134 | -0.142 | -0.019 | 0.027 | -0.133 | 0.014 | 0.101 | 0.102 | 0.099 | 0.167 | 0.162 | 0.102 | 0.000 | 0.025 | 0.028 | 0.030 | 0.028 | 0.027 | 0.028 | 1.000 | 0.007 | 1.000 | 0.527 | 0.429 | 0.339 | 0.979 |  |  |  |
|  |  | 1.3 | 0.8 | 0.5 | 0 | 0.3 | 0.2 | 0.023 | 0.120 | -0.098 | -0.101 | -0.031 | 0.034 | -0.098 | 0.012 | 0.097 | 0.097 | 0.094 | 0.159 | 0.159 | 0.097 | 0.001 | 0.024 | 0.019 | 0.019 | 0.026 | 0.026 | 0.019 | 1.000 | 1.000 | 0.578 | 0.004 | 0.432 | 0.364 | 0.046 |  |  |  |
|  |  | 1.5 | 0.8 | 0.7 | 0.5 | 0 | 0.2 | -0.014 | 0.132 | -0.146 | -0.140 | -0.042 | 0.036 | -0.145 | 0.014 | 0.095 | 0.095 | 0.098 | 0.162 | 0.163 | 0.095 | 0.000 | 0.026 | 0.030 | 0.029 | 0.028 | 0.028 | 0.030 | 1.000 | 0.999 | 0.851 | 0.210 | 0.077 | 0.367 | 0.246 |  |  |  |
|  |  | 1 | 0.8 | 0.2 | 0 | 0 | 0.2 | 0.023 | 0.128 | -0.106 | -0.098 | -0.037 | 0.030 | -0.106 | 0.012 | 0.091 | 0.091 | 0.096 | 0.156 | 0.159 | 0.091 | 0.001 | 0.025 | 0.019 | 0.019 | 0.026 | 0.026 | 0.019 | 1.000 | 1.000 | 0.014 | 0.013 | 0.076 | 0.340 | 0.000 |  |  |  |
|  |  | 0 | -1 | 1 | 0.5 | 0.3 | 0.2 | -0.015 | 0.124 | -0.139 | -0.142 | -0.026 | 0.029 | -0.139 | 0.015 | 0.100 | 0.101 | 0.098 | 0.158 | 0.154 | 0.101 | 0.000 | 0.025 | 0.029 | 0.030 | 0.026 | 0.025 | 0.029 | 0.165 | 1.000 | 1.000 | 0.891 | 0.435 | 0.330 | 0.993 |  |  |  |
|  | Egger | 1.8 | 0.8 | 1 | 0.5 | 0.3 | 0.2 | 0.000 | -0.060 | 0.060 | 0.049 | -0.062 | 0.074 | 0.060 | 0.034 | 0.111 | 0.116 | 0.114 | 0.211 | 0.211 | 0.116 | 0.001 | 0.016 | 0.017 | 0.015 | 0.048 | 0.050 | 0.017 | 1.000 | 0.587 | 0.983 | 0.198 | 0.198 | 0.263 | 0.796 | 0.001 | 0.056 | 0.834 |
|  |  | 0.8 | 0.8 | 0 | -0.5 | 0.3 | 0.2 | 0.000 | -0.057 | 0.057 | 0.045 | -0.036 | 0.048 | 0.057 | 0.030 | 0.113 | 0.116 | 0.112 | 0.202 | 0.199 | 0.116 | 0.001 | 0.016 | 0.017 | 0.015 | 0.042 | 0.042 | 0.017 | 1.000 | 0.998 | 0.012 | 0.871 | 0.285 | 0.262 | 0.000 | 0.388 | 0.061 | 0.853 |
|  |  | 1.2 | 0.8 | 0.4 | 0.5 | -0.3 | 0.2 | 0.001 | -0.056 | 0.057 | 0.054 | 0.072 | -0.069 | 0.057 | 0.033 | 0.112 | 0.114 | 0.110 | 0.199 | 0.195 | 0.114 | 0.001 | 0.016 | 0.016 | 0.015 | 0.045 | 0.043 | 0.016 | 1.000 | 0.981 | 0.588 | 0.849 | 0.256 | 0.128 | 0.050 | 0.117 | 0.064 | 0.821 |
|  |  | 0.2 | 0.8 | -0.6 | -0.5 | -0.3 | 0.2 | 0.000 | -0.055 | 0.056 | 0.052 | 0.062 | -0.060 | 0.055 | 0.029 | 0.113 | 0.115 | 0.110 | 0.183 | 0.179 | 0.115 | 0.001 | 0.016 | 0.016 | 0.015 | 0.037 | 0.036 | 0.016 | 1.000 | 1.000 | 0.993 | 0.976 | 0.262 | 0.118 | 0.465 | 0.724 | 0.052 | 0.796 |
|  |  | 0.2 | -0.8 | 1 | 0.5 | 0.3 | 0.2 | 0.001 | -0.058 | 0.059 | 0.041 | -0.047 | 0.065 | 0.059 | 0.036 | 0.116 | 0.121 | 0.118 | 0.213 | 0.213 | 0.121 | 0.001 | 0.017 | 0.018 | 0.016 | 0.048 | 0.049 | 0.018 | 1.000 | 0.999 | 1.000 | 0.961 | 0.254 | 0.285 | 0.990 | 0.427 | 0.054 | 0.822 |
|  |  | -1.8 | -0.8 | -1 | -0.5 | -0.3 | -0.2 | -0.001 | -0.058 | 0.057 | 0.045 | 0.065 | -0.054 | 0.057 | 0.032 | 0.113 | 0.117 | 0.117 | 0.198 | 0.199 | 0.117 | 0.001 | 0.016 | 0.017 | 0.016 | 0.044 | 0.043 | 0.017 | 1.000 | 0.933 | 0.993 | 0.189 | 0.234 | 0.278 | 0.791 | 0.009 | 0.045 | 0.836 |
|  |  | 1 | 0 | 1 | 0.5 | 0.3 | 0.2 | -0.001 | -0.058 | 0.057 | 0.042 | -0.050 | 0.066 | 0.057 | 0.035 | 0.113 | 0.116 | 0.114 | 0.208 | 0.206 | 0.116 | 0.001 | 0.016 | 0.017 | 0.015 | 0.046 | 0.047 | 0.017 | 1.000 | 0.001 | 0.999 | 0.741 | 0.255 | 0.294 | 0.962 | 0.070 | 0.062 | 0.830 |
|  |  | 1.3 | 0.8 | 0.5 | 0 | 0.3 | 0.2 | 0.002 | -0.056 | 0.058 | 0.043 | -0.052 | 0.067 | 0.058 | 0.029 | 0.111 | 0.113 | 0.110 | 0.202 | 0.202 | 0.113 | 0.001 | 0.015 | 0.016 | 0.014 | 0.044 | 0.045 | 0.016 | 1.000 | 0.950 | 0.698 | 0.000 | 0.258 | 0.285 | 0.076 | 0.037 | 0.060 | 0.835 |
|  |  | 1.5 | 0.8 | 0.7 | 0.5 | 0 | 0.2 | 0.002 | -0.045 | 0.047 | 0.040 | -0.065 | 0.072 | 0.047 | 0.037 | 0.115 | 0.120 | 0.119 | 0.204 | 0.205 | 0.120 | 0.001 | 0.015 | 0.017 | 0.016 | 0.046 | 0.047 | 0.017 | 1.000 | 0.854 | 0.876 | 0.395 | 0.074 | 0.285 | 0.292 | 0.005 | 0.039 | 0.822 |
|  |  | 1 | 0.8 | 0.2 | 0 | 0 | 0.2 | 0.001 | -0.053 | 0.054 | 0.048 | -0.055 | 0.062 | 0.054 | 0.030 | 0.106 | 0.108 | 0.109 | 0.203 | 0.204 | 0.108 | 0.001 | 0.014 | 0.015 | 0.014 | 0.044 | 0.046 | 0.015 | 1.000 | 0.994 | 0.111 | 0.000 | 0.087 | 0.282 | 0.000 | 0.155 | 0.055 | 0.823 |
|  |  | 0 | -1 | 1 | 0.5 | 0.3 | 0.2 | 0.001 | -0.058 | 0.059 | 0.045 | -0.046 | 0.059 | 0.059 | 0.036 | 0.114 | 0.116 | 0.114 | 0.207 | 0.201 | 0.116 | 0.001 | 0.016 | 0.017 | 0.015 | 0.045 | 0.044 | 0.017 | 0.043 | 1.000 | 1.000 | 0.976 | 0.269 | 0.282 | 0.993 | 0.511 | 0.056 | 0.821 |

TE: total effect; DE: direct effect; IE: indirect effect; IE_d: indirect effect calculated by difference method; IE_p: indirect effect calculated by product method; IE1: *X*→*M_1_*→*Y*; IE2: *X*→*M_2_*→*Y*; IE3: *X*→*M_1_*→*M_2_*→*Y*.

**eTable 18** Simulation of PSE-Egger in two ordered mediators setting with different effects (case(a),sc22,sc23).

|  |  | true | | | | | | BIAS | | | | | | | SE | | | | | | | MSE | | | | | | | Power/Type I error | | | | | | | | | |
| --- | --- | --- | --- | --- | --- | --- | --- | --- | --- | --- | --- | --- | --- | --- | --- | --- | --- | --- | --- | --- | --- | --- | --- | --- | --- | --- | --- | --- | --- | --- | --- | --- | --- | --- | --- | --- | --- | --- |
|  |  | TE | DE | IE | IE1 | IE2 | IE3 | TE | DE | IE_d | IE1 | IE2 | IE3 | IE_p | TE | DE | IE_d | IE1 | IE2 | IE3 | IE_p | TE | DE | IE_d | IE1 | IE2 | IE3 | IE_p | TE | DE | IE_d | IE1 | IE2 | IE3 | IE_p | GY | GM1 | GM2 |
| Sc22 | IVW | 1.8 | 0.8 | 1 | 0.5 | 0.3 | 0.2 | 0.001 | 0.014 | -0.013 | 0.066 | -0.050 | -0.030 | -0.013 | 0.017 | 0.095 | 0.096 | 0.084 | 0.086 | 0.072 | 0.096 | 0.000 | 0.009 | 0.009 | 0.011 | 0.010 | 0.006 | 0.009 | 1.000 | 0.983 | 1.000 | 0.908 | 0.096 | 0.068 | 0.998 |  |  |  |
|  |  | 0.8 | 0.8 | 0 | -0.5 | 0.3 | 0.2 | 0.000 | 0.013 | -0.013 | 0.060 | -0.042 | -0.031 | -0.013 | 0.009 | 0.096 | 0.097 | 0.083 | 0.088 | 0.070 | 0.097 | 0.000 | 0.009 | 0.010 | 0.010 | 0.010 | 0.006 | 0.010 | 1.000 | 1.000 | 0.021 | 0.997 | 0.897 | 0.777 | 0.001 |  |  |  |
|  |  | 1.2 | 0.8 | 0.4 | 0.5 | -0.3 | 0.2 | -0.074 | 0.010 | -0.084 | -0.019 | -0.088 | 0.022 | -0.084 | 0.016 | 0.098 | 0.099 | 0.079 | 0.106 | 0.090 | 0.099 | 0.006 | 0.010 | 0.017 | 0.007 | 0.019 | 0.009 | 0.017 | 1.000 | 0.999 | 0.566 | 0.996 | 0.973 | 0.769 | 0.195 |  |  |  |
|  |  | 0.2 | 0.8 | -0.6 | -0.5 | -0.3 | 0.2 | -0.074 | 0.020 | -0.095 | -0.027 | -0.094 | 0.026 | -0.095 | 0.011 | 0.096 | 0.097 | 0.079 | 0.107 | 0.092 | 0.097 | 0.006 | 0.010 | 0.018 | 0.007 | 0.020 | 0.009 | 0.018 | 1.000 | 1.000 | 1.000 | 1.000 | 0.991 | 0.779 | 1.000 |  |  |  |
|  |  | 0.2 | -0.8 | 1 | 0.5 | 0.3 | 0.2 | 0.001 | 0.018 | -0.017 | 0.057 | -0.046 | -0.028 | -0.017 | 0.016 | 0.097 | 0.096 | 0.082 | 0.084 | 0.068 | 0.096 | 0.000 | 0.010 | 0.010 | 0.010 | 0.009 | 0.005 | 0.010 | 1.000 | 1.000 | 1.000 | 1.000 | 0.906 | 0.792 | 1.000 |  |  |  |
|  |  | -1.8 | -0.8 | -1 | -0.5 | -0.3 | -0.2 | -0.074 | 0.017 | -0.091 | 0.059 | -0.090 | -0.060 | -0.091 | 0.016 | 0.095 | 0.096 | 0.082 | 0.112 | 0.095 | 0.096 | 0.006 | 0.009 | 0.017 | 0.010 | 0.021 | 0.013 | 0.017 | 1.000 | 0.986 | 1.000 | 0.740 | 0.780 | 0.640 | 1.000 |  |  |  |
|  |  | 1 | 0 | 1 | 0.5 | 0.3 | 0.2 | 0.000 | 0.014 | -0.014 | 0.065 | -0.047 | -0.032 | -0.014 | 0.017 | 0.095 | 0.096 | 0.078 | 0.084 | 0.067 | 0.096 | 0.000 | 0.009 | 0.009 | 0.010 | 0.009 | 0.005 | 0.009 | 1.000 | 0.000 | 1.000 | 0.994 | 0.577 | 0.468 | 1.000 |  |  |  |
|  |  | 1.3 | 0.8 | 0.5 | 0 | 0.3 | 0.2 | 0.001 | 0.017 | -0.016 | 0.061 | -0.045 | -0.032 | -0.016 | 0.009 | 0.097 | 0.097 | 0.081 | 0.086 | 0.072 | 0.097 | 0.000 | 0.010 | 0.010 | 0.010 | 0.009 | 0.006 | 0.010 | 1.000 | 0.999 | 0.881 | 0.000 | 0.532 | 0.419 | 0.627 |  |  |  |
|  |  | 1.5 | 0.8 | 0.7 | 0.5 | 0 | 0.2 | 0.000 | -0.048 | 0.048 | 0.059 | 0.016 | -0.027 | 0.048 | 0.017 | 0.066 | 0.068 | 0.080 | 0.059 | 0.072 | 0.068 | 0.000 | 0.007 | 0.007 | 0.010 | 0.004 | 0.006 | 0.007 | 1.000 | 1.000 | 1.000 | 0.959 | 0.011 | 0.181 | 0.991 |  |  |  |
|  |  | 1 | 0.8 | 0.2 | 0 | 0 | 0.2 | 0.000 | -0.051 | 0.051 | 0.062 | 0.015 | -0.027 | 0.051 | 0.009 | 0.068 | 0.068 | 0.080 | 0.057 | 0.070 | 0.068 | 0.000 | 0.007 | 0.007 | 0.010 | 0.003 | 0.006 | 0.007 | 1.000 | 1.000 | 0.737 | 0.011 | 0.030 | 0.651 | 0.162 |  |  |  |
|  |  | 0 | -1 | 1 | 0.5 | 0.3 | 0.2 | 0.000 | 0.018 | -0.017 | 0.062 | -0.046 | -0.033 | -0.017 | 0.016 | 0.097 | 0.098 | 0.080 | 0.086 | 0.070 | 0.098 | 0.000 | 0.010 | 0.010 | 0.010 | 0.010 | 0.006 | 0.010 | 0.047 | 1.000 | 1.000 | 1.000 | 0.924 | 0.801 | 1.000 |  |  |  |
|  | Egger | 1.8 | 0.8 | 1 | 0.5 | 0.3 | 0.2 | 0.002 | -0.045 | 0.046 | -0.002 | 0.010 | 0.038 | 0.047 | 0.042 | 0.088 | 0.095 | 0.082 | 0.093 | 0.081 | 0.095 | 0.002 | 0.010 | 0.011 | 0.007 | 0.009 | 0.008 | 0.011 | 1.000 | 0.941 | 1.000 | 0.666 | 0.213 | 0.188 | 0.992 | 0.001 | 0.769 | 0.050 |
|  |  | 0.8 | 0.8 | 0 | -0.5 | 0.3 | 0.2 | 0.001 | -0.045 | 0.046 | -0.009 | 0.018 | 0.037 | 0.046 | 0.023 | 0.089 | 0.090 | 0.080 | 0.092 | 0.084 | 0.090 | 0.001 | 0.010 | 0.010 | 0.007 | 0.009 | 0.008 | 0.010 | 1.000 | 1.000 | 0.009 | 0.999 | 0.964 | 0.906 | 0.000 | 0.464 | 0.773 | 0.050 |
|  |  | 1.2 | 0.8 | 0.4 | 0.5 | -0.3 | 0.2 | 0.001 | -0.047 | 0.048 | 0.038 | 0.046 | -0.035 | 0.048 | 0.040 | 0.089 | 0.095 | 0.077 | 0.084 | 0.069 | 0.095 | 0.002 | 0.010 | 0.011 | 0.007 | 0.009 | 0.006 | 0.011 | 1.000 | 1.000 | 0.882 | 0.999 | 0.673 | 0.558 | 0.621 | 0.151 | 0.810 | 0.056 |
|  |  | 0.2 | 0.8 | -0.6 | -0.5 | -0.3 | 0.2 | 0.001 | -0.040 | 0.041 | 0.031 | 0.043 | -0.033 | 0.041 | 0.024 | 0.089 | 0.090 | 0.078 | 0.083 | 0.068 | 0.090 | 0.001 | 0.010 | 0.010 | 0.007 | 0.009 | 0.006 | 0.010 | 1.000 | 1.000 | 1.000 | 1.000 | 0.971 | 0.859 | 0.999 | 0.743 | 0.815 | 0.056 |
|  |  | 0.2 | -0.8 | 1 | 0.5 | 0.3 | 0.2 | 0.000 | -0.041 | 0.041 | -0.009 | 0.013 | 0.037 | 0.042 | 0.043 | 0.089 | 0.095 | 0.080 | 0.092 | 0.077 | 0.095 | 0.002 | 0.010 | 0.011 | 0.006 | 0.009 | 0.007 | 0.011 | 0.994 | 1.000 | 1.000 | 0.999 | 0.971 | 0.915 | 1.000 | 0.435 | 0.761 | 0.053 |
|  |  | -1.8 | -0.8 | -1 | -0.5 | -0.3 | -0.2 | -0.001 | -0.042 | 0.041 | -0.010 | 0.045 | 0.006 | 0.041 | 0.039 | 0.088 | 0.094 | 0.080 | 0.084 | 0.071 | 0.094 | 0.002 | 0.010 | 0.011 | 0.007 | 0.009 | 0.005 | 0.011 | 1.000 | 0.997 | 1.000 | 0.835 | 0.136 | 0.103 | 0.997 | 0.001 | 0.777 | 0.048 |
|  |  | 1 | 0 | 1 | 0.5 | 0.3 | 0.2 | 0.002 | -0.044 | 0.046 | -0.003 | 0.011 | 0.038 | 0.047 | 0.041 | 0.085 | 0.094 | 0.077 | 0.091 | 0.078 | 0.094 | 0.002 | 0.009 | 0.011 | 0.006 | 0.008 | 0.008 | 0.011 | 1.000 | 0.000 | 1.000 | 0.974 | 0.794 | 0.725 | 1.000 | 0.066 | 0.766 | 0.042 |
|  |  | 1.3 | 0.8 | 0.5 | 0 | 0.3 | 0.2 | 0.002 | -0.041 | 0.043 | -0.007 | 0.014 | 0.035 | 0.043 | 0.023 | 0.089 | 0.091 | 0.077 | 0.095 | 0.084 | 0.091 | 0.001 | 0.010 | 0.010 | 0.006 | 0.009 | 0.008 | 0.010 | 1.000 | 0.998 | 0.928 | 0.000 | 0.733 | 0.652 | 0.669 | 0.048 | 0.747 | 0.047 |
|  |  | 1.5 | 0.8 | 0.7 | 0.5 | 0 | 0.2 | 0.000 | -0.014 | 0.014 | -0.007 | -0.016 | 0.037 | 0.014 | 0.042 | 0.061 | 0.071 | 0.077 | 0.071 | 0.083 | 0.071 | 0.002 | 0.004 | 0.005 | 0.006 | 0.005 | 0.008 | 0.005 | 1.000 | 1.000 | 1.000 | 0.849 | 0.020 | 0.423 | 0.959 | 0.006 | 0.786 | 0.047 |
|  |  | 1 | 0.8 | 0.2 | 0 | 0 | 0.2 | 0.001 | -0.015 | 0.016 | -0.006 | -0.019 | 0.042 | 0.016 | 0.023 | 0.061 | 0.062 | 0.075 | 0.068 | 0.081 | 0.062 | 0.001 | 0.004 | 0.004 | 0.006 | 0.005 | 0.008 | 0.004 | 1.000 | 1.000 | 0.476 | 0.000 | 0.051 | 0.866 | 0.018 | 0.173 | 0.804 | 0.043 |
|  |  | 0 | -1 | 1 | 0.5 | 0.3 | 0.2 | 0.002 | -0.043 | 0.044 | -0.006 | 0.015 | 0.035 | 0.045 | 0.041 | 0.089 | 0.095 | 0.079 | 0.089 | 0.081 | 0.095 | 0.002 | 0.010 | 0.011 | 0.006 | 0.008 | 0.008 | 0.011 | 0.048 | 1.000 | 1.000 | 1.000 | 0.973 | 0.923 | 1.000 | 0.529 | 0.757 | 0.050 |
| Sc23 | IVW | 1.8 | 0.8 | 1 | 0.5 | 0.3 | 0.2 | -0.015 | -0.083 | 0.068 | -0.010 | 0.177 | -0.099 | 0.068 | 0.013 | 0.207 | 0.207 | 0.154 | 0.183 | 0.114 | 0.207 | 0.000 | 0.050 | 0.047 | 0.024 | 0.065 | 0.023 | 0.047 | 1.000 | 0.516 | 0.953 | 0.490 | 0.538 | 0.057 | 0.928 |  |  |  |
|  |  | 0.8 | 0.8 | 0 | -0.5 | 0.3 | 0.2 | 0.060 | -0.089 | 0.149 | 0.077 | 0.175 | -0.103 | 0.149 | 0.012 | 0.212 | 0.212 | 0.158 | 0.181 | 0.109 | 0.212 | 0.004 | 0.053 | 0.067 | 0.031 | 0.063 | 0.023 | 0.067 | 1.000 | 0.888 | 0.117 | 0.767 | 0.865 | 0.125 | 0.035 |  |  |  |
|  |  | 1.2 | 0.8 | 0.4 | 0.5 | -0.3 | 0.2 | -0.089 | -0.082 | -0.007 | 0.029 | -0.082 | 0.045 | -0.007 | 0.015 | 0.218 | 0.218 | 0.185 | 0.168 | 0.126 | 0.218 | 0.008 | 0.054 | 0.047 | 0.035 | 0.035 | 0.018 | 0.047 | 1.000 | 0.880 | 0.428 | 0.794 | 0.710 | 0.572 | 0.231 |  |  |  |
|  |  | 0.2 | 0.8 | -0.6 | -0.5 | -0.3 | 0.2 | -0.015 | -0.073 | 0.058 | 0.097 | -0.096 | 0.057 | 0.058 | 0.011 | 0.209 | 0.209 | 0.178 | 0.172 | 0.132 | 0.209 | 0.000 | 0.049 | 0.047 | 0.041 | 0.039 | 0.021 | 0.047 | 1.000 | 0.907 | 0.730 | 0.613 | 0.774 | 0.642 | 0.494 |  |  |  |
|  |  | 0.2 | -0.8 | 1 | 0.5 | 0.3 | 0.2 | -0.015 | -0.086 | 0.071 | -0.011 | 0.179 | -0.096 | 0.071 | 0.014 | 0.213 | 0.213 | 0.160 | 0.178 | 0.113 | 0.213 | 0.000 | 0.053 | 0.050 | 0.026 | 0.064 | 0.022 | 0.050 | 1.000 | 0.985 | 0.999 | 0.860 | 0.892 | 0.134 | 0.998 |  |  |  |
|  |  | -1.8 | -0.8 | -1 | -0.5 | -0.3 | -0.2 | 0.015 | -0.082 | 0.097 | 0.072 | -0.092 | 0.117 | 0.097 | 0.013 | 0.209 | 0.209 | 0.154 | 0.168 | 0.093 | 0.209 | 0.000 | 0.050 | 0.053 | 0.029 | 0.037 | 0.022 | 0.053 | 1.000 | 0.844 | 0.896 | 0.464 | 0.425 | 0.047 | 0.861 |  |  |  |
|  |  | 1 | 0 | 1 | 0.5 | 0.3 | 0.2 | -0.015 | -0.085 | 0.070 | -0.007 | 0.175 | -0.098 | 0.070 | 0.014 | 0.211 | 0.212 | 0.153 | 0.179 | 0.108 | 0.212 | 0.000 | 0.052 | 0.050 | 0.023 | 0.063 | 0.021 | 0.050 | 1.000 | 0.027 | 0.993 | 0.835 | 0.815 | 0.114 | 0.989 |  |  |  |
|  |  | 1.3 | 0.8 | 0.5 | 0 | 0.3 | 0.2 | 0.022 | -0.085 | 0.107 | 0.032 | 0.173 | -0.098 | 0.107 | 0.012 | 0.209 | 0.209 | 0.155 | 0.181 | 0.110 | 0.209 | 0.001 | 0.051 | 0.055 | 0.025 | 0.063 | 0.022 | 0.055 | 1.000 | 0.803 | 0.690 | 0.024 | 0.779 | 0.092 | 0.533 |  |  |  |
|  |  | 1.5 | 0.8 | 0.7 | 0.5 | 0 | 0.2 | -0.015 | -0.046 | 0.031 | -0.011 | 0.147 | -0.105 | 0.032 | 0.015 | 0.162 | 0.162 | 0.160 | 0.113 | 0.108 | 0.162 | 0.000 | 0.028 | 0.027 | 0.026 | 0.034 | 0.023 | 0.027 | 1.000 | 0.928 | 0.934 | 0.627 | 0.165 | 0.060 | 0.822 |  |  |  |
|  |  | 1 | 0.8 | 0.2 | 0 | 0 | 0.2 | 0.023 | -0.047 | 0.070 | 0.025 | 0.143 | -0.098 | 0.071 | 0.012 | 0.163 | 0.163 | 0.159 | 0.118 | 0.113 | 0.163 | 0.001 | 0.029 | 0.031 | 0.026 | 0.034 | 0.022 | 0.031 | 1.000 | 0.991 | 0.372 | 0.049 | 0.237 | 0.147 | 0.140 |  |  |  |
|  |  | 0 | -1 | 1 | 0.5 | 0.3 | 0.2 | -0.015 | -0.070 | 0.055 | -0.018 | 0.168 | -0.095 | 0.055 | 0.013 | 0.217 | 0.217 | 0.153 | 0.177 | 0.111 | 0.217 | 0.000 | 0.052 | 0.050 | 0.024 | 0.060 | 0.021 | 0.050 | 0.138 | 0.993 | 0.995 | 0.869 | 0.890 | 0.149 | 0.989 |  |  |  |
|  | Egger | 1.8 | 0.8 | 1 | 0.5 | 0.3 | 0.2 | 0.001 | -0.081 | 0.082 | 0.029 | -0.030 | 0.084 | 0.082 | 0.036 | 0.219 | 0.218 | 0.220 | 0.157 | 0.159 | 0.218 | 0.001 | 0.055 | 0.054 | 0.049 | 0.026 | 0.032 | 0.054 | 1.000 | 0.444 | 0.929 | 0.219 | 0.149 | 0.178 | 0.739 | 0.000 | 0.716 | 0.845 |
|  |  | 0.8 | 0.8 | 0 | -0.5 | 0.3 | 0.2 | 0.003 | -0.083 | 0.086 | 0.035 | -0.025 | 0.076 | 0.086 | 0.029 | 0.224 | 0.223 | 0.219 | 0.152 | 0.159 | 0.223 | 0.001 | 0.057 | 0.057 | 0.049 | 0.024 | 0.031 | 0.057 | 1.000 | 0.868 | 0.078 | 0.560 | 0.447 | 0.457 | 0.011 | 0.042 | 0.712 | 0.822 |
|  |  | 1.2 | 0.8 | 0.4 | 0.5 | -0.3 | 0.2 | -0.001 | -0.079 | 0.078 | 0.069 | 0.080 | -0.071 | 0.078 | 0.032 | 0.227 | 0.228 | 0.212 | 0.149 | 0.122 | 0.228 | 0.001 | 0.058 | 0.058 | 0.049 | 0.029 | 0.020 | 0.058 | 1.000 | 0.841 | 0.549 | 0.746 | 0.327 | 0.150 | 0.346 | 0.019 | 0.732 | 0.838 |
|  |  | 0.2 | 0.8 | -0.6 | -0.5 | -0.3 | 0.2 | 0.000 | -0.070 | 0.071 | 0.062 | 0.071 | -0.063 | 0.070 | 0.028 | 0.222 | 0.222 | 0.207 | 0.151 | 0.126 | 0.222 | 0.001 | 0.054 | 0.054 | 0.047 | 0.028 | 0.020 | 0.054 | 1.000 | 0.870 | 0.654 | 0.564 | 0.395 | 0.172 | 0.466 | 0.047 | 0.771 | 0.808 |
|  |  | 0.2 | -0.8 | 1 | 0.5 | 0.3 | 0.2 | 0.001 | -0.088 | 0.089 | 0.029 | -0.024 | 0.084 | 0.089 | 0.036 | 0.224 | 0.226 | 0.229 | 0.164 | 0.155 | 0.226 | 0.001 | 0.058 | 0.059 | 0.053 | 0.027 | 0.031 | 0.059 | 0.999 | 0.966 | 0.997 | 0.643 | 0.443 | 0.457 | 0.982 | 0.044 | 0.701 | 0.827 |
|  |  | -1.8 | -0.8 | -1 | -0.5 | -0.3 | -0.2 | 0.001 | -0.081 | 0.082 | 0.038 | 0.076 | -0.032 | 0.082 | 0.032 | 0.218 | 0.219 | 0.220 | 0.146 | 0.148 | 0.219 | 0.001 | 0.054 | 0.055 | 0.050 | 0.027 | 0.023 | 0.055 | 1.000 | 0.797 | 0.868 | 0.216 | 0.140 | 0.148 | 0.653 | 0.001 | 0.728 | 0.817 |
|  |  | 1 | 0 | 1 | 0.5 | 0.3 | 0.2 | 0.001 | -0.085 | 0.086 | 0.037 | -0.028 | 0.078 | 0.086 | 0.037 | 0.226 | 0.226 | 0.226 | 0.159 | 0.159 | 0.226 | 0.001 | 0.058 | 0.059 | 0.052 | 0.026 | 0.032 | 0.059 | 1.000 | 0.030 | 0.991 | 0.574 | 0.381 | 0.405 | 0.953 | 0.020 | 0.713 | 0.812 |
|  |  | 1.3 | 0.8 | 0.5 | 0 | 0.3 | 0.2 | 0.000 | -0.087 | 0.088 | 0.042 | -0.025 | 0.071 | 0.088 | 0.031 | 0.225 | 0.224 | 0.232 | 0.162 | 0.161 | 0.224 | 0.001 | 0.058 | 0.058 | 0.055 | 0.027 | 0.031 | 0.058 | 1.000 | 0.746 | 0.597 | 0.029 | 0.341 | 0.357 | 0.270 | 0.011 | 0.698 | 0.819 |
|  |  | 1.5 | 0.8 | 0.7 | 0.5 | 0 | 0.2 | 0.001 | -0.044 | 0.046 | 0.028 | -0.054 | 0.072 | 0.046 | 0.037 | 0.203 | 0.203 | 0.226 | 0.120 | 0.160 | 0.203 | 0.001 | 0.043 | 0.043 | 0.052 | 0.017 | 0.031 | 0.043 | 1.000 | 0.774 | 0.783 | 0.351 | 0.016 | 0.272 | 0.453 | 0.002 | 0.744 | 0.824 |
|  |  | 1 | 0.8 | 0.2 | 0 | 0 | 0.2 | 0.002 | -0.045 | 0.047 | 0.024 | -0.058 | 0.081 | 0.047 | 0.029 | 0.207 | 0.206 | 0.227 | 0.129 | 0.159 | 0.206 | 0.001 | 0.045 | 0.045 | 0.052 | 0.020 | 0.032 | 0.045 | 1.000 | 0.918 | 0.209 | 0.051 | 0.062 | 0.485 | 0.052 | 0.034 | 0.766 | 0.830 |
|  |  | 0 | -1 | 1 | 0.5 | 0.3 | 0.2 | 0.001 | -0.070 | 0.071 | 0.025 | -0.033 | 0.079 | 0.071 | 0.037 | 0.231 | 0.231 | 0.222 | 0.162 | 0.167 | 0.231 | 0.001 | 0.058 | 0.058 | 0.050 | 0.027 | 0.034 | 0.058 | 0.064 | 0.990 | 0.993 | 0.661 | 0.432 | 0.457 | 0.974 | 0.048 | 0.719 | 0.836 |

TE: total effect; DE: direct effect; IE: indirect effect; IE_d: indirect effect calculated by difference method; IE_p: indirect effect calculated by product method; IE1: *X*→*M_1_*→*Y*; IE2: *X*→*M_2_*→*Y*; IE3: *X*→*M_1_*→*M_2_*→*Y*.

**eTable 19** Simulation of PSE-Egger in two ordered mediators setting with different effects (case(b),sc3,sc31).

|  |  | true | | | | | | BIAS | | | | | | | SE | | | | | | | MSE | | | | | | | Power/Type I error | | | | | | | | | |
| --- | --- | --- | --- | --- | --- | --- | --- | --- | --- | --- | --- | --- | --- | --- | --- | --- | --- | --- | --- | --- | --- | --- | --- | --- | --- | --- | --- | --- | --- | --- | --- | --- | --- | --- | --- | --- | --- | --- |
|  |  | TE | DE | IE | IE1 | IE2 | IE3 | TE | DE | IE_d | IE1 | IE2 | IE3 | IE_p | TE | DE | IE_d | IE1 | IE2 | IE3 | IE_p | TE | DE | IE_d | IE1 | IE2 | IE3 | IE_p | TE | DE | IE_d | IE1 | IE2 | IE3 | IE_p | GY | GM1 | GM2 |
| Sc3 | IVW | 1.8 | 0.8 | 1 | 0.5 | 0.3 | 0.2 | 0.127 | -0.195 | 0.323 | 0.121 | -0.113 | 0.315 | 0.323 | 0.015 | 0.105 | 0.108 | 0.129 | 0.138 | 0.160 | 0.108 | 0.000 | 0.050 | 0.047 | 0.024 | 0.065 | 0.023 | 0.047 | 1.000 | 0.355 | 1.000 | 0.206 | 0.118 | 0.570 | 0.946 |  |  |  |
|  |  | 0.8 | 0.8 | 0 | -0.5 | 0.3 | 0.2 | 0.052 | -0.191 | 0.243 | 0.042 | -0.112 | 0.313 | 0.243 | 0.009 | 0.108 | 0.109 | 0.132 | 0.137 | 0.153 | 0.109 | 0.004 | 0.053 | 0.067 | 0.031 | 0.063 | 0.023 | 0.067 | 1.000 | 0.988 | 0.370 | 0.787 | 0.280 | 0.957 | 0.003 |  |  |  |
|  |  | 1.2 | 0.8 | 0.4 | 0.5 | -0.3 | 0.2 | 0.052 | -0.190 | 0.242 | 0.231 | 0.198 | -0.188 | 0.242 | 0.011 | 0.105 | 0.106 | 0.102 | 0.082 | 0.076 | 0.106 | 0.008 | 0.054 | 0.047 | 0.035 | 0.035 | 0.018 | 0.047 | 1.000 | 0.933 | 0.966 | 0.991 | 0.082 | 0.014 | 0.857 |  |  |  |
|  |  | 0.2 | 0.8 | -0.6 | -0.5 | -0.3 | 0.2 | -0.022 | -0.189 | 0.167 | 0.158 | 0.191 | -0.182 | 0.167 | 0.008 | 0.103 | 0.103 | 0.099 | 0.086 | 0.080 | 0.103 | 0.000 | 0.049 | 0.047 | 0.041 | 0.039 | 0.021 | 0.047 | 1.000 | 1.000 | 0.973 | 0.912 | 0.267 | 0.056 | 0.874 |  |  |  |
|  |  | 0.2 | -0.8 | 1 | 0.5 | 0.3 | 0.2 | 0.127 | -0.188 | 0.315 | 0.119 | -0.114 | 0.311 | 0.315 | 0.015 | 0.107 | 0.109 | 0.130 | 0.132 | 0.152 | 0.109 | 0.000 | 0.053 | 0.050 | 0.026 | 0.064 | 0.022 | 0.050 | 1.000 | 1.000 | 1.000 | 0.964 | 0.261 | 0.954 | 1.000 |  |  |  |
|  |  | -1.8 | -0.8 | -1 | -0.5 | -0.3 | -0.2 | -0.052 | -0.191 | 0.139 | 0.054 | 0.190 | -0.105 | 0.139 | 0.011 | 0.103 | 0.103 | 0.127 | 0.083 | 0.113 | 0.103 | 0.000 | 0.050 | 0.053 | 0.029 | 0.037 | 0.022 | 0.053 | 1.000 | 0.987 | 0.979 | 0.057 | 0.010 | 0.081 | 0.540 |  |  |  |
|  |  | 1 | 0 | 1 | 0.5 | 0.3 | 0.2 | 0.127 | -0.191 | 0.319 | 0.120 | -0.111 | 0.310 | 0.319 | 0.015 | 0.112 | 0.113 | 0.135 | 0.137 | 0.151 | 0.112 | 0.000 | 0.052 | 0.050 | 0.023 | 0.063 | 0.021 | 0.050 | 1.000 | 0.009 | 1.000 | 0.706 | 0.239 | 0.909 | 0.999 |  |  |  |
|  |  | 1.3 | 0.8 | 0.5 | 0 | 0.3 | 0.2 | 0.090 | -0.190 | 0.280 | 0.083 | -0.112 | 0.309 | 0.280 | 0.011 | 0.104 | 0.104 | 0.128 | 0.142 | 0.159 | 0.104 | 0.001 | 0.051 | 0.055 | 0.025 | 0.063 | 0.022 | 0.055 | 1.000 | 0.828 | 0.983 | 0.000 | 0.244 | 0.871 | 0.549 |  |  |  |
|  |  | 1.5 | 0.8 | 0.7 | 0.5 | 0 | 0.2 | 0.126 | -0.112 | 0.239 | 0.120 | -0.194 | 0.312 | 0.239 | 0.015 | 0.108 | 0.110 | 0.132 | 0.137 | 0.159 | 0.109 | 0.000 | 0.028 | 0.027 | 0.026 | 0.034 | 0.023 | 0.027 | 1.000 | 0.786 | 0.987 | 0.377 | 0.189 | 0.728 | 0.580 |  |  |  |
|  |  | 1 | 0.8 | 0.2 | 0 | 0 | 0.2 | 0.090 | -0.109 | 0.199 | 0.079 | -0.200 | 0.320 | 0.199 | 0.010 | 0.106 | 0.107 | 0.131 | 0.136 | 0.157 | 0.107 | 0.001 | 0.029 | 0.031 | 0.026 | 0.034 | 0.022 | 0.031 | 1.000 | 0.986 | 0.555 | 0.002 | 0.312 | 0.946 | 0.026 |  |  |  |
|  |  | 0 | -1 | 1 | 0.5 | 0.3 | 0.2 | 0.128 | -0.194 | 0.322 | 0.128 | -0.123 | 0.317 | 0.322 | 0.014 | 0.107 | 0.108 | 0.138 | 0.130 | 0.155 | 0.108 | 0.000 | 0.052 | 0.050 | 0.024 | 0.060 | 0.021 | 0.050 | 1.000 | 1.000 | 1.000 | 0.972 | 0.257 | 0.965 | 1.000 |  |  |  |
|  | Egger | 1.8 | 0.8 | 1 | 0.5 | 0.3 | 0.2 | 0.002 | -0.080 | 0.082 | 0.027 | -0.027 | 0.083 | 0.082 | 0.029 | 0.123 | 0.125 | 0.123 | 0.130 | 0.134 | 0.125 | 0.001 | 0.055 | 0.054 | 0.049 | 0.026 | 0.032 | 0.054 | 1.000 | 0.306 | 0.950 | 0.082 | 0.080 | 0.094 | 0.732 | 0.000 | 0.522 | 0.801 |
|  |  | 0.8 | 0.8 | 0 | -0.5 | 0.3 | 0.2 | 0.000 | -0.079 | 0.079 | 0.027 | -0.026 | 0.079 | 0.079 | 0.021 | 0.126 | 0.127 | 0.123 | 0.135 | 0.136 | 0.127 | 0.001 | 0.057 | 0.057 | 0.049 | 0.024 | 0.031 | 0.057 | 1.000 | 0.988 | 0.017 | 0.846 | 0.567 | 0.601 | 0.000 | 0.125 | 0.534 | 0.789 |
|  |  | 1.2 | 0.8 | 0.4 | 0.5 | -0.3 | 0.2 | 0.000 | -0.076 | 0.077 | 0.066 | 0.084 | -0.074 | 0.077 | 0.025 | 0.121 | 0.122 | 0.113 | 0.113 | 0.103 | 0.122 | 0.001 | 0.058 | 0.058 | 0.049 | 0.029 | 0.020 | 0.058 | 1.000 | 0.922 | 0.525 | 0.838 | 0.324 | 0.100 | 0.222 | 0.008 | 0.552 | 0.818 |
|  |  | 0.2 | 0.8 | -0.6 | -0.5 | -0.3 | 0.2 | 0.000 | -0.076 | 0.076 | 0.067 | 0.079 | -0.071 | 0.076 | 0.020 | 0.121 | 0.122 | 0.114 | 0.113 | 0.104 | 0.122 | 0.001 | 0.054 | 0.054 | 0.047 | 0.028 | 0.020 | 0.054 | 1.000 | 1.000 | 0.984 | 0.970 | 0.619 | 0.281 | 0.895 | 0.407 | 0.556 | 0.788 |
|  |  | 0.2 | -0.8 | 1 | 0.5 | 0.3 | 0.2 | 0.004 | -0.073 | 0.077 | 0.025 | -0.033 | 0.084 | 0.077 | 0.029 | 0.117 | 0.119 | 0.122 | 0.130 | 0.129 | 0.119 | 0.001 | 0.058 | 0.059 | 0.053 | 0.027 | 0.031 | 0.059 | 1.000 | 0.999 | 1.000 | 0.905 | 0.528 | 0.621 | 1.000 | 0.140 | 0.477 | 0.806 |
|  |  | -1.8 | -0.8 | -1 | -0.5 | -0.3 | -0.2 | -0.001 | -0.078 | 0.077 | 0.033 | 0.075 | -0.031 | 0.077 | 0.026 | 0.120 | 0.121 | 0.119 | 0.111 | 0.115 | 0.121 | 0.001 | 0.054 | 0.055 | 0.050 | 0.027 | 0.023 | 0.055 | 1.000 | 0.847 | 0.938 | 0.083 | 0.056 | 0.060 | 0.702 | 0.000 | 0.512 | 0.803 |
|  |  | 1 | 0 | 1 | 0.5 | 0.3 | 0.2 | 0.004 | -0.078 | 0.082 | 0.031 | -0.027 | 0.078 | 0.082 | 0.031 | 0.125 | 0.127 | 0.127 | 0.132 | 0.129 | 0.127 | 0.001 | 0.058 | 0.059 | 0.052 | 0.026 | 0.032 | 0.059 | 1.000 | 0.000 | 0.997 | 0.539 | 0.367 | 0.398 | 0.986 | 0.005 | 0.535 | 0.786 |
|  |  | 1.3 | 0.8 | 0.5 | 0 | 0.3 | 0.2 | 0.003 | -0.079 | 0.081 | 0.030 | -0.027 | 0.079 | 0.081 | 0.022 | 0.119 | 0.120 | 0.118 | 0.135 | 0.131 | 0.120 | 0.001 | 0.058 | 0.058 | 0.055 | 0.027 | 0.031 | 0.058 | 1.000 | 0.821 | 0.592 | 0.000 | 0.346 | 0.379 | 0.129 | 0.001 | 0.514 | 0.776 |
|  |  | 1.5 | 0.8 | 0.7 | 0.5 | 0 | 0.2 | 0.002 | -0.046 | 0.049 | 0.027 | -0.060 | 0.082 | 0.049 | 0.030 | 0.112 | 0.116 | 0.126 | 0.123 | 0.135 | 0.116 | 0.001 | 0.043 | 0.043 | 0.052 | 0.017 | 0.031 | 0.043 | 1.000 | 0.798 | 0.830 | 0.212 | 0.015 | 0.205 | 0.365 | 0.000 | 0.575 | 0.797 |
|  |  | 1 | 0.8 | 0.2 | 0 | 0 | 0.2 | 0.002 | -0.046 | 0.048 | 0.027 | -0.061 | 0.081 | 0.048 | 0.021 | 0.108 | 0.108 | 0.121 | 0.124 | 0.137 | 0.108 | 0.001 | 0.045 | 0.045 | 0.052 | 0.020 | 0.032 | 0.045 | 1.000 | 0.992 | 0.071 | 0.000 | 0.068 | 0.523 | 0.001 | 0.023 | 0.597 | 0.796 |
|  |  | 0 | -1 | 1 | 0.5 | 0.3 | 0.2 | 0.001 | -0.077 | 0.079 | 0.031 | -0.037 | 0.085 | 0.079 | 0.030 | 0.125 | 0.127 | 0.125 | 0.133 | 0.136 | 0.127 | 0.001 | 0.058 | 0.058 | 0.050 | 0.027 | 0.034 | 0.058 | 0.050 | 1.000 | 1.000 | 0.935 | 0.550 | 0.612 | 0.998 | 0.187 | 0.481 | 0.828 |
| Sc31 | IVW | 1.8 | 0.8 | 1 | 0.5 | 0.3 | 0.2 | 0.038 | 0.013 | 0.025 | 0.010 | -0.006 | 0.021 | 0.025 | 0.019 | 0.081 | 0.082 | 0.077 | 0.116 | 0.114 | 0.082 | 0.002 | 0.007 | 0.007 | 0.006 | 0.013 | 0.014 | 0.007 | 1.000 | 0.999 | 1.000 | 0.910 | 0.741 | 0.514 | 0.998 |  |  |  |
|  |  | 0.8 | 0.8 | 0 | -0.5 | 0.3 | 0.2 | 0.038 | 0.012 | 0.026 | 0.010 | -0.005 | 0.021 | 0.026 | 0.013 | 0.078 | 0.079 | 0.072 | 0.116 | 0.115 | 0.079 | 0.002 | 0.006 | 0.007 | 0.005 | 0.013 | 0.014 | 0.007 | 1.000 | 1.000 | 0.018 | 1.000 | 0.764 | 0.560 | 0.000 |  |  |  |
|  |  | 1.2 | 0.8 | 0.4 | 0.5 | -0.3 | 0.2 | 0.038 | 0.012 | 0.026 | 0.023 | 0.018 | -0.016 | 0.026 | 0.017 | 0.083 | 0.084 | 0.077 | 0.107 | 0.102 | 0.084 | 0.002 | 0.007 | 0.008 | 0.006 | 0.012 | 0.011 | 0.008 | 1.000 | 1.000 | 0.952 | 1.000 | 0.807 | 0.470 | 0.574 |  |  |  |
|  |  | 0.2 | 0.8 | -0.6 | -0.5 | -0.3 | 0.2 | 0.038 | 0.019 | 0.019 | 0.016 | 0.018 | -0.016 | 0.018 | 0.013 | 0.078 | 0.078 | 0.073 | 0.117 | 0.112 | 0.078 | 0.002 | 0.006 | 0.006 | 0.006 | 0.014 | 0.013 | 0.006 | 1.000 | 1.000 | 1.000 | 1.000 | 0.763 | 0.437 | 0.995 |  |  |  |
|  |  | 0.2 | -0.8 | 1 | 0.5 | 0.3 | 0.2 | 0.038 | 0.018 | 0.020 | 0.008 | -0.017 | 0.030 | 0.020 | 0.019 | 0.080 | 0.081 | 0.076 | 0.110 | 0.110 | 0.081 | 0.002 | 0.007 | 0.007 | 0.006 | 0.012 | 0.013 | 0.007 | 1.000 | 1.000 | 1.000 | 1.000 | 0.753 | 0.592 | 1.000 |  |  |  |
|  |  | -1.8 | -0.8 | -1 | -0.5 | -0.3 | -0.2 | 0.037 | 0.017 | 0.020 | 0.005 | 0.024 | -0.009 | 0.020 | 0.017 | 0.078 | 0.079 | 0.074 | 0.112 | 0.108 | 0.079 | 0.002 | 0.006 | 0.007 | 0.006 | 0.013 | 0.012 | 0.007 | 1.000 | 0.999 | 1.000 | 0.960 | 0.754 | 0.531 | 1.000 |  |  |  |
|  |  | 1 | 0 | 1 | 0.5 | 0.3 | 0.2 | 0.037 | 0.013 | 0.024 | 0.011 | -0.011 | 0.025 | 0.025 | 0.018 | 0.077 | 0.079 | 0.075 | 0.115 | 0.113 | 0.079 | 0.002 | 0.006 | 0.007 | 0.006 | 0.013 | 0.013 | 0.007 | 1.000 | 0.000 | 1.000 | 0.990 | 0.763 | 0.581 | 1.000 |  |  |  |
|  |  | 1.3 | 0.8 | 0.5 | 0 | 0.3 | 0.2 | 0.038 | 0.012 | 0.026 | 0.013 | -0.009 | 0.023 | 0.026 | 0.013 | 0.081 | 0.080 | 0.078 | 0.113 | 0.109 | 0.080 | 0.002 | 0.007 | 0.007 | 0.006 | 0.013 | 0.012 | 0.007 | 1.000 | 1.000 | 0.986 | 0.001 | 0.764 | 0.549 | 0.855 |  |  |  |
|  |  | 1.5 | 0.8 | 0.7 | 0.5 | 0 | 0.2 | 0.038 | 0.025 | 0.013 | 0.006 | -0.016 | 0.024 | 0.013 | 0.019 | 0.068 | 0.070 | 0.075 | 0.110 | 0.113 | 0.070 | 0.002 | 0.005 | 0.005 | 0.006 | 0.012 | 0.013 | 0.005 | 1.000 | 1.000 | 1.000 | 0.964 | 0.062 | 0.526 | 0.980 |  |  |  |
|  |  | 1 | 0.8 | 0.2 | 0 | 0 | 0.2 | 0.038 | 0.022 | 0.015 | 0.010 | -0.014 | 0.020 | 0.016 | 0.012 | 0.068 | 0.069 | 0.076 | 0.107 | 0.111 | 0.069 | 0.002 | 0.005 | 0.005 | 0.006 | 0.012 | 0.013 | 0.005 | 1.000 | 1.000 | 0.519 | 0.004 | 0.057 | 0.549 | 0.022 |  |  |  |
|  |  | 0 | -1 | 1 | 0.5 | 0.3 | 0.2 | 0.037 | 0.011 | 0.026 | 0.010 | -0.005 | 0.022 | 0.027 | 0.018 | 0.080 | 0.083 | 0.074 | 0.118 | 0.115 | 0.082 | 0.002 | 0.006 | 0.008 | 0.006 | 0.014 | 0.014 | 0.008 | 0.476 | 1.000 | 1.000 | 1.000 | 0.778 | 0.540 | 1.000 |  |  |  |
|  | Egger | 1.8 | 0.8 | 1 | 0.5 | 0.3 | 0.2 | 0.000 | -0.021 | 0.021 | 0.007 | -0.007 | 0.022 | 0.021 | 0.045 | 0.069 | 0.079 | 0.069 | 0.118 | 0.116 | 0.079 | 0.002 | 0.005 | 0.007 | 0.005 | 0.014 | 0.014 | 0.007 | 1.000 | 0.997 | 1.000 | 0.907 | 0.688 | 0.491 | 1.000 | 0.002 | 0.044 | 0.038 |
|  |  | 0.8 | 0.8 | 0 | -0.5 | 0.3 | 0.2 | 0.002 | -0.024 | 0.026 | 0.010 | -0.004 | 0.020 | 0.026 | 0.031 | 0.071 | 0.074 | 0.066 | 0.120 | 0.117 | 0.074 | 0.001 | 0.006 | 0.006 | 0.005 | 0.015 | 0.014 | 0.006 | 1.000 | 1.000 | 0.006 | 1.000 | 0.730 | 0.530 | 0.000 | 0.463 | 0.050 | 0.048 |
|  |  | 1.2 | 0.8 | 0.4 | 0.5 | -0.3 | 0.2 | 0.001 | -0.026 | 0.027 | 0.024 | 0.017 | -0.014 | 0.027 | 0.044 | 0.072 | 0.082 | 0.069 | 0.110 | 0.104 | 0.082 | 0.002 | 0.006 | 0.007 | 0.005 | 0.012 | 0.011 | 0.007 | 1.000 | 1.000 | 0.932 | 1.000 | 0.759 | 0.459 | 0.506 | 0.130 | 0.055 | 0.049 |
|  |  | 0.2 | 0.8 | -0.6 | -0.5 | -0.3 | 0.2 | -0.002 | -0.023 | 0.021 | 0.019 | 0.018 | -0.017 | 0.020 | 0.031 | 0.070 | 0.074 | 0.067 | 0.119 | 0.112 | 0.074 | 0.001 | 0.005 | 0.006 | 0.005 | 0.014 | 0.013 | 0.006 | 1.000 | 1.000 | 1.000 | 1.000 | 0.730 | 0.420 | 0.991 | 0.783 | 0.057 | 0.056 |
|  |  | 0.2 | -0.8 | 1 | 0.5 | 0.3 | 0.2 | 0.004 | -0.022 | 0.026 | 0.009 | -0.015 | 0.032 | 0.026 | 0.047 | 0.071 | 0.082 | 0.068 | 0.113 | 0.112 | 0.082 | 0.002 | 0.005 | 0.007 | 0.005 | 0.013 | 0.014 | 0.007 | 0.981 | 1.000 | 1.000 | 1.000 | 0.717 | 0.581 | 1.000 | 0.467 | 0.059 | 0.050 |
|  |  | -1.8 | -0.8 | -1 | -0.5 | -0.3 | -0.2 | -0.001 | -0.022 | 0.020 | 0.005 | 0.024 | -0.009 | 0.020 | 0.042 | 0.070 | 0.080 | 0.069 | 0.114 | 0.108 | 0.080 | 0.002 | 0.005 | 0.007 | 0.005 | 0.014 | 0.012 | 0.007 | 1.000 | 0.998 | 1.000 | 0.967 | 0.693 | 0.511 | 0.999 | 0.013 | 0.053 | 0.038 |
|  |  | 1 | 0 | 1 | 0.5 | 0.3 | 0.2 | 0.001 | -0.023 | 0.024 | 0.007 | -0.009 | 0.026 | 0.024 | 0.045 | 0.070 | 0.080 | 0.069 | 0.120 | 0.114 | 0.080 | 0.002 | 0.005 | 0.007 | 0.005 | 0.014 | 0.014 | 0.007 | 1.000 | 0.000 | 1.000 | 0.993 | 0.725 | 0.573 | 1.000 | 0.078 | 0.046 | 0.042 |
|  |  | 1.3 | 0.8 | 0.5 | 0 | 0.3 | 0.2 | 0.002 | -0.024 | 0.026 | 0.012 | -0.009 | 0.024 | 0.026 | 0.031 | 0.071 | 0.075 | 0.066 | 0.116 | 0.113 | 0.076 | 0.001 | 0.006 | 0.006 | 0.005 | 0.014 | 0.013 | 0.006 | 1.000 | 1.000 | 0.991 | 0.000 | 0.735 | 0.537 | 0.808 | 0.067 | 0.054 | 0.056 |
|  |  | 1.5 | 0.8 | 0.7 | 0.5 | 0 | 0.2 | 0.002 | -0.012 | 0.014 | 0.007 | -0.018 | 0.025 | 0.014 | 0.047 | 0.061 | 0.075 | 0.070 | 0.113 | 0.116 | 0.075 | 0.002 | 0.004 | 0.006 | 0.005 | 0.013 | 0.014 | 0.006 | 1.000 | 1.000 | 1.000 | 0.960 | 0.057 | 0.506 | 0.976 | 0.007 | 0.051 | 0.051 |
|  |  | 1 | 0.8 | 0.2 | 0 | 0 | 0.2 | 0.003 | -0.017 | 0.020 | 0.011 | -0.013 | 0.022 | 0.020 | 0.030 | 0.062 | 0.067 | 0.067 | 0.113 | 0.114 | 0.067 | 0.001 | 0.004 | 0.005 | 0.005 | 0.013 | 0.013 | 0.005 | 1.000 | 1.000 | 0.450 | 0.000 | 0.066 | 0.525 | 0.013 | 0.201 | 0.046 | 0.054 |
|  |  | 0 | -1 | 1 | 0.5 | 0.3 | 0.2 | -0.002 | -0.025 | 0.024 | 0.009 | -0.006 | 0.021 | 0.024 | 0.044 | 0.069 | 0.079 | 0.068 | 0.121 | 0.116 | 0.079 | 0.002 | 0.005 | 0.007 | 0.005 | 0.015 | 0.014 | 0.007 | 0.038 | 1.000 | 1.000 | 1.000 | 0.738 | 0.523 | 1.000 | 0.593 | 0.058 | 0.045 |

TE: total effect; DE: direct effect; IE: indirect effect; IE_d: indirect effect calculated by difference method; IE_p: indirect effect calculated by product method; IE1: *X*→*M_1_*→*Y*; IE2: *X*→*M_2_*→*Y*; IE3: *X*→*M_1_*→*M_2_*→*Y*.

**eTable 20** Simulation of PSE-Egger in two ordered mediators setting with different effects (case(b),sc32,sc33).

|  |  | true | | | | | | BIAS | | | | | | | SE | | | | | | | MSE | | | | | | | Power/Type I error | | | | | | | | | |
| --- | --- | --- | --- | --- | --- | --- | --- | --- | --- | --- | --- | --- | --- | --- | --- | --- | --- | --- | --- | --- | --- | --- | --- | --- | --- | --- | --- | --- | --- | --- | --- | --- | --- | --- | --- | --- | --- | --- |
|  |  | TE | DE | IE | IE1 | IE2 | IE3 | TE | DE | IE_d | IE1 | IE2 | IE3 | IE_p | TE | DE | IE_d | IE1 | IE2 | IE3 | IE_p | TE | DE | IE_d | IE1 | IE2 | IE3 | IE_p | TE | DE | IE_d | IE1 | IE2 | IE3 | IE_p | GY | GM1 | GM2 |
| Sc32 | IVW | 1.8 | 0.8 | 1 | 0.5 | 0.3 | 0.2 | 0.038 | -0.041 | 0.078 | 0.007 | 0.043 | 0.028 | 0.079 | 0.018 | 0.148 | 0.147 | 0.127 | 0.132 | 0.102 | 0.147 | 0.002 | 0.023 | 0.028 | 0.016 | 0.019 | 0.011 | 0.028 | 1.000 | 0.917 | 0.998 | 0.748 | 0.486 | 0.396 | 0.991 |  |  |  |
|  |  | 0.8 | 0.8 | 0 | -0.5 | 0.3 | 0.2 | 0.038 | -0.040 | 0.078 | 0.001 | 0.041 | 0.035 | 0.077 | 0.013 | 0.152 | 0.153 | 0.124 | 0.129 | 0.102 | 0.153 | 0.002 | 0.025 | 0.029 | 0.015 | 0.018 | 0.012 | 0.029 | 1.000 | 0.992 | 0.099 | 0.971 | 0.890 | 0.796 | 0.008 |  |  |  |
|  |  | 1.2 | 0.8 | 0.4 | 0.5 | -0.3 | 0.2 | -0.037 | -0.056 | 0.020 | 0.043 | 0.011 | -0.035 | 0.019 | 0.018 | 0.149 | 0.149 | 0.121 | 0.124 | 0.090 | 0.149 | 0.002 | 0.025 | 0.022 | 0.017 | 0.015 | 0.009 | 0.023 | 1.000 | 0.990 | 0.753 | 0.987 | 0.736 | 0.542 | 0.523 |  |  |  |
|  |  | 0.2 | 0.8 | -0.6 | -0.5 | -0.3 | 0.2 | -0.037 | -0.038 | 0.001 | 0.028 | 0.003 | -0.030 | 0.001 | 0.013 | 0.152 | 0.151 | 0.124 | 0.125 | 0.090 | 0.151 | 0.002 | 0.025 | 0.023 | 0.016 | 0.016 | 0.009 | 0.023 | 1.000 | 0.995 | 0.971 | 0.962 | 0.796 | 0.598 | 0.917 |  |  |  |
|  |  | 0.2 | -0.8 | 1 | 0.5 | 0.3 | 0.2 | 0.037 | -0.034 | 0.071 | -0.002 | 0.042 | 0.031 | 0.071 | 0.018 | 0.152 | 0.152 | 0.127 | 0.129 | 0.107 | 0.152 | 0.002 | 0.024 | 0.028 | 0.016 | 0.019 | 0.012 | 0.028 | 1.000 | 0.998 | 1.000 | 0.961 | 0.880 | 0.764 | 1.000 |  |  |  |
|  |  | -1.8 | -0.8 | -1 | -0.5 | -0.3 | -0.2 | -0.036 | -0.044 | 0.007 | -0.001 | 0.006 | 0.002 | 0.007 | 0.017 | 0.156 | 0.156 | 0.129 | 0.123 | 0.100 | 0.156 | 0.002 | 0.026 | 0.025 | 0.017 | 0.015 | 0.010 | 0.025 | 1.000 | 0.980 | 0.996 | 0.812 | 0.433 | 0.363 | 0.992 |  |  |  |
|  |  | 1 | 0 | 1 | 0.5 | 0.3 | 0.2 | 0.038 | -0.036 | 0.074 | 0.008 | 0.041 | 0.025 | 0.074 | 0.018 | 0.154 | 0.154 | 0.130 | 0.128 | 0.104 | 0.154 | 0.002 | 0.025 | 0.029 | 0.017 | 0.018 | 0.011 | 0.029 | 1.000 | 0.025 | 0.999 | 0.948 | 0.799 | 0.688 | 0.999 |  |  |  |
|  |  | 1.3 | 0.8 | 0.5 | 0 | 0.3 | 0.2 | 0.037 | -0.045 | 0.082 | -0.002 | 0.050 | 0.034 | 0.082 | 0.012 | 0.151 | 0.151 | 0.126 | 0.131 | 0.101 | 0.151 | 0.002 | 0.025 | 0.030 | 0.016 | 0.020 | 0.011 | 0.030 | 1.000 | 0.983 | 0.909 | 0.015 | 0.796 | 0.683 | 0.754 |  |  |  |
|  |  | 1.5 | 0.8 | 0.7 | 0.5 | 0 | 0.2 | 0.038 | -0.014 | 0.052 | -0.008 | 0.026 | 0.034 | 0.052 | 0.020 | 0.109 | 0.109 | 0.129 | 0.077 | 0.104 | 0.109 | 0.002 | 0.012 | 0.015 | 0.017 | 0.007 | 0.012 | 0.015 | 1.000 | 1.000 | 1.000 | 0.814 | 0.029 | 0.553 | 0.970 |  |  |  |
|  |  | 1 | 0.8 | 0.2 | 0 | 0 | 0.2 | 0.038 | -0.019 | 0.057 | 0.001 | 0.024 | 0.032 | 0.057 | 0.013 | 0.103 | 0.103 | 0.129 | 0.075 | 0.105 | 0.103 | 0.002 | 0.011 | 0.014 | 0.017 | 0.006 | 0.012 | 0.014 | 1.000 | 1.000 | 0.642 | 0.045 | 0.049 | 0.756 | 0.176 |  |  |  |
|  |  | 0 | -1 | 1 | 0.5 | 0.3 | 0.2 | 0.038 | -0.043 | 0.081 | 0.003 | 0.048 | 0.031 | 0.082 | 0.019 | 0.150 | 0.151 | 0.130 | 0.134 | 0.107 | 0.151 | 0.002 | 0.024 | 0.029 | 0.017 | 0.020 | 0.013 | 0.029 | 0.499 | 1.000 | 1.000 | 0.963 | 0.882 | 0.758 | 1.000 |  |  |  |
|  | Egger | 1.8 | 0.8 | 1 | 0.5 | 0.3 | 0.2 | 0.002 | -0.047 | 0.050 | -0.002 | 0.013 | 0.039 | 0.050 | 0.046 | 0.156 | 0.161 | 0.144 | 0.136 | 0.113 | 0.161 | 0.002 | 0.027 | 0.028 | 0.021 | 0.019 | 0.014 | 0.028 | 1.000 | 0.860 | 0.994 | 0.605 | 0.329 | 0.303 | 0.968 | 0.000 | 0.768 | 0.047 |
|  |  | 0.8 | 0.8 | 0 | -0.5 | 0.3 | 0.2 | 0.002 | -0.045 | 0.047 | -0.003 | 0.010 | 0.041 | 0.047 | 0.032 | 0.160 | 0.159 | 0.134 | 0.134 | 0.110 | 0.159 | 0.001 | 0.028 | 0.028 | 0.018 | 0.018 | 0.014 | 0.028 | 1.000 | 0.987 | 0.063 | 0.946 | 0.775 | 0.733 | 0.012 | 0.042 | 0.756 | 0.047 |
|  |  | 1.2 | 0.8 | 0.4 | 0.5 | -0.3 | 0.2 | 0.000 | -0.060 | 0.060 | 0.047 | 0.053 | -0.040 | 0.060 | 0.044 | 0.157 | 0.158 | 0.131 | 0.126 | 0.093 | 0.158 | 0.002 | 0.028 | 0.029 | 0.019 | 0.019 | 0.010 | 0.029 | 1.000 | 0.988 | 0.764 | 0.970 | 0.534 | 0.443 | 0.579 | 0.021 | 0.801 | 0.049 |
|  |  | 0.2 | 0.8 | -0.6 | -0.5 | -0.3 | 0.2 | 0.000 | -0.044 | 0.044 | 0.034 | 0.046 | -0.036 | 0.044 | 0.031 | 0.160 | 0.159 | 0.135 | 0.124 | 0.092 | 0.159 | 0.001 | 0.028 | 0.027 | 0.019 | 0.017 | 0.010 | 0.027 | 1.000 | 0.988 | 0.919 | 0.928 | 0.639 | 0.541 | 0.844 | 0.065 | 0.798 | 0.056 |
|  |  | 0.2 | -0.8 | 1 | 0.5 | 0.3 | 0.2 | 0.002 | -0.041 | 0.043 | -0.009 | 0.014 | 0.038 | 0.043 | 0.045 | 0.162 | 0.163 | 0.137 | 0.130 | 0.113 | 0.163 | 0.002 | 0.028 | 0.028 | 0.019 | 0.017 | 0.014 | 0.028 | 0.989 | 0.995 | 1.000 | 0.937 | 0.788 | 0.727 | 1.000 | 0.052 | 0.751 | 0.048 |
|  |  | -1.8 | -0.8 | -1 | -0.5 | -0.3 | -0.2 | -0.002 | -0.050 | 0.048 | -0.010 | 0.050 | 0.008 | 0.048 | 0.043 | 0.167 | 0.168 | 0.141 | 0.129 | 0.107 | 0.168 | 0.002 | 0.030 | 0.031 | 0.020 | 0.019 | 0.012 | 0.031 | 1.000 | 0.964 | 0.992 | 0.746 | 0.257 | 0.235 | 0.967 | 0.001 | 0.746 | 0.055 |
|  |  | 1 | 0 | 1 | 0.5 | 0.3 | 0.2 | 0.003 | -0.040 | 0.042 | 0.003 | 0.007 | 0.032 | 0.042 | 0.045 | 0.162 | 0.165 | 0.140 | 0.131 | 0.112 | 0.165 | 0.002 | 0.028 | 0.029 | 0.020 | 0.017 | 0.014 | 0.029 | 1.000 | 0.025 | 1.000 | 0.907 | 0.656 | 0.612 | 1.000 | 0.009 | 0.739 | 0.045 |
|  |  | 1.3 | 0.8 | 0.5 | 0 | 0.3 | 0.2 | 0.001 | -0.048 | 0.049 | -0.010 | 0.014 | 0.045 | 0.049 | 0.032 | 0.159 | 0.158 | 0.136 | 0.130 | 0.111 | 0.158 | 0.001 | 0.028 | 0.027 | 0.018 | 0.017 | 0.014 | 0.027 | 1.000 | 0.967 | 0.834 | 0.017 | 0.658 | 0.622 | 0.593 | 0.008 | 0.770 | 0.050 |
|  |  | 1.5 | 0.8 | 0.7 | 0.5 | 0 | 0.2 | 0.003 | -0.010 | 0.013 | -0.014 | -0.015 | 0.043 | 0.013 | 0.048 | 0.114 | 0.118 | 0.143 | 0.074 | 0.114 | 0.118 | 0.002 | 0.013 | 0.014 | 0.021 | 0.006 | 0.015 | 0.014 | 1.000 | 0.999 | 0.997 | 0.730 | 0.025 | 0.444 | 0.912 | 0.002 | 0.810 | 0.048 |
|  |  | 1 | 0.8 | 0.2 | 0 | 0 | 0.2 | 0.002 | -0.014 | 0.016 | -0.009 | -0.017 | 0.043 | 0.016 | 0.032 | 0.108 | 0.107 | 0.142 | 0.073 | 0.117 | 0.107 | 0.001 | 0.012 | 0.012 | 0.020 | 0.006 | 0.016 | 0.012 | 1.000 | 1.000 | 0.428 | 0.046 | 0.044 | 0.700 | 0.069 | 0.045 | 0.810 | 0.043 |
|  |  | 0 | -1 | 1 | 0.5 | 0.3 | 0.2 | 0.003 | -0.047 | 0.050 | -0.003 | 0.013 | 0.041 | 0.050 | 0.047 | 0.155 | 0.157 | 0.141 | 0.131 | 0.114 | 0.157 | 0.002 | 0.026 | 0.027 | 0.020 | 0.017 | 0.015 | 0.027 | 0.054 | 1.000 | 1.000 | 0.925 | 0.776 | 0.718 | 1.000 | 0.048 | 0.770 | 0.057 |
| Sc33 | IVW | 1.8 | 0.8 | 1 | 0.5 | 0.3 | 0.2 | 0.053 | -0.057 | 0.110 | 0.076 | -0.028 | 0.062 | 0.110 | 0.017 | 0.187 | 0.187 | 0.191 | 0.189 | 0.186 | 0.187 | 0.003 | 0.038 | 0.047 | 0.042 | 0.036 | 0.039 | 0.047 | 1.000 | 0.669 | 0.981 | 0.416 | 0.270 | 0.251 | 0.887 |  |  |  |
|  |  | 0.8 | 0.8 | 0 | -0.5 | 0.3 | 0.2 | -0.021 | -0.056 | 0.034 | 0.003 | -0.032 | 0.063 | 0.034 | 0.014 | 0.183 | 0.183 | 0.180 | 0.191 | 0.185 | 0.183 | 0.001 | 0.036 | 0.035 | 0.032 | 0.038 | 0.038 | 0.035 | 1.000 | 0.966 | 0.056 | 0.770 | 0.313 | 0.317 | 0.003 |  |  |  |
|  |  | 1.2 | 0.8 | 0.4 | 0.5 | -0.3 | 0.2 | 0.053 | -0.045 | 0.098 | 0.081 | 0.039 | -0.023 | 0.098 | 0.016 | 0.195 | 0.195 | 0.190 | 0.189 | 0.183 | 0.195 | 0.003 | 0.040 | 0.047 | 0.043 | 0.037 | 0.034 | 0.047 | 1.000 | 0.926 | 0.674 | 0.814 | 0.340 | 0.191 | 0.248 |  |  |  |
|  |  | 0.2 | 0.8 | -0.6 | -0.5 | -0.3 | 0.2 | -0.022 | -0.038 | 0.016 | -0.001 | 0.037 | -0.020 | 0.015 | 0.014 | 0.192 | 0.192 | 0.186 | 0.192 | 0.186 | 0.192 | 0.001 | 0.038 | 0.037 | 0.035 | 0.038 | 0.035 | 0.037 | 1.000 | 0.964 | 0.863 | 0.789 | 0.347 | 0.188 | 0.442 |  |  |  |
|  |  | 0.2 | -0.8 | 1 | 0.5 | 0.3 | 0.2 | 0.053 | -0.048 | 0.100 | 0.068 | -0.032 | 0.064 | 0.100 | 0.017 | 0.185 | 0.185 | 0.187 | 0.203 | 0.199 | 0.185 | 0.003 | 0.037 | 0.044 | 0.040 | 0.042 | 0.044 | 0.044 | 1.000 | 0.988 | 0.999 | 0.849 | 0.330 | 0.346 | 0.982 |  |  |  |
|  |  | -1.8 | -0.8 | -1 | -0.5 | -0.3 | -0.2 | -0.052 | -0.057 | 0.005 | 0.001 | 0.055 | -0.051 | 0.005 | 0.016 | 0.195 | 0.195 | 0.190 | 0.182 | 0.190 | 0.195 | 0.003 | 0.041 | 0.038 | 0.036 | 0.036 | 0.039 | 0.038 | 1.000 | 0.888 | 0.965 | 0.411 | 0.271 | 0.300 | 0.839 |  |  |  |
|  |  | 1 | 0 | 1 | 0.5 | 0.3 | 0.2 | 0.053 | -0.057 | 0.110 | 0.078 | -0.035 | 0.066 | 0.110 | 0.017 | 0.184 | 0.184 | 0.185 | 0.210 | 0.204 | 0.184 | 0.003 | 0.037 | 0.046 | 0.040 | 0.045 | 0.046 | 0.046 | 1.000 | 0.018 | 0.998 | 0.776 | 0.338 | 0.342 | 0.969 |  |  |  |
|  |  | 1.3 | 0.8 | 0.5 | 0 | 0.3 | 0.2 | 0.016 | -0.057 | 0.073 | 0.045 | -0.039 | 0.067 | 0.073 | 0.014 | 0.195 | 0.195 | 0.188 | 0.192 | 0.189 | 0.195 | 0.000 | 0.041 | 0.043 | 0.037 | 0.038 | 0.040 | 0.043 | 1.000 | 0.906 | 0.740 | 0.024 | 0.303 | 0.337 | 0.286 |  |  |  |
|  |  | 1.5 | 0.8 | 0.7 | 0.5 | 0 | 0.2 | 0.052 | -0.030 | 0.083 | 0.059 | -0.040 | 0.064 | 0.083 | 0.016 | 0.181 | 0.182 | 0.187 | 0.187 | 0.194 | 0.182 | 0.003 | 0.034 | 0.040 | 0.038 | 0.037 | 0.042 | 0.040 | 1.000 | 0.862 | 0.900 | 0.549 | 0.068 | 0.299 | 0.524 |  |  |  |
|  |  | 1 | 0.8 | 0.2 | 0 | 0 | 0.2 | 0.015 | -0.043 | 0.058 | 0.038 | -0.048 | 0.069 | 0.058 | 0.015 | 0.183 | 0.183 | 0.191 | 0.182 | 0.189 | 0.183 | 0.000 | 0.035 | 0.037 | 0.038 | 0.035 | 0.040 | 0.037 | 1.000 | 0.969 | 0.279 | 0.061 | 0.058 | 0.343 | 0.035 |  |  |  |
|  |  | 0 | -1 | 1 | 0.5 | 0.3 | 0.2 | 0.051 | -0.044 | 0.095 | 0.063 | -0.024 | 0.056 | 0.095 | 0.017 | 0.194 | 0.195 | 0.191 | 0.204 | 0.201 | 0.195 | 0.003 | 0.040 | 0.047 | 0.041 | 0.042 | 0.044 | 0.047 | 0.834 | 0.999 | 1.000 | 0.835 | 0.363 | 0.321 | 0.979 |  |  |  |
|  | Egger | 1.8 | 0.8 | 1 | 0.5 | 0.3 | 0.2 | 0.003 | -0.063 | 0.066 | 0.046 | -0.043 | 0.063 | 0.067 | 0.040 | 0.205 | 0.206 | 0.212 | 0.213 | 0.213 | 0.206 | 0.002 | 0.046 | 0.047 | 0.047 | 0.047 | 0.049 | 0.047 | 1.000 | 0.513 | 0.940 | 0.271 | 0.208 | 0.210 | 0.698 | 0.001 | 0.054 | 0.761 |
|  |  | 0.8 | 0.8 | 0 | -0.5 | 0.3 | 0.2 | -0.002 | -0.065 | 0.063 | 0.047 | -0.039 | 0.055 | 0.063 | 0.035 | 0.210 | 0.213 | 0.205 | 0.214 | 0.204 | 0.213 | 0.001 | 0.048 | 0.049 | 0.044 | 0.047 | 0.045 | 0.049 | 1.000 | 0.920 | 0.082 | 0.605 | 0.247 | 0.256 | 0.004 | 0.049 | 0.055 | 0.810 |
|  |  | 1.2 | 0.8 | 0.4 | 0.5 | -0.3 | 0.2 | 0.001 | -0.057 | 0.058 | 0.056 | 0.053 | -0.051 | 0.058 | 0.038 | 0.218 | 0.220 | 0.213 | 0.205 | 0.197 | 0.220 | 0.001 | 0.051 | 0.052 | 0.049 | 0.045 | 0.041 | 0.052 | 1.000 | 0.860 | 0.493 | 0.704 | 0.248 | 0.121 | 0.154 | 0.019 | 0.054 | 0.786 |
|  |  | 0.2 | 0.8 | -0.6 | -0.5 | -0.3 | 0.2 | 0.000 | -0.050 | 0.049 | 0.048 | 0.048 | -0.047 | 0.049 | 0.035 | 0.211 | 0.211 | 0.204 | 0.209 | 0.201 | 0.211 | 0.001 | 0.047 | 0.047 | 0.044 | 0.046 | 0.043 | 0.047 | 1.000 | 0.922 | 0.739 | 0.636 | 0.270 | 0.129 | 0.274 | 0.057 | 0.053 | 0.794 |
|  |  | 0.2 | -0.8 | 1 | 0.5 | 0.3 | 0.2 | 0.003 | -0.063 | 0.066 | 0.050 | -0.048 | 0.064 | 0.066 | 0.042 | 0.204 | 0.205 | 0.208 | 0.220 | 0.216 | 0.205 | 0.002 | 0.046 | 0.047 | 0.046 | 0.050 | 0.051 | 0.047 | 0.994 | 0.972 | 0.998 | 0.749 | 0.255 | 0.283 | 0.930 | 0.052 | 0.042 | 0.822 |
|  |  | -1.8 | -0.8 | -1 | -0.5 | -0.3 | -0.2 | -0.001 | -0.066 | 0.065 | 0.048 | 0.068 | -0.051 | 0.065 | 0.040 | 0.214 | 0.213 | 0.210 | 0.205 | 0.212 | 0.213 | 0.002 | 0.050 | 0.050 | 0.046 | 0.047 | 0.048 | 0.050 | 1.000 | 0.816 | 0.899 | 0.241 | 0.198 | 0.251 | 0.659 | 0.000 | 0.044 | 0.805 |
|  |  | 1 | 0 | 1 | 0.5 | 0.3 | 0.2 | 0.003 | -0.068 | 0.071 | 0.054 | -0.047 | 0.064 | 0.071 | 0.041 | 0.206 | 0.207 | 0.208 | 0.224 | 0.220 | 0.207 | 0.002 | 0.047 | 0.048 | 0.046 | 0.052 | 0.052 | 0.048 | 1.000 | 0.018 | 0.991 | 0.643 | 0.262 | 0.272 | 0.906 | 0.012 | 0.040 | 0.779 |
|  |  | 1.3 | 0.8 | 0.5 | 0 | 0.3 | 0.2 | 0.002 | -0.066 | 0.069 | 0.054 | -0.058 | 0.073 | 0.069 | 0.036 | 0.215 | 0.216 | 0.212 | 0.211 | 0.210 | 0.216 | 0.001 | 0.051 | 0.051 | 0.048 | 0.048 | 0.050 | 0.051 | 1.000 | 0.838 | 0.636 | 0.027 | 0.215 | 0.260 | 0.184 | 0.016 | 0.038 | 0.798 |
|  |  | 1.5 | 0.8 | 0.7 | 0.5 | 0 | 0.2 | 0.002 | -0.043 | 0.045 | 0.036 | -0.055 | 0.063 | 0.045 | 0.042 | 0.196 | 0.197 | 0.199 | 0.209 | 0.213 | 0.197 | 0.002 | 0.040 | 0.041 | 0.041 | 0.047 | 0.049 | 0.041 | 1.000 | 0.753 | 0.766 | 0.392 | 0.061 | 0.256 | 0.321 | 0.000 | 0.051 | 0.813 |
|  |  | 1 | 0.8 | 0.2 | 0 | 0 | 0.2 | 0.001 | -0.057 | 0.058 | 0.053 | -0.059 | 0.064 | 0.058 | 0.036 | 0.203 | 0.204 | 0.209 | 0.204 | 0.208 | 0.204 | 0.001 | 0.045 | 0.045 | 0.047 | 0.045 | 0.047 | 0.045 | 1.000 | 0.921 | 0.227 | 0.055 | 0.073 | 0.277 | 0.022 | 0.040 | 0.053 | 0.792 |
|  |  | 0 | -1 | 1 | 0.5 | 0.3 | 0.2 | 0.003 | -0.054 | 0.056 | 0.039 | -0.041 | 0.058 | 0.057 | 0.041 | 0.213 | 0.214 | 0.210 | 0.221 | 0.217 | 0.214 | 0.002 | 0.048 | 0.049 | 0.046 | 0.050 | 0.050 | 0.049 | 0.040 | 0.996 | 0.998 | 0.733 | 0.263 | 0.271 | 0.931 | 0.039 | 0.039 | 0.797 |

TE: total effect; DE: direct effect; IE: indirect effect; IE_d: indirect effect calculated by difference method; IE_p: indirect effect calculated by product method; IE1: *X*→*M_1_*→*Y*; IE2: *X*→*M_2_*→*Y*; IE3: *X*→*M_1_*→*M_2_*→*Y*.

**eTable 21** Simulation of PSE-Egger in two ordered mediators setting with different effects (case(b),sc34,sc35).

|  |  | true | | | | | | BIAS | | | | | | | SE | | | | | | | MSE | | | | | | | Power/Type I error | | | | | | | | | |
| --- | --- | --- | --- | --- | --- | --- | --- | --- | --- | --- | --- | --- | --- | --- | --- | --- | --- | --- | --- | --- | --- | --- | --- | --- | --- | --- | --- | --- | --- | --- | --- | --- | --- | --- | --- | --- | --- | --- |
|  |  | TE | DE | IE | IE1 | IE2 | IE3 | TE | DE | IE_d | IE1 | IE2 | IE3 | IE_p | TE | DE | IE_d | IE1 | IE2 | IE3 | IE_p | TE | DE | IE_d | IE1 | IE2 | IE3 | IE_p | TE | DE | IE_d | IE1 | IE2 | IE3 | IE_p | GY | GM1 | GM2 |
| Sc34 | IVW | 1.8 | 0.8 | 1 | 0.5 | 0.3 | 0.2 | 0.090 | -0.036 | 0.126 | 0.038 | -0.151 | 0.239 | 0.126 | 0.016 | 0.180 | 0.180 | 0.238 | 0.122 | 0.190 | 0.180 | 0.008 | 0.034 | 0.048 | 0.058 | 0.038 | 0.093 | 0.048 | 1.000 | 0.718 | 0.984 | 0.181 | 0.069 | 0.346 | 0.722 |  |  |  |
|  |  | 0.8 | 0.8 | 0 | -0.5 | 0.3 | 0.2 | 0.015 | -0.044 | 0.059 | -0.025 | -0.146 | 0.230 | 0.059 | 0.012 | 0.183 | 0.183 | 0.235 | 0.125 | 0.186 | 0.183 | 0.000 | 0.036 | 0.037 | 0.056 | 0.037 | 0.088 | 0.037 | 1.000 | 0.963 | 0.066 | 0.650 | 0.199 | 0.800 | 0.004 |  |  |  |
|  |  | 1.2 | 0.8 | 0.4 | 0.5 | -0.3 | 0.2 | 0.015 | -0.048 | 0.062 | 0.078 | 0.165 | -0.180 | 0.062 | 0.013 | 0.195 | 0.196 | 0.187 | 0.113 | 0.103 | 0.196 | 0.000 | 0.040 | 0.042 | 0.041 | 0.040 | 0.043 | 0.042 | 1.000 | 0.939 | 0.636 | 0.848 | 0.158 | 0.027 | 0.425 |  |  |  |
|  |  | 0.2 | 0.8 | -0.6 | -0.5 | -0.3 | 0.2 | -0.060 | -0.060 | 0.000 | 0.014 | 0.167 | -0.181 | 0.000 | 0.012 | 0.187 | 0.187 | 0.180 | 0.112 | 0.102 | 0.187 | 0.004 | 0.039 | 0.035 | 0.033 | 0.040 | 0.043 | 0.035 | 1.000 | 0.968 | 0.898 | 0.782 | 0.178 | 0.033 | 0.753 |  |  |  |
|  |  | 0.2 | -0.8 | 1 | 0.5 | 0.3 | 0.2 | 0.090 | -0.053 | 0.142 | 0.053 | -0.147 | 0.236 | 0.142 | 0.016 | 0.182 | 0.182 | 0.230 | 0.129 | 0.192 | 0.182 | 0.008 | 0.036 | 0.054 | 0.056 | 0.038 | 0.093 | 0.053 | 1.000 | 0.987 | 0.999 | 0.698 | 0.202 | 0.794 | 0.988 |  |  |  |
|  |  | -1.8 | -0.8 | -1 | -0.5 | -0.3 | -0.2 | -0.090 | -0.051 | -0.038 | -0.003 | 0.156 | -0.192 | -0.038 | 0.015 | 0.183 | 0.183 | 0.222 | 0.121 | 0.182 | 0.183 | 0.008 | 0.036 | 0.035 | 0.049 | 0.039 | 0.070 | 0.035 | 1.000 | 0.878 | 0.984 | 0.241 | 0.075 | 0.397 | 0.764 |  |  |  |
|  |  | 1 | 0 | 1 | 0.5 | 0.3 | 0.2 | 0.090 | -0.049 | 0.139 | 0.052 | -0.145 | 0.231 | 0.139 | 0.016 | 0.184 | 0.185 | 0.223 | 0.125 | 0.181 | 0.185 | 0.008 | 0.036 | 0.053 | 0.053 | 0.037 | 0.086 | 0.053 | 1.000 | 0.018 | 0.997 | 0.551 | 0.169 | 0.704 | 0.977 |  |  |  |
|  |  | 1.3 | 0.8 | 0.5 | 0 | 0.3 | 0.2 | 0.052 | -0.051 | 0.103 | 0.025 | -0.150 | 0.228 | 0.103 | 0.013 | 0.183 | 0.183 | 0.231 | 0.126 | 0.181 | 0.183 | 0.003 | 0.036 | 0.044 | 0.054 | 0.038 | 0.085 | 0.044 | 1.000 | 0.904 | 0.798 | 0.019 | 0.164 | 0.660 | 0.263 |  |  |  |
|  |  | 1.5 | 0.8 | 0.7 | 0.5 | 0 | 0.2 | 0.089 | -0.031 | 0.120 | 0.053 | -0.162 | 0.229 | 0.120 | 0.016 | 0.187 | 0.188 | 0.232 | 0.125 | 0.181 | 0.188 | 0.008 | 0.036 | 0.050 | 0.056 | 0.042 | 0.085 | 0.050 | 1.000 | 0.825 | 0.888 | 0.341 | 0.128 | 0.494 | 0.424 |  |  |  |
|  |  | 1 | 0.8 | 0.2 | 0 | 0 | 0.2 | 0.052 | -0.028 | 0.080 | 0.016 | -0.163 | 0.226 | 0.080 | 0.012 | 0.191 | 0.191 | 0.237 | 0.131 | 0.195 | 0.191 | 0.003 | 0.037 | 0.043 | 0.056 | 0.044 | 0.089 | 0.043 | 1.000 | 0.966 | 0.314 | 0.049 | 0.244 | 0.755 | 0.050 |  |  |  |
|  |  | 0 | -1 | 1 | 0.5 | 0.3 | 0.2 | 0.089 | -0.043 | 0.132 | 0.044 | -0.143 | 0.231 | 0.132 | 0.016 | 0.187 | 0.187 | 0.234 | 0.125 | 0.188 | 0.187 | 0.008 | 0.037 | 0.053 | 0.057 | 0.036 | 0.089 | 0.052 | 1.000 | 0.998 | 1.000 | 0.681 | 0.211 | 0.794 | 0.989 |  |  |  |
|  | Egger | 1.8 | 0.8 | 1 | 0.5 | 0.3 | 0.2 | 0.003 | -0.061 | 0.064 | 0.014 | -0.041 | 0.091 | 0.064 | 0.034 | 0.218 | 0.217 | 0.234 | 0.153 | 0.164 | 0.217 | 0.001 | 0.051 | 0.051 | 0.055 | 0.025 | 0.035 | 0.051 | 1.000 | 0.435 | 0.899 | 0.163 | 0.132 | 0.154 | 0.649 | 0.001 | 0.516 | 0.789 |
|  |  | 0.8 | 0.8 | 0 | -0.5 | 0.3 | 0.2 | 0.001 | -0.072 | 0.073 | 0.024 | -0.031 | 0.080 | 0.073 | 0.029 | 0.220 | 0.220 | 0.231 | 0.163 | 0.165 | 0.220 | 0.001 | 0.054 | 0.054 | 0.054 | 0.028 | 0.034 | 0.054 | 1.000 | 0.882 | 0.067 | 0.589 | 0.417 | 0.478 | 0.005 | 0.054 | 0.528 | 0.805 |
|  |  | 1.2 | 0.8 | 0.4 | 0.5 | -0.3 | 0.2 | 0.003 | -0.077 | 0.079 | 0.069 | 0.079 | -0.068 | 0.079 | 0.033 | 0.226 | 0.223 | 0.207 | 0.145 | 0.121 | 0.223 | 0.001 | 0.057 | 0.056 | 0.048 | 0.027 | 0.019 | 0.056 | 1.000 | 0.815 | 0.506 | 0.738 | 0.314 | 0.123 | 0.330 | 0.019 | 0.594 | 0.789 |
|  |  | 0.2 | 0.8 | -0.6 | -0.5 | -0.3 | 0.2 | -0.001 | -0.092 | 0.091 | 0.080 | 0.084 | -0.073 | 0.091 | 0.028 | 0.221 | 0.219 | 0.206 | 0.138 | 0.115 | 0.219 | 0.001 | 0.057 | 0.056 | 0.049 | 0.026 | 0.019 | 0.056 | 1.000 | 0.850 | 0.617 | 0.520 | 0.346 | 0.137 | 0.425 | 0.049 | 0.570 | 0.809 |
|  |  | 0.2 | -0.8 | 1 | 0.5 | 0.3 | 0.2 | 0.002 | -0.083 | 0.085 | 0.030 | -0.032 | 0.087 | 0.085 | 0.036 | 0.216 | 0.216 | 0.224 | 0.162 | 0.164 | 0.216 | 0.001 | 0.054 | 0.054 | 0.051 | 0.027 | 0.034 | 0.054 | 0.999 | 0.963 | 0.993 | 0.673 | 0.421 | 0.467 | 0.973 | 0.056 | 0.494 | 0.800 |
|  |  | -1.8 | -0.8 | -1 | -0.5 | -0.3 | -0.2 | 0.000 | -0.080 | 0.080 | 0.045 | 0.067 | -0.032 | 0.080 | 0.032 | 0.224 | 0.225 | 0.217 | 0.150 | 0.149 | 0.225 | 0.001 | 0.057 | 0.057 | 0.049 | 0.027 | 0.023 | 0.057 | 1.000 | 0.743 | 0.828 | 0.186 | 0.139 | 0.136 | 0.597 | 0.002 | 0.520 | 0.808 |
|  |  | 1 | 0 | 1 | 0.5 | 0.3 | 0.2 | 0.003 | -0.083 | 0.086 | 0.029 | -0.027 | 0.084 | 0.086 | 0.036 | 0.222 | 0.222 | 0.220 | 0.163 | 0.160 | 0.222 | 0.001 | 0.056 | 0.057 | 0.049 | 0.027 | 0.033 | 0.057 | 1.000 | 0.020 | 0.988 | 0.520 | 0.368 | 0.391 | 0.939 | 0.018 | 0.489 | 0.811 |
|  |  | 1.3 | 0.8 | 0.5 | 0 | 0.3 | 0.2 | 0.002 | -0.083 | 0.085 | 0.038 | -0.036 | 0.084 | 0.085 | 0.028 | 0.226 | 0.226 | 0.224 | 0.165 | 0.161 | 0.226 | 0.001 | 0.058 | 0.058 | 0.051 | 0.029 | 0.033 | 0.058 | 1.000 | 0.755 | 0.580 | 0.021 | 0.314 | 0.352 | 0.264 | 0.014 | 0.503 | 0.803 |
|  |  | 1.5 | 0.8 | 0.7 | 0.5 | 0 | 0.2 | 0.002 | -0.047 | 0.049 | 0.028 | -0.061 | 0.083 | 0.049 | 0.035 | 0.197 | 0.199 | 0.224 | 0.131 | 0.165 | 0.199 | 0.001 | 0.041 | 0.042 | 0.051 | 0.021 | 0.034 | 0.042 | 1.000 | 0.720 | 0.767 | 0.307 | 0.030 | 0.252 | 0.399 | 0.002 | 0.555 | 0.806 |
|  |  | 1 | 0.8 | 0.2 | 0 | 0 | 0.2 | 0.002 | -0.045 | 0.048 | 0.029 | -0.064 | 0.083 | 0.048 | 0.030 | 0.210 | 0.209 | 0.232 | 0.135 | 0.173 | 0.209 | 0.001 | 0.046 | 0.046 | 0.055 | 0.022 | 0.037 | 0.046 | 1.000 | 0.921 | 0.199 | 0.047 | 0.067 | 0.457 | 0.056 | 0.033 | 0.537 | 0.770 |
|  |  | 0 | -1 | 1 | 0.5 | 0.3 | 0.2 | 0.002 | -0.076 | 0.079 | 0.022 | -0.027 | 0.084 | 0.079 | 0.036 | 0.227 | 0.226 | 0.227 | 0.162 | 0.167 | 0.226 | 0.001 | 0.058 | 0.057 | 0.052 | 0.027 | 0.035 | 0.057 | 0.052 | 0.991 | 0.992 | 0.654 | 0.444 | 0.473 | 0.975 | 0.053 | 0.495 | 0.814 |
| Sc35 | IVW | 1.8 | 0.8 | 1 | 0.5 | 0.3 | 0.2 | 0.089 | -0.160 | 0.250 | 0.220 | -0.039 | 0.068 | 0.250 | 0.016 | 0.112 | 0.114 | 0.110 | 0.184 | 0.184 | 0.114 | 0.008 | 0.038 | 0.075 | 0.061 | 0.035 | 0.038 | 0.075 | 1.000 | 0.447 | 1.000 | 0.737 | 0.274 | 0.298 | 0.981 |  |  |  |
|  |  | 0.8 | 0.8 | 0 | -0.5 | 0.3 | 0.2 | 0.015 | -0.163 | 0.178 | 0.156 | -0.034 | 0.056 | 0.178 | 0.012 | 0.103 | 0.103 | 0.102 | 0.185 | 0.185 | 0.103 | 0.000 | 0.037 | 0.042 | 0.035 | 0.035 | 0.037 | 0.042 | 1.000 | 0.999 | 0.179 | 0.725 | 0.352 | 0.351 | 0.001 |  |  |  |
|  |  | 1.2 | 0.8 | 0.4 | 0.5 | -0.3 | 0.2 | 0.089 | -0.160 | 0.249 | 0.232 | 0.046 | -0.030 | 0.249 | 0.014 | 0.116 | 0.117 | 0.113 | 0.174 | 0.173 | 0.117 | 0.008 | 0.039 | 0.076 | 0.067 | 0.033 | 0.031 | 0.076 | 1.000 | 0.937 | 0.960 | 0.989 | 0.351 | 0.197 | 0.510 |  |  |  |
|  |  | 0.2 | 0.8 | -0.6 | -0.5 | -0.3 | 0.2 | 0.015 | -0.157 | 0.172 | 0.156 | 0.044 | -0.028 | 0.172 | 0.011 | 0.111 | 0.110 | 0.107 | 0.174 | 0.172 | 0.110 | 0.000 | 0.037 | 0.042 | 0.036 | 0.032 | 0.030 | 0.042 | 1.000 | 1.000 | 0.960 | 0.861 | 0.341 | 0.191 | 0.221 |  |  |  |
|  |  | 0.2 | -0.8 | 1 | 0.5 | 0.3 | 0.2 | 0.089 | -0.157 | 0.246 | 0.220 | -0.045 | 0.070 | 0.246 | 0.016 | 0.109 | 0.110 | 0.106 | 0.182 | 0.181 | 0.110 | 0.008 | 0.036 | 0.073 | 0.060 | 0.035 | 0.038 | 0.072 | 1.000 | 1.000 | 1.000 | 1.000 | 0.318 | 0.343 | 1.000 |  |  |  |
|  |  | -1.8 | -0.8 | -1 | -0.5 | -0.3 | -0.2 | -0.014 | -0.155 | 0.141 | 0.149 | 0.037 | -0.045 | 0.141 | 0.013 | 0.110 | 0.110 | 0.109 | 0.179 | 0.177 | 0.110 | 0.000 | 0.036 | 0.032 | 0.034 | 0.033 | 0.033 | 0.032 | 1.000 | 0.986 | 0.981 | 0.077 | 0.345 | 0.298 | 0.770 |  |  |  |
|  |  | 1 | 0 | 1 | 0.5 | 0.3 | 0.2 | 0.089 | -0.163 | 0.252 | 0.223 | -0.032 | 0.060 | 0.252 | 0.015 | 0.109 | 0.109 | 0.108 | 0.180 | 0.179 | 0.109 | 0.008 | 0.038 | 0.075 | 0.062 | 0.034 | 0.036 | 0.075 | 1.000 | 0.007 | 1.000 | 0.976 | 0.331 | 0.333 | 1.000 |  |  |  |
|  |  | 1.3 | 0.8 | 0.5 | 0 | 0.3 | 0.2 | 0.052 | -0.166 | 0.218 | 0.189 | -0.030 | 0.058 | 0.218 | 0.013 | 0.106 | 0.106 | 0.104 | 0.181 | 0.179 | 0.106 | 0.003 | 0.039 | 0.059 | 0.047 | 0.034 | 0.035 | 0.059 | 1.000 | 0.899 | 0.974 | 0.017 | 0.349 | 0.327 | 0.614 |  |  |  |
|  |  | 1.5 | 0.8 | 0.7 | 0.5 | 0 | 0.2 | 0.089 | -0.150 | 0.239 | 0.221 | -0.050 | 0.067 | 0.239 | 0.016 | 0.106 | 0.108 | 0.111 | 0.185 | 0.187 | 0.108 | 0.008 | 0.034 | 0.069 | 0.061 | 0.037 | 0.040 | 0.069 | 1.000 | 0.766 | 0.990 | 0.869 | 0.068 | 0.333 | 0.830 |  |  |  |
|  |  | 1 | 0.8 | 0.2 | 0 | 0 | 0.2 | 0.052 | -0.150 | 0.203 | 0.183 | -0.036 | 0.055 | 0.203 | 0.013 | 0.110 | 0.111 | 0.116 | 0.182 | 0.184 | 0.111 | 0.003 | 0.035 | 0.053 | 0.047 | 0.034 | 0.037 | 0.053 | 1.000 | 0.978 | 0.631 | 0.065 | 0.075 | 0.322 | 0.064 |  |  |  |
|  |  | 0 | -1 | 1 | 0.5 | 0.3 | 0.2 | 0.090 | -0.157 | 0.247 | 0.221 | -0.030 | 0.056 | 0.247 | 0.016 | 0.112 | 0.113 | 0.111 | 0.182 | 0.183 | 0.113 | 0.008 | 0.037 | 0.074 | 0.061 | 0.034 | 0.037 | 0.074 | 1.000 | 1.000 | 1.000 | 0.999 | 0.357 | 0.320 | 0.998 |  |  |  |
|  | Egger | 1.8 | 0.8 | 1 | 0.5 | 0.3 | 0.2 | 0.001 | -0.060 | 0.061 | 0.045 | -0.046 | 0.063 | 0.062 | 0.037 | 0.118 | 0.123 | 0.118 | 0.211 | 0.211 | 0.123 | 0.001 | 0.018 | 0.019 | 0.016 | 0.047 | 0.049 | 0.019 | 1.000 | 0.512 | 0.974 | 0.186 | 0.219 | 0.250 | 0.747 | 0.000 | 0.062 | 0.806 |
|  |  | 0.8 | 0.8 | 0 | -0.5 | 0.3 | 0.2 | 0.001 | -0.066 | 0.068 | 0.054 | -0.048 | 0.061 | 0.068 | 0.029 | 0.109 | 0.110 | 0.111 | 0.210 | 0.210 | 0.110 | 0.001 | 0.016 | 0.017 | 0.015 | 0.046 | 0.048 | 0.017 | 1.000 | 0.998 | 0.022 | 0.878 | 0.271 | 0.298 | 0.000 | 0.238 | 0.057 | 0.818 |
|  |  | 1.2 | 0.8 | 0.4 | 0.5 | -0.3 | 0.2 | 0.001 | -0.065 | 0.066 | 0.063 | 0.062 | -0.059 | 0.066 | 0.033 | 0.120 | 0.122 | 0.116 | 0.193 | 0.190 | 0.122 | 0.001 | 0.019 | 0.019 | 0.018 | 0.041 | 0.040 | 0.019 | 1.000 | 0.950 | 0.545 | 0.816 | 0.249 | 0.131 | 0.053 | 0.019 | 0.053 | 0.793 |
|  |  | 0.2 | 0.8 | -0.6 | -0.5 | -0.3 | 0.2 | 0.001 | -0.059 | 0.059 | 0.057 | 0.058 | -0.056 | 0.059 | 0.029 | 0.116 | 0.116 | 0.112 | 0.190 | 0.187 | 0.116 | 0.001 | 0.017 | 0.017 | 0.016 | 0.040 | 0.038 | 0.017 | 1.000 | 1.000 | 0.992 | 0.972 | 0.257 | 0.126 | 0.416 | 0.556 | 0.046 | 0.835 |
|  |  | 0.2 | -0.8 | 1 | 0.5 | 0.3 | 0.2 | 0.002 | -0.061 | 0.063 | 0.048 | -0.058 | 0.073 | 0.063 | 0.036 | 0.115 | 0.118 | 0.113 | 0.206 | 0.207 | 0.118 | 0.001 | 0.017 | 0.018 | 0.015 | 0.046 | 0.048 | 0.018 | 0.999 | 1.000 | 1.000 | 0.970 | 0.250 | 0.303 | 0.991 | 0.221 | 0.055 | 0.812 |
|  |  | -1.8 | -0.8 | -1 | -0.5 | -0.3 | -0.2 | -0.001 | -0.059 | 0.058 | 0.048 | 0.052 | -0.042 | 0.058 | 0.033 | 0.112 | 0.114 | 0.111 | 0.197 | 0.195 | 0.114 | 0.001 | 0.016 | 0.016 | 0.015 | 0.042 | 0.040 | 0.016 | 1.000 | 0.927 | 0.981 | 0.150 | 0.254 | 0.240 | 0.755 | 0.000 | 0.066 | 0.795 |
|  |  | 1 | 0 | 1 | 0.5 | 0.3 | 0.2 | 0.001 | -0.063 | 0.064 | 0.047 | -0.042 | 0.059 | 0.064 | 0.036 | 0.117 | 0.120 | 0.117 | 0.202 | 0.200 | 0.120 | 0.001 | 0.018 | 0.018 | 0.016 | 0.043 | 0.043 | 0.018 | 1.000 | 0.000 | 0.998 | 0.703 | 0.255 | 0.269 | 0.960 | 0.014 | 0.049 | 0.801 |
|  |  | 1.3 | 0.8 | 0.5 | 0 | 0.3 | 0.2 | 0.001 | -0.069 | 0.070 | 0.053 | -0.039 | 0.056 | 0.070 | 0.030 | 0.113 | 0.116 | 0.110 | 0.202 | 0.200 | 0.116 | 0.001 | 0.017 | 0.018 | 0.015 | 0.042 | 0.043 | 0.018 | 1.000 | 0.921 | 0.712 | 0.000 | 0.269 | 0.262 | 0.090 | 0.007 | 0.034 | 0.831 |
|  |  | 1.5 | 0.8 | 0.7 | 0.5 | 0 | 0.2 | 0.002 | -0.048 | 0.051 | 0.046 | -0.061 | 0.065 | 0.051 | 0.036 | 0.110 | 0.115 | 0.115 | 0.205 | 0.205 | 0.115 | 0.001 | 0.015 | 0.016 | 0.015 | 0.046 | 0.046 | 0.016 | 1.000 | 0.801 | 0.847 | 0.321 | 0.067 | 0.257 | 0.261 | 0.000 | 0.056 | 0.802 |
|  |  | 1 | 0.8 | 0.2 | 0 | 0 | 0.2 | 0.003 | -0.051 | 0.053 | 0.046 | -0.052 | 0.060 | 0.053 | 0.030 | 0.114 | 0.115 | 0.118 | 0.209 | 0.210 | 0.115 | 0.001 | 0.016 | 0.016 | 0.016 | 0.046 | 0.048 | 0.016 | 1.000 | 0.986 | 0.100 | 0.000 | 0.085 | 0.287 | 0.000 | 0.052 | 0.051 | 0.774 |
|  |  | 0 | -1 | 1 | 0.5 | 0.3 | 0.2 | 0.001 | -0.057 | 0.058 | 0.044 | -0.039 | 0.054 | 0.058 | 0.037 | 0.118 | 0.122 | 0.115 | 0.202 | 0.200 | 0.122 | 0.001 | 0.017 | 0.018 | 0.015 | 0.042 | 0.043 | 0.018 | 0.054 | 1.000 | 1.000 | 0.976 | 0.259 | 0.258 | 0.990 | 0.302 | 0.067 | 0.816 |

TE: total effect; DE: direct effect; IE: indirect effect; IE_d: indirect effect calculated by difference method; IE_p: indirect effect calculated by product method; IE1: *X*→*M_1_*→*Y*; IE2: *X*→*M_2_*→*Y*; IE3: *X*→*M_1_*→*M_2_*→*Y*.

**eTable 22** Simulation of PSE-Egger in two ordered mediators setting with different effects (case(b),sc36,sc37).

|  |  | true | | | | | | BIAS | | | | | | | SE | | | | | | | MSE | | | | | | | Power/Type I error | | | | | | | | | |
| --- | --- | --- | --- | --- | --- | --- | --- | --- | --- | --- | --- | --- | --- | --- | --- | --- | --- | --- | --- | --- | --- | --- | --- | --- | --- | --- | --- | --- | --- | --- | --- | --- | --- | --- | --- | --- | --- | --- |
|  |  | TE | DE | IE | IE1 | IE2 | IE3 | TE | DE | IE_d | IE1 | IE2 | IE3 | IE_p | TE | DE | IE_d | IE1 | IE2 | IE3 | IE_p | TE | DE | IE_d | IE1 | IE2 | IE3 | IE_p | TE | DE | IE_d | IE1 | IE2 | IE3 | IE_p | GY | GM1 | GM2 |
| Sc36 | IVW | 1.8 | 0.8 | 1 | 0.5 | 0.3 | 0.2 | 0.075 | -0.095 | 0.170 | -0.059 | 0.139 | 0.090 | 0.170 | 0.017 | 0.096 | 0.097 | 0.075 | 0.115 | 0.102 | 0.097 | 0.006 | 0.018 | 0.038 | 0.009 | 0.032 | 0.019 | 0.038 | 1.000 | 0.915 | 1.000 | 0.580 | 0.819 | 0.689 | 1.000 |  |  |  |
|  |  | 0.8 | 0.8 | 0 | -0.5 | 0.3 | 0.2 | 0.075 | -0.088 | 0.163 | -0.068 | 0.135 | 0.095 | 0.163 | 0.011 | 0.091 | 0.091 | 0.077 | 0.110 | 0.104 | 0.092 | 0.006 | 0.016 | 0.035 | 0.011 | 0.030 | 0.020 | 0.035 | 1.000 | 1.000 | 0.233 | 0.999 | 0.989 | 0.875 | 0.006 |  |  |  |
|  |  | 1.2 | 0.8 | 0.4 | 0.5 | -0.3 | 0.2 | -0.001 | -0.097 | 0.096 | 0.084 | 0.092 | -0.081 | 0.096 | 0.016 | 0.087 | 0.089 | 0.073 | 0.075 | 0.056 | 0.089 | 0.000 | 0.017 | 0.017 | 0.013 | 0.014 | 0.010 | 0.017 | 1.000 | 0.997 | 0.950 | 1.000 | 0.408 | 0.279 | 0.809 |  |  |  |
|  |  | 0.2 | 0.8 | -0.6 | -0.5 | -0.3 | 0.2 | 0.000 | -0.091 | 0.091 | 0.079 | 0.091 | -0.080 | 0.090 | 0.009 | 0.090 | 0.090 | 0.075 | 0.076 | 0.057 | 0.090 | 0.000 | 0.016 | 0.016 | 0.012 | 0.014 | 0.010 | 0.016 | 1.000 | 0.999 | 0.998 | 0.998 | 0.892 | 0.698 | 0.993 |  |  |  |
|  |  | 0.2 | -0.8 | 1 | 0.5 | 0.3 | 0.2 | 0.075 | -0.087 | 0.162 | -0.065 | 0.133 | 0.095 | 0.162 | 0.017 | 0.091 | 0.092 | 0.076 | 0.112 | 0.104 | 0.092 | 0.006 | 0.016 | 0.035 | 0.010 | 0.030 | 0.020 | 0.035 | 1.000 | 1.000 | 1.000 | 0.994 | 0.992 | 0.877 | 1.000 |  |  |  |
|  |  | -1.8 | -0.8 | -1 | -0.5 | -0.3 | -0.2 | 0.001 | -0.094 | 0.095 | -0.061 | 0.094 | 0.061 | 0.094 | 0.016 | 0.088 | 0.089 | 0.078 | 0.079 | 0.059 | 0.089 | 0.000 | 0.017 | 0.017 | 0.010 | 0.015 | 0.007 | 0.017 | 1.000 | 0.999 | 1.000 | 0.972 | 0.067 | 0.046 | 0.998 |  |  |  |
|  |  | 1 | 0 | 1 | 0.5 | 0.3 | 0.2 | 0.075 | -0.093 | 0.168 | -0.055 | 0.132 | 0.091 | 0.168 | 0.018 | 0.090 | 0.091 | 0.075 | 0.113 | 0.102 | 0.091 | 0.006 | 0.017 | 0.036 | 0.009 | 0.030 | 0.019 | 0.036 | 1.000 | 0.000 | 1.000 | 0.948 | 0.975 | 0.864 | 1.000 |  |  |  |
|  |  | 1.3 | 0.8 | 0.5 | 0 | 0.3 | 0.2 | 0.075 | -0.090 | 0.165 | -0.063 | 0.135 | 0.093 | 0.165 | 0.010 | 0.091 | 0.091 | 0.078 | 0.113 | 0.101 | 0.092 | 0.006 | 0.016 | 0.036 | 0.010 | 0.031 | 0.019 | 0.036 | 1.000 | 0.994 | 0.990 | 0.000 | 0.963 | 0.859 | 0.939 |  |  |  |
|  |  | 1.5 | 0.8 | 0.7 | 0.5 | 0 | 0.2 | 0.074 | 0.015 | 0.059 | -0.061 | 0.029 | 0.091 | 0.059 | 0.017 | 0.065 | 0.066 | 0.077 | 0.097 | 0.102 | 0.066 | 0.006 | 0.004 | 0.008 | 0.010 | 0.010 | 0.019 | 0.008 | 1.000 | 1.000 | 1.000 | 0.755 | 0.063 | 0.777 | 0.988 |  |  |  |
|  |  | 1 | 0.8 | 0.2 | 0 | 0 | 0.2 | 0.074 | 0.017 | 0.058 | -0.066 | 0.027 | 0.097 | 0.058 | 0.011 | 0.065 | 0.066 | 0.076 | 0.096 | 0.104 | 0.066 | 0.006 | 0.005 | 0.008 | 0.010 | 0.010 | 0.020 | 0.008 | 1.000 | 1.000 | 0.726 | 0.000 | 0.076 | 0.879 | 0.052 |  |  |  |
|  |  | 0 | -1 | 1 | 0.5 | 0.3 | 0.2 | 0.075 | -0.095 | 0.171 | -0.064 | 0.138 | 0.097 | 0.171 | 0.017 | 0.087 | 0.090 | 0.080 | 0.112 | 0.103 | 0.090 | 0.006 | 0.017 | 0.037 | 0.011 | 0.032 | 0.020 | 0.037 | 0.989 | 1.000 | 1.000 | 0.995 | 0.997 | 0.897 | 1.000 |  |  |  |
|  | Egger | 1.8 | 0.8 | 1 | 0.5 | 0.3 | 0.2 | 0.002 | -0.049 | 0.051 | -0.004 | 0.018 | 0.038 | 0.052 | 0.041 | 0.092 | 0.097 | 0.079 | 0.095 | 0.081 | 0.097 | 0.002 | 0.011 | 0.012 | 0.006 | 0.009 | 0.008 | 0.012 | 1.000 | 0.916 | 0.999 | 0.649 | 0.225 | 0.187 | 0.995 | 0.000 | 0.753 | 0.046 |
|  |  | 0.8 | 0.8 | 0 | -0.5 | 0.3 | 0.2 | 0.000 | -0.044 | 0.044 | -0.015 | 0.014 | 0.046 | 0.044 | 0.023 | 0.086 | 0.087 | 0.081 | 0.091 | 0.084 | 0.087 | 0.001 | 0.009 | 0.010 | 0.007 | 0.008 | 0.009 | 0.010 | 1.000 | 1.000 | 0.011 | 0.998 | 0.959 | 0.902 | 0.000 | 0.196 | 0.769 | 0.038 |
|  |  | 1.2 | 0.8 | 0.4 | 0.5 | -0.3 | 0.2 | 0.001 | -0.052 | 0.053 | 0.042 | 0.046 | -0.036 | 0.053 | 0.037 | 0.081 | 0.087 | 0.072 | 0.079 | 0.065 | 0.087 | 0.001 | 0.009 | 0.010 | 0.007 | 0.008 | 0.005 | 0.010 | 1.000 | 0.998 | 0.880 | 0.998 | 0.613 | 0.503 | 0.596 | 0.048 | 0.783 | 0.037 |
|  |  | 0.2 | 0.8 | -0.6 | -0.5 | -0.3 | 0.2 | 0.000 | -0.043 | 0.043 | 0.032 | 0.045 | -0.035 | 0.042 | 0.022 | 0.088 | 0.089 | 0.077 | 0.082 | 0.067 | 0.089 | 0.000 | 0.010 | 0.010 | 0.007 | 0.009 | 0.006 | 0.010 | 1.000 | 1.000 | 0.998 | 0.998 | 0.970 | 0.860 | 0.998 | 0.526 | 0.810 | 0.044 |
|  |  | 0.2 | -0.8 | 1 | 0.5 | 0.3 | 0.2 | 0.003 | -0.043 | 0.046 | -0.010 | 0.013 | 0.043 | 0.046 | 0.040 | 0.088 | 0.093 | 0.080 | 0.092 | 0.083 | 0.093 | 0.002 | 0.010 | 0.011 | 0.007 | 0.009 | 0.009 | 0.011 | 0.995 | 1.000 | 1.000 | 0.998 | 0.963 | 0.912 | 1.000 | 0.223 | 0.748 | 0.043 |
|  |  | -1.8 | -0.8 | -1 | -0.5 | -0.3 | -0.2 | -0.001 | -0.049 | 0.049 | -0.008 | 0.049 | 0.007 | 0.049 | 0.039 | 0.086 | 0.095 | 0.079 | 0.082 | 0.072 | 0.095 | 0.002 | 0.010 | 0.011 | 0.006 | 0.009 | 0.005 | 0.011 | 1.000 | 0.996 | 0.999 | 0.854 | 0.139 | 0.125 | 0.993 | 0.001 | 0.737 | 0.050 |
|  |  | 1 | 0 | 1 | 0.5 | 0.3 | 0.2 | 0.000 | -0.044 | 0.044 | -0.007 | 0.016 | 0.035 | 0.044 | 0.041 | 0.088 | 0.094 | 0.078 | 0.092 | 0.082 | 0.094 | 0.002 | 0.010 | 0.011 | 0.006 | 0.009 | 0.008 | 0.011 | 1.000 | 0.000 | 1.000 | 0.970 | 0.761 | 0.685 | 1.000 | 0.019 | 0.730 | 0.049 |
|  |  | 1.3 | 0.8 | 0.5 | 0 | 0.3 | 0.2 | 0.001 | -0.044 | 0.046 | -0.008 | 0.012 | 0.042 | 0.046 | 0.023 | 0.085 | 0.086 | 0.079 | 0.090 | 0.084 | 0.086 | 0.001 | 0.009 | 0.009 | 0.006 | 0.008 | 0.009 | 0.009 | 1.000 | 0.997 | 0.928 | 0.000 | 0.715 | 0.671 | 0.652 | 0.006 | 0.757 | 0.046 |
|  |  | 1.5 | 0.8 | 0.7 | 0.5 | 0 | 0.2 | 0.000 | -0.012 | 0.012 | -0.008 | -0.017 | 0.037 | 0.012 | 0.041 | 0.061 | 0.070 | 0.078 | 0.069 | 0.081 | 0.070 | 0.002 | 0.004 | 0.005 | 0.006 | 0.005 | 0.008 | 0.005 | 1.000 | 1.000 | 1.000 | 0.814 | 0.008 | 0.362 | 0.943 | 0.001 | 0.803 | 0.043 |
|  |  | 1 | 0.8 | 0.2 | 0 | 0 | 0.2 | 0.001 | -0.012 | 0.013 | -0.009 | -0.020 | 0.042 | 0.013 | 0.023 | 0.061 | 0.063 | 0.076 | 0.067 | 0.082 | 0.063 | 0.001 | 0.004 | 0.004 | 0.006 | 0.005 | 0.009 | 0.004 | 1.000 | 1.000 | 0.382 | 0.000 | 0.047 | 0.833 | 0.021 | 0.049 | 0.808 | 0.052 |
|  |  | 0 | -1 | 1 | 0.5 | 0.3 | 0.2 | 0.000 | -0.046 | 0.046 | -0.012 | 0.018 | 0.041 | 0.046 | 0.039 | 0.088 | 0.098 | 0.081 | 0.095 | 0.084 | 0.098 | 0.002 | 0.010 | 0.012 | 0.007 | 0.009 | 0.009 | 0.012 | 0.045 | 1.000 | 1.000 | 0.998 | 0.969 | 0.916 | 1.000 | 0.303 | 0.760 | 0.046 |
| Sc37 | IVW | 1.8 | 0.8 | 1 | 0.5 | 0.3 | 0.2 | 0.053 | -0.102 | 0.155 | 0.243 | 0.023 | -0.111 | 0.155 | 0.012 | 0.133 | 0.134 | 0.111 | 0.121 | 0.099 | 0.134 | 0.003 | 0.028 | 0.042 | 0.071 | 0.015 | 0.022 | 0.042 | 1.000 | 0.364 | 0.988 | 0.842 | 0.122 | 0.009 | 0.972 |  |  |  |
|  |  | 0.8 | 0.8 | 0 | -0.5 | 0.3 | 0.2 | -0.022 | -0.105 | 0.083 | 0.165 | 0.027 | -0.110 | 0.083 | 0.008 | 0.127 | 0.127 | 0.108 | 0.119 | 0.095 | 0.127 | 0.001 | 0.027 | 0.023 | 0.039 | 0.015 | 0.021 | 0.023 | 1.000 | 0.989 | 0.026 | 0.738 | 0.872 | 0.137 | 0.001 |  |  |  |
|  |  | 1.2 | 0.8 | 0.4 | 0.5 | -0.3 | 0.2 | 0.127 | -0.110 | 0.237 | 0.191 | -0.118 | 0.164 | 0.237 | 0.014 | 0.127 | 0.128 | 0.122 | 0.144 | 0.140 | 0.128 | 0.016 | 0.028 | 0.072 | 0.051 | 0.035 | 0.047 | 0.073 | 1.000 | 0.897 | 0.838 | 0.939 | 0.851 | 0.769 | 0.382 |  |  |  |
|  |  | 0.2 | 0.8 | -0.6 | -0.5 | -0.3 | 0.2 | 0.052 | -0.100 | 0.152 | 0.106 | -0.128 | 0.174 | 0.152 | 0.009 | 0.127 | 0.127 | 0.121 | 0.147 | 0.143 | 0.127 | 0.003 | 0.026 | 0.039 | 0.026 | 0.038 | 0.051 | 0.039 | 1.000 | 0.997 | 0.930 | 0.888 | 0.901 | 0.816 | 0.422 |  |  |  |
|  |  | 0.2 | -0.8 | 1 | 0.5 | 0.3 | 0.2 | 0.053 | -0.102 | 0.155 | 0.238 | 0.029 | -0.112 | 0.155 | 0.012 | 0.127 | 0.127 | 0.106 | 0.123 | 0.097 | 0.127 | 0.003 | 0.027 | 0.040 | 0.068 | 0.016 | 0.022 | 0.040 | 1.000 | 1.000 | 1.000 | 1.000 | 0.868 | 0.122 | 1.000 |  |  |  |
|  |  | -1.8 | -0.8 | -1 | -0.5 | -0.3 | -0.2 | 0.022 | -0.110 | 0.131 | 0.172 | -0.117 | 0.076 | 0.131 | 0.010 | 0.123 | 0.124 | 0.101 | 0.143 | 0.124 | 0.124 | 0.001 | 0.027 | 0.033 | 0.040 | 0.034 | 0.021 | 0.033 | 1.000 | 0.927 | 0.946 | 0.045 | 0.636 | 0.086 | 0.855 |  |  |  |
|  |  | 1 | 0 | 1 | 0.5 | 0.3 | 0.2 | 0.052 | -0.105 | 0.157 | 0.239 | 0.031 | -0.113 | 0.157 | 0.013 | 0.130 | 0.130 | 0.108 | 0.127 | 0.102 | 0.130 | 0.003 | 0.028 | 0.041 | 0.069 | 0.017 | 0.023 | 0.041 | 1.000 | 0.001 | 0.998 | 0.978 | 0.591 | 0.078 | 0.997 |  |  |  |
|  |  | 1.3 | 0.8 | 0.5 | 0 | 0.3 | 0.2 | 0.015 | -0.104 | 0.119 | 0.203 | 0.026 | -0.110 | 0.119 | 0.008 | 0.131 | 0.132 | 0.108 | 0.127 | 0.100 | 0.132 | 0.000 | 0.028 | 0.031 | 0.053 | 0.017 | 0.022 | 0.031 | 1.000 | 0.854 | 0.736 | 0.032 | 0.552 | 0.067 | 0.565 |  |  |  |
|  |  | 1.5 | 0.8 | 0.7 | 0.5 | 0 | 0.2 | 0.052 | -0.138 | 0.190 | 0.236 | 0.067 | -0.114 | 0.190 | 0.012 | 0.106 | 0.107 | 0.106 | 0.095 | 0.097 | 0.107 | 0.003 | 0.030 | 0.048 | 0.067 | 0.014 | 0.022 | 0.048 | 1.000 | 0.812 | 0.983 | 0.927 | 0.021 | 0.018 | 0.948 |  |  |  |
|  |  | 1 | 0.8 | 0.2 | 0 | 0 | 0.2 | 0.015 | -0.142 | 0.157 | 0.203 | 0.068 | -0.115 | 0.157 | 0.009 | 0.106 | 0.106 | 0.107 | 0.098 | 0.098 | 0.106 | 0.000 | 0.031 | 0.036 | 0.053 | 0.014 | 0.023 | 0.036 | 1.000 | 0.987 | 0.517 | 0.099 | 0.072 | 0.095 | 0.198 |  |  |  |
|  |  | 0 | -1 | 1 | 0.5 | 0.3 | 0.2 | 0.052 | -0.109 | 0.161 | 0.243 | 0.026 | -0.108 | 0.161 | 0.012 | 0.127 | 0.127 | 0.104 | 0.123 | 0.095 | 0.127 | 0.003 | 0.028 | 0.042 | 0.070 | 0.016 | 0.021 | 0.042 | 0.992 | 1.000 | 1.000 | 1.000 | 0.860 | 0.136 | 1.000 |  |  |  |
|  | Egger | 1.8 | 0.8 | 1 | 0.5 | 0.3 | 0.2 | 0.005 | -0.080 | 0.084 | 0.032 | -0.027 | 0.079 | 0.084 | 0.028 | 0.127 | 0.128 | 0.127 | 0.135 | 0.141 | 0.128 | 0.001 | 0.022 | 0.023 | 0.017 | 0.019 | 0.026 | 0.023 | 1.000 | 0.387 | 0.968 | 0.108 | 0.084 | 0.100 | 0.797 | 0.000 | 0.765 | 0.809 |
|  |  | 0.8 | 0.8 | 0 | -0.5 | 0.3 | 0.2 | 0.001 | -0.082 | 0.083 | 0.029 | -0.022 | 0.076 | 0.083 | 0.020 | 0.121 | 0.121 | 0.125 | 0.130 | 0.131 | 0.121 | 0.000 | 0.021 | 0.022 | 0.016 | 0.017 | 0.023 | 0.022 | 1.000 | 0.992 | 0.018 | 0.852 | 0.616 | 0.615 | 0.000 | 0.235 | 0.743 | 0.787 |
|  |  | 1.2 | 0.8 | 0.4 | 0.5 | -0.3 | 0.2 | 0.002 | -0.085 | 0.087 | 0.077 | 0.082 | -0.072 | 0.087 | 0.026 | 0.119 | 0.120 | 0.111 | 0.111 | 0.103 | 0.120 | 0.001 | 0.021 | 0.022 | 0.018 | 0.019 | 0.016 | 0.022 | 1.000 | 0.904 | 0.494 | 0.821 | 0.268 | 0.105 | 0.219 | 0.017 | 0.772 | 0.798 |
|  |  | 0.2 | 0.8 | -0.6 | -0.5 | -0.3 | 0.2 | -0.001 | -0.076 | 0.075 | 0.065 | 0.073 | -0.063 | 0.075 | 0.020 | 0.123 | 0.123 | 0.114 | 0.110 | 0.100 | 0.123 | 0.000 | 0.021 | 0.021 | 0.017 | 0.018 | 0.014 | 0.021 | 1.000 | 0.999 | 0.987 | 0.965 | 0.640 | 0.286 | 0.898 | 0.548 | 0.764 | 0.795 |
|  |  | 0.2 | -0.8 | 1 | 0.5 | 0.3 | 0.2 | 0.003 | -0.078 | 0.080 | 0.026 | -0.024 | 0.079 | 0.081 | 0.030 | 0.121 | 0.124 | 0.122 | 0.132 | 0.131 | 0.124 | 0.001 | 0.021 | 0.022 | 0.016 | 0.018 | 0.023 | 0.022 | 1.000 | 0.998 | 1.000 | 0.917 | 0.590 | 0.626 | 1.000 | 0.232 | 0.748 | 0.799 |
|  |  | -1.8 | -0.8 | -1 | -0.5 | -0.3 | -0.2 | 0.000 | -0.082 | 0.083 | 0.036 | 0.080 | -0.033 | 0.083 | 0.026 | 0.115 | 0.118 | 0.118 | 0.109 | 0.112 | 0.118 | 0.001 | 0.020 | 0.021 | 0.015 | 0.018 | 0.014 | 0.021 | 1.000 | 0.892 | 0.962 | 0.099 | 0.080 | 0.079 | 0.762 | 0.002 | 0.733 | 0.846 |
|  |  | 1 | 0 | 1 | 0.5 | 0.3 | 0.2 | 0.003 | -0.081 | 0.084 | 0.026 | -0.023 | 0.082 | 0.084 | 0.029 | 0.125 | 0.126 | 0.124 | 0.139 | 0.139 | 0.126 | 0.001 | 0.022 | 0.023 | 0.016 | 0.020 | 0.026 | 0.023 | 1.000 | 0.000 | 0.995 | 0.551 | 0.420 | 0.443 | 0.981 | 0.006 | 0.767 | 0.805 |
|  |  | 1.3 | 0.8 | 0.5 | 0 | 0.3 | 0.2 | 0.001 | -0.075 | 0.076 | 0.026 | -0.030 | 0.080 | 0.076 | 0.021 | 0.128 | 0.128 | 0.126 | 0.140 | 0.142 | 0.128 | 0.000 | 0.022 | 0.022 | 0.017 | 0.021 | 0.027 | 0.022 | 1.000 | 0.866 | 0.614 | 0.000 | 0.364 | 0.409 | 0.159 | 0.012 | 0.737 | 0.818 |
|  |  | 1.5 | 0.8 | 0.7 | 0.5 | 0 | 0.2 | 0.002 | -0.044 | 0.045 | 0.026 | -0.050 | 0.070 | 0.045 | 0.029 | 0.110 | 0.110 | 0.120 | 0.117 | 0.127 | 0.110 | 0.001 | 0.014 | 0.014 | 0.015 | 0.016 | 0.021 | 0.014 | 1.000 | 0.845 | 0.865 | 0.209 | 0.016 | 0.198 | 0.395 | 0.000 | 0.760 | 0.786 |
|  |  | 1 | 0.8 | 0.2 | 0 | 0 | 0.2 | 0.002 | -0.049 | 0.050 | 0.031 | -0.049 | 0.069 | 0.050 | 0.022 | 0.111 | 0.113 | 0.123 | 0.118 | 0.127 | 0.113 | 0.000 | 0.015 | 0.015 | 0.016 | 0.016 | 0.021 | 0.015 | 1.000 | 0.991 | 0.111 | 0.000 | 0.060 | 0.547 | 0.008 | 0.065 | 0.755 | 0.803 |
|  |  | 0 | -1 | 1 | 0.5 | 0.3 | 0.2 | 0.002 | -0.081 | 0.082 | 0.033 | -0.028 | 0.078 | 0.082 | 0.030 | 0.121 | 0.124 | 0.122 | 0.132 | 0.128 | 0.124 | 0.001 | 0.021 | 0.022 | 0.016 | 0.018 | 0.022 | 0.022 | 0.054 | 1.000 | 1.000 | 0.939 | 0.580 | 0.614 | 1.000 | 0.270 | 0.746 | 0.786 |

TE: total effect; DE: direct effect; IE: indirect effect; IE_d: indirect effect calculated by difference method; IE_p: indirect effect calculated by product method; IE1: *X*→*M_1_*→*Y*; IE2: *X*→*M_2_*→*Y*; IE3: *X*→*M_1_*→*M_2_*→*Y*.

**eTable 23** Simulation of PSE-Egger in two ordered mediators setting with different effects (case(b),sc38,sc39).

|  |  | true | | | | | | BIAS | | | | | | | SE | | | | | | | MSE | | | | | | | Power/Type I error | | | | | | | | | |
| --- | --- | --- | --- | --- | --- | --- | --- | --- | --- | --- | --- | --- | --- | --- | --- | --- | --- | --- | --- | --- | --- | --- | --- | --- | --- | --- | --- | --- | --- | --- | --- | --- | --- | --- | --- | --- | --- | --- |
|  |  | TE | DE | IE | IE1 | IE2 | IE3 | TE | DE | IE_d | IE1 | IE2 | IE3 | IE_p | TE | DE | IE_d | IE1 | IE2 | IE3 | IE_p | TE | DE | IE_d | IE1 | IE2 | IE3 | IE_p | TE | DE | IE_d | IE1 | IE2 | IE3 | IE_p | GY | GM1 | GM2 |
| Sc38 | IVW | 1.8 | 0.8 | 1 | 0.5 | 0.3 | 0.2 | 0.053 | 0.098 | -0.045 | -0.015 | -0.176 | 0.146 | -0.045 | 0.012 | 0.115 | 0.115 | 0.148 | 0.093 | 0.122 | 0.115 | 0.003 | 0.023 | 0.015 | 0.022 | 0.040 | 0.036 | 0.015 | 1.000 | 0.933 | 0.976 | 0.048 | 0.010 | 0.090 | 0.524 |  |  |  |
|  |  | 0.8 | 0.8 | 0 | -0.5 | 0.3 | 0.2 | -0.022 | 0.101 | -0.123 | -0.088 | -0.173 | 0.138 | -0.123 | 0.008 | 0.115 | 0.115 | 0.146 | 0.095 | 0.126 | 0.115 | 0.001 | 0.024 | 0.028 | 0.029 | 0.039 | 0.035 | 0.028 | 1.000 | 1.000 | 0.115 | 0.949 | 0.242 | 0.863 | 0.001 |  |  |  |
|  |  | 1.2 | 0.8 | 0.4 | 0.5 | -0.3 | 0.2 | -0.022 | 0.103 | -0.124 | -0.083 | 0.128 | -0.169 | -0.125 | 0.010 | 0.114 | 0.115 | 0.110 | 0.130 | 0.124 | 0.115 | 0.001 | 0.024 | 0.029 | 0.019 | 0.033 | 0.044 | 0.029 | 1.000 | 0.998 | 0.185 | 0.675 | 0.250 | 0.044 | 0.028 |  |  |  |
|  |  | 0.2 | 0.8 | -0.6 | -0.5 | -0.3 | 0.2 | -0.097 | 0.100 | -0.197 | -0.156 | 0.134 | -0.175 | -0.197 | 0.011 | 0.115 | 0.115 | 0.112 | 0.123 | 0.120 | 0.115 | 0.010 | 0.023 | 0.052 | 0.037 | 0.033 | 0.045 | 0.052 | 1.000 | 0.999 | 0.999 | 0.999 | 0.287 | 0.059 | 0.993 |  |  |  |
|  |  | 0.2 | -0.8 | 1 | 0.5 | 0.3 | 0.2 | 0.052 | 0.096 | -0.044 | -0.015 | -0.173 | 0.144 | -0.045 | 0.013 | 0.117 | 0.117 | 0.140 | 0.091 | 0.126 | 0.117 | 0.003 | 0.023 | 0.016 | 0.020 | 0.038 | 0.037 | 0.016 | 1.000 | 0.999 | 1.000 | 0.823 | 0.222 | 0.874 | 0.998 |  |  |  |
|  |  | -1.8 | -0.8 | -1 | -0.5 | -0.3 | -0.2 | -0.126 | 0.102 | -0.229 | -0.088 | 0.132 | -0.273 | -0.229 | 0.013 | 0.113 | 0.114 | 0.147 | 0.127 | 0.153 | 0.114 | 0.016 | 0.023 | 0.065 | 0.029 | 0.034 | 0.098 | 0.065 | 1.000 | 0.754 | 1.000 | 0.279 | 0.104 | 0.573 | 0.946 |  |  |  |
|  |  | 1 | 0 | 1 | 0.5 | 0.3 | 0.2 | 0.053 | 0.096 | -0.043 | -0.010 | -0.176 | 0.142 | -0.043 | 0.012 | 0.113 | 0.112 | 0.138 | 0.094 | 0.126 | 0.112 | 0.003 | 0.022 | 0.014 | 0.019 | 0.040 | 0.036 | 0.014 | 1.000 | 0.000 | 0.999 | 0.427 | 0.118 | 0.544 | 0.966 |  |  |  |
|  |  | 1.3 | 0.8 | 0.5 | 0 | 0.3 | 0.2 | 0.015 | 0.097 | -0.082 | -0.050 | -0.180 | 0.147 | -0.082 | 0.008 | 0.115 | 0.115 | 0.145 | 0.095 | 0.128 | 0.115 | 0.000 | 0.023 | 0.020 | 0.023 | 0.041 | 0.038 | 0.020 | 1.000 | 0.995 | 0.453 | 0.000 | 0.092 | 0.505 | 0.017 |  |  |  |
|  |  | 1.5 | 0.8 | 0.7 | 0.5 | 0 | 0.2 | 0.053 | 0.050 | 0.003 | -0.011 | -0.133 | 0.147 | 0.003 | 0.013 | 0.120 | 0.119 | 0.149 | 0.100 | 0.135 | 0.119 | 0.003 | 0.017 | 0.014 | 0.022 | 0.028 | 0.040 | 0.014 | 1.000 | 0.957 | 0.872 | 0.140 | 0.050 | 0.230 | 0.203 |  |  |  |
|  |  | 1 | 0.8 | 0.2 | 0 | 0 | 0.2 | 0.015 | 0.053 | -0.038 | -0.052 | -0.134 | 0.149 | -0.038 | 0.009 | 0.116 | 0.116 | 0.143 | 0.096 | 0.129 | 0.116 | 0.000 | 0.016 | 0.015 | 0.023 | 0.027 | 0.039 | 0.015 | 1.000 | 0.997 | 0.038 | 0.004 | 0.222 | 0.702 | 0.000 |  |  |  |
|  |  | 0 | -1 | 1 | 0.5 | 0.3 | 0.2 | 0.053 | 0.100 | -0.047 | -0.016 | -0.172 | 0.141 | -0.047 | 0.012 | 0.116 | 0.116 | 0.149 | 0.094 | 0.129 | 0.116 | 0.003 | 0.023 | 0.016 | 0.023 | 0.038 | 0.037 | 0.016 | 0.984 | 1.000 | 1.000 | 0.836 | 0.247 | 0.881 | 0.999 |  |  |  |
|  | Egger | 1.8 | 0.8 | 1 | 0.5 | 0.3 | 0.2 | 0.003 | -0.074 | 0.078 | 0.026 | -0.029 | 0.081 | 0.078 | 0.029 | 0.125 | 0.127 | 0.127 | 0.132 | 0.128 | 0.127 | 0.001 | 0.021 | 0.022 | 0.017 | 0.018 | 0.023 | 0.022 | 1.000 | 0.379 | 0.962 | 0.076 | 0.100 | 0.083 | 0.764 | 0.000 | 0.519 | 0.802 |
|  |  | 0.8 | 0.8 | 0 | -0.5 | 0.3 | 0.2 | 0.001 | -0.072 | 0.073 | 0.025 | -0.025 | 0.072 | 0.072 | 0.021 | 0.123 | 0.123 | 0.122 | 0.132 | 0.131 | 0.123 | 0.000 | 0.020 | 0.020 | 0.016 | 0.018 | 0.022 | 0.020 | 1.000 | 0.995 | 0.017 | 0.875 | 0.590 | 0.592 | 0.000 | 0.444 | 0.536 | 0.807 |
|  |  | 1.2 | 0.8 | 0.4 | 0.5 | -0.3 | 0.2 | 0.002 | -0.068 | 0.070 | 0.062 | 0.075 | -0.067 | 0.070 | 0.025 | 0.123 | 0.126 | 0.116 | 0.118 | 0.108 | 0.126 | 0.001 | 0.020 | 0.021 | 0.017 | 0.019 | 0.016 | 0.021 | 1.000 | 0.957 | 0.532 | 0.844 | 0.367 | 0.141 | 0.242 | 0.148 | 0.564 | 0.789 |
|  |  | 0.2 | 0.8 | -0.6 | -0.5 | -0.3 | 0.2 | 0.000 | -0.070 | 0.070 | 0.060 | 0.076 | -0.066 | 0.070 | 0.019 | 0.123 | 0.123 | 0.115 | 0.114 | 0.105 | 0.123 | 0.000 | 0.020 | 0.020 | 0.017 | 0.019 | 0.015 | 0.020 | 1.000 | 0.999 | 0.982 | 0.962 | 0.637 | 0.285 | 0.885 | 0.712 | 0.583 | 0.807 |
|  |  | 0.2 | -0.8 | 1 | 0.5 | 0.3 | 0.2 | 0.002 | -0.078 | 0.080 | 0.026 | -0.024 | 0.078 | 0.080 | 0.030 | 0.127 | 0.129 | 0.121 | 0.129 | 0.132 | 0.129 | 0.001 | 0.022 | 0.023 | 0.015 | 0.017 | 0.023 | 0.023 | 1.000 | 0.998 | 1.000 | 0.919 | 0.591 | 0.585 | 1.000 | 0.431 | 0.540 | 0.780 |
|  |  | -1.8 | -0.8 | -1 | -0.5 | -0.3 | -0.2 | 0.000 | -0.074 | 0.074 | 0.028 | 0.077 | -0.031 | 0.074 | 0.027 | 0.121 | 0.123 | 0.127 | 0.110 | 0.113 | 0.123 | 0.001 | 0.020 | 0.021 | 0.017 | 0.018 | 0.014 | 0.021 | 1.000 | 0.806 | 0.924 | 0.073 | 0.037 | 0.049 | 0.623 | 0.002 | 0.527 | 0.806 |
|  |  | 1 | 0 | 1 | 0.5 | 0.3 | 0.2 | 0.002 | -0.074 | 0.076 | 0.031 | -0.032 | 0.078 | 0.077 | 0.028 | 0.123 | 0.125 | 0.121 | 0.136 | 0.134 | 0.125 | 0.001 | 0.021 | 0.022 | 0.016 | 0.019 | 0.024 | 0.022 | 1.000 | 0.000 | 1.000 | 0.581 | 0.399 | 0.422 | 0.989 | 0.042 | 0.519 | 0.809 |
|  |  | 1.3 | 0.8 | 0.5 | 0 | 0.3 | 0.2 | 0.001 | -0.072 | 0.073 | 0.025 | -0.035 | 0.083 | 0.073 | 0.021 | 0.124 | 0.125 | 0.122 | 0.132 | 0.133 | 0.125 | 0.000 | 0.021 | 0.021 | 0.016 | 0.019 | 0.025 | 0.021 | 1.000 | 0.879 | 0.646 | 0.000 | 0.338 | 0.384 | 0.145 | 0.049 | 0.517 | 0.794 |
|  |  | 1.5 | 0.8 | 0.7 | 0.5 | 0 | 0.2 | 0.003 | -0.048 | 0.051 | 0.031 | -0.060 | 0.080 | 0.051 | 0.030 | 0.112 | 0.116 | 0.126 | 0.129 | 0.139 | 0.116 | 0.001 | 0.015 | 0.016 | 0.017 | 0.020 | 0.026 | 0.016 | 1.000 | 0.812 | 0.860 | 0.216 | 0.025 | 0.207 | 0.379 | 0.004 | 0.572 | 0.792 |
|  |  | 1 | 0.8 | 0.2 | 0 | 0 | 0.2 | 0.000 | -0.043 | 0.043 | 0.023 | -0.061 | 0.082 | 0.043 | 0.021 | 0.110 | 0.110 | 0.120 | 0.122 | 0.135 | 0.110 | 0.000 | 0.014 | 0.014 | 0.015 | 0.019 | 0.025 | 0.014 | 1.000 | 0.987 | 0.096 | 0.000 | 0.073 | 0.583 | 0.005 | 0.148 | 0.597 | 0.804 |
|  |  | 0 | -1 | 1 | 0.5 | 0.3 | 0.2 | 0.004 | -0.075 | 0.079 | 0.027 | -0.025 | 0.077 | 0.080 | 0.029 | 0.123 | 0.125 | 0.124 | 0.135 | 0.134 | 0.125 | 0.001 | 0.021 | 0.022 | 0.016 | 0.019 | 0.024 | 0.022 | 0.052 | 1.000 | 1.000 | 0.927 | 0.577 | 0.598 | 1.000 | 0.515 | 0.522 | 0.804 |
| Sc39 | IVW | 1.8 | 0.8 | 1 | 0.5 | 0.3 | 0.2 | 0.022 | -0.012 | 0.034 | -0.157 | 0.279 | -0.088 | 0.034 | 0.012 | 0.125 | 0.126 | 0.092 | 0.157 | 0.133 | 0.126 | 0.001 | 0.016 | 0.017 | 0.033 | 0.103 | 0.026 | 0.017 | 1.000 | 0.644 | 0.965 | 0.070 | 0.773 | 0.082 | 0.925 |  |  |  |
|  |  | 0.8 | 0.8 | 0 | -0.5 | 0.3 | 0.2 | 0.097 | -0.011 | 0.107 | -0.085 | 0.268 | -0.076 | 0.107 | 0.011 | 0.124 | 0.124 | 0.092 | 0.156 | 0.129 | 0.124 | 0.009 | 0.016 | 0.027 | 0.016 | 0.096 | 0.022 | 0.027 | 1.000 | 0.998 | 0.035 | 0.998 | 0.983 | 0.186 | 0.000 |  |  |  |
|  |  | 1.2 | 0.8 | 0.4 | 0.5 | -0.3 | 0.2 | -0.052 | -0.012 | -0.040 | -0.038 | 0.010 | -0.012 | -0.040 | 0.011 | 0.121 | 0.121 | 0.104 | 0.105 | 0.085 | 0.121 | 0.003 | 0.015 | 0.016 | 0.012 | 0.011 | 0.007 | 0.016 | 1.000 | 0.983 | 0.311 | 0.805 | 0.486 | 0.352 | 0.105 |  |  |  |
|  |  | 0.2 | 0.8 | -0.6 | -0.5 | -0.3 | 0.2 | 0.023 | -0.011 | 0.034 | 0.035 | 0.007 | -0.009 | 0.034 | 0.008 | 0.127 | 0.127 | 0.109 | 0.109 | 0.087 | 0.127 | 0.001 | 0.016 | 0.017 | 0.013 | 0.012 | 0.008 | 0.017 | 1.000 | 1.000 | 0.984 | 0.979 | 0.900 | 0.749 | 0.945 |  |  |  |
|  |  | 0.2 | -0.8 | 1 | 0.5 | 0.3 | 0.2 | 0.022 | -0.011 | 0.033 | -0.157 | 0.267 | -0.077 | 0.033 | 0.012 | 0.126 | 0.127 | 0.094 | 0.146 | 0.124 | 0.127 | 0.001 | 0.016 | 0.017 | 0.033 | 0.093 | 0.021 | 0.017 | 1.000 | 0.997 | 1.000 | 0.867 | 0.987 | 0.164 | 1.000 |  |  |  |
|  |  | -1.8 | -0.8 | -1 | -0.5 | -0.3 | -0.2 | 0.052 | 0.002 | 0.050 | -0.090 | 0.003 | 0.136 | 0.050 | 0.011 | 0.130 | 0.131 | 0.095 | 0.111 | 0.066 | 0.130 | 0.003 | 0.017 | 0.020 | 0.017 | 0.012 | 0.023 | 0.020 | 1.000 | 0.840 | 0.975 | 0.864 | 0.117 | 0.001 | 0.955 |  |  |  |
|  |  | 1 | 0 | 1 | 0.5 | 0.3 | 0.2 | 0.022 | -0.012 | 0.034 | -0.163 | 0.275 | -0.078 | 0.034 | 0.012 | 0.125 | 0.126 | 0.093 | 0.152 | 0.125 | 0.126 | 0.001 | 0.016 | 0.017 | 0.035 | 0.099 | 0.022 | 0.017 | 1.000 | 0.000 | 1.000 | 0.473 | 0.977 | 0.154 | 0.999 |  |  |  |
|  |  | 1.3 | 0.8 | 0.5 | 0 | 0.3 | 0.2 | 0.059 | -0.013 | 0.072 | -0.120 | 0.268 | -0.075 | 0.072 | 0.009 | 0.125 | 0.124 | 0.093 | 0.151 | 0.129 | 0.124 | 0.004 | 0.016 | 0.021 | 0.023 | 0.094 | 0.022 | 0.021 | 1.000 | 0.948 | 0.677 | 0.003 | 0.960 | 0.166 | 0.337 |  |  |  |
|  |  | 1.5 | 0.8 | 0.7 | 0.5 | 0 | 0.2 | 0.023 | 0.089 | -0.066 | -0.162 | 0.172 | -0.076 | -0.066 | 0.012 | 0.094 | 0.094 | 0.093 | 0.131 | 0.130 | 0.094 | 0.001 | 0.017 | 0.013 | 0.035 | 0.047 | 0.023 | 0.013 | 1.000 | 0.997 | 0.927 | 0.179 | 0.241 | 0.131 | 0.624 |  |  |  |
|  |  | 1 | 0.8 | 0.2 | 0 | 0 | 0.2 | 0.060 | 0.088 | -0.027 | -0.121 | 0.177 | -0.084 | -0.027 | 0.010 | 0.094 | 0.094 | 0.091 | 0.131 | 0.129 | 0.094 | 0.004 | 0.017 | 0.010 | 0.023 | 0.049 | 0.024 | 0.010 | 1.000 | 1.000 | 0.061 | 0.023 | 0.310 | 0.169 | 0.001 |  |  |  |
|  |  | 0 | -1 | 1 | 0.5 | 0.3 | 0.2 | 0.023 | -0.015 | 0.038 | -0.158 | 0.272 | -0.076 | 0.038 | 0.012 | 0.131 | 0.132 | 0.096 | 0.149 | 0.125 | 0.132 | 0.001 | 0.017 | 0.019 | 0.034 | 0.096 | 0.021 | 0.019 | 0.468 | 1.000 | 1.000 | 0.886 | 0.989 | 0.174 | 1.000 |  |  |  |
|  | Egger | 1.8 | 0.8 | 1 | 0.5 | 0.3 | 0.2 | 0.001 | -0.077 | 0.078 | 0.032 | -0.022 | 0.068 | 0.078 | 0.029 | 0.124 | 0.126 | 0.126 | 0.136 | 0.135 | 0.126 | 0.001 | 0.021 | 0.022 | 0.017 | 0.019 | 0.023 | 0.022 | 1.000 | 0.381 | 0.961 | 0.114 | 0.107 | 0.094 | 0.770 | 0.000 | 0.712 | 0.833 |
|  |  | 0.8 | 0.8 | 0 | -0.5 | 0.3 | 0.2 | 0.002 | -0.078 | 0.080 | 0.028 | -0.025 | 0.076 | 0.080 | 0.021 | 0.121 | 0.122 | 0.125 | 0.131 | 0.131 | 0.122 | 0.000 | 0.021 | 0.021 | 0.016 | 0.018 | 0.023 | 0.021 | 1.000 | 0.993 | 0.010 | 0.800 | 0.590 | 0.592 | 0.000 | 0.166 | 0.683 | 0.809 |
|  |  | 1.2 | 0.8 | 0.4 | 0.5 | -0.3 | 0.2 | -0.001 | -0.078 | 0.077 | 0.068 | 0.077 | -0.068 | 0.077 | 0.026 | 0.116 | 0.117 | 0.109 | 0.113 | 0.106 | 0.117 | 0.001 | 0.020 | 0.020 | 0.017 | 0.019 | 0.016 | 0.020 | 1.000 | 0.943 | 0.582 | 0.874 | 0.355 | 0.161 | 0.270 | 0.037 | 0.751 | 0.828 |
|  |  | 0.2 | 0.8 | -0.6 | -0.5 | -0.3 | 0.2 | 0.000 | -0.077 | 0.077 | 0.066 | 0.072 | -0.062 | 0.077 | 0.019 | 0.122 | 0.123 | 0.116 | 0.112 | 0.103 | 0.123 | 0.000 | 0.021 | 0.021 | 0.018 | 0.018 | 0.014 | 0.021 | 1.000 | 1.000 | 0.976 | 0.952 | 0.650 | 0.323 | 0.872 | 0.508 | 0.729 | 0.838 |
|  |  | 0.2 | -0.8 | 1 | 0.5 | 0.3 | 0.2 | 0.002 | -0.077 | 0.079 | 0.029 | -0.030 | 0.080 | 0.079 | 0.029 | 0.123 | 0.126 | 0.122 | 0.129 | 0.134 | 0.126 | 0.001 | 0.021 | 0.022 | 0.016 | 0.018 | 0.024 | 0.022 | 1.000 | 1.000 | 1.000 | 0.920 | 0.583 | 0.619 | 1.000 | 0.205 | 0.722 | 0.824 |
|  |  | -1.8 | -0.8 | -1 | -0.5 | -0.3 | -0.2 | 0.001 | -0.063 | 0.064 | 0.021 | 0.074 | -0.032 | 0.064 | 0.026 | 0.125 | 0.128 | 0.124 | 0.114 | 0.116 | 0.128 | 0.001 | 0.020 | 0.020 | 0.016 | 0.019 | 0.015 | 0.020 | 1.000 | 0.865 | 0.961 | 0.141 | 0.082 | 0.080 | 0.783 | 0.000 | 0.696 | 0.803 |
|  |  | 1 | 0 | 1 | 0.5 | 0.3 | 0.2 | 0.000 | -0.076 | 0.076 | 0.025 | -0.027 | 0.078 | 0.076 | 0.029 | 0.121 | 0.124 | 0.124 | 0.133 | 0.132 | 0.124 | 0.001 | 0.020 | 0.021 | 0.016 | 0.018 | 0.024 | 0.021 | 1.000 | 0.000 | 1.000 | 0.568 | 0.427 | 0.455 | 0.992 | 0.018 | 0.719 | 0.822 |
|  |  | 1.3 | 0.8 | 0.5 | 0 | 0.3 | 0.2 | 0.000 | -0.078 | 0.078 | 0.031 | -0.027 | 0.074 | 0.078 | 0.022 | 0.118 | 0.119 | 0.118 | 0.128 | 0.125 | 0.119 | 0.000 | 0.020 | 0.020 | 0.015 | 0.017 | 0.021 | 0.020 | 1.000 | 0.866 | 0.624 | 0.000 | 0.358 | 0.391 | 0.144 | 0.006 | 0.710 | 0.806 |
|  |  | 1.5 | 0.8 | 0.7 | 0.5 | 0 | 0.2 | 0.001 | -0.043 | 0.044 | 0.023 | -0.057 | 0.078 | 0.044 | 0.030 | 0.107 | 0.110 | 0.121 | 0.120 | 0.131 | 0.110 | 0.001 | 0.013 | 0.014 | 0.015 | 0.018 | 0.023 | 0.014 | 1.000 | 0.856 | 0.868 | 0.248 | 0.014 | 0.232 | 0.408 | 0.000 | 0.767 | 0.819 |
|  |  | 1 | 0.8 | 0.2 | 0 | 0 | 0.2 | 0.002 | -0.052 | 0.054 | 0.035 | -0.053 | 0.072 | 0.054 | 0.022 | 0.109 | 0.109 | 0.121 | 0.122 | 0.133 | 0.109 | 0.000 | 0.015 | 0.015 | 0.016 | 0.018 | 0.023 | 0.015 | 1.000 | 0.992 | 0.104 | 0.000 | 0.062 | 0.516 | 0.000 | 0.040 | 0.727 | 0.848 |
|  |  | 0 | -1 | 1 | 0.5 | 0.3 | 0.2 | 0.003 | -0.083 | 0.086 | 0.030 | -0.021 | 0.077 | 0.086 | 0.029 | 0.127 | 0.131 | 0.127 | 0.138 | 0.129 | 0.131 | 0.001 | 0.023 | 0.025 | 0.017 | 0.020 | 0.023 | 0.025 | 0.046 | 0.999 | 1.000 | 0.945 | 0.608 | 0.602 | 0.999 | 0.267 | 0.712 | 0.843 |

TE: total effect; DE: direct effect; IE: indirect effect; IE_d: indirect effect calculated by difference method; IE_p: indirect effect calculated by product method; IE1: *X*→*M_1_*→*Y*; IE2: *X*→*M_2_*→*Y*; IE3: *X*→*M_1_*→*M_2_*→*Y*.

**eTable 24** Simulation of PSE-Egger in two ordered mediators setting with different effects (case(c),sc4).

|  |  | true | | | | | | BIAS | | | | | | | SE | | | | | | | MSE | | | | | | | Power/Type I error | | | | | | | | | |
| --- | --- | --- | --- | --- | --- | --- | --- | --- | --- | --- | --- | --- | --- | --- | --- | --- | --- | --- | --- | --- | --- | --- | --- | --- | --- | --- | --- | --- | --- | --- | --- | --- | --- | --- | --- | --- | --- | --- |
|  |  | TE | DE | IE | IE1 | IE2 | IE3 | TE | DE | IE_d | IE1 | IE2 | IE3 | IE_p | TE | DE | IE_d | IE1 | IE2 | IE3 | IE_p | TE | DE | IE_d | IE1 | IE2 | IE3 | IE_p | TE | DE | IE_d | IE1 | IE2 | IE3 | IE_p | GY | GM1 | GM2 |
| Sc4 | IVW | 1.8 | 0.8 | 1 | 0.5 | 0.3 | 0.2 | 0.418 | -0.371 | 0.789 | 0.296 | -0.503 | 0.995 | 0.789 | 0.044 | 0.058 | 0.073 | 0.160 | 0.090 | 0.175 | 0.073 | 0.176 | 0.141 | 0.627 | 0.113 | 0.261 | 1.021 | 0.628 | 1.000 | 0.715 | 1.000 | 0.139 | 0.505 | 0.854 | 0.918 |  |  |  |
|  |  | 0.8 | 0.8 | 0 | -0.5 | 0.3 | 0.2 | 0.173 | -0.371 | 0.544 | 0.052 | -0.505 | 0.998 | 0.545 | 0.019 | 0.058 | 0.060 | 0.153 | 0.087 | 0.170 | 0.060 | 0.030 | 0.141 | 0.300 | 0.026 | 0.263 | 1.025 | 0.300 | 1.000 | 1.000 | 1.000 | 0.483 | 0.662 | 1.000 | 0.095 |  |  |  |
|  |  | 1.2 | 0.8 | 0.4 | 0.5 | -0.3 | 0.2 | 0.172 | -0.373 | 0.545 | 0.538 | 0.374 | -0.366 | 0.546 | 0.020 | 0.056 | 0.059 | 0.076 | 0.040 | 0.063 | 0.059 | 0.030 | 0.142 | 0.301 | 0.295 | 0.142 | 0.138 | 0.301 | 1.000 | 0.991 | 1.000 | 1.000 | 0.076 | 0.162 | 1.000 |  |  |  |
|  |  | 0.2 | 0.8 | -0.6 | -0.5 | -0.3 | 0.2 | -0.074 | -0.371 | 0.297 | 0.289 | 0.371 | -0.363 | 0.297 | 0.010 | 0.056 | 0.056 | 0.075 | 0.039 | 0.063 | 0.056 | 0.006 | 0.141 | 0.092 | 0.089 | 0.139 | 0.135 | 0.091 | 1.000 | 1.000 | 0.998 | 0.783 | 0.503 | 0.800 | 0.885 |  |  |  |
|  |  | 0.2 | -0.8 | 1 | 0.5 | 0.3 | 0.2 | 0.421 | -0.372 | 0.793 | 0.303 | -0.501 | 0.989 | 0.791 | 0.043 | 0.057 | 0.072 | 0.158 | 0.090 | 0.179 | 0.072 | 0.179 | 0.141 | 0.633 | 0.117 | 0.259 | 1.009 | 0.631 | 1.000 | 1.000 | 1.000 | 0.963 | 0.635 | 1.000 | 1.000 |  |  |  |
|  |  | -1.8 | -0.8 | -1 | -0.5 | -0.3 | -0.2 | -0.173 | -0.372 | 0.199 | 0.054 | 0.373 | -0.228 | 0.199 | 0.020 | 0.057 | 0.059 | 0.150 | 0.039 | 0.146 | 0.059 | 0.030 | 0.142 | 0.043 | 0.026 | 0.141 | 0.073 | 0.043 | 1.000 | 1.000 | 1.000 | 0.006 | 0.004 | 0.008 | 0.004 |  |  |  |
|  |  | 1 | 0 | 1 | 0.5 | 0.3 | 0.2 | 0.420 | -0.372 | 0.792 | 0.299 | -0.505 | 0.998 | 0.792 | 0.044 | 0.058 | 0.073 | 0.155 | 0.089 | 0.178 | 0.073 | 0.179 | 0.142 | 0.633 | 0.114 | 0.263 | 1.028 | 0.632 | 1.000 | 0.905 | 1.000 | 0.685 | 0.641 | 0.991 | 0.998 |  |  |  |
|  |  | 1.3 | 0.8 | 0.5 | 0 | 0.3 | 0.2 | 0.296 | -0.373 | 0.669 | 0.180 | -0.503 | 0.993 | 0.669 | 0.032 | 0.058 | 0.065 | 0.149 | 0.087 | 0.166 | 0.065 | 0.089 | 0.142 | 0.452 | 0.055 | 0.261 | 1.013 | 0.452 | 1.000 | 0.973 | 1.000 | 0.000 | 0.635 | 0.986 | 0.772 |  |  |  |
|  |  | 1.5 | 0.8 | 0.7 | 0.5 | 0 | 0.2 | 0.420 | -0.231 | 0.651 | 0.302 | -0.643 | 0.992 | 0.651 | 0.043 | 0.096 | 0.107 | 0.158 | 0.114 | 0.168 | 0.107 | 0.178 | 0.062 | 0.435 | 0.116 | 0.427 | 1.013 | 0.435 | 1.000 | 0.498 | 1.000 | 0.332 | 0.932 | 0.935 | 0.546 |  |  |  |
|  |  | 1 | 0.8 | 0.2 | 0 | 0 | 0.2 | 0.295 | -0.228 | 0.523 | 0.170 | -0.647 | 1.000 | 0.523 | 0.030 | 0.094 | 0.099 | 0.157 | 0.117 | 0.175 | 0.099 | 0.088 | 0.061 | 0.283 | 0.054 | 0.432 | 1.031 | 0.283 | 1.000 | 0.943 | 0.994 | 0.005 | 0.999 | 0.997 | 0.089 |  |  |  |
|  |  | 0 | -1 | 1 | 0.5 | 0.3 | 0.2 | 0.421 | -0.373 | 0.794 | 0.303 | -0.502 | 0.991 | 0.792 | 0.044 | 0.058 | 0.073 | 0.154 | 0.089 | 0.177 | 0.073 | 0.179 | 0.142 | 0.636 | 0.116 | 0.259 | 1.013 | 0.633 | 1.000 | 1.000 | 1.000 | 0.987 | 0.661 | 1.000 | 1.000 |  |  |  |
|  | Egger | 1.8 | 0.8 | 1 | 0.5 | 0.3 | 0.2 | 0.067 | -0.364 | 0.432 | 0.159 | -0.478 | 0.751 | 0.432 | 0.107 | 0.061 | 0.120 | 0.142 | 0.092 | 0.161 | 0.120 | 0.016 | 0.136 | 0.201 | 0.045 | 0.237 | 0.590 | 0.201 | 1.000 | 0.641 | 1.000 | 0.117 | 0.337 | 0.804 | 0.839 | 0.000 | 0.090 | 0.928 |
|  |  | 0.8 | 0.8 | 0 | -0.5 | 0.3 | 0.2 | 0.031 | -0.366 | 0.397 | 0.122 | -0.482 | 0.758 | 0.397 | 0.046 | 0.061 | 0.072 | 0.135 | 0.092 | 0.164 | 0.072 | 0.003 | 0.138 | 0.163 | 0.033 | 0.241 | 0.601 | 0.163 | 1.000 | 1.000 | 0.994 | 0.461 | 0.489 | 1.000 | 0.009 | 0.006 | 0.090 | 0.912 |
|  |  | 1.2 | 0.8 | 0.4 | 0.5 | -0.3 | 0.2 | 0.028 | -0.367 | 0.395 | 0.356 | 0.367 | -0.328 | 0.395 | 0.048 | 0.058 | 0.072 | 0.083 | 0.041 | 0.055 | 0.072 | 0.003 | 0.138 | 0.161 | 0.133 | 0.136 | 0.110 | 0.161 | 1.000 | 0.988 | 1.000 | 1.000 | 0.041 | 0.135 | 1.000 | 0.001 | 0.117 | 0.942 |
|  |  | 0.2 | 0.8 | -0.6 | -0.5 | -0.3 | 0.2 | -0.011 | -0.366 | 0.355 | 0.315 | 0.365 | -0.325 | 0.355 | 0.026 | 0.059 | 0.062 | 0.069 | 0.039 | 0.052 | 0.062 | 0.001 | 0.137 | 0.130 | 0.104 | 0.134 | 0.108 | 0.130 | 1.000 | 1.000 | 0.964 | 0.756 | 0.366 | 0.781 | 0.756 | 0.064 | 0.097 | 0.934 |
|  |  | 0.2 | -0.8 | 1 | 0.5 | 0.3 | 0.2 | 0.074 | -0.366 | 0.440 | 0.168 | -0.478 | 0.751 | 0.441 | 0.103 | 0.060 | 0.115 | 0.142 | 0.093 | 0.168 | 0.115 | 0.016 | 0.138 | 0.207 | 0.048 | 0.237 | 0.592 | 0.207 | 0.722 | 1.000 | 1.000 | 0.961 | 0.498 | 1.000 | 1.000 | 0.003 | 0.090 | 0.920 |
|  |  | -1.8 | -0.8 | -1 | -0.5 | -0.3 | -0.2 | -0.030 | -0.366 | 0.336 | 0.123 | 0.367 | -0.154 | 0.335 | 0.048 | 0.060 | 0.072 | 0.132 | 0.040 | 0.126 | 0.072 | 0.003 | 0.137 | 0.118 | 0.032 | 0.136 | 0.040 | 0.118 | 1.000 | 1.000 | 1.000 | 0.007 | 0.001 | 0.008 | 0.007 | 0.000 | 0.107 | 0.917 |
|  |  | 1 | 0 | 1 | 0.5 | 0.3 | 0.2 | 0.069 | -0.366 | 0.436 | 0.163 | -0.479 | 0.752 | 0.436 | 0.104 | 0.062 | 0.121 | 0.143 | 0.092 | 0.162 | 0.121 | 0.016 | 0.138 | 0.204 | 0.047 | 0.238 | 0.592 | 0.204 | 1.000 | 0.853 | 1.000 | 0.620 | 0.467 | 0.986 | 0.994 | 0.000 | 0.094 | 0.919 |
|  |  | 1.3 | 0.8 | 0.5 | 0 | 0.3 | 0.2 | 0.048 | -0.366 | 0.414 | 0.145 | -0.478 | 0.747 | 0.414 | 0.075 | 0.060 | 0.092 | 0.128 | 0.090 | 0.155 | 0.092 | 0.008 | 0.138 | 0.180 | 0.037 | 0.236 | 0.582 | 0.180 | 1.000 | 0.966 | 1.000 | 0.000 | 0.466 | 0.984 | 0.572 | 0.000 | 0.096 | 0.914 |
|  |  | 1.5 | 0.8 | 0.7 | 0.5 | 0 | 0.2 | 0.070 | -0.228 | 0.298 | 0.165 | -0.612 | 0.746 | 0.298 | 0.099 | 0.098 | 0.139 | 0.141 | 0.117 | 0.158 | 0.139 | 0.015 | 0.062 | 0.108 | 0.047 | 0.389 | 0.581 | 0.108 | 1.000 | 0.467 | 0.993 | 0.286 | 0.907 | 0.910 | 0.195 | 0.000 | 0.110 | 0.923 |
|  |  | 1 | 0.8 | 0.2 | 0 | 0 | 0.2 | 0.047 | -0.227 | 0.274 | 0.139 | -0.616 | 0.751 | 0.274 | 0.070 | 0.097 | 0.118 | 0.136 | 0.118 | 0.160 | 0.118 | 0.007 | 0.061 | 0.089 | 0.038 | 0.394 | 0.590 | 0.089 | 1.000 | 0.927 | 0.703 | 0.002 | 0.996 | 0.996 | 0.008 | 0.001 | 0.121 | 0.927 |
|  |  | 0 | -1 | 1 | 0.5 | 0.3 | 0.2 | 0.065 | -0.366 | 0.431 | 0.164 | -0.478 | 0.745 | 0.431 | 0.104 | 0.061 | 0.120 | 0.139 | 0.090 | 0.156 | 0.120 | 0.015 | 0.138 | 0.200 | 0.046 | 0.237 | 0.579 | 0.200 | 0.096 | 1.000 | 1.000 | 0.986 | 0.497 | 1.000 | 1.000 | 0.009 | 0.083 | 0.932 |

TE: total effect; DE: direct effect; IE: indirect effect; IE_d: indirect effect calculated by difference method; IE_p: indirect effect calculated by product method; IE1: *X*→*M_1_*→*Y*; IE2: *X*→*M_2_*→*Y*; IE3: *X*→*M_1_*→*M_2_*→*Y*.

#### 7 Additional simulation scenario 1: Bidirectional causal effects between risk factor and mediator

There might be a case that the risk factor had a causal eﬀect on the mediator, as well as the mediator had a causal eﬀect on the risk factor. To consider this scenario, we simulated causal eﬀects in both directions and consider direct and indirect estimate. The data-generating models were:

**eTable 25** Data generation in the additional simulation scenario1.

| Causally ordered mediators PSE-IVW | Causally ordered mediators PSE-Egger |
| --- | --- |
| Causally non-ordered mediators PSE-IVW | Causally non-ordered mediators PSE-Egger |

This was the same as the previous data-generating model, except that we ﬁrstly generated and then generated a second risk factor variable that had a causal eﬀect from the mediator. These could be thought of as values of the risk factor at diﬀerent time points. We considered cases where the mediator had a positive and a negative eﬀect on the risk factor. For MR-Egger, we only considered the case where all the pathway had directional pleiotropy. All other aspects of this simulation were the same as the original.

The total effect varied depending on whether the effect of the mediator on the risk factor was positive or negative, and cannot be calculated as previous (as there were additional components of the total effect via the effect of the mediator on the risk factor). However, the direct effect as estimated by multivariable Mendelian randomization was invariant to any bidirectional effect. Results were listed in **eTable 26-29**.

**eTable 26**. Simulation results of PSE-IVW for two causally ordered mediators when there exist bidirectional causal effects between risk factor and mediators

|  | TRUE | | | | | | BIAS | | | | | | | SE | | | | | | | Power/Type I error | | | | | | |
| --- | --- | --- | --- | --- | --- | --- | --- | --- | --- | --- | --- | --- | --- | --- | --- | --- | --- | --- | --- | --- | --- | --- | --- | --- | --- | --- | --- |
|  | TE | DE | IE | IE1 | IE2 | IE3 | TE | DE | IE_d | IE1 | IE2 | IE3 | IE_p | TE | DE | IE_d | IE1 | IE2 | IE3 | IE_p | TE | DE | IE_d | IE1 | IE2 | IE3 | IE_p |
| Negative | 1.8 | 0.8 | 1 | 0.5 | 0.3 | 0.2 | 0.424 | -0.049 | 0.473 | 0.296 | -0.418 | 0.595 | 0.473 | 0.249 | 0.046 | 0.250 | 0.140 | 0.141 | 0.177 | 0.250 | 1.000 | 1.000 | 1.000 | 1.000 | 0.165 | 1.000 | 1.000 |
|  | 0.8 | 0.8 | 0 | -0.5 | 0.3 | 0.2 | -0.111 | -0.045 | -0.066 | -0.236 | -0.415 | 0.585 | -0.066 | 0.087 | 0.046 | 0.092 | 0.136 | 0.140 | 0.162 | 0.092 | 1.000 | 1.000 | 0.059 | 1.000 | 0.159 | 1.000 | 0.000 |
|  | 1.2 | 0.8 | 0.4 | 0.5 | -0.3 | 0.2 | 0.096 | -0.048 | 0.144 | 0.096 | 0.420 | -0.373 | 0.144 | 0.064 | 0.046 | 0.075 | 0.067 | 0.139 | 0.131 | 0.075 | 1.000 | 1.000 | 0.999 | 1.000 | 0.171 | 0.314 | 0.821 |
|  | 0.2 | 0.8 | -0.6 | -0.5 | -0.3 | 0.2 | 0.003 | -0.046 | 0.048 | 0.001 | 0.411 | -0.364 | 0.048 | 0.072 | 0.048 | 0.085 | 0.064 | 0.135 | 0.128 | 0.085 | 0.726 | 1.000 | 1.000 | 1.000 | 0.154 | 0.279 | 0.915 |
|  | 0.2 | -0.8 | 1 | 0.5 | 0.3 | 0.2 | 0.426 | -0.048 | 0.473 | 0.299 | -0.414 | 0.589 | 0.474 | 0.239 | 0.046 | 0.241 | 0.134 | 0.139 | 0.157 | 0.241 | 0.566 | 1.000 | 1.000 | 0.985 | 0.138 | 0.997 | 1.000 |
|  | -1.8 | -0.8 | -1 | -0.5 | -0.3 | -0.2 | -0.449 | -0.045 | -0.403 | -0.243 | 0.413 | -0.574 | -0.403 | 0.254 | 0.047 | 0.252 | 0.135 | 0.142 | 0.171 | 0.252 | 1.000 | 1.000 | 1.000 | 1.000 | 0.165 | 1.000 | 1.000 |
|  | 1 | 0 | 1 | 0.5 | 0.3 | 0.2 | 0.429 | -0.047 | 0.476 | 0.297 | -0.414 | 0.594 | 0.477 | 0.259 | 0.047 | 0.261 | 0.147 | 0.142 | 0.173 | 0.261 | 1.000 | 0.000 | 1.000 | 0.994 | 0.146 | 1.000 | 1.000 |
|  | 1.3 | 0.8 | 0.5 | 0 | 0.3 | 0.2 | 0.166 | -0.047 | 0.214 | 0.032 | -0.418 | 0.600 | 0.214 | 0.151 | 0.044 | 0.150 | 0.065 | 0.141 | 0.172 | 0.150 | 1.000 | 1.000 | 0.997 | 0.000 | 0.175 | 1.000 | 0.946 |
|  | 1.5 | 0.8 | 0.7 | 0.5 | 0 | 0.2 | 0.013 | -0.029 | 0.042 | 0.076 | -0.246 | 0.212 | 0.042 | 0.130 | 0.041 | 0.133 | 0.087 | 0.088 | 0.096 | 0.133 | 1.000 | 1.000 | 0.999 | 0.999 | 0.798 | 0.996 | 0.993 |
|  | 1 | 0.8 | 0.2 | 0 | 0 | 0.2 | -0.045 | -0.028 | -0.017 | 0.020 | -0.242 | 0.205 | -0.017 | 0.067 | 0.041 | 0.074 | 0.047 | 0.086 | 0.094 | 0.074 | 1.000 | 1.000 | 0.352 | 0.002 | 0.800 | 0.999 | 0.025 |
|  | 0 | -1 | 1 | 0.5 | 0.3 | 0.2 | 0.413 | -0.047 | 0.460 | 0.294 | -0.422 | 0.589 | 0.461 | 0.248 | 0.047 | 0.249 | 0.138 | 0.142 | 0.170 | 0.249 | 0.242 | 1.000 | 1.000 | 0.975 | 0.155 | 0.998 | 0.998 |
| Positive | 1.8 | 0.8 | 1 | 0.5 | 0.3 | 0.2 | -0.313 | -0.045 | -0.268 | -0.153 | 0.066 | -0.181 | -0.268 | 0.035 | 0.046 | 0.056 | 0.039 | 0.075 | 0.063 | 0.056 | 1.000 | 0.982 | 0.992 | 0.812 | 0.476 | 0.011 | 0.988 |
|  | 0.8 | 0.8 | 0 | -0.5 | 0.3 | 0.2 | 0.020 | -0.044 | 0.064 | 0.176 | 0.071 | -0.184 | 0.064 | 0.033 | 0.046 | 0.055 | 0.037 | 0.075 | 0.064 | 0.055 | 1.000 | 0.999 | 0.000 | 1.000 | 0.994 | 0.070 | 0.000 |
|  | 1.2 | 0.8 | 0.4 | 0.5 | -0.3 | 0.2 | -0.093 | -0.043 | -0.051 | -0.038 | -0.001 | -0.011 | -0.051 | 0.076 | 0.047 | 0.090 | 0.063 | 0.069 | 0.061 | 0.090 | 1.000 | 1.000 | 0.759 | 0.999 | 0.980 | 0.940 | 0.515 |
|  | 0.2 | 0.8 | -0.6 | -0.5 | -0.3 | 0.2 | 0.051 | -0.048 | 0.099 | 0.110 | 0.003 | -0.013 | 0.099 | 0.036 | 0.045 | 0.054 | 0.052 | 0.065 | 0.056 | 0.054 | 1.000 | 1.000 | 1.000 | 1.000 | 1.000 | 0.965 | 1.000 |
|  | 0.2 | -0.8 | 1 | 0.5 | 0.3 | 0.2 | -0.312 | -0.046 | -0.265 | -0.152 | 0.071 | -0.184 | -0.265 | 0.036 | 0.046 | 0.056 | 0.038 | 0.079 | 0.065 | 0.057 | 0.847 | 1.000 | 1.000 | 1.000 | 0.996 | 0.086 | 1.000 |
|  | -1.8 | -0.8 | -1 | -0.5 | -0.3 | -0.2 | 0.318 | -0.047 | 0.365 | 0.177 | 0.001 | 0.188 | 0.365 | 0.033 | 0.047 | 0.058 | 0.038 | 0.067 | 0.051 | 0.058 | 1.000 | 0.999 | 0.993 | 0.957 | 0.530 | 0.014 | 0.991 |
|  | 1 | 0 | 1 | 0.5 | 0.3 | 0.2 | -0.312 | -0.045 | -0.266 | -0.155 | 0.069 | -0.181 | -0.266 | 0.038 | 0.047 | 0.057 | 0.039 | 0.076 | 0.064 | 0.057 | 1.000 | 0.000 | 1.000 | 0.998 | 0.972 | 0.063 | 1.000 |
|  | 1.3 | 0.8 | 0.5 | 0 | 0.3 | 0.2 | -0.147 | -0.045 | -0.102 | 0.010 | 0.070 | -0.183 | -0.102 | 0.024 | 0.045 | 0.049 | 0.028 | 0.073 | 0.063 | 0.049 | 1.000 | 0.999 | 0.506 | 0.000 | 0.877 | 0.040 | 0.404 |
|  | 1.5 | 0.8 | 0.7 | 0.5 | 0 | 0.2 | -0.081 | -0.026 | -0.054 | -0.069 | 0.234 | -0.219 | -0.054 | 0.047 | 0.042 | 0.061 | 0.048 | 0.097 | 0.090 | 0.061 | 1.000 | 1.000 | 0.990 | 0.907 | 0.573 | 0.033 | 0.951 |
|  | 1 | 0.8 | 0.2 | 0 | 0 | 0.2 | 0.002 | -0.029 | 0.031 | 0.017 | 0.232 | -0.218 | 0.031 | 0.035 | 0.041 | 0.050 | 0.034 | 0.102 | 0.095 | 0.050 | 1.000 | 1.000 | 0.053 | 0.000 | 0.619 | 0.059 | 0.006 |
|  | 0 | -1 | 1 | 0.5 | 0.3 | 0.2 | -0.313 | -0.044 | -0.269 | -0.156 | 0.066 | -0.179 | -0.269 | 0.036 | 0.045 | 0.056 | 0.038 | 0.071 | 0.060 | 0.056 | 1.000 | 1.000 | 1.000 | 1.000 | 0.998 | 0.067 | 1.000 |
|  | 1.8 | 0.8 | 1 | 0.5 | 0.3 | 0.2 | 0.424 | -0.049 | 0.473 | 0.296 | -0.418 | 0.595 | 0.473 | 0.249 | 0.046 | 0.250 | 0.140 | 0.141 | 0.177 | 0.250 | 1.000 | 1.000 | 1.000 | 1.000 | 0.165 | 1.000 | 1.000 |

TE: total effect; DE: direct effect; IE: indirect effect; IE_d: indirect effect calculated by difference method; IE_p: indirect effect calculated by product method; IE1: *X*→*M_1_*→*Y*; IE2: *X*→*M_2_*→*Y*; IE3: *X*→*M_1_*→*M_2_*→*Y*.

**eTable 27**. Simulation results of PSE-IVW for two causally non-ordered mediators when there exist bidirectional causal effects between risk factor and mediators

|  | TRUE | | | | | BIAS | | | | | | SE | | | | | | Power/Type I error | | | | | |
| --- | --- | --- | --- | --- | --- | --- | --- | --- | --- | --- | --- | --- | --- | --- | --- | --- | --- | --- | --- | --- | --- | --- | --- |
|  | TE | DE | IE | IE1 | IE2 | TE | DE | IE_d | IE1 | IE2 | IE_p | TE | DE | IE_d | IE1 | IE2 | IE_p | TE | DE | IE_d | IE1 | IE2 | IE_p |
| Negative | 1.6 | 0.8 | 0.8 | 0.5 | 0.3 | 0.173 | -0.048 | 0.222 | 0.198 | 0.023 | 0.222 | 0.137 | 0.046 | 0.143 | 0.093 | 0.080 | 0.143 | 1 | 1 | 1 | 1 | 0.997 | 1 |
|  | 0.6 | 0.8 | -0.2 | -0.5 | 0.3 | -0.142 | -0.047 | -0.094 | -0.119 | 0.024 | -0.095 | 0.068 | 0.046 | 0.076 | 0.082 | 0.081 | 0.076 | 1 | 1 | 0.928 | 1 | 0.995 | 0.729 |
|  | 0.7 | 0.5 | 0.2 | 0.5 | -0.3 | 0.135 | -0.045 | 0.180 | 0.195 | -0.015 | 0.180 | 0.071 | 0.044 | 0.079 | 0.091 | 0.077 | 0.079 | 1 | 1 | 0.983 | 1 | 0.994 | 0.878 |
|  | -0.3 | 0.5 | -0.8 | -0.5 | -0.3 | -0.178 | -0.048 | -0.131 | -0.117 | -0.014 | -0.131 | 0.128 | 0.046 | 0.133 | 0.084 | 0.079 | 0.133 | 0.959 | 1 | 1 | 1 | 0.994 | 1 |
|  | 0 | -0.8 | 0.8 | 0.5 | 0.3 | 0.175 | -0.048 | 0.223 | 0.200 | 0.024 | 0.224 | 0.143 | 0.046 | 0.145 | 0.093 | 0.081 | 0.145 | 0.174 | 1 | 1 | 0.999 | 0.996 | 1 |
|  | -1.6 | -0.8 | -0.8 | -0.5 | -0.3 | -0.184 | -0.045 | -0.139 | -0.122 | -0.017 | -0.139 | 0.130 | 0.047 | 0.134 | 0.084 | 0.079 | 0.134 | 1 | 1 | 1 | 1 | 0.989 | 1 |
|  | 0.8 | 0 | 0.8 | 0.5 | 0.3 | 0.170 | -0.044 | 0.214 | 0.194 | 0.020 | 0.214 | 0.137 | 0.047 | 0.139 | 0.089 | 0.080 | 0.139 | 1 | 0 | 1 | 1 | 0.994 | 1 |
|  | 1.3 | 0.8 | 0.5 | 0.5 | 0 | 0.155 | -0.045 | 0.200 | 0.195 | 0.005 | 0.200 | 0.082 | 0.047 | 0.088 | 0.092 | 0.022 | 0.088 | 1 | 1 | 1 | 1 | 0 | 1 |
|  | 1.3 | 0.8 | 0.5 | 0.5 | 0 | -0.053 | -0.026 | -0.028 | 0.011 | -0.039 | -0.028 | 0.081 | 0.042 | 0.089 | 0.063 | 0.041 | 0.089 | 1 | 1 | 0.982 | 1 | 0.169 | 0.997 |
|  | 0.8 | 0.8 | 0 | 0 | 0 | -0.002 | -0.047 | 0.045 | 0.041 | 0.004 | 0.045 | 0.022 | 0.047 | 0.042 | 0.051 | 0.023 | 0.042 | 1 | 1 | 0.042 | 0.074 | 0.027 | 0.029 |
| Positive | 1.6 | 0.8 | 0.8 | 0.5 | 0.3 | -0.168 | -0.049 | -0.119 | -0.097 | -0.021 | -0.119 | 0.038 | 0.048 | 0.060 | 0.047 | 0.037 | 0.060 | 1 | 0.995 | 0.997 | 0.989 | 0.812 | 1 |
|  | 0.6 | 0.8 | -0.2 | -0.5 | 0.3 | 0.075 | -0.047 | 0.122 | 0.144 | -0.023 | 0.121 | 0.050 | 0.045 | 0.066 | 0.044 | 0.037 | 0.066 | 1 | 1 | 0.007 | 1 | 1 | 0.026 |
|  | 0.7 | 0.5 | 0.2 | 0.5 | -0.3 | -0.069 | -0.049 | -0.020 | -0.097 | 0.077 | -0.021 | 0.047 | 0.047 | 0.064 | 0.045 | 0.034 | 0.064 | 1 | 1 | 0.295 | 1 | 0.996 | 0.468 |
|  | -0.3 | 0.5 | -0.8 | -0.5 | -0.3 | 0.174 | -0.047 | 0.221 | 0.144 | 0.077 | 0.221 | 0.035 | 0.047 | 0.057 | 0.044 | 0.034 | 0.057 | 0.937 | 1 | 1 | 1 | 1 | 1 |
|  | 0 | -0.8 | 0.8 | 0.5 | 0.3 | -0.168 | -0.045 | -0.123 | -0.099 | -0.023 | -0.123 | 0.042 | 0.047 | 0.061 | 0.047 | 0.038 | 0.061 | 0.982 | 1 | 1 | 1 | 1 | 1 |
|  | -1.6 | -0.8 | -0.8 | -0.5 | -0.3 | 0.173 | -0.047 | 0.220 | 0.144 | 0.076 | 0.220 | 0.036 | 0.048 | 0.057 | 0.044 | 0.035 | 0.057 | 1 | 1 | 0.998 | 0.999 | 0.875 | 1 |
|  | 0.8 | 0 | 0.8 | 0.5 | 0.3 | -0.167 | -0.045 | -0.122 | -0.099 | -0.023 | -0.122 | 0.039 | 0.046 | 0.057 | 0.046 | 0.037 | 0.057 | 1 | 0 | 1 | 1 | 0.996 | 1 |
|  | 1.3 | 0.8 | 0.5 | 0.5 | 0 | -0.119 | -0.046 | -0.072 | -0.099 | 0.027 | -0.072 | 0.037 | 0.047 | 0.059 | 0.047 | 0.027 | 0.059 | 1 | 1 | 0.898 | 1 | 0 | 0.98 |
|  | 1.3 | 0.8 | 0.5 | 0.5 | 0 | 0.033 | -0.027 | 0.061 | 0.014 | 0.047 | 0.061 | 0.052 | 0.039 | 0.064 | 0.059 | 0.042 | 0.064 | 1 | 1 | 0.994 | 0.996 | 0.203 | 0.996 |
|  | 0.8 | 0.8 | 0 | 0 | 0 | 0.004 | -0.047 | 0.050 | 0.023 | 0.027 | 0.051 | 0.011 | 0.045 | 0.044 | 0.029 | 0.025 | 0.044 | 1 | 1 | 0 | 0 | 0 | 0.002 |

TE: total effect; DE: direct effect; IE: indirect effect; IE_d: indirect effect calculated by difference method; IE_p: indirect effect calculated by product method; IE1: *X*→*M_1_*→*Y*; IE2: *X*→*M_2_*→*Y*.

**eTable 28**. Simulation results of PE-Egger for two causally ordered mediators when there exist bidirectional causal effects between risk factor and mediators

|  |  | TRUE | | | | | | BIAS | | | | | | | SE | | | | | | | Power/Type I error | | | | | | | | | |
| --- | --- | --- | --- | --- | --- | --- | --- | --- | --- | --- | --- | --- | --- | --- | --- | --- | --- | --- | --- | --- | --- | --- | --- | --- | --- | --- | --- | --- | --- | --- | --- |
|  |  | TE | DE | IE | IE1 | IE2 | IE3 | TE | DE | IE_d | IE1 | IE2 | IE3 | IE_p | TE | DE | IE_d | IE1 | IE2 | IE3 | IE_p | TE | DE | IE_d | IE1 | IE2 | IE3 | IE_p | GY | GM1 | GM2 |
| Negative | IVW | 1.8 | 0.8 | 1 | 0.5 | 0.3 | 0.2 | 1.486 | -0.198 | 1.684 | 0.885 | -0.445 | 1.246 | 1.685 | 0.066 | 0.110 | 0.129 | 0.281 | 0.114 | 0.293 | 0.129 | 1.000 | 0.787 | 1.000 | 0.618 | 0.150 | 0.777 | 0.990 |  |  |  |
|  |  | 0.8 | 0.8 | 0 | -0.5 | 0.3 | 0.2 | 0.111 | -0.196 | 0.307 | -0.512 | -0.444 | 1.263 | 0.307 | 0.021 | 0.106 | 0.107 | 0.300 | 0.116 | 0.314 | 0.107 | 1.000 | 0.997 | 0.799 | 0.911 | 0.214 | 0.996 | 0.000 |  |  |  |
|  |  | 1.2 | 0.8 | 0.4 | 0.5 | -0.3 | 0.2 | 0.115 | -0.194 | 0.309 | 0.338 | 0.420 | -0.449 | 0.309 | 0.012 | 0.110 | 0.110 | 0.120 | 0.095 | 0.103 | 0.110 | 1.000 | 0.932 | 0.989 | 0.997 | 0.140 | 0.555 | 0.845 |  |  |  |
|  |  | 0.2 | 0.8 | -0.6 | -0.5 | -0.3 | 0.2 | -0.107 | -0.188 | 0.081 | 0.109 | 0.418 | -0.446 | 0.081 | 0.012 | 0.107 | 0.107 | 0.115 | 0.093 | 0.101 | 0.107 | 1.000 | 0.999 | 0.994 | 0.898 | 0.209 | 0.704 | 0.844 |  |  |  |
|  |  | 0.2 | -0.8 | 1 | 0.5 | 0.3 | 0.2 | 1.485 | -0.190 | 1.675 | 0.859 | -0.442 | 1.257 | 1.674 | 0.067 | 0.106 | 0.125 | 0.283 | 0.111 | 0.287 | 0.125 | 1.000 | 1.000 | 1.000 | 0.736 | 0.144 | 0.888 | 0.998 |  |  |  |
|  |  | -1.8 | -0.8 | -1 | -0.5 | -0.3 | -0.2 | -1.324 | -0.190 | -1.134 | -0.510 | 0.415 | -1.041 | -1.135 | 0.054 | 0.106 | 0.120 | 0.290 | 0.094 | 0.267 | 0.120 | 1.000 | 1.000 | 1.000 | 0.398 | 0.103 | 0.752 | 0.975 |  |  |  |
|  |  | 1 | 0 | 1 | 0.5 | 0.3 | 0.2 | 1.487 | -0.192 | 1.679 | 0.880 | -0.437 | 1.235 | 1.678 | 0.068 | 0.109 | 0.129 | 0.303 | 0.117 | 0.313 | 0.129 | 1.000 | 0.008 | 1.000 | 0.738 | 0.151 | 0.843 | 0.997 |  |  |  |
|  |  | 1.3 | 0.8 | 0.5 | 0 | 0.3 | 0.2 | 0.800 | -0.193 | 0.993 | 0.191 | -0.443 | 1.245 | 0.993 | 0.040 | 0.107 | 0.113 | 0.280 | 0.113 | 0.293 | 0.113 | 1.000 | 0.982 | 1.000 | 0.010 | 0.216 | 0.987 | 0.939 |  |  |  |
|  |  | 1.5 | 0.8 | 0.7 | 0.5 | 0 | 0.2 | 0.398 | -0.118 | 0.516 | 0.330 | -0.315 | 0.502 | 0.516 | 0.027 | 0.105 | 0.109 | 0.173 | 0.108 | 0.180 | 0.109 | 1.000 | 0.923 | 1.000 | 0.661 | 0.550 | 0.577 | 0.754 |  |  |  |
|  |  | 1 | 0.8 | 0.2 | 0 | 0 | 0.2 | 0.182 | -0.115 | 0.297 | 0.119 | -0.309 | 0.487 | 0.297 | 0.018 | 0.109 | 0.111 | 0.175 | 0.101 | 0.169 | 0.111 | 1.000 | 0.999 | 0.943 | 0.028 | 0.921 | 0.953 | 0.117 |  |  |  |
|  |  | 0 | -1 | 1 | 0.5 | 0.3 | 0.2 | 1.487 | -0.191 | 1.678 | 0.859 | -0.448 | 1.266 | 1.677 | 0.067 | 0.108 | 0.126 | 0.300 | 0.116 | 0.313 | 0.125 | 1.000 | 1.000 | 1.000 | 0.705 | 0.162 | 0.856 | 0.996 |  |  |  |
|  | Egger | 1.8 | 0.8 | 1 | 0.5 | 0.3 | 0.2 | 0.939 | -0.083 | 1.022 | 0.535 | -0.386 | 0.874 | 1.022 | 0.111 | 0.123 | 0.164 | 0.232 | 0.123 | 0.237 | 0.164 | 1.000 | 0.762 | 1.000 | 0.403 | 0.019 | 0.690 | 0.941 | 0.000 | 0.060 | 0.996 |
|  |  | 0.8 | 0.8 | 0 | -0.5 | 0.3 | 0.2 | -0.012 | -0.082 | 0.070 | -0.426 | -0.383 | 0.879 | 0.069 | 0.041 | 0.121 | 0.121 | 0.248 | 0.123 | 0.256 | 0.121 | 1.000 | 0.999 | 0.083 | 0.959 | 0.064 | 0.998 | 0.000 | 0.419 | 0.065 | 0.994 |
|  |  | 1.2 | 0.8 | 0.4 | 0.5 | -0.3 | 0.2 | 0.051 | -0.084 | 0.135 | 0.139 | 0.375 | -0.379 | 0.135 | 0.025 | 0.129 | 0.129 | 0.133 | 0.116 | 0.117 | 0.129 | 1.000 | 0.936 | 0.727 | 0.870 | 0.039 | 0.198 | 0.251 | 0.018 | 0.085 | 0.957 |
|  |  | 0.2 | 0.8 | -0.6 | -0.5 | -0.3 | 0.2 | -0.060 | -0.072 | 0.012 | 0.015 | 0.372 | -0.375 | 0.012 | 0.029 | 0.123 | 0.126 | 0.128 | 0.112 | 0.115 | 0.126 | 0.996 | 1.000 | 0.995 | 0.964 | 0.072 | 0.298 | 0.819 | 0.440 | 0.090 | 0.964 |
|  |  | 0.2 | -0.8 | 1 | 0.5 | 0.3 | 0.2 | 0.936 | -0.073 | 1.009 | 0.518 | -0.378 | 0.869 | 1.009 | 0.110 | 0.123 | 0.159 | 0.239 | 0.122 | 0.245 | 0.159 | 1.000 | 0.968 | 1.000 | 0.554 | 0.043 | 0.826 | 0.982 | 0.002 | 0.083 | 0.994 |
|  |  | -1.8 | -0.8 | -1 | -0.5 | -0.3 | -0.2 | -0.943 | -0.076 | -0.867 | -0.424 | 0.372 | -0.816 | -0.867 | 0.098 | 0.123 | 0.156 | 0.240 | 0.112 | 0.218 | 0.156 | 1.000 | 0.982 | 1.000 | 0.490 | 0.020 | 0.792 | 0.980 | 0.009 | 0.070 | 0.993 |
|  |  | 1 | 0 | 1 | 0.5 | 0.3 | 0.2 | 0.936 | -0.075 | 1.011 | 0.530 | -0.382 | 0.863 | 1.011 | 0.111 | 0.120 | 0.157 | 0.244 | 0.123 | 0.252 | 0.157 | 1.000 | 0.000 | 1.000 | 0.535 | 0.033 | 0.753 | 0.982 | 0.004 | 0.058 | 0.991 |
|  |  | 1.3 | 0.8 | 0.5 | 0 | 0.3 | 0.2 | 0.465 | -0.077 | 0.542 | 0.062 | -0.383 | 0.863 | 0.542 | 0.066 | 0.126 | 0.138 | 0.230 | 0.118 | 0.237 | 0.138 | 1.000 | 0.984 | 1.000 | 0.002 | 0.040 | 0.973 | 0.600 | 0.116 | 0.062 | 0.990 |
|  |  | 1.5 | 0.8 | 0.7 | 0.5 | 0 | 0.2 | 0.158 | -0.054 | 0.213 | 0.157 | -0.239 | 0.295 | 0.213 | 0.050 | 0.109 | 0.119 | 0.151 | 0.098 | 0.144 | 0.119 | 1.000 | 0.940 | 0.993 | 0.466 | 0.250 | 0.390 | 0.454 | 0.002 | 0.161 | 0.989 |
|  |  | 1 | 0.8 | 0.2 | 0 | 0 | 0.2 | 0.042 | -0.049 | 0.091 | 0.039 | -0.237 | 0.289 | 0.091 | 0.030 | 0.113 | 0.114 | 0.153 | 0.093 | 0.140 | 0.114 | 1.000 | 0.999 | 0.405 | 0.007 | 0.711 | 0.898 | 0.011 | 0.120 | 0.140 | 0.977 |
|  |  | 0 | -1 | 1 | 0.5 | 0.3 | 0.2 | 0.937 | -0.079 | 1.016 | 0.521 | -0.388 | 0.882 | 1.016 | 0.111 | 0.122 | 0.161 | 0.243 | 0.120 | 0.254 | 0.161 | 1.000 | 0.998 | 1.000 | 0.516 | 0.026 | 0.784 | 0.969 | 0.005 | 0.070 | 0.996 |
| Positive | IVW | 1.8 | 0.8 | 1 | 0.5 | 0.3 | 0.2 | -0.274 | -0.194 | -0.080 | -0.101 | 0.093 | -0.072 | -0.080 | 0.007 | 0.106 | 0.106 | 0.082 | 0.127 | 0.101 | 0.106 | 1.000 | 0.058 | 0.787 | 0.019 | 0.117 | 0.017 | 0.580 |  |  |  |
|  |  | 0.8 | 0.8 | 0 | -0.5 | 0.3 | 0.2 | 0.034 | -0.185 | 0.219 | 0.201 | 0.086 | -0.068 | 0.219 | 0.006 | 0.105 | 0.105 | 0.086 | 0.122 | 0.102 | 0.105 | 1.000 | 0.874 | 0.025 | 0.296 | 0.803 | 0.148 | 0.000 |  |  |  |
|  |  | 1.2 | 0.8 | 0.4 | 0.5 | -0.3 | 0.2 | 0.002 | -0.196 | 0.198 | 0.158 | 0.156 | -0.116 | 0.198 | 0.011 | 0.109 | 0.109 | 0.093 | 0.075 | 0.053 | 0.109 | 1.000 | 0.888 | 0.905 | 0.990 | 0.040 | 0.016 | 0.742 |  |  |  |
|  |  | 0.2 | 0.8 | -0.6 | -0.5 | -0.3 | 0.2 | 0.045 | -0.194 | 0.239 | 0.199 | 0.154 | -0.114 | 0.239 | 0.006 | 0.109 | 0.108 | 0.093 | 0.077 | 0.055 | 0.108 | 1.000 | 0.999 | 0.902 | 0.890 | 0.546 | 0.331 | 0.815 |  |  |  |
|  |  | 0.2 | -0.8 | 1 | 0.5 | 0.3 | 0.2 | -0.273 | -0.192 | -0.081 | -0.098 | 0.092 | -0.075 | -0.081 | 0.007 | 0.106 | 0.107 | 0.083 | 0.122 | 0.095 | 0.107 | 1.000 | 1.000 | 1.000 | 0.994 | 0.944 | 0.167 | 1.000 |  |  |  |
|  |  | -1.8 | -0.8 | -1 | -0.5 | -0.3 | -0.2 | 0.322 | -0.190 | 0.511 | 0.209 | 0.154 | 0.149 | 0.511 | 0.005 | 0.108 | 0.108 | 0.085 | 0.078 | 0.048 | 0.108 | 1.000 | 0.905 | 0.059 | 0.002 | 0.000 | 0.000 | 0.012 |  |  |  |
|  |  | 1 | 0 | 1 | 0.5 | 0.3 | 0.2 | -0.273 | -0.193 | -0.081 | -0.094 | 0.085 | -0.072 | -0.081 | 0.007 | 0.106 | 0.106 | 0.084 | 0.116 | 0.093 | 0.106 | 1.000 | 0.007 | 1.000 | 0.739 | 0.826 | 0.134 | 0.997 |  |  |  |
|  |  | 1.3 | 0.8 | 0.5 | 0 | 0.3 | 0.2 | -0.120 | -0.193 | 0.073 | 0.056 | 0.089 | -0.071 | 0.073 | 0.006 | 0.108 | 0.108 | 0.087 | 0.117 | 0.098 | 0.108 | 1.000 | 0.392 | 0.406 | 0.000 | 0.459 | 0.048 | 0.153 |  |  |  |
|  |  | 1.5 | 0.8 | 0.7 | 0.5 | 0 | 0.2 | -0.039 | -0.119 | 0.080 | 0.003 | 0.173 | -0.095 | 0.081 | 0.009 | 0.110 | 0.110 | 0.107 | 0.158 | 0.153 | 0.110 | 1.000 | 0.452 | 0.787 | 0.147 | 0.118 | 0.050 | 0.420 |  |  |  |
|  |  | 1 | 0.8 | 0.2 | 0 | 0 | 0.2 | 0.033 | -0.111 | 0.144 | 0.064 | 0.172 | -0.092 | 0.144 | 0.007 | 0.109 | 0.109 | 0.106 | 0.149 | 0.145 | 0.109 | 1.000 | 0.910 | 0.127 | 0.000 | 0.186 | 0.100 | 0.003 |  |  |  |
|  |  | 0 | -1 | 1 | 0.5 | 0.3 | 0.2 | -0.274 | -0.197 | -0.077 | -0.096 | 0.087 | -0.067 | -0.076 | 0.007 | 0.109 | 0.109 | 0.084 | 0.126 | 0.100 | 0.109 | 1.000 | 1.000 | 1.000 | 0.991 | 0.929 | 0.198 | 1.000 |  |  |  |
|  | Egger | 1.8 | 0.8 | 1 | 0.5 | 0.3 | 0.2 | -0.331 | -0.078 | -0.253 | -0.149 | 0.065 | -0.168 | -0.253 | 0.016 | 0.125 | 0.125 | 0.081 | 0.119 | 0.071 | 0.125 | 1.000 | 0.042 | 0.125 | 0.001 | 0.002 | 0.000 | 0.054 | 0.000 | 0.544 | 0.335 |
|  |  | 0.8 | 0.8 | 0 | -0.5 | 0.3 | 0.2 | 0.002 | -0.071 | 0.073 | 0.179 | 0.060 | -0.166 | 0.073 | 0.014 | 0.122 | 0.122 | 0.083 | 0.114 | 0.069 | 0.122 | 1.000 | 0.865 | 0.000 | 0.361 | 0.397 | 0.005 | 0.000 | 0.003 | 0.599 | 0.300 |
|  |  | 1.2 | 0.8 | 0.4 | 0.5 | -0.3 | 0.2 | -0.040 | -0.084 | 0.044 | 0.021 | 0.049 | -0.027 | 0.044 | 0.027 | 0.123 | 0.125 | 0.104 | 0.102 | 0.079 | 0.125 | 1.000 | 0.878 | 0.320 | 0.773 | 0.096 | 0.087 | 0.108 | 0.005 | 0.815 | 0.419 |
|  |  | 0.2 | 0.8 | -0.6 | -0.5 | -0.3 | 0.2 | 0.054 | -0.081 | 0.135 | 0.112 | 0.047 | -0.025 | 0.135 | 0.016 | 0.121 | 0.120 | 0.102 | 0.101 | 0.078 | 0.120 | 1.000 | 0.998 | 0.956 | 0.955 | 0.812 | 0.754 | 0.874 | 0.389 | 0.831 | 0.472 |
|  |  | 0.2 | -0.8 | 1 | 0.5 | 0.3 | 0.2 | -0.330 | -0.075 | -0.255 | -0.148 | 0.062 | -0.169 | -0.255 | 0.015 | 0.124 | 0.125 | 0.082 | 0.119 | 0.070 | 0.125 | 1.000 | 1.000 | 0.999 | 0.985 | 0.949 | 0.017 | 0.999 | 0.401 | 0.518 | 0.325 |
|  |  | -1.8 | -0.8 | -1 | -0.5 | -0.3 | -0.2 | 0.332 | -0.077 | 0.409 | 0.187 | 0.046 | 0.177 | 0.409 | 0.013 | 0.121 | 0.120 | 0.083 | 0.102 | 0.053 | 0.120 | 1.000 | 0.453 | 0.049 | 0.005 | 0.000 | 0.000 | 0.021 | 0.000 | 0.547 | 0.297 |
|  |  | 1 | 0 | 1 | 0.5 | 0.3 | 0.2 | -0.330 | -0.077 | -0.254 | -0.143 | 0.057 | -0.168 | -0.254 | 0.016 | 0.123 | 0.123 | 0.083 | 0.110 | 0.066 | 0.123 | 1.000 | 0.000 | 0.914 | 0.549 | 0.386 | 0.001 | 0.858 | 0.005 | 0.549 | 0.314 |
|  |  | 1.3 | 0.8 | 0.5 | 0 | 0.3 | 0.2 | -0.164 | -0.078 | -0.086 | 0.022 | 0.057 | -0.165 | -0.086 | 0.012 | 0.122 | 0.121 | 0.083 | 0.112 | 0.070 | 0.121 | 1.000 | 0.385 | 0.015 | 0.000 | 0.043 | 0.000 | 0.005 | 0.000 | 0.556 | 0.274 |
|  |  | 1.5 | 0.8 | 0.7 | 0.5 | 0 | 0.2 | -0.111 | -0.057 | -0.054 | -0.050 | 0.193 | -0.198 | -0.054 | 0.019 | 0.113 | 0.114 | 0.106 | 0.116 | 0.107 | 0.114 | 1.000 | 0.463 | 0.319 | 0.052 | 0.027 | 0.002 | 0.146 | 0.000 | 0.850 | 0.176 |
|  |  | 1 | 0.8 | 0.2 | 0 | 0 | 0.2 | -0.028 | -0.045 | 0.017 | 0.021 | 0.200 | -0.204 | 0.017 | 0.017 | 0.113 | 0.112 | 0.105 | 0.109 | 0.099 | 0.112 | 1.000 | 0.918 | 0.003 | 0.000 | 0.199 | 0.012 | 0.000 | 0.000 | 0.846 | 0.178 |
|  |  | 0 | -1 | 1 | 0.5 | 0.3 | 0.2 | -0.331 | -0.087 | -0.244 | -0.143 | 0.062 | -0.163 | -0.244 | 0.015 | 0.122 | 0.122 | 0.080 | 0.118 | 0.072 | 0.122 | 1.000 | 1.000 | 1.000 | 0.983 | 0.937 | 0.011 | 0.998 | 0.367 | 0.527 | 0.322 |

TE: total effect; DE: direct effect; IE: indirect effect; IE_d: indirect effect calculated by difference method; IE_p: indirect effect calculated by product method; IE1: *X*→*M_1_*→*Y*; IE2: *X*→*M_2_*→*Y*; IE3: *X*→*M_1_*→*M_2_*→*Y*.

**eTable 29**. Simulation results of PE-Egger for two causally non-ordered mediators when there exist bidirectional causal effects between risk factor and mediators

|  |  | TE | DE | IE | IE1 | IE2 | TE | DE | IE_d | IE1 | IE2 | IE_p | TE | DE | IE_d | IE1 | IE2 | IE_p | TE | DE | IE_d | IE1 | IE2 | IE_p | GY | GM1 | GM2 |
| --- | --- | --- | --- | --- | --- | --- | --- | --- | --- | --- | --- | --- | --- | --- | --- | --- | --- | --- | --- | --- | --- | --- | --- | --- | --- | --- | --- |
| Negative | IVW | 1.6 | 0.8 | 0.8 | 0.5 | 0.3 | 0.567 | -0.195 | 0.762 | 0.521 | 0.242 | 0.763 | 0.028 | 0.104 | 0.109 | 0.167 | 0.104 | 0.109 | 1.000 | 0.816 | 1.000 | 0.924 | 0.802 | 1.000 |  |  |  |
|  |  | 0.6 | 0.8 | -0.2 | -0.5 | 0.3 | -0.047 | -0.192 | 0.145 | -0.097 | 0.242 | 0.145 | 0.013 | 0.106 | 0.107 | 0.163 | 0.103 | 0.107 | 1.000 | 0.997 | 0.079 | 0.937 | 0.999 | 0.005 |  |  |  |
|  |  | 0.7 | 0.5 | 0.2 | 0.5 | -0.3 | 0.158 | -0.189 | 0.348 | 0.516 | -0.169 | 0.347 | 0.014 | 0.111 | 0.112 | 0.172 | 0.105 | 0.112 | 1.000 | 0.741 | 0.995 | 0.999 | 0.983 | 0.867 |  |  |  |
|  |  | -0.3 | 0.5 | -0.8 | -0.5 | -0.3 | -0.459 | -0.187 | -0.271 | -0.103 | -0.168 | -0.271 | 0.023 | 0.105 | 0.107 | 0.167 | 0.104 | 0.107 | 1.000 | 0.745 | 1.000 | 0.927 | 0.975 | 0.999 |  |  |  |
|  |  | 0 | -0.8 | 0.8 | 0.5 | 0.3 | 0.570 | -0.197 | 0.767 | 0.522 | 0.244 | 0.767 | 0.029 | 0.113 | 0.118 | 0.175 | 0.105 | 0.118 | 1.000 | 1.000 | 1.000 | 0.994 | 0.978 | 1.000 |  |  |  |
|  |  | -1.6 | -0.8 | -0.8 | -0.5 | -0.3 | -0.457 | -0.189 | -0.269 | -0.101 | -0.168 | -0.269 | 0.023 | 0.110 | 0.110 | 0.165 | 0.099 | 0.110 | 1.000 | 1.000 | 1.000 | 0.534 | 0.806 | 0.983 |  |  |  |
|  |  | 0.8 | 0 | 0.8 | 0.5 | 0.3 | 0.571 | -0.197 | 0.768 | 0.527 | 0.241 | 0.768 | 0.030 | 0.108 | 0.113 | 0.172 | 0.101 | 0.113 | 1.000 | 0.048 | 1.000 | 0.981 | 0.954 | 1.000 |  |  |  |
|  |  | 1.3 | 0.8 | 0.5 | 0.5 | 0 | 0.363 | -0.190 | 0.553 | 0.516 | 0.037 | 0.553 | 0.018 | 0.104 | 0.106 | 0.166 | 0.104 | 0.106 | 1.000 | 0.978 | 1.000 | 0.985 | 0.004 | 0.990 |  |  |  |
|  |  | 1.3 | 0.8 | 0.5 | 0.5 | 0 | 0.133 | -0.112 | 0.245 | 0.202 | 0.043 | 0.245 | 0.016 | 0.105 | 0.107 | 0.113 | 0.012 | 0.107 | 1.000 | 0.958 | 0.981 | 0.941 | 0.629 | 0.970 |  |  |  |
|  |  | 0.8 | 0.8 | 0 | 0 | 0 | 0.055 | -0.189 | 0.245 | 0.214 | 0.031 | 0.245 | 0.010 | 0.102 | 0.102 | 0.159 | 0.101 | 0.102 | 1.000 | 0.998 | 0.612 | 0.268 | 0.063 | 0.130 |  |  |  |
|  | Egger | 1.6 | 0.8 | 0.8 | 0.5 | 0.3 | 0.327 | -0.081 | 0.409 | 0.281 | 0.128 | 0.409 | 0.054 | 0.122 | 0.132 | 0.162 | 0.085 | 0.132 | 1.000 | 0.820 | 0.999 | 0.653 | 0.714 | 0.990 | 0.001 | 0.913 | 0.980 |
|  |  | 0.6 | 0.8 | -0.2 | -0.5 | 0.3 | -0.090 | -0.079 | -0.011 | -0.140 | 0.129 | -0.011 | 0.028 | 0.122 | 0.123 | 0.153 | 0.089 | 0.123 | 1.000 | 1.000 | 0.423 | 0.976 | 0.998 | 0.152 | 0.407 | 0.918 | 0.984 |
|  |  | 0.7 | 0.5 | 0.2 | 0.5 | -0.3 | 0.091 | -0.076 | 0.167 | 0.275 | -0.108 | 0.167 | 0.029 | 0.125 | 0.126 | 0.163 | 0.090 | 0.126 | 1.000 | 0.874 | 0.779 | 0.989 | 0.990 | 0.434 | 0.309 | 0.912 | 0.986 |
|  |  | -0.3 | 0.5 | -0.8 | -0.5 | -0.3 | -0.324 | -0.077 | -0.247 | -0.143 | -0.104 | -0.247 | 0.047 | 0.120 | 0.127 | 0.160 | 0.087 | 0.127 | 1.000 | 0.855 | 1.000 | 0.961 | 0.988 | 1.000 | 0.241 | 0.924 | 0.983 |
|  |  | 0 | -0.8 | 0.8 | 0.5 | 0.3 | 0.326 | -0.081 | 0.407 | 0.278 | 0.129 | 0.407 | 0.054 | 0.130 | 0.139 | 0.165 | 0.091 | 0.139 | 1.000 | 0.996 | 1.000 | 0.932 | 0.955 | 0.999 | 0.065 | 0.916 | 0.987 |
|  |  | -1.6 | -0.8 | -0.8 | -0.5 | -0.3 | -0.326 | -0.074 | -0.252 | -0.146 | -0.107 | -0.252 | 0.050 | 0.129 | 0.134 | 0.162 | 0.083 | 0.134 | 1.000 | 0.990 | 0.998 | 0.625 | 0.840 | 0.980 | 0.017 | 0.928 | 0.983 |
|  |  | 0.8 | 0 | 0.8 | 0.5 | 0.3 | 0.327 | -0.086 | 0.413 | 0.288 | 0.125 | 0.413 | 0.053 | 0.124 | 0.134 | 0.163 | 0.084 | 0.134 | 1.000 | 0.002 | 1.000 | 0.891 | 0.911 | 0.999 | 0.019 | 0.915 | 0.979 |
|  |  | 1.3 | 0.8 | 0.5 | 0.5 | 0 | 0.210 | -0.076 | 0.287 | 0.279 | 0.008 | 0.287 | 0.033 | 0.117 | 0.120 | 0.155 | 0.084 | 0.120 | 1.000 | 0.975 | 0.994 | 0.910 | 0.001 | 0.890 | 0.039 | 0.906 | 0.980 |
|  |  | 1.3 | 0.8 | 0.5 | 0.5 | 0 | -0.005 | -0.047 | 0.042 | 0.045 | -0.003 | 0.042 | 0.030 | 0.111 | 0.113 | 0.112 | 0.014 | 0.113 | 1.000 | 0.964 | 0.671 | 0.721 | 0.007 | 0.702 | 0.001 | 0.906 | 0.976 |
|  |  | 0.8 | 0.8 | 0 | 0 | 0 | 0.000 | -0.076 | 0.076 | 0.071 | 0.005 | 0.076 | 0.019 | 0.119 | 0.119 | 0.151 | 0.081 | 0.119 | 1.000 | 0.997 | 0.075 | 0.075 | 0.057 | 0.016 | 0.382 | 0.904 | 0.987 |
| Positive | IVW | 1.6 | 0.8 | 0.8 | 0.5 | 0.3 | -0.117 | -0.189 | 0.072 | 0.006 | 0.066 | 0.072 | 0.008 | 0.106 | 0.107 | 0.080 | 0.065 | 0.107 | 1.000 | 0.274 | 0.904 | 0.510 | 0.364 | 0.949 |  |  |  |
|  |  | 0.6 | 0.8 | -0.2 | -0.5 | 0.3 | 0.079 | -0.194 | 0.273 | 0.206 | 0.067 | 0.273 | 0.007 | 0.106 | 0.106 | 0.083 | 0.064 | 0.106 | 1.000 | 0.971 | 0.006 | 0.686 | 0.982 | 0.011 |  |  |  |
|  |  | 0.7 | 0.5 | 0.2 | 0.5 | -0.3 | -0.023 | -0.192 | 0.169 | 0.005 | 0.164 | 0.169 | 0.007 | 0.106 | 0.106 | 0.083 | 0.062 | 0.106 | 1.000 | 0.474 | 0.718 | 0.990 | 0.235 | 0.732 |  |  |  |
|  |  | -0.3 | 0.5 | -0.8 | -0.5 | -0.3 | 0.173 | -0.195 | 0.369 | 0.206 | 0.162 | 0.369 | 0.006 | 0.106 | 0.106 | 0.080 | 0.066 | 0.106 | 1.000 | 0.786 | 0.972 | 0.944 | 0.576 | 0.980 |  |  |  |
|  |  | 0 | -0.8 | 0.8 | 0.5 | 0.3 | -0.118 | -0.191 | 0.073 | 0.003 | 0.070 | 0.073 | 0.008 | 0.100 | 0.101 | 0.078 | 0.063 | 0.101 | 1.000 | 1.000 | 1.000 | 1.000 | 0.999 | 1.000 |  |  |  |
|  |  | -1.6 | -0.8 | -0.8 | -0.5 | -0.3 | 0.174 | -0.192 | 0.366 | 0.203 | 0.163 | 0.366 | 0.006 | 0.102 | 0.102 | 0.079 | 0.064 | 0.102 | 1.000 | 0.985 | 0.124 | 0.053 | 0.003 | 0.126 |  |  |  |
|  |  | 0.8 | 0 | 0.8 | 0.5 | 0.3 | -0.117 | -0.195 | 0.077 | 0.009 | 0.069 | 0.077 | 0.008 | 0.108 | 0.109 | 0.084 | 0.065 | 0.109 | 1.000 | 0.051 | 1.000 | 0.990 | 0.963 | 1.000 |  |  |  |
|  |  | 1.3 | 0.8 | 0.5 | 0.5 | 0 | -0.071 | -0.192 | 0.122 | 0.009 | 0.113 | 0.122 | 0.007 | 0.102 | 0.102 | 0.079 | 0.063 | 0.102 | 1.000 | 0.643 | 0.756 | 0.837 | 0.000 | 0.803 |  |  |  |
|  |  | 1.3 | 0.8 | 0.5 | 0.5 | 0 | 0.091 | -0.109 | 0.200 | 0.150 | 0.050 | 0.200 | 0.011 | 0.113 | 0.113 | 0.110 | 0.011 | 0.113 | 1.000 | 0.829 | 0.859 | 0.806 | 0.905 | 0.872 |  |  |  |
|  |  | 0.8 | 0.8 | 0 | 0 | 0 | 0.028 | -0.192 | 0.220 | 0.104 | 0.116 | 0.220 | 0.004 | 0.102 | 0.102 | 0.078 | 0.062 | 0.102 | 1.000 | 0.968 | 0.098 | 0.017 | 0.064 | 0.107 |  |  |  |
|  | Egger | 1.6 | 0.8 | 0.8 | 0.5 | 0.3 | -0.181 | -0.077 | -0.104 | -0.079 | -0.025 | -0.104 | 0.017 | 0.127 | 0.127 | 0.086 | 0.071 | 0.127 | 1.000 | 0.230 | 0.276 | 0.124 | 0.016 | 0.458 | 0.000 | 0.722 | 0.398 |
|  |  | 0.6 | 0.8 | -0.2 | -0.5 | 0.3 | 0.049 | -0.079 | 0.129 | 0.156 | -0.027 | 0.129 | 0.018 | 0.116 | 0.118 | 0.083 | 0.068 | 0.118 | 1.000 | 0.969 | 0.003 | 0.780 | 0.734 | 0.005 | 0.038 | 0.734 | 0.363 |
|  |  | 0.7 | 0.5 | 0.2 | 0.5 | -0.3 | -0.048 | -0.077 | 0.029 | -0.079 | 0.108 | 0.029 | 0.018 | 0.121 | 0.120 | 0.083 | 0.068 | 0.120 | 1.000 | 0.614 | 0.150 | 0.956 | 0.428 | 0.224 | 0.092 | 0.732 | 0.383 |
|  |  | -0.3 | 0.5 | -0.8 | -0.5 | -0.3 | 0.183 | -0.084 | 0.268 | 0.158 | 0.110 | 0.268 | 0.014 | 0.125 | 0.125 | 0.084 | 0.072 | 0.125 | 1.000 | 0.903 | 0.984 | 0.978 | 0.791 | 0.991 | 0.395 | 0.708 | 0.371 |
|  |  | 0 | -0.8 | 0.8 | 0.5 | 0.3 | -0.182 | -0.077 | -0.105 | -0.080 | -0.025 | -0.104 | 0.016 | 0.115 | 0.117 | 0.081 | 0.066 | 0.117 | 1.000 | 1.000 | 0.999 | 0.998 | 0.970 | 1.000 | 0.387 | 0.740 | 0.369 |
|  |  | -1.6 | -0.8 | -0.8 | -0.5 | -0.3 | 0.183 | -0.077 | 0.260 | 0.153 | 0.107 | 0.260 | 0.014 | 0.120 | 0.121 | 0.083 | 0.071 | 0.121 | 1.000 | 0.793 | 0.143 | 0.084 | 0.007 | 0.267 | 0.000 | 0.723 | 0.387 |
|  |  | 0.8 | 0 | 0.8 | 0.5 | 0.3 | -0.183 | -0.076 | -0.107 | -0.079 | -0.028 | -0.107 | 0.017 | 0.124 | 0.124 | 0.087 | 0.070 | 0.124 | 1.000 | 0.001 | 0.951 | 0.889 | 0.638 | 0.986 | 0.032 | 0.745 | 0.400 |
|  |  | 1.3 | 0.8 | 0.5 | 0.5 | 0 | -0.115 | -0.080 | -0.035 | -0.076 | 0.041 | -0.035 | 0.015 | 0.119 | 0.119 | 0.082 | 0.067 | 0.120 | 1.000 | 0.605 | 0.104 | 0.445 | 0.000 | 0.199 | 0.000 | 0.697 | 0.397 |
|  |  | 1.3 | 0.8 | 0.5 | 0.5 | 0 | 0.005 | -0.043 | 0.048 | 0.043 | 0.005 | 0.048 | 0.023 | 0.115 | 0.115 | 0.114 | 0.016 | 0.115 | 1.000 | 0.846 | 0.394 | 0.420 | 0.022 | 0.434 | 0.000 | 0.716 | 0.282 |

TE: total effect; DE: direct effect; IE: indirect effect; IE_d: indirect effect calculated by difference method; IE_p: indirect effect calculated by product method; IE1: *X*→*M_1_*→*Y*; IE2: *X*→*M_2_*→*Y*.

#### 8 Additional simulation scenario 2: Robustness of PSE-MR for unmeasured confounders of three relationships

Our method was robust for all sources of unmeasured confounders, because, as you mentioned, the IV was an IV for the exposure-mediator, exposure-outcome, and mediator-outcome relationships. We chose IV satisfied three assumptions and these IV was valid for the exposure. Thus PSE-MR was robust for all confounders of exposure-mediator and exposure-outcome relationships. In addition, a valid IV for the exposure was also valid for the mediator, thus PSE-MR was robust for unmeasured confounders of mediator-outcome relationship.

To validate the robustness of PSE-MR for unmeasured confounders of three relationships. We varied the magnitude of effect of unmeasured confounders on the risk factor, mediator and the outcome. The data generating process was the same as that in the Simulation A. Results revealed that PSE-MR were robust for all the three type of unmeasured confounders. Results were listed in **eTable 30-33**.

**eTable 30.** Simulation results of robustness of PE-Egger for unmeasured confounders between the risk factor and outcome

|  |  | TRUE | | BIAS | | | | SE | | | | Power | | | | | |
| --- | --- | --- | --- | --- | --- | --- | --- | --- | --- | --- | --- | --- | --- | --- | --- | --- | --- |
|  |  | uy | ux | TE | DE | IE_d | IE_p | TE | DE | IE_d | IE_p | TE | DE | IE_d | IE_p | GM | GY |
| Sc2 | IVW | -1 | -1 | 0.000 | 0.000 | 0.000 | 0.000 | 0.014 | 0.064 | 0.064 | 0.064 | 1.000 | 1.000 | 0.994 | 0.995 |  |  |
|  |  | -0.8 | -0.8 | 0.001 | 0.001 | 0.000 | 0.000 | 0.014 | 0.064 | 0.064 | 0.064 | 1.000 | 1.000 | 0.995 | 0.995 |  |  |
|  |  | -0.6 | -0.6 | 0.001 | 0.003 | -0.002 | -0.002 | 0.013 | 0.063 | 0.064 | 0.064 | 1.000 | 1.000 | 0.996 | 0.996 |  |  |
|  |  | -0.4 | -0.4 | 0.001 | 0.000 | 0.001 | 0.001 | 0.013 | 0.063 | 0.063 | 0.063 | 1.000 | 1.000 | 0.996 | 0.997 |  |  |
|  |  | -0.2 | -0.2 | 0.000 | -0.002 | 0.002 | 0.002 | 0.013 | 0.062 | 0.063 | 0.063 | 1.000 | 1.000 | 0.997 | 0.997 |  |  |
|  |  | 0 | 0 | 0.000 | -0.001 | 0.001 | 0.001 | 0.013 | 0.065 | 0.065 | 0.065 | 1.000 | 1.000 | 0.994 | 0.994 |  |  |
|  |  | 0.2 | 0.2 | 0.000 | -0.002 | 0.002 | 0.002 | 0.014 | 0.063 | 0.062 | 0.062 | 1.000 | 1.000 | 0.995 | 0.995 |  |  |
|  |  | 0.4 | 0.4 | 0.001 | 0.003 | -0.002 | -0.002 | 0.014 | 0.066 | 0.065 | 0.065 | 1.000 | 1.000 | 0.994 | 0.995 |  |  |
|  |  | 0.6 | 0.6 | 0.000 | -0.001 | 0.002 | 0.002 | 0.014 | 0.064 | 0.064 | 0.064 | 1.000 | 1.000 | 0.994 | 0.995 |  |  |
|  |  | 0.8 | 0.8 | 0.000 | 0.001 | -0.001 | -0.001 | 0.014 | 0.063 | 0.064 | 0.064 | 1.000 | 1.000 | 0.999 | 0.999 |  |  |
|  |  | 1 | 1 | 0.000 | 0.001 | -0.001 | -0.001 | 0.014 | 0.061 | 0.061 | 0.062 | 1.000 | 1.000 | 0.996 | 0.996 |  |  |
|  | Egger | -1 | -1 | 0.000 | 0.001 | 0.000 | 0.000 | 0.035 | 0.069 | 0.070 | 0.070 | 1.000 | 1.000 | 0.990 | 0.995 | 0.038 | 0.049 |
|  |  | -0.8 | -0.8 | 0.002 | 0.001 | 0.001 | 0.001 | 0.034 | 0.068 | 0.070 | 0.070 | 1.000 | 1.000 | 0.986 | 0.994 | 0.044 | 0.059 |
|  |  | -0.6 | -0.6 | 0.003 | 0.005 | -0.003 | -0.002 | 0.035 | 0.069 | 0.068 | 0.068 | 1.000 | 1.000 | 0.991 | 0.995 | 0.046 | 0.042 |
|  |  | -0.4 | -0.4 | 0.001 | 0.001 | 0.000 | 0.000 | 0.033 | 0.068 | 0.068 | 0.068 | 1.000 | 1.000 | 0.990 | 0.993 | 0.046 | 0.040 |
|  |  | -0.2 | -0.2 | 0.000 | -0.001 | 0.001 | 0.001 | 0.035 | 0.069 | 0.069 | 0.069 | 1.000 | 1.000 | 0.996 | 0.997 | 0.042 | 0.053 |
|  |  | 0 | 0 | 0.001 | -0.001 | 0.002 | 0.002 | 0.034 | 0.069 | 0.069 | 0.069 | 1.000 | 1.000 | 0.987 | 0.992 | 0.057 | 0.044 |
|  |  | 0.2 | 0.2 | 0.001 | -0.001 | 0.002 | 0.002 | 0.035 | 0.069 | 0.068 | 0.068 | 1.000 | 1.000 | 0.988 | 0.991 | 0.055 | 0.064 |
|  |  | 0.4 | 0.4 | 0.001 | 0.005 | -0.004 | -0.004 | 0.035 | 0.072 | 0.070 | 0.070 | 1.000 | 1.000 | 0.986 | 0.991 | 0.051 | 0.046 |
|  |  | 0.6 | 0.6 | 0.000 | -0.002 | 0.001 | 0.002 | 0.034 | 0.070 | 0.070 | 0.070 | 1.000 | 1.000 | 0.989 | 0.993 | 0.039 | 0.049 |
|  |  | 0.8 | 0.8 | 0.001 | 0.002 | -0.001 | -0.001 | 0.037 | 0.068 | 0.069 | 0.069 | 1.000 | 1.000 | 0.991 | 0.997 | 0.062 | 0.060 |
|  |  | 1 | 1 | 0.001 | 0.001 | 0.000 | 0.000 | 0.034 | 0.067 | 0.068 | 0.068 | 1.000 | 1.000 | 0.989 | 0.991 | 0.054 | 0.046 |
| Sc3 | IVW | -1 | -1 | 0.074 | -0.065 | 0.140 | 0.140 | 0.009 | 0.067 | 0.068 | 0.068 | 1.000 | 1.000 | 0.999 | 0.999 |  |  |
|  |  | -0.8 | -0.8 | 0.075 | -0.065 | 0.140 | 0.140 | 0.009 | 0.066 | 0.066 | 0.066 | 1.000 | 0.999 | 0.999 | 0.999 |  |  |
|  |  | -0.6 | -0.6 | 0.075 | -0.064 | 0.139 | 0.139 | 0.009 | 0.065 | 0.065 | 0.065 | 1.000 | 0.999 | 0.999 | 0.999 |  |  |
|  |  | -0.4 | -0.4 | 0.075 | -0.066 | 0.141 | 0.141 | 0.009 | 0.063 | 0.063 | 0.063 | 1.000 | 0.998 | 1.000 | 1.000 |  |  |
|  |  | -0.2 | -0.2 | 0.074 | -0.063 | 0.137 | 0.138 | 0.009 | 0.064 | 0.065 | 0.065 | 1.000 | 1.000 | 0.998 | 0.998 |  |  |
|  |  | 0 | 0 | 0.074 | -0.065 | 0.139 | 0.139 | 0.009 | 0.063 | 0.063 | 0.063 | 1.000 | 1.000 | 1.000 | 1.000 |  |  |
|  |  | 0.2 | 0.2 | 0.074 | -0.066 | 0.139 | 0.140 | 0.009 | 0.063 | 0.064 | 0.064 | 1.000 | 0.999 | 1.000 | 1.000 |  |  |
|  |  | 0.4 | 0.4 | 0.074 | -0.065 | 0.139 | 0.140 | 0.009 | 0.066 | 0.067 | 0.067 | 1.000 | 1.000 | 1.000 | 1.000 |  |  |
|  |  | 0.6 | 0.6 | 0.075 | -0.062 | 0.137 | 0.137 | 0.009 | 0.064 | 0.063 | 0.063 | 1.000 | 0.999 | 1.000 | 1.000 |  |  |
|  |  | 0.8 | 0.8 | 0.074 | -0.065 | 0.139 | 0.139 | 0.009 | 0.067 | 0.068 | 0.068 | 1.000 | 0.998 | 1.000 | 1.000 |  |  |
|  |  | 1 | 1 | 0.075 | -0.060 | 0.135 | 0.135 | 0.009 | 0.070 | 0.071 | 0.071 | 1.000 | 0.999 | 0.996 | 0.996 |  |  |
|  | Egger | -1 | -1 | 0.000 | -0.001 | 0.001 | 0.001 | 0.018 | 0.070 | 0.071 | 0.071 | 1.000 | 1.000 | 0.918 | 0.929 | 0.335 | 0.873 |
|  |  | -0.8 | -0.8 | 0.000 | 0.000 | 0.000 | 0.000 | 0.019 | 0.069 | 0.069 | 0.069 | 1.000 | 0.999 | 0.942 | 0.948 | 0.390 | 0.862 |
|  |  | -0.6 | -0.6 | 0.001 | 0.002 | -0.001 | -0.001 | 0.019 | 0.066 | 0.066 | 0.066 | 1.000 | 0.999 | 0.942 | 0.952 | 0.434 | 0.856 |
|  |  | -0.4 | -0.4 | 0.000 | 0.000 | 0.000 | 0.000 | 0.018 | 0.063 | 0.064 | 0.064 | 1.000 | 0.999 | 0.966 | 0.973 | 0.435 | 0.880 |
|  |  | -0.2 | -0.2 | 0.001 | 0.000 | 0.002 | 0.002 | 0.018 | 0.066 | 0.065 | 0.065 | 1.000 | 1.000 | 0.969 | 0.971 | 0.432 | 0.867 |
|  |  | 0 | 0 | 0.000 | -0.002 | 0.002 | 0.002 | 0.018 | 0.063 | 0.064 | 0.064 | 1.000 | 1.000 | 0.966 | 0.972 | 0.430 | 0.865 |
|  |  | 0.2 | 0.2 | 0.000 | 0.000 | 0.000 | 0.000 | 0.018 | 0.066 | 0.067 | 0.067 | 1.000 | 0.999 | 0.961 | 0.971 | 0.428 | 0.843 |
|  |  | 0.4 | 0.4 | 0.000 | 0.001 | 0.000 | 0.000 | 0.018 | 0.067 | 0.068 | 0.068 | 1.000 | 1.000 | 0.958 | 0.964 | 0.452 | 0.872 |
|  |  | 0.6 | 0.6 | 0.001 | 0.004 | -0.003 | -0.003 | 0.019 | 0.066 | 0.065 | 0.065 | 1.000 | 1.000 | 0.957 | 0.960 | 0.407 | 0.880 |
|  |  | 0.8 | 0.8 | 0.001 | 0.001 | 0.000 | 0.000 | 0.018 | 0.070 | 0.069 | 0.069 | 1.000 | 0.999 | 0.941 | 0.951 | 0.397 | 0.865 |
|  |  | 1 | 1 | 0.001 | 0.003 | -0.001 | -0.001 | 0.019 | 0.069 | 0.068 | 0.068 | 1.000 | 0.999 | 0.931 | 0.948 | 0.331 | 0.847 |
| Sc4 | IVW | -1 | -1 | -0.016 | 0.170 | -0.185 | -0.185 | 0.015 | 0.158 | 0.158 | 0.158 | 1.000 | 0.965 | 0.103 | 0.112 |  |  |
|  |  | -0.8 | -0.8 | 0.002 | 0.107 | -0.105 | -0.105 | 0.013 | 0.123 | 0.123 | 0.123 | 1.000 | 0.989 | 0.320 | 0.325 |  |  |
|  |  | -0.6 | -0.6 | 0.021 | 0.058 | -0.037 | -0.037 | 0.011 | 0.108 | 0.107 | 0.107 | 1.000 | 0.994 | 0.717 | 0.723 |  |  |
|  |  | -0.4 | -0.4 | 0.038 | 0.009 | 0.029 | 0.030 | 0.009 | 0.081 | 0.080 | 0.080 | 1.000 | 1.000 | 0.975 | 0.973 |  |  |
|  |  | -0.2 | -0.2 | 0.057 | -0.032 | 0.089 | 0.089 | 0.008 | 0.067 | 0.068 | 0.068 | 1.000 | 1.000 | 0.999 | 0.999 |  |  |
|  |  | 0 | 0 | 0.074 | -0.069 | 0.143 | 0.143 | 0.009 | 0.065 | 0.066 | 0.066 | 1.000 | 1.000 | 1.000 | 1.000 |  |  |
|  |  | 0.2 | 0.2 | 0.092 | -0.090 | 0.182 | 0.182 | 0.010 | 0.078 | 0.079 | 0.079 | 1.000 | 0.995 | 0.995 | 0.995 |  |  |
|  |  | 0.4 | 0.4 | 0.108 | -0.111 | 0.219 | 0.220 | 0.011 | 0.093 | 0.094 | 0.094 | 1.000 | 0.959 | 0.995 | 0.996 |  |  |
|  |  | 0.6 | 0.6 | 0.126 | -0.130 | 0.255 | 0.255 | 0.013 | 0.121 | 0.121 | 0.121 | 1.000 | 0.803 | 0.982 | 0.984 |  |  |
|  |  | 0.8 | 0.8 | 0.141 | -0.134 | 0.276 | 0.276 | 0.014 | 0.144 | 0.144 | 0.144 | 1.000 | 0.639 | 0.953 | 0.959 |  |  |
|  |  | 1 | 1 | 0.158 | -0.141 | 0.299 | 0.299 | 0.015 | 0.172 | 0.173 | 0.173 | 1.000 | 0.486 | 0.904 | 0.906 |  |  |
|  | Egger | -1 | -1 | 0.020 | 0.017 | 0.002 | 0.002 | 0.034 | 0.172 | 0.170 | 0.170 | 1.000 | 0.840 | 0.425 | 0.455 | 0.538 | 0.925 |
|  |  | -0.8 | -0.8 | 0.012 | 0.012 | 0.000 | 0.000 | 0.029 | 0.139 | 0.138 | 0.138 | 1.000 | 0.923 | 0.571 | 0.607 | 0.350 | 0.903 |
|  |  | -0.6 | -0.6 | 0.008 | 0.011 | -0.003 | -0.003 | 0.026 | 0.114 | 0.112 | 0.112 | 1.000 | 0.985 | 0.741 | 0.763 | 0.153 | 0.898 |
|  |  | -0.4 | -0.4 | 0.003 | 0.003 | -0.001 | -0.001 | 0.021 | 0.090 | 0.089 | 0.089 | 1.000 | 0.999 | 0.889 | 0.901 | 0.036 | 0.904 |
|  |  | -0.2 | -0.2 | 0.001 | 0.002 | -0.001 | 0.000 | 0.019 | 0.074 | 0.075 | 0.075 | 1.000 | 0.999 | 0.951 | 0.959 | 0.107 | 0.890 |
|  |  | 0 | 0 | 0.000 | -0.005 | 0.005 | 0.005 | 0.018 | 0.066 | 0.066 | 0.066 | 1.000 | 1.000 | 0.966 | 0.972 | 0.431 | 0.854 |
|  |  | 0.2 | 0.2 | 0.002 | 0.000 | 0.002 | 0.002 | 0.019 | 0.071 | 0.072 | 0.072 | 1.000 | 1.000 | 0.945 | 0.958 | 0.793 | 0.838 |
|  |  | 0.4 | 0.4 | 0.003 | 0.008 | -0.005 | -0.005 | 0.020 | 0.092 | 0.091 | 0.091 | 1.000 | 0.999 | 0.853 | 0.881 | 0.896 | 0.835 |
|  |  | 0.6 | 0.6 | 0.008 | 0.007 | 0.001 | 0.001 | 0.025 | 0.118 | 0.115 | 0.115 | 1.000 | 0.986 | 0.730 | 0.752 | 0.896 | 0.830 |
|  |  | 0.8 | 0.8 | 0.014 | 0.019 | -0.004 | -0.004 | 0.030 | 0.141 | 0.137 | 0.137 | 1.000 | 0.942 | 0.552 | 0.589 | 0.861 | 0.816 |
|  |  | 1 | 1 | 0.023 | 0.020 | 0.003 | 0.003 | 0.034 | 0.169 | 0.166 | 0.166 | 1.000 | 0.840 | 0.445 | 0.477 | 0.807 | 0.798 |

TE: total effect; DE: direct effect; IE: indirect effect; IE_d: indirect effect calculated by difference method; IE_p: indirect effect calculated by product method.

**eTable 31.** Simulation results of robustness of PE-Egger for unmeasured confounders between the risk factor and mediator

|  |  | TRUE | | BIAS | | | | SE | | | | Power | | | | | |
| --- | --- | --- | --- | --- | --- | --- | --- | --- | --- | --- | --- | --- | --- | --- | --- | --- | --- |
|  |  | um | ux | TE | DE | IE_d | IE_p | TE | DE | IE_d | IE_p | TE | DE | IE_d | IE_p | GM | GY |
| Sc2 | IVW | -1 | -1 | 0.000 | 0.001 | -0.001 | -0.001 | 0.014 | 0.063 | 0.063 | 0.063 | 1.000 | 1.000 | 0.999 | 0.998 |  |  |
|  |  | -0.8 | -0.8 | 0.001 | 0.003 | -0.002 | -0.002 | 0.014 | 0.063 | 0.063 | 0.063 | 1.000 | 1.000 | 0.997 | 0.997 |  |  |
|  |  | -0.6 | -0.6 | 0.000 | 0.000 | 0.000 | 0.000 | 0.014 | 0.062 | 0.063 | 0.063 | 1.000 | 1.000 | 0.997 | 0.997 |  |  |
|  |  | -0.4 | -0.4 | 0.000 | -0.001 | 0.002 | 0.002 | 0.014 | 0.064 | 0.064 | 0.064 | 1.000 | 1.000 | 0.997 | 0.995 |  |  |
|  |  | -0.2 | -0.2 | 0.001 | 0.000 | 0.000 | 0.000 | 0.013 | 0.064 | 0.064 | 0.064 | 1.000 | 1.000 | 0.997 | 0.998 |  |  |
|  |  | 0 | 0 | 0.001 | -0.001 | 0.002 | 0.002 | 0.014 | 0.063 | 0.063 | 0.063 | 1.000 | 1.000 | 0.996 | 0.996 |  |  |
|  |  | 0.2 | 0.2 | 0.001 | 0.000 | 0.000 | 0.001 | 0.014 | 0.061 | 0.061 | 0.061 | 1.000 | 1.000 | 0.998 | 0.998 |  |  |
|  |  | 0.4 | 0.4 | 0.000 | -0.001 | 0.001 | 0.001 | 0.014 | 0.064 | 0.065 | 0.065 | 1.000 | 1.000 | 0.997 | 0.997 |  |  |
|  |  | 0.6 | 0.6 | 0.000 | 0.001 | 0.000 | 0.000 | 0.014 | 0.065 | 0.065 | 0.065 | 1.000 | 1.000 | 0.994 | 0.994 |  |  |
|  |  | 0.8 | 0.8 | 0.001 | 0.004 | -0.002 | -0.002 | 0.014 | 0.061 | 0.061 | 0.061 | 1.000 | 1.000 | 0.997 | 0.999 |  |  |
|  |  | 1 | 1 | 0.000 | 0.003 | -0.003 | -0.003 | 0.014 | 0.062 | 0.062 | 0.062 | 1.000 | 1.000 | 0.998 | 0.998 |  |  |
|  | Egger | -1 | -1 | 0.002 | 0.002 | 0.000 | 0.000 | 0.034 | 0.068 | 0.069 | 0.069 | 1.000 | 1.000 | 0.993 | 0.996 | 0.039 | 0.050 |
|  |  | -0.8 | -0.8 | 0.003 | 0.004 | -0.002 | -0.002 | 0.035 | 0.068 | 0.069 | 0.069 | 1.000 | 1.000 | 0.987 | 0.995 | 0.055 | 0.043 |
|  |  | -0.6 | -0.6 | 0.002 | 0.002 | 0.000 | 0.000 | 0.034 | 0.068 | 0.068 | 0.068 | 1.000 | 1.000 | 0.990 | 0.994 | 0.043 | 0.051 |
|  |  | -0.4 | -0.4 | 0.002 | -0.001 | 0.002 | 0.002 | 0.035 | 0.072 | 0.070 | 0.070 | 1.000 | 1.000 | 0.991 | 0.995 | 0.048 | 0.052 |
|  |  | -0.2 | -0.2 | 0.001 | 0.001 | 0.000 | 0.000 | 0.035 | 0.070 | 0.072 | 0.072 | 1.000 | 1.000 | 0.991 | 0.995 | 0.041 | 0.050 |
|  |  | 0 | 0 | 0.001 | -0.001 | 0.001 | 0.002 | 0.034 | 0.069 | 0.069 | 0.069 | 1.000 | 1.000 | 0.986 | 0.994 | 0.049 | 0.058 |
|  |  | 0.2 | 0.2 | 0.002 | 0.001 | 0.001 | 0.001 | 0.034 | 0.068 | 0.066 | 0.066 | 1.000 | 1.000 | 0.994 | 0.996 | 0.048 | 0.050 |
|  |  | 0.4 | 0.4 | 0.000 | -0.001 | 0.001 | 0.002 | 0.033 | 0.071 | 0.070 | 0.070 | 1.000 | 1.000 | 0.992 | 0.995 | 0.047 | 0.038 |
|  |  | 0.6 | 0.6 | 0.001 | 0.002 | 0.000 | 0.000 | 0.034 | 0.070 | 0.069 | 0.069 | 1.000 | 1.000 | 0.989 | 0.993 | 0.043 | 0.052 |
|  |  | 0.8 | 0.8 | 0.001 | 0.004 | -0.002 | -0.002 | 0.034 | 0.066 | 0.067 | 0.067 | 1.000 | 1.000 | 0.990 | 0.996 | 0.050 | 0.048 |
|  |  | 1 | 1 | 0.001 | 0.002 | -0.001 | -0.001 | 0.033 | 0.067 | 0.068 | 0.068 | 1.000 | 1.000 | 0.993 | 0.996 | 0.046 | 0.041 |
| Sc3 | IVW | -1 | -1 | 0.074 | -0.058 | 0.132 | 0.132 | 0.009 | 0.066 | 0.067 | 0.067 | 1.000 | 0.999 | 1.000 | 0.999 |  |  |
|  |  | -0.8 | -0.8 | 0.075 | -0.059 | 0.134 | 0.134 | 0.009 | 0.064 | 0.065 | 0.065 | 1.000 | 0.999 | 0.999 | 0.999 |  |  |
|  |  | -0.6 | -0.6 | 0.075 | -0.065 | 0.140 | 0.140 | 0.009 | 0.063 | 0.064 | 0.064 | 1.000 | 1.000 | 1.000 | 1.000 |  |  |
|  |  | -0.4 | -0.4 | 0.074 | -0.065 | 0.139 | 0.139 | 0.009 | 0.064 | 0.065 | 0.065 | 1.000 | 1.000 | 0.999 | 0.999 |  |  |
|  |  | -0.2 | -0.2 | 0.074 | -0.063 | 0.137 | 0.137 | 0.009 | 0.066 | 0.067 | 0.067 | 1.000 | 0.998 | 1.000 | 1.000 |  |  |
|  |  | 0 | 0 | 0.074 | -0.064 | 0.139 | 0.139 | 0.009 | 0.064 | 0.064 | 0.064 | 1.000 | 1.000 | 0.999 | 0.999 |  |  |
|  |  | 0.2 | 0.2 | 0.075 | -0.067 | 0.141 | 0.141 | 0.009 | 0.066 | 0.065 | 0.065 | 1.000 | 0.999 | 0.999 | 0.999 |  |  |
|  |  | 0.4 | 0.4 | 0.074 | -0.063 | 0.137 | 0.138 | 0.009 | 0.064 | 0.065 | 0.065 | 1.000 | 0.999 | 1.000 | 1.000 |  |  |
|  |  | 0.6 | 0.6 | 0.074 | -0.062 | 0.136 | 0.136 | 0.009 | 0.066 | 0.065 | 0.065 | 1.000 | 0.999 | 0.999 | 0.999 |  |  |
|  |  | 0.8 | 0.8 | 0.074 | -0.059 | 0.133 | 0.133 | 0.009 | 0.067 | 0.067 | 0.067 | 1.000 | 1.000 | 0.999 | 0.998 |  |  |
|  |  | 1 | 1 | 0.075 | -0.054 | 0.129 | 0.129 | 0.009 | 0.061 | 0.062 | 0.062 | 1.000 | 1.000 | 1.000 | 1.000 |  |  |
|  | Egger | -1 | -1 | 0.001 | 0.000 | 0.001 | 0.001 | 0.019 | 0.066 | 0.066 | 0.066 | 1.000 | 0.999 | 0.952 | 0.962 | 0.377 | 0.815 |
|  |  | -0.8 | -0.8 | 0.002 | 0.001 | 0.001 | 0.001 | 0.018 | 0.064 | 0.065 | 0.065 | 1.000 | 1.000 | 0.956 | 0.969 | 0.377 | 0.842 |
|  |  | -0.6 | -0.6 | 0.000 | -0.003 | 0.002 | 0.003 | 0.018 | 0.064 | 0.063 | 0.063 | 1.000 | 1.000 | 0.964 | 0.972 | 0.418 | 0.847 |
|  |  | -0.4 | -0.4 | 0.000 | 0.000 | 0.000 | 0.000 | 0.018 | 0.064 | 0.064 | 0.064 | 1.000 | 1.000 | 0.971 | 0.974 | 0.435 | 0.870 |
|  |  | -0.2 | -0.2 | 0.001 | 0.002 | -0.001 | -0.001 | 0.018 | 0.068 | 0.068 | 0.068 | 1.000 | 0.999 | 0.950 | 0.959 | 0.469 | 0.851 |
|  |  | 0 | 0 | 0.000 | 0.001 | 0.000 | 0.000 | 0.018 | 0.066 | 0.067 | 0.067 | 1.000 | 1.000 | 0.956 | 0.961 | 0.435 | 0.869 |
|  |  | 0.2 | 0.2 | 0.001 | -0.001 | 0.002 | 0.002 | 0.018 | 0.066 | 0.066 | 0.066 | 1.000 | 1.000 | 0.968 | 0.975 | 0.443 | 0.860 |
|  |  | 0.4 | 0.4 | 0.001 | -0.001 | 0.002 | 0.002 | 0.019 | 0.065 | 0.066 | 0.066 | 1.000 | 1.000 | 0.966 | 0.967 | 0.415 | 0.853 |
|  |  | 0.6 | 0.6 | 0.001 | 0.000 | 0.002 | 0.002 | 0.019 | 0.065 | 0.065 | 0.065 | 1.000 | 1.000 | 0.964 | 0.969 | 0.409 | 0.827 |
|  |  | 0.8 | 0.8 | 0.000 | 0.001 | -0.001 | -0.001 | 0.019 | 0.066 | 0.066 | 0.066 | 1.000 | 1.000 | 0.946 | 0.955 | 0.403 | 0.834 |
|  |  | 1 | 1 | 0.001 | 0.001 | 0.000 | 0.000 | 0.018 | 0.060 | 0.060 | 0.060 | 1.000 | 1.000 | 0.965 | 0.973 | 0.380 | 0.815 |
| Sc4 | IVW | -1 | -1 | -0.015 | 0.069 | -0.084 | -0.085 | 0.016 | 0.022 | 0.025 | 0.025 | 1.000 | 1.000 | 1.000 | 1.000 |  |  |
|  |  | -0.8 | -0.8 | 0.003 | 0.071 | -0.068 | -0.068 | 0.013 | 0.028 | 0.029 | 0.029 | 1.000 | 1.000 | 1.000 | 1.000 |  |  |
|  |  | -0.6 | -0.6 | 0.020 | 0.067 | -0.046 | -0.046 | 0.011 | 0.039 | 0.039 | 0.039 | 1.000 | 1.000 | 1.000 | 1.000 |  |  |
|  |  | -0.4 | -0.4 | 0.039 | 0.045 | -0.006 | -0.006 | 0.009 | 0.058 | 0.058 | 0.058 | 1.000 | 1.000 | 0.996 | 0.995 |  |  |
|  |  | -0.2 | -0.2 | 0.057 | -0.019 | 0.075 | 0.075 | 0.008 | 0.069 | 0.069 | 0.069 | 1.000 | 1.000 | 0.997 | 0.996 |  |  |
|  |  | 0 | 0 | 0.074 | -0.069 | 0.143 | 0.143 | 0.009 | 0.065 | 0.066 | 0.066 | 1.000 | 1.000 | 0.999 | 0.999 |  |  |
|  |  | 0.2 | 0.2 | 0.091 | -0.068 | 0.159 | 0.159 | 0.010 | 0.055 | 0.055 | 0.055 | 1.000 | 1.000 | 1.000 | 1.000 |  |  |
|  |  | 0.4 | 0.4 | 0.109 | -0.051 | 0.159 | 0.160 | 0.011 | 0.047 | 0.049 | 0.049 | 1.000 | 1.000 | 1.000 | 1.000 |  |  |
|  |  | 0.6 | 0.6 | 0.126 | -0.035 | 0.161 | 0.161 | 0.013 | 0.041 | 0.042 | 0.042 | 1.000 | 1.000 | 1.000 | 1.000 |  |  |
|  |  | 0.8 | 0.8 | 0.141 | -0.022 | 0.164 | 0.164 | 0.014 | 0.036 | 0.038 | 0.038 | 1.000 | 1.000 | 1.000 | 1.000 |  |  |
|  |  | 1 | 1 | 0.158 | -0.014 | 0.172 | 0.172 | 0.015 | 0.030 | 0.033 | 0.033 | 1.000 | 1.000 | 1.000 | 1.000 |  |  |
|  | Egger | -1 | -1 | 0.020 | 0.000 | 0.020 | 0.020 | 0.034 | 0.029 | 0.041 | 0.041 | 1.000 | 1.000 | 1.000 | 1.000 | 0.619 | 0.710 |
|  |  | -0.8 | -0.8 | 0.012 | 0.001 | 0.011 | 0.011 | 0.029 | 0.034 | 0.040 | 0.040 | 1.000 | 1.000 | 1.000 | 1.000 | 0.682 | 0.481 |
|  |  | -0.6 | -0.6 | 0.006 | 0.000 | 0.007 | 0.007 | 0.026 | 0.039 | 0.043 | 0.043 | 1.000 | 1.000 | 1.000 | 1.000 | 0.718 | 0.258 |
|  |  | -0.4 | -0.4 | 0.003 | 0.000 | 0.002 | 0.002 | 0.022 | 0.049 | 0.050 | 0.050 | 1.000 | 1.000 | 0.999 | 0.999 | 0.730 | 0.062 |
|  |  | -0.2 | -0.2 | 0.001 | -0.003 | 0.004 | 0.004 | 0.019 | 0.060 | 0.060 | 0.060 | 1.000 | 1.000 | 0.992 | 0.995 | 0.622 | 0.296 |
|  |  | 0 | 0 | 0.000 | -0.004 | 0.004 | 0.004 | 0.018 | 0.066 | 0.066 | 0.066 | 1.000 | 1.000 | 0.964 | 0.969 | 0.413 | 0.883 |
|  |  | 0.2 | 0.2 | 0.003 | -0.001 | 0.004 | 0.004 | 0.019 | 0.059 | 0.059 | 0.059 | 1.000 | 1.000 | 0.980 | 0.987 | 0.292 | 0.978 |
|  |  | 0.4 | 0.4 | 0.005 | -0.002 | 0.007 | 0.007 | 0.022 | 0.050 | 0.051 | 0.051 | 1.000 | 1.000 | 0.993 | 0.996 | 0.254 | 0.984 |
|  |  | 0.6 | 0.6 | 0.008 | 0.000 | 0.008 | 0.008 | 0.025 | 0.042 | 0.045 | 0.045 | 1.000 | 1.000 | 1.000 | 1.000 | 0.216 | 0.971 |
|  |  | 0.8 | 0.8 | 0.013 | 0.000 | 0.012 | 0.012 | 0.029 | 0.035 | 0.043 | 0.043 | 1.000 | 1.000 | 1.000 | 1.000 | 0.192 | 0.951 |
|  |  | 1 | 1 | 0.020 | 0.000 | 0.020 | 0.020 | 0.034 | 0.029 | 0.041 | 0.041 | 1.000 | 1.000 | 1.000 | 1.000 | 0.165 | 0.932 |

TE: total effect; DE: direct effect; IE: indirect effect; IE_d: indirect effect calculated by difference method; IE_p: indirect effect calculated by product method.

**eTable 32.** Simulation results of robustness of PE-Egger for unmeasured confounders between the mediator and outcome

|  |  | TRUE | | BIAS | | | | SE | | | | Power | | | | | |
| --- | --- | --- | --- | --- | --- | --- | --- | --- | --- | --- | --- | --- | --- | --- | --- | --- | --- |
|  |  | um | uy | TE | DE | IE_d | IE_p | TE | DE | IE_d | IE_p | TE | DE | IE_d | IE_p | GM | GY |
| Sc2 | IVW | -1 | -1 | 0.000 | -0.009 | 0.009 | 0.009 | 0.015 | 0.064 | 0.064 | 0.064 | 1.000 | 1.000 | 0.998 | 0.997 |  |  |
|  |  | -0.8 | -0.8 | 0.000 | -0.007 | 0.007 | 0.007 | 0.014 | 0.064 | 0.064 | 0.064 | 1.000 | 1.000 | 0.998 | 0.997 |  |  |
|  |  | -0.6 | -0.6 | 0.000 | -0.005 | 0.005 | 0.005 | 0.014 | 0.061 | 0.061 | 0.061 | 1.000 | 1.000 | 0.997 | 0.999 |  |  |
|  |  | -0.4 | -0.4 | 0.000 | -0.002 | 0.002 | 0.002 | 0.014 | 0.061 | 0.061 | 0.061 | 1.000 | 1.000 | 0.997 | 0.997 |  |  |
|  |  | -0.2 | -0.2 | 0.000 | 0.002 | -0.002 | -0.002 | 0.013 | 0.063 | 0.063 | 0.063 | 1.000 | 1.000 | 0.992 | 0.993 |  |  |
|  |  | 0 | 0 | 0.000 | -0.002 | 0.002 | 0.002 | 0.014 | 0.063 | 0.062 | 0.062 | 1.000 | 1.000 | 0.994 | 0.994 |  |  |
|  |  | 0.2 | 0.2 | 0.001 | -0.001 | 0.002 | 0.002 | 0.014 | 0.062 | 0.062 | 0.062 | 1.000 | 1.000 | 0.996 | 0.997 |  |  |
|  |  | 0.4 | 0.4 | 0.000 | -0.001 | 0.002 | 0.002 | 0.013 | 0.064 | 0.064 | 0.064 | 1.000 | 1.000 | 0.994 | 0.994 |  |  |
|  |  | 0.6 | 0.6 | 0.001 | -0.004 | 0.005 | 0.006 | 0.014 | 0.064 | 0.065 | 0.065 | 1.000 | 1.000 | 0.995 | 0.996 |  |  |
|  |  | 0.8 | 0.8 | 0.000 | -0.005 | 0.006 | 0.006 | 0.014 | 0.063 | 0.063 | 0.063 | 1.000 | 1.000 | 0.998 | 0.997 |  |  |
|  |  | 1 | 1 | 0.000 | -0.012 | 0.012 | 0.012 | 0.014 | 0.066 | 0.066 | 0.066 | 1.000 | 1.000 | 0.999 | 1.000 |  |  |
|  | Egger | -1 | -1 | 0.001 | -0.011 | 0.011 | 0.011 | 0.034 | 0.069 | 0.071 | 0.071 | 1.000 | 1.000 | 0.992 | 0.997 | 0.044 | 0.049 |
|  |  | -0.8 | -0.8 | 0.000 | -0.007 | 0.007 | 0.007 | 0.035 | 0.069 | 0.070 | 0.070 | 1.000 | 1.000 | 0.995 | 0.998 | 0.051 | 0.055 |
|  |  | -0.6 | -0.6 | 0.001 | -0.003 | 0.005 | 0.005 | 0.035 | 0.067 | 0.066 | 0.066 | 1.000 | 1.000 | 0.990 | 0.994 | 0.052 | 0.046 |
|  |  | -0.4 | -0.4 | 0.000 | -0.002 | 0.001 | 0.001 | 0.033 | 0.065 | 0.066 | 0.066 | 1.000 | 1.000 | 0.993 | 0.996 | 0.045 | 0.045 |
|  |  | -0.2 | -0.2 | -0.001 | 0.001 | -0.002 | -0.002 | 0.034 | 0.068 | 0.069 | 0.069 | 1.000 | 1.000 | 0.988 | 0.992 | 0.052 | 0.054 |
|  |  | 0 | 0 | 0.000 | -0.003 | 0.002 | 0.002 | 0.035 | 0.068 | 0.070 | 0.070 | 1.000 | 1.000 | 0.992 | 0.993 | 0.053 | 0.040 |
|  |  | 0.2 | 0.2 | 0.000 | -0.003 | 0.002 | 0.002 | 0.033 | 0.066 | 0.067 | 0.067 | 1.000 | 1.000 | 0.993 | 0.997 | 0.043 | 0.043 |
|  |  | 0.4 | 0.4 | 0.000 | -0.002 | 0.002 | 0.002 | 0.033 | 0.068 | 0.069 | 0.069 | 1.000 | 1.000 | 0.990 | 0.995 | 0.052 | 0.046 |
|  |  | 0.6 | 0.6 | 0.002 | -0.005 | 0.007 | 0.007 | 0.035 | 0.068 | 0.071 | 0.071 | 1.000 | 1.000 | 0.986 | 0.992 | 0.046 | 0.047 |
|  |  | 0.8 | 0.8 | 0.001 | -0.006 | 0.007 | 0.007 | 0.034 | 0.067 | 0.069 | 0.069 | 1.000 | 1.000 | 0.989 | 0.995 | 0.044 | 0.054 |
|  |  | 1 | 1 | 0.001 | -0.012 | 0.012 | 0.012 | 0.035 | 0.071 | 0.070 | 0.070 | 1.000 | 1.000 | 0.997 | 0.999 | 0.048 | 0.053 |
| Sc3 | IVW | -1 | -1 | 0.074 | -0.084 | 0.158 | 0.158 | 0.010 | 0.064 | 0.064 | 0.064 | 1.000 | 1.000 | 1.000 | 1.000 |  |  |
|  |  | -0.8 | -0.8 | 0.074 | -0.079 | 0.153 | 0.153 | 0.009 | 0.067 | 0.067 | 0.067 | 1.000 | 0.999 | 0.999 | 0.999 |  |  |
|  |  | -0.6 | -0.6 | 0.074 | -0.070 | 0.144 | 0.144 | 0.009 | 0.064 | 0.064 | 0.064 | 1.000 | 1.000 | 1.000 | 1.000 |  |  |
|  |  | -0.4 | -0.4 | 0.074 | -0.067 | 0.141 | 0.141 | 0.009 | 0.065 | 0.065 | 0.065 | 1.000 | 0.998 | 1.000 | 1.000 |  |  |
|  |  | -0.2 | -0.2 | 0.074 | -0.068 | 0.142 | 0.142 | 0.009 | 0.065 | 0.065 | 0.065 | 1.000 | 1.000 | 1.000 | 1.000 |  |  |
|  |  | 0 | 0 | 0.075 | -0.062 | 0.137 | 0.137 | 0.009 | 0.066 | 0.067 | 0.067 | 1.000 | 1.000 | 0.998 | 0.999 |  |  |
|  |  | 0.2 | 0.2 | 0.074 | -0.070 | 0.144 | 0.144 | 0.009 | 0.065 | 0.065 | 0.065 | 1.000 | 1.000 | 0.999 | 0.999 |  |  |
|  |  | 0.4 | 0.4 | 0.074 | -0.072 | 0.147 | 0.147 | 0.009 | 0.063 | 0.064 | 0.064 | 1.000 | 1.000 | 0.999 | 0.999 |  |  |
|  |  | 0.6 | 0.6 | 0.074 | -0.073 | 0.148 | 0.148 | 0.009 | 0.063 | 0.063 | 0.063 | 1.000 | 1.000 | 1.000 | 1.000 |  |  |
|  |  | 0.8 | 0.8 | 0.075 | -0.078 | 0.152 | 0.152 | 0.009 | 0.065 | 0.065 | 0.065 | 1.000 | 0.999 | 0.999 | 0.999 |  |  |
|  |  | 1 | 1 | 0.074 | -0.087 | 0.162 | 0.162 | 0.010 | 0.067 | 0.067 | 0.067 | 1.000 | 0.997 | 0.999 | 0.999 |  |  |
|  | Egger | -1 | -1 | 0.000 | -0.032 | 0.032 | 0.032 | 0.020 | 0.064 | 0.065 | 0.065 | 1.000 | 1.000 | 0.980 | 0.980 | 0.242 | 0.826 |
|  |  | -0.8 | -0.8 | 0.000 | -0.022 | 0.022 | 0.022 | 0.019 | 0.068 | 0.068 | 0.068 | 1.000 | 1.000 | 0.966 | 0.971 | 0.317 | 0.855 |
|  |  | -0.6 | -0.6 | 0.001 | -0.011 | 0.012 | 0.012 | 0.018 | 0.066 | 0.066 | 0.066 | 1.000 | 1.000 | 0.963 | 0.969 | 0.341 | 0.845 |
|  |  | -0.4 | -0.4 | 0.002 | -0.006 | 0.008 | 0.008 | 0.018 | 0.067 | 0.068 | 0.068 | 1.000 | 0.999 | 0.963 | 0.970 | 0.401 | 0.850 |
|  |  | -0.2 | -0.2 | 0.000 | -0.002 | 0.003 | 0.003 | 0.018 | 0.067 | 0.067 | 0.067 | 1.000 | 1.000 | 0.958 | 0.967 | 0.434 | 0.870 |
|  |  | 0 | 0 | 0.001 | 0.002 | 0.000 | 0.000 | 0.019 | 0.064 | 0.064 | 0.064 | 1.000 | 1.000 | 0.966 | 0.971 | 0.439 | 0.853 |
|  |  | 0.2 | 0.2 | 0.000 | -0.005 | 0.005 | 0.005 | 0.018 | 0.067 | 0.067 | 0.067 | 1.000 | 0.999 | 0.959 | 0.970 | 0.434 | 0.872 |
|  |  | 0.4 | 0.4 | 0.001 | -0.009 | 0.010 | 0.010 | 0.018 | 0.065 | 0.065 | 0.065 | 1.000 | 1.000 | 0.967 | 0.974 | 0.405 | 0.859 |
|  |  | 0.6 | 0.6 | 0.001 | -0.013 | 0.014 | 0.014 | 0.019 | 0.065 | 0.066 | 0.066 | 1.000 | 1.000 | 0.965 | 0.967 | 0.377 | 0.824 |
|  |  | 0.8 | 0.8 | 0.001 | -0.022 | 0.023 | 0.023 | 0.020 | 0.066 | 0.066 | 0.066 | 1.000 | 1.000 | 0.965 | 0.972 | 0.303 | 0.825 |
|  |  | 1 | 1 | -0.001 | -0.034 | 0.033 | 0.033 | 0.021 | 0.068 | 0.068 | 0.068 | 1.000 | 0.998 | 0.972 | 0.978 | 0.277 | 0.838 |
| Sc4 | IVW | -1 | -1 | -0.110 | -0.268 | 0.158 | 0.158 | 0.027 | 0.025 | 0.035 | 0.035 | 1.000 | 1.000 | 1.000 | 1.000 |  |  |
|  |  | -0.8 | -0.8 | -0.074 | -0.249 | 0.176 | 0.175 | 0.021 | 0.033 | 0.038 | 0.038 | 1.000 | 1.000 | 1.000 | 1.000 |  |  |
|  |  | -0.6 | -0.6 | -0.036 | -0.212 | 0.175 | 0.175 | 0.016 | 0.040 | 0.042 | 0.042 | 1.000 | 1.000 | 1.000 | 1.000 |  |  |
|  |  | -0.4 | -0.4 | 0.000 | -0.144 | 0.144 | 0.144 | 0.012 | 0.050 | 0.051 | 0.051 | 1.000 | 1.000 | 1.000 | 1.000 |  |  |
|  |  | -0.2 | -0.2 | 0.038 | -0.064 | 0.101 | 0.101 | 0.009 | 0.061 | 0.061 | 0.061 | 1.000 | 1.000 | 1.000 | 1.000 |  |  |
|  |  | 0 | 0 | 0.074 | -0.063 | 0.137 | 0.138 | 0.009 | 0.065 | 0.065 | 0.065 | 1.000 | 1.000 | 1.000 | 1.000 |  |  |
|  |  | 0.2 | 0.2 | 0.111 | -0.140 | 0.251 | 0.251 | 0.011 | 0.059 | 0.060 | 0.060 | 1.000 | 0.998 | 1.000 | 1.000 |  |  |
|  |  | 0.4 | 0.4 | 0.148 | -0.196 | 0.344 | 0.344 | 0.015 | 0.051 | 0.053 | 0.053 | 1.000 | 0.996 | 1.000 | 1.000 |  |  |
|  |  | 0.6 | 0.6 | 0.187 | -0.230 | 0.417 | 0.417 | 0.021 | 0.044 | 0.049 | 0.049 | 1.000 | 0.997 | 1.000 | 1.000 |  |  |
|  |  | 0.8 | 0.8 | 0.224 | -0.253 | 0.477 | 0.477 | 0.026 | 0.040 | 0.047 | 0.047 | 1.000 | 0.998 | 1.000 | 1.000 |  |  |
|  |  | 1 | 1 | 0.259 | -0.265 | 0.524 | 0.524 | 0.030 | 0.037 | 0.047 | 0.047 | 1.000 | 0.999 | 1.000 | 1.000 |  |  |
|  | Egger | -1 | -1 | 0.001 | -0.256 | 0.257 | 0.257 | 0.062 | 0.038 | 0.067 | 0.067 | 1.000 | 0.999 | 1.000 | 1.000 | 0.070 | 0.428 |
|  |  | -0.8 | -0.8 | 0.001 | -0.238 | 0.239 | 0.239 | 0.052 | 0.043 | 0.062 | 0.062 | 1.000 | 0.998 | 1.000 | 1.000 | 0.056 | 0.316 |
|  |  | -0.6 | -0.6 | 0.001 | -0.202 | 0.203 | 0.203 | 0.040 | 0.048 | 0.060 | 0.060 | 1.000 | 1.000 | 1.000 | 1.000 | 0.056 | 0.147 |
|  |  | -0.4 | -0.4 | 0.000 | -0.144 | 0.143 | 0.143 | 0.030 | 0.054 | 0.058 | 0.058 | 1.000 | 1.000 | 1.000 | 1.000 | 0.018 | 0.039 |
|  |  | -0.2 | -0.2 | 0.001 | -0.057 | 0.058 | 0.058 | 0.022 | 0.062 | 0.063 | 0.063 | 1.000 | 1.000 | 0.996 | 0.998 | 0.111 | 0.291 |
|  |  | 0 | 0 | 0.001 | 0.000 | 0.001 | 0.001 | 0.018 | 0.063 | 0.063 | 0.063 | 1.000 | 1.000 | 0.975 | 0.982 | 0.461 | 0.849 |
|  |  | 0.2 | 0.2 | 0.001 | -0.055 | 0.056 | 0.056 | 0.022 | 0.065 | 0.064 | 0.064 | 1.000 | 1.000 | 0.993 | 0.994 | 0.484 | 0.982 |
|  |  | 0.4 | 0.4 | 0.002 | -0.140 | 0.141 | 0.142 | 0.029 | 0.055 | 0.059 | 0.059 | 1.000 | 0.999 | 1.000 | 1.000 | 0.311 | 0.988 |
|  |  | 0.6 | 0.6 | 0.004 | -0.199 | 0.203 | 0.203 | 0.040 | 0.047 | 0.058 | 0.058 | 1.000 | 0.997 | 1.000 | 1.000 | 0.160 | 0.991 |
|  |  | 0.8 | 0.8 | 0.003 | -0.238 | 0.241 | 0.241 | 0.053 | 0.042 | 0.064 | 0.064 | 1.000 | 0.995 | 1.000 | 1.000 | 0.067 | 0.985 |
|  |  | 1 | 1 | 0.003 | -0.256 | 0.259 | 0.259 | 0.064 | 0.038 | 0.069 | 0.069 | 1.000 | 0.999 | 1.000 | 1.000 | 0.031 | 0.976 |

TE: total effect; DE: direct effect; IE: indirect effect; IE_d: indirect effect calculated by difference method; IE_p: indirect effect calculated by product method.

**eTable 33.** Simulation results of robustness of PE-Egger for all the unmeasured confounders

|  |  | TRUE | | | BIAS | | | | SE | | | | Power | | | | | |
| --- | --- | --- | --- | --- | --- | --- | --- | --- | --- | --- | --- | --- | --- | --- | --- | --- | --- | --- |
|  |  | um | ux | uy | TE | DE | IE_d | IE_p | TE | DE | IE_d | IE_p | TE | DE | IE_d | IE_p | GM | GY |
| Sc2 | IVW | -1 | -1 | -1 | 0.000 | -0.006 | 0.006 | 0.021 | 0.000 | 0.006 | 0.014 | 0.063 | 0.063 | 0.063 | 1.000 | 1.000 | 0.994 | 0.995 |
|  |  | -0.8 | -0.8 | -0.8 | 0.001 | -0.006 | 0.006 | 0.021 | 0.000 | 0.006 | 0.014 | 0.061 | 0.061 | 0.061 | 1.000 | 1.000 | 0.997 | 0.997 |
|  |  | -0.6 | -0.6 | -0.6 | 0.001 | -0.005 | 0.006 | 0.018 | 0.001 | 0.006 | 0.014 | 0.063 | 0.063 | 0.063 | 1.000 | 1.000 | 0.999 | 0.999 |
|  |  | -0.4 | -0.4 | -0.4 | 0.001 | 0.000 | 0.001 | 0.002 | 0.001 | 0.001 | 0.013 | 0.063 | 0.063 | 0.063 | 1.000 | 1.000 | 0.994 | 0.996 |
|  |  | -0.2 | -0.2 | -0.2 | 0.001 | 0.001 | 0.000 | 0.000 | 0.000 | 0.000 | 0.014 | 0.066 | 0.066 | 0.066 | 1.000 | 1.000 | 0.991 | 0.990 |
|  |  | 0 | 0 | 0 | 0.001 | 0.001 | 0.000 | -0.001 | 0.001 | 0.000 | 0.014 | 0.061 | 0.061 | 0.061 | 1.000 | 1.000 | 0.996 | 0.997 |
|  |  | 0.2 | 0.2 | 0.2 | 0.000 | -0.003 | 0.003 | 0.012 | 0.000 | 0.004 | 0.014 | 0.067 | 0.066 | 0.066 | 1.000 | 1.000 | 0.995 | 0.995 |
|  |  | 0.4 | 0.4 | 0.4 | 0.001 | 0.003 | -0.003 | -0.010 | 0.000 | -0.002 | 0.014 | 0.065 | 0.066 | 0.066 | 1.000 | 1.000 | 0.991 | 0.994 |
|  |  | 0.6 | 0.6 | 0.6 | 0.000 | -0.005 | 0.005 | 0.018 | 0.000 | 0.005 | 0.014 | 0.063 | 0.063 | 0.063 | 1.000 | 1.000 | 0.998 | 0.998 |
|  |  | 0.8 | 0.8 | 0.8 | 0.000 | -0.007 | 0.007 | 0.024 | 0.000 | 0.007 | 0.014 | 0.063 | 0.063 | 0.063 | 1.000 | 1.000 | 0.996 | 0.996 |
|  |  | 1 | 1 | 1 | 0.001 | -0.010 | 0.011 | 0.035 | 0.000 | 0.011 | 0.014 | 0.064 | 0.063 | 0.063 | 1.000 | 1.000 | 0.998 | 0.999 |
|  | Egger | -1 | -1 | -1 | 0.001 | -0.006 | 0.007 | 0.020 | 0.001 | 0.007 | 0.036 | 0.069 | 0.070 | 0.070 | 1.000 | 1.000 | 0.991 | 0.994 |
|  |  | -0.8 | -0.8 | -0.8 | 0.002 | -0.005 | 0.007 | 0.023 | 0.000 | 0.007 | 0.034 | 0.065 | 0.065 | 0.065 | 1.000 | 1.000 | 0.992 | 0.997 |
|  |  | -0.6 | -0.6 | -0.6 | 0.002 | -0.004 | 0.006 | 0.016 | 0.001 | 0.006 | 0.035 | 0.069 | 0.070 | 0.070 | 1.000 | 1.000 | 0.996 | 0.998 |
|  |  | -0.4 | -0.4 | -0.4 | 0.000 | -0.001 | 0.001 | 0.001 | 0.000 | 0.001 | 0.035 | 0.070 | 0.071 | 0.071 | 1.000 | 1.000 | 0.992 | 0.993 |
|  |  | -0.2 | -0.2 | -0.2 | 0.001 | 0.000 | 0.002 | 0.003 | 0.001 | 0.002 | 0.033 | 0.070 | 0.071 | 0.071 | 1.000 | 1.000 | 0.983 | 0.989 |
|  |  | 0 | 0 | 0 | 0.002 | 0.001 | 0.001 | -0.001 | 0.002 | 0.001 | 0.034 | 0.067 | 0.067 | 0.067 | 1.000 | 1.000 | 0.991 | 0.996 |
|  |  | 0.2 | 0.2 | 0.2 | 0.000 | -0.003 | 0.003 | 0.011 | 0.000 | 0.003 | 0.035 | 0.071 | 0.070 | 0.070 | 1.000 | 1.000 | 0.989 | 0.994 |
|  |  | 0.4 | 0.4 | 0.4 | 0.001 | 0.004 | -0.003 | -0.009 | 0.000 | -0.002 | 0.034 | 0.071 | 0.072 | 0.072 | 1.000 | 1.000 | 0.984 | 0.993 |
|  |  | 0.6 | 0.6 | 0.6 | -0.001 | -0.007 | 0.006 | 0.020 | 0.000 | 0.006 | 0.033 | 0.067 | 0.068 | 0.068 | 1.000 | 1.000 | 0.993 | 0.996 |
|  |  | 0.8 | 0.8 | 0.8 | -0.002 | -0.008 | 0.006 | 0.023 | -0.001 | 0.006 | 0.036 | 0.069 | 0.068 | 0.068 | 1.000 | 1.000 | 0.989 | 0.997 |
|  |  | 1 | 1 | 1 | 0.002 | -0.009 | 0.011 | 0.034 | 0.001 | 0.011 | 0.036 | 0.071 | 0.071 | 0.071 | 1.000 | 1.000 | 0.993 | 0.996 |
| Sc3 | IVW | -1 | -1 | -1 | 0.075 | -0.084 | 0.159 | 0.359 | 0.038 | 0.159 | 0.010 | 0.066 | 0.066 | 0.066 | 1.000 | 0.997 | 0.998 | 0.999 |
|  |  | -0.8 | -0.8 | -0.8 | 0.075 | -0.079 | 0.154 | 0.344 | 0.038 | 0.154 | 0.009 | 0.063 | 0.063 | 0.063 | 1.000 | 0.998 | 1.000 | 1.000 |
|  |  | -0.6 | -0.6 | -0.6 | 0.075 | -0.073 | 0.148 | 0.328 | 0.038 | 0.148 | 0.009 | 0.066 | 0.066 | 0.066 | 1.000 | 1.000 | 1.000 | 1.000 |
|  |  | -0.4 | -0.4 | -0.4 | 0.074 | -0.069 | 0.142 | 0.314 | 0.037 | 0.142 | 0.009 | 0.066 | 0.066 | 0.066 | 1.000 | 1.000 | 1.000 | 1.000 |
|  |  | -0.2 | -0.2 | -0.2 | 0.074 | -0.065 | 0.139 | 0.303 | 0.037 | 0.139 | 0.009 | 0.066 | 0.066 | 0.066 | 1.000 | 1.000 | 1.000 | 1.000 |
|  |  | 0 | 0 | 0 | 0.074 | -0.067 | 0.141 | 0.309 | 0.037 | 0.142 | 0.009 | 0.061 | 0.061 | 0.061 | 1.000 | 1.000 | 1.000 | 1.000 |
|  |  | 0.2 | 0.2 | 0.2 | 0.074 | -0.065 | 0.139 | 0.303 | 0.037 | 0.139 | 0.009 | 0.064 | 0.064 | 0.064 | 1.000 | 1.000 | 0.999 | 0.999 |
|  |  | 0.4 | 0.4 | 0.4 | 0.074 | -0.068 | 0.142 | 0.311 | 0.037 | 0.142 | 0.009 | 0.066 | 0.066 | 0.066 | 1.000 | 1.000 | 1.000 | 1.000 |
|  |  | 0.6 | 0.6 | 0.6 | 0.074 | -0.068 | 0.142 | 0.313 | 0.037 | 0.142 | 0.009 | 0.063 | 0.063 | 0.063 | 1.000 | 0.999 | 0.999 | 0.998 |
|  |  | 0.8 | 0.8 | 0.8 | 0.074 | -0.078 | 0.152 | 0.341 | 0.037 | 0.152 | 0.009 | 0.066 | 0.066 | 0.066 | 1.000 | 0.997 | 1.000 | 0.999 |
|  |  | 1 | 1 | 1 | 0.075 | -0.080 | 0.155 | 0.349 | 0.037 | 0.155 | 0.009 | 0.063 | 0.064 | 0.064 | 1.000 | 0.998 | 1.000 | 1.000 |
|  | Egger | -1 | -1 | -1 | 0.002 | -0.032 | 0.034 | 0.108 | 0.001 | 0.034 | 0.020 | 0.067 | 0.068 | 0.068 | 1.000 | 1.000 | 0.957 | 0.965 |
|  |  | -0.8 | -0.8 | -0.8 | 0.001 | -0.023 | 0.024 | 0.076 | 0.001 | 0.024 | 0.019 | 0.066 | 0.067 | 0.067 | 1.000 | 0.998 | 0.952 | 0.959 |
|  |  | -0.6 | -0.6 | -0.6 | 0.000 | -0.014 | 0.014 | 0.048 | 0.000 | 0.014 | 0.019 | 0.066 | 0.066 | 0.066 | 1.000 | 1.000 | 0.965 | 0.965 |
|  |  | -0.4 | -0.4 | -0.4 | 0.000 | -0.006 | 0.006 | 0.022 | 0.000 | 0.006 | 0.018 | 0.066 | 0.065 | 0.065 | 1.000 | 1.000 | 0.971 | 0.972 |
|  |  | -0.2 | -0.2 | -0.2 | 0.000 | -0.002 | 0.002 | 0.007 | 0.000 | 0.002 | 0.019 | 0.066 | 0.066 | 0.066 | 1.000 | 1.000 | 0.968 | 0.975 |
|  |  | 0 | 0 | 0 | -0.001 | 0.001 | -0.002 | -0.006 | 0.000 | -0.002 | 0.018 | 0.062 | 0.061 | 0.061 | 1.000 | 1.000 | 0.966 | 0.971 |
|  |  | 0.2 | 0.2 | 0.2 | 0.001 | -0.001 | 0.002 | 0.004 | 0.001 | 0.002 | 0.018 | 0.064 | 0.064 | 0.064 | 1.000 | 1.000 | 0.966 | 0.970 |
|  |  | 0.4 | 0.4 | 0.4 | 0.001 | -0.007 | 0.008 | 0.023 | 0.001 | 0.008 | 0.018 | 0.068 | 0.068 | 0.068 | 1.000 | 0.999 | 0.964 | 0.967 |
|  |  | 0.6 | 0.6 | 0.6 | 0.001 | -0.010 | 0.011 | 0.035 | 0.000 | 0.011 | 0.019 | 0.065 | 0.065 | 0.065 | 1.000 | 0.999 | 0.953 | 0.961 |
|  |  | 0.8 | 0.8 | 0.8 | 0.001 | -0.021 | 0.021 | 0.069 | 0.001 | 0.021 | 0.019 | 0.065 | 0.065 | 0.065 | 1.000 | 0.999 | 0.968 | 0.975 |
|  |  | 1 | 1 | 1 | 0.002 | -0.031 | 0.032 | 0.106 | 0.000 | 0.032 | 0.020 | 0.064 | 0.064 | 0.064 | 1.000 | 1.000 | 0.967 | 0.971 |
| Sc4 | IVW | -1 | -1 | -1 | -0.110 | -0.271 | 0.161 | 0.882 | -0.055 | 0.161 | 0.030 | 0.025 | 0.038 | 0.038 | 1.000 | 1.000 | 1.000 | 1.000 |
|  |  | -0.8 | -0.8 | -0.8 | -0.073 | -0.252 | 0.179 | 0.816 | -0.037 | 0.179 | 0.023 | 0.033 | 0.039 | 0.039 | 1.000 | 1.000 | 1.000 | 1.000 |
|  |  | -0.6 | -0.6 | -0.6 | -0.035 | -0.212 | 0.177 | 0.688 | -0.018 | 0.177 | 0.018 | 0.042 | 0.045 | 0.045 | 1.000 | 1.000 | 1.000 | 1.000 |
|  |  | -0.4 | -0.4 | -0.4 | 0.001 | -0.141 | 0.143 | 0.473 | 0.001 | 0.143 | 0.012 | 0.049 | 0.050 | 0.050 | 1.000 | 1.000 | 1.000 | 1.000 |
|  |  | -0.2 | -0.2 | -0.2 | 0.038 | -0.058 | 0.096 | 0.240 | 0.019 | 0.096 | 0.009 | 0.065 | 0.066 | 0.066 | 1.000 | 1.000 | 0.998 | 0.998 |
|  |  | 0 | 0 | 0 | 0.074 | -0.066 | 0.140 | 0.306 | 0.037 | 0.141 | 0.009 | 0.065 | 0.065 | 0.065 | 1.000 | 1.000 | 0.999 | 0.999 |
|  |  | 0.2 | 0.2 | 0.2 | 0.110 | -0.135 | 0.245 | 0.534 | 0.055 | 0.245 | 0.011 | 0.061 | 0.063 | 0.063 | 1.000 | 0.998 | 1.000 | 1.000 |
|  |  | 0.4 | 0.4 | 0.4 | 0.145 | -0.192 | 0.337 | 0.709 | 0.073 | 0.337 | 0.015 | 0.053 | 0.055 | 0.055 | 1.000 | 0.997 | 1.000 | 1.000 |
|  |  | 0.6 | 0.6 | 0.6 | 0.180 | -0.223 | 0.403 | 0.802 | 0.090 | 0.403 | 0.019 | 0.047 | 0.051 | 0.051 | 1.000 | 0.998 | 1.000 | 1.000 |
|  |  | 0.8 | 0.8 | 0.8 | 0.215 | -0.246 | 0.461 | 0.868 | 0.108 | 0.461 | 0.023 | 0.042 | 0.048 | 0.048 | 1.000 | 0.993 | 1.000 | 1.000 |
|  |  | 1 | 1 | 1 | 0.247 | -0.258 | 0.505 | 0.901 | 0.124 | 0.505 | 0.025 | 0.040 | 0.047 | 0.047 | 1.000 | 0.992 | 1.000 | 1.000 |
|  | Egger | -1 | -1 | -1 | 0.040 | -0.253 | 0.292 | 0.850 | 0.020 | 0.292 | 0.062 | 0.040 | 0.070 | 0.070 | 1.000 | 1.000 | 1.000 | 1.000 |
|  |  | -0.8 | -0.8 | -0.8 | 0.024 | -0.233 | 0.257 | 0.783 | 0.012 | 0.257 | 0.051 | 0.044 | 0.061 | 0.061 | 1.000 | 0.999 | 1.000 | 1.000 |
|  |  | -0.6 | -0.6 | -0.6 | 0.014 | -0.197 | 0.212 | 0.667 | 0.007 | 0.212 | 0.041 | 0.048 | 0.060 | 0.060 | 1.000 | 0.997 | 1.000 | 1.000 |
|  |  | -0.4 | -0.4 | -0.4 | 0.006 | -0.141 | 0.147 | 0.472 | 0.004 | 0.147 | 0.030 | 0.052 | 0.056 | 0.056 | 1.000 | 1.000 | 1.000 | 1.000 |
|  |  | -0.2 | -0.2 | -0.2 | 0.002 | -0.052 | 0.054 | 0.176 | 0.001 | 0.054 | 0.022 | 0.066 | 0.067 | 0.067 | 1.000 | 1.000 | 0.997 | 0.997 |
|  |  | 0 | 0 | 0 | 0.000 | 0.000 | 0.000 | -0.001 | 0.000 | 0.000 | 0.018 | 0.067 | 0.068 | 0.068 | 1.000 | 1.000 | 0.964 | 0.973 |
|  |  | 0.2 | 0.2 | 0.2 | 0.003 | -0.054 | 0.058 | 0.186 | 0.002 | 0.058 | 0.022 | 0.064 | 0.065 | 0.065 | 1.000 | 0.999 | 0.996 | 0.997 |
|  |  | 0.4 | 0.4 | 0.4 | 0.007 | -0.141 | 0.148 | 0.475 | 0.004 | 0.148 | 0.030 | 0.055 | 0.059 | 0.059 | 1.000 | 0.996 | 1.000 | 1.000 |
|  |  | 0.6 | 0.6 | 0.6 | 0.016 | -0.199 | 0.214 | 0.673 | 0.007 | 0.214 | 0.039 | 0.048 | 0.057 | 0.057 | 1.000 | 0.997 | 1.000 | 1.000 |
|  |  | 0.8 | 0.8 | 0.8 | 0.026 | -0.234 | 0.260 | 0.789 | 0.013 | 0.260 | 0.053 | 0.042 | 0.063 | 0.063 | 1.000 | 0.994 | 1.000 | 1.000 |
|  |  | 1 | 1 | 1 | 0.041 | -0.252 | 0.293 | 0.849 | 0.021 | 0.293 | 0.061 | 0.040 | 0.069 | 0.069 | 1.000 | 0.988 | 1.000 | 1.000 |

TE: total effect; DE: direct effect; IE: indirect effect; IE_d: indirect effect calculated by difference method; IE_p: indirect effect calculated by product method.

#### 9 Additional simulation scenario 3: PSE-MR with time varying exposure and mediator

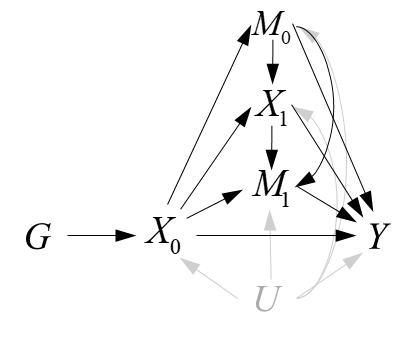

**eFigure 8.** Graphical diagram of time-varying exposure and mediators

The data-generating models with no pleiotropy were:

The data-generating models with pleiotropy were:

Based on the DAG in Labrecque and Swanson (2020), we performed our method to estimate lifetime effects of *A* and *B* on *Y*. The definition of a total lifetime effect of a variable was the same as the Labrecque and Swanson (2020), and it was the effect of shifting the trajectory of the exposure by one unit on the outcome measured at a specific point in time. We regarded *B_0_*, *A_1_* and *B_1_* as the three causally ordered mediators from *A_0_* on Y (**eFigure 8**). The parameters’ settings were the same as that in Labrecque and Swanson (2020). And we only considered the case where the direct effects denoted by the gray arrows were non-zero. Results of simulation were listed in the **eTable 34.** In one sample setting, PSE-MR obtained unbiased estimates of the lifetime effects of *A* and *B* on *Y* when there was no pleiotropy, balanced and directional pleiotropy with the InSIDE assumption was satisfied. However, the estimates were biased in two sample MR setting.

**eTable 34.** Simulation results of with time varying exposure and mediator.

|  |  | BIAS | | SE | | MSE | |
| --- | --- | --- | --- | --- | --- | --- | --- |
|  |  | A-Y | B-Y | A-Y | B-Y | A-Y | B-Y |
| PSE-IVW |  | -0.011 | 0.002 | 0.456 | 0.438 | 0.208 | 0.192 |
| PSE-Egger | sc2 | -0.028 | -0.002 | 0.373 | 0.335 | 0.140 | 0.112 |
|  | sc3 | -0.076 | 0.007 | 0.389 | 0.326 | 0.157 | 0.106 |
|  | sc4 | -0.477 | 0.009 | 0.358 | 0.361 | 0.355 | 0.131 |

#### 10 Additional simulation scenario 4: PSE-MR with Heterogeneous populations

The genetic composition of different human populations varied because of, among other factors, their differing evolutionary histories and patterns of dispersal and interbreeding. Balding D J and Nichols R A proposed a method for quantifying the effect of genetic differentiation in terms of a single parameter *F*_st_ [ref 1], which can often be interpreted in terms of coancestry. Thus, we generated heterogeneous populations with different values of *F*_st_. For heterogeneous populations, we conducted simulations in four populations (the exposure, outcome and two mediators are from four different populations) with different values of *F*_st_: 0.008 , 0.027, 0.043 and 0.07, in two mediators setting. These indexes were obtained by a survey (Cavalli-Sforza & Piazza, 1993), which reported F_st_ estimates among Europeans with a median of 0.8% while the 90th percentile was about 2.8%. The corresponding values were 2.7% and 14% among Africans and 4.3% and 12% among Asians. Within Europe, the most divergent ethnic groups had been found to have values of the order of 7%.

Compared with four homogeneous populations or one population, heterogeneous populations sometimes brought larger bias for non-ordered and ordered mediators. For non-ordered mediators, PSE-Egger can obtain unbiased estimates of the total effect, direct or the indirect effect when they were null. When the causal effects were non-zero, for PSE-IVW, the bias of total effect was smaller than that of direct and indirect effect. Because the direction of the bias of the direct and indirect effects were opposite. This seemingly canceled out much of the bias. For PSE-Egger, when the direction of true direct and indirect effects were the same, the bias of total effect was larger than that of direct and indirect effect, and the direction of the bias of the direct and indirect effects were also same. In the contrast, if the direction of true direct and indirect effects were opposite, the bias was similar to that of PSE-IVW. In this case, PSE-IVW was more plausible than PSE-Egger. For ordered mediators, the bias of total effect was smaller than that of direct and indirect effect in PSE-IVW. And it was also in PSE-Egger except one case that the exposure had no direct effect on *M*_2_ and the *M*_1_ had no direct on the outcome, that is, the only indirect pathway was *X*→*M*_1_→*M*_2_→*Y*. In conclusion, if there was no pleiotropy, PSE-IVW was more robust than PSE-IVW in heterogeneous populations.

For both causally ordered and non-ordered mediators, when the causal order was misspecified, total effect, direct effect and indirect effect (through all the mediators) can be unbiasedly estimated. This was because, taking two mediators as an example, the total effect was estimated by the weighted regression , which didn’t use the information of mediators. Similarly, the direct effect was estimated by the weighted regression , which was not also influenced by the order of mediators. Moreover, the indirect effect can be estimated by the difference method (). In two non-ordered mediators setting, the order of mediators was independent of estimation and path-specific effects (*X*→*M*_1_→*Y* and *X*→*M*_2_→*Y*) were unbiased. However, in two ordered mediators setting, path-specific effects (*X*→*M*_1_→*Y*, *X*→*M*_2_→*Y* and *X*→*M*_1_→*M*_2_→*Y*) would be biased when the causal order was misspecified. If the correct order was *M*_2_→*M*_1_, we could obtain the estimate of parameters in the indirect pathway by the weighted regression . However, if we were misspecified this order, we would regress the on the and the . Thus the above three path-specific effects would be biased. Results were listed in e**Table 35-36**.

**eTable 35** Simulation of heterogeneous populations with two non-ordered mediators

|  |  | TRUE | | | | | BIAS | | | | | | SE | | | | | | MSE | | | | | | Power/Type I error | | | | | | | | |
| --- | --- | --- | --- | --- | --- | --- | --- | --- | --- | --- | --- | --- | --- | --- | --- | --- | --- | --- | --- | --- | --- | --- | --- | --- | --- | --- | --- | --- | --- | --- | --- | --- | --- |
|  |  | TE | DE | IE | IE1 | IE2 | TE | DE | IE_d | IE1 | IE2 | IE_p | TE | DE | IE_d | IE1 | IE2 | IE_p | TE | DE | IE_d | IE1 | IE2 | IE_p | TE | DE | IE_d | IE1 | IE2 | IE_p | GY | GM1 | GM2 |
| Standard MR | IVW | 1.6 | 0.8 | 0.8 | 0.5 | 0.3 | -0.153 | 0.334 | -0.487 | -0.487 | -0.254 | -0.232 | 0.429 | 0.830 | 0.710 | 0.710 | 0.643 | 0.371 | 0.208 | 0.799 | 0.741 | 0.741 | 0.478 | 0.192 | 0.855 | 0.237 | 0.013 | 0.064 | 0.084 | 0.043 |  |  |  |
|  |  | 0.6 | 0.8 | -0.2 | -0.5 | 0.3 | -0.071 | -0.379 | 0.308 | 0.308 | 0.594 | -0.286 | 0.209 | 0.383 | 0.324 | 0.324 | 0.296 | 0.174 | 0.049 | 0.290 | 0.200 | 0.200 | 0.441 | 0.112 | 0.634 | 0.153 | 0.009 | 0.045 | 0.055 | 0.034 |  |  |  |
|  |  | 0.7 | 0.5 | 0.2 | 0.5 | -0.3 | -0.065 | 0.006 | -0.071 | -0.071 | -0.404 | 0.333 | 0.215 | 0.419 | 0.357 | 0.357 | 0.312 | 0.188 | 0.050 | 0.176 | 0.133 | 0.133 | 0.260 | 0.146 | 0.792 | 0.200 | 0.018 | 0.068 | 0.081 | 0.053 |  |  |  |
|  |  | -0.3 | 0.5 | -0.8 | -0.5 | -0.3 | 0.033 | -0.703 | 0.736 | 0.736 | 0.447 | 0.289 | 0.180 | 0.335 | 0.292 | 0.292 | 0.266 | 0.165 | 0.033 | 0.606 | 0.626 | 0.626 | 0.271 | 0.111 | 0.246 | 0.062 | 0.008 | 0.042 | 0.055 | 0.053 |  |  |  |
|  |  | 0 | -0.8 | 0.8 | 0.5 | 0.3 | 0.005 | 0.822 | -0.817 | -0.817 | -0.509 | -0.307 | 0.294 | 0.542 | 0.468 | 0.468 | 0.418 | 0.252 | 0.086 | 0.969 | 0.886 | 0.886 | 0.434 | 0.158 | 0.031 | 0.029 | 0.010 | 0.040 | 0.063 | 0.040 |  |  |  |
|  |  | -1.6 | -0.8 | -0.8 | -0.5 | -0.3 | 0.143 | -0.322 | 0.464 | 0.464 | 0.238 | 0.226 | 0.313 | 0.562 | 0.486 | 0.487 | 0.431 | 0.267 | 0.118 | 0.420 | 0.452 | 0.452 | 0.243 | 0.122 | 0.994 | 0.475 | 0.021 | 0.091 | 0.095 | 0.061 |  |  |  |
|  |  | 0.8 | 0 | 0.8 | 0.5 | 0.3 | -0.068 | 0.540 | -0.607 | -0.607 | -0.354 | -0.254 | 0.356 | 0.640 | 0.564 | 0.564 | 0.513 | 0.292 | 0.132 | 0.701 | 0.687 | 0.687 | 0.388 | 0.150 | 0.459 | 0.093 | 0.010 | 0.048 | 0.061 | 0.035 |  |  |  |
|  |  | 1.3 | 0.8 | 0.5 | 0.5 | 0 | -0.112 | 0.110 | -0.222 | -0.222 | -0.286 | 0.063 | 0.341 | 0.564 | 0.470 | 0.470 | 0.428 | 0.271 | 0.129 | 0.330 | 0.270 | 0.270 | 0.265 | 0.078 | 0.916 | 0.277 | 0.005 | 0.049 | 0.065 | 0.052 |  |  |  |
|  |  | 1.3 | 0.8 | 0.5 | 0.5 | 0 | -0.097 | 0.197 | -0.293 | -0.293 | -0.288 | -0.005 | 0.417 | 0.694 | 0.582 | 0.582 | 0.574 | 0.090 | 0.184 | 0.521 | 0.425 | 0.425 | 0.413 | 0.008 | 0.771 | 0.261 | 0.008 | 0.069 | 0.075 | 0.000 |  |  |  |
|  |  | 0.8 | 0.8 | 0 | 0 | 0 | -0.086 | -0.244 | 0.157 | 0.157 | 0.122 | 0.035 | 0.191 | 0.353 | 0.298 | 0.298 | 0.269 | 0.168 | 0.044 | 0.184 | 0.113 | 0.113 | 0.087 | 0.029 | 0.950 | 0.294 | 0.020 | 0.069 | 0.083 | 0.049 |  |  |  |
| Sc2 | IVW | 1.6 | 0.8 | 0.8 | 0.5 | 0.3 | -0.245 | -0.237 | -0.008 | -0.008 | -0.070 | 0.063 | 0.182 | 0.340 | 0.319 | 0.319 | 0.288 | 0.296 | 0.093 | 0.172 | 0.102 | 0.102 | 0.088 | 0.092 | 1.000 | 0.347 | 0.514 | 0.495 | 0.329 | 0.288 |  |  |  |
|  |  | 0.6 | 0.8 | -0.2 | -0.5 | 0.3 | -0.100 | -0.601 | 0.502 | 0.502 | 0.666 | -0.164 | 0.074 | 0.145 | 0.134 | 0.134 | 0.117 | 0.109 | 0.015 | 0.383 | 0.269 | 0.270 | 0.457 | 0.039 | 1.000 | 0.284 | 0.455 | 0.447 | 0.316 | 0.241 |  |  |  |
|  |  | 0.7 | 0.5 | 0.2 | 0.5 | -0.3 | -0.111 | -0.252 | 0.140 | 0.140 | -0.313 | 0.453 | 0.082 | 0.159 | 0.147 | 0.147 | 0.140 | 0.131 | 0.019 | 0.089 | 0.041 | 0.041 | 0.118 | 0.223 | 1.000 | 0.333 | 0.460 | 0.434 | 0.320 | 0.258 |  |  |  |
|  |  | -0.3 | 0.5 | -0.8 | -0.5 | -0.3 | 0.047 | -0.604 | 0.651 | 0.651 | 0.418 | 0.233 | 0.048 | 0.095 | 0.087 | 0.087 | 0.080 | 0.076 | 0.004 | 0.374 | 0.431 | 0.431 | 0.181 | 0.060 | 0.999 | 0.205 | 0.242 | 0.246 | 0.208 | 0.182 |  |  |  |
|  |  | 0 | -0.8 | 0.8 | 0.5 | 0.3 | 0.001 | 0.803 | -0.802 | -0.802 | -0.503 | -0.298 | 0.052 | 0.105 | 0.095 | 0.095 | 0.093 | 0.085 | 0.003 | 0.656 | 0.652 | 0.652 | 0.262 | 0.096 | 0.031 | 0.036 | 0.008 | 0.009 | 0.057 | 0.057 |  |  |  |
|  |  | -1.6 | -0.8 | -0.8 | -0.5 | -0.3 | 0.269 | 0.256 | 0.013 | 0.012 | 0.063 | -0.051 | 0.167 | 0.337 | 0.313 | 0.313 | 0.277 | 0.268 | 0.100 | 0.179 | 0.098 | 0.098 | 0.081 | 0.075 | 1.000 | 0.343 | 0.575 | 0.571 | 0.375 | 0.301 |  |  |  |
|  |  | 0.8 | 0 | 0.8 | 0.5 | 0.3 | -0.128 | 0.274 | -0.402 | -0.402 | -0.287 | -0.115 | 0.107 | 0.198 | 0.180 | 0.180 | 0.176 | 0.165 | 0.028 | 0.115 | 0.194 | 0.194 | 0.113 | 0.040 | 1.000 | 0.267 | 0.372 | 0.384 | 0.266 | 0.249 |  |  |  |
|  |  | 1.3 | 0.8 | 0.5 | 0.5 | 0 | -0.219 | -0.355 | 0.137 | 0.137 | -0.146 | 0.282 | 0.144 | 0.277 | 0.253 | 0.253 | 0.230 | 0.224 | 0.069 | 0.203 | 0.083 | 0.083 | 0.074 | 0.130 | 1.000 | 0.340 | 0.508 | 0.500 | 0.337 | 0.286 |  |  |  |
|  |  | 1.3 | 0.8 | 0.5 | 0.5 | 0 | -0.213 | -0.182 | -0.031 | -0.031 | -0.032 | 0.001 | 0.147 | 0.256 | 0.226 | 0.226 | 0.224 | 0.031 | 0.067 | 0.098 | 0.052 | 0.052 | 0.051 | 0.001 | 1.000 | 0.570 | 0.229 | 0.525 | 0.541 | 0.001 |  |  |  |
|  |  | 0.8 | 0.8 | 0 | 0 | 0 | -0.126 | -0.506 | 0.380 | 0.380 | 0.209 | 0.171 | 0.087 | 0.170 | 0.158 | 0.158 | 0.150 | 0.141 | 0.023 | 0.285 | 0.169 | 0.169 | 0.066 | 0.049 | 1.000 | 0.382 | 0.510 | 0.487 | 0.346 | 0.283 |  |  |  |
|  | Egger | 1.6 | 0.8 | 0.8 | 0.5 | 0.3 | -1.579 | -0.779 | -0.800 | -0.800 | -0.502 | -0.298 | 0.377 | 0.403 | 0.127 | 0.127 | 0.088 | 0.088 | 2.634 | 0.770 | 0.656 | 0.656 | 0.260 | 0.096 | 0.045 | 0.057 | 0.000 | 0.004 | 0.004 | 0.005 | 0.631 | 0.945 | 0.973 |
|  |  | 0.6 | 0.8 | -0.2 | -0.5 | 0.3 | -0.607 | -0.809 | 0.202 | 0.202 | 0.502 | -0.300 | 0.155 | 0.162 | 0.052 | 0.052 | 0.036 | 0.037 | 0.392 | 0.680 | 0.043 | 0.043 | 0.253 | 0.091 | 0.045 | 0.052 | 0.000 | 0.003 | 0.004 | 0.003 | 0.551 | 0.961 | 0.986 |
|  |  | 0.7 | 0.5 | 0.2 | 0.5 | -0.3 | -0.695 | -0.493 | -0.202 | -0.202 | -0.500 | 0.298 | 0.181 | 0.189 | 0.058 | 0.058 | 0.042 | 0.039 | 0.516 | 0.279 | 0.044 | 0.044 | 0.251 | 0.090 | 0.067 | 0.066 | 0.000 | 0.003 | 0.003 | 0.002 | 0.595 | 0.953 | 0.980 |
|  |  | -0.3 | 0.5 | -0.8 | -0.5 | -0.3 | 0.298 | -0.502 | 0.800 | 0.800 | 0.500 | 0.300 | 0.105 | 0.112 | 0.033 | 0.033 | 0.025 | 0.023 | 0.100 | 0.265 | 0.642 | 0.642 | 0.251 | 0.091 | 0.038 | 0.041 | 0.000 | 0.003 | 0.004 | 0.002 | 0.346 | 0.934 | 0.982 |
|  |  | 0 | -0.8 | 0.8 | 0.5 | 0.3 | 0.007 | 0.805 | -0.798 | -0.798 | -0.499 | -0.299 | 0.125 | 0.131 | 0.042 | 0.042 | 0.029 | 0.031 | 0.016 | 0.666 | 0.639 | 0.639 | 0.250 | 0.090 | 0.034 | 0.031 | 0.000 | 0.003 | 0.000 | 0.004 | 0.040 | 0.940 | 0.974 |
|  |  | -1.6 | -0.8 | -0.8 | -0.5 | -0.3 | 1.594 | 0.793 | 0.801 | 0.801 | 0.502 | 0.300 | 0.345 | 0.360 | 0.111 | 0.111 | 0.079 | 0.078 | 2.660 | 0.758 | 0.654 | 0.654 | 0.258 | 0.096 | 0.046 | 0.041 | 0.000 | 0.002 | 0.003 | 0.002 | 0.672 | 0.963 | 0.981 |
|  |  | 0.8 | 0 | 0.8 | 0.5 | 0.3 | -0.789 | 0.010 | -0.799 | -0.799 | -0.499 | -0.300 | 0.220 | 0.228 | 0.067 | 0.067 | 0.048 | 0.050 | 0.671 | 0.052 | 0.643 | 0.643 | 0.252 | 0.092 | 0.037 | 0.039 | 0.000 | 0.003 | 0.001 | 0.003 | 0.480 | 0.955 | 0.980 |
|  |  | 1.3 | 0.8 | 0.5 | 0.5 | 0 | -1.301 | -0.798 | -0.503 | -0.503 | -0.503 | -0.001 | 0.309 | 0.323 | 0.096 | 0.096 | 0.066 | 0.068 | 1.789 | 0.741 | 0.262 | 0.262 | 0.257 | 0.005 | 0.063 | 0.063 | 0.000 | 0.003 | 0.002 | 0.001 | 0.641 | 0.960 | 0.985 |
|  |  | 1.3 | 0.8 | 0.5 | 0.5 | 0 | -1.301 | -0.801 | -0.500 | -0.500 | -0.498 | -0.002 | 0.312 | 0.328 | 0.101 | 0.101 | 0.076 | 0.070 | 1.791 | 0.750 | 0.260 | 0.260 | 0.254 | 0.005 | 0.041 | 0.041 | 0.000 | 0.002 | 0.006 | 0.001 | 0.723 | 0.039 | 0.970 |
|  |  | 0.8 | 0.8 | 0 | 0 | 0 | -0.791 | -0.786 | -0.005 | -0.005 | -0.002 | -0.003 | 0.187 | 0.194 | 0.061 | 0.061 | 0.040 | 0.044 | 0.660 | 0.655 | 0.004 | 0.004 | 0.002 | 0.002 | 0.058 | 0.056 | 0.000 | 0.004 | 0.005 | 0.001 | 0.653 | 0.940 | 0.983 |
| Sc3 | IVW | 1.6 | 0.8 | 0.8 | 0.5 | 0.3 | -0.150 | -0.291 | 0.142 | 0.142 | -0.018 | 0.160 | 0.189 | 0.361 | 0.324 | 0.324 | 0.305 | 0.316 | 0.058 | 0.215 | 0.125 | 0.125 | 0.094 | 0.126 | 1.000 | 0.268 | 0.657 | 0.552 | 0.345 | 0.338 |  |  |  |
|  |  | 0.6 | 0.8 | -0.2 | -0.5 | 0.3 | -0.058 | -0.607 | 0.549 | 0.549 | 0.675 | -0.125 | 0.072 | 0.139 | 0.132 | 0.132 | 0.127 | 0.129 | 0.009 | 0.388 | 0.319 | 0.319 | 0.472 | 0.032 | 1.000 | 0.254 | 0.579 | 0.500 | 0.304 | 0.319 |  |  |  |
|  |  | 0.7 | 0.5 | 0.2 | 0.5 | -0.3 | -0.074 | -0.281 | 0.207 | 0.207 | -0.299 | 0.505 | 0.081 | 0.157 | 0.150 | 0.150 | 0.148 | 0.141 | 0.012 | 0.104 | 0.065 | 0.065 | 0.111 | 0.275 | 1.000 | 0.243 | 0.620 | 0.506 | 0.316 | 0.334 |  |  |  |
|  |  | -0.3 | 0.5 | -0.8 | -0.5 | -0.3 | 0.014 | -0.598 | 0.612 | 0.612 | 0.407 | 0.205 | 0.047 | 0.093 | 0.083 | 0.083 | 0.083 | 0.081 | 0.002 | 0.366 | 0.382 | 0.381 | 0.173 | 0.048 | 1.000 | 0.186 | 0.433 | 0.351 | 0.249 | 0.251 |  |  |  |
|  |  | 0 | -0.8 | 0.8 | 0.5 | 0.3 | 0.113 | 0.838 | -0.725 | -0.725 | -0.458 | -0.266 | 0.050 | 0.104 | 0.093 | 0.093 | 0.091 | 0.090 | 0.015 | 0.712 | 0.534 | 0.534 | 0.218 | 0.079 | 0.551 | 0.041 | 0.027 | 0.017 | 0.065 | 0.056 |  |  |  |
|  |  | -1.6 | -0.8 | -0.8 | -0.5 | -0.3 | 0.221 | 0.319 | -0.098 | -0.098 | 0.050 | -0.148 | 0.172 | 0.343 | 0.323 | 0.323 | 0.307 | 0.297 | 0.078 | 0.219 | 0.114 | 0.114 | 0.096 | 0.110 | 1.000 | 0.268 | 0.667 | 0.579 | 0.349 | 0.333 |  |  |  |
|  |  | 0.8 | 0 | 0.8 | 0.5 | 0.3 | -0.018 | 0.278 | -0.295 | -0.295 | -0.251 | -0.044 | 0.102 | 0.207 | 0.192 | 0.192 | 0.192 | 0.183 | 0.011 | 0.120 | 0.124 | 0.124 | 0.100 | 0.036 | 1.000 | 0.250 | 0.571 | 0.487 | 0.303 | 0.302 |  |  |  |
|  |  | 1.3 | 0.8 | 0.5 | 0.5 | 0 | -0.139 | -0.410 | 0.271 | 0.271 | -0.093 | 0.364 | 0.147 | 0.290 | 0.276 | 0.276 | 0.264 | 0.268 | 0.041 | 0.252 | 0.150 | 0.150 | 0.078 | 0.205 | 1.000 | 0.255 | 0.687 | 0.588 | 0.393 | 0.328 |  |  |  |
|  |  | 1.3 | 0.8 | 0.5 | 0.5 | 0 | -0.106 | -0.220 | 0.114 | 0.115 | 0.043 | 0.071 | 0.151 | 0.270 | 0.250 | 0.250 | 0.242 | 0.123 | 0.034 | 0.121 | 0.076 | 0.076 | 0.060 | 0.020 | 1.000 | 0.490 | 0.410 | 0.614 | 0.604 | 0.084 |  |  |  |
|  |  | 0.8 | 0.8 | 0 | 0 | 0 | -0.089 | -0.543 | 0.454 | 0.454 | 0.230 | 0.223 | 0.094 | 0.174 | 0.157 | 0.157 | 0.154 | 0.151 | 0.017 | 0.325 | 0.231 | 0.231 | 0.077 | 0.073 | 1.000 | 0.275 | 0.629 | 0.541 | 0.343 | 0.326 |  |  |  |
|  | Egger | 1.6 | 0.8 | 0.8 | 0.5 | 0.3 | -1.591 | -0.789 | -0.802 | -0.802 | -0.502 | -0.300 | 0.387 | 0.400 | 0.114 | 0.114 | 0.079 | 0.085 | 2.681 | 0.782 | 0.657 | 0.657 | 0.259 | 0.097 | 0.060 | 0.054 | 0.000 | 0.000 | 0.003 | 0.001 | 0.608 | 0.986 | 0.994 |
|  |  | 0.6 | 0.8 | -0.2 | -0.5 | 0.3 | -0.592 | -0.789 | 0.197 | 0.197 | 0.500 | -0.302 | 0.152 | 0.157 | 0.046 | 0.046 | 0.032 | 0.033 | 0.374 | 0.648 | 0.041 | 0.041 | 0.251 | 0.092 | 0.049 | 0.047 | 0.000 | 0.001 | 0.001 | 0.002 | 0.538 | 0.991 | 0.995 |
|  |  | 0.7 | 0.5 | 0.2 | 0.5 | -0.3 | -0.697 | -0.498 | -0.199 | -0.199 | -0.500 | 0.301 | 0.166 | 0.175 | 0.056 | 0.056 | 0.039 | 0.038 | 0.513 | 0.278 | 0.043 | 0.043 | 0.252 | 0.092 | 0.043 | 0.047 | 0.000 | 0.003 | 0.004 | 0.004 | 0.591 | 0.991 | 0.993 |
|  |  | -0.3 | 0.5 | -0.8 | -0.5 | -0.3 | 0.301 | -0.501 | 0.801 | 0.801 | 0.501 | 0.300 | 0.103 | 0.107 | 0.032 | 0.032 | 0.023 | 0.023 | 0.101 | 0.262 | 0.643 | 0.643 | 0.251 | 0.091 | 0.055 | 0.049 | 0.000 | 0.003 | 0.003 | 0.004 | 0.403 | 0.989 | 0.995 |
|  |  | 0 | -0.8 | 0.8 | 0.5 | 0.3 | 0.002 | 0.803 | -0.801 | -0.801 | -0.501 | -0.300 | 0.120 | 0.126 | 0.038 | 0.038 | 0.027 | 0.026 | 0.014 | 0.661 | 0.643 | 0.643 | 0.251 | 0.091 | 0.029 | 0.028 | 0.000 | 0.000 | 0.000 | 0.000 | 0.060 | 0.986 | 0.994 |
|  |  | -1.6 | -0.8 | -0.8 | -0.5 | -0.3 | 1.591 | 0.792 | 0.799 | 0.799 | 0.502 | 0.297 | 0.355 | 0.374 | 0.118 | 0.118 | 0.082 | 0.085 | 2.656 | 0.766 | 0.653 | 0.653 | 0.258 | 0.096 | 0.058 | 0.057 | 0.000 | 0.005 | 0.001 | 0.008 | 0.622 | 0.989 | 0.992 |
|  |  | 0.8 | 0 | 0.8 | 0.5 | 0.3 | -0.805 | -0.002 | -0.803 | -0.803 | -0.500 | -0.303 | 0.218 | 0.231 | 0.075 | 0.075 | 0.055 | 0.053 | 0.696 | 0.053 | 0.650 | 0.650 | 0.253 | 0.094 | 0.051 | 0.051 | 0.000 | 0.002 | 0.002 | 0.003 | 0.573 | 0.989 | 0.990 |
|  |  | 1.3 | 0.8 | 0.5 | 0.5 | 0 | -1.299 | -0.803 | -0.496 | -0.496 | -0.498 | 0.002 | 0.308 | 0.323 | 0.091 | 0.091 | 0.065 | 0.065 | 1.782 | 0.749 | 0.254 | 0.254 | 0.253 | 0.004 | 0.055 | 0.061 | 0.000 | 0.002 | 0.001 | 0.001 | 0.609 | 0.988 | 0.993 |
|  |  | 1.3 | 0.8 | 0.5 | 0.5 | 0 | -1.297 | -0.799 | -0.498 | -0.498 | -0.502 | 0.004 | 0.312 | 0.325 | 0.100 | 0.100 | 0.067 | 0.073 | 1.780 | 0.744 | 0.258 | 0.258 | 0.256 | 0.005 | 0.051 | 0.049 | 0.000 | 0.006 | 0.003 | 0.005 | 0.746 | 0.364 | 0.992 |
|  |  | 0.8 | 0.8 | 0 | 0 | 0 | -0.799 | -0.797 | -0.002 | -0.002 | -0.003 | 0.001 | 0.187 | 0.197 | 0.057 | 0.057 | 0.038 | 0.042 | 0.673 | 0.674 | 0.003 | 0.003 | 0.001 | 0.002 | 0.055 | 0.054 | 0.000 | 0.000 | 0.000 | 0.004 | 0.653 | 0.989 | 0.992 |
| Sc4 | IVW | 1.6 | 0.8 | 0.8 | 0.5 | 0.3 | 0.147 | -0.288 | 0.435 | 0.436 | 0.105 | 0.331 | 0.187 | 0.392 | 0.378 | 0.378 | 0.396 | 0.382 | 0.057 | 0.236 | 0.332 | 0.333 | 0.168 | 0.255 | 1.000 | 0.232 | 0.849 | 0.659 | 0.400 | 0.430 |  |  |  |
|  |  | 0.6 | 0.8 | -0.2 | -0.5 | 0.3 | 0.035 | -0.615 | 0.650 | 0.650 | 0.723 | -0.073 | 0.071 | 0.156 | 0.147 | 0.147 | 0.143 | 0.146 | 0.006 | 0.403 | 0.444 | 0.444 | 0.543 | 0.027 | 1.000 | 0.231 | 0.811 | 0.627 | 0.363 | 0.374 |  |  |  |
|  |  | 0.7 | 0.5 | 0.2 | 0.5 | -0.3 | 0.024 | -0.293 | 0.317 | 0.317 | -0.246 | 0.563 | 0.081 | 0.173 | 0.167 | 0.167 | 0.165 | 0.169 | 0.007 | 0.116 | 0.128 | 0.128 | 0.088 | 0.346 | 1.000 | 0.201 | 0.800 | 0.615 | 0.351 | 0.384 |  |  |  |
|  |  | -0.3 | 0.5 | -0.8 | -0.5 | -0.3 | -0.080 | -0.610 | 0.529 | 0.529 | 0.367 | 0.162 | 0.049 | 0.106 | 0.100 | 0.100 | 0.097 | 0.097 | 0.009 | 0.383 | 0.290 | 0.290 | 0.144 | 0.036 | 1.000 | 0.201 | 0.695 | 0.500 | 0.300 | 0.298 |  |  |  |
|  |  | 0 | -0.8 | 0.8 | 0.5 | 0.3 | 0.363 | 0.906 | -0.543 | -0.543 | -0.374 | -0.169 | 0.060 | 0.138 | 0.129 | 0.129 | 0.124 | 0.130 | 0.135 | 0.840 | 0.312 | 0.312 | 0.155 | 0.045 | 1.000 | 0.112 | 0.393 | 0.233 | 0.183 | 0.196 |  |  |  |
|  |  | -1.6 | -0.8 | -0.8 | -0.5 | -0.3 | 0.095 | 0.341 | -0.246 | -0.246 | -0.010 | -0.236 | 0.168 | 0.346 | 0.334 | 0.334 | 0.349 | 0.352 | 0.037 | 0.236 | 0.172 | 0.172 | 0.122 | 0.180 | 1.000 | 0.250 | 0.818 | 0.625 | 0.361 | 0.404 |  |  |  |
|  |  | 0.8 | 0 | 0.8 | 0.5 | 0.3 | 0.254 | 0.319 | -0.065 | -0.064 | -0.141 | 0.077 | 0.115 | 0.247 | 0.240 | 0.240 | 0.245 | 0.243 | 0.078 | 0.163 | 0.062 | 0.062 | 0.080 | 0.065 | 1.000 | 0.245 | 0.809 | 0.636 | 0.380 | 0.386 |  |  |  |
|  |  | 1.3 | 0.8 | 0.5 | 0.5 | 0 | 0.064 | -0.409 | 0.472 | 0.473 | -0.026 | 0.499 | 0.151 | 0.301 | 0.291 | 0.291 | 0.300 | 0.301 | 0.027 | 0.258 | 0.308 | 0.308 | 0.091 | 0.339 | 1.000 | 0.227 | 0.844 | 0.670 | 0.377 | 0.428 |  |  |  |
|  |  | 1.3 | 0.8 | 0.5 | 0.5 | 0 | 0.194 | -0.260 | 0.453 | 0.454 | 0.159 | 0.295 | 0.168 | 0.330 | 0.309 | 0.309 | 0.318 | 0.250 | 0.066 | 0.176 | 0.301 | 0.301 | 0.126 | 0.149 | 1.000 | 0.363 | 0.768 | 0.714 | 0.593 | 0.252 |  |  |  |
|  |  | 0.8 | 0.8 | 0 | 0 | 0 | 0.016 | -0.561 | 0.577 | 0.577 | 0.278 | 0.299 | 0.091 | 0.187 | 0.179 | 0.179 | 0.187 | 0.180 | 0.009 | 0.350 | 0.365 | 0.365 | 0.112 | 0.122 | 1.000 | 0.229 | 0.841 | 0.671 | 0.352 | 0.418 |  |  |  |
|  | Egger | 1.6 | 0.8 | 0.8 | 0.5 | 0.3 | -1.613 | -0.810 | -0.803 | -0.803 | -0.501 | -0.302 | 0.406 | 0.425 | 0.130 | 0.130 | 0.088 | 0.097 | 2.766 | 0.837 | 0.662 | 0.662 | 0.258 | 0.101 | 0.060 | 0.058 | 0.000 | 0.000 | 0.002 | 0.004 | 0.671 | 0.997 | 0.998 |
|  |  | 0.6 | 0.8 | -0.2 | -0.5 | 0.3 | -0.598 | -0.796 | 0.198 | 0.198 | 0.499 | -0.301 | 0.157 | 0.165 | 0.050 | 0.050 | 0.037 | 0.035 | 0.382 | 0.661 | 0.042 | 0.042 | 0.250 | 0.092 | 0.052 | 0.046 | 0.000 | 0.005 | 0.003 | 0.004 | 0.569 | 0.999 | 0.999 |
|  |  | 0.7 | 0.5 | 0.2 | 0.5 | -0.3 | -0.705 | -0.506 | -0.199 | -0.199 | -0.500 | 0.301 | 0.177 | 0.187 | 0.057 | 0.057 | 0.042 | 0.039 | 0.528 | 0.291 | 0.043 | 0.043 | 0.252 | 0.092 | 0.049 | 0.047 | 0.000 | 0.002 | 0.006 | 0.001 | 0.603 | 0.999 | 0.996 |
|  |  | -0.3 | 0.5 | -0.8 | -0.5 | -0.3 | 0.298 | -0.502 | 0.801 | 0.801 | 0.500 | 0.301 | 0.110 | 0.117 | 0.034 | 0.034 | 0.023 | 0.025 | 0.101 | 0.266 | 0.642 | 0.642 | 0.250 | 0.091 | 0.052 | 0.053 | 0.000 | 0.002 | 0.000 | 0.003 | 0.500 | 1.000 | 0.998 |
|  |  | 0 | -0.8 | 0.8 | 0.5 | 0.3 | 0.000 | 0.801 | -0.801 | -0.801 | -0.501 | -0.300 | 0.155 | 0.159 | 0.046 | 0.046 | 0.032 | 0.034 | 0.024 | 0.667 | 0.644 | 0.643 | 0.252 | 0.091 | 0.058 | 0.050 | 0.000 | 0.000 | 0.001 | 0.003 | 0.255 | 0.998 | 1.000 |
|  |  | -1.6 | -0.8 | -0.8 | -0.5 | -0.3 | 1.562 | 0.761 | 0.801 | 0.801 | 0.498 | 0.303 | 0.351 | 0.373 | 0.118 | 0.118 | 0.083 | 0.085 | 2.565 | 0.719 | 0.656 | 0.656 | 0.255 | 0.099 | 0.048 | 0.048 | 0.000 | 0.005 | 0.004 | 0.004 | 0.592 | 0.998 | 0.996 |
|  |  | 0.8 | 0 | 0.8 | 0.5 | 0.3 | -0.793 | 0.007 | -0.800 | -0.800 | -0.500 | -0.300 | 0.250 | 0.262 | 0.081 | 0.081 | 0.058 | 0.055 | 0.691 | 0.069 | 0.647 | 0.647 | 0.253 | 0.093 | 0.058 | 0.047 | 0.000 | 0.002 | 0.003 | 0.002 | 0.636 | 0.995 | 0.996 |
|  |  | 1.3 | 0.8 | 0.5 | 0.5 | 0 | -1.305 | -0.808 | -0.497 | -0.497 | -0.499 | 0.002 | 0.306 | 0.316 | 0.101 | 0.101 | 0.069 | 0.074 | 1.797 | 0.753 | 0.257 | 0.257 | 0.254 | 0.005 | 0.048 | 0.048 | 0.000 | 0.002 | 0.002 | 0.004 | 0.655 | 0.996 | 0.996 |
|  |  | 1.3 | 0.8 | 0.5 | 0.5 | 0 | -1.301 | -0.808 | -0.494 | -0.494 | -0.497 | 0.003 | 0.347 | 0.364 | 0.106 | 0.106 | 0.073 | 0.076 | 1.814 | 0.785 | 0.255 | 0.255 | 0.252 | 0.006 | 0.052 | 0.053 | 0.000 | 0.002 | 0.001 | 0.002 | 0.744 | 0.900 | 0.998 |
|  |  | 0.8 | 0.8 | 0 | 0 | 0 | -0.802 | -0.801 | -0.001 | -0.001 | -0.001 | 0.000 | 0.188 | 0.200 | 0.060 | 0.060 | 0.041 | 0.040 | 0.679 | 0.681 | 0.004 | 0.004 | 0.002 | 0.002 | 0.051 | 0.050 | 0.000 | 0.002 | 0.003 | 0.002 | 0.661 | 1.000 | 0.997 |

TE: total effect; DE: direct effect; IE: indirect effect; IE_d: indirect effect calculated by difference method; IE_p: indirect effect calculated by product method; IE1: *X*→*M_1_*→*Y*; IE2: *X*→*M_2_*→*Y*.

**eTable 36** Simulation of heterogeneous populations with two ordered mediators

|  |  | TRUE | | | | | | BIAS | | | | | | | SE | | | | | | | MSE | | | | | | | Power/Type I error | | | | | | | | | |
| --- | --- | --- | --- | --- | --- | --- | --- | --- | --- | --- | --- | --- | --- | --- | --- | --- | --- | --- | --- | --- | --- | --- | --- | --- | --- | --- | --- | --- | --- | --- | --- | --- | --- | --- | --- | --- | --- | --- |
|  |  | TE | DE | IE | IE1 | IE2 | IE3 | TE | DE | IE_d | IE1 | IE2 | IE3 | IE_p | TE | DE | IE_d | IE1 | IE2 | IE3 | IE_p | TE | DE | IE_d | IE1 | IE2 | IE3 | IE_p | TE | DE | IE_d | IE1 | IE2 | IE3 | IE_p | GY | GM1 | GM2 |
| Standard MR | IVW | 1.8 | 0.8 | 1 | 0.5 | 0.3 | 0.2 | -0.172 | 0.390 | -0.562 | -0.562 | -0.209 | -0.182 | -0.172 | 0.502 | 0.959 | 0.813 | 0.813 | 0.705 | 0.453 | 0.185 | 0.281 | 1.073 | 0.977 | 0.977 | 0.540 | 0.238 | 0.064 | 0.867 | 0.198 | 0.013 | 0.044 | 0.059 | 0.020 | 0.006 |  |  |  |
|  |  | 0.8 | 0.8 | 0 | -0.5 | 0.3 | 0.2 | -0.074 | -0.254 | 0.180 | 0.180 | 0.620 | -0.252 | -0.188 | 0.242 | 0.484 | 0.428 | 0.428 | 0.354 | 0.221 | 0.087 | 0.064 | 0.299 | 0.215 | 0.215 | 0.510 | 0.113 | 0.043 | 0.822 | 0.203 | 0.018 | 0.051 | 0.079 | 0.024 | 0.003 |  |  |  |
|  |  | 1.2 | 0.8 | 0.4 | 0.5 | -0.3 | 0.2 | -0.127 | 0.067 | -0.195 | -0.195 | -0.302 | 0.302 | -0.194 | 0.297 | 0.509 | 0.419 | 0.419 | 0.408 | 0.140 | 0.105 | 0.105 | 0.264 | 0.213 | 0.213 | 0.258 | 0.111 | 0.049 | 0.928 | 0.340 | 0.009 | 0.043 | 0.072 | 0.004 | 0.003 |  |  |  |
|  |  | 0.2 | 0.8 | -0.6 | -0.5 | -0.3 | 0.2 | -0.020 | -0.653 | 0.632 | 0.633 | 0.530 | 0.303 | -0.201 | 0.135 | 0.244 | 0.199 | 0.199 | 0.193 | 0.061 | 0.042 | 0.019 | 0.486 | 0.440 | 0.440 | 0.318 | 0.096 | 0.042 | 0.209 | 0.073 | 0.009 | 0.035 | 0.060 | 0.003 | 0.001 |  |  |  |
|  |  | 0.2 | -0.8 | 1 | 0.5 | 0.3 | 0.2 | -0.003 | 0.954 | -0.957 | -0.957 | -0.480 | -0.273 | -0.204 | 0.356 | 0.649 | 0.550 | 0.550 | 0.465 | 0.307 | 0.120 | 0.127 | 1.330 | 1.218 | 1.218 | 0.446 | 0.169 | 0.056 | 0.058 | 0.036 | 0.004 | 0.020 | 0.033 | 0.025 | 0.001 |  |  |  |
|  |  | -1.8 | -0.8 | -1 | -0.5 | -0.3 | -0.2 | 0.171 | -0.418 | 0.589 | 0.588 | 0.231 | 0.182 | 0.176 | 0.378 | 0.718 | 0.625 | 0.625 | 0.534 | 0.365 | 0.140 | 0.172 | 0.690 | 0.737 | 0.737 | 0.339 | 0.166 | 0.051 | 0.988 | 0.385 | 0.027 | 0.073 | 0.105 | 0.041 | 0.008 |  |  |  |
|  |  | 1 | 0 | 1 | 0.5 | 0.3 | 0.2 | -0.097 | 0.693 | -0.790 | -0.790 | -0.378 | -0.233 | -0.179 | 0.427 | 0.810 | 0.690 | 0.690 | 0.602 | 0.406 | 0.158 | 0.192 | 1.137 | 1.101 | 1.101 | 0.505 | 0.219 | 0.057 | 0.515 | 0.111 | 0.013 | 0.030 | 0.062 | 0.027 | 0.005 |  |  |  |
|  |  | 1.3 | 0.8 | 0.5 | 0 | 0.3 | 0.2 | -0.125 | 0.083 | -0.209 | -0.209 | 0.182 | -0.208 | -0.182 | 0.352 | 0.684 | 0.608 | 0.608 | 0.506 | 0.353 | 0.134 | 0.139 | 0.474 | 0.413 | 0.413 | 0.289 | 0.168 | 0.051 | 0.879 | 0.213 | 0.017 | 0.057 | 0.063 | 0.037 | 0.005 |  |  |  |
|  |  | 1.5 | 0.8 | 0.7 | 0.5 | 0 | 0.2 | -0.137 | 0.289 | -0.427 | -0.426 | -0.257 | 0.025 | -0.194 | 0.453 | 0.802 | 0.678 | 0.678 | 0.652 | 0.252 | 0.145 | 0.224 | 0.726 | 0.642 | 0.642 | 0.491 | 0.064 | 0.059 | 0.786 | 0.213 | 0.007 | 0.033 | 0.061 | 0.012 | 0.001 |  |  |  |
|  |  | 1 | 0.8 | 0.2 | 0 | 0 | 0.2 | -0.111 | -0.060 | -0.050 | -0.050 | 0.143 | 0.006 | -0.199 | 0.323 | 0.528 | 0.441 | 0.441 | 0.427 | 0.174 | 0.098 | 0.117 | 0.283 | 0.197 | 0.197 | 0.203 | 0.030 | 0.049 | 0.734 | 0.197 | 0.002 | 0.016 | 0.049 | 0.010 | 0.005 |  |  |  |
|  |  | 0 | -1 | 1 | 0.5 | 0.3 | 0.2 | 0.016 | 1.032 | -1.016 | -1.016 | -0.508 | -0.305 | -0.203 | 0.340 | 0.655 | 0.577 | 0.577 | 0.488 | 0.312 | 0.113 | 0.116 | 1.494 | 1.365 | 1.365 | 0.496 | 0.190 | 0.054 | 0.033 | 0.038 | 0.017 | 0.043 | 0.059 | 0.029 | 0.005 |  |  |  |
| Sc2 | IVW | 1.8 | 0.8 | 1 | 0.5 | 0.3 | 0.2 | -0.064 | 0.568 | -0.632 | -0.632 | -0.288 | -0.162 | -0.182 | 0.179 | 0.468 | 0.434 | 0.434 | 0.344 | 0.260 | 0.087 | 0.036 | 0.542 | 0.588 | 0.588 | 0.201 | 0.094 | 0.041 | 1.000 | 0.805 | 0.082 | 0.115 | 0.123 | 0.072 | 0.005 |  |  |  |
|  |  | 0.8 | 0.8 | 0 | -0.5 | 0.3 | 0.2 | -0.028 | -0.176 | 0.148 | 0.149 | 0.581 | -0.241 | -0.191 | 0.088 | 0.217 | 0.201 | 0.201 | 0.168 | 0.124 | 0.043 | 0.009 | 0.078 | 0.062 | 0.062 | 0.365 | 0.073 | 0.038 | 1.000 | 0.779 | 0.060 | 0.079 | 0.101 | 0.064 | 0.004 |  |  |  |
|  |  | 1.2 | 0.8 | 0.4 | 0.5 | -0.3 | 0.2 | -0.050 | 0.195 | -0.245 | -0.245 | -0.348 | 0.302 | -0.199 | 0.109 | 0.233 | 0.209 | 0.209 | 0.198 | 0.077 | 0.044 | 0.014 | 0.092 | 0.104 | 0.104 | 0.160 | 0.097 | 0.041 | 1.000 | 0.977 | 0.045 | 0.080 | 0.129 | 0.008 | 0.001 |  |  |  |
|  |  | 0.2 | 0.8 | -0.6 | -0.5 | -0.3 | 0.2 | -0.008 | -0.622 | 0.614 | 0.614 | 0.517 | 0.299 | -0.201 | 0.044 | 0.100 | 0.091 | 0.091 | 0.087 | 0.031 | 0.019 | 0.002 | 0.397 | 0.385 | 0.385 | 0.275 | 0.090 | 0.041 | 0.983 | 0.335 | 0.013 | 0.022 | 0.049 | 0.005 | 0.001 |  |  |  |
|  |  | 0.2 | -0.8 | 1 | 0.5 | 0.3 | 0.2 | -0.007 | 0.921 | -0.928 | -0.928 | -0.457 | -0.277 | -0.195 | 0.118 | 0.300 | 0.276 | 0.276 | 0.222 | 0.172 | 0.052 | 0.014 | 0.939 | 0.938 | 0.938 | 0.258 | 0.106 | 0.041 | 0.323 | 0.053 | 0.016 | 0.032 | 0.054 | 0.038 | 0.001 |  |  |  |
|  |  | -1.8 | -0.8 | -1 | -0.5 | -0.3 | -0.2 | 0.075 | -0.560 | 0.635 | 0.635 | 0.289 | 0.168 | 0.179 | 0.136 | 0.344 | 0.318 | 0.318 | 0.251 | 0.198 | 0.076 | 0.024 | 0.432 | 0.505 | 0.505 | 0.146 | 0.067 | 0.038 | 1.000 | 0.959 | 0.130 | 0.173 | 0.152 | 0.082 | 0.009 |  |  |  |
|  |  | 1 | 0 | 1 | 0.5 | 0.3 | 0.2 | -0.032 | 0.761 | -0.793 | -0.793 | -0.379 | -0.227 | -0.188 | 0.146 | 0.372 | 0.340 | 0.341 | 0.273 | 0.213 | 0.067 | 0.022 | 0.718 | 0.745 | 0.745 | 0.218 | 0.097 | 0.040 | 1.000 | 0.523 | 0.041 | 0.071 | 0.076 | 0.056 | 0.001 |  |  |  |
|  |  | 1.3 | 0.8 | 0.5 | 0 | 0.3 | 0.2 | -0.052 | 0.212 | -0.264 | -0.264 | 0.129 | -0.204 | -0.189 | 0.127 | 0.321 | 0.303 | 0.303 | 0.246 | 0.186 | 0.059 | 0.019 | 0.148 | 0.162 | 0.162 | 0.077 | 0.076 | 0.039 | 1.000 | 0.840 | 0.060 | 0.082 | 0.103 | 0.071 | 0.002 |  |  |  |
|  |  | 1.5 | 0.8 | 0.7 | 0.5 | 0 | 0.2 | -0.058 | 0.429 | -0.487 | -0.487 | -0.319 | 0.027 | -0.194 | 0.165 | 0.355 | 0.315 | 0.315 | 0.293 | 0.135 | 0.069 | 0.030 | 0.310 | 0.336 | 0.336 | 0.188 | 0.019 | 0.042 | 1.000 | 0.891 | 0.038 | 0.063 | 0.082 | 0.015 | 0.001 |  |  |  |
|  |  | 1 | 0.8 | 0.2 | 0 | 0 | 0.2 | -0.035 | 0.028 | -0.062 | -0.062 | 0.116 | 0.016 | -0.195 | 0.114 | 0.252 | 0.232 | 0.232 | 0.223 | 0.088 | 0.047 | 0.014 | 0.064 | 0.058 | 0.058 | 0.063 | 0.008 | 0.040 | 1.000 | 0.863 | 0.031 | 0.064 | 0.098 | 0.011 | 0.003 |  |  |  |
|  |  | 0 | -1 | 1 | 0.5 | 0.3 | 0.2 | 0.010 | 0.981 | -0.970 | -0.970 | -0.493 | -0.279 | -0.198 | 0.109 | 0.289 | 0.266 | 0.266 | 0.214 | 0.172 | 0.050 | 0.012 | 1.045 | 1.012 | 1.012 | 0.289 | 0.108 | 0.042 | 0.026 | 0.032 | 0.018 | 0.030 | 0.042 | 0.036 | 0.003 |  |  |  |
|  | Egger | 1.8 | 0.8 | 1 | 0.5 | 0.3 | 0.2 | -0.331 | 0.385 | -0.716 | -0.716 | -0.341 | -0.187 | -0.188 | 0.410 | 0.574 | 0.395 | 0.395 | 0.302 | 0.254 | 0.075 | 0.278 | 0.478 | 0.669 | 0.669 | 0.207 | 0.100 | 0.041 | 0.922 | 0.539 | 0.008 | 0.059 | 0.083 | 0.028 | 0.002 | 0.083 | 0.060 | 0.087 |
|  |  | 0.8 | 0.8 | 0 | -0.5 | 0.3 | 0.2 | -0.140 | -0.248 | 0.108 | 0.108 | 0.560 | -0.258 | -0.195 | 0.199 | 0.272 | 0.184 | 0.184 | 0.155 | 0.115 | 0.039 | 0.059 | 0.135 | 0.046 | 0.046 | 0.338 | 0.080 | 0.039 | 0.908 | 0.508 | 0.005 | 0.043 | 0.067 | 0.027 | 0.001 | 0.071 | 0.077 | 0.089 |
|  |  | 1.2 | 0.8 | 0.4 | 0.5 | -0.3 | 0.2 | -0.223 | 0.065 | -0.289 | -0.289 | -0.387 | 0.297 | -0.199 | 0.251 | 0.309 | 0.196 | 0.196 | 0.181 | 0.087 | 0.037 | 0.113 | 0.100 | 0.122 | 0.122 | 0.182 | 0.096 | 0.041 | 0.966 | 0.748 | 0.002 | 0.023 | 0.080 | 0.004 | 0.000 | 0.081 | 0.029 | 0.079 |
|  |  | 0.2 | 0.8 | -0.6 | -0.5 | -0.3 | 0.2 | -0.044 | -0.654 | 0.610 | 0.610 | 0.512 | 0.299 | -0.201 | 0.101 | 0.133 | 0.085 | 0.085 | 0.080 | 0.036 | 0.018 | 0.012 | 0.445 | 0.379 | 0.379 | 0.268 | 0.091 | 0.041 | 0.239 | 0.141 | 0.001 | 0.012 | 0.039 | 0.003 | 0.002 | 0.040 | 0.039 | 0.088 |
|  |  | 0.2 | -0.8 | 1 | 0.5 | 0.3 | 0.2 | -0.045 | 0.897 | -0.942 | -0.942 | -0.466 | -0.280 | -0.196 | 0.275 | 0.385 | 0.264 | 0.264 | 0.205 | 0.169 | 0.049 | 0.078 | 0.953 | 0.957 | 0.957 | 0.259 | 0.107 | 0.041 | 0.067 | 0.041 | 0.000 | 0.022 | 0.045 | 0.021 | 0.001 | 0.036 | 0.054 | 0.083 |
|  |  | -1.8 | -0.8 | -1 | -0.5 | -0.3 | -0.2 | 0.336 | -0.390 | 0.726 | 0.726 | 0.342 | 0.200 | 0.185 | 0.302 | 0.416 | 0.294 | 0.294 | 0.226 | 0.190 | 0.065 | 0.204 | 0.326 | 0.614 | 0.614 | 0.168 | 0.076 | 0.038 | 0.995 | 0.770 | 0.006 | 0.096 | 0.094 | 0.033 | 0.005 | 0.102 | 0.057 | 0.086 |
|  |  | 1 | 0 | 1 | 0.5 | 0.3 | 0.2 | -0.189 | 0.653 | -0.842 | -0.842 | -0.409 | -0.243 | -0.190 | 0.334 | 0.467 | 0.314 | 0.314 | 0.252 | 0.199 | 0.064 | 0.147 | 0.645 | 0.808 | 0.808 | 0.231 | 0.099 | 0.040 | 0.641 | 0.261 | 0.001 | 0.026 | 0.053 | 0.019 | 0.003 | 0.045 | 0.052 | 0.082 |
|  |  | 1.3 | 0.8 | 0.5 | 0 | 0.3 | 0.2 | -0.237 | 0.087 | -0.324 | -0.324 | 0.094 | -0.224 | -0.194 | 0.295 | 0.395 | 0.278 | 0.278 | 0.223 | 0.174 | 0.051 | 0.143 | 0.164 | 0.183 | 0.183 | 0.059 | 0.081 | 0.040 | 0.935 | 0.558 | 0.003 | 0.048 | 0.071 | 0.029 | 0.000 | 0.076 | 0.051 | 0.078 |
|  |  | 1.5 | 0.8 | 0.7 | 0.5 | 0 | 0.2 | -0.292 | 0.250 | -0.542 | -0.542 | -0.367 | 0.021 | -0.196 | 0.377 | 0.471 | 0.304 | 0.304 | 0.273 | 0.141 | 0.065 | 0.227 | 0.284 | 0.386 | 0.386 | 0.209 | 0.020 | 0.043 | 0.863 | 0.535 | 0.000 | 0.031 | 0.057 | 0.004 | 0.001 | 0.069 | 0.037 | 0.080 |
|  |  | 1 | 0.8 | 0.2 | 0 | 0 | 0.2 | -0.186 | -0.087 | -0.099 | -0.098 | 0.084 | 0.014 | -0.196 | 0.257 | 0.325 | 0.210 | 0.210 | 0.196 | 0.094 | 0.042 | 0.100 | 0.113 | 0.054 | 0.054 | 0.046 | 0.009 | 0.040 | 0.821 | 0.524 | 0.000 | 0.024 | 0.071 | 0.003 | 0.001 | 0.059 | 0.028 | 0.070 |
|  |  | 0 | -1 | 1 | 0.5 | 0.3 | 0.2 | 0.018 | 0.987 | -0.969 | -0.969 | -0.489 | -0.281 | -0.199 | 0.256 | 0.357 | 0.244 | 0.244 | 0.190 | 0.164 | 0.043 | 0.066 | 1.101 | 0.999 | 0.999 | 0.276 | 0.106 | 0.041 | 0.034 | 0.029 | 0.000 | 0.019 | 0.036 | 0.026 | 0.001 | 0.035 | 0.064 | 0.081 |
| Sc3 | IVW | 1.8 | 0.8 | 1 | 0.5 | 0.3 | 0.2 | 0.050 | 0.646 | -0.596 | -0.596 | -0.261 | -0.160 | -0.174 | 0.186 | 0.489 | 0.449 | 0.449 | 0.353 | 0.273 | 0.094 | 0.037 | 0.656 | 0.556 | 0.556 | 0.193 | 0.100 | 0.039 | 1.000 | 0.803 | 0.080 | 0.103 | 0.097 | 0.070 | 0.006 |  |  |  |
|  |  | 0.8 | 0.8 | 0 | -0.5 | 0.3 | 0.2 | 0.024 | -0.152 | 0.176 | 0.176 | 0.593 | -0.229 | -0.188 | 0.088 | 0.226 | 0.207 | 0.207 | 0.169 | 0.129 | 0.042 | 0.008 | 0.074 | 0.074 | 0.074 | 0.380 | 0.069 | 0.037 | 1.000 | 0.771 | 0.066 | 0.102 | 0.100 | 0.066 | 0.002 |  |  |  |
|  |  | 1.2 | 0.8 | 0.4 | 0.5 | -0.3 | 0.2 | 0.009 | 0.222 | -0.213 | -0.213 | -0.326 | 0.311 | -0.198 | 0.113 | 0.241 | 0.224 | 0.224 | 0.212 | 0.083 | 0.049 | 0.013 | 0.108 | 0.095 | 0.095 | 0.151 | 0.104 | 0.042 | 1.000 | 0.961 | 0.047 | 0.086 | 0.136 | 0.006 | 0.001 |  |  |  |
|  |  | 0.2 | 0.8 | -0.6 | -0.5 | -0.3 | 0.2 | -0.032 | -0.653 | 0.621 | 0.621 | 0.521 | 0.300 | -0.200 | 0.047 | 0.105 | 0.097 | 0.097 | 0.093 | 0.036 | 0.020 | 0.003 | 0.437 | 0.395 | 0.395 | 0.280 | 0.091 | 0.041 | 0.929 | 0.225 | 0.021 | 0.031 | 0.053 | 0.005 | 0.003 |  |  |  |
|  |  | 0.2 | -0.8 | 1 | 0.5 | 0.3 | 0.2 | 0.113 | 1.021 | -0.908 | -0.908 | -0.439 | -0.275 | -0.194 | 0.112 | 0.311 | 0.295 | 0.295 | 0.236 | 0.188 | 0.060 | 0.025 | 1.140 | 0.911 | 0.911 | 0.249 | 0.111 | 0.041 | 0.709 | 0.081 | 0.026 | 0.039 | 0.053 | 0.039 | 0.003 |  |  |  |
|  |  | -1.8 | -0.8 | -1 | -0.5 | -0.3 | -0.2 | 0.013 | -0.602 | 0.615 | 0.615 | 0.282 | 0.160 | 0.173 | 0.139 | 0.362 | 0.334 | 0.334 | 0.259 | 0.218 | 0.075 | 0.019 | 0.494 | 0.490 | 0.490 | 0.147 | 0.073 | 0.035 | 1.000 | 0.947 | 0.150 | 0.178 | 0.128 | 0.095 | 0.008 |  |  |  |
|  |  | 1 | 0 | 1 | 0.5 | 0.3 | 0.2 | 0.083 | 0.821 | -0.738 | -0.738 | -0.359 | -0.198 | -0.182 | 0.142 | 0.401 | 0.374 | 0.374 | 0.301 | 0.242 | 0.076 | 0.027 | 0.835 | 0.685 | 0.685 | 0.219 | 0.098 | 0.039 | 1.000 | 0.531 | 0.063 | 0.081 | 0.075 | 0.058 | 0.003 |  |  |  |
|  |  | 1.3 | 0.8 | 0.5 | 0 | 0.3 | 0.2 | 0.042 | 0.262 | -0.220 | -0.220 | 0.163 | -0.201 | -0.182 | 0.129 | 0.346 | 0.320 | 0.320 | 0.251 | 0.201 | 0.066 | 0.018 | 0.188 | 0.151 | 0.151 | 0.090 | 0.081 | 0.038 | 1.000 | 0.835 | 0.076 | 0.105 | 0.092 | 0.061 | 0.002 |  |  |  |
|  |  | 1.5 | 0.8 | 0.7 | 0.5 | 0 | 0.2 | 0.056 | 0.463 | -0.407 | -0.407 | -0.264 | 0.048 | -0.191 | 0.172 | 0.403 | 0.367 | 0.367 | 0.339 | 0.154 | 0.077 | 0.033 | 0.377 | 0.301 | 0.301 | 0.185 | 0.026 | 0.043 | 1.000 | 0.843 | 0.061 | 0.107 | 0.124 | 0.010 | 0.004 |  |  |  |
|  |  | 1 | 0.8 | 0.2 | 0 | 0 | 0.2 | 0.053 | 0.062 | -0.009 | -0.009 | 0.153 | 0.034 | -0.196 | 0.111 | 0.274 | 0.255 | 0.255 | 0.238 | 0.106 | 0.053 | 0.015 | 0.079 | 0.065 | 0.065 | 0.080 | 0.012 | 0.041 | 1.000 | 0.829 | 0.050 | 0.077 | 0.111 | 0.014 | 0.002 |  |  |  |
|  |  | 0 | -1 | 1 | 0.5 | 0.3 | 0.2 | 0.117 | 1.076 | -0.959 | -0.959 | -0.476 | -0.287 | -0.197 | 0.108 | 0.307 | 0.285 | 0.285 | 0.230 | 0.178 | 0.057 | 0.025 | 1.253 | 1.002 | 1.002 | 0.279 | 0.114 | 0.042 | 0.136 | 0.040 | 0.018 | 0.028 | 0.056 | 0.035 | 0.000 |  |  |  |
|  | Egger | 1.8 | 0.8 | 1 | 0.5 | 0.3 | 0.2 | -0.348 | 0.422 | -0.771 | -0.771 | -0.359 | -0.222 | -0.189 | 0.398 | 0.558 | 0.390 | 0.390 | 0.298 | 0.238 | 0.068 | 0.279 | 0.490 | 0.746 | 0.746 | 0.218 | 0.106 | 0.040 | 0.923 | 0.560 | 0.002 | 0.039 | 0.053 | 0.016 | 0.001 | 0.115 | 0.116 | 0.149 |
|  |  | 0.8 | 0.8 | 0 | -0.5 | 0.3 | 0.2 | -0.152 | -0.253 | 0.101 | 0.101 | 0.555 | -0.258 | -0.196 | 0.192 | 0.264 | 0.176 | 0.176 | 0.142 | 0.110 | 0.032 | 0.060 | 0.134 | 0.041 | 0.041 | 0.328 | 0.079 | 0.039 | 0.888 | 0.496 | 0.000 | 0.043 | 0.059 | 0.022 | 0.001 | 0.093 | 0.110 | 0.130 |
|  |  | 1.2 | 0.8 | 0.4 | 0.5 | -0.3 | 0.2 | -0.217 | 0.069 | -0.286 | -0.286 | -0.391 | 0.304 | -0.198 | 0.235 | 0.296 | 0.197 | 0.197 | 0.182 | 0.085 | 0.041 | 0.102 | 0.093 | 0.120 | 0.120 | 0.186 | 0.100 | 0.041 | 0.967 | 0.757 | 0.000 | 0.019 | 0.078 | 0.006 | 0.001 | 0.098 | 0.054 | 0.153 |
|  |  | 0.2 | 0.8 | -0.6 | -0.5 | -0.3 | 0.2 | -0.042 | -0.659 | 0.617 | 0.617 | 0.517 | 0.301 | -0.201 | 0.105 | 0.133 | 0.086 | 0.086 | 0.080 | 0.038 | 0.018 | 0.013 | 0.452 | 0.388 | 0.388 | 0.273 | 0.092 | 0.041 | 0.280 | 0.145 | 0.000 | 0.017 | 0.043 | 0.004 | 0.001 | 0.040 | 0.055 | 0.138 |
|  |  | 0.2 | -0.8 | 1 | 0.5 | 0.3 | 0.2 | -0.035 | 0.927 | -0.962 | -0.962 | -0.474 | -0.291 | -0.197 | 0.264 | 0.364 | 0.256 | 0.256 | 0.197 | 0.169 | 0.049 | 0.071 | 0.992 | 0.991 | 0.991 | 0.264 | 0.113 | 0.041 | 0.072 | 0.039 | 0.001 | 0.019 | 0.038 | 0.015 | 0.002 | 0.067 | 0.110 | 0.155 |
|  |  | -1.8 | -0.8 | -1 | -0.5 | -0.3 | -0.2 | 0.347 | -0.423 | 0.769 | 0.769 | 0.368 | 0.214 | 0.188 | 0.300 | 0.416 | 0.290 | 0.290 | 0.214 | 0.196 | 0.059 | 0.210 | 0.352 | 0.676 | 0.676 | 0.181 | 0.084 | 0.039 | 0.995 | 0.817 | 0.007 | 0.063 | 0.076 | 0.045 | 0.003 | 0.128 | 0.108 | 0.151 |
|  |  | 1 | 0 | 1 | 0.5 | 0.3 | 0.2 | -0.183 | 0.665 | -0.848 | -0.848 | -0.417 | -0.240 | -0.191 | 0.338 | 0.471 | 0.324 | 0.324 | 0.255 | 0.211 | 0.058 | 0.148 | 0.664 | 0.824 | 0.824 | 0.239 | 0.102 | 0.040 | 0.637 | 0.283 | 0.002 | 0.030 | 0.054 | 0.024 | 0.002 | 0.090 | 0.093 | 0.130 |
|  |  | 1.3 | 0.8 | 0.5 | 0 | 0.3 | 0.2 | -0.235 | 0.102 | -0.337 | -0.337 | 0.096 | -0.240 | -0.192 | 0.287 | 0.396 | 0.279 | 0.279 | 0.208 | 0.180 | 0.050 | 0.138 | 0.167 | 0.191 | 0.191 | 0.053 | 0.090 | 0.040 | 0.928 | 0.578 | 0.002 | 0.043 | 0.055 | 0.023 | 0.000 | 0.107 | 0.108 | 0.163 |
|  |  | 1.5 | 0.8 | 0.7 | 0.5 | 0 | 0.2 | -0.303 | 0.227 | -0.530 | -0.530 | -0.359 | 0.025 | -0.196 | 0.357 | 0.468 | 0.312 | 0.312 | 0.288 | 0.146 | 0.062 | 0.219 | 0.271 | 0.379 | 0.378 | 0.212 | 0.022 | 0.042 | 0.858 | 0.503 | 0.000 | 0.027 | 0.078 | 0.007 | 0.002 | 0.106 | 0.074 | 0.158 |
|  |  | 1 | 0.8 | 0.2 | 0 | 0 | 0.2 | -0.198 | -0.100 | -0.098 | -0.098 | 0.087 | 0.015 | -0.200 | 0.250 | 0.323 | 0.213 | 0.213 | 0.193 | 0.094 | 0.043 | 0.102 | 0.114 | 0.055 | 0.055 | 0.045 | 0.009 | 0.042 | 0.818 | 0.493 | 0.000 | 0.016 | 0.062 | 0.003 | 0.000 | 0.115 | 0.066 | 0.147 |
|  |  | 0 | -1 | 1 | 0.5 | 0.3 | 0.2 | -0.004 | 0.993 | -0.997 | -0.997 | -0.496 | -0.301 | -0.200 | 0.258 | 0.356 | 0.247 | 0.247 | 0.199 | 0.157 | 0.046 | 0.067 | 1.112 | 1.055 | 1.055 | 0.285 | 0.116 | 0.042 | 0.038 | 0.035 | 0.000 | 0.016 | 0.044 | 0.016 | 0.000 | 0.052 | 0.101 | 0.149 |
| Sc4 | IVW | 1.8 | 0.8 | 1 | 0.5 | 0.3 | 0.2 | 0.346 | 0.595 | -0.249 | -0.249 | -0.111 | -0.036 | -0.102 | 0.167 | 0.504 | 0.482 | 0.482 | 0.401 | 0.293 | 0.149 | 0.148 | 0.609 | 0.295 | 0.295 | 0.173 | 0.087 | 0.033 | 1.000 | 0.747 | 0.272 | 0.266 | 0.186 | 0.129 | 0.031 |  |  |  |
|  |  | 0.8 | 0.8 | 0 | -0.5 | 0.3 | 0.2 | 0.151 | -0.176 | 0.327 | 0.327 | 0.667 | -0.179 | -0.161 | 0.082 | 0.246 | 0.238 | 0.238 | 0.200 | 0.150 | 0.073 | 0.029 | 0.092 | 0.164 | 0.164 | 0.485 | 0.055 | 0.031 | 1.000 | 0.678 | 0.219 | 0.214 | 0.139 | 0.113 | 0.016 |  |  |  |
|  |  | 1.2 | 0.8 | 0.4 | 0.5 | -0.3 | 0.2 | 0.132 | 0.218 | -0.086 | -0.086 | -0.231 | 0.332 | -0.188 | 0.107 | 0.271 | 0.252 | 0.252 | 0.236 | 0.099 | 0.059 | 0.029 | 0.121 | 0.071 | 0.071 | 0.109 | 0.120 | 0.039 | 1.000 | 0.924 | 0.158 | 0.178 | 0.214 | 0.018 | 0.003 |  |  |  |
|  |  | 0.2 | 0.8 | -0.6 | -0.5 | -0.3 | 0.2 | -0.080 | -0.673 | 0.593 | 0.593 | 0.500 | 0.295 | -0.202 | 0.042 | 0.117 | 0.109 | 0.109 | 0.103 | 0.040 | 0.025 | 0.008 | 0.467 | 0.364 | 0.364 | 0.261 | 0.089 | 0.041 | 0.766 | 0.175 | 0.026 | 0.029 | 0.044 | 0.014 | 0.002 |  |  |  |
|  |  | 0.2 | -0.8 | 1 | 0.5 | 0.3 | 0.2 | 0.403 | 1.046 | -0.643 | -0.643 | -0.322 | -0.165 | -0.155 | 0.116 | 0.355 | 0.339 | 0.339 | 0.273 | 0.207 | 0.096 | 0.176 | 1.220 | 0.528 | 0.528 | 0.178 | 0.070 | 0.033 | 1.000 | 0.095 | 0.133 | 0.136 | 0.097 | 0.085 | 0.016 |  |  |  |
|  |  | -1.8 | -0.8 | -1 | -0.5 | -0.3 | -0.2 | -0.117 | -0.540 | 0.423 | 0.423 | 0.174 | 0.114 | 0.135 | 0.134 | 0.384 | 0.365 | 0.365 | 0.312 | 0.223 | 0.114 | 0.032 | 0.439 | 0.312 | 0.312 | 0.128 | 0.063 | 0.031 | 1.000 | 0.894 | 0.289 | 0.272 | 0.211 | 0.125 | 0.025 |  |  |  |
|  |  | 1 | 0 | 1 | 0.5 | 0.3 | 0.2 | 0.386 | 0.822 | -0.436 | -0.435 | -0.210 | -0.093 | -0.133 | 0.140 | 0.423 | 0.404 | 0.404 | 0.329 | 0.260 | 0.119 | 0.169 | 0.854 | 0.353 | 0.353 | 0.152 | 0.076 | 0.032 | 1.000 | 0.433 | 0.206 | 0.215 | 0.134 | 0.114 | 0.020 |  |  |  |
|  |  | 1.3 | 0.8 | 0.5 | 0 | 0.3 | 0.2 | 0.248 | 0.196 | 0.052 | 0.052 | 0.284 | -0.098 | -0.134 | 0.125 | 0.368 | 0.353 | 0.353 | 0.279 | 0.225 | 0.109 | 0.077 | 0.173 | 0.127 | 0.127 | 0.158 | 0.060 | 0.030 | 1.000 | 0.717 | 0.278 | 0.275 | 0.155 | 0.139 | 0.030 |  |  |  |
|  |  | 1.5 | 0.8 | 0.7 | 0.5 | 0 | 0.2 | 0.370 | 0.498 | -0.128 | -0.128 | -0.104 | 0.120 | -0.144 | 0.163 | 0.450 | 0.428 | 0.428 | 0.389 | 0.190 | 0.119 | 0.164 | 0.451 | 0.200 | 0.200 | 0.162 | 0.051 | 0.035 | 1.000 | 0.808 | 0.234 | 0.235 | 0.200 | 0.041 | 0.010 |  |  |  |
|  |  | 1 | 0.8 | 0.2 | 0 | 0 | 0.2 | 0.255 | 0.067 | 0.188 | 0.188 | 0.263 | 0.090 | -0.165 | 0.110 | 0.299 | 0.279 | 0.279 | 0.261 | 0.140 | 0.088 | 0.077 | 0.094 | 0.114 | 0.114 | 0.137 | 0.028 | 0.035 | 1.000 | 0.775 | 0.186 | 0.182 | 0.170 | 0.058 | 0.015 |  |  |  |
|  |  | 0 | -1 | 1 | 0.5 | 0.3 | 0.2 | 0.401 | 1.083 | -0.681 | -0.681 | -0.346 | -0.176 | -0.159 | 0.111 | 0.340 | 0.314 | 0.314 | 0.266 | 0.193 | 0.087 | 0.173 | 1.288 | 0.563 | 0.563 | 0.191 | 0.068 | 0.033 | 0.931 | 0.051 | 0.121 | 0.117 | 0.095 | 0.090 | 0.016 |  |  |  |
|  | Egger | 1.8 | 0.8 | 1 | 0.5 | 0.3 | 0.2 | -0.276 | 0.367 | -0.643 | -0.643 | -0.309 | -0.165 | -0.168 | 0.408 | 0.548 | 0.383 | 0.383 | 0.305 | 0.242 | 0.090 | 0.242 | 0.434 | 0.560 | 0.560 | 0.189 | 0.086 | 0.036 | 0.936 | 0.518 | 0.003 | 0.075 | 0.093 | 0.036 | 0.005 | 0.174 | 0.208 | 0.320 |
|  |  | 0.8 | 0.8 | 0 | -0.5 | 0.3 | 0.2 | -0.127 | -0.281 | 0.154 | 0.154 | 0.581 | -0.238 | -0.189 | 0.192 | 0.267 | 0.191 | 0.191 | 0.153 | 0.123 | 0.044 | 0.053 | 0.150 | 0.060 | 0.060 | 0.361 | 0.072 | 0.038 | 0.908 | 0.456 | 0.004 | 0.080 | 0.080 | 0.042 | 0.003 | 0.143 | 0.235 | 0.320 |
|  |  | 1.2 | 0.8 | 0.4 | 0.5 | -0.3 | 0.2 | -0.191 | 0.056 | -0.248 | -0.248 | -0.362 | 0.311 | -0.197 | 0.254 | 0.321 | 0.199 | 0.199 | 0.184 | 0.081 | 0.041 | 0.101 | 0.106 | 0.101 | 0.101 | 0.165 | 0.104 | 0.041 | 0.966 | 0.743 | 0.003 | 0.044 | 0.091 | 0.003 | 0.003 | 0.166 | 0.099 | 0.323 |
|  |  | 0.2 | 0.8 | -0.6 | -0.5 | -0.3 | 0.2 | -0.052 | -0.655 | 0.603 | 0.603 | 0.506 | 0.297 | -0.201 | 0.106 | 0.139 | 0.087 | 0.087 | 0.081 | 0.037 | 0.019 | 0.014 | 0.448 | 0.371 | 0.371 | 0.263 | 0.090 | 0.041 | 0.254 | 0.153 | 0.000 | 0.019 | 0.033 | 0.003 | 0.001 | 0.046 | 0.115 | 0.289 |
|  |  | 0.2 | -0.8 | 1 | 0.5 | 0.3 | 0.2 | 0.029 | 0.889 | -0.860 | -0.860 | -0.429 | -0.242 | -0.189 | 0.273 | 0.384 | 0.266 | 0.266 | 0.202 | 0.169 | 0.057 | 0.075 | 0.938 | 0.811 | 0.811 | 0.225 | 0.087 | 0.039 | 0.106 | 0.050 | 0.002 | 0.042 | 0.041 | 0.033 | 0.001 | 0.146 | 0.222 | 0.315 |
|  |  | -1.8 | -0.8 | -1 | -0.5 | -0.3 | -0.2 | 0.310 | -0.396 | 0.707 | 0.707 | 0.319 | 0.209 | 0.179 | 0.300 | 0.411 | 0.299 | 0.299 | 0.249 | 0.182 | 0.072 | 0.186 | 0.326 | 0.589 | 0.589 | 0.164 | 0.077 | 0.037 | 0.998 | 0.758 | 0.006 | 0.102 | 0.117 | 0.029 | 0.008 | 0.139 | 0.233 | 0.281 |
|  |  | 1 | 0 | 1 | 0.5 | 0.3 | 0.2 | -0.134 | 0.623 | -0.757 | -0.757 | -0.371 | -0.205 | -0.181 | 0.346 | 0.460 | 0.316 | 0.316 | 0.250 | 0.207 | 0.069 | 0.138 | 0.599 | 0.673 | 0.673 | 0.200 | 0.085 | 0.038 | 0.668 | 0.243 | 0.003 | 0.054 | 0.060 | 0.026 | 0.003 | 0.184 | 0.225 | 0.303 |
|  |  | 1.3 | 0.8 | 0.5 | 0 | 0.3 | 0.2 | -0.188 | 0.045 | -0.233 | -0.233 | 0.145 | -0.199 | -0.179 | 0.300 | 0.397 | 0.280 | 0.280 | 0.216 | 0.185 | 0.065 | 0.125 | 0.160 | 0.133 | 0.133 | 0.068 | 0.074 | 0.036 | 0.949 | 0.515 | 0.004 | 0.086 | 0.076 | 0.035 | 0.007 | 0.167 | 0.260 | 0.276 |
|  |  | 1.5 | 0.8 | 0.7 | 0.5 | 0 | 0.2 | -0.225 | 0.232 | -0.457 | -0.457 | -0.313 | 0.041 | -0.185 | 0.375 | 0.504 | 0.342 | 0.342 | 0.305 | 0.156 | 0.075 | 0.191 | 0.308 | 0.326 | 0.326 | 0.191 | 0.026 | 0.040 | 0.883 | 0.520 | 0.003 | 0.054 | 0.097 | 0.010 | 0.001 | 0.188 | 0.211 | 0.304 |
|  |  | 1 | 0.8 | 0.2 | 0 | 0 | 0.2 | -0.146 | -0.116 | -0.030 | -0.030 | 0.128 | 0.034 | -0.192 | 0.262 | 0.343 | 0.226 | 0.226 | 0.206 | 0.108 | 0.052 | 0.090 | 0.131 | 0.052 | 0.052 | 0.059 | 0.013 | 0.040 | 0.857 | 0.475 | 0.003 | 0.047 | 0.084 | 0.009 | 0.000 | 0.189 | 0.192 | 0.296 |
|  |  | 0 | -1 | 1 | 0.5 | 0.3 | 0.2 | 0.048 | 0.934 | -0.886 | -0.886 | -0.450 | -0.249 | -0.187 | 0.264 | 0.364 | 0.248 | 0.248 | 0.199 | 0.157 | 0.055 | 0.072 | 1.005 | 0.846 | 0.846 | 0.242 | 0.086 | 0.038 | 0.040 | 0.046 | 0.001 | 0.034 | 0.046 | 0.024 | 0.003 | 0.156 | 0.201 | 0.288 |

TE: total effect; DE: direct effect; IE: indirect effect; IE_d: indirect effect calculated by difference method; IE_p: indirect effect calculated by product method; IE1: *X*→*M_1_*→*Y*; IE2: *X*→*M_2_*→*Y*; IE3: *X*→*M_1_*→*M_2_*→*Y*.

#### 11 Additional simulation scenario 5: PSE-MR with causal order of multiple mediators is misspecified

For both causally ordered and non-ordered mediators, when the causal order was misspecified, total effect, direct effect and indirect effect (through all the mediators) can be unbiasedly estimated. This was because, taking two mediators as an example, the total effect was estimated by the weighted regression , which didn’t use the information of mediators. Similarly, the direct effect was estimated by the weighted regression , which was not also influenced by the order of mediators. Moreover, the indirect effect can be estimated by the difference method (). In two non-ordered mediators setting, the order of mediators was independent of estimation and path-specific effects (*X*→*M*_1_→*Y* and *X*→*M*_2_→*Y*) are unbiased. However, in two ordered mediators setting, path-specific effects (*X*→*M*_1_→*Y*, *X*→*M*_2_→*Y* and *X*→*M*_1_→*M*_2_→*Y*) would be biased when the causal order was misspecified. If the correct order was *M*_2_→*M*_1_, we can obtain the estimate of parameters in the indirect pathway by the weighted regression . However, if we were misspecified this order, we would regress the on the and the . Thus the above three path-specific effects would be biased. Results were listed in **eTable 37**.

**eTable 37.** Simulation of PSE-MR when causal order of multiple mediators is misspecified

|  |  | TRUIE | | | | | | BIAS | | | | | | | SE | | | | | | | MSE | | | | | | | Power/Type I error | | | | | | | | | |
| --- | --- | --- | --- | --- | --- | --- | --- | --- | --- | --- | --- | --- | --- | --- | --- | --- | --- | --- | --- | --- | --- | --- | --- | --- | --- | --- | --- | --- | --- | --- | --- | --- | --- | --- | --- | --- | --- | --- |
|  |  | TE | DE | IE | IE1 | IE2 | IE3 | TE | DE | IE_d | IE1 | IE2 | IE3 | IE_p | TE | DE | IE_d | IE1 | IE2 | IE3 | IE_p | TE | DE | IE_d | IE1 | IE2 | IE3 | IE_p | TE | DE | IE_d | IE1 | IE2 | IE3 | IE_p | GY | GM1 | GM2 |
| Standard MR | IVW | 1.8 | 0.8 | 1 | 0.5 | 0.3 | 0.2 | 0.007 | -0.049 | 0.056 | 0.056 | 0.036 | 0.021 | 0.000 | 0.079 | 0.047 | 0.089 | 0.089 | 0.064 | 0.100 | 0.095 | 0.006 | 0.005 | 0.011 | 0.011 | 0.005 | 0.010 | 0.009 | 1.000 | 1.000 | 1.000 | 1.000 | 0.995 | 0.871 | 0.572 |  |  |  |
|  |  | 0.8 | 0.8 | 0 | -0.5 | 0.3 | 0.2 | 0.004 | -0.044 | 0.047 | 0.047 | 1.035 | -0.600 | -0.388 | 0.050 | 0.043 | 0.063 | 0.063 | 0.064 | 0.094 | 0.086 | 0.003 | 0.004 | 0.006 | 0.006 | 1.075 | 0.368 | 0.158 | 1.000 | 1.000 | 0.005 | 0.000 | 1.000 | 0.900 | 0.617 |  |  |  |
|  |  | 1.2 | 0.8 | 0.4 | 0.5 | -0.3 | 0.2 | 0.008 | -0.048 | 0.055 | 0.055 | -0.593 | 0.880 | -0.233 | 0.074 | 0.045 | 0.083 | 0.083 | 0.042 | 0.067 | 0.026 | 0.006 | 0.004 | 0.010 | 0.010 | 0.353 | 0.779 | 0.055 | 1.000 | 1.000 | 0.987 | 0.987 | 0.608 | 1.000 | 0.185 |  |  |  |
|  |  | 0.2 | 0.8 | -0.6 | -0.5 | -0.3 | 0.2 | -0.002 | -0.048 | 0.046 | 0.045 | 0.405 | -0.187 | -0.173 | 0.049 | 0.045 | 0.064 | 0.064 | 0.042 | 0.059 | 0.023 | 0.002 | 0.004 | 0.006 | 0.006 | 0.166 | 0.038 | 0.030 | 0.964 | 1.000 | 1.000 | 1.000 | 0.628 | 1.000 | 0.210 |  |  |  |
|  |  | 0.2 | -0.8 | 1 | 0.5 | 0.3 | 0.2 | 0.014 | -0.045 | 0.059 | 0.059 | 0.039 | 0.023 | -0.003 | 0.082 | 0.048 | 0.091 | 0.091 | 0.064 | 0.099 | 0.090 | 0.007 | 0.004 | 0.012 | 0.012 | 0.006 | 0.010 | 0.008 | 0.714 | 1.000 | 1.000 | 1.000 | 1.000 | 0.905 | 0.604 |  |  |  |
|  |  | -1.8 | -0.8 | -1 | -0.5 | -0.3 | -0.2 | -0.006 | -0.047 | 0.041 | 0.041 | 0.025 | 0.005 | 0.011 | 0.075 | 0.046 | 0.084 | 0.084 | 0.062 | 0.099 | 0.089 | 0.006 | 0.004 | 0.009 | 0.009 | 0.004 | 0.010 | 0.008 | 1.000 | 1.000 | 1.000 | 1.000 | 0.996 | 0.871 | 0.631 |  |  |  |
|  |  | 1 | 0 | 1 | 0.5 | 0.3 | 0.2 | 0.012 | -0.046 | 0.058 | 0.058 | 0.036 | 0.026 | -0.004 | 0.082 | 0.048 | 0.088 | 0.088 | 0.062 | 0.104 | 0.095 | 0.007 | 0.004 | 0.011 | 0.011 | 0.005 | 0.012 | 0.009 | 1.000 | 0.000 | 1.000 | 1.000 | 1.000 | 0.890 | 0.592 |  |  |  |
|  |  | 1.3 | 0.8 | 0.5 | 0 | 0.3 | 0.2 | 0.006 | -0.048 | 0.054 | 0.054 | 0.536 | -0.289 | -0.193 | 0.052 | 0.049 | 0.069 | 0.069 | 0.065 | 0.028 | 0.017 | 0.003 | 0.005 | 0.008 | 0.008 | 0.291 | 0.084 | 0.038 | 1.000 | 1.000 | 0.999 | 0.999 | 1.000 | 0.000 | 0.000 |  |  |  |
|  |  | 1.5 | 0.8 | 0.7 | 0.5 | 0 | 0.2 | 0.008 | -0.029 | 0.036 | 0.036 | -0.286 | 0.441 | -0.119 | 0.084 | 0.044 | 0.092 | 0.092 | 0.051 | 0.065 | 0.044 | 0.007 | 0.003 | 0.010 | 0.010 | 0.084 | 0.199 | 0.016 | 1.000 | 1.000 | 1.000 | 1.000 | 0.989 | 0.989 | 0.590 |  |  |  |
|  |  | 1 | 0.8 | 0.2 | 0 | 0 | 0.2 | 0.005 | -0.029 | 0.033 | 0.033 | 0.213 | 0.017 | -0.197 | 0.053 | 0.041 | 0.063 | 0.063 | 0.053 | 0.036 | 0.007 | 0.003 | 0.002 | 0.005 | 0.005 | 0.048 | 0.002 | 0.039 | 1.000 | 1.000 | 0.406 | 0.477 | 0.990 | 0.000 | 0.000 |  |  |  |
|  |  | 0 | -1 | 1 | 0.5 | 0.3 | 0.2 | 0.010 | -0.046 | 0.056 | 0.056 | 0.035 | 0.022 | -0.001 | 0.081 | 0.046 | 0.091 | 0.091 | 0.063 | 0.100 | 0.096 | 0.007 | 0.004 | 0.011 | 0.011 | 0.005 | 0.010 | 0.009 | 0.058 | 1.000 | 1.000 | 1.000 | 1.000 | 0.894 | 0.607 |  |  |  |
| Sc2 | Egger | 1.8 | 0.8 | 1 | 0.5 | 0.3 | 0.2 | 0.001 | -0.026 | 0.028 | 0.028 | 0.016 | 0.030 | -0.018 | 0.050 | 0.127 | 0.132 | 0.132 | 0.108 | 0.115 | 0.100 | 0.003 | 0.017 | 0.018 | 0.018 | 0.012 | 0.014 | 0.010 | 1.000 | 0.981 | 1.000 | 0.999 | 0.917 | 0.790 | 0.395 | 0.000 | 0.042 | 0.055 |
|  |  | 0.8 | 0.8 | 0 | -0.5 | 0.3 | 0.2 | 0.001 | -0.020 | 0.021 | 0.021 | 1.022 | -0.620 | -0.380 | 0.039 | 0.125 | 0.128 | 0.128 | 0.116 | 0.115 | 0.095 | 0.001 | 0.016 | 0.017 | 0.017 | 1.057 | 0.398 | 0.153 | 1.000 | 1.000 | 0.042 | 0.006 | 0.991 | 0.932 | 0.512 | 0.042 | 0.051 | 0.046 |
|  |  | 1.2 | 0.8 | 0.4 | 0.5 | -0.3 | 0.2 | 0.004 | -0.019 | 0.023 | 0.022 | -0.597 | 0.851 | -0.233 | 0.047 | 0.134 | 0.136 | 0.136 | 0.033 | 0.133 | 0.021 | 0.002 | 0.018 | 0.019 | 0.019 | 0.357 | 0.743 | 0.055 | 1.000 | 1.000 | 0.828 | 0.841 | 0.954 | 0.982 | 0.392 | 0.021 | 0.042 | 0.042 |
|  |  | 0.2 | 0.8 | -0.6 | -0.5 | -0.3 | 0.2 | 0.002 | -0.021 | 0.024 | 0.023 | 0.403 | -0.211 | -0.169 | 0.037 | 0.132 | 0.131 | 0.131 | 0.032 | 0.131 | 0.021 | 0.001 | 0.018 | 0.018 | 0.018 | 0.163 | 0.062 | 0.029 | 0.998 | 1.000 | 0.990 | 0.990 | 0.959 | 0.979 | 0.375 | 0.038 | 0.045 | 0.047 |
|  |  | 0.2 | -0.8 | 1 | 0.5 | 0.3 | 0.2 | 0.002 | -0.026 | 0.027 | 0.028 | 0.015 | 0.028 | -0.015 | 0.050 | 0.127 | 0.132 | 0.132 | 0.113 | 0.119 | 0.100 | 0.002 | 0.017 | 0.018 | 0.018 | 0.013 | 0.015 | 0.010 | 0.963 | 0.999 | 1.000 | 1.000 | 0.995 | 0.940 | 0.539 | 0.047 | 0.054 | 0.041 |
|  |  | -1.8 | -0.8 | -1 | -0.5 | -0.3 | -0.2 | -0.001 | -0.025 | 0.024 | 0.024 | 0.013 | -0.016 | 0.026 | 0.047 | 0.130 | 0.134 | 0.134 | 0.110 | 0.115 | 0.092 | 0.002 | 0.018 | 0.019 | 0.019 | 0.012 | 0.014 | 0.009 | 1.000 | 0.993 | 1.000 | 0.999 | 0.922 | 0.810 | 0.439 | 0.002 | 0.039 | 0.053 |
|  |  | 1 | 0 | 1 | 0.5 | 0.3 | 0.2 | 0.001 | -0.024 | 0.025 | 0.025 | 0.020 | 0.029 | -0.024 | 0.052 | 0.131 | 0.136 | 0.136 | 0.112 | 0.118 | 0.094 | 0.003 | 0.018 | 0.019 | 0.019 | 0.013 | 0.015 | 0.009 | 1.000 | 0.019 | 1.000 | 1.000 | 0.995 | 0.923 | 0.489 | 0.012 | 0.054 | 0.055 |
|  |  | 1.3 | 0.8 | 0.5 | 0 | 0.3 | 0.2 | 0.001 | -0.027 | 0.029 | 0.029 | 0.520 | -0.294 | -0.197 | 0.038 | 0.128 | 0.128 | 0.128 | 0.108 | 0.078 | 0.048 | 0.001 | 0.017 | 0.017 | 0.017 | 0.282 | 0.093 | 0.041 | 1.000 | 0.999 | 0.953 | 0.907 | 0.990 | 0.018 | 0.011 | 0.015 | 0.050 | 0.048 |
|  |  | 1.5 | 0.8 | 0.7 | 0.5 | 0 | 0.2 | 0.001 | -0.014 | 0.015 | 0.015 | -0.294 | 0.438 | -0.129 | 0.049 | 0.115 | 0.119 | 0.119 | 0.052 | 0.112 | 0.038 | 0.002 | 0.013 | 0.014 | 0.015 | 0.089 | 0.204 | 0.018 | 1.000 | 1.000 | 1.000 | 1.000 | 0.963 | 0.903 | 0.446 | 0.002 | 0.041 | 0.053 |
|  |  | 1 | 0.8 | 0.2 | 0 | 0 | 0.2 | 0.002 | -0.019 | 0.020 | 0.020 | 0.206 | 0.012 | -0.198 | 0.037 | 0.119 | 0.119 | 0.119 | 0.051 | 0.106 | 0.019 | 0.001 | 0.014 | 0.015 | 0.015 | 0.045 | 0.011 | 0.040 | 1.000 | 1.000 | 0.419 | 0.425 | 0.990 | 0.051 | 0.020 | 0.039 | 0.049 | 0.048 |
|  |  | 0 | -1 | 1 | 0.5 | 0.3 | 0.2 | 0.000 | -0.027 | 0.028 | 0.028 | 0.018 | 0.031 | -0.021 | 0.051 | 0.127 | 0.135 | 0.135 | 0.113 | 0.112 | 0.095 | 0.003 | 0.017 | 0.019 | 0.019 | 0.013 | 0.014 | 0.009 | 0.052 | 1.000 | 1.000 | 1.000 | 0.993 | 0.946 | 0.526 | 0.043 | 0.047 | 0.052 |
| Sc3 | Egger | 1.8 | 0.8 | 1 | 0.5 | 0.3 | 0.2 | 0.003 | -0.079 | 0.081 | 0.081 | 0.051 | 0.010 | 0.021 | 0.029 | 0.124 | 0.125 | 0.125 | 0.111 | 0.115 | 0.106 | 0.001 | 0.022 | 0.022 | 0.022 | 0.015 | 0.013 | 0.012 | 1.000 | 0.323 | 0.951 | 0.828 | 0.183 | 0.043 | 0.029 | 0.000 | 0.141 | 0.942 |
|  |  | 0.8 | 0.8 | 0 | -0.5 | 0.3 | 0.2 | 0.001 | -0.078 | 0.079 | 0.079 | 1.044 | -0.581 | -0.384 | 0.020 | 0.119 | 0.118 | 0.118 | 0.109 | 0.108 | 0.088 | 0.000 | 0.020 | 0.020 | 0.020 | 1.103 | 0.349 | 0.155 | 1.000 | 0.995 | 0.012 | 0.001 | 0.965 | 0.758 | 0.452 | 0.099 | 0.127 | 0.947 |
|  |  | 1.2 | 0.8 | 0.4 | 0.5 | -0.3 | 0.2 | 0.000 | -0.081 | 0.081 | 0.081 | -0.591 | 0.902 | -0.230 | 0.026 | 0.125 | 0.128 | 0.128 | 0.026 | 0.126 | 0.024 | 0.001 | 0.022 | 0.023 | 0.023 | 0.350 | 0.830 | 0.054 | 1.000 | 0.921 | 0.532 | 0.531 | 0.581 | 0.831 | 0.174 | 0.007 | 0.872 | 0.374 |
|  |  | 0.2 | 0.8 | -0.6 | -0.5 | -0.3 | 0.2 | 0.000 | -0.081 | 0.081 | 0.081 | 0.409 | -0.152 | -0.177 | 0.020 | 0.125 | 0.124 | 0.124 | 0.026 | 0.122 | 0.018 | 0.000 | 0.022 | 0.022 | 0.022 | 0.168 | 0.038 | 0.032 | 1.000 | 0.998 | 0.984 | 0.984 | 0.970 | 0.955 | 0.220 | 0.409 | 0.867 | 0.367 |
|  |  | 0.2 | -0.8 | 1 | 0.5 | 0.3 | 0.2 | 0.001 | -0.084 | 0.085 | 0.085 | 0.056 | 0.013 | 0.016 | 0.030 | 0.127 | 0.128 | 0.128 | 0.107 | 0.114 | 0.100 | 0.001 | 0.023 | 0.023 | 0.024 | 0.015 | 0.013 | 0.010 | 1.000 | 0.998 | 1.000 | 0.999 | 0.979 | 0.827 | 0.545 | 0.128 | 0.121 | 0.934 |
|  |  | -1.8 | -0.8 | -1 | -0.5 | -0.3 | -0.2 | 0.000 | -0.077 | 0.077 | 0.077 | 0.051 | 0.019 | 0.007 | 0.026 | 0.121 | 0.122 | 0.122 | 0.107 | 0.107 | 0.091 | 0.001 | 0.021 | 0.021 | 0.021 | 0.014 | 0.012 | 0.008 | 1.000 | 0.855 | 0.938 | 0.790 | 0.133 | 0.085 | 0.037 | 0.000 | 0.130 | 0.948 |
|  |  | 1 | 0 | 1 | 0.5 | 0.3 | 0.2 | 0.002 | -0.079 | 0.082 | 0.082 | 0.051 | 0.010 | 0.021 | 0.029 | 0.122 | 0.122 | 0.122 | 0.111 | 0.109 | 0.103 | 0.001 | 0.021 | 0.022 | 0.022 | 0.015 | 0.012 | 0.011 | 1.000 | 0.000 | 0.999 | 0.994 | 0.745 | 0.456 | 0.272 | 0.002 | 0.123 | 0.943 |
|  |  | 1.3 | 0.8 | 0.5 | 0 | 0.3 | 0.2 | 0.002 | -0.083 | 0.086 | 0.086 | 0.554 | -0.283 | -0.186 | 0.022 | 0.118 | 0.119 | 0.119 | 0.109 | 0.072 | 0.055 | 0.000 | 0.021 | 0.022 | 0.022 | 0.319 | 0.085 | 0.038 | 1.000 | 0.845 | 0.627 | 0.326 | 0.708 | 0.000 | 0.000 | 0.002 | 0.123 | 0.935 |
|  |  | 1.5 | 0.8 | 0.7 | 0.5 | 0 | 0.2 | 0.003 | -0.052 | 0.055 | 0.055 | -0.279 | 0.447 | -0.113 | 0.029 | 0.109 | 0.112 | 0.112 | 0.046 | 0.105 | 0.042 | 0.001 | 0.015 | 0.015 | 0.015 | 0.080 | 0.211 | 0.015 | 1.000 | 0.785 | 0.849 | 0.826 | 0.368 | 0.197 | 0.090 | 0.001 | 0.135 | 0.934 |
|  |  | 1 | 0.8 | 0.2 | 0 | 0 | 0.2 | 0.002 | -0.052 | 0.054 | 0.054 | 0.221 | 0.027 | -0.194 | 0.021 | 0.112 | 0.113 | 0.113 | 0.047 | 0.103 | 0.022 | 0.000 | 0.015 | 0.016 | 0.016 | 0.051 | 0.011 | 0.038 | 1.000 | 0.987 | 0.100 | 0.080 | 0.860 | 0.000 | 0.000 | 0.013 | 0.126 | 0.935 |
|  |  | 0 | -1 | 1 | 0.5 | 0.3 | 0.2 | 0.004 | -0.078 | 0.082 | 0.082 | 0.048 | 0.016 | 0.019 | 0.029 | 0.122 | 0.122 | 0.122 | 0.115 | 0.109 | 0.099 | 0.001 | 0.021 | 0.022 | 0.022 | 0.016 | 0.012 | 0.010 | 0.042 | 1.000 | 1.000 | 1.000 | 0.982 | 0.860 | 0.596 | 0.177 | 0.118 | 0.933 |
| Sc4 | Egger | 1.8 | 0.8 | 1 | 0.5 | 0.3 | 0.2 | 0.072 | -0.366 | 0.438 | 0.438 | 0.269 | -0.091 | 0.261 | 0.104 | 0.060 | 0.117 | 0.117 | 0.122 | 0.062 | 0.108 | 0.016 | 0.138 | 0.206 | 0.206 | 0.087 | 0.012 | 0.080 | 1.000 | 0.622 | 1.000 | 0.993 | 0.804 | 0.124 | 0.134 | 0.000 | 0.029 | 0.934 |
|  |  | 0.8 | 0.8 | 0 | -0.5 | 0.3 | 0.2 | 0.031 | -0.367 | 0.398 | 0.398 | 1.275 | -0.416 | -0.462 | 0.044 | 0.060 | 0.071 | 0.071 | 0.119 | 0.051 | 0.097 | 0.003 | 0.138 | 0.163 | 0.163 | 1.641 | 0.176 | 0.222 | 1.000 | 0.999 | 0.997 | 0.264 | 1.000 | 0.471 | 0.484 | 0.004 | 0.022 | 0.936 |
|  |  | 1.2 | 0.8 | 0.4 | 0.5 | -0.3 | 0.2 | 0.029 | -0.365 | 0.393 | 0.393 | -0.560 | 0.983 | -0.030 | 0.047 | 0.061 | 0.075 | 0.075 | 0.025 | 0.079 | 0.055 | 0.003 | 0.137 | 0.160 | 0.160 | 0.314 | 0.972 | 0.004 | 1.000 | 0.988 | 1.000 | 1.000 | 0.131 | 1.000 | 0.974 | 0.000 | 0.359 | 0.840 |
|  |  | 0.2 | 0.8 | -0.6 | -0.5 | -0.3 | 0.2 | -0.012 | -0.369 | 0.356 | 0.356 | 0.438 | 0.154 | -0.236 | 0.026 | 0.063 | 0.064 | 0.064 | 0.026 | 0.059 | 0.018 | 0.001 | 0.140 | 0.131 | 0.131 | 0.193 | 0.027 | 0.056 | 1.000 | 0.999 | 0.950 | 0.954 | 0.793 | 0.727 | 0.709 | 0.070 | 0.322 | 0.846 |
|  |  | 0.2 | -0.8 | 1 | 0.5 | 0.3 | 0.2 | 0.071 | -0.367 | 0.438 | 0.438 | 0.270 | -0.093 | 0.261 | 0.102 | 0.059 | 0.116 | 0.116 | 0.115 | 0.058 | 0.109 | 0.016 | 0.138 | 0.205 | 0.206 | 0.086 | 0.012 | 0.080 | 0.712 | 1.000 | 1.000 | 1.000 | 1.000 | 0.950 | 0.965 | 0.001 | 0.022 | 0.932 |
|  |  | -1.8 | -0.8 | -1 | -0.5 | -0.3 | -0.2 | -0.029 | -0.365 | 0.335 | 0.335 | 0.217 | 0.181 | -0.063 | 0.046 | 0.057 | 0.071 | 0.071 | 0.097 | 0.048 | 0.094 | 0.003 | 0.136 | 0.118 | 0.118 | 0.057 | 0.035 | 0.013 | 1.000 | 1.000 | 1.000 | 0.156 | 0.004 | 0.002 | 0.004 | 0.000 | 0.022 | 0.938 |
|  |  | 1 | 0 | 1 | 0.5 | 0.3 | 0.2 | 0.072 | -0.364 | 0.436 | 0.436 | 0.276 | -0.096 | 0.257 | 0.102 | 0.060 | 0.114 | 0.114 | 0.118 | 0.057 | 0.108 | 0.016 | 0.136 | 0.203 | 0.203 | 0.090 | 0.012 | 0.078 | 1.000 | 0.849 | 1.000 | 1.000 | 0.988 | 0.628 | 0.646 | 0.000 | 0.025 | 0.932 |
|  |  | 1.3 | 0.8 | 0.5 | 0 | 0.3 | 0.2 | 0.049 | -0.366 | 0.416 | 0.416 | 0.772 | -0.256 | -0.101 | 0.072 | 0.058 | 0.089 | 0.089 | 0.117 | 0.041 | 0.090 | 0.008 | 0.137 | 0.181 | 0.181 | 0.610 | 0.067 | 0.018 | 1.000 | 0.955 | 1.000 | 0.952 | 0.984 | 0.000 | 0.000 | 0.000 | 0.017 | 0.935 |
|  |  | 1.5 | 0.8 | 0.7 | 0.5 | 0 | 0.2 | 0.066 | -0.233 | 0.299 | 0.299 | -0.170 | 0.471 | -0.003 | 0.101 | 0.098 | 0.141 | 0.141 | 0.074 | 0.098 | 0.058 | 0.015 | 0.064 | 0.109 | 0.109 | 0.034 | 0.232 | 0.003 | 1.000 | 0.433 | 0.990 | 0.968 | 0.912 | 0.285 | 0.291 | 0.000 | 0.021 | 0.947 |
|  |  | 1 | 0.8 | 0.2 | 0 | 0 | 0.2 | 0.056 | -0.225 | 0.281 | 0.280 | 0.340 | 0.098 | -0.158 | 0.072 | 0.099 | 0.119 | 0.119 | 0.077 | 0.095 | 0.043 | 0.008 | 0.060 | 0.093 | 0.093 | 0.122 | 0.019 | 0.027 | 1.000 | 0.918 | 0.710 | 0.570 | 0.993 | 0.004 | 0.004 | 0.000 | 0.028 | 0.933 |
|  |  | 0 | -1 | 1 | 0.5 | 0.3 | 0.2 | 0.070 | -0.367 | 0.436 | 0.437 | 0.269 | -0.095 | 0.262 | 0.099 | 0.058 | 0.112 | 0.112 | 0.117 | 0.058 | 0.109 | 0.015 | 0.138 | 0.203 | 0.203 | 0.086 | 0.012 | 0.081 | 0.097 | 1.000 | 1.000 | 1.000 | 1.000 | 0.967 | 0.984 | 0.013 | 0.019 | 0.948 |

TE: total effect; DE: direct effect; IE: indirect effect; IE_d: indirect effect calculated by difference method; IE_p: indirect effect calculated by product method; IE1: *X*→*M_1_*→*Y*; IE2: *X*→*M_2_*→*Y*; IE3: *X*→*M_1_*→*M_2_*→*Y*.

#### 12 Additional simulation scenario 6: PSE-MR when missing one of the multiple mediators

We conducted a simulation that the data-generating model was the same as the **Simulation C-F**. Taking two mediators as an example. In two non-ordered mediators setting (Figure 1 (e)), if we were missing *M*_1_ (or *M*_2_), the new indirect pathway of *X* on *Y* was *X*→*M*_2_→*Y* (or *X*→*M*_1_→*Y*) with effect (or ) and the new direct effect was (or ). Thus the PSE (*X*→*M*_2_→*Y*) can be unbiasedly estimated. Original direct effect and PSE (*X*→*M*_1_→*Y*) were merged into new direct effect, which can also be correctly estimated.

In two ordered mediators setting (Figure 1 (f)), we also supposed that *M*_1_ (or *M*_2_) was missing, the new indirect pathway of *X* on *Y* through *M*_2_ (or *M*_1_) consisted of original PSEs: *X*→*M*_2_→*Y* and *X*→*M*_1_→*M*_2_→*Y* (or *X*→*M*_1_→*Y* and *X*→*M*_1_→*M*_2_→*Y*), with effect (or ) and the new direct effect became (or ). When *M*_1_ was missing, the estimation of new direct and indirect effects were biased (eTable 38) regardless of the existence of pleiotropy, and the biases became larger as the increase of (eTable 39). In this case, *M*_1_ was the confounder of *M*_2_ and Y and it was affected by X (i.e. X–induced unmeasured confounder of *M*_2_ and *Y*), thus the assumption of cross-world independence assumption was violated. In addition, as you said, it might violate the InSIDE assumption. If we were missing *M*_1_, the InSIDE assumption would be , which wasn’t satisfied because of the common factor . Nevertheless when the causal effect of *M*_1_ on *Y* () or *M*_1_ on *M*_2_ () was zero, PSE-MR can still obtain unbiased estimates of new direct and indirect effects. If we were missing *M*_2_ (eTable 40), the estimation of new direct and indirect effects were unbiased regardless of the existence of pleiotropy. The InSIDE assumption can be satisfied. Therefore, we only allowed the mediator to be missed if it had no direct effects on either other mediators or Y. Results were listed in **eTable 38-40**.

**eTable 38.** Simulation of PSE-MR when missing M1

|  |  | TRUE | | | | | | | | | BIAS | | | | SE | | | | MSE | | | | Power/Type I error | | | | | |
| --- | --- | --- | --- | --- | --- | --- | --- | --- | --- | --- | --- | --- | --- | --- | --- | --- | --- | --- | --- | --- | --- | --- | --- | --- | --- | --- | --- | --- |
|  |  | TE | DE | IE | $\theta_{0}$ | $\alpha_{1}$ | $\delta_{1}$ | $\alpha_{2}$ | $\delta_{2}$ | $r_{12}$ | TE | DE | IE_d | IE_p | TE | DE | IE_d | IE_p | TE | DE | IE_d | IE_p | TE | DE | IE_d | IE_p | GY | GM |
| PSE-MR | Standard MR | 0.8 | 0.5 | 0.3 | 0 | 0.3 | 1 | 0.5 | 1 | 0.4 | 0.007 | -0.227 | 0.234 | 0.234 | 0.079 | 0.101 | 0.115 | 0.115 | 0.006 | 0.062 | 0.068 | 0.068 | 1.000 | 0.510 | 1.000 | 1.000 |  |  |
|  |  | 1.3 | 1.3 | 0 | 0.8 | 0 | 1 | 0.5 | 1 | 0.4 | 0.012 | -0.085 | 0.096 | 0.096 | 0.083 | 0.058 | 0.079 | 0.079 | 0.007 | 0.010 | 0.016 | 0.016 | 1.000 | 1.000 | 0.851 | 0.988 |  |  |
|  |  | 1.3 | 1.3 | 0 | 0.8 | 0.3 | 0 | 0.5 | 1 | 0.4 | 0.004 | -0.224 | 0.228 | 0.228 | 0.048 | 0.107 | 0.100 | 0.100 | 0.002 | 0.062 | 0.062 | 0.062 | 1.000 | 1.000 | 0.492 | 0.641 |  |  |
|  |  | 1.3 | 1.3 | 0 | 0.8 | 0 | 0 | 0.5 | 1 | 0.4 | 0.007 | -0.085 | 0.091 | 0.091 | 0.049 | 0.059 | 0.043 | 0.043 | 0.002 | 0.011 | 0.010 | 0.010 | 1.000 | 1.000 | 0.055 | 0.623 |  |  |
|  |  | -0.7 | -1 | 0.3 | -1 | 0.3 | 1 | 0 | 1 | 0.4 | 0.007 | -0.132 | 0.139 | 0.139 | 0.079 | 0.072 | 0.086 | 0.086 | 0.006 | 0.023 | 0.027 | 0.027 | 1.000 | 1.000 | 0.999 | 1.000 |  |  |
|  |  | 1.1 | 0.8 | 0.3 | 0.8 | 0.3 | 1 | 0.5 | 0 | 0.4 | 0.007 | -0.035 | 0.042 | 0.042 | 0.052 | 0.038 | 0.060 | 0.060 | 0.003 | 0.003 | 0.005 | 0.005 | 1.000 | 1.000 | 1.000 | 1.000 |  |  |
|  |  | 1.1 | 0.8 | 0.3 | 0.8 | 0.3 | 1 | 0 | 0 | 0.4 | 0.007 | -0.021 | 0.028 | 0.028 | 0.054 | 0.026 | 0.056 | 0.056 | 0.003 | 0.001 | 0.004 | 0.004 | 1.000 | 1.000 | 0.998 | 1.000 |  |  |
| PSE-Egger | sc2 | 0.8 | 0.5 | 0.3 | 0 | 0.3 | 1 | 0.5 | 1 | 0.4 | 0.001 | -0.194 | 0.195 | 0.195 | 0.050 | 0.135 | 0.138 | 0.138 | 0.002 | 0.056 | 0.057 | 0.057 | 1.000 | 0.568 | 0.996 | 0.996 | 0.037 | 0.055 |
|  |  | 1.3 | 1.3 | 0 | 0.8 | 0 | 1 | 0.5 | 1 | 0.4 | 0.001 | -0.076 | 0.077 | 0.077 | 0.049 | 0.061 | 0.064 | 0.064 | 0.002 | 0.009 | 0.010 | 0.010 | 1.000 | 1.000 | 0.968 | 0.994 | 0.020 | 0.051 |
|  |  | 1.3 | 1.3 | 0 | 0.8 | 0.3 | 0 | 0.5 | 1 | 0.4 | 0.003 | -0.200 | 0.202 | 0.202 | 0.036 | 0.133 | 0.128 | 0.128 | 0.001 | 0.058 | 0.057 | 0.057 | 1.000 | 1.000 | 0.301 | 0.345 | 0.052 | 0.050 |
|  |  | 1.3 | 1.3 | 0 | 0.8 | 0 | 0 | 0.5 | 1 | 0.4 | -0.001 | -0.083 | 0.082 | 0.082 | 0.034 | 0.061 | 0.053 | 0.053 | 0.001 | 0.011 | 0.010 | 0.010 | 1.000 | 1.000 | 0.126 | 0.332 | 0.036 | 0.061 |
|  |  | -0.7 | -1 | 0.3 | -1 | 0.3 | 1 | 0 | 1 | 0.4 | 0.001 | -0.115 | 0.116 | 0.116 | 0.048 | 0.086 | 0.086 | 0.086 | 0.002 | 0.021 | 0.021 | 0.021 | 1.000 | 1.000 | 0.990 | 0.996 | 0.043 | 0.045 |
|  |  | 1.1 | 0.8 | 0.3 | 0.8 | 0.3 | 1 | 0.5 | 0 | 0.4 | 0.002 | -0.013 | 0.015 | 0.015 | 0.039 | 0.101 | 0.102 | 0.102 | 0.002 | 0.010 | 0.011 | 0.011 | 1.000 | 1.000 | 0.990 | 0.993 | 0.018 | 0.050 |
|  |  | 1.1 | 0.8 | 0.3 | 0.8 | 0.3 | 1 | 0 | 0 | 0.4 | 0.001 | -0.009 | 0.010 | 0.010 | 0.038 | 0.064 | 0.066 | 0.066 | 0.001 | 0.004 | 0.004 | 0.004 | 1.000 | 1.000 | 0.994 | 0.998 | 0.027 | 0.045 |
|  | sc3 | 0.8 | 0.5 | 0.3 | 0 | 0.3 | 1 | 0.5 | 1 | 0.4 | 0.002 | -0.264 | 0.266 | 0.266 | 0.031 | 0.132 | 0.131 | 0.131 | 0.001 | 0.087 | 0.088 | 0.088 | 1.000 | 0.060 | 0.979 | 0.980 | 0.121 | 0.944 |
|  |  | 1.3 | 1.3 | 0 | 0.8 | 0 | 1 | 0.5 | 1 | 0.4 | 0.003 | -0.101 | 0.104 | 0.104 | 0.031 | 0.056 | 0.058 | 0.058 | 0.001 | 0.013 | 0.014 | 0.014 | 1.000 | 1.000 | 0.855 | 0.914 | 0.030 | 0.957 |
|  |  | 1.3 | 1.3 | 0 | 0.8 | 0.3 | 0 | 0.5 | 1 | 0.4 | 0.000 | -0.264 | 0.265 | 0.265 | 0.020 | 0.130 | 0.129 | 0.129 | 0.000 | 0.087 | 0.087 | 0.087 | 1.000 | 0.999 | 0.148 | 0.167 | 0.134 | 0.928 |
|  |  | 1.3 | 1.3 | 0 | 0.8 | 0 | 0 | 0.5 | 1 | 0.4 | 0.002 | -0.105 | 0.107 | 0.107 | 0.021 | 0.052 | 0.049 | 0.049 | 0.000 | 0.014 | 0.014 | 0.014 | 1.000 | 1.000 | 0.074 | 0.149 | 0.128 | 0.939 |
|  |  | -0.7 | -1 | 0.3 | -1 | 0.3 | 1 | 0 | 1 | 0.4 | 0.003 | -0.158 | 0.161 | 0.161 | 0.030 | 0.078 | 0.080 | 0.080 | 0.001 | 0.031 | 0.032 | 0.032 | 1.000 | 1.000 | 0.999 | 0.999 | 0.482 | 0.950 |
|  |  | 1.1 | 0.8 | 0.3 | 0.8 | 0.3 | 1 | 0.5 | 0 | 0.4 | 0.003 | -0.060 | 0.063 | 0.063 | 0.022 | 0.094 | 0.094 | 0.094 | 0.000 | 0.012 | 0.013 | 0.013 | 1.000 | 0.986 | 0.867 | 0.879 | 0.000 | 0.927 |
|  |  | 1.1 | 0.8 | 0.3 | 0.8 | 0.3 | 1 | 0 | 0 | 0.4 | 0.000 | -0.038 | 0.039 | 0.039 | 0.022 | 0.058 | 0.060 | 0.060 | 0.000 | 0.005 | 0.005 | 0.005 | 1.000 | 1.000 | 0.920 | 0.936 | 0.016 | 0.946 |
|  | sc4 | 0.8 | 0.5 | 0.3 | 0 | 0.3 | 1 | 0.5 | 1 | 0.4 | 0.074 | -0.659 | 0.733 | 0.733 | 0.103 | 0.059 | 0.114 | 0.114 | 0.016 | 0.438 | 0.551 | 0.551 | 1.000 | 0.250 | 1.000 | 1.000 | 0.004 | 0.939 |
|  |  | 1.3 | 1.3 | 0 | 0.8 | 0 | 1 | 0.5 | 1 | 0.4 | 0.074 | -0.260 | 0.334 | 0.334 | 0.096 | 0.033 | 0.097 | 0.097 | 0.015 | 0.069 | 0.121 | 0.121 | 1.000 | 1.000 | 0.998 | 0.999 | 0.000 | 0.949 |
|  |  | 1.3 | 1.3 | 0 | 0.8 | 0.3 | 0 | 0.5 | 1 | 0.4 | 0.044 | -0.663 | 0.707 | 0.707 | 0.060 | 0.064 | 0.080 | 0.080 | 0.006 | 0.444 | 0.506 | 0.506 | 1.000 | 1.000 | 1.000 | 1.000 | 0.004 | 0.938 |
|  |  | 1.3 | 1.3 | 0 | 0.8 | 0 | 0 | 0.5 | 1 | 0.4 | 0.043 | -0.262 | 0.305 | 0.305 | 0.059 | 0.033 | 0.060 | 0.060 | 0.005 | 0.070 | 0.096 | 0.096 | 1.000 | 1.000 | 0.994 | 0.999 | 0.003 | 0.944 |
|  |  | -0.7 | -1 | 0.3 | -1 | 0.3 | 1 | 0 | 1 | 0.4 | 0.065 | -0.395 | 0.460 | 0.463 | 0.104 | 0.042 | 0.108 | 0.108 | 0.015 | 0.158 | 0.223 | 0.226 | 1.000 | 1.000 | 1.000 | 1.000 | 0.086 | 0.941 |
|  |  | 1.1 | 0.8 | 0.3 | 0.8 | 0.3 | 1 | 0.5 | 0 | 0.4 | 0.050 | -0.320 | 0.370 | 0.370 | 0.073 | 0.042 | 0.081 | 0.081 | 0.008 | 0.104 | 0.144 | 0.144 | 1.000 | 1.000 | 1.000 | 1.000 | 0.000 | 0.942 |
|  |  | 1.1 | 0.8 | 0.3 | 0.8 | 0.3 | 1 | 0 | 0 | 0.4 | 0.046 | -0.193 | 0.238 | 0.238 | 0.074 | 0.030 | 0.076 | 0.076 | 0.008 | 0.038 | 0.063 | 0.063 | 1.000 | 1.000 | 1.000 | 1.000 | 0.000 | 0.947 |
|  | GM2 | 0.8 | 0.5 | 0.3 | 0 | 0.3 | 1 | 0.5 | 1 | 0.4 | -0.001 | -0.495 | 0.494 | 0.494 | 0.046 | 0.188 | 0.188 | 0.188 | 0.002 | 0.280 | 0.280 | 0.280 | 1.000 | 0.033 | 0.993 | 0.993 | 0.192 | 0.641 |
|  |  | 1.3 | 1.3 | 0 | 0.8 | 0 | 1 | 0.5 | 1 | 0.4 | 0.002 | -0.199 | 0.201 | 0.201 | 0.046 | 0.085 | 0.086 | 0.086 | 0.002 | 0.047 | 0.048 | 0.048 | 1.000 | 1.000 | 0.984 | 0.989 | 0.108 | 0.625 |
|  |  | 1.3 | 1.3 | 0 | 0.8 | 0.3 | 0 | 0.5 | 1 | 0.4 | 0.002 | -0.490 | 0.492 | 0.492 | 0.036 | 0.187 | 0.185 | 0.185 | 0.001 | 0.275 | 0.276 | 0.276 | 1.000 | 0.973 | 0.696 | 0.717 | 0.187 | 0.621 |
|  |  | 1.3 | 1.3 | 0 | 0.8 | 0 | 0 | 0.5 | 1 | 0.4 | 0.000 | -0.200 | 0.200 | 0.200 | 0.035 | 0.079 | 0.074 | 0.074 | 0.001 | 0.046 | 0.045 | 0.045 | 1.000 | 1.000 | 0.612 | 0.733 | 0.182 | 0.609 |
|  |  | -0.7 | -1 | 0.3 | -1 | 0.3 | 1 | 0 | 1 | 0.4 | 0.000 | -0.292 | 0.292 | 0.293 | 0.046 | 0.114 | 0.114 | 0.114 | 0.002 | 0.098 | 0.098 | 0.099 | 1.000 | 1.000 | 0.994 | 0.995 | 0.155 | 0.595 |
|  |  | 1.1 | 0.8 | 0.3 | 0.8 | 0.3 | 1 | 0.5 | 0 | 0.4 | 0.001 | -0.044 | 0.045 | 0.045 | 0.032 | 0.161 | 0.160 | 0.160 | 0.001 | 0.028 | 0.028 | 0.028 | 1.000 | 0.977 | 0.834 | 0.847 | 0.014 | 0.620 |
|  |  | 1.1 | 0.8 | 0.3 | 0.8 | 0.3 | 1 | 0 | 0 | 0.4 | 0.000 | -0.026 | 0.026 | 0.026 | 0.031 | 0.095 | 0.093 | 0.093 | 0.001 | 0.010 | 0.009 | 0.009 | 1.000 | 1.000 | 0.876 | 0.905 | 0.020 | 0.624 |
|  | GM1 | 0.8 | 0.5 | 0.3 | 0 | 0.3 | 1 | 0.5 | 1 | 0.4 | 0.002 | -0.092 | 0.094 | 0.094 | 0.042 | 0.120 | 0.122 | 0.122 | 0.002 | 0.023 | 0.024 | 0.024 | 1.000 | 0.885 | 0.986 | 0.988 | 0.175 | 0.095 |
|  |  | 1.3 | 1.3 | 0 | 0.8 | 0 | 1 | 0.5 | 1 | 0.4 | 0.004 | -0.034 | 0.038 | 0.038 | 0.041 | 0.056 | 0.055 | 0.055 | 0.002 | 0.004 | 0.004 | 0.004 | 1.000 | 1.000 | 0.931 | 0.987 | 0.080 | 0.082 |
|  |  | 1.3 | 1.3 | 0 | 0.8 | 0.3 | 0 | 0.5 | 1 | 0.4 | 0.001 | -0.085 | 0.086 | 0.086 | 0.029 | 0.121 | 0.118 | 0.118 | 0.001 | 0.022 | 0.021 | 0.021 | 1.000 | 1.000 | 0.065 | 0.080 | 0.184 | 0.103 |
|  |  | 1.3 | 1.3 | 0 | 0.8 | 0 | 0 | 0.5 | 1 | 0.4 | 0.000 | -0.036 | 0.036 | 0.036 | 0.028 | 0.055 | 0.047 | 0.047 | 0.001 | 0.004 | 0.003 | 0.003 | 1.000 | 1.000 | 0.025 | 0.100 | 0.176 | 0.085 |
|  |  | -0.7 | -1 | 0.3 | -1 | 0.3 | 1 | 0 | 1 | 0.4 | 0.003 | -0.049 | 0.052 | 0.053 | 0.043 | 0.075 | 0.076 | 0.076 | 0.002 | 0.008 | 0.008 | 0.009 | 1.000 | 1.000 | 0.988 | 0.994 | 0.189 | 0.100 |
|  |  | 1.1 | 0.8 | 0.3 | 0.8 | 0.3 | 1 | 0.5 | 0 | 0.4 | 0.000 | -0.018 | 0.018 | 0.018 | 0.036 | 0.109 | 0.110 | 0.110 | 0.001 | 0.012 | 0.012 | 0.012 | 1.000 | 1.000 | 0.987 | 0.990 | 0.008 | 0.097 |
|  |  | 1.1 | 0.8 | 0.3 | 0.8 | 0.3 | 1 | 0 | 0 | 0.4 | 0.003 | -0.014 | 0.017 | 0.017 | 0.036 | 0.066 | 0.066 | 0.066 | 0.001 | 0.004 | 0.005 | 0.005 | 1.000 | 1.000 | 0.989 | 0.994 | 0.024 | 0.082 |
|  | GY | 0.8 | 0.5 | 0.3 | 0 | 0.3 | 1 | 0.5 | 1 | 0.4 | 0.003 | -0.196 | 0.199 | 0.199 | 0.047 | 0.105 | 0.109 | 0.109 | 0.002 | 0.049 | 0.051 | 0.051 | 1.000 | 0.710 | 0.998 | 0.998 | 0.221 | 0.052 |
|  |  | 1.3 | 1.3 | 0 | 0.8 | 0 | 1 | 0.5 | 1 | 0.4 | 0.001 | -0.079 | 0.080 | 0.080 | 0.046 | 0.047 | 0.054 | 0.055 | 0.002 | 0.009 | 0.009 | 0.009 | 1.000 | 1.000 | 0.993 | 0.999 | 0.094 | 0.051 |
|  |  | 1.3 | 1.3 | 0 | 0.8 | 0.3 | 0 | 0.5 | 1 | 0.4 | 0.002 | -0.194 | 0.195 | 0.195 | 0.029 | 0.101 | 0.098 | 0.098 | 0.001 | 0.048 | 0.048 | 0.048 | 1.000 | 1.000 | 0.339 | 0.404 | 0.220 | 0.041 |
|  |  | 1.3 | 1.3 | 0 | 0.8 | 0 | 0 | 0.5 | 1 | 0.4 | 0.002 | -0.078 | 0.079 | 0.079 | 0.030 | 0.049 | 0.043 | 0.043 | 0.001 | 0.008 | 0.008 | 0.008 | 1.000 | 1.000 | 0.133 | 0.416 | 0.190 | 0.060 |
|  |  | -0.7 | -1 | 0.3 | -1 | 0.3 | 1 | 0 | 1 | 0.4 | 0.004 | -0.119 | 0.122 | 0.123 | 0.047 | 0.065 | 0.072 | 0.072 | 0.002 | 0.018 | 0.020 | 0.020 | 1.000 | 1.000 | 1.000 | 1.000 | 0.282 | 0.050 |
|  |  | 1.1 | 0.8 | 0.3 | 0.8 | 0.3 | 1 | 0.5 | 0 | 0.4 | 0.002 | -0.020 | 0.021 | 0.021 | 0.030 | 0.058 | 0.062 | 0.062 | 0.001 | 0.004 | 0.004 | 0.004 | 1.000 | 1.000 | 1.000 | 1.000 | 0.065 | 0.051 |
|  |  | 1.1 | 0.8 | 0.3 | 0.8 | 0.3 | 1 | 0 | 0 | 0.4 | 0.002 | -0.010 | 0.012 | 0.012 | 0.029 | 0.034 | 0.041 | 0.041 | 0.001 | 0.001 | 0.002 | 0.002 | 1.000 | 1.000 | 0.999 | 0.999 | 0.130 | 0.045 |
|  | GM1+GM2 | 0.8 | 0.5 | 0.3 | 0 | 0.3 | 1 | 0.5 | 1 | 0.4 | 0.001 | -0.277 | 0.278 | 0.278 | 0.037 | 0.197 | 0.195 | 0.195 | 0.001 | 0.115 | 0.115 | 0.115 | 1.000 | 0.124 | 0.941 | 0.947 | 0.023 | 0.946 |
|  |  | 1.3 | 1.3 | 0 | 0.8 | 0 | 1 | 0.5 | 1 | 0.4 | 0.002 | -0.103 | 0.105 | 0.105 | 0.036 | 0.083 | 0.083 | 0.083 | 0.001 | 0.017 | 0.018 | 0.018 | 1.000 | 1.000 | 0.789 | 0.869 | 0.005 | 0.948 |
|  |  | 1.3 | 1.3 | 0 | 0.8 | 0.3 | 0 | 0.5 | 1 | 0.4 | 0.001 | -0.268 | 0.269 | 0.269 | 0.028 | 0.191 | 0.191 | 0.191 | 0.001 | 0.109 | 0.109 | 0.109 | 1.000 | 0.997 | 0.222 | 0.239 | 0.034 | 0.935 |
|  |  | 1.3 | 1.3 | 0 | 0.8 | 0 | 0 | 0.5 | 1 | 0.4 | -0.001 | -0.106 | 0.105 | 0.105 | 0.029 | 0.083 | 0.079 | 0.079 | 0.001 | 0.018 | 0.017 | 0.017 | 1.000 | 1.000 | 0.143 | 0.231 | 0.039 | 0.932 |
|  |  | -0.7 | -1 | 0.3 | -1 | 0.3 | 1 | 0 | 1 | 0.4 | 0.001 | -0.161 | 0.162 | 0.162 | 0.037 | 0.116 | 0.115 | 0.115 | 0.001 | 0.039 | 0.039 | 0.039 | 1.000 | 1.000 | 0.963 | 0.968 | 0.057 | 0.935 |
|  |  | 1.1 | 0.8 | 0.3 | 0.8 | 0.3 | 1 | 0.5 | 0 | 0.4 | 0.001 | -0.058 | 0.060 | 0.060 | 0.030 | 0.175 | 0.172 | 0.172 | 0.001 | 0.034 | 0.033 | 0.033 | 1.000 | 0.939 | 0.780 | 0.798 | 0.008 | 0.931 |
|  |  | 1.1 | 0.8 | 0.3 | 0.8 | 0.3 | 1 | 0 | 0 | 0.4 | 0.001 | -0.039 | 0.040 | 0.040 | 0.030 | 0.105 | 0.106 | 0.106 | 0.001 | 0.013 | 0.013 | 0.013 | 1.000 | 1.000 | 0.827 | 0.855 | 0.032 | 0.940 |
|  | GM2+GY | 0.8 | 0.5 | 0.3 | 0 | 0.3 | 1 | 0.5 | 1 | 0.4 | 0.000 | -0.498 | 0.498 | 0.498 | 0.043 | 0.133 | 0.134 | 0.134 | 0.002 | 0.266 | 0.266 | 0.266 | 1.000 | 0.001 | 1.000 | 1.000 | 0.003 | 0.623 |
|  |  | 1.3 | 1.3 | 0 | 0.8 | 0 | 1 | 0.5 | 1 | 0.4 | 0.003 | -0.196 | 0.199 | 0.199 | 0.040 | 0.056 | 0.061 | 0.061 | 0.002 | 0.042 | 0.043 | 0.043 | 1.000 | 1.000 | 0.996 | 0.998 | 0.003 | 0.623 |
|  |  | 1.3 | 1.3 | 0 | 0.8 | 0.3 | 0 | 0.5 | 1 | 0.4 | 0.003 | -0.483 | 0.486 | 0.486 | 0.029 | 0.134 | 0.134 | 0.134 | 0.001 | 0.251 | 0.254 | 0.254 | 1.000 | 0.991 | 0.802 | 0.824 | 0.006 | 0.599 |
|  |  | 1.3 | 1.3 | 0 | 0.8 | 0 | 0 | 0.5 | 1 | 0.4 | 0.002 | -0.199 | 0.201 | 0.201 | 0.029 | 0.056 | 0.054 | 0.054 | 0.001 | 0.043 | 0.043 | 0.043 | 1.000 | 1.000 | 0.758 | 0.853 | 0.004 | 0.609 |
|  |  | -0.7 | -1 | 0.3 | -1 | 0.3 | 1 | 0 | 1 | 0.4 | 0.003 | -0.294 | 0.297 | 0.297 | 0.042 | 0.081 | 0.086 | 0.086 | 0.002 | 0.093 | 0.095 | 0.096 | 1.000 | 1.000 | 1.000 | 1.000 | 0.051 | 0.623 |
|  |  | 1.1 | 0.8 | 0.3 | 0.8 | 0.3 | 1 | 0.5 | 0 | 0.4 | 0.002 | -0.046 | 0.048 | 0.048 | 0.024 | 0.085 | 0.088 | 0.088 | 0.001 | 0.009 | 0.010 | 0.010 | 1.000 | 0.998 | 0.949 | 0.955 | 0.018 | 0.610 |
|  |  | 1.1 | 0.8 | 0.3 | 0.8 | 0.3 | 1 | 0 | 0 | 0.4 | 0.001 | -0.029 | 0.030 | 0.030 | 0.024 | 0.054 | 0.054 | 0.054 | 0.001 | 0.004 | 0.004 | 0.004 | 1.000 | 1.000 | 0.961 | 0.974 | 0.061 | 0.612 |
|  | GM1+GY | 0.8 | 0.5 | 0.3 | 0 | 0.3 | 1 | 0.5 | 1 | 0.4 | 0.004 | -0.089 | 0.093 | 0.093 | 0.035 | 0.085 | 0.086 | 0.086 | 0.001 | 0.015 | 0.016 | 0.016 | 1.000 | 0.967 | 0.994 | 0.995 | 0.749 | 0.091 |
|  |  | 1.3 | 1.3 | 0 | 0.8 | 0 | 1 | 0.5 | 1 | 0.4 | 0.001 | -0.035 | 0.036 | 0.036 | 0.035 | 0.039 | 0.044 | 0.044 | 0.001 | 0.003 | 0.003 | 0.003 | 1.000 | 1.000 | 0.973 | 0.992 | 0.492 | 0.092 |
|  |  | 1.3 | 1.3 | 0 | 0.8 | 0.3 | 0 | 0.5 | 1 | 0.4 | 0.001 | -0.087 | 0.087 | 0.087 | 0.021 | 0.081 | 0.079 | 0.079 | 0.000 | 0.014 | 0.014 | 0.014 | 1.000 | 1.000 | 0.023 | 0.037 | 0.753 | 0.096 |
|  |  | 1.3 | 1.3 | 0 | 0.8 | 0 | 0 | 0.5 | 1 | 0.4 | 0.001 | -0.036 | 0.037 | 0.037 | 0.020 | 0.038 | 0.033 | 0.033 | 0.000 | 0.003 | 0.002 | 0.002 | 1.000 | 1.000 | 0.001 | 0.043 | 0.740 | 0.107 |
|  |  | -0.7 | -1 | 0.3 | -1 | 0.3 | 1 | 0 | 1 | 0.4 | 0.000 | -0.053 | 0.052 | 0.053 | 0.037 | 0.051 | 0.056 | 0.056 | 0.001 | 0.005 | 0.006 | 0.006 | 1.000 | 1.000 | 1.000 | 1.000 | 0.950 | 0.106 |
|  |  | 1.1 | 0.8 | 0.3 | 0.8 | 0.3 | 1 | 0.5 | 0 | 0.4 | 0.002 | -0.022 | 0.023 | 0.023 | 0.030 | 0.058 | 0.063 | 0.063 | 0.001 | 0.004 | 0.004 | 0.004 | 1.000 | 1.000 | 1.000 | 1.000 | 0.056 | 0.097 |
|  |  | 1.1 | 0.8 | 0.3 | 0.8 | 0.3 | 1 | 0 | 0 | 0.4 | 0.001 | -0.011 | 0.012 | 0.012 | 0.029 | 0.037 | 0.043 | 0.043 | 0.001 | 0.001 | 0.002 | 0.002 | 1.000 | 1.000 | 0.999 | 1.000 | 0.134 | 0.083 |

TE: total effect; DE: direct effect; IE: indirect effect; IE_d: indirect effect calculated by difference method; IE_p: indirect effect calculated by product method; GM2: there is a direct effect from G to M2; GM1: there is a direct effect from G to M1; GY: there is a direct effect from G to Y; GM1+GM2: there is a direct effect from G to M1 and M2; GM1+GY: there is a direct effect from G to M1 and Y; GM2+GY: there is a direct effect from G to M2 and Y.

**eTable 39.** Simulation of PSE-MR when missing M1 for varying $r_{12}$

| $r_{12}$ | BIAS | | | | SE | | | | MSE | | | | Power | | | |
| --- | --- | --- | --- | --- | --- | --- | --- | --- | --- | --- | --- | --- | --- | --- | --- | --- |
|  | TE | DE | IE_d | IE_p | TE | DE | IE_d | IE_p | TE | DE | IE_d | IE_p | TE | DE | IE_d | IE_p |
| 0 | 0.009 | -0.031 | 0.041 | 0.041 | 0.069 | 0.088 | 0.087 | 0.087 | 0.005 | 0.009 | 0.009 | 0.009 | 1.000 | 0.996 | 0.948 | 0.992 |
| 0.1 | 0.011 | -0.076 | 0.087 | 0.087 | 0.073 | 0.092 | 0.101 | 0.101 | 0.005 | 0.014 | 0.018 | 0.018 | 1.000 | 0.975 | 0.983 | 0.995 |
| 0.2 | 0.010 | -0.126 | 0.136 | 0.137 | 0.076 | 0.098 | 0.109 | 0.109 | 0.006 | 0.026 | 0.030 | 0.031 | 1.000 | 0.906 | 0.998 | 0.999 |
| 0.3 | 0.005 | -0.171 | 0.176 | 0.176 | 0.078 | 0.103 | 0.111 | 0.111 | 0.006 | 0.040 | 0.043 | 0.043 | 1.000 | 0.775 | 1.000 | 1.000 |
| 0.4 | 0.011 | -0.222 | 0.233 | 0.233 | 0.084 | 0.110 | 0.125 | 0.125 | 0.007 | 0.061 | 0.070 | 0.070 | 1.000 | 0.513 | 0.999 | 0.999 |
| 0.5 | 0.009 | -0.277 | 0.286 | 0.287 | 0.082 | 0.109 | 0.124 | 0.124 | 0.007 | 0.089 | 0.098 | 0.098 | 1.000 | 0.274 | 1.000 | 1.000 |
| 0.6 | 0.012 | -0.317 | 0.329 | 0.329 | 0.086 | 0.107 | 0.127 | 0.127 | 0.008 | 0.112 | 0.124 | 0.124 | 1.000 | 0.109 | 1.000 | 1.000 |
| 0.7 | 0.012 | -0.358 | 0.370 | 0.370 | 0.091 | 0.103 | 0.126 | 0.126 | 0.008 | 0.139 | 0.153 | 0.153 | 1.000 | 0.030 | 1.000 | 1.000 |
| 0.8 | 0.010 | -0.397 | 0.407 | 0.407 | 0.095 | 0.101 | 0.130 | 0.130 | 0.009 | 0.168 | 0.182 | 0.183 | 1.000 | 0.005 | 1.000 | 1.000 |
| 0.9 | 0.012 | -0.421 | 0.433 | 0.433 | 0.102 | 0.095 | 0.131 | 0.131 | 0.010 | 0.186 | 0.204 | 0.204 | 1.000 | 0.002 | 1.000 | 1.000 |
| 1 | 0.013 | -0.440 | 0.452 | 0.452 | 0.104 | 0.102 | 0.142 | 0.142 | 0.011 | 0.204 | 0.225 | 0.225 | 1.000 | 0.000 | 1.000 | 1.000 |

TE: total effect; DE: direct effect; IE: indirect effect; IE_d: indirect effect calculated by difference method; IE_p: indirect effect calculated by product method.

**eTable 40.** Simulation of PSE-MR when missing M2

|  |  | TRUE | | | | | | | | | BIAS | | | | SE | | | | MSE | | | | Power/Type I error | | | | | |
| --- | --- | --- | --- | --- | --- | --- | --- | --- | --- | --- | --- | --- | --- | --- | --- | --- | --- | --- | --- | --- | --- | --- | --- | --- | --- | --- | --- | --- |
|  |  | TE | DE | IE | $\theta_{0}$ | $\alpha_{1}$ | $\delta_{1}$ | $\alpha_{2}$ | $\delta_{2}$ | $r_{12}$ | TE | DE | IE_d | IE_p | TE | DE | IE_d | IE_p | TE | DE | IE_d | IE_p | TE | DE | IE_d | IE_p | GY | GM |
| PSE-MR | Standard MR | 0.8 | 0.5 | 0.3 | 0 | 0.3 | 1 | 0.5 | 1 | 0.4 | 0.007 | -0.063 | 0.070 | 0.070 | 0.079 | 0.123 | 0.133 | 0.133 | 0.006 | 0.019 | 0.023 | 0.023 | 1.000 | 0.318 | 1.000 | 1.000 |  |  |
|  |  | 1.3 | 1.3 | 0 | 0.8 | 0 | 1 | 0.5 | 1 | 0.4 | 0.012 | -0.064 | 0.076 | 0.076 | 0.083 | 0.126 | 0.135 | 0.135 | 0.007 | 0.020 | 0.024 | 0.024 | 1.000 | 0.993 | 0.999 | 1.000 |  |  |
|  |  | 1.3 | 1.3 | 0 | 0.8 | 0.3 | 0 | 0.5 | 1 | 0.4 | 0.004 | -0.030 | 0.034 | 0.034 | 0.048 | 0.042 | 0.059 | 0.059 | 0.002 | 0.003 | 0.005 | 0.005 | 1.000 | 1.000 | 1.000 | 1.000 |  |  |
|  |  | 1.3 | 1.3 | 0 | 0.8 | 0 | 0 | 0.5 | 1 | 0.4 | 0.007 | -0.029 | 0.035 | 0.035 | 0.049 | 0.040 | 0.060 | 0.060 | 0.002 | 0.002 | 0.005 | 0.005 | 1.000 | 1.000 | 1.000 | 1.000 |  |  |
|  |  | -0.7 | -1 | 0.3 | -1 | 0.3 | 1 | 0 | 1 | 0.4 | 0.007 | 0.002 | 0.005 | 0.006 | 0.079 | 0.049 | 0.066 | 0.066 | 0.006 | 0.002 | 0.004 | 0.004 | 1.000 | 1.000 | 0.004 | 0.059 |  |  |
|  |  | 1.1 | 0.8 | 0.3 | 0.8 | 0.3 | 1 | 0.5 | 0 | 0.4 | 0.007 | -0.063 | 0.069 | 0.069 | 0.052 | 0.129 | 0.119 | 0.119 | 0.003 | 0.021 | 0.019 | 0.019 | 1.000 | 1.000 | 0.472 | 0.604 |  |  |
|  |  | 1.1 | 0.8 | 0.3 | 0.8 | 0.3 | 1 | 0 | 0 | 0.4 | 0.007 | 0.006 | 0.001 | 0.001 | 0.054 | 0.049 | 0.025 | 0.025 | 0.003 | 0.002 | 0.001 | 0.001 | 1.000 | 1.000 | 0.000 | 0.028 |  |  |
| PSE-Egger | sc2 | 0.8 | 0.5 | 0.3 | 0 | 0.3 | 1 | 0.5 | 1 | 0.4 | 0.001 | -0.028 | 0.029 | 0.029 | 0.050 | 0.163 | 0.165 | 0.165 | 0.002 | 0.027 | 0.028 | 0.028 | 1.000 | 0.369 | 0.990 | 0.992 | 0.045 | 0.047 |
|  |  | 1.3 | 1.3 | 0 | 0.8 | 0 | 1 | 0.5 | 1 | 0.4 | 0.001 | -0.029 | 0.030 | 0.030 | 0.049 | 0.156 | 0.157 | 0.157 | 0.002 | 0.025 | 0.025 | 0.025 | 1.000 | 0.996 | 0.991 | 0.995 | 0.019 | 0.037 |
|  |  | 1.3 | 1.3 | 0 | 0.8 | 0.3 | 0 | 0.5 | 1 | 0.4 | 0.003 | -0.009 | 0.012 | 0.012 | 0.036 | 0.107 | 0.107 | 0.107 | 0.001 | 0.011 | 0.012 | 0.012 | 1.000 | 1.000 | 0.993 | 0.994 | 0.017 | 0.042 |
|  |  | 1.3 | 1.3 | 0 | 0.8 | 0 | 0 | 0.5 | 1 | 0.4 | -0.001 | -0.021 | 0.020 | 0.020 | 0.034 | 0.106 | 0.107 | 0.107 | 0.001 | 0.012 | 0.012 | 0.012 | 1.000 | 1.000 | 0.992 | 0.995 | 0.012 | 0.039 |
|  |  | -0.7 | -1 | 0.3 | -1 | 0.3 | 1 | 0 | 1 | 0.4 | 0.001 | 0.001 | -0.001 | -0.001 | 0.048 | 0.036 | 0.035 | 0.035 | 0.002 | 0.001 | 0.001 | 0.001 | 1.000 | 1.000 | 0.001 | 0.051 | 0.050 | 0.041 |
|  |  | 1.1 | 0.8 | 0.3 | 0.8 | 0.3 | 1 | 0.5 | 0 | 0.4 | 0.002 | -0.031 | 0.033 | 0.033 | 0.039 | 0.158 | 0.152 | 0.152 | 0.002 | 0.026 | 0.024 | 0.024 | 1.000 | 1.000 | 0.314 | 0.370 | 0.055 | 0.063 |
|  |  | 1.1 | 0.8 | 0.3 | 0.8 | 0.3 | 1 | 0 | 0 | 0.4 | 0.001 | 0.001 | 0.000 | 0.000 | 0.038 | 0.037 | 0.012 | 0.012 | 0.001 | 0.001 | 0.000 | 0.000 | 1.000 | 1.000 | 0.000 | 0.009 | 0.054 | 0.044 |
|  | sc3 | 0.8 | 0.5 | 0.3 | 0 | 0.3 | 1 | 0.5 | 1 | 0.4 | 0.002 | -0.110 | 0.111 | 0.111 | 0.031 | 0.155 | 0.154 | 0.154 | 0.001 | 0.036 | 0.036 | 0.036 | 1.000 | 0.023 | 0.967 | 0.970 | 0.453 | 0.809 |
|  |  | 1.3 | 1.3 | 0 | 0.8 | 0 | 1 | 0.5 | 1 | 0.4 | 0.003 | -0.104 | 0.107 | 0.107 | 0.031 | 0.161 | 0.161 | 0.161 | 0.001 | 0.037 | 0.037 | 0.037 | 1.000 | 0.670 | 0.875 | 0.884 | 0.192 | 0.823 |
|  |  | 1.3 | 1.3 | 0 | 0.8 | 0.3 | 0 | 0.5 | 1 | 0.4 | 0.000 | -0.057 | 0.057 | 0.057 | 0.020 | 0.108 | 0.108 | 0.108 | 0.000 | 0.015 | 0.015 | 0.015 | 1.000 | 0.973 | 0.781 | 0.798 | 0.016 | 0.787 |
|  |  | 1.3 | 1.3 | 0 | 0.8 | 0 | 0 | 0.5 | 1 | 0.4 | 0.002 | -0.052 | 0.054 | 0.054 | 0.021 | 0.106 | 0.106 | 0.106 | 0.000 | 0.014 | 0.014 | 0.014 | 1.000 | 0.982 | 0.768 | 0.790 | 0.019 | 0.783 |
|  |  | -0.7 | -1 | 0.3 | -1 | 0.3 | 1 | 0 | 1 | 0.4 | 0.003 | 0.002 | 0.001 | 0.001 | 0.030 | 0.020 | 0.023 | 0.023 | 0.001 | 0.000 | 0.001 | 0.001 | 1.000 | 1.000 | 0.004 | 0.068 | 0.791 | 0.810 |
|  |  | 1.1 | 0.8 | 0.3 | 0.8 | 0.3 | 1 | 0.5 | 0 | 0.4 | 0.003 | -0.117 | 0.119 | 0.119 | 0.022 | 0.162 | 0.160 | 0.160 | 0.000 | 0.040 | 0.040 | 0.040 | 1.000 | 0.997 | 0.168 | 0.187 | 0.399 | 0.797 |
|  |  | 1.1 | 0.8 | 0.3 | 0.8 | 0.3 | 1 | 0 | 0 | 0.4 | 0.000 | 0.000 | 0.000 | 0.000 | 0.022 | 0.021 | 0.010 | 0.010 | 0.000 | 0.000 | 0.000 | 0.000 | 1.000 | 1.000 | 0.000 | 0.007 | 0.573 | 0.801 |
|  | sc4 | 0.8 | 0.5 | 0.3 | 0 | 0.3 | 1 | 0.5 | 1 | 0.4 | 0.074 | -0.841 | 0.914 | 0.914 | 0.103 | 0.100 | 0.137 | 0.137 | 0.016 | 0.717 | 0.855 | 0.855 | 1.000 | 0.989 | 1.000 | 1.000 | 0.050 | 0.915 |
|  |  | 1.3 | 1.3 | 0 | 0.8 | 0 | 1 | 0.5 | 1 | 0.4 | 0.074 | -0.846 | 0.920 | 0.920 | 0.096 | 0.105 | 0.134 | 0.134 | 0.015 | 0.727 | 0.864 | 0.864 | 1.000 | 0.003 | 1.000 | 1.000 | 0.008 | 0.920 |
|  |  | 1.3 | 1.3 | 0 | 0.8 | 0.3 | 0 | 0.5 | 1 | 0.4 | 0.044 | -0.424 | 0.468 | 0.468 | 0.060 | 0.061 | 0.081 | 0.081 | 0.006 | 0.184 | 0.226 | 0.226 | 1.000 | 0.945 | 1.000 | 1.000 | 0.000 | 0.920 |
|  |  | 1.3 | 1.3 | 0 | 0.8 | 0 | 0 | 0.5 | 1 | 0.4 | 0.043 | -0.422 | 0.465 | 0.465 | 0.059 | 0.059 | 0.082 | 0.082 | 0.005 | 0.182 | 0.223 | 0.223 | 1.000 | 0.941 | 1.000 | 1.000 | 0.000 | 0.920 |
|  |  | -0.7 | -1 | 0.3 | -1 | 0.3 | 1 | 0 | 1 | 0.4 | 0.065 | 0.006 | 0.059 | 0.061 | 0.104 | 0.029 | 0.099 | 0.099 | 0.015 | 0.001 | 0.013 | 0.013 | 1.000 | 1.000 | 0.082 | 0.124 | 0.145 | 0.931 |
|  |  | 1.1 | 0.8 | 0.3 | 0.8 | 0.3 | 1 | 0.5 | 0 | 0.4 | 0.050 | -0.847 | 0.897 | 0.897 | 0.073 | 0.094 | 0.109 | 0.109 | 0.008 | 0.726 | 0.816 | 0.816 | 1.000 | 0.467 | 1.000 | 1.000 | 0.051 | 0.920 |
|  |  | 1.1 | 0.8 | 0.3 | 0.8 | 0.3 | 1 | 0 | 0 | 0.4 | 0.046 | 0.006 | 0.040 | 0.039 | 0.074 | 0.031 | 0.069 | 0.069 | 0.008 | 0.001 | 0.006 | 0.006 | 1.000 | 1.000 | 0.066 | 0.127 | 0.082 | 0.931 |
|  | GM_2_ | 0.8 | 0.5 | 0.3 | 0 | 0.3 | 1 | 0.5 | 1 | 0.4 | -0.001 | -0.033 | 0.032 | 0.032 | 0.046 | 0.127 | 0.129 | 0.129 | 0.002 | 0.017 | 0.018 | 0.018 | 1.000 | 0.500 | 1.000 | 1.000 | 0.243 | 0.047 |
|  |  | 1.3 | 1.3 | 0 | 0.8 | 0 | 1 | 0.5 | 1 | 0.4 | 0.002 | -0.034 | 0.037 | 0.037 | 0.046 | 0.131 | 0.134 | 0.134 | 0.002 | 0.018 | 0.019 | 0.019 | 1.000 | 0.998 | 0.997 | 0.997 | 0.102 | 0.049 |
|  |  | 1.3 | 1.3 | 0 | 0.8 | 0.3 | 0 | 0.5 | 1 | 0.4 | 0.002 | -0.012 | 0.014 | 0.014 | 0.036 | 0.109 | 0.110 | 0.110 | 0.001 | 0.012 | 0.012 | 0.012 | 1.000 | 1.000 | 0.985 | 0.987 | 0.013 | 0.047 |
|  |  | 1.3 | 1.3 | 0 | 0.8 | 0 | 0 | 0.5 | 1 | 0.4 | 0.000 | -0.017 | 0.017 | 0.017 | 0.035 | 0.110 | 0.110 | 0.110 | 0.001 | 0.012 | 0.012 | 0.012 | 1.000 | 1.000 | 0.988 | 0.992 | 0.017 | 0.055 |
|  |  | -0.7 | -1 | 0.3 | -1 | 0.3 | 1 | 0 | 1 | 0.4 | 0.000 | 0.000 | 0.000 | 0.000 | 0.046 | 0.028 | 0.037 | 0.037 | 0.002 | 0.001 | 0.001 | 0.001 | 1.000 | 1.000 | 0.004 | 0.063 | 0.251 | 0.050 |
|  |  | 1.1 | 0.8 | 0.3 | 0.8 | 0.3 | 1 | 0.5 | 0 | 0.4 | 0.001 | -0.028 | 0.029 | 0.029 | 0.032 | 0.132 | 0.128 | 0.128 | 0.001 | 0.018 | 0.017 | 0.017 | 1.000 | 1.000 | 0.361 | 0.406 | 0.224 | 0.049 |
|  |  | 1.1 | 0.8 | 0.3 | 0.8 | 0.3 | 1 | 0 | 0 | 0.4 | 0.000 | 0.000 | 0.000 | 0.000 | 0.031 | 0.030 | 0.013 | 0.013 | 0.001 | 0.001 | 0.000 | 0.000 | 1.000 | 1.000 | 0.000 | 0.024 | 0.253 | 0.053 |
|  | GM_1_ | 0.8 | 0.5 | 0.3 | 0 | 0.3 | 1 | 0.5 | 1 | 0.4 | 0.002 | -0.094 | 0.096 | 0.096 | 0.042 | 0.277 | 0.278 | 0.278 | 0.002 | 0.086 | 0.086 | 0.086 | 1.000 | 0.094 | 0.797 | 0.807 | 0.054 | 0.791 |
|  |  | 1.3 | 1.3 | 0 | 0.8 | 0 | 1 | 0.5 | 1 | 0.4 | 0.004 | -0.093 | 0.096 | 0.096 | 0.041 | 0.284 | 0.280 | 0.280 | 0.002 | 0.089 | 0.088 | 0.088 | 1.000 | 0.629 | 0.756 | 0.771 | 0.029 | 0.798 |
|  |  | 1.3 | 1.3 | 0 | 0.8 | 0.3 | 0 | 0.5 | 1 | 0.4 | 0.001 | -0.048 | 0.049 | 0.049 | 0.029 | 0.204 | 0.203 | 0.203 | 0.001 | 0.044 | 0.043 | 0.043 | 1.000 | 0.886 | 0.666 | 0.684 | 0.017 | 0.790 |
|  |  | 1.3 | 1.3 | 0 | 0.8 | 0 | 0 | 0.5 | 1 | 0.4 | 0.000 | -0.052 | 0.051 | 0.051 | 0.028 | 0.202 | 0.199 | 0.199 | 0.001 | 0.043 | 0.042 | 0.042 | 1.000 | 0.898 | 0.677 | 0.696 | 0.020 | 0.803 |
|  |  | -0.7 | -1 | 0.3 | -1 | 0.3 | 1 | 0 | 1 | 0.4 | 0.003 | 0.003 | 0.000 | 0.000 | 0.043 | 0.037 | 0.023 | 0.023 | 0.002 | 0.001 | 0.001 | 0.001 | 1.000 | 1.000 | 0.000 | 0.042 | 0.056 | 0.814 |
|  |  | 1.1 | 0.8 | 0.3 | 0.8 | 0.3 | 1 | 0.5 | 0 | 0.4 | 0.000 | -0.115 | 0.115 | 0.115 | 0.036 | 0.286 | 0.283 | 0.283 | 0.001 | 0.095 | 0.093 | 0.093 | 1.000 | 0.911 | 0.216 | 0.222 | 0.046 | 0.811 |
|  |  | 1.1 | 0.8 | 0.3 | 0.8 | 0.3 | 1 | 0 | 0 | 0.4 | 0.003 | 0.002 | 0.001 | 0.001 | 0.036 | 0.036 | 0.012 | 0.012 | 0.001 | 0.001 | 0.000 | 0.000 | 1.000 | 1.000 | 0.000 | 0.011 | 0.054 | 0.815 |
|  | GY | 0.8 | 0.5 | 0.3 | 0 | 0.3 | 1 | 0.5 | 1 | 0.4 | 0.003 | -0.028 | 0.031 | 0.031 | 0.047 | 0.124 | 0.128 | 0.128 | 0.002 | 0.016 | 0.017 | 0.017 | 1.000 | 0.488 | 0.999 | 1.000 | 0.218 | 0.050 |
|  |  | 1.3 | 1.3 | 0 | 0.8 | 0 | 1 | 0.5 | 1 | 0.4 | 0.001 | -0.035 | 0.035 | 0.036 | 0.046 | 0.129 | 0.132 | 0.132 | 0.002 | 0.018 | 0.019 | 0.019 | 1.000 | 0.997 | 0.995 | 0.997 | 0.109 | 0.049 |
|  |  | 1.3 | 1.3 | 0 | 0.8 | 0.3 | 0 | 0.5 | 1 | 0.4 | 0.002 | -0.014 | 0.016 | 0.016 | 0.029 | 0.058 | 0.063 | 0.063 | 0.001 | 0.004 | 0.004 | 0.004 | 1.000 | 1.000 | 0.999 | 1.000 | 0.082 | 0.049 |
|  |  | 1.3 | 1.3 | 0 | 0.8 | 0 | 0 | 0.5 | 1 | 0.4 | 0.002 | -0.017 | 0.019 | 0.019 | 0.030 | 0.060 | 0.065 | 0.065 | 0.001 | 0.004 | 0.005 | 0.005 | 1.000 | 1.000 | 1.000 | 1.000 | 0.071 | 0.058 |
|  |  | -0.7 | -1 | 0.3 | -1 | 0.3 | 1 | 0 | 1 | 0.4 | 0.004 | 0.001 | 0.003 | 0.003 | 0.047 | 0.030 | 0.036 | 0.036 | 0.002 | 0.001 | 0.001 | 0.001 | 1.000 | 1.000 | 0.005 | 0.069 | 0.261 | 0.052 |
|  |  | 1.1 | 0.8 | 0.3 | 0.8 | 0.3 | 1 | 0.5 | 0 | 0.4 | 0.002 | -0.034 | 0.036 | 0.036 | 0.030 | 0.132 | 0.130 | 0.130 | 0.001 | 0.019 | 0.018 | 0.018 | 1.000 | 1.000 | 0.385 | 0.432 | 0.198 | 0.050 |
|  |  | 1.1 | 0.8 | 0.3 | 0.8 | 0.3 | 1 | 0 | 0 | 0.4 | 0.002 | 0.002 | 0.000 | 0.000 | 0.029 | 0.028 | 0.012 | 0.012 | 0.001 | 0.001 | 0.000 | 0.000 | 1.000 | 1.000 | 0.000 | 0.015 | 0.217 | 0.048 |
|  | GM_1_+GM_2_ | 0.8 | 0.5 | 0.3 | 0 | 0.3 | 1 | 0.5 | 1 | 0.4 | 0.001 | -0.106 | 0.108 | 0.108 | 0.037 | 0.229 | 0.228 | 0.228 | 0.001 | 0.064 | 0.064 | 0.064 | 1.000 | 0.091 | 0.904 | 0.905 | 0.102 | 0.812 |
|  |  | 1.3 | 1.3 | 0 | 0.8 | 0 | 1 | 0.5 | 1 | 0.4 | 0.002 | -0.113 | 0.116 | 0.116 | 0.036 | 0.232 | 0.232 | 0.232 | 0.001 | 0.067 | 0.067 | 0.067 | 1.000 | 0.624 | 0.825 | 0.828 | 0.046 | 0.814 |
|  |  | 1.3 | 1.3 | 0 | 0.8 | 0.3 | 0 | 0.5 | 1 | 0.4 | 0.001 | -0.053 | 0.054 | 0.054 | 0.028 | 0.197 | 0.196 | 0.196 | 0.001 | 0.042 | 0.042 | 0.042 | 1.000 | 0.895 | 0.699 | 0.712 | 0.011 | 0.795 |
|  |  | 1.3 | 1.3 | 0 | 0.8 | 0 | 0 | 0.5 | 1 | 0.4 | -0.001 | -0.050 | 0.049 | 0.049 | 0.029 | 0.202 | 0.201 | 0.201 | 0.001 | 0.043 | 0.043 | 0.043 | 1.000 | 0.897 | 0.676 | 0.693 | 0.021 | 0.808 |
|  |  | -0.7 | -1 | 0.3 | -1 | 0.3 | 1 | 0 | 1 | 0.4 | 0.001 | 0.000 | 0.001 | 0.001 | 0.037 | 0.030 | 0.022 | 0.022 | 0.001 | 0.001 | 0.001 | 0.001 | 1.000 | 1.000 | 0.001 | 0.048 | 0.127 | 0.803 |
|  |  | 1.1 | 0.8 | 0.3 | 0.8 | 0.3 | 1 | 0.5 | 0 | 0.4 | 0.001 | -0.117 | 0.118 | 0.118 | 0.030 | 0.228 | 0.226 | 0.226 | 0.001 | 0.066 | 0.065 | 0.065 | 1.000 | 0.970 | 0.249 | 0.260 | 0.090 | 0.821 |
|  |  | 1.1 | 0.8 | 0.3 | 0.8 | 0.3 | 1 | 0 | 0 | 0.4 | 0.001 | 0.000 | 0.001 | 0.001 | 0.030 | 0.030 | 0.011 | 0.011 | 0.001 | 0.001 | 0.000 | 0.000 | 1.000 | 1.000 | 0.000 | 0.015 | 0.122 | 0.797 |
|  | GM_2_+GY | 0.8 | 0.5 | 0.3 | 0 | 0.3 | 1 | 0.5 | 1 | 0.4 | 0.000 | -0.035 | 0.035 | 0.036 | 0.043 | 0.087 | 0.093 | 0.093 | 0.002 | 0.009 | 0.010 | 0.010 | 1.000 | 0.507 | 1.000 | 1.000 | 0.829 | 0.051 |
|  |  | 1.3 | 1.3 | 0 | 0.8 | 0 | 1 | 0.5 | 1 | 0.4 | 0.003 | -0.030 | 0.033 | 0.033 | 0.040 | 0.091 | 0.097 | 0.097 | 0.002 | 0.009 | 0.010 | 0.010 | 1.000 | 1.000 | 1.000 | 1.000 | 0.567 | 0.054 |
|  |  | 1.3 | 1.3 | 0 | 0.8 | 0.3 | 0 | 0.5 | 1 | 0.4 | 0.003 | -0.015 | 0.018 | 0.018 | 0.029 | 0.063 | 0.068 | 0.068 | 0.001 | 0.004 | 0.005 | 0.005 | 1.000 | 1.000 | 0.999 | 0.999 | 0.087 | 0.058 |
|  |  | 1.3 | 1.3 | 0 | 0.8 | 0 | 0 | 0.5 | 1 | 0.4 | 0.002 | -0.014 | 0.016 | 0.016 | 0.029 | 0.059 | 0.064 | 0.064 | 0.001 | 0.004 | 0.004 | 0.004 | 1.000 | 1.000 | 1.000 | 1.000 | 0.071 | 0.048 |
|  |  | -0.7 | -1 | 0.3 | -1 | 0.3 | 1 | 0 | 1 | 0.4 | 0.003 | 0.001 | 0.002 | 0.002 | 0.042 | 0.021 | 0.037 | 0.037 | 0.002 | 0.000 | 0.001 | 0.001 | 1.000 | 1.000 | 0.015 | 0.073 | 0.957 | 0.048 |
|  |  | 1.1 | 0.8 | 0.3 | 0.8 | 0.3 | 1 | 0.5 | 0 | 0.4 | 0.002 | -0.034 | 0.036 | 0.036 | 0.024 | 0.089 | 0.089 | 0.089 | 0.001 | 0.009 | 0.009 | 0.009 | 1.000 | 1.000 | 0.346 | 0.403 | 0.775 | 0.047 |
|  |  | 1.1 | 0.8 | 0.3 | 0.8 | 0.3 | 1 | 0 | 0 | 0.4 | 0.001 | 0.001 | 0.000 | 0.000 | 0.024 | 0.021 | 0.012 | 0.012 | 0.001 | 0.000 | 0.000 | 0.000 | 1.000 | 1.000 | 0.000 | 0.028 | 0.851 | 0.056 |
|  | GM_1_+GY | 0.8 | 0.5 | 0.3 | 0 | 0.3 | 1 | 0.5 | 1 | 0.4 | 0.004 | -0.110 | 0.114 | 0.114 | 0.035 | 0.221 | 0.219 | 0.219 | 0.001 | 0.061 | 0.061 | 0.061 | 1.000 | 0.084 | 0.929 | 0.931 | 0.093 | 0.799 |
|  |  | 1.3 | 1.3 | 0 | 0.8 | 0 | 1 | 0.5 | 1 | 0.4 | 0.001 | -0.111 | 0.113 | 0.113 | 0.035 | 0.231 | 0.229 | 0.229 | 0.001 | 0.066 | 0.065 | 0.065 | 1.000 | 0.644 | 0.841 | 0.850 | 0.033 | 0.810 |
|  |  | 1.3 | 1.3 | 0 | 0.8 | 0.3 | 0 | 0.5 | 1 | 0.4 | 0.001 | -0.055 | 0.056 | 0.056 | 0.021 | 0.108 | 0.109 | 0.109 | 0.000 | 0.015 | 0.015 | 0.015 | 1.000 | 0.962 | 0.775 | 0.784 | 0.016 | 0.805 |
|  |  | 1.3 | 1.3 | 0 | 0.8 | 0 | 0 | 0.5 | 1 | 0.4 | 0.001 | -0.055 | 0.057 | 0.057 | 0.020 | 0.111 | 0.113 | 0.113 | 0.000 | 0.015 | 0.016 | 0.016 | 1.000 | 0.967 | 0.767 | 0.781 | 0.013 | 0.803 |
|  |  | -0.7 | -1 | 0.3 | -1 | 0.3 | 1 | 0 | 1 | 0.4 | 0.000 | -0.001 | 0.000 | 0.000 | 0.037 | 0.029 | 0.023 | 0.023 | 0.001 | 0.001 | 0.001 | 0.001 | 1.000 | 1.000 | 0.001 | 0.057 | 0.152 | 0.825 |
|  |  | 1.1 | 0.8 | 0.3 | 0.8 | 0.3 | 1 | 0.5 | 0 | 0.4 | 0.002 | -0.116 | 0.117 | 0.117 | 0.030 | 0.227 | 0.224 | 0.224 | 0.001 | 0.065 | 0.064 | 0.064 | 1.000 | 0.975 | 0.235 | 0.240 | 0.093 | 0.798 |
|  |  | 1.1 | 0.8 | 0.3 | 0.8 | 0.3 | 1 | 0 | 0 | 0.4 | 0.001 | 0.001 | 0.000 | 0.000 | 0.029 | 0.030 | 0.011 | 0.011 | 0.001 | 0.001 | 0.000 | 0.000 | 1.000 | 1.000 | 0.000 | 0.019 | 0.142 | 0.830 |

TE: total effect; DE: direct effect; IE: indirect effect; IE_d: indirect effect calculated by difference method; IE_p: indirect effect calculated by product method.

#### 13 Additional simulation scenario 7: Performance of PSE-MR with different number of SNPs and mediators

We also simulated the performance of PSE-MR with different number of SNPs and mediators. Then we found the minimize number of SNPs with different number of mediators. Results were presented in eFigure 9-10. Left was the performance of PSE-IVW in standard MR and the right was PSE-Egger with directional pleiotropy and the InSIDE assumption was satisfied. The number of genetic variants required for power greater than 80% increasing with the number of mediators. PSE-Egger needed more genetic variants than PSE-IVW to reach 80% of power.

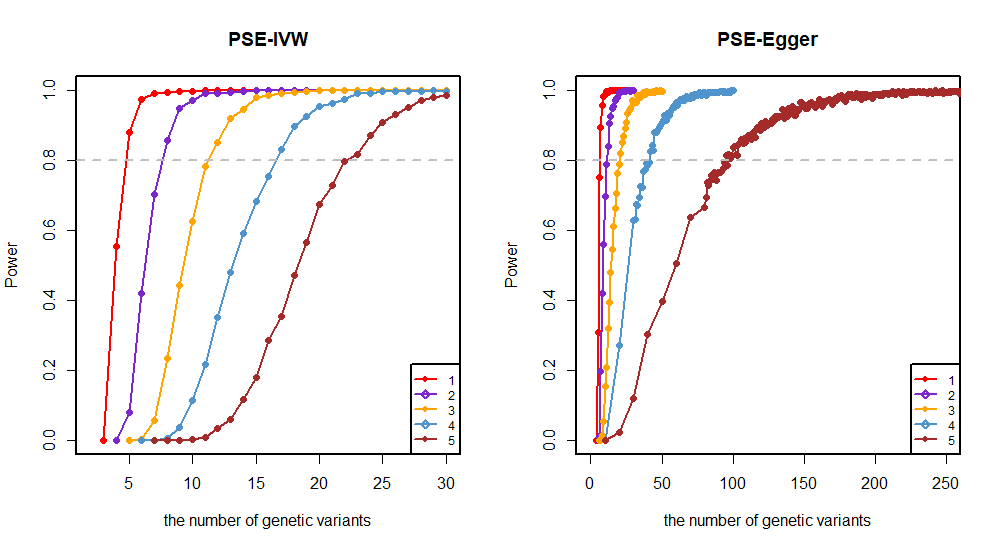

**eFigure 9.** Performance of PSE-MR with different number of SNPs and mediators (causally non-ordered mediators).

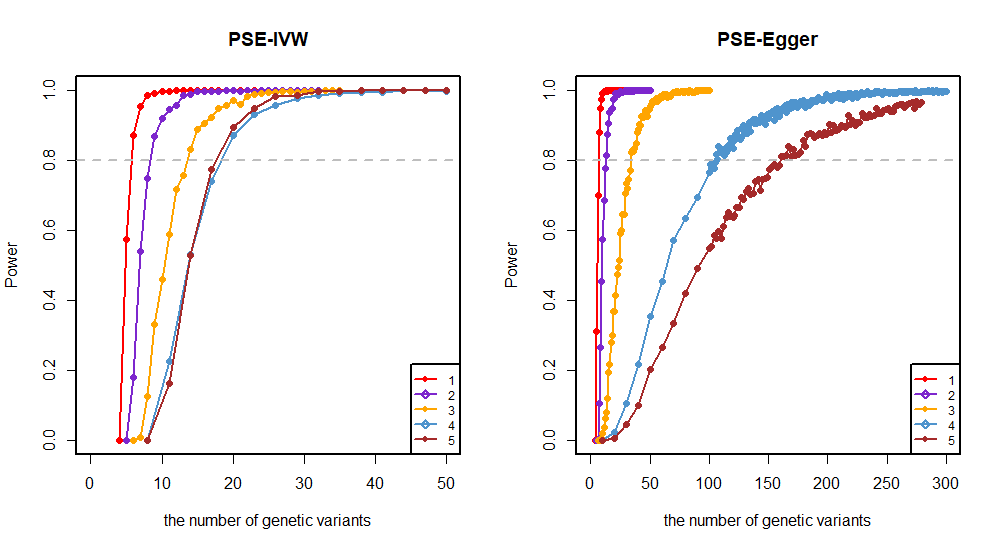

**eFigure 10.** Performance of PSE-MR with different number of SNPs and mediators (causally ordered mediators).

**eTable 41.** Performance of PSE-MR with different number of SNPs and mediators (causally non-ordered mediators) (N=20000).

| The number of instrumental variables | Power of direct effect (PSE-IVW) | Power of direct effect (PSE-Egger) |
| --- | --- | --- |
| One mediator | | |
| 4 | 0.554 | 0 |
| 5 | 0.881 | 0.31 |
| 6 | 0.975 | 0.751 |
| 7 | 0.992 | 0.894 |
| 8 | 0.996 | 0.957 |
| 9 | 0.999 | 0.984 |
| 10 | 0.999 | 0.989 |
| Two mediators | | |
| 7 | 0.704 | 0.198 |
| 8 | 0.858 | 0.419 |
| 9 | 0.95 | 0.561 |
| 10 | 0.973 | 0.698 |
| 11 | 0.992 | 0.788 |
| 12 | 0.993 | 0.84 |
| 13 | 0.996 | 0.907 |
| 15 | 1 | 0.949 |
| 25 | 1 | 1 |
| Three mediators | | |
| 11 | 0.782 | 0.209 |
| 12 | 0.851 | 0.321 |
| 13 | 0.921 | 0.394 |
| 20 | 1 | 0.788 |
| 22 | 1 | 0.822 |
| 25 | 1 | 0.908 |
| 28 | 1 | 0.95 |
| 36 | 1 | 0.99 |
| Four mediators | | |
| 16 | 0.755 | - |
| 17 | 0.831 | - |
| 18 | 0.898 | - |
| 20 | 0.955 | 0.271 |
| 25 | 0.999 | 0.5 |
| 42 | 1 | 0.827 |
| 50 | 1 | 0.9 |
| 80 | 1 | 1 |
| Five mediators | | |
| 22 | 0.797 | - |
| 24 | 0.871 | - |
| 25 | 0.91 | - |
| 27 | 0.953 | - |
| 30 | 0.986 | 0.12 |
| 96 | 1 | 0.785 |
| 97 | 1 | 0.818 |
| 105 | 1 | 0.852 |
| 120 | 1 | 0.906 |
| 142 | 1 | 0.955 |
| 200 | 1 | 0.99 |

**eTable 42.** Performance of PSE-MR with different number of SNPs and mediators (causally ordered mediators).

| The number of instrumental variables | Power of direct effect (PSE-IVW) | Power of direct effect (PSE-Egger) |
| --- | --- | --- |
| One mediator (N=20000) | | |
| 5 | 0.575 | 0.312 |
| 6 | 0.873 | 0.701 |
| 7 | 0.956 | 0.882 |
| 8 | 0.985 | 0.949 |
| 9 | 0.991 | 0.974 |
| 10 | 0.997 | 0.991 |
| 13 | 1 | 1 |
| Two mediators (N=20000) | | |
| 8 | 0.75 | 0.265 |
| 9 | 0.868 | 0.456 |
| 10 | 0.921 | 0.575 |
| 11 | 0.945 | 0.687 |
| 12 | 0.959 | 0.777 |
| 13 | 0.985 | 0.815 |
| 15 | 0.997 | 0.906 |
| 20 | 1 | 0.979 |
| 25 | 1 | 0.998 |
| Three mediators (N=20000) | | |
| 13 | 0.759 | 0.08 |
| 14 | 0.832 | 0.121 |
| 15 | 0.888 | 0.195 |
| 16 | 0.905 | 0.218 |
| 18 | 0.95 | 0.299 |
| 22 | 0.983 | 0.474 |
| 25 | 0.995 | 0.592 |
| 28 | 0.999 | 0.646 |
| 34 | 1 | 0.823 |
| 41 | 1 | 0.903 |
| 50 | 1 | 0.955 |
| 64 | 1 | 0.99 |
| Four mediators (N=30000) | | |
| 17 | 0.74 | - |
| 20 | 0.873 | - |
| 23 | 0.931 | - |
| 26 | 0.959 | - |
| 90 | 1 | 0.694 |
| 106 | 1 | 0.817 |
| 119 | 1 | 0.853 |
| 134 | 1 | 0.908 |
| 163 | 1 | 0.956 |
| 230 | 1 | 0.99 |
| Five mediators (N=70000) | | |
| 17 | 0.774 | - |
| 20 | 0.895 | - |
| 23 | 0.948 | - |
| 35 | 1 | - |
| 140 | 1 | 0.74 |
| 161 | 1 | 0.812 |
| 179 | 1 | 0.859 |
| 201 | 1 | 0.904 |
| 257 | 1 | 0.954 |
| 300 | 1 | 0.99 |
